# Untargeted Screening of Environmental and Endogenous Metabolites and Risk of Incident COPD: A Prospective Study in Three European Populations

**DOI:** 10.64898/2026.03.20.26348905

**Authors:** Max J. Oosterwegel, Anastasia Chrysovalantou Chatziioannou, Nivonirina Robinot, Pekka Keski-Rahkonen, Brooklynn R. McNeil, Randolph R. Singh, Gary W. Miller, Ayoung Jeong, Medea Imboden, Megi Vogli, Regina Pickford, Peter E. Engelfriet, W.M.Monique Verschuren, Annette Peters, Roel Vermeulen, Nicole Probst-Hensch, Jelle Vlaanderen, the EXPANSE consortium

## Abstract

**Objective:** Chronic obstructive pulmonary disease (COPD) is a leading cause of death in the industrialized world. Although smoking, air pollution, and occupational exposures are well established risk factors, the molecular pathways linking environmental exposures and biological susceptibility to COPD remain incompletely understood. Untargeted metabolomics offers a unique opportunity to simultaneously capture internalized environmental chemicals and endogenous metabolic perturbations. However, large prospective studies integrating broad exposomic and metabolic screening prior to COPD onset are lacking.

**Methods:** We conducted a nested case-control study within three European population-based cohorts (Doetinchem Cohort Study, KORA, SAPALDIA) and analyzed 1473 prospectively collected plasma samples. COPD was defined by a pre-bronchodilation FEV1/FVC ratio below 0.7 at follow-up (4–16 years after blood sample collection). We applied complementary untargeted liquid- and gas chromatography high-resolution mass spectrometry (LC- and GC-HRMS), enabling extensive coverage of endogenous metabolism and exogenous environmental contaminants, including pesticides, plastic-related chemicals, and polycyclic aromatic hydrocarbons. Controls maintained normal lung function and were matched to cases on age, sex, follow-up time, and sample collection round. We performed separate conditional logistic regression models for each metabolomic feature, and used Mummichog for prediction of biological pathways involved. The false discovery rate (FDR) was controlled using the Benjamini-Hochberg procedure. Long-term measurement reliability was evaluated using intraclass correlation coefficients (ICCs) from repeat samples in the Doetinchem Cohort Study.

**Results:** In total, thousands of metabolomic features were screened, including 724 annotated exogenous compounds, 13 endogenous metabolites, and 197 features that could be derived as both. Nicotine and cotinine intensity levels were statistically significantly associated with COPD incidence at an FDR of 10%, validating the analytical and epidemiologic framework. Beyond smoking-related markers, lower levels of butyrylcarnitine were associated with increased COPD risk among never-smokers, implicating altered mitochondrial fatty-acid metabolism as a potential early pathway independent of tobacco exposure. Although most screened environmental contaminants, including PAHs and pesticides, were not associated with COPD at stringent significance thresholds, restricting analyses to temporally stable metabolites identified the insecticide metabolite phenyl N-methylcarbamate as a predictor.

**Conclusion:** This large-scale, prospective untargeted metabolomics study represents one of the most comprehensive assessments to date of both environmental and endogenous metabolic predictors of COPD. Our findings demonstrate the feasibility of exposome-wide molecular screening years before disease onset, identify butyrylcarnitine as a novel metabolic predictor in never-smokers, and highlight the importance of accounting for temporal variability in metabolomic epidemiology.

## Introduction

Chronic obstructive pulmonary disease (COPD) is a progressive lung disease with chronic respiratory symptoms and obstructed airflow. It is a leading cause of morbidity and mortality worldwide and imposes a substantial and growing public health burden, particularly in aging populations.^1^ Despite its clinical importance, COPD is typically diagnosed late in its course, after irreversible lung damage has occurred. As a result, identifying early biological and environmental determinants of disease onset remains a critical priority for prevention.

While smoking, air pollution, and occupational exposures are well-established contributors^2^, these factors do not fully explain disease occurrence, and other environmental determinants are likely involved but remain poorly characterized. For example, a scoping review identified associations between COPD and pollutants including insecticides, herbicides, and polycyclic aromatic hydrocarbons (PAHs).^3^ As smoking prevalence and air pollution levels decline in many high-income countries, these additional exposures may account for an increasing share of incident COPD. However, identifying such exposures and understanding their biological consequences has been challenging, in part because traditional exposure assessment methods rely on external measurements or self-report and do not capture the internal biological response. Approaches that simultaneously characterize internalized environmental exposures and their metabolic effects could provide critical insights into early disease mechanisms and inform preventive strategies.

Untargeted metabolomics offers a powerful approach to address this gap by enabling simultaneous measurement of thousands of small molecules in biological samples, including both exogenous environmental chemicals and endogenous metabolites reflecting host physiological responses. Through combining high-resolution mass spectrometry platforms such as liquid chromatography (LC-HRMS) and gas chromatography (GC-HRMS), metabolomic profiling can capture a broad spectrum of the internal exposome and provide insights into mechanisms linking environmental exposures to disease onset. However, large prospective studies integrating comprehensive exogenous and endogenous metabolomic profiling in relation to incident COPD remain scarce.

In this study, we conducted a nested case-control analysis within three population-based European cohorts (Doetinchem Cohort Study, KORA, and SAPALDIA) and analyzed 1,473 prospectively collected plasma samples from 747 cases and 726 matched controls, using complementary untargeted LC- and GC-HRMS platforms. Our aim was to identify metabolomic predictors of incident COPD and to characterize the reliability of circulating metabolomic markers in prospective epidemiologic studies.

## Methods

### Study population and design

This case-control study was nested in three prospective European cohorts (Doetinchem Cohort Study, KORA, SAPALDIA), which have been previously described.^4–7^ Briefly, all cohorts are population-based cohorts that collected blood samples and spirometry measurements repeatedly during baseline and follow-up. In these cohorts, participants were originally recruited between 1987–91 (SAPALDIA, Doetinchem Cohort Study) and 1999-2001 (KORA S4).

COPD diagnosis was defined according to the GOLD criteria with a pre-bronchodilation FEV1/FVC below 0.7. Cases were then defined as those participants that were free of COPD during sample collection (baseline) but developed COPD (i.e. FEV1/FVC below 0.7) during follow-up (1-20 years after sample collection). Cases were included if they had EDTA plasma available, and were between 25 and 80 years old at the time of diagnosis.

Controls were required to be free of COPD during both sample collection and follow-up. The cases were 1:1 matched to controls on age (+-5 years), sex, follow-up time, and sample collection round. When possible, the cohorts additionally matched on season of blood collection, time of day sample was collected (morning, afternoon, evening), and fasting state (fasting/non-fasting). Supplementary Methods I describes the matching variables per cohort.

To assess temporal stability of metabolomic features, a subset of participants (n = 295: 148 cases, 147 controls) from the Doetinchem Cohort Study provided repeat blood samples collected approximately five years earlier.

The nested case-control study was approved by the Ethics Committee of International Agency for Research on Cancer (IARC).

### Metabolomic analyses

Detailed protocols for sample preparation and metabolomic analyses are provided in Supplementary Methods II, with key procedural steps summarized below.

#### Batch design and quality control

To avoid bias by technical variation, matched pairs were randomly allocated across plates while the order of case and control within a pair was randomized (block randomization). All samples (including repeats) from a subject were considered part of the same block. Case–control pairs from each cohort were distributed evenly across plates and batches, such that each plate contained the same proportion of pairs from each cohort.

For the LC analysis, quality control (QC) samples were prepared from a sample pool that was made by mixing 50 μL aliquots of samples of the Moli-Sani cohort (another EXPANSE cohort) and extracted along the study samples. Blank samples, which contained all reagents except for plasma, were prepared and extracted in the same manner as the study samples. Each 96-well plate included four individually prepared QCs and two blanks. A QC sample was injected every ca. 20 samples, with a repetition of each QC taking place after ca. 10 injections. In total, all samples were analyzed in four analytical batches, each consisting of five 96-well plates.

For the GC analysis, QC samples were prepared along with the study samples which consisted of blanks, two age defined pooled plasma samples and a standard reference material (SRM). A blank and pooled sample were injected every 20 samples. In total, all samples were processed in 22 batches of 96-well plates – each containing up to 82 study samples and up to 14 QAQC samples – using OT-2 liquid handling robot (Opentrons Labworks Inc.).

#### LC-HRMS analysis and data pre-processing

An UHPLC-QTOF-MS system consisted of a 1290 Binary LC system, and a 6550 QTOF mass spectrometer with Jet Stream electrospray ionization source (Agilent Technologies).

Pre-processing was performed using Profinder 10.0.2 and Mass Profiler Professional B.14.9.1 software (Agilent Technologies). A “Batch Recursive Feature Extraction (small molecules/peptides)” process was employed to find [M+H]^+^ ions utilizing the study samples and the blanks of each batch. Mass peak height threshold of 1500 counts and chromatographic peaks threshold of 8000 counts were used, while the minimum quality score was 70.

Targeted feature extraction from all study samples and QCs of each batch was then performed in Profinder in a “Batch targeted feature extraction” process using a match tolerance of ±10 ppm for masses and ±0.03 min for retention times. Ion species were limited to [M+H]^+^ and the integrator was Agile 2 with no smoothing applied to the chromatograms. Peak areas were used as a measure of feature intensity.

To avoid redundancy caused by multiple features corresponding to the same compound (e.g., proton and sodium adducts or fragments), a clustering-based method was applied to identify highly correlated features (Pearson correlation coefficient of intensities higher than 0.85) with similar retention times (±0.05 min). Within each cluster, the feature with the highest average intensity and least nondetects was retained for subsequent statistical analyses as the cluster representative.

#### GC-HRMS analysis and data pre-processing

Data acquisition was performed using a Trace 1310 gas chromatography system coupled to an Exploris GC 240-HRMS system (Thermo Fisher Scientific, Rockford, IL, USA).

Features were extracted using XCMS^8^ 4.4.0 using an area threshold of 10,000, filtered by at least 10% presence and clustered into spectra and abundances using RAMClustR^9^ 1.3.1 with correlation parameters set to 8 and 1 for retention time and abundance, respectively. After clustering the features, RAMClustR uses a weighted mean function to combine the features of a cluster into an aggregated signal where the more abundant features contribute more to the spectral intensity of the cluster.

### Metabolomic annotation

#### LC-HRMS

To provide annotations of known metabolites, a compound library was created using Agilent MassHunter PCDL Manager B.08.00 software, by compiling information from the following sources: (1) Past published studies performed by the IARC laboratory, using the same analytical method and instrumentation (2) In-house list of analyzed chemical standards (3) List of lipids found through lipidomics profiling of two random study samples (case and control) subjected to data-dependent fragmentation with 10, 20 and 40 V collision energies. The lipidomics analysis was performed using Lipid Annotator 1.0 software (Agilent). Following the creation of the compound library, data from a representative QC sample was used to confirm and adjust the retention times of the current study. The library was then used in Agilent IDBrowser B.08.00 module of the Mass Profiler Professional 14.9.1 software to annotate the extracted features using matching tolerances of ±10 ppm and ±0.3 min. The matching considered singly charged [M+H]^+^ ions. Metabolites known to be detected with different ions were included in the library in a format allowing to be detected using the described method. Additionally, the entries of PubChemLite v.1.39 were filtered for the presence of Br and Cl in their chemical formula. The 25,874 brominated and the 54,572 chlorinated entries populated two PCDL libraries, which were matched with IDBrowser to the feature table using only the requirement of matching the accurate mass (±10 ppm). The matching features were further filtered based on their isotopic pattern, using three requirements: a) at least 3 isotope peaks detected, b) M+2 isotope peak with greater intensity than M+1 and c) IDBrowser peak score of 76 %, when the contribution to overall score in IDBrowser were: Mass score 100.00, Isotope abundance score 60.00, Isotope spacing score 50.00.

All features indicated by the statistical analysis as significantly deviating between the COPD and the control group (previously annotated or unannotated) were confirmed by reanalysis of the sample with the highest intensity of the corresponding feature together with the analytical standard, allowing the confirmation of the exact retention time and shape of the peak. MS/MS spectra were also acquired for a representative sample and the analytical standard, allowing the confirmation of the identification of the metabolite to Level 1 according to the Metabolomics Standards Initiative (MSI).^10^

#### GC-HRMS

Spectral matching was performed with MetabonAnnotatoR^11^ and followed the criteria suggested by the Koelmel scale^12^. For initial level 1 annotations, experimental spectra were matched to an in-house library of 2206 compounds using a 5 ppm fragment matching tolerance, reverse match score of 0.6, and delta retention index of 50. For initial level 2 annotations, experimental spectra were matched to 3 external databases: Thermo GC-Orbitrap libraries (756 compounds – high resolution), RECETOX Public Library (384 compounds – high resolution)^13^, and the Wiley Registry/Nist Mass Spectral Library (533,125 compounds – unit resolution). Unit resolution spectral matching was done with a mass tolerance of 0.75 amu, reverse matching score of 0.7 and delta retention index of 50.

### Statistical methods

#### Partitioning of features into sets

After the tentative annotation of the LC-HRMS and the initial annotation of the GC-HRMS data, we divided the features from both platforms into two tiers to guide the statistical analysis. Only features detected in at least 10% of the samples were included in this process. The first tier contained features that were tentatively annotated based on library matching (LC-HRMS) or annotated with confidence levels 1 and 2 (GC-HRMS)^12^, while the second tier contained the features that could not be identified initially. The identified features (tier A) were further partitioned into exogenous, endogenous, and compounds that could be both exogenously and endogenously derived. Lastly, all resulting partitions were divided into a set ‘a’ and ‘b’. Set ‘a’ contained features that were detected in at least 40% of the samples while set ‘b’ contained the remaining features. Features from set ‘a’ were imputed (see ‘Imputation and batch effect adjustment’) and treated as continuous variable while features from set ‘b’ were converted to a binary indicator of detected versus not detected. In Figure S1 this partitioning of the LC- and GC-HRMS data into sets is visually explained. Moreover, a global overview of the study design and conducted analyses is shown in Figure 1.

**Figure 1:**
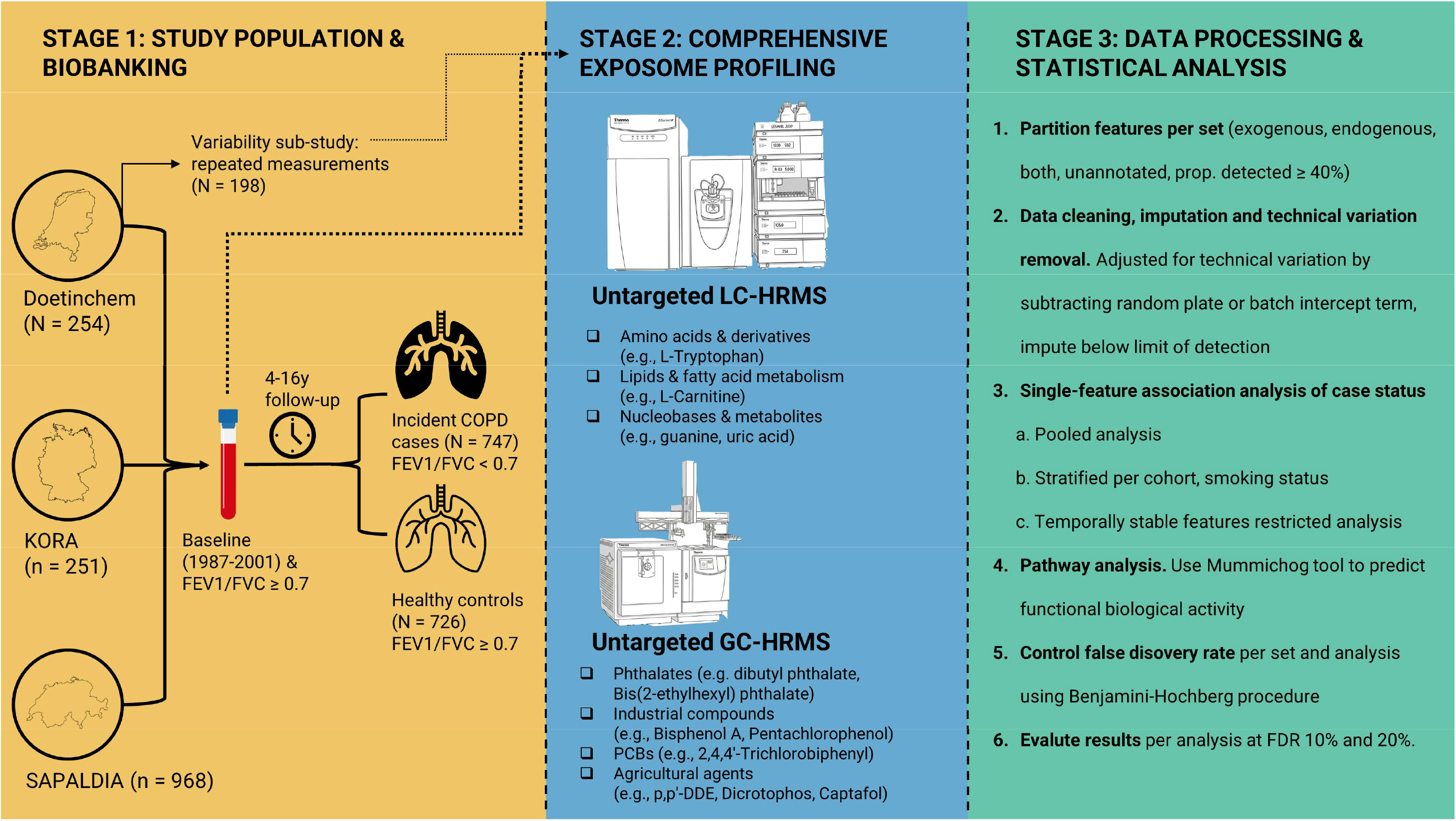
global overview of study design and analyses. LC-HRMS = liquid chromatography high resolution mass spectrometry; GC-HRMS = gas chromatography high resolution mass spectrometry; COPD = chronic obstructive pulmonary disease; FEV1 = forced expiratory volume in 1 second; FVC = forced vital capacity; y = years; N = tally.

Each of the features was analyzed in a single feature at a time approach by regressing COPD on the feature in a conditional logistic regression model. For results from features in set ‘A’, Rubin’s rules were used to combine coefficients from each imputed version of a feature into a single estimate for a feature. The false discovery rate (FDR) was controlled at 10% per set using the Benjamini-Hochberg procedure.

#### Imputation and batch effect adjustment

We assumed all features with nondetects were left-censored at the limit of detection (defined as the minimum detected value on that batch/plate) and nondetects were not due to interfering analytes. To adjust for technical variation and impute values when nondetects were present, we built a statistical model of each feature that models the batch effect and left-censoring. Details on this model are available in Supplementary Methods III. In brief, we fitted a multilevel, Bayesian version of the left-censored imputation procedure described by Lubin et al. (2004) that also models the technical variation.^14^ In other words, we regressed each of the (natural log-transformed) features on a random effect for plate and fixed effects for COPD, the matching variables, and smoking to ensure congeniality. After fitting the model, we imputed values for the nondetects by drawing a random value (using the parameters of the model we fitted) between minus infinity and (the batch adjusted) limit of detection (natural logarithmic scale). Lastly, each feature was adjusted for possible technical variation by subtracting the random plate (or batch for GC) intercept term from the observed (or imputed) value.

To represent the inherent uncertainty on the exact value of the nondetects, 60 imputations were drawn from the posterior of the imputed estimates of the model.

### Variability calculation

We estimated the variability of a single measurement over approximately 5 years using the repeated samples from the Doetinchem Cohort Study. Specifically, we calculated an intraclass correlation coefficient (ICC) for each feature using the methodology from Oosterwegel et al.^15^ (Supplementary Methods IV).

### Stratified analysis

In addition to the main analysis, we conducted a stratified analysis by smoking status (never, former, current). While the primary analysis employed conditional logistic regression, the smoking-stratified analysis used Firth’s logistic regression to account for data sparsity. This approach was necessary because stratification by smoking status often resulted in matched pairs with only one member (e.g., a control in the never-smoker group whose matched case was a smoker). Such incomplete pairs cannot contribute information to conditional logistic regression, as no discordance is possible. Therefore, in the smoking-stratified analysis, we broke the matching and instead modeled case status as a function of sex, age, diagnosis round, and metabolite intensity, using Firth’s correction to reduce potential sparse data bias.

#### Pathway analyses

To aid in interpreting the unidentified features, we performed a pathway analysis on all results from the LC-HRMS platform (both identified and unidentified) for the pooled and smoking stratified analysis. This was done with the mummichog tool (via MetaboAnalyst^16^). Mummichog predicts functional activity directly from the feature’s accurate mass without a certain identification of the features.^17^ The p-value of a feature’s relationship with case status, the feature’s *m/z* value and retention time were used as input. Parameters of mummichog were set to a mass tolerance of 10 ppm, a retention time window of 6 seconds, and [M+Na]^+^ and [M+H]^+^ adducts. The features that were nominally statistically significant (i.e. p-value smaller than 0.05) were considered enriched, whereas the other features were considered as background. Subsequently, we used Fisher’s exact test to analyze pathway enrichment, considering only pathways with at least four matches. We controlled the false discovery rate at 10% using the Benjamini-Hochberg procedure.

### Sensitivity analysis

To assess differences in signals across cohort we performed an analysis stratified by cohort (Doetinchem Cohort Study, SAPALDIA, KORA).

## Results

### Study population

The final nested case-control study included 1,473 plasma samples, consisting of 747 cases and 726 controls. The unequal group sizes are the result of post-hoc sample exclusions based on quality control criteria, which are detailed in Supplementary Methods I. Baseline characteristics of the participants are shown in Table 1. In brief, most participants were from the SAPALDIA study (n = 968), followed by KORA (n = 251) and Doetinchem Cohort Study (n = 254). The average age was 52 (IQR 13).

**Table 1:** population characteristics of cases and controls with pre-diagnostic blood samples included in our analyses, per cohort. n (%); Median [IQR] (Min-Max).

|  | **Overall** (N = 1473) |  | **Doetinchem Cohort Study** (N = 254) |  | **KORA** (N = 251) |  | **SAPALDIA** (N = 968) |  |
|---|---|---|---|---|---|---|---|---|
| **Characteristic** | **Case**<br>(N = 747)<sup>*1*</sup> | **Control**<br>(N = 726)<sup>*1*</sup> | **Case**<br>(N = 139)<sup>*1*</sup> | **Control**<br>(N = 115)<sup>*1*</sup> | **Case**<br>(N = 124)<sup>*1*</sup> | **Control**<br>(N = 127)<sup>*1*</sup> | **Case**<br>(N = 484)<sup>*1*</sup> | **Control**<br>(N = 484)<sup>*1*</sup> |
| Sex |  |  |  |  |  |  |  |  |
| female | 351 (47%) | 340 (47%) | 74 (53%) | 63 (55%) | 50 (40%) | 50 (39%) | 227 (47%) | 227 (47%) |
| male | 396 (53%) | 386 (53%) | 65 (47%) | 52 (45%) | 74 (60%) | 77 (61%) | 257 (53%) | 257 (53%) |
| Age | 52.2 [13.0]<br>(29.3-71.6) | 50.0 [13.0]<br>(30.0-72.2) | 52.4 [11.1]<br>(36.9-65.2) | 52.0 [10.5]<br>(36.6-66.6) | 46.0 [8.0]<br>(35.0-55.0) | 46.0 [9.0]<br>(34.0-55.0) | 55.0 [13.4]<br>(29.3-71.6) | 52.2 [14.1]<br>(30.0-72.2) |
| Smoking |  |  |  |  |  |  |  |  |
| never | 267 (36%) | 295 (41%) | 34 (24%) | 47 (41%) | 40 (32%) | 46 (36%) | 193 (40%) | 202 (42%) |
| current | 228 (31%) | 156 (21%) | 55 (40%) | 16 (14%) | 54 (44%) | 22 (17%) | 119 (25%) | 118 (24%) |
| ex | 252 (34%) | 275 (38%) | 50 (36%) | 52 (45%) | 30 (24%) | 59 (46%) | 172 (36%) | 164 (34%) |
| BMI | 25.2 [5.2]<br>(16.5-44.8) | 26.1 [5.3]<br>(17.0-55.1) | 25.3 [4.3]<br>(16.5-37.2) | 26.2 [4.5]<br>(19.3-42.0) | 26.6 [6.0]<br>(19.8-42.6) | 26.9 [6.0]<br>(19.1-55.1) | 24.8 [5.1]<br>(17.5-44.8) | 25.7 [5.0]<br>(17.0-42.7) |
| Diabetes | 16 (2.1%) | 14 (1.9%) | 3 (2.2%) | 2 (1.7%) | 1 (0.8%) | 1 (0.8%) | 12 (2.5%) | 11 (2.3%) |
| Unknown | 2 | 2 | 1 | 0 | 1 | 1 | 0 | 1 |
| Asthma | 57 (7.7%) | 51 (7.0%) | 5 (3.6%) | 4 (3.5%) | 6 (5.0%) | 4 (3.2%) | 46 (9.5%) | 43 (8.9%) |
| Unknown | 3 | 2 |  |  | 3 | 2 |  |  |
| Wheeze | 106 (14%) | 68 (9.4%) | 14 (10%) | 11 (9.6%) | 19 (16%) | 9 (7.2%) | 73 (15%) | 48 (9.9%) |
| Unknown | 3 | 2 |  |  | 3 | 2 |  |  |
| Wheeze without cold | 66 (8.9%) | 39 (5.4%) | 10 (7.2%) | 8 (7.0%) | 11 (9.1%) | 4 (3.2%) | 45 (9.3%) | 27 (5.6%) |
| Unknown | 4 | 3 | 0 | 1 | 3 | 2 | 1 | 0 |
| Stroke | 4 (0.5%) | 8 (1.1%) | 1 (0.7%) | 2 (1.7%) | 0 (0%) | 1 (0.8%) | 3 (0.6%) | 5 (1.0%) |
| Unknown | 1 | 2 |  |  | 1 | 1 | 0 | 1 |
| Years between baseline and follow-up | 8 [5] (4-16) | 8 [5] (4-16) | 5 [0] (4-6) | 5 [0] (4-6) | 13 [4] (9-14) | 13 [3] (9-15) | 8 [1] (7-16) | 8 [1] (7-16) |
| FEV1 at baseline | 3.14 [1.01]<br>(1.22-5.69) | 3.37 [1.08]<br>(1.11-6.08) | 3.15 [0.86]<br>(1.98-5.50) | 3.40 [0.91]<br>(1.93-5.22) | 3.21 [1.04]<br>(1.70-4.99) | 3.55 [1.03]<br>(1.11-5.86) | 3.12 [1.04]<br>(1.22-5.69) | 3.32 [1.08]<br>(1.89-6.08) |
| FEV1 at follow-up | 2.73 [0.92]<br>(0.96-5.16) | 3.10 [1.10]<br>(1.32-5.80) | 2.92 [0.79]<br>(1.48-5.10) | 3.25 [1.03]<br>(1.68-5.10) | 2.75 [0.94]<br>(1.38-4.83) | 3.34 [1.05]<br>(1.32-5.80) | 2.60 [0.91]<br>(0.96-5.16) | 3.00 [1.04]<br>(1.41-5.63) |
| FVC at baseline | 4.27 [1.41]<br>(1.70-8.08) | 4.33 [1.39]<br>(1.37-7.63) | 4.31 [1.19]<br>(2.80-7.38) | 4.34 [1.32]<br>(2.36-6.37) | 4.32 [1.36]<br>(2.26-6.93) | 4.49 [1.31]<br>(1.37-7.63) | 4.24 [1.46]<br>(1.70-8.08) | 4.30 [1.42]<br>(2.14-7.57) |
| FVC at follow-up | 4.02 [1.32]<br>(1.72-7.40) | 4.06 [1.39]<br>(1.63-7.72) | 4.34 [1.15]<br>(2.34-7.32) | 4.19 [1.37]<br>(2.14-6.26) | 4.20 [1.38]<br>(2.11-7.27) | 4.28 [1.16]<br>(1.63-7.72) | 3.86 [1.29]<br>(1.72-7.40) | 3.97 [1.38]<br>(1.73-7.06) |
| FEV1/FVC ratio at baseline | 0.73 [0.03]<br>(0.70-0.87) | 0.78 [0.06]<br>(0.70-0.99) | 0.72 [0.03]<br>(0.70-0.86) | 0.79 [0.05]<br>(0.71-0.89) | 0.73 [0.03]<br>(0.70-0.82) | 0.79 [0.04]<br>(0.71-0.87) | 0.73 [0.04]<br>(0.70-0.87) | 0.77 [0.06]<br>(0.70-0.99) |
| FEV1/FVC ratio at follow-up | 0.67 [0.02]<br>(0.48-0.69) | 0.76 [0.06]<br>(0.70-0.93) | 0.67 [0.02]<br>(0.48-0.69) | 0.78 [0.05]<br>(0.70-0.88) | 0.68 [0.02]<br>(0.56-0.69) | 0.78 [0.06]<br>(0.70-0.90) | 0.67 [0.02]<br>(0.51-0.69) | 0.75 [0.05]<br>(0.70-0.93) |

<sup>*1*</sup>n (%); Median [IQR] (Min-Max)

COPD diagnosis occurred on average 8 years after blood collection, ranging from 5 years in Doetinchem to 13 years in KORA. At baseline, cases had lower mean FEV₁/FVC ratios than controls (0.73 vs 0.78), and this difference widened during follow-up (0.67 vs 0.76), reflecting both lower initial lung function and faster decline among cases (Figure S2).

Comorbidities were uncommon. Smoking prevalence varied substantially across cohorts: in KORA, 44% of cases and 17% of controls were current smokers, compared with 40% and 14% in Doetinchem and 25% and 24% in SAPALDIA, respectively. A similar variation in asthma prevalence could be observed with SAPALDIA having a notably higher asthma prevalence of circa 9% (cases 9.5%, controls 8.9%) vs 4% for the other cohorts (Doetinchem Cohort Study: cases 3.6%, controls 3.5%; KORA: cases 5%, controls 3.2%).

### Assessment of blood metabolome

We acquired metabolomic data based on 1,473 plasma samples with LC-HRMS methodology and 1,436 plasma samples with GC-HRMS based methodology due to insufficient sample volume in 37 samples.

LC-HRMS detected 4,679 features grouped into 2,580 retention-time clusters, of which 219 were annotated (12 endogenous, 41 exogenous, 166 mixed origin). Set ‘a’ (features detected in ≥40% of samples) contained 3904 features (193 annotated) while set ‘b’ (features detected in <40% of samples) contained 498 features (12 annotated). Examples of compounds detected using the LC platform include amino acids and their derivatives (e.g., L-tryptophan, L-phenylalanine), a broad spectrum of lipids and molecules related to fatty acid metabolism (e.g., L-carnitine), and key nucleobases and their metabolites (e.g., guanine, uric acid). See Table S4 for a list of the precise endogenous, exogenous classifications per compound.

GC-HRMS detected 73,710 features, which clustered into 5,923 distinct chemical entities, including 715 annotated compounds (114 level 1, 601 level 2). 684 of these clusters belonged to set ‘a’, while 31 belonged to set ‘b’. 683 of the annotations were classified as exogenous compounds, while 31 were categorized as both endogenously and exogenously and 1 as endogenous. Examples of compounds detected using the GC platform include major classes of environmental contaminants, such as phthalates (e.g. dibutyl phthalate, Bis(2-ethylhexyl) phthalate), industrial compounds and their byproducts like phenols (e.g., Bisphenol A, Pentachlorophenol), persistent organic pollutants such as polychlorinated biphenyls (PCBs) (e.g., 2,4,4’-Trichlorobiphenyl, 2,2’,4,4’,5,5’-Hexachlorobiphenyl), and various agricultural agents, including pesticides, herbicides, and fungicides (e.g., p,p’-DDE, Dicrotophos, Captafol).

### Associations of single metabolites with COPD incidence

In the pooled analysis, (S)-nicotine and cotinine were FDR significant (Figure 2). The feature corresponding to cotinine shows a minor interference from serotonin, which was considered negligible in this study, as presented in Supporting Information I. An unidentified feature (monoisotopic mass 192.0892, retention time 0.92 minutes; 192.0892@0.92 hereafter) was also significant and strongly correlated with smoking behavior (Table S2). In a separate multivariable model that included all three (single cotinine feature) features and self-reported smoking status, only nicotine remained statistically significantly associated with COPD; smoking status itself was no longer statistically significant. However, when pack-years was used as the smoking metric, both nicotine and pack-years were independently associated with COPD (Table S1a, S1b).

**Figure 2:**
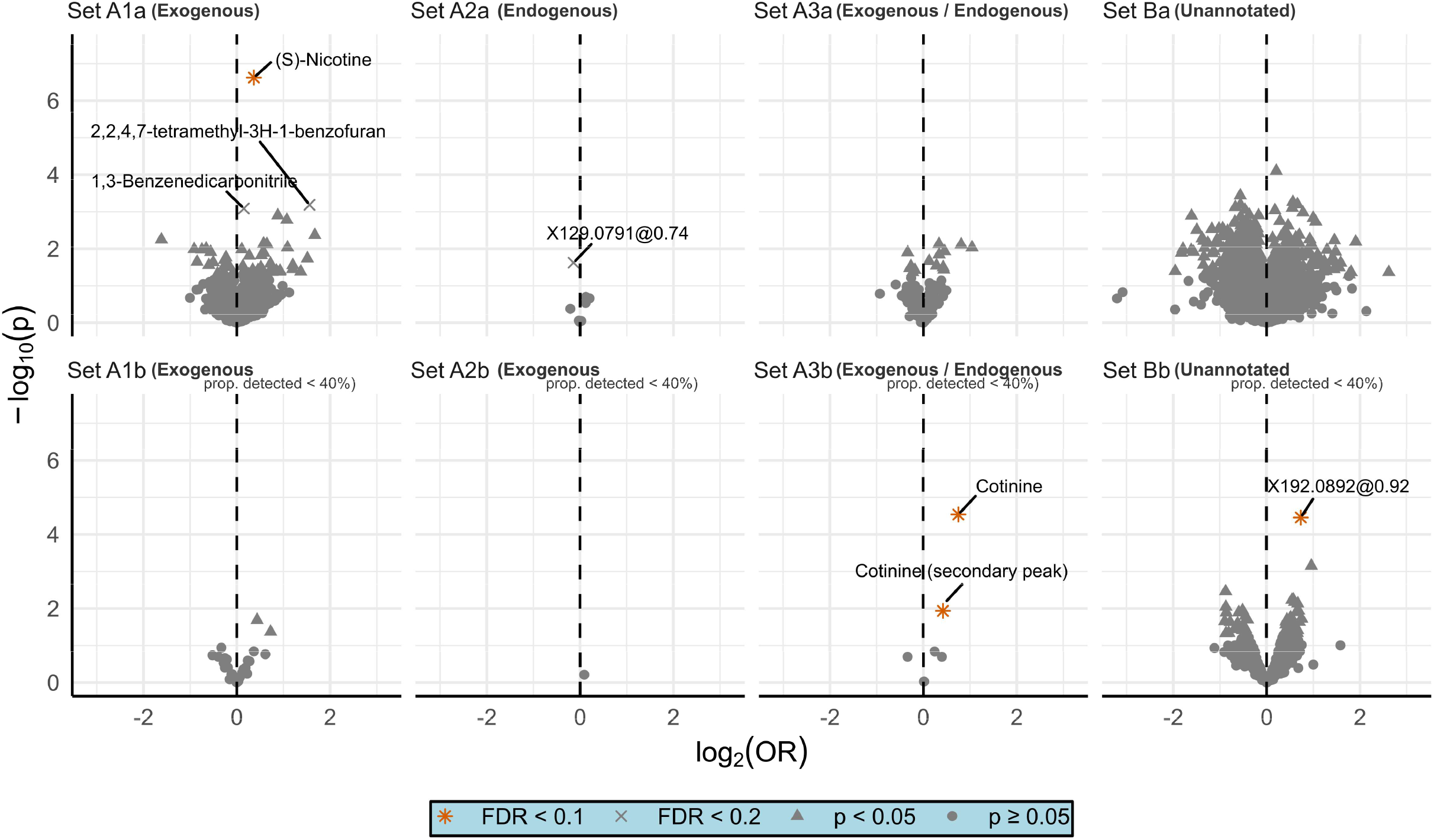
volcano plot of the results from the pooled analysis. Set A describes the annotated metabolites, Set B indicates the initially unidentified features/clusters. 1, 2, 3 describes the exogenous, endogenous, both endogenous/exogenously derived metabolites respectively. The last letter indicates if the feature was analyzed as continuous variable (lower case ‘a’) or as binary variable of detected, not detected (lower case ‘b’). 129.0791@0.74 is in set A because it was tentatively annotated but could not be validated in the end. Cotinine is in the endogenous/exogenously derived set because initially it was thought to be a combination of cotinine and serotonin. p = p-value; FDR = false discovery rate; OR = odds ratio.

At a more lenient FDR threshold of 20%, additional smoking-related compounds were associated with COPD, including 1,3-benzenedicarbonitrile and 2,2,4,7-tetramethyl-3H-1-benzofuran (Table S2, S3).

#### Smoking-stratified analysis

When we stratified the data by smoking behavior (Figure 3, S3, Table S5 for initially annotated compounds), (S)-nicotine was FDR significant in current smokers (current: OR = 1.4, p < 0.0001, FDR = 0.03; ex: OR =1.2, p = 0.37, FDR = 0.97; never: OR = 1.17, p = 0.47, FDR = 0.97), while ACar 4:0 (from the continuous both endogenous/exogenously derived set), also known as butyrylcarnitine, was FDR significant in never-smokers (never: OR = 0.72, p = <0.0005, FDR = 0.07; ex: OR =1.10, p = 0.34, FDR = 0.99; current: OR = 0.95, p = 0.66, FDR = 0.96).

**Figure 3:**
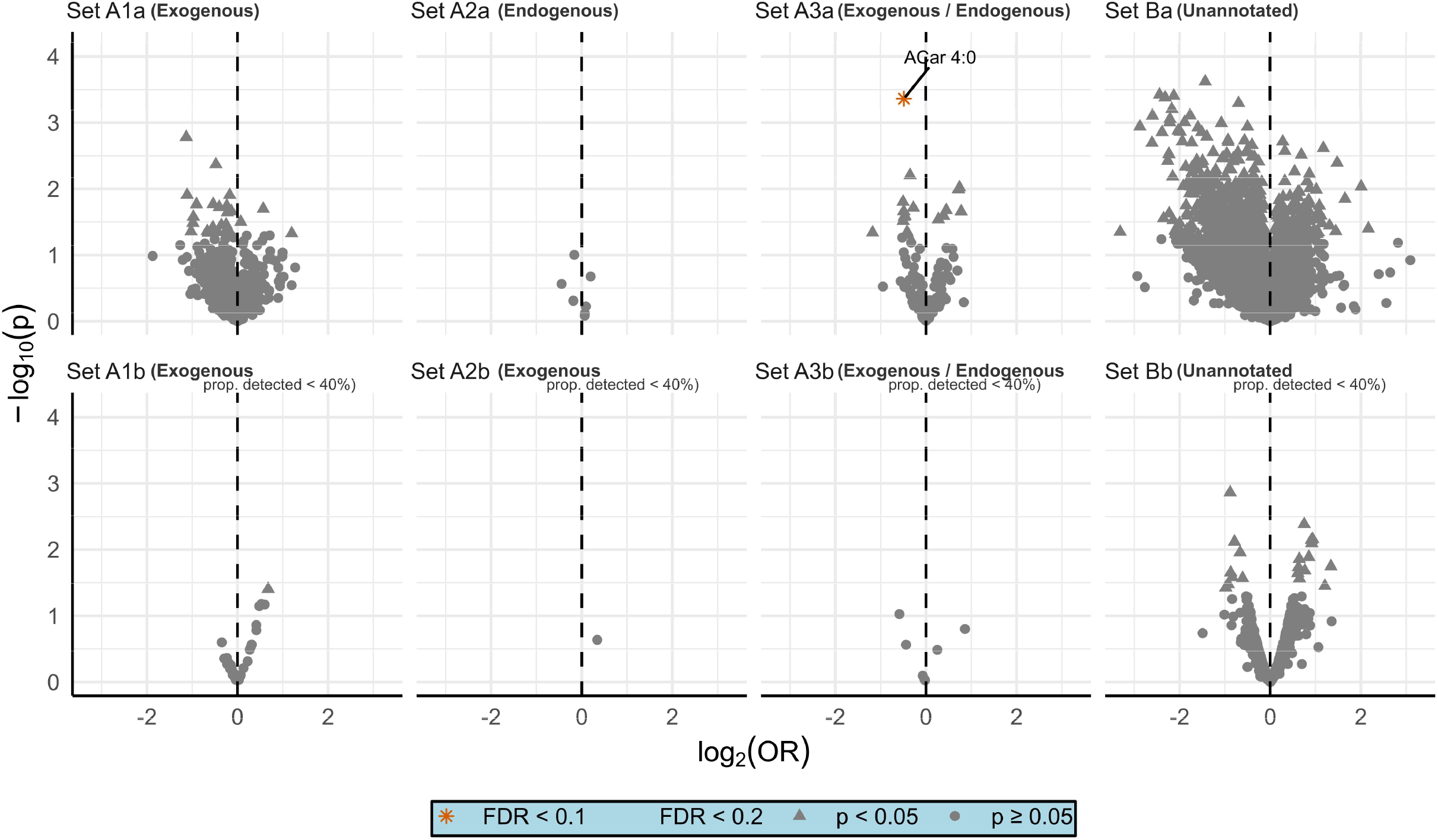
volcano plot of the results from the analysis stratified by smoking status (never). Set A describes the annotated metabolites, Set B indicates the initially unidentified features/clusters. 1, 2, 3 describes the exogenous, endogenous, both endogenous/exogenously derived metabolites respectively. The last letter indicates if the feature was analyzed as continuous variable (lower case ‘a’) or as binary variable of detected, not detected (lower case ‘b’). p = p-value; FDR = false discovery rate; OR = odds ratio; ACar 4:0 = butyrylcarnitine.

At FDR < 0.2, two additional unannotated peaks were predictive of COPD in ex-smokers.

#### Cohort-stratified analysis

In the analysis stratified by cohort (Figure S4, Table S6 for initially annotated compounds), we found no FDR significant compounds in the SAPALDIA cohort, while (S)-nicotine, 192.0892@0.92, and cotinine were FDR significant in the KORA and Doetinchem Cohort Study.

Several cohort-specific associations were observed, particularly in the Doetinchem Cohort Study: LysoPC (20:3) (OR 0.1), LysoPC (20:4) (OR 1.9), 2-methyl-5-phenyl-1H-pyrrole (OR 8.3, level 2 GC-annotation), 1,3-benzenedicarbonitrile (OR 1.6), and the 7-isopropyl-3,4-dihydro-2H-cyclohepta[b]pyrazine (OR 2.4, level 2 GC-annotation) compound were FDR significant. 1,3-benzenedicarbonitrile (OR: 1.2) and 2-methyl-5-phenyl-1H-pyrrole (OR: 4.3) were also nominally significant in the KORA cohort. These differences in statistically significant metabolites are also described in Figure S5. All these compounds except the lipids were strongly related to smoking behavior (Table S2).

At an FDR of 20%, ACar 10:1 (decanoylcarnitine), and 145.0839@0.83 were statistically significant in the SAPALDIA cohort, while 4-chloro-2-(6-chlorohex-1-yn-1-yl)pyridine, 2-[(3R)-3,7-dimethyloct-6-enylidene]malononitrile, and 4-Thiazolidinecarboxylic acid (Thioproline) were statistically significant in the Doetinchem Cohort Study. In KORA, LysoPC (20:3), cortisone, and glycochenodeoxycholic acid were related to COPD at an FDR of 20%. Four of these cohort-specific markers were very strongly related to smoking behavior (Table S3).

#### Exogenous compounds previously associated with COPD

Given the environmental focus of the GC-HRMS platform, we specifically examined previously implicated compound classes. None of the detected PAHs (i.e. pyrene, phenanthrene, and fluorene) were nominally associated with COPD, either in the pooled analysis or in any cohort-specific analysis. Among the herbicides and pesticides (i.e. molinate, p,p’-DDE, bromoxynil, carbaryl, phenyl N-methylcarbamate), phenyl N-methylcarbamate showed a nominally significant positive association with COPD in the pooled analysis (OR = 2.1, p < 0.05, FDR = 0.23). p,p’-DDE was nominally significant in the KORA cohort (OR = 1.39, p < 0.05, FDR = 0.95) but was not replicated in the other cohorts or in the pooled analysis. None of these associations survived FDR correction.

### Pathway analysis

Pathway enrichment analysis identified the fatty acid activation pathway as significantly enriched in pooled analyses (Table S7). This pathway stemmed from 9 putatively annotated compounds from the Mummichog program (Table S7c), six of which belong to the fatty acid activation pathway. Through follow-up validation, we elevated two of these (arachidonic, and docosahexaenoic acid) to MSI level 1 annotations, while one other was annotated as Linoleic acid (instead of Linoelaidic acid).

### Variability single measurement over 5 years

Long-term repeatability was modest, with median ICCs of 0.20 for LC features and 0.26 for GC features. Notably, the repeatability across the clusters from the GC platform was greater than for its features, with a median ICC of 0.34 vs 0.26, while the median ICC for the clusters from the LC platform was similar to the repeatability for its features (0.18 vs 0.20). For both platforms, annotated features or clusters had higher ICCs than unidentified peaks. Figure S6a and S6b shows the full distribution of ICC values over 5 years per platform and annotation status for the features and clusters.

Among annotated peaks and across the platforms, repeatability of endogenous compounds was 0.33 (n = 11), while the repeatability of the exogenous compounds was 0.37 (n = 692) on average. Compounds classified as both had a median ICC of 0.36. Exogenous compounds from the LC platform had a lower average ICC than those from the GC platform (0.28 vs 0.37) despite the ICC calculation adjusting away technical variation. See Figure S7 for more detailed ICC values of the annotated compounds per exogenous, endogenous class.

When we restricted our analysis to more temporally stable features of our data (i.e. ICC ≥ 0.5), four metabolites besides nicotine (ICC = 0.58) and cotinine (secondary peak) (ICC = 0.76) of the 900 remaining features (804 GC, 96 LC) were FDR significant. Namely, 1,3 benzenedicarbonitrile, 2,2,4,7-tetramethyl-3H-1-benzofuran (GC-level 2), phenyl N-methylcarbamate (GC-level 2), and one unidentified compound. A scatter plot that compares the FDR significance of metabolites in both the ICC restricted and pooled analysis is shown in Figure S8.

A comprehensive list of all statistically significant features across all analyses, including their respective identification confidence levels, is provided in Table S8.

## Discussion

In this large prospective metabolomics study integrating broad screening of both endogenous metabolites and exogenous environmental contaminants, we identified several molecular predictors of incident COPD years before diagnosis. Tobacco-related metabolites, including nicotine and cotinine, were robustly associated with COPD incidence, validating both the analytical platform and epidemiologic design. Beyond smoking-related markers, we identified lower plasma butyrylcarnitine as a predictor of COPD among never-smokers, implicating non-smoking-related metabolic pathways in disease development. In addition, analyses restricted to temporally stable metabolites identified the insecticide metabolite phenyl N-methylcarbamate as a potential predictor.

The strong associations observed for nicotine and cotinine are consistent with the established causal role of tobacco smoking in COPD and serve as an important internal validation of our untargeted exposomic approach. Notably, nicotine remained predictive even after adjustment for self-reported smoking, suggesting that metabolomic biomarkers may capture aspects of tobacco exposure not fully reflected by questionnaire-based metrics, such as inhalation patterns, nicotine metabolism, or unreported exposure. We also identified additional exogenous compounds associated with COPD, including 1,3-benzenedicarbonitrile and 2-methyl-5-phenyl-1H-pyrrole, which were strongly correlated with smoking behavior. 1,3-benzenedicarbonitrile (isophthalonitrile) is an industrial intermediate that can also arise from combustion processes, while 2-methyl-5-phenyl-1H-pyrrole is a heterocyclic compound commonly detected in combustion products, including tobacco smoke. Their strong correlation with smoking behavior in our data suggests that these associations most likely reflect smoking-related combustion exposures rather than independent environmental risk factors. These findings highlight the ability of untargeted metabolomics to capture a broader spectrum of smoking-related exposures, including combustion byproducts and correlated environmental co-exposures.

A key novel finding of this study was the association between lower butyrylcarnitine levels and COPD incidence among never-smokers. Butyrylcarnitine is involved in mitochondrial fatty-acid oxidation and reflects systemic energy metabolism. While our use of EDTA plasma limits interpretation to systemic rather than tissue-specific metabolite levels, several biological hypotheses might explain this novel finding. Given C4’s role as a marker of mitochondrial β-oxidation, a systemic deficit may reflect a potential disruption in this critical energy pathway.

This could reflect chronically insufficient substrate availability from sources like the gut, or a reduced mitochondrial capacity to process available fuel.^18^ Regardless of the precise origin, the downstream consequence could be a compromised ability of lung epithelial cells to maintain cellular homeostasis and effectively repair environmental challenges.^19^ Alternatively, reduced plasma C4 could serve as a systemic indicator of perturbations in SCFA metabolism. This may reflect altered gut microbiome function, though plasma C4 represents host metabolism of butyrate rather than direct microbial production.^19^ These two potential vulnerabilities of cellular energy and of gut-derived signaling could be driven or exacerbated by environmental insults such as air pollution.^19,20^ The oxidative burden imposed by pollutants requires robust cellular repair mechanisms and individuals with pre-existing metabolic limitations may be particularly vulnerable to cumulative lung damage from such exposures. The specificity of this association to never-smokers suggests that metabolic factors might play a more prominent etiologic role when direct tobacco toxicity is absent. However, an association with a single metabolite warrants cautious interpretation. Moreover, our hypotheses frame lower C4 as a systemic shortage, but it could also indicate heightened consumption by tissues as a result of metabolic stress or altered renal clearance.

Our study also identified phenyl N-methylcarbamate, an insecticide metabolite^21^, as a predictor of COPD when analyses were restricted to metabolites with higher long-term stability. Carbamate insecticides have previously been linked to impaired lung function and respiratory disease, particularly in occupational settings.^3,22^ Although this association did not meet statistical significance in the primary analysis, its emergence in the stability-restricted analysis highlights the importance of accounting for temporal variability when evaluating environmental exposures using metabolomics. More broadly, while our GC-HRMS platform detected a wide range of environmental contaminants, including phthalates, phenols such as bisphenol A, persistent organic pollutants such as polychlorinated biphenyls, and multiple pesticides, most did not show robust associations with COPD incidence. This may reflect relatively low exposure levels in general population cohorts, limited statistical power after multiple testing correction, or substantial temporal variability in exposure.

Consistent with this interpretation, we observed generally modest long-term reproducibility of metabolomic features, particularly for LC-derived metabolites. This temporal variability likely attenuated associations and reduced power to detect true relationships. Restricting analyses to more stable metabolites identified additional associations, underscoring the importance of incorporating exposure stability into metabolomic epidemiology. Previous research has reported moderate to fair repeatability for LC-HRMS measurements from the same analytical platform at IARC over shorter time frames (e.g., a median ICC of 0.51 over 100 days^15^). In contrast, we observed substantially lower repeatability over a five-year period, with a median ICC of 0.18 for LC-based features and 0.34 for GC-based clusters. The higher ICCs for GC clusters likely reflect differences in chemical classes captured by each platform, rather than technical noise, as technical variation was explicitly adjusted for in our analyses. The generally low long-term reproducibility of many features suggests that temporal variability may have attenuated our ability to detect true associations with COPD risk, particularly for more biologically or exposure dynamic compounds.

We also observed substantial heterogeneity across cohorts. Several metabolites, including lipid species and exogenous compounds, were associated with COPD in individual cohorts but did not replicate consistently. These differences may reflect variation in exposure patterns, smoking behaviors, follow-up duration, or statistical power. For example, the relatively limited smoking contrast in SAPALDIA may have reduced the ability to detect smoking-related and other metabolomic associations. Notably, some environmental contaminants previously implicated in COPD or respiratory decline (e.g. p,p’-DDE^3^), showed a nominal or cohort-specific association, but did not replicate consistently or survive correction for multiple testing.

Our findings should also be interpreted in the context of COPD pathogenesis. Although all participants had normal lung function at baseline, cases had lower initial FEV₁/FVC ratios and experienced faster decline during follow-up. Therefore, identified metabolites could reflect early disease processes, susceptibility to accelerated lung function decline, or both. Future studies with more detailed longitudinal lung function data could help distinguish these possibilities.

This study has several important strengths. Our study represents the largest prospective investigation that combines both LC- and GC-HRMS platforms to simultaneously assess exogenous environmental exposures and endogenous metabolic perturbations in relation to COPD incidence. Most existing metabolomic studies focus on established COPD in cross-sectional designs, where reverse causation and confounding are more difficult to address. In contrast, our prospective design captures metabolomic profiles before clinical thresholds are crossed, potentially reflecting early pathophysiological changes or upstream risk factors.

Several limitations should also be considered. First, despite extensive screening, relatively few metabolites were associated with COPD after multiple testing correction, which may reflect limited power, exposure variability, or modest effect sizes. Second, while the pre-bronchodilation FEV1/FVC ratio is a recognized metric in the diagnosis of COPD, it does not capture all aspects of an official diagnosis. Therefore, this may have led to outcome misclassification. Moreover, some meaningful biological signals may lie in relationships between metabolites (e.g., ratios or co-expression networks) rather than in individual features. While this study focused on single-metabolite associations, future work could explore these more complex interactions. Lastly, although we employed a dual-platform approach, there are still gaps in our coverage of all possible circulating metabolites. Consequently, our findings should be interpreted as reflecting a subset of the complete metabolome. Future studies incorporating additional analytical approaches may provide more comprehensive metabolome coverage and potentially reveal additional associations with COPD incidence. For example, the addition of hydrophilic interaction liquid chromatography (HILIC) for highly polar metabolites or additional phases optimized for specific compound classes, could help dissect a validated association of glycoprotein acetyls with COPD incidence at the molecular level, potentially identifying more specific and mechanistically informative biomarkers for COPD development.^23^

In conclusion, this large prospective exposome-scale metabolomics study demonstrates the feasibility of detecting environmental contaminants and metabolic alterations years before COPD onset. In addition to confirming tobacco-related biomarkers, we identified butyrylcarnitine as a novel metabolic predictor of COPD among never-smokers and observed suggestive associations with environmental contaminants such as phenyl N-methylcarbamate. These findings highlight the potential of untargeted metabolomics to identify early molecular markers of COPD and provide a foundation for future replication, mechanistic studies, and exposomic investigations of disease etiology.

## Data Availability

All data produced in the present study are available upon reasonable request to the authors

## Acknowledgements

This work was supported by the EXPANSE and EXPOSOME-NL projects. The EXPANSE project is funded by the European Union’s Horizon 2020 research and innovation programme under grand agreement No 874627. The EXPOSOME-NL project is funded through the Gravitation program of the Dutch Ministry of Education, Culture, and Science and the Netherlands Organization for Scientific Research (NWO grant number 024.004.017).

The Swiss National Science Foundation (grants no 33CS30-177506/1, 33CS30-148470/1C2, 33CSCO-134276/1, 33CSCO-108796,, 324730_135673, 3247BO-104283, 3247BO-104288, 3247BO-104284, 3247-065896, 3100-059302, 3200-052720, 3200-042532, 4026-028099, PMPDP3_129021/1, PMPDP3_141671/1).

Where authors are identified as personnel of the International Agency for Research on Cancer/World Health Organization, the authors alone are responsible for the views expressed in this article and they do not necessarily represent the decisions, policy or views of the International Agency for Research on Cancer/World Health Organization.

## KORA

The KORA study was initiated and financed by the Helmholtz Zentrum München – German Research Center for Environmental Health, which is funded by the German Federal Ministry of Education and Research (BMBF) and by the State of Bavaria. Data collection in the KORA study is done in cooperation with the University Hospital of Augsburg.

We thank all participants for their long-term commitment to the KORA study, the staff for data collection and research data management and the members of the KORA Study Group (https://www.helmholtz-munich.de/en/epi/cohort/kora) who are responsible for the design and conduct of the study.

## SAPALDIA

The study could not have been done without the help of the study participants, technical and administrative support and the medical teams and field workers at the local study sites.

## Authorship contribution

|  |  |
|---|---|
| Max J. Oosterwegel | Conceptualization, data curation, formal analysis, investigation, software, validation, visualization, methodology, writing – original draft, writing – review & editing |
| Anastasia Chrysovalantou Chatziioannou | Data curation, investigation, methodology, validation, resources, writing – review & editing |
| Nivonirina Robinot | Investigation, methodology, validation, writing - review & editing |
| Pekka Keski-Rahkonen | Funding acquisition, Methodology, Supervision, writing – review & editing |
| Brooklynn R. McNeil | Data curation, investigation, methodology, resources, writing – review & editing |
| Randolph R. Singh | Data curation, investigation, methodology, resources, writing – review & editing |
| Gary W. Miller | Data curation, investigation, methodology, resources, writing – review & editing |
| Ayoung Jeong | Conceptualization, data curation, project administration, resources, writing – review & editing |
| Medea Imboden | Investigation, writing - review & editing - preparation |
| Megi Vogli | Project administration, writing – review & editing |
| Regina Pickford | Project administration, Resources, writing – review & editing |
| Annette Peters | Funding acquisition, conceptualization, writing – review & editing |
| Peter E. Engelfriet | Resources, writing – review & editing |
| W.M.Monique Verschuren | Funding acquisition, writing – review & editing |
| Roel Vermeulen | Funding acquisition, conceptualization, supervision, writing – review & editing |
| Nicole Probst-Hensch | Funding acquisition, investigation, methodology, writing – review & editing |
| Jelle Vlaanderen | Funding acquisition, conceptualization, supervision, writing – review & editing |

## Supplementary Methods I: Matching procedure information per cohort (case-control selection)

The table below shows what variables each of the cohorts matched on:

|  | **Sex** | **Age (±5 y)** | **Diagnosis and sample round** | **Season (month ±1)** | **Part of day** | **Fasting** |
|---|---|---|---|---|---|---|
| **Doetinchem Cohort Study** | X | X | X | X | X |  |
| **SAPALDIA** | X | X | X |  |  |  |
| **KORA** | X | X | X | X** | X | X |

\*\*one subject with 3 months and three subjects with two months difference.

### KORA

The KORA samples were from S4 (1999-2001), but spirometry collection started only during a later follow-up. Therefore, baseline (i.e. where both cases and controls are above threshold) used the spirometry from KORA F4 (2006-2008, first round after S4). The follow-up where cases became cases was in the FuL (2009-2010) or the FF4 (2013-2014) follow-up. Some participants took part in only one of these follow-ups, most of them in both. The FEV1, FVC ratio was derived manually and did not necessarily come from the same maneuver.

Availability of genetics or methylation data were not taken into account when selecting the participants.

### SAPALDIA

The SAPALDIA samples from SAP2 were used as baseline. Follow-up was either SAP3 or SAP4. The FEV1, FVC parameters used to calculate their ratio came from the same maneuver. The FEV1 and FVC measurements from SAP3 and SAP4 were adjusted for change in spirometry device during follow-up.^24^

Cases were selected regardless of genome/methylome data availability. Controls were prioritized for those with genome data, and if possible methylome as well, as follows:

1. Matching was first conducted among the controls with genome and methylome available.
2. When (1) failed, search was broadened to those with genome only.
3. When (2) failed, search was broadened to all controls regardless of genome/methylome availability.

### Doetinchem Cohort Study

The Doetinchem Cohort Study case-control samples were from round 4 and were used as baseline. The repeats (not included in case-control study) were from round 3. Follow-up was round 5. The FEV1, FVC ratio was derived manually and did not necessarily come from the same maneuver.

### Post-processing

During initial data analysis we discovered that some samples from KORA and the Doetinchem Cohort Study did not meet the prescribed case and control definitions. Specifically, control subjects from the Doetinchem Cohort Study sometimes also fell below the threshold during follow-up, and some cases did not have spirometry available at baseline or follow-up. 9 cases, 32 controls were removed. In the KORA cohort, two cases had a spirometry of 0.70 during follow-up, and one case had no spirometry measurements during baseline. These subjects from these cohorts were excluded from the case-control analysis upon discovery.

## Supplementary Methods II: LC- and GC-HRMS analysis

### Sample preparation LC

For the LC analysis, samples were prepared by mixing 35 µL of plasma with 200 µL of cold acetonitrile and filtering the precipitate with 0.2 µm Captiva ND plates (Agilent Technologies). 100 µL of the filtrate was mixed with equal volume of ultrapure water, sealed (Rapid EPS, BioChromato, Fujisawa, Japan), and analyzed. Quality control (QC) samples were prepared from a sample pool that was made by mixing 50 μL aliquots of all AMI samples of the Moli-Sani cohort (another EXPANSE cohort) and extracted along the study samples. Blank samples were prepared along the plasma samples in an identical manner, only leaving the plasma out of the process. Each well plate included four individually prepared QCs and two blanks. A QC sample was injected every ca. 20 samples, with a repetition of each QC taking place after ca. 10 injections. In total, all samples were processed in four analytical batches, each consisting of five 96-well plates.

### Sample preparation GC

For the GC analysis, samples were thawed at 4°C, and 100 μL plasma was treated with 25 μL formic acid and extracted by adding 200 μL (4:1 hexane/ethyl acetate) containing 13C labeled internal standards (ISs). Treated samples were vortex mixed for 1 hour, and then centrifuged for 10 min at 4000rpm, at 4°C. After centrifuging, 150 μL of supernatant was transferred to a new 96-well plate containing ∼20mg of MgSO4, PSA, and C-18, vortex mixed for 2 minutes, and centrifuged for 10 min at 4000rpm. The resulting supernatant was transferred to a 96-well plate containing glass inserts for analysis. QAQC samples were prepared along with the study samples consisted of blanks, two pooled plasma samples and a standard reference material (SRM). For the process blanks 100 μL HPLC grade water was used instead of plasma. The pooled plasma consisted of ‘Pool 1’ (20-35 years old), and ‘Pool 2’ (50+ years old). The SRM used was NIST 1958, freeze-dried plasma fortified with environmental contaminants. All QAQC samples were injected at the beginning and end of the sequence. A blank and Pool 2 sample were injected every 20 samples. In total, all samples were processed in 22 batches of 96-well plates – each containing up to 82 study samples and up to 14 QAQC samples – using OT-2 liquid handling robot (Opentrons Labworks Inc., New York, NY, USA).

### LC analysis and processing

A UHPLC-QTOF-MS system consisted of a 1290 Binary LC system, and a 6550 QTOF mass spectrometer with Jet Stream electrospray ionization source (Agilent Technologies). Samples were kept at 4 °C and 2 µL was injected. An ACQUITY UHPLC HSS T3 column (2.1 × 100 mm, 1.8 μm) was used at 45 °C and the mobile phase consisted of ultrapure water and LC-MS grade methanol, both with 0.05 % (v/v) of formic acid. The gradient profile was as follows: 0–6 min: 5% to 100% methanol, 6–10.5 min: 100% methanol, 10.5–13 min: 5 % methanol. The flow rate was 0.4 ml/min. Mass spectrometer drying gas temperature was 175 °C and flow 12 L/min, with capillary, nozzle, and fragmentor voltages of 3500 V, 300 V, and 175 V, respectively. The sheath gas temperature was 350 °C and flow 11 L/min, and nebulizer pressure 45 psi. MS/MS analyses were performed with an isolation width of 1.3 Da and collision energies of 10 V, 20 V, and 40 V. Continuous mass axis calibration was employed using lock mass ions m/z 121.0509 and 922.0098. Data was acquired in centroid format using an extended dynamic range mode, and acquisition rate of 1.67 Hz, over the mass range of 50-1200 Da (MassHunter Acquisition 10.1, Agilent Technologies).

Pre-processing was performed using Profinder 10.0.2 and Mass Profiler Professional B.14.9.1 software (Agilent Technologies). A “Batch Recursive Feature Extraction (small molecules/peptides)” process was employed to find [M+H]^+^ ions utilizing the study samples and the blanks of each batch. Mass peak height threshold of 1500 counts and chromatographic peaks threshold of 8000 counts were used, while the minimum quality score was 70. The isotope grouping model used was for common organic molecules. Feature alignment between samples was performed with retention time and mass windows of ±0.03 min and ±(10 ppm + 1 mDa), respectively. Features detected in at least 25 % of the samples within a batch were recursively extracted using ±25 ppm m/z width to draw chromatographic peaks, Agile 2 integrator, and no smoothing of the chromatograms. Mass was calculated as an average from the spectra at >50 % peak heights. The recursive feature extraction was repeated as above but with a mass peak height threshold of 10 000 counts and chromatographic peaks threshold of 50,000 counts, and the frequency of detection filter set to 2.5%. The resulting features were matched against the targets using tolerances for the mass and retention time at ±10 ppm and ±0.03 min, respectively.

The four batches were processed separately, exporting the detected features as a .pfa files which were then aligned in Mass Profiler Professional. Before the alignment the background for each batch was excluded based on leaving out features present in every blank sample of the batch, unless 5-fold greater in average intensity in samples. The remaining features were exported as a single .cef file from each batch and feature alignment between samples of all batches was performed with retention time and mass windows of ±0.07 min and ±(15 ppm + 2 mDa). A single target list for the recursive extraction was created combining the features found in at least one of the four batches. Targeted feature extraction from all study samples and QCs

of each batch was then performed in Profinder in a “Batch targeted feature extraction” process using a match tolerance of ±10 ppm for masses and ±0.03 min for retention times. Ion species were limited to [M+H]^+^ and the integrator was Agile 2. No smoothing was applied on the chromatogram peaks and no further filtration took place. The resulting features from each sample were exported as an individual .cef file for alignment in Mass Profiler Professional. Alignment was performed using an RT window of ±0.1 min and a mass window of ±(15 ppm + 2 mDa). Chromatographic peak areas were used as a measurement of intensity.

Since the workflow can result in some duplicate features, a clustering-based method was applied to identify features with close retention time (±0.01 min) and mass (±10 ppm) that are also highly correlated (Pearson correlation >0.95). From the pairs of features matching these criteria, the one with the least nondetects and highest average intensity was retained. This workflow can result in few negative intensity values, which were replaced with nondetect values.

### GC analysis and processing

Data acquisition was performed using a Trace 1310 gas chromatography system coupled to an Exploris GC 240-HRMS system (Thermo Fisher Scientific, Rockford, IL, USA). Separation was accomplished by injecting 6 µL of extract into a PTV inlet with a variable temperature program connected to a 15m DB-5MS column equipped with a 10m guard column (Restek), 1.2 mL/min flow rate, and the following oven temperature gradient: 50°C hold for 1.25 min, increase to 180°C at 50°C/min and hold for 0.5 min, increase to 250°C at 12°C/min, final temperature ramp to 320°C at 40°C/min and hold for 6 min. The HRMS resolution was set at 60,000 FWHM (at m/z 200). The scan range was set from 70 to 700 m/z; the automatic gain control (AGC) target and maximum injection time in full-scan MS settings were set to ‘Standard’ and ‘Auto’, respectively. This allowed the instrument to automatically adjust the injection time for an ion count of ∼1 × 106 for MS. The electron ionization source was maintained at 280°C and 70eV during the run. With every batch, the source was cleaned, mass spectrometer tuned and calibrated, and the system checked for leaks. The column was changed roughly every 700 injections by monitoring peak shape of internal standards as reference. Data was qualified using median relative standard deviation (RSD) for 13C labelled internal standards: RSD for internal standards for all samples in batch, RSD for spiked analytes in pooled reference samples; and overall study QA/QC (RSD and intensity distribution for common analytes across all batches, distribution of the sum of feature intensity across all samples and batch, intensity of analytes in method blanks). QA/QC analysis was completed after every batch using TraceFinder and batches not meeting acceptance criteria were discarded and re-analyzed.

Following analysis of all study and QAQC samples, raw instrument files were converted to .mzML for using Proteowizard. Raw retention times were converted to their respective indices using alkanes and Kovats scale before extraction. Peaks were extracted using the XCMS package^8^ with a signal to noise threshold of 5, intensity threshold of 10,000 and mass accuracy of 5 ppm. Features that did not meet a 5-fold intensity change from blank to average sample intensity were marked and removed before clustering. The extracted feature table was filtered by at least 10% presence and clustered into spectra and abundances using RAMClustR^9^ with correlation parameters set to 8 and 1 for retention time and abundance, respectively. After clustering the features, RAMClustR uses a weighted mean function to combine the features of a cluster into an aggregated signal where the more abundant features contribute more to the spectral intensity of the cluster.

## Supplementary Methods III: Congenial imputation model

For any imputation method it is important that the imputation method (adequately) captures all aspects of the data that you may be interested in later on. In other words, you need to include all variables (including the outcome in stochastic multiple imputation like ours) in the imputation model that will be used the final analyses. In the statistics jargon this is usually referred to as a congeniality. To this end, we a priori identified that we would want to do a pooled analysis of case-status using conditional logistic regression, and potentially carry out analyses stratified by cohort, smoking, and length of follow-up time. We then built the following regression model to adequately capture aspect of the data important to these analyses (for example allowing the effect of case-status on the feature to differ by different strata of smoking and cohort):

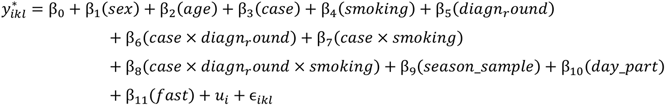

Where:

– The indices are defined as: *i* for batch (plate for LC, as described later), *k* for subject, and *l* for the individual observation/measurement. In this dataset, *k* and *l* were identical (each subject one measurement).
– *y*^∗^ represents the natural log-transformed intensity for a single, potentially left-censored observation. Observations are assumed to be censored at the minimum value of that batch/plate.
– *β*_0_ is the overall intercept.
– *β*_1_, …, *β*_11_ are the coefficients for the fixed effects, with the variable name in parentheses. All variables except age are coded as factor variables. The additional coefficients as a result of the dummy representation are omitted here for brevity.
– 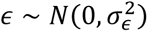 is the random intercept for the *i*-th plate / batch.
– 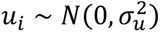 is the error term for the *l*-th observation from the *k*-th subject within the *i*- th batch/plate.

All matching variables (age, sex, season, part of day, fasting, diagnose round) are included to be consistent with the matched analysis later on. Diagnose round is a more granular description of a cohort and thereby functions as a substitute for the cohort variable. It is also a proxy for length of follow-up time. Given the stratified analyses of case-status later on we also allow the effect of case-status to vary by smoking and diagnose round. Since models would not converge when we added all the interactions of case-status with all variables, we induced some sparsity by selecting the interactions most important to our subsequent analyses. Moreover, we added a prior to these interactions to help with convergence and to prevent extremely large coefficient when a feature was not detected in a certain stratum. Specifically, we used a Cauchy prior with a mean of zero, sigma of 2 on the random effects and residual error parameters, a Student-t prior with 3 degrees of freedom, mean zero and sigma of 0.5 of on the coefficients involving an interaction (sigma 1 on the main effect coefficients). Lastly, we had a Student-t prior with 3 degrees of freedom, mean of 11 and sigma of 2.5 on the intercept term.

We did not model the full hierarchical technical structure (plates nested in batches batch) of the LC data to improve the speed of the model and left out the highest level.

These models were fitted using brms / STAN. We imputed values for the nondetects by drawing a random value (using the parameters of the model we fitted) between minus infinity and the batch adjusted limit of detection, meaning we adjusted the limit of detection for technical variation by subtracting the random batch intercept. And as explained earlier in the manuscript, we also subtracted the random intercepts from the imputations to remove these sources of technical variation from the data – thereby adjusting for technical variation and imputing nondetects in one and the same model.

## Supplementary Methods IV: ICC calculation

We calculated ICC values using all samples from the Doetinchem Cohort Study (original case-control samples (n = 295) + repeats (n = 198)). The ICC quantifies the proportion of the total variance that is attributable to differences between subjects. This is commonly referred to as repeatability, and can also be interpreted as the correlation between two different observations from the same class (i.e. samples from the same subject). Using the variance components of a random intercept model, we derived these values for the features/clusters that were found in at least 40% of Doetinchem Cohort Study samples. Following methodology from Oosterwegel et al. (2023), we fit a left-censored model to each of these natural log-transformed features using brms / STAN with an adapt_delta of 0.95. Other options were kept at the tools their default values (4 chains, 2000 samples per chain with 50% being burn in samples).

As the Gas Chromatography (GC) and Liquid Chromatography (LC) platforms had different experimental designs, we specified slightly different models for each. Namely, for the GC data we fit a model containing only a random intercept that describes the batch membership of the sample and a random intercept for the subject. The experimental design for the LC platform was hierarchical, with analytical plates nested within batches. Therefore, the technical variation was modelled as a plate nested in a batch for data from the LC platform. For both LC and GC data, we calculate the ICC by dividing the between subject variance by the between subject variance plus the residual variance (within-subject variance). The variation between batches was thus modelled but treated as nuisance variance and excluded from the ICC denominator.

In more precise math terms: for the GC data that involved crossed a subject and batch design, the model was:

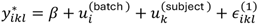

For data measured by LC that involved a hierarchical technical structure (plates nested in batches) design crossed with a subject effect, the model was:

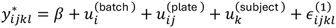

Where the terms for both models are defined as:

– The indices are defined as: *i* for batch, *j* for plate, *k* for subject, and *l* for the individual observation/measurement.
– *y*^∗^ represents the natural log-transformed intensity for a single, potentially left-censored observation. Observations are assumed to be censored at the minimum value of that batch.
– *β* is the fixed overall mean intensity for the feature.
– 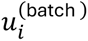 is the random intercept for batch *i*, assumed to be distributed as 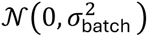.
– 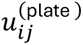 is the random intercept for plate *j* nested within batch *i*, used only in the LC model and assumed to be distributed as 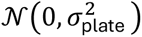.
– 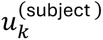 is the random intercept for subject *k*, assumed to be distributed as 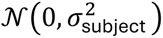.
– *ε*^(1)^ is the residual error term for an individual observation, assumed to be distributed as 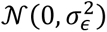.

For both the GC and LC models, the (adjusted) ICC could then be calculated as:

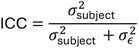

**Figure S1:**
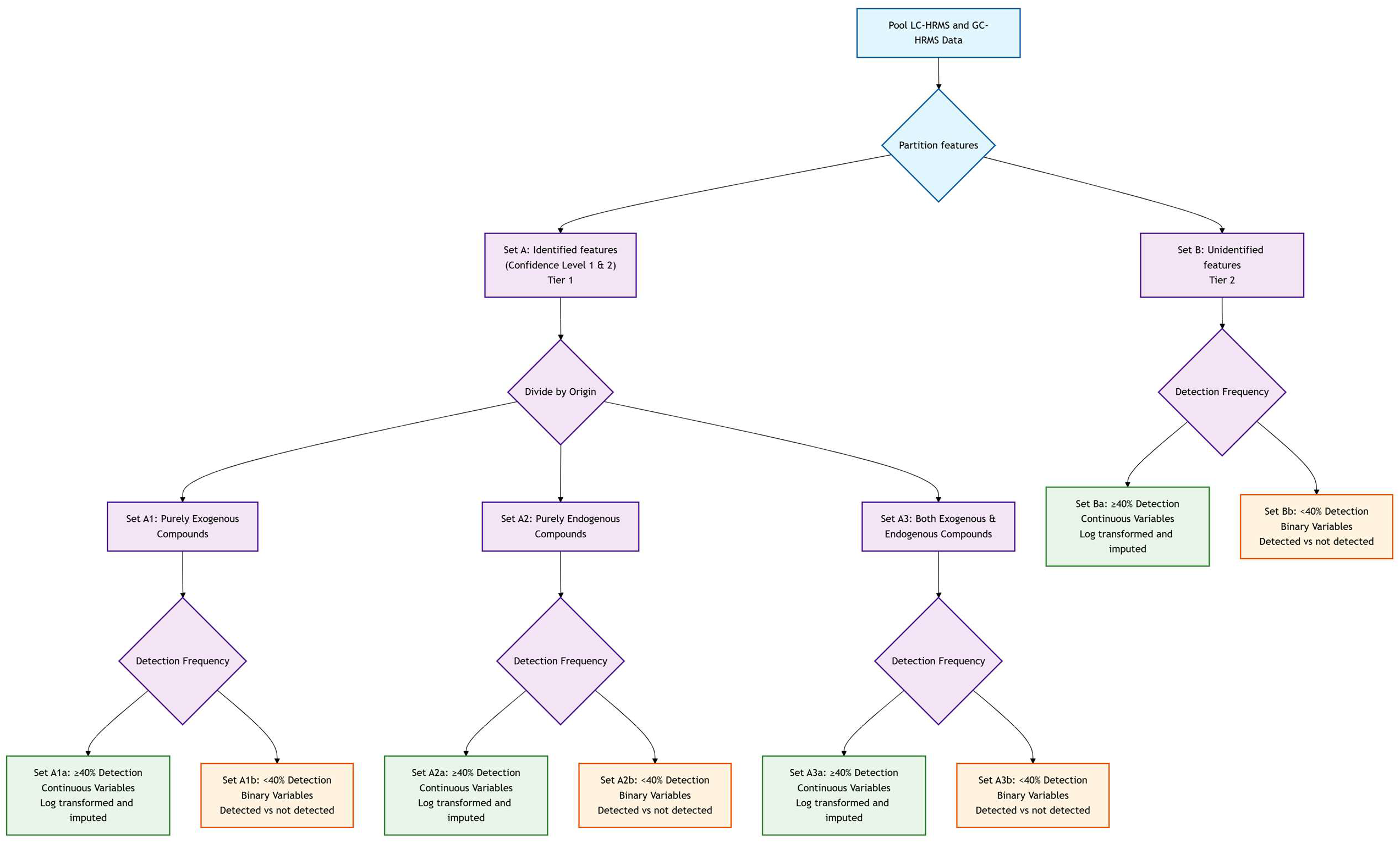
Mermaid diagram of portioning of LC- and GC-features into sets. Rectangles represent sets, diamonds represent decision points where data gets partitioned.

**Figure S2:**
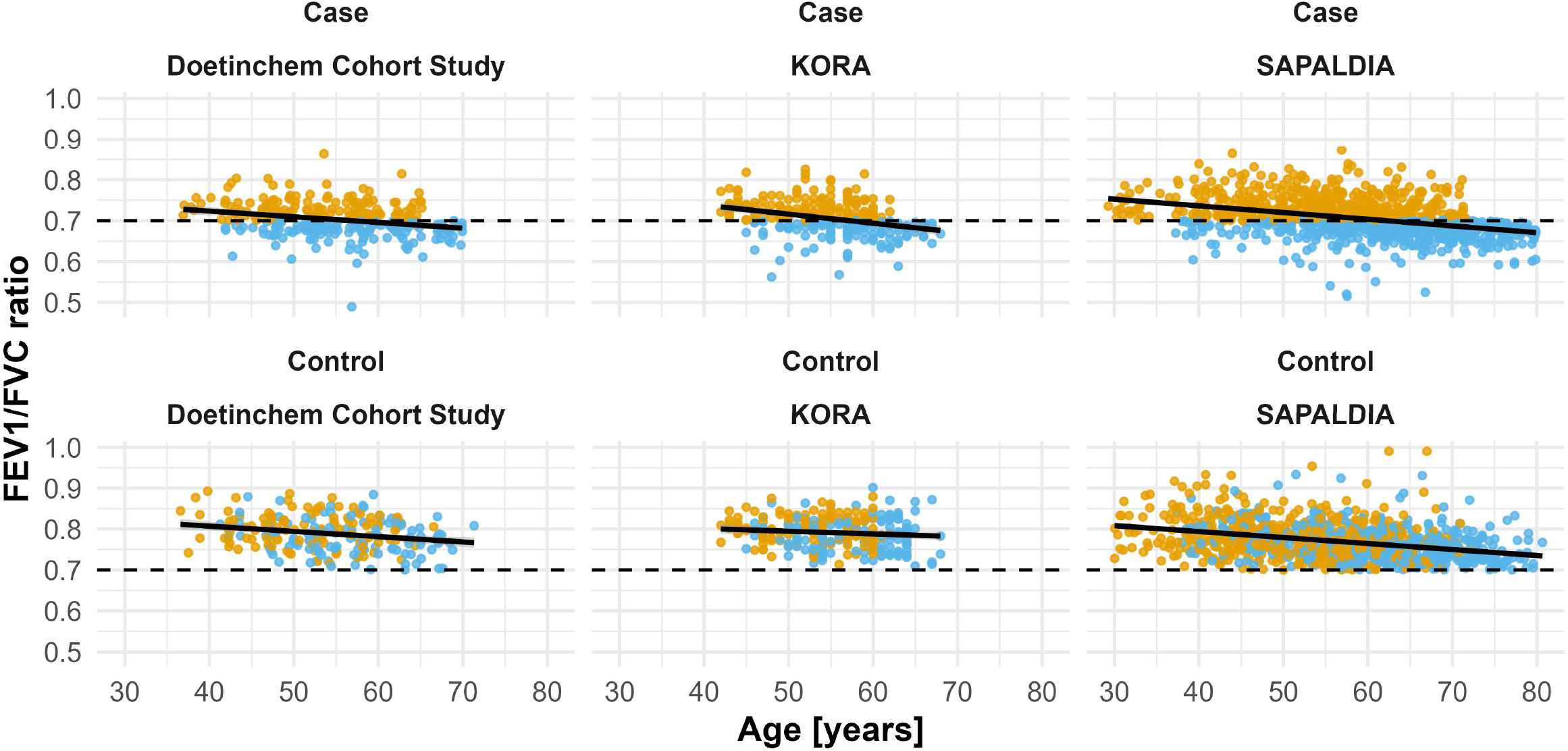
trajectories of FEV1/FVC ratio for the case-control study participants. The trend line is from a simple regression of the ratio on age per facet. The orange color indicates spirometry at baseline, the blue color at follow-up.

**Figure S3a:**
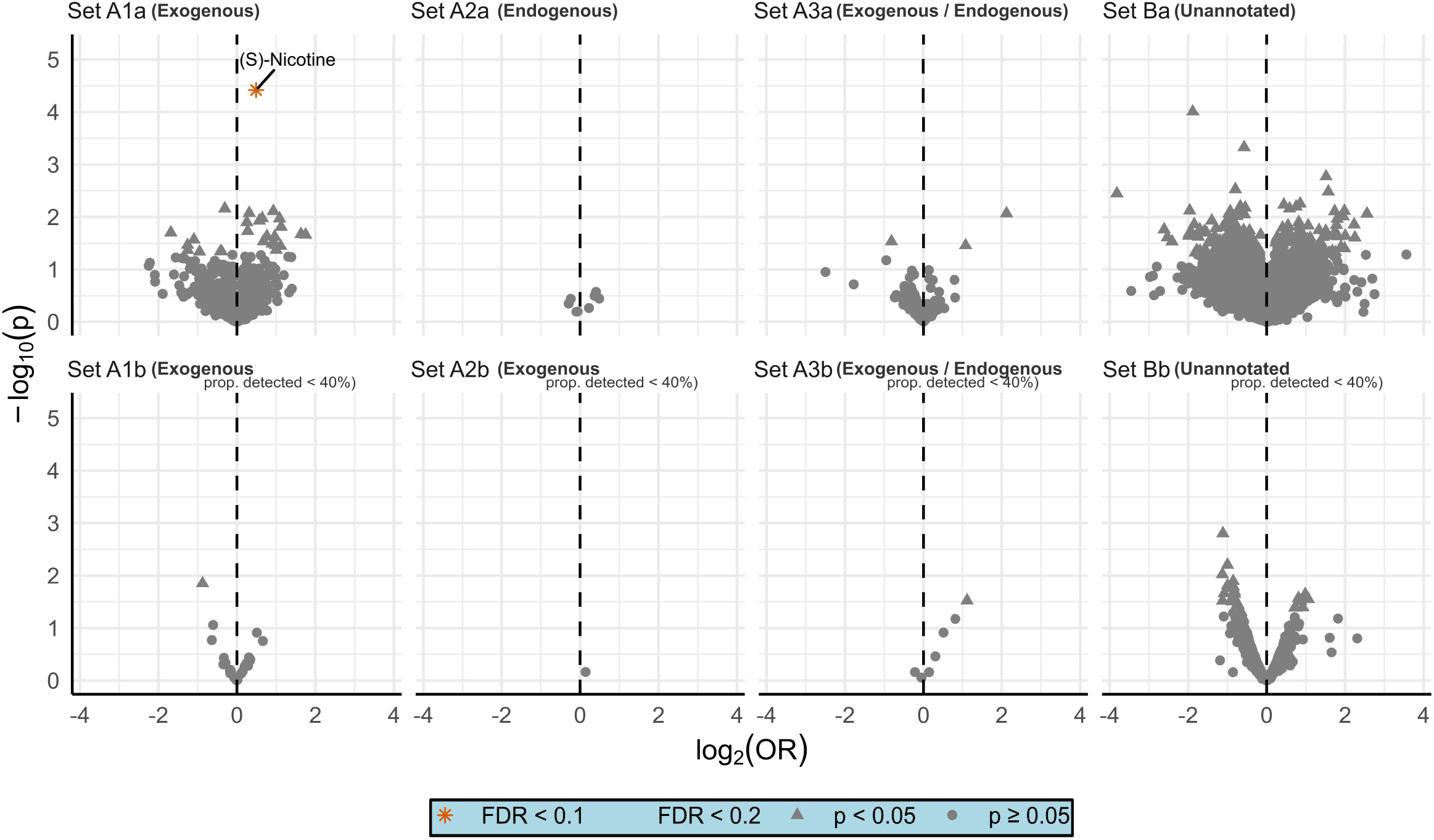
volcano plot of the results from the analysis stratified by smoking status (current). Set A describes the annotated metabolites, Set B indicates the initially unidentified features/clusters. 1, 2, 3 describes the exogenous, endogenous, both endogenous/exogenously derived metabolites respectively. The last letter indicates if the feature was analyzed as continuous variable (lower case ‘a’) or as binary variable of detected, not detected (lower case ‘b’). p = p-value; FDR = false discovery rate; OR = odds ratio.

**Figure S3b:**
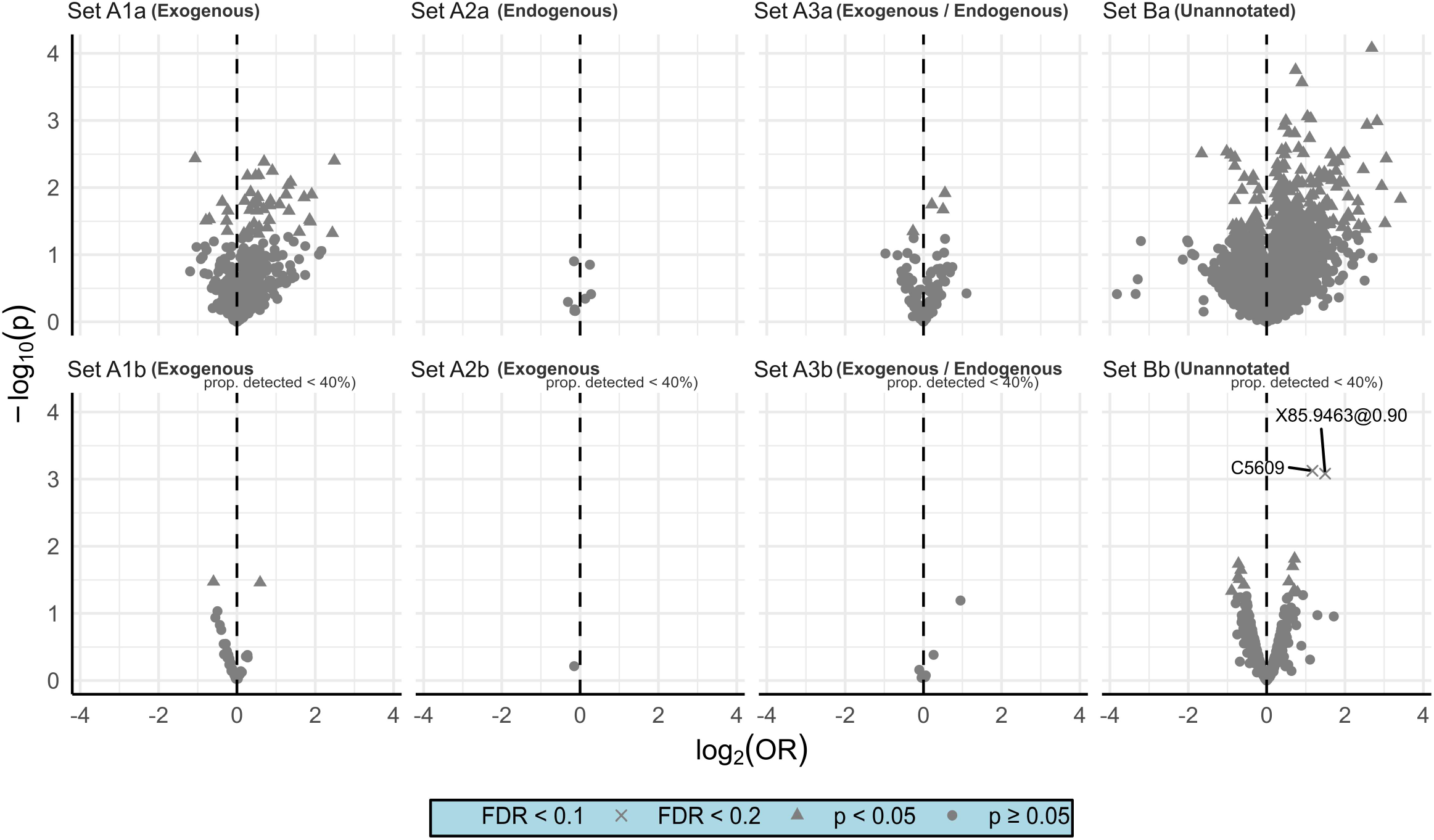
volcano plot of the results from the analysis stratified by smoking status (ex). Set A describes the annotated metabolites, Set B indicates the initially unidentified features/clusters. 1, 2, 3 describes the exogenous, endogenous, both endogenous/exogenously derived metabolites respectively. The last letter indicates if the feature was analyzed as continuous variable (lower case ‘a’) or as binary variable of detected, not detected (lower case ‘b’). p = p-value; FDR = false discovery rate; OR = odds ratio.

**Figure S4a:**
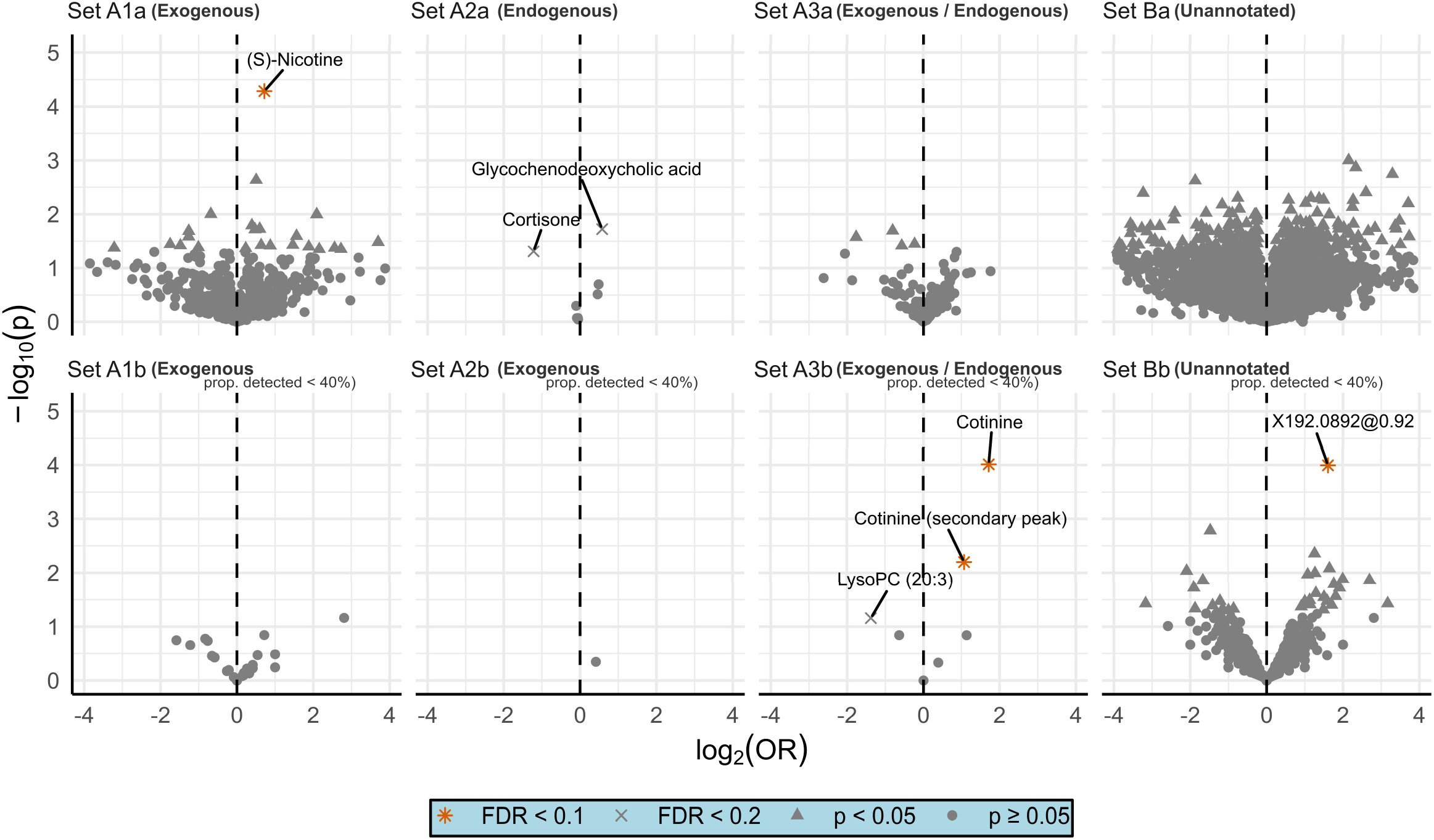
volcano plot of the results from the cohort stratified analysis (KORA cohort). Set A describes the annotated metabolites, Set B indicates the initially unidentified features/clusters. 1, 2, 3 describes the exogenous, endogenous, both endogenous/exogenously derived metabolites respectively. The last letter indicates if the feature was analyzed as continuous variable (lower case ‘a’) or as binary variable of detected, not detected (lower case ‘b’). p = p-value; FDR = false discovery rate; OR = odds ratio.

**Figure S4b:**
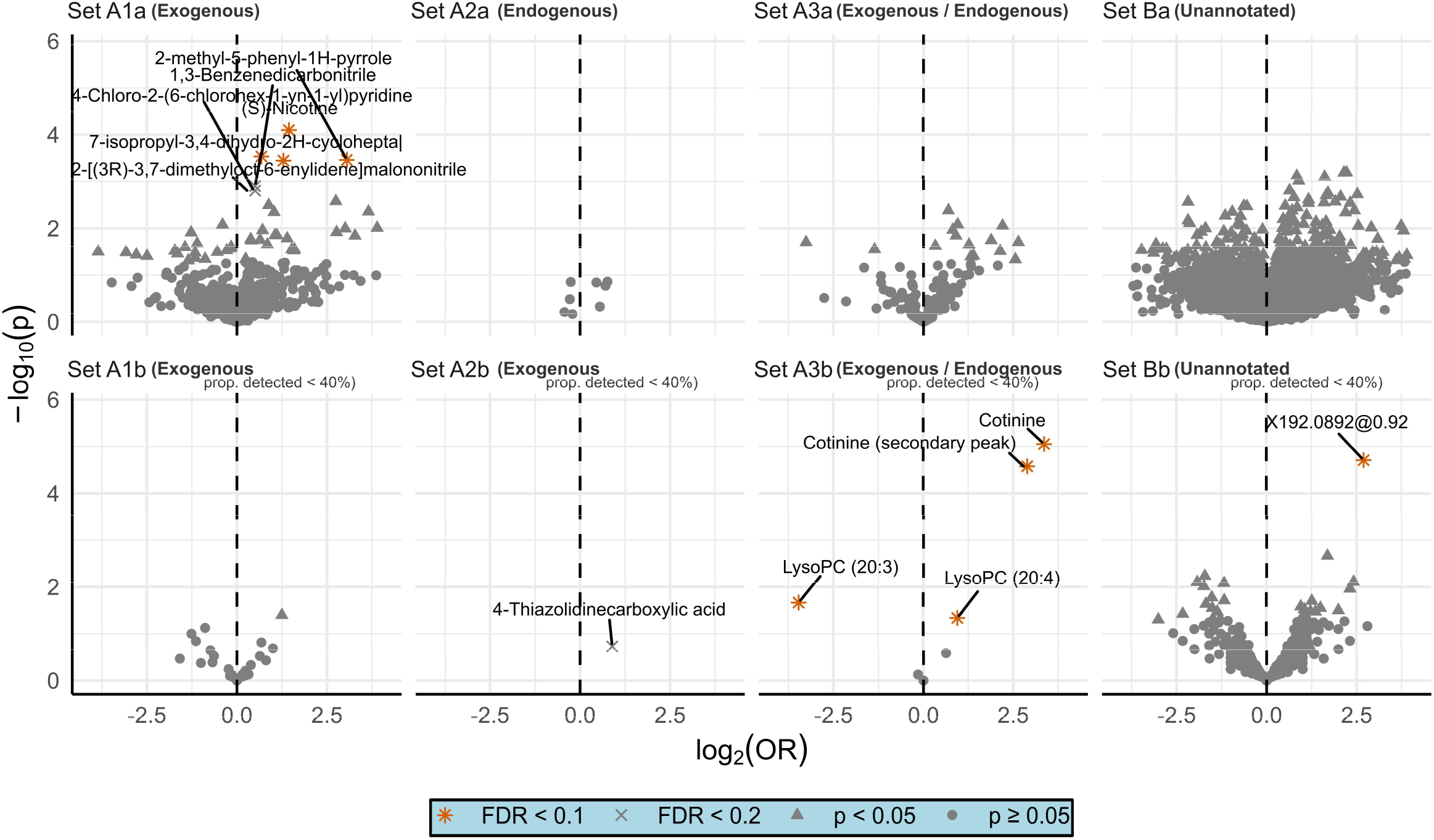
volcano plot of the results from the cohort stratified analysis (Doetinchem Cohort Study). Set A describes the annotated metabolites, Set B indicates the initially unidentified features/clusters. 1, 2, 3 describes the exogenous, endogenous, both endogenous/exogenously derived metabolites respectively. The last letter indicates if the feature was analyzed as continuous variable (lower case ‘a’) or as binary variable of detected, not detected (lower case ‘b’). p = p-value; FDR = false discovery rate; OR = odds ratio.:

**Figure S4c:**
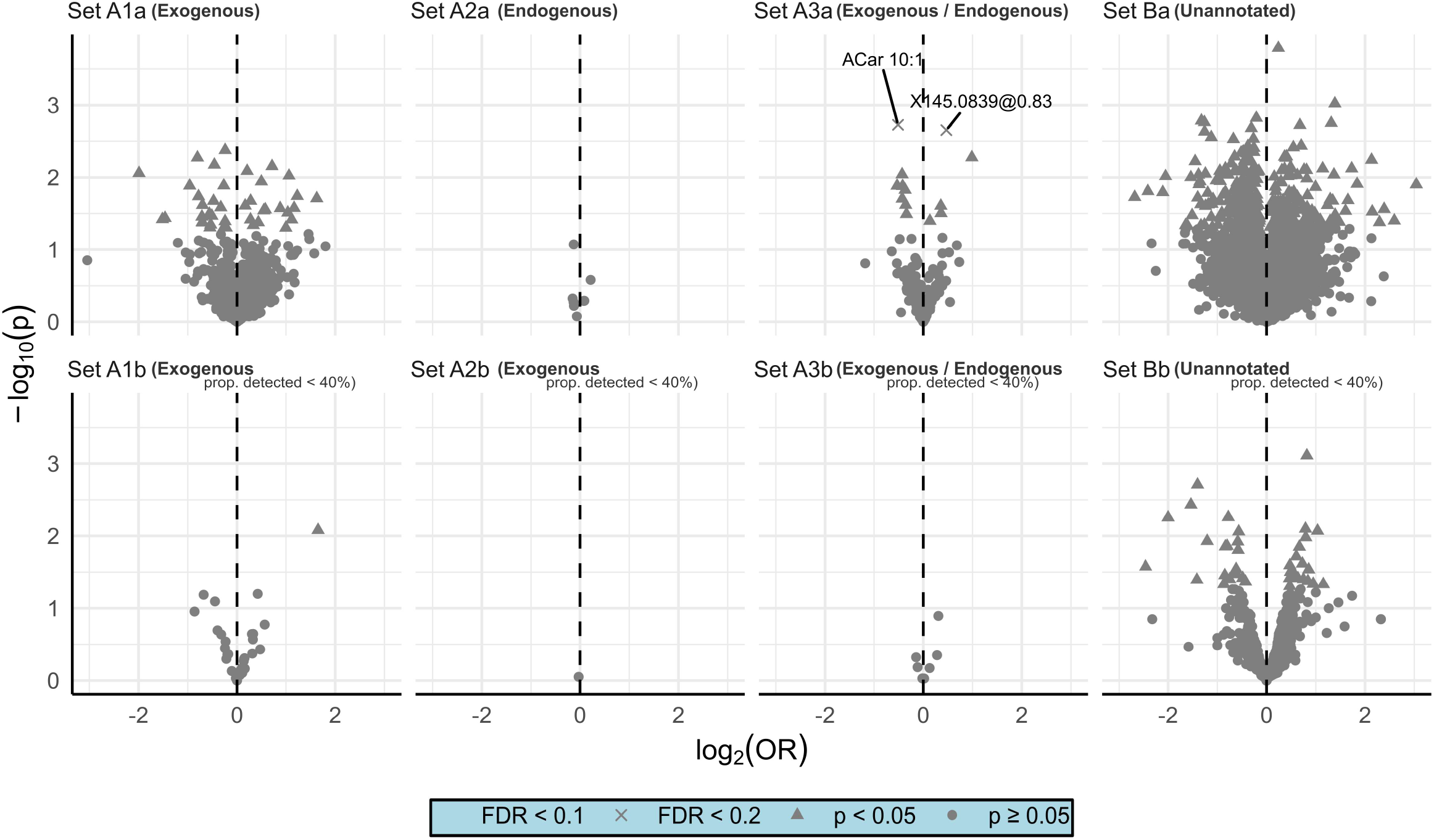
volcano plot of the results from the cohort stratified analysis (SAPALDIA cohort). 145.0839@0.83 was tentatively annotated but could not be validated in the end. Set A describes the annotated metabolites, Set B indicates the initially unidentified features/clusters. 1, 2, 3 describes the exogenous, endogenous, both endogenous/exogenously derived metabolites respectively. The last letter indicates if the feature was analyzed as continuous variable (lower case ‘a’) or as binary variable of detected, not detected (lower case ‘b’). p = p-value; FDR = false discovery rate; OR = odds ratio.

**Figure S5:**
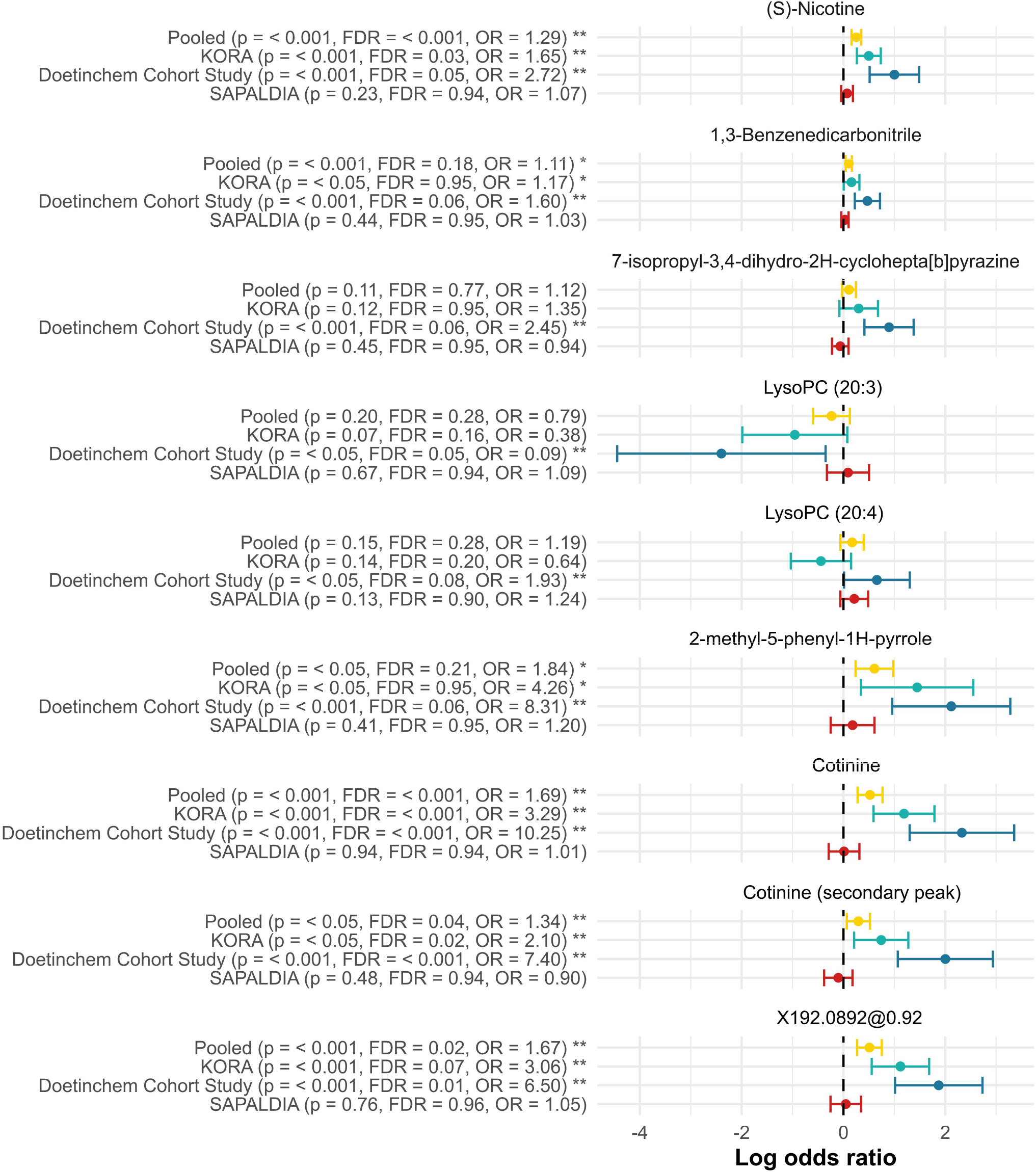
all compounds that were FDR significant at 10% in either the pooled or in one of the analyses stratified by cohort. p = p-value; FDR = false discovery rate; OR = odds ratio.

**Table S1a:**
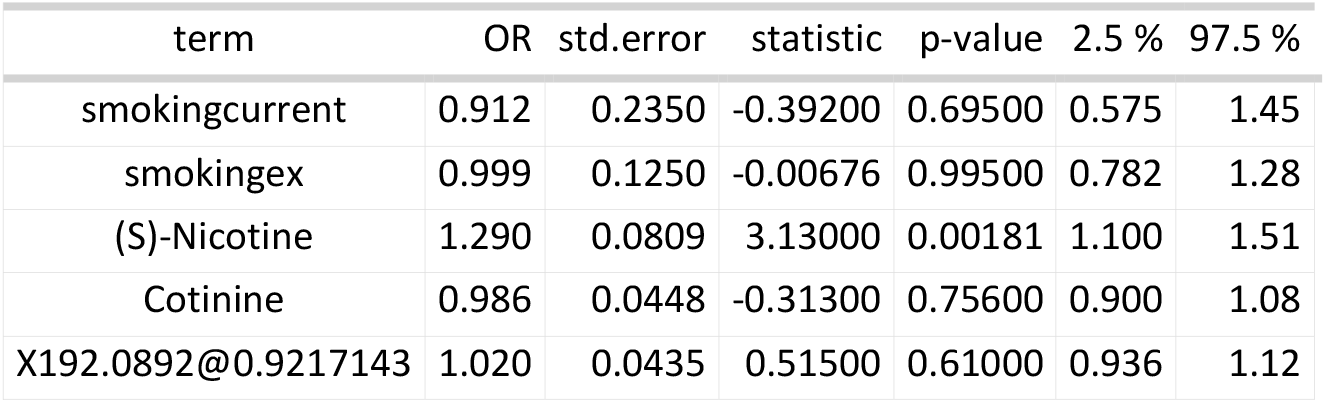
results of regressing COPD on each of these terms in a conditional logistic regression. Prior to this regression, batch effects were removed in a procedure similar to the one described in Supplementary Methods III. However, contrary to the main analysis imputation was done in a separate subsequent mice procedure with all these variables as predictor of each other. This was necessary to take the covariance between the features into account. OR = odds ratio.

| term | OR | std.error | statistic | p-value | 2.5 % | 97.5 % |
|---|---|---|---|---|---|---|
| smokingcurrent | 0.912 | 0.2350 | -0.39200 | 0.69500 | 0.575 | 1.45 |
| smokingex | 0.999 | 0.1250 | -0.00676 | 0.99500 | 0.782 | 1.28 |
| (S)-Nicotine | 1.290 | 0.0809 | 3.13000 | 0.00181 | 1.100 | 1.51 |
| Cotinine | 0.986 | 0.0448 | -0.31300 | 0.75600 | 0.900 | 1.08 |
| X192.0892@0.9217143 | 1.020 | 0.0435 | 0.51500 | 0.61000 | 0.936 | 1.12 |

**Table S1b:** results of regressing COPD on each of these terms in a conditional logistic regression. Prior to this regression, batch effects were removed in a procedure similar to the one described in Supplementary Methods III. However, contrary to the main analysis imputation was done in a separate subsequent mice procedure with all these variables as predictor of each other. This was necessary to take the covariance between the features into account. OR = odds ratio.

| term | OR | std.error | statistic | p-value | 2.5 % | 97.5 % |
|---|---|---|---|---|---|---|
| packyears | 1.010 | 0.00293 | 3.290 | 0.00105 | 1.000 | 1.02 |
| (S)-Nicotine | 1.200 | 0.08250 | 2.240 | 0.02550 | 1.020 | 1.41 |
| Cotinine | 0.975 | 0.04590 | -0.550 | 0.58700 | 0.888 | 1.07 |
| X192.0892@0.9217143 | 1.010 | 0.04200 | 0.298 | 0.76800 | 0.929 | 1.10 |

**Table S2:**
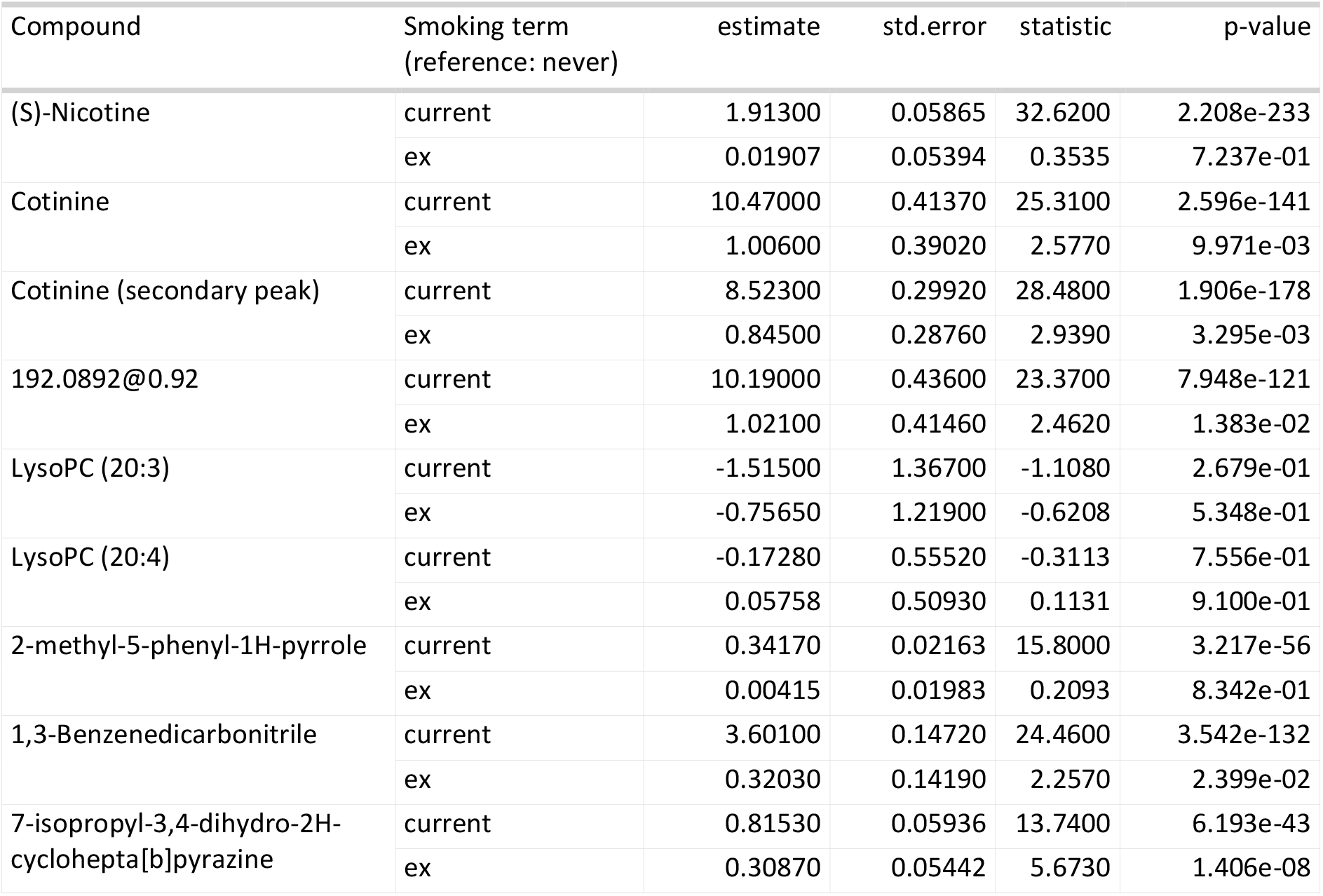
in the first column: all compounds that were FDR significant at 10% in either the pooled, or in one of the analyses stratified by cohort. The rest of the table describes the statistical relationship of these compounds with smoking behavior. All of these compounds were thus also related to COPD in (part of) the data. Smoking behavior is, of course, also related to COPD. Therefore we are at risk of increased false positives of the relationship of these compounds with smoking (secondary analysis of case-control data). It is very likely, however, that this increased false positive rate is negligible. See for example the difference in the p-value magnitude known smoking related compounds (e.g. nicotine) and the lipids. The lipids essentially function as a negative control here. The p-value of Pyrrole, 2-methyl-5-phenyl-, 1,3-Benzenedicarbonitrile, 7-isopropyl-3,4-dihydro-2H-cyclohepta[b]pyrazine is extremely small – contrary to the lipids, but similar to known smoking related compounds – suggesting these compounds are also strongly related to smoking (and not a false positive through relationship with COPD). The relationship of these compounds was estimated by fitting a left-censored model with Gaussian errors to the compound with smoking, batch/plate id and an intercept as main effects (using all case-control data samples). Nondetects were assumed to be left-censored at the minimum value of the batch/plate.

**Table S3:** in the first column: all compounds that were FDR significant at 20% (but not at 10%) in the pooled analysis, or at 10% after the ICC filter. The rest of the table describes the statistical relationship of these compounds with smoking behavior. All of these compounds were thus also related to COPD in (part of) the data. The relationship of these compounds was estimated by fitting a left-censored model with Gaussian errors to the compound with smoking, batch/plate id and an intercept as main effects (using all case-control data samples). Nondetects were assumed to be left-censored at the minimum value of the batch/plate. (Same as Table S1)

| Compound | Smoking term<br>(reference: never) | estimate | std.error | statistic | p-value |
|---|---|---|---|---|---|
| Glycochenodeoxycholic acid | current | -0.0836200 | 0.05204 | -1.6070 | 1.081e-01 |
|  | ex | -0.0712200 | 0.04785 | -1.4880 | 1.367e-01 |
| 129.0791@0.74 | current | -0.1831000 | 0.09042 | -2.0250 | 4.284e-02 |
|  | ex | -0.2077000 | 0.08313 | -2.4980 | 1.248e-02 |
| Cortisone | current | 0.0680500 | 0.02137 | 3.1850 | 1.450e-03 |
|  | ex | 0.0592700 | 0.01965 | 3.0160 | 2.558e-03 |
| ACar 10:1 | current | 0.0474500 | 0.03969 | 1.1960 | 2.319e-01 |
|  | ex | 0.0932100 | 0.03649 | 2.5540 | 1.064e-02 |
| 145.0839@0.83 | current | -0.0577600 | 0.04873 | -1.1850 | 2.358e-01 |
|  | ex | 0.0540900 | 0.04480 | 1.2070 | 2.273e-01 |
| LysoPC (20:3) | current | -1.4440000 | 1.26800 | -1.1390 | 2.548e-01 |
|  | ex | -0.7093000 | 1.13000 | -0.6278 | 5.301e-01 |
| 2,2,4,7-tetramethyl-3H-1-benzofuran | current | 0.1834000 | 0.01396 | 13.1400 | 1.981e-39 |
|  | ex | 0.0147600 | 0.01279 | 1.1540 | 2.486e-01 |
| 1,3-Benzenedicarbonitrile | current | 4.2690000 | 0.17800 | 23.9800 | 4.037e-127 |
|  | ex | 0.4041000 | 0.17040 | 2.3720 | 1.770e-02 |
| 4-Chloro-2-(6-chlorohex-1-yn-1-yl)pyridine | current | 1.7540000 | 0.10170 | 17.2500 | 1.114e-66 |
|  | ex | 0.1897000 | 0.09376 | 2.0230 | 4.309e-02 |
| 2-[(3R)-3,7-dimethyloct-6-enylidene]malononitrile | current | 2.4230000 | 0.12250 | 19.7800 | 4.312e-87 |
|  | ex | 0.8950000 | 0.11350 | 7.8840 | 3.167e-15 |
| 4-Thiazolidinecarboxylic acid | current | 0.7413000 | 0.33080 | 2.2410 | 2.503e-02 |
|  | ex | 0.3238000 | 0.30650 | 1.0560 | 2.908e-01 |
| phenyl N-methylcarbamate | current | -0.0003887 | 0.02300 | -0.0169 | 9.865e-01 |
|  | ex | -0.0450000 | 0.02109 | -2.1340 | 3.283e-02 |
| C5155 | current | 0.2366000 | 0.24330 | 0.9727 | 3.307e-01 |
|  | ex | -0.0759900 | 0.22660 | -0.3353 | 7.374e-01 |

**Figure S6a:**
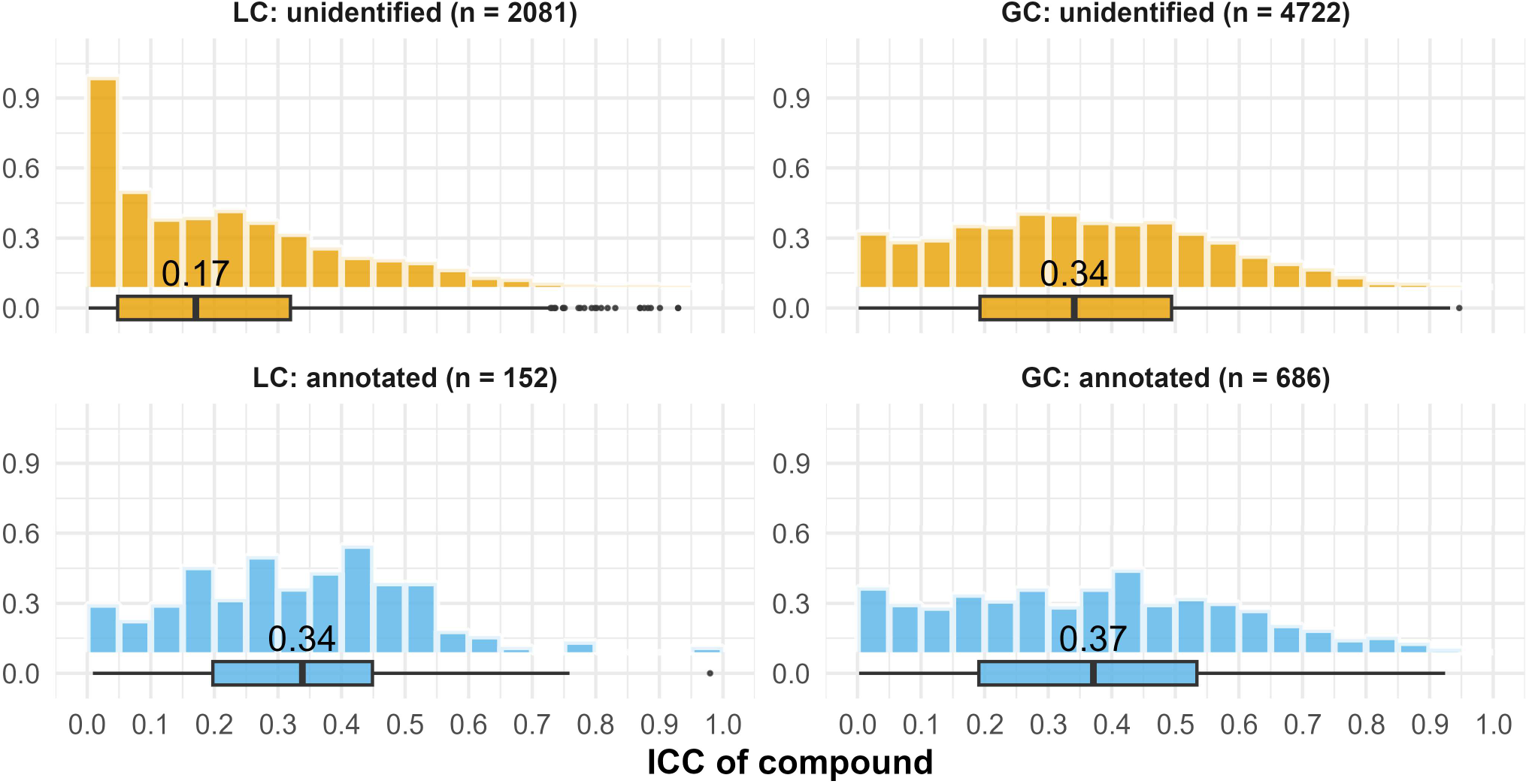
temporal repeatability clusters per platform and annotation status. ICC = intraclass correlation coefficient; GC = Gas Chromatography. LC = Liquid Chromatography.

**Figure S6b:**
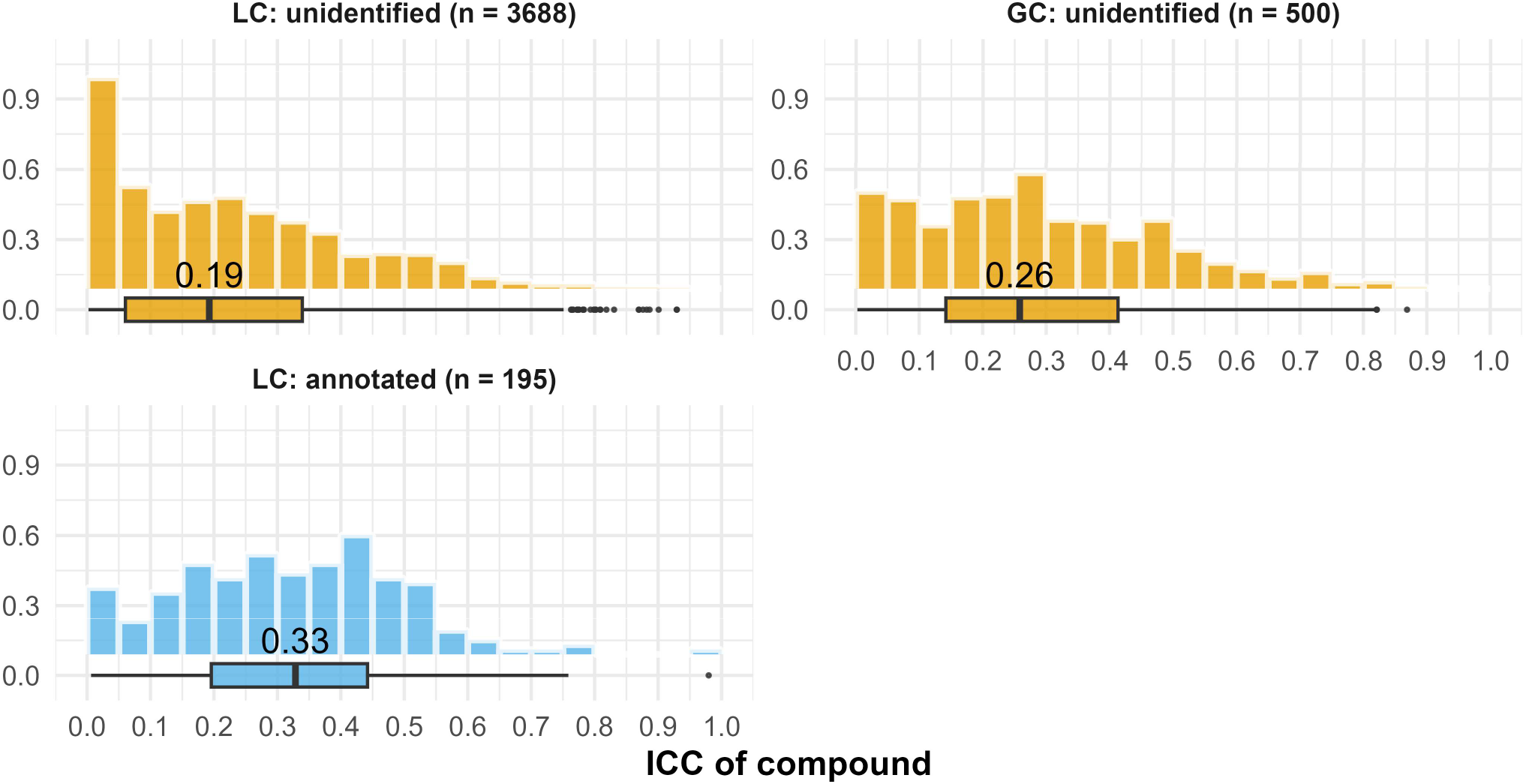
temporal repeatability features per platform and annotation status. Annotation was not on feature level for the GC platform. A random sample of 500 features was taken for the GC features. ICC = intraclass correlation coefficient; GC = Gas Chromatography. LC = Liquid Chromatography.

**Figure S7a:**
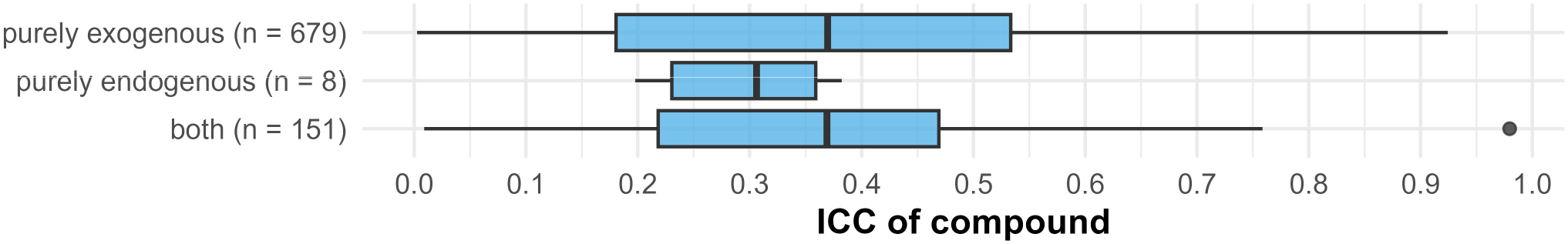
distribution ICC values across endogenous, exogenous categories for the annotated clusters / clusters with an annotation (both platforms). ICC = intraclass correlation coefficient.

**Figure S7b:**
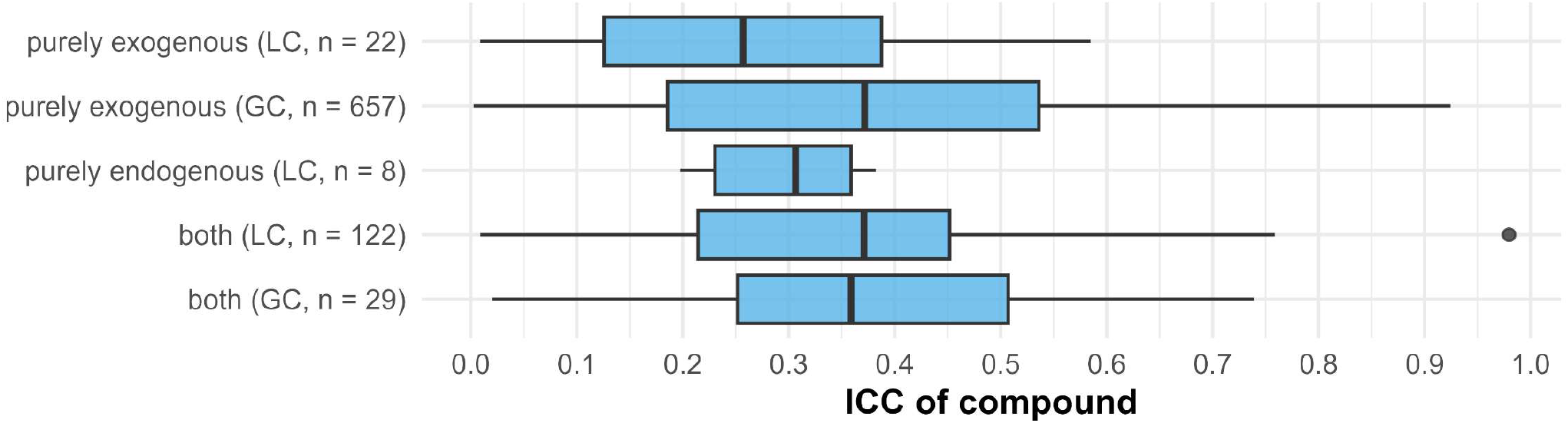
distribution ICC values across endogenous, exogenous categories for the annotated clusters / clusters with an annotation. Stratified per platforms. ICC = intraclass correlation coefficient. GC = Gas Chromatography. LC = Liquid Chromatography.

**Figure S8:**
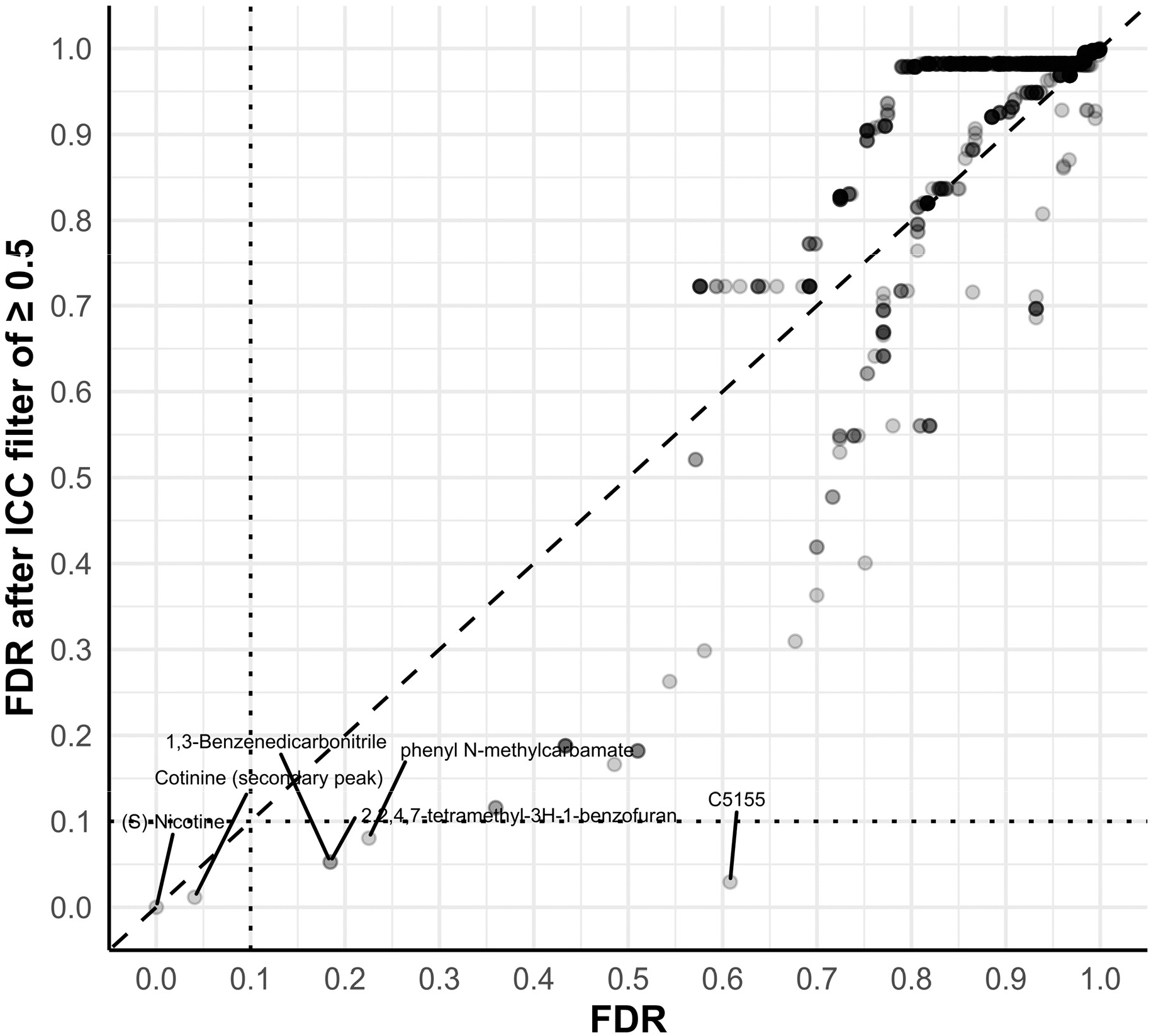
a scatter plot of the FDR per metabolite in both the main (pooled) analysis from the paper and the analysis after excluding metabolites with an ICC < 0.5 before controlling the FDR. The dotted line indicates the FDR at 10%. The dashed line is the ab line where the FDR for a metabolite is identical in both analyses. FDR = false discovery rate; ICC = intraclass correlation coefficient.

**Table S4:**
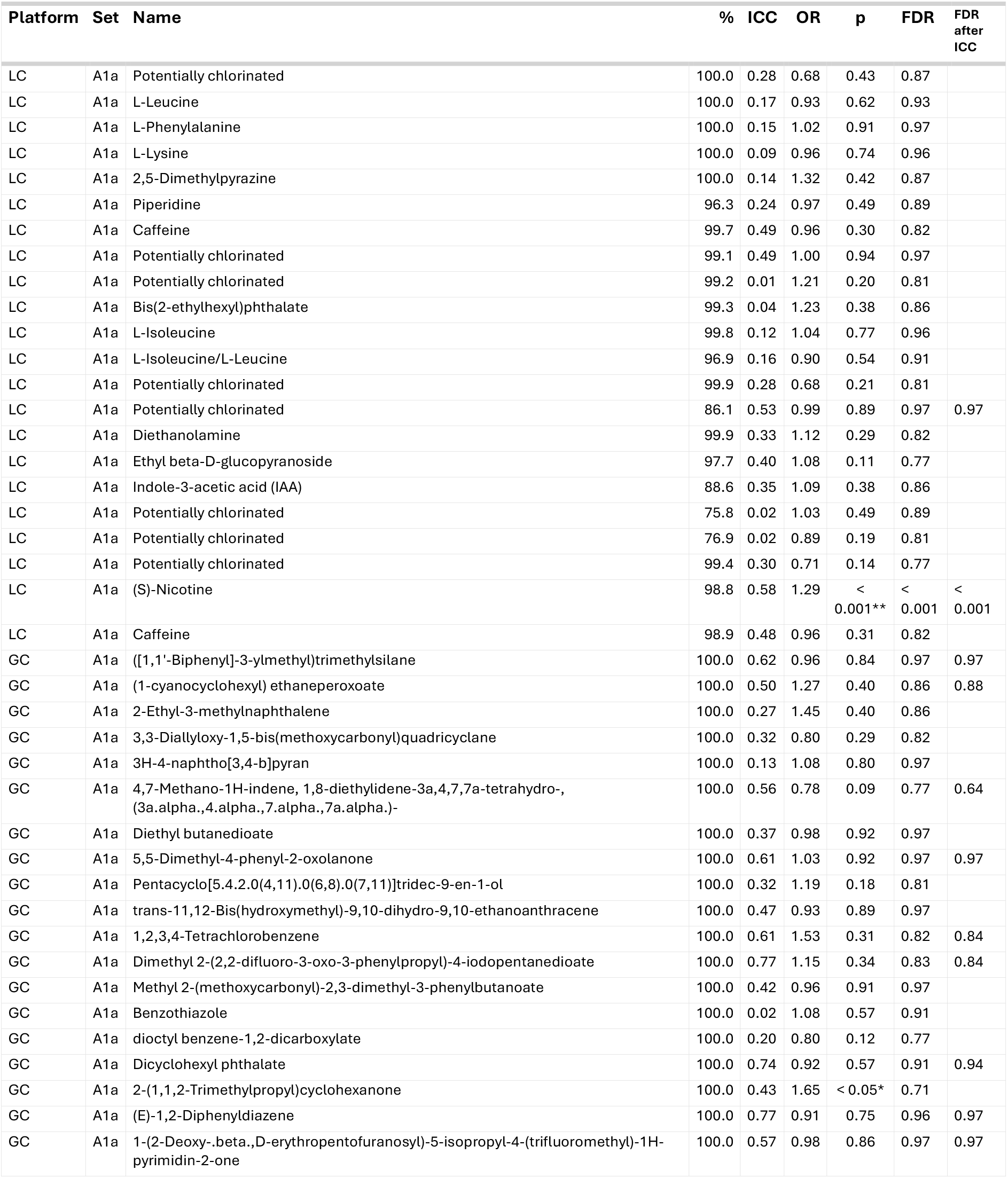
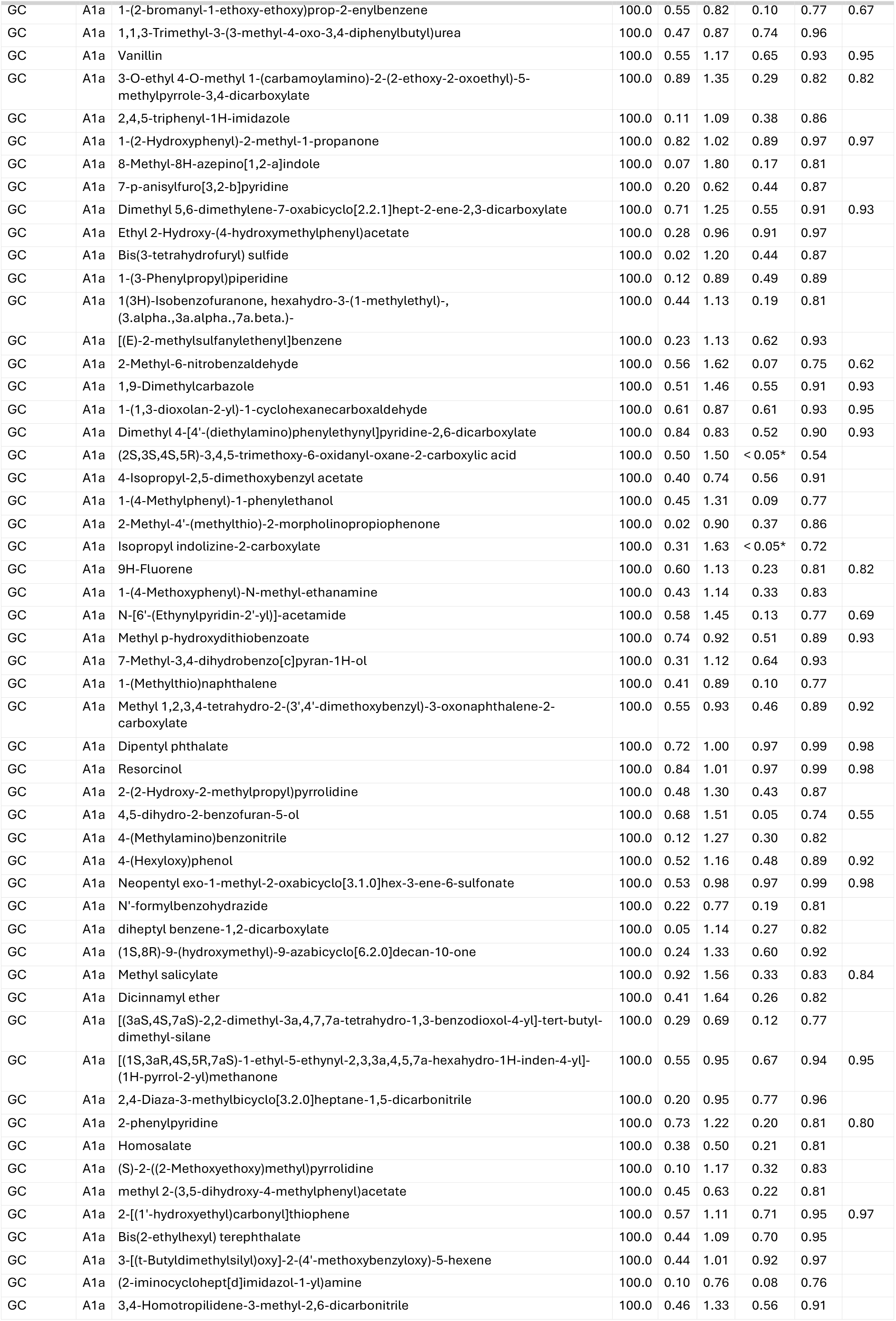
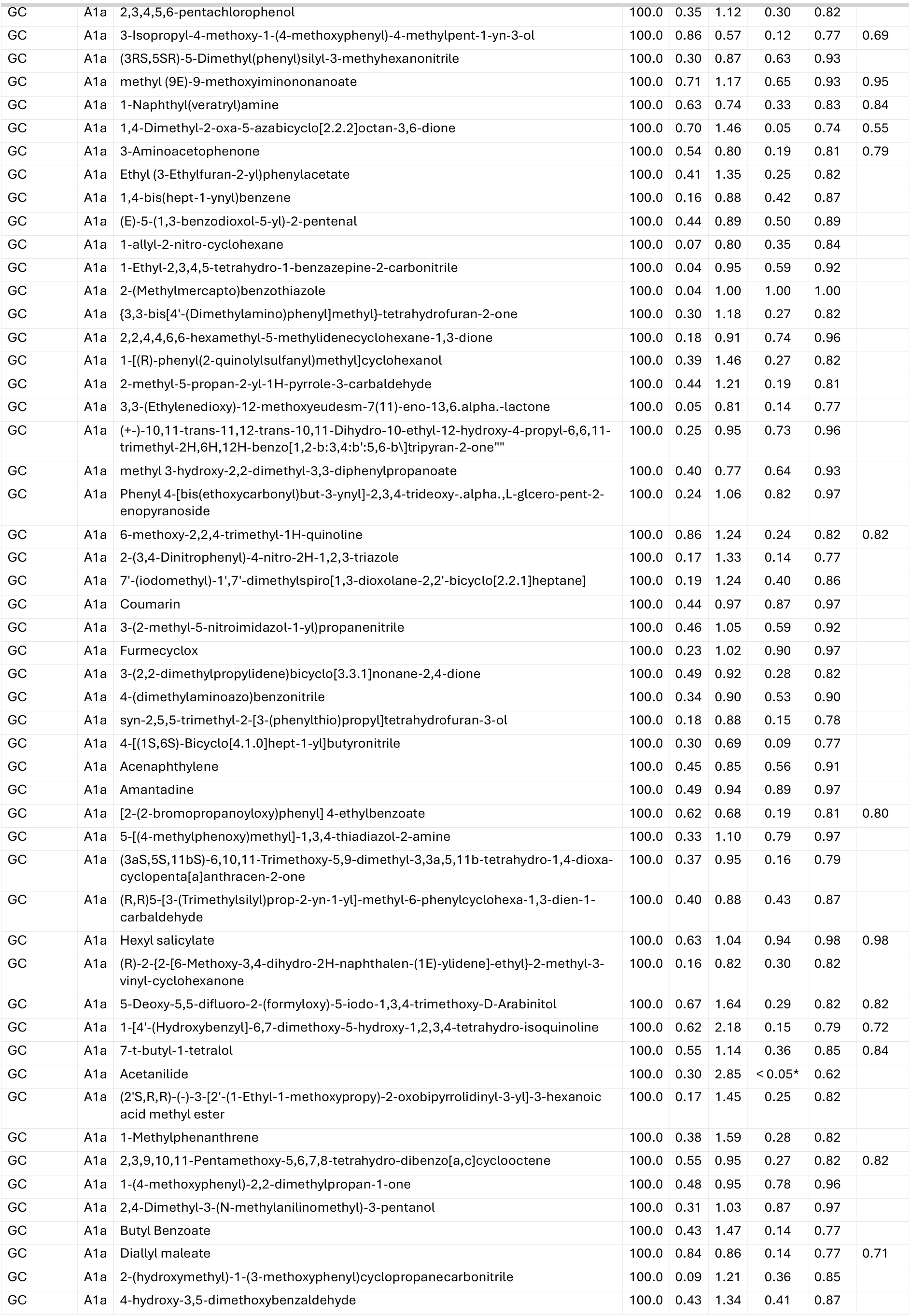
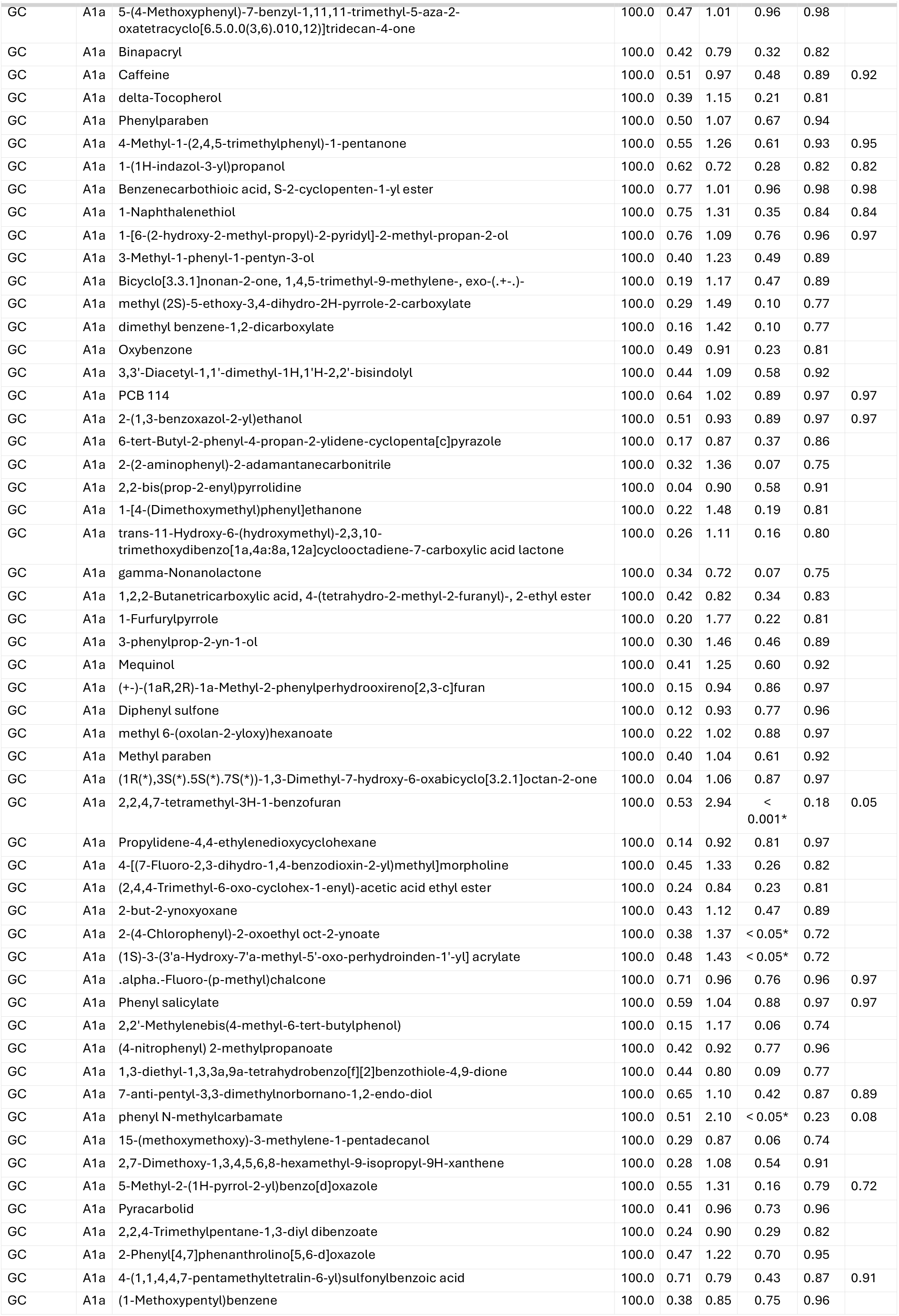
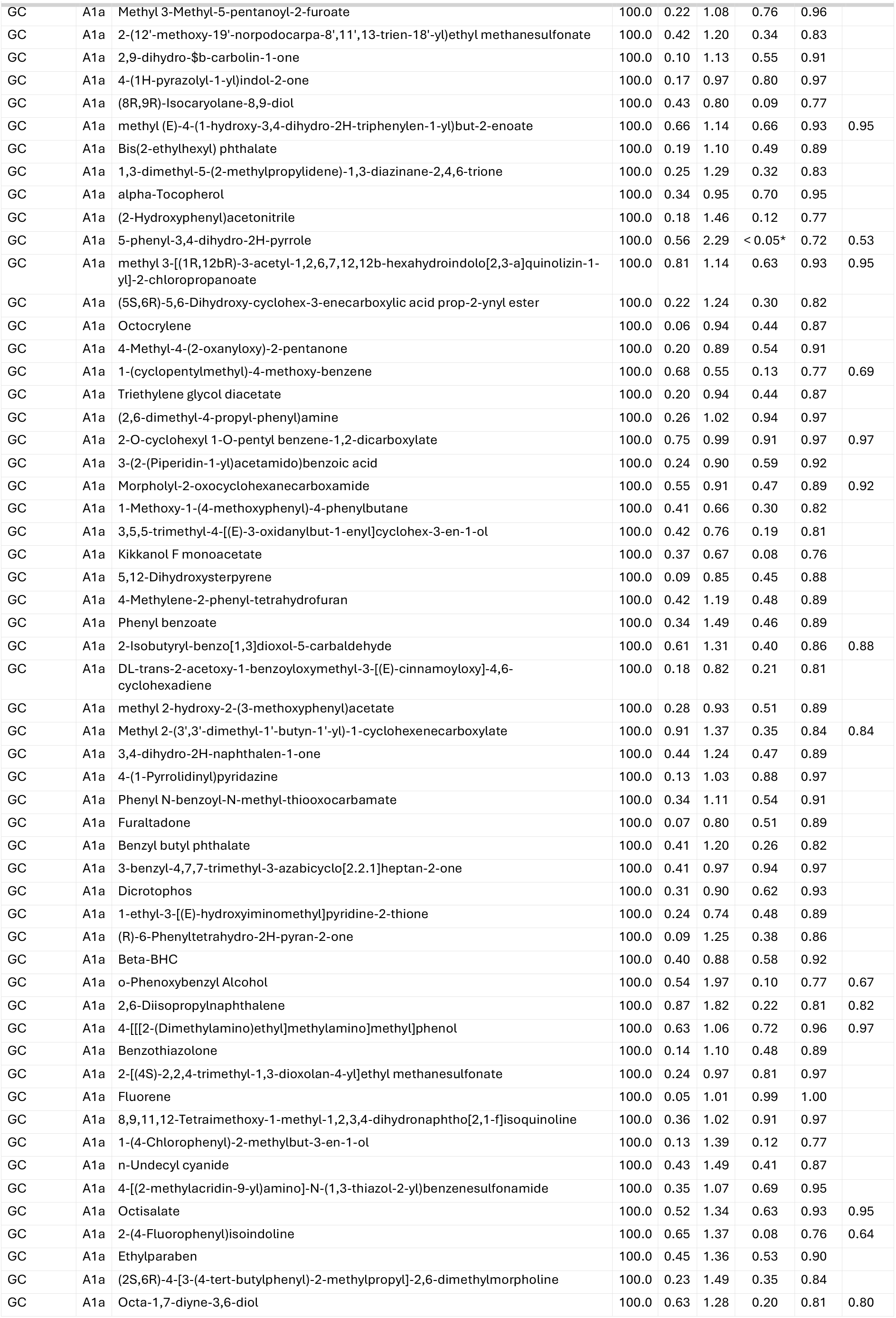
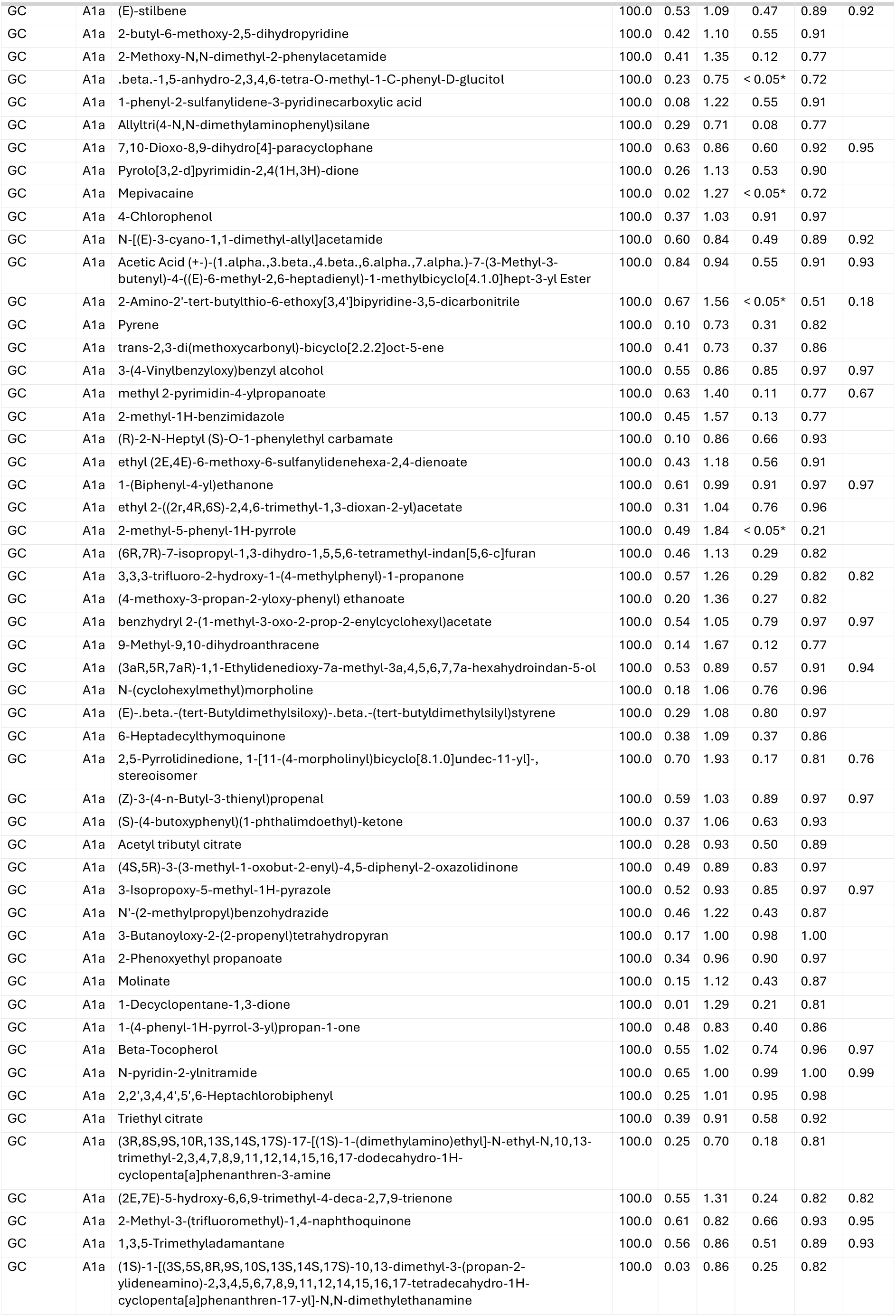
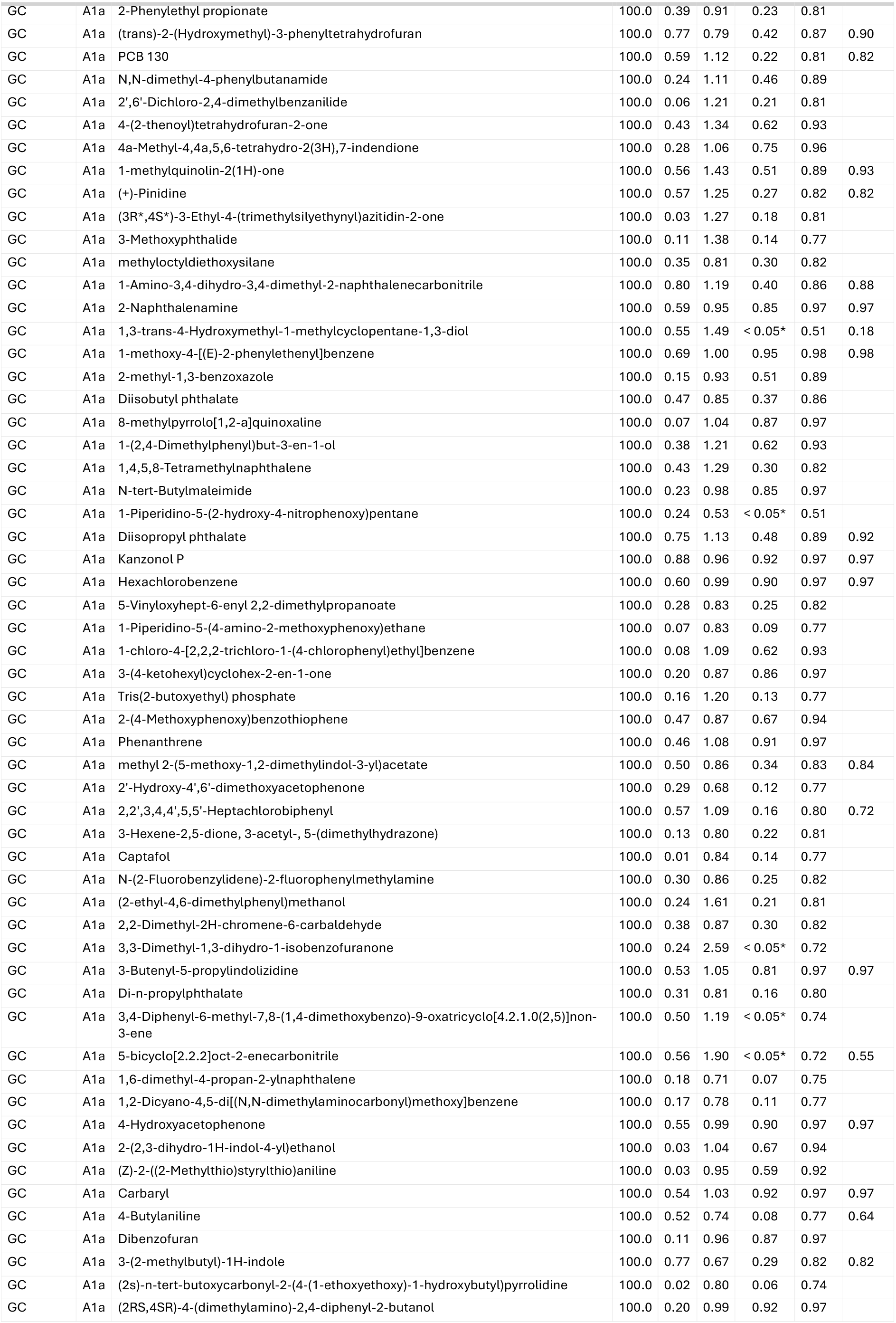
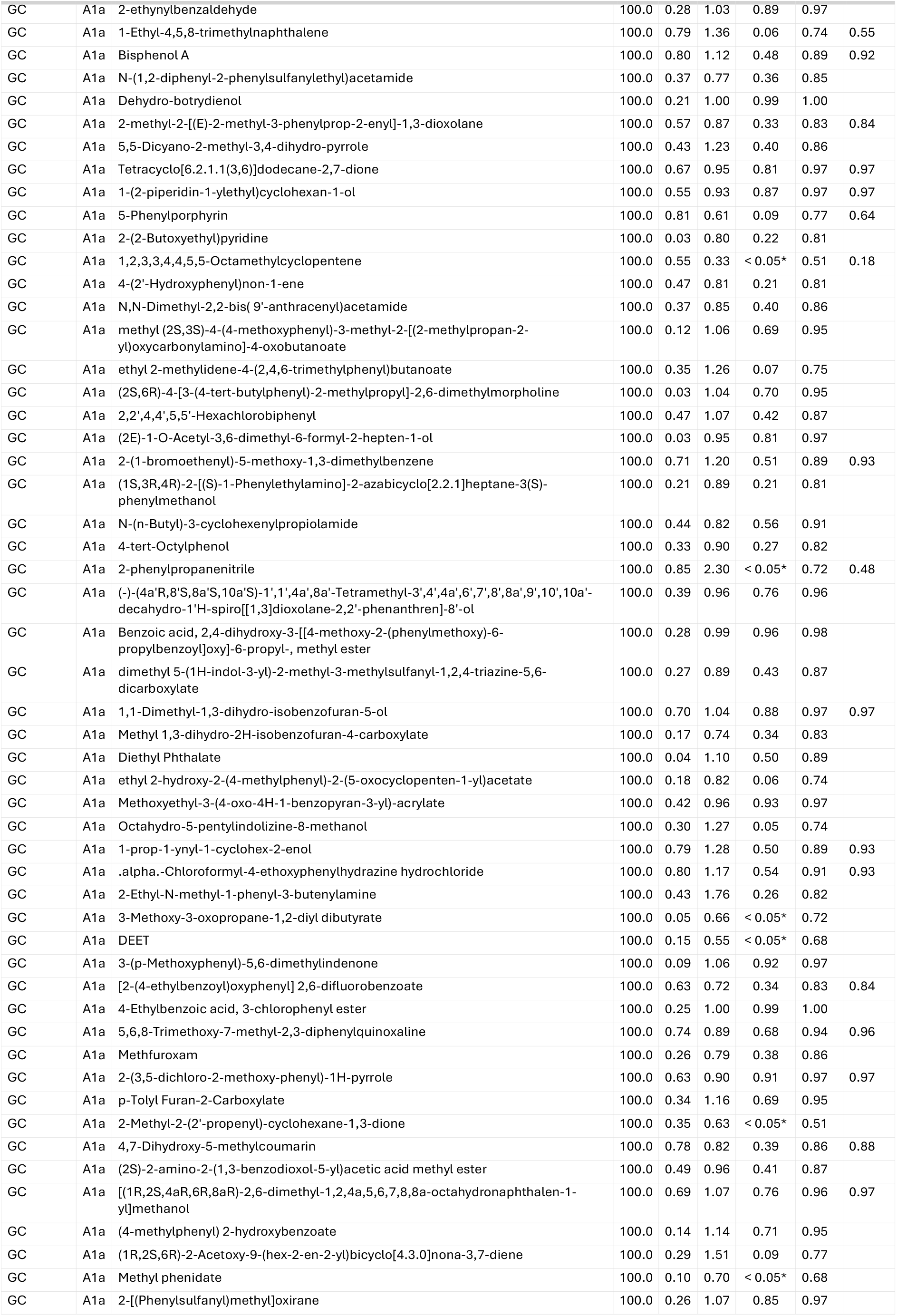
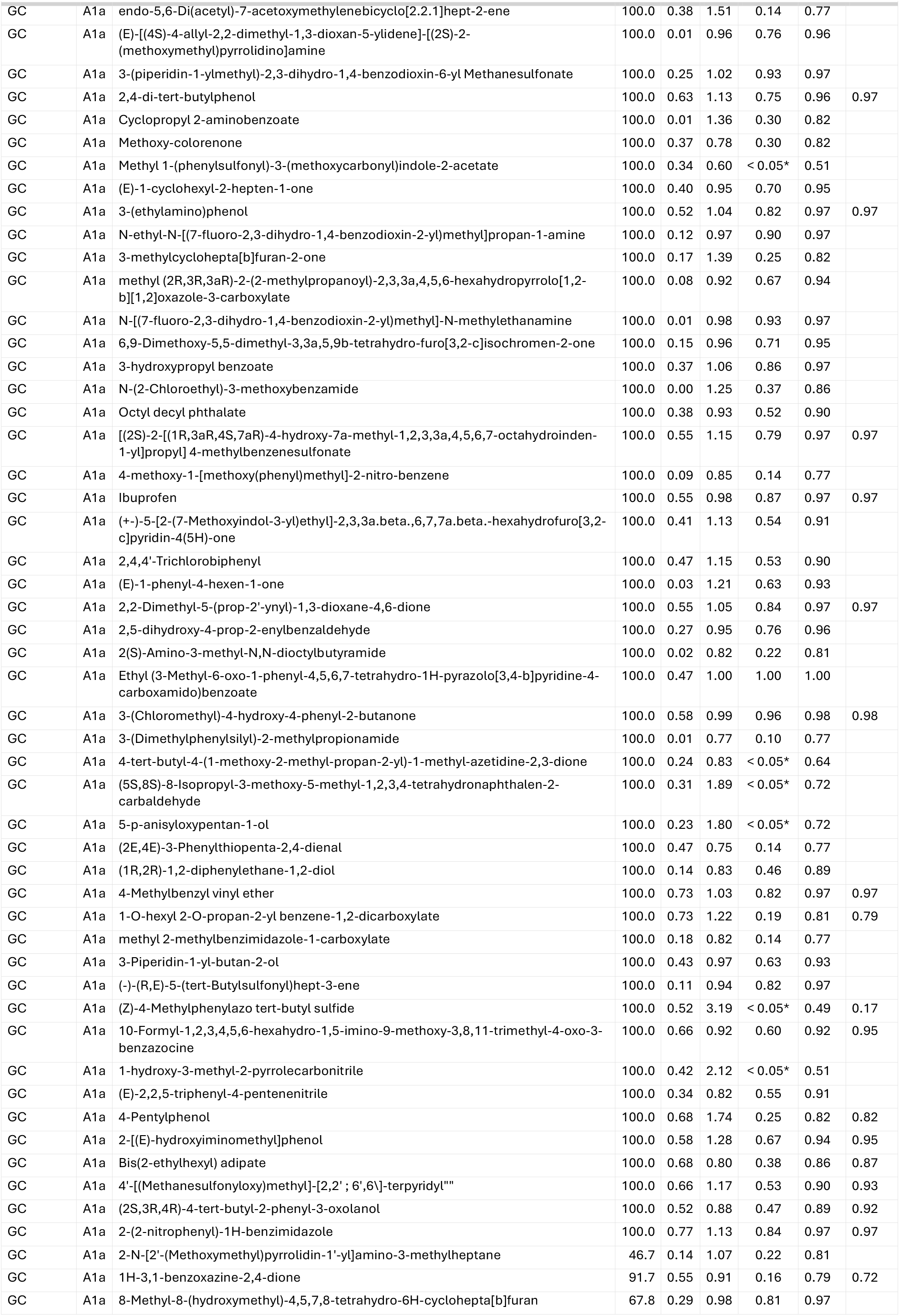
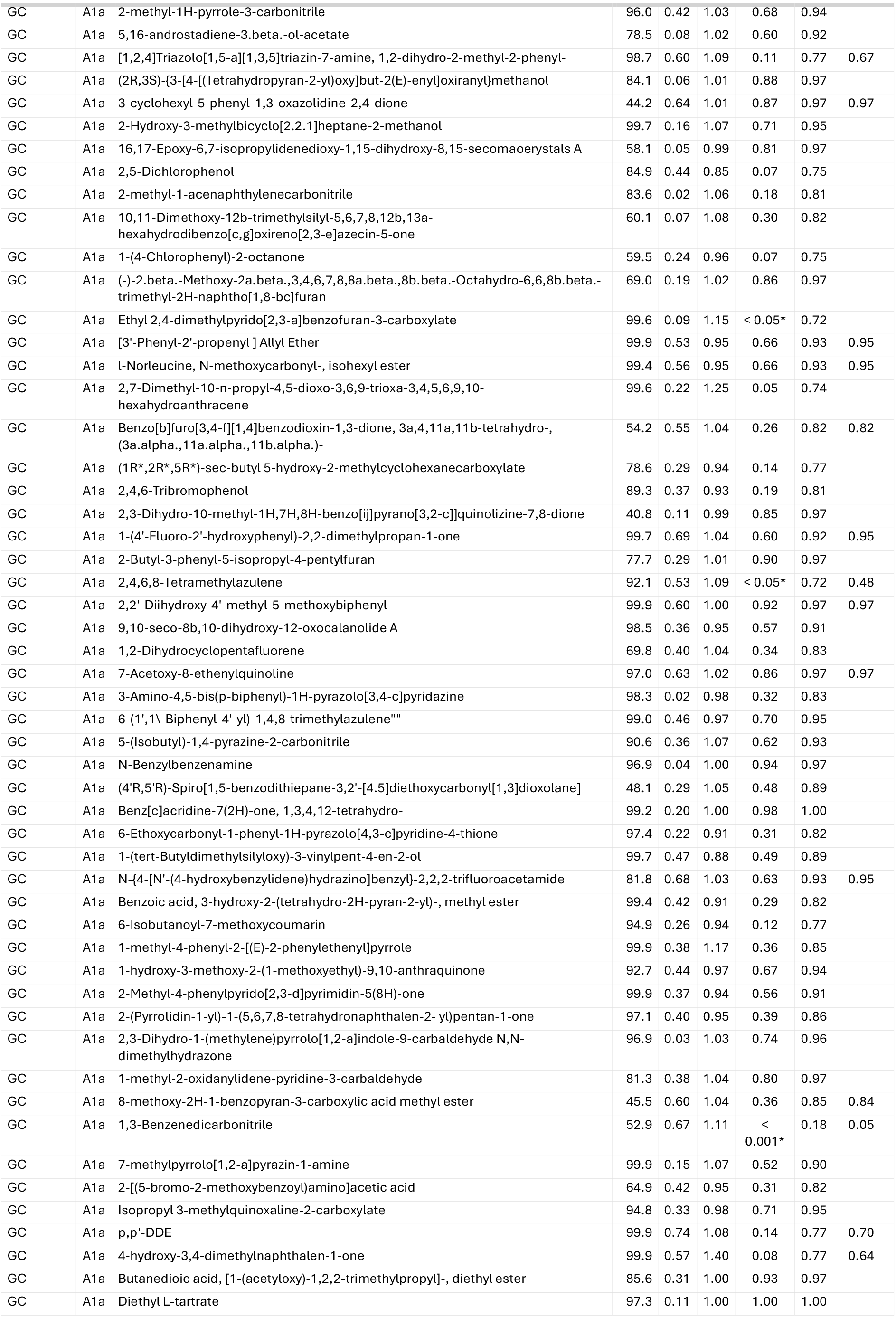
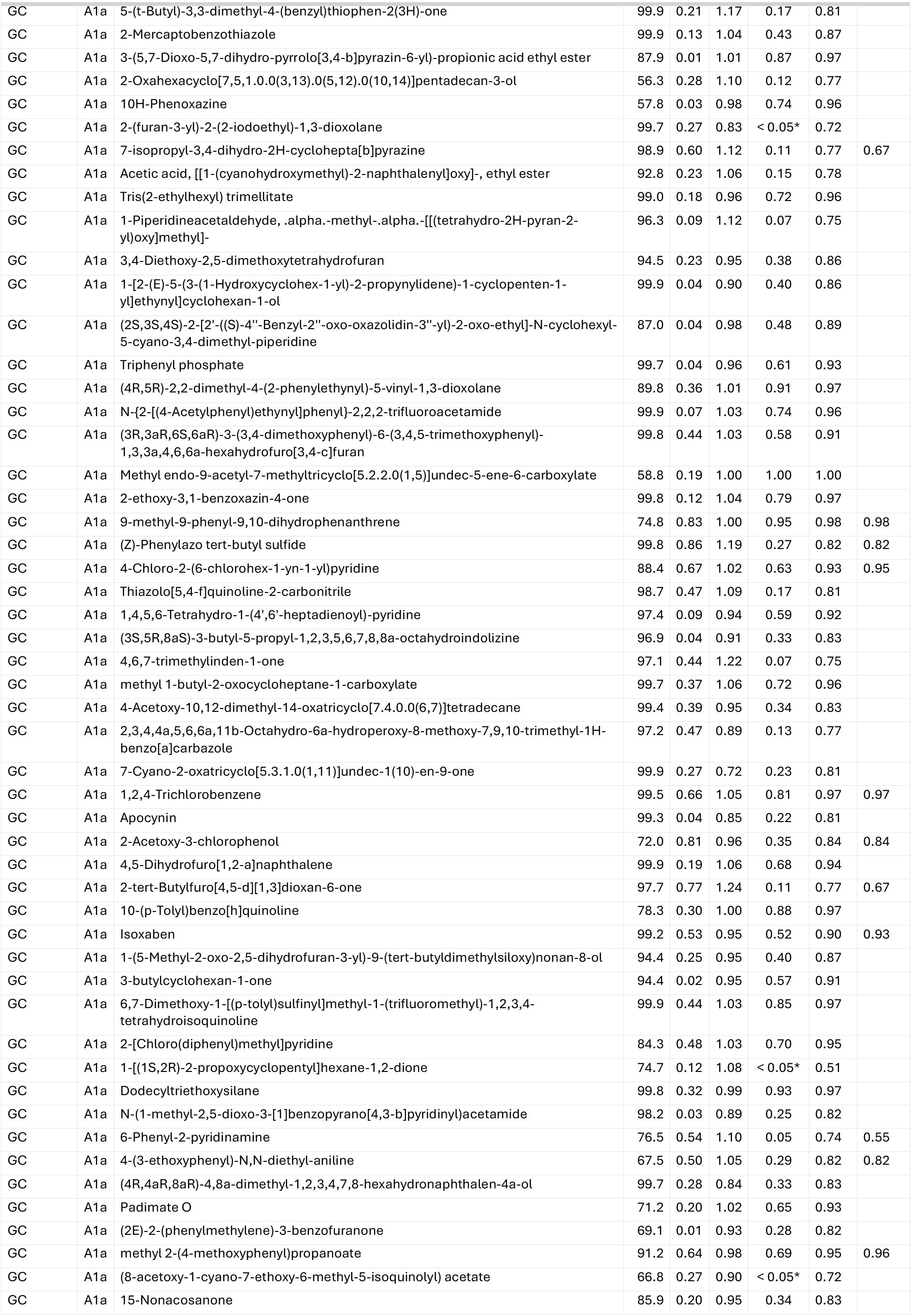
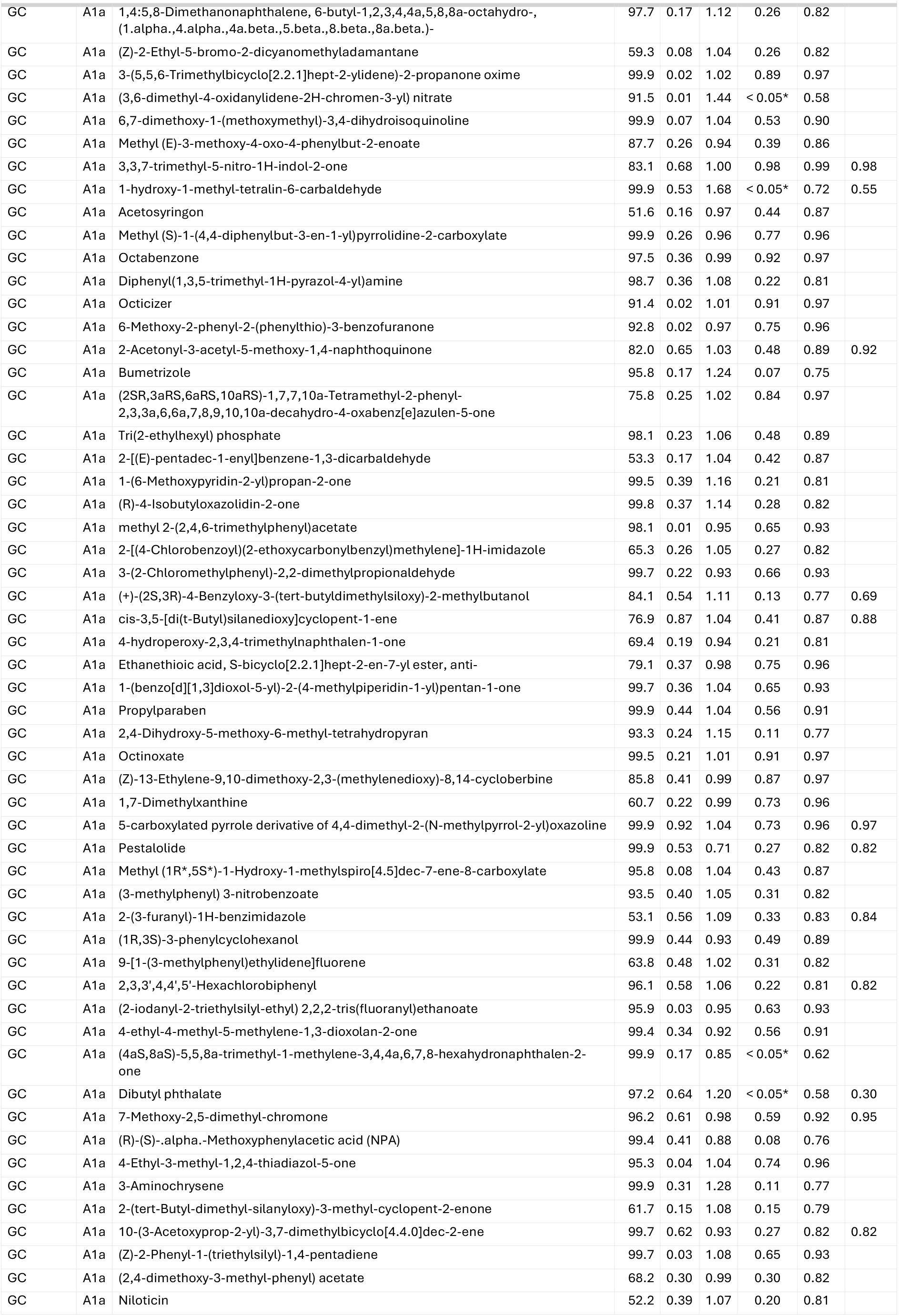
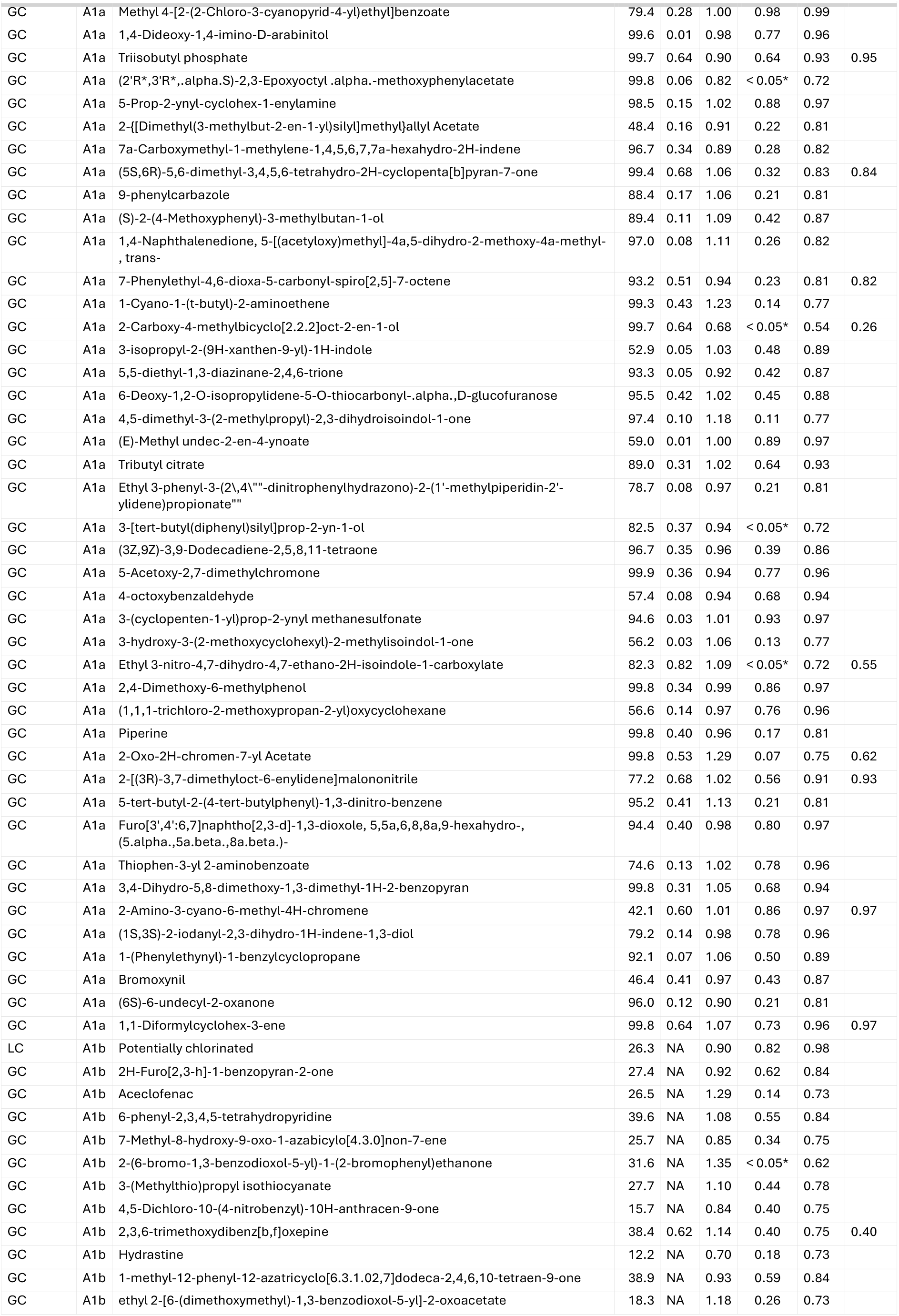
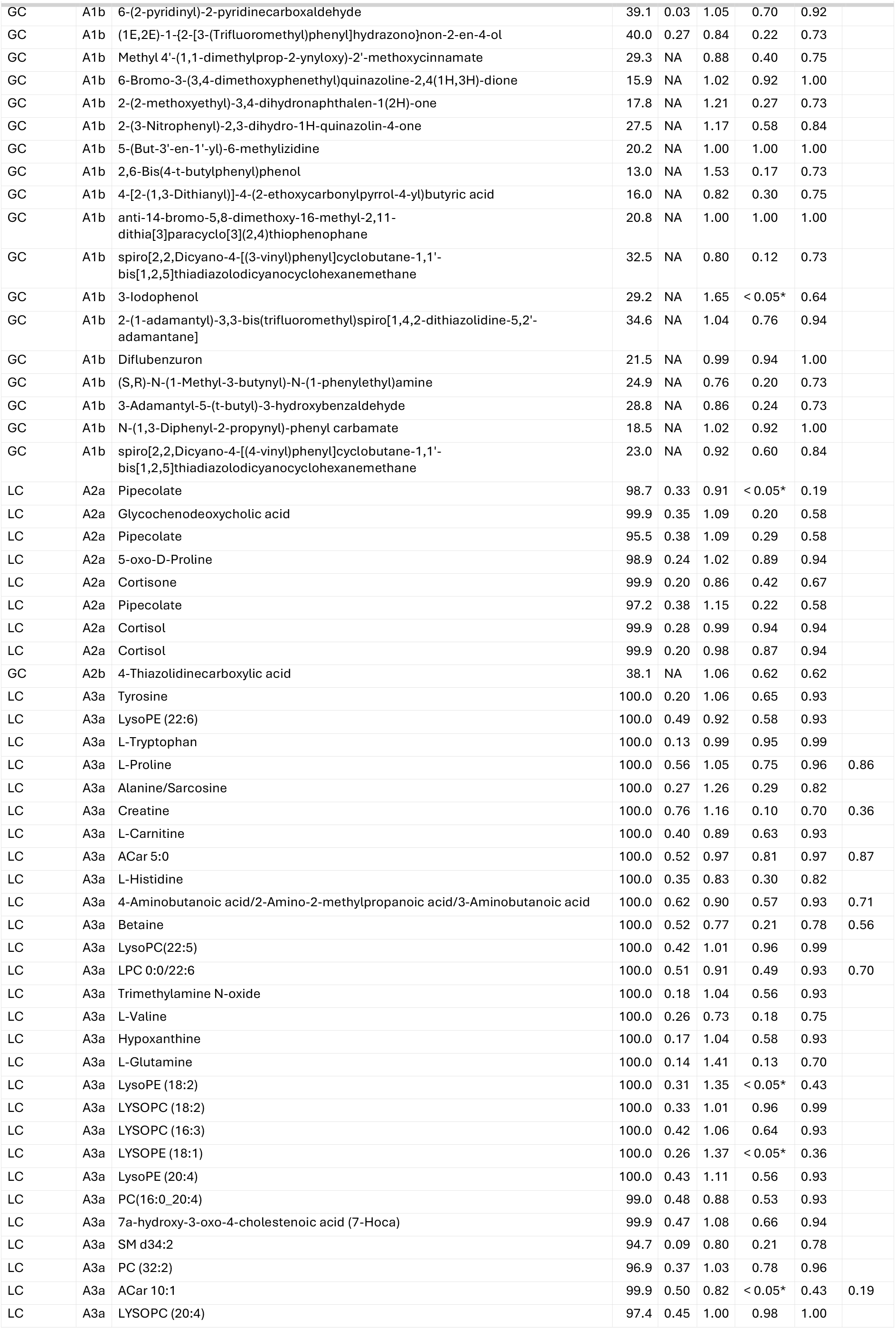
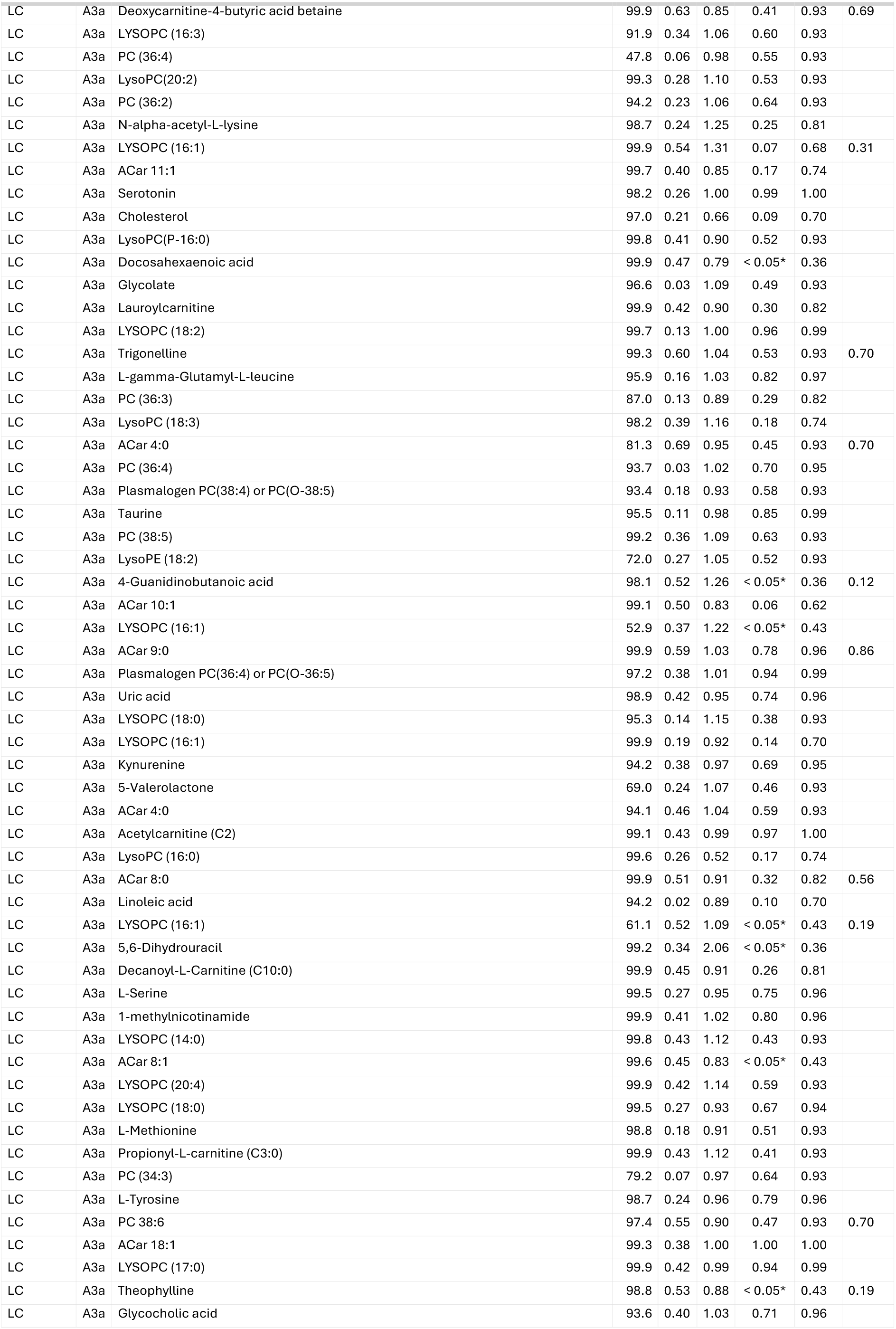
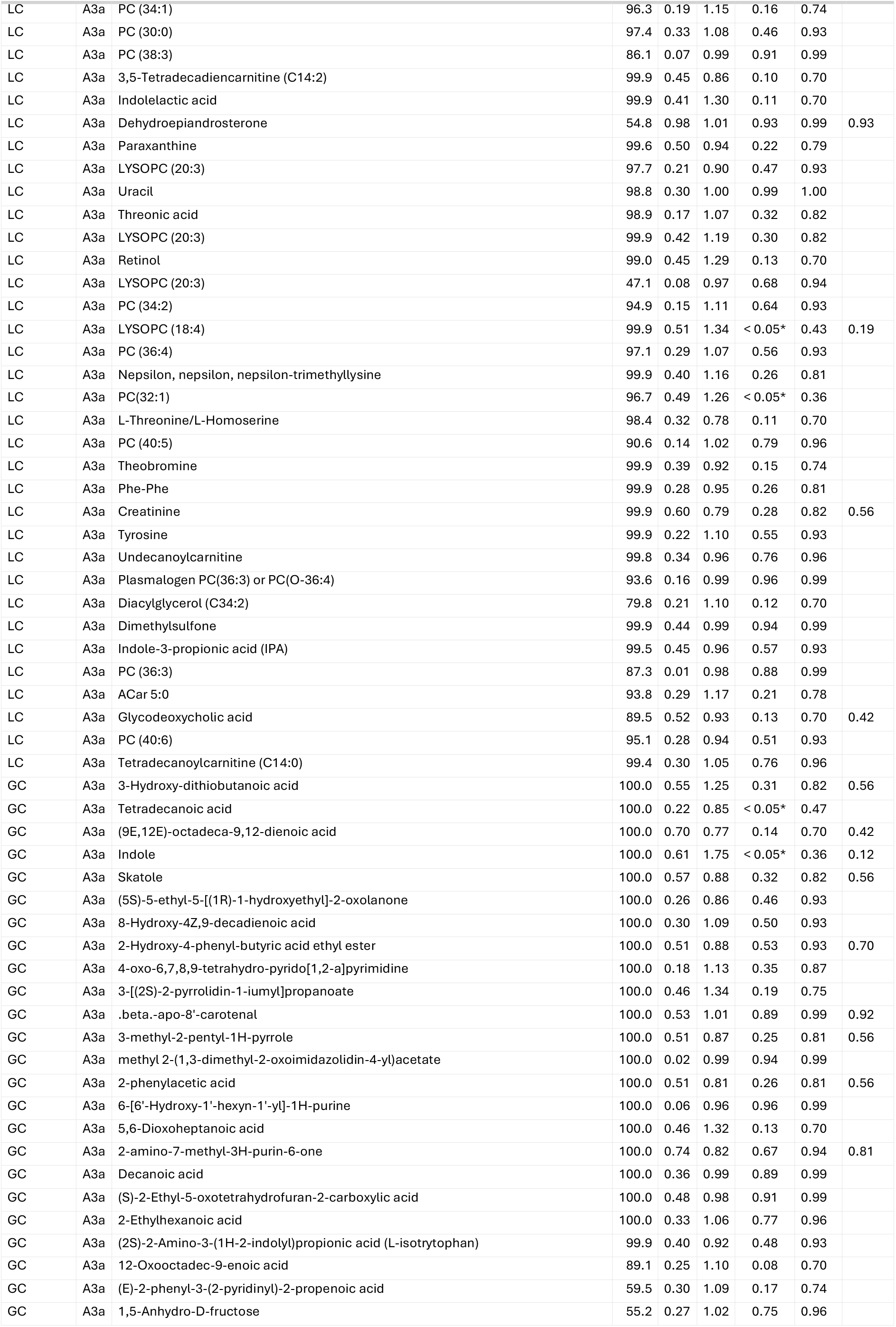
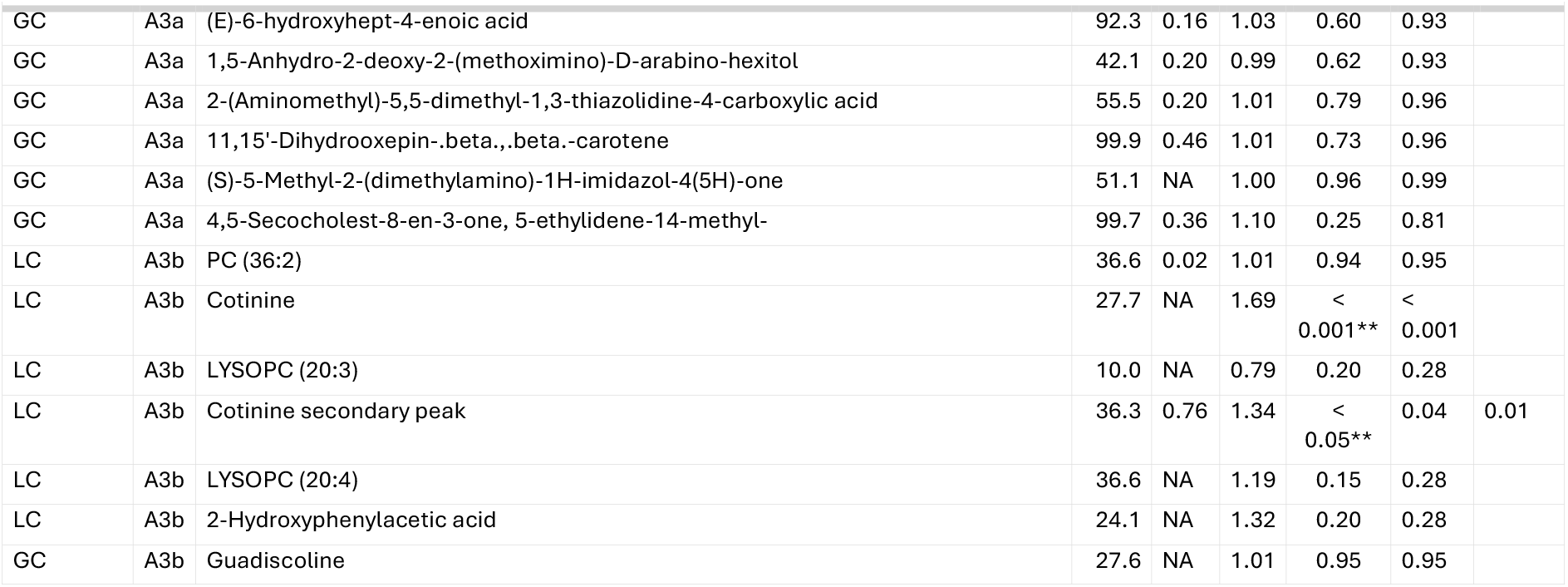
results for the initially annotated peaks in the main (pooled) analysis and the ICC restricted analysis. NA in the ICC column means that the feature was excluded from the ICC calculation because it was not present in sufficient samples of the Doetinchem Cohort Study. Set A describes the annotated metabolites, Set B indicates the initially unidentified features/clusters. 1, 2, 3 describes the exogenous, endogenous, both endogenous/exogenously derived metabolites respectively. The last letter indicates if the feature was analyzed as continuous variable (lower case ‘a’) or as binary variable of detected, not detected (lower case ‘b’). % = proportion of samples case-control study feature was detected (note: not proportion of Doetinchem Cohort Study samples where feature was detected (used for inclusion ICC analysis)); ICC = intraclass correlation coefficient; OR = odds ratio; p = p-value; FDR = false discovery rate; FDR after ICC = resulting FDR when controlling FDR after excluding features with ICC below 0.5.

| Platform | Set | Name | % | ICC | OR | p | FDR | FDR after ICC |
|---|---|---|---|---|---|---|---|---|
| LC | A1a | Potentially chlorinated | 100.0 | 0.28 | 0.68 | 0.43 | 0.87 |  |
| LC | A1a | L-Leucine | 100.0 | 0.17 | 0.93 | 0.62 | 0.93 |  |
| LC | A1a | L-Phenylalanine | 100.0 | 0.15 | 1.02 | 0.91 | 0.97 |  |
| LC | A1a | L-Lysine | 100.0 | 0.09 | 0.96 | 0.74 | 0.96 |  |
| LC | A1a | 2,5-Dimethylpyrazine | 100.0 | 0.14 | 1.32 | 0.42 | 0.87 |  |
| LC | A1a | Piperidine | 96.3 | 0.24 | 0.97 | 0.49 | 0.89 |  |
| LC | A1a | Caffeine | 99.7 | 0.49 | 0.96 | 0.30 | 0.82 |  |
| LC | A1a | Potentially chlorinated | 99.1 | 0.49 | 1.00 | 0.94 | 0.97 |  |
| LC | A1a | Potentially chlorinated | 99.2 | 0.01 | 1.21 | 0.20 | 0.81 |  |
| LC | A1a | Bis(2-ethylhexyl)phthalate | 99.3 | 0.04 | 1.23 | 0.38 | 0.86 |  |
| LC | A1a | L-Isoleucine | 99.8 | 0.12 | 1.04 | 0.77 | 0.96 |  |
| LC | A1a | L-Isoleucine/L-Leucine | 96.9 | 0.16 | 0.90 | 0.54 | 0.91 |  |
| LC | A1a | Potentially chlorinated | 99.9 | 0.28 | 0.68 | 0.21 | 0.81 |  |
| LC | A1a | Potentially chlorinated | 86.1 | 0.53 | 0.99 | 0.89 | 0.97 | 0.97 |
| LC | A1a | Diethanolamine | 99.9 | 0.33 | 1.12 | 0.29 | 0.82 |  |
| LC | A1a | Ethyl beta-D-glucopyranoside | 97.7 | 0.40 | 1.08 | 0.11 | 0.77 |  |
| LC | A1a | Indole-3-acetic acid (IAA) | 88.6 | 0.35 | 1.09 | 0.38 | 0.86 |  |
| LC | A1a | Potentially chlorinated | 75.8 | 0.02 | 1.03 | 0.49 | 0.89 |  |
| LC | A1a | Potentially chlorinated | 76.9 | 0.02 | 0.89 | 0.19 | 0.81 |  |
| LC | A1a | Potentially chlorinated | 99.4 | 0.30 | 0.71 | 0.14 | 0.77 |  |
| LC | A1a | (S)-Nicotine | 98.8 | 0.58 | 1.29 | < 0.001** | < 0.001 | < 0.001 |
| LC | A1a | Caffeine | 98.9 | 0.48 | 0.96 | 0.31 | 0.82 |  |
| GC | A1a | ([1,1'-Biphenyl]-3-ylmethyl)trimethylsilane | 100.0 | 0.62 | 0.96 | 0.84 | 0.97 | 0.97 |
| GC | A1a | (1-cyanocyclohexyl) ethaneperoxoate | 100.0 | 0.50 | 1.27 | 0.40 | 0.86 | 0.88 |
| GC | A1a | 2-Ethyl-3-methylnaphthalene | 100.0 | 0.27 | 1.45 | 0.40 | 0.86 |  |
| GC | A1a | 3,3-Diallyloxy-1,5-bis(methoxycarbonyl)quadricyclane | 100.0 | 0.32 | 0.80 | 0.29 | 0.82 |  |
| GC | A1a | 3H-4-naphtho[3,4-b]pyran | 100.0 | 0.13 | 1.08 | 0.80 | 0.97 |  |
| GC | A1a | 4,7-Methano-1H-indene, 1,8-diethylidene-3a,4,7,7a-tetrahydro-, (3a.alpha.,4.alpha.,7.alpha.,7a.alpha.)- | 100.0 | 0.56 | 0.78 | 0.09 | 0.77 | 0.64 |
| GC | A1a | Diethyl butanedioate | 100.0 | 0.37 | 0.98 | 0.92 | 0.97 |  |
| GC | A1a | 5,5-Dimethyl-4-phenyl-2-oxolanone | 100.0 | 0.61 | 1.03 | 0.92 | 0.97 | 0.97 |
| GC | A1a | Pentacyclo[5.4.2.0(4,11).0(6,8).0(7,11)]tridec-9-en-1-ol | 100.0 | 0.32 | 1.19 | 0.18 | 0.81 |  |
| GC | A1a | trans-11,12-Bis(hydroxymethyl)-9,10-dihydro-9,10-ethanoanthracene | 100.0 | 0.47 | 0.93 | 0.89 | 0.97 |  |
| GC | A1a | 1,2,3,4-Tetrachlorobenzene | 100.0 | 0.61 | 1.53 | 0.31 | 0.82 | 0.84 |
| GC | A1a | Dimethyl 2-(2,2-difluoro-3-oxo-3-phenylpropyl)-4-iodopentanedioate | 100.0 | 0.77 | 1.15 | 0.34 | 0.83 | 0.84 |
| GC | A1a | Methyl 2-(methoxycarbonyl)-2,3-dimethyl-3-phenylbutanoate | 100.0 | 0.42 | 0.96 | 0.91 | 0.97 |  |
| GC | A1a | Benzothiazole | 100.0 | 0.02 | 1.08 | 0.57 | 0.91 |  |
| GC | A1a | dioctyl benzene-1,2-dicarboxylate | 100.0 | 0.20 | 0.80 | 0.12 | 0.77 |  |
| GC | A1a | Dicyclohexyl phthalate | 100.0 | 0.74 | 0.92 | 0.57 | 0.91 | 0.94 |
| GC | A1a | 2-(1,1,2-Trimethylpropyl)cyclohexanone | 100.0 | 0.43 | 1.65 | < 0.05* | 0.71 |  |
| GC | A1a | (E)-1,2-Diphenyldiazene | 100.0 | 0.77 | 0.91 | 0.75 | 0.96 | 0.97 |
| GC | A1a | 1-(2-Deoxy-.beta.,D-erythropentofuranosyl)-5-isopropyl-4-(trifluoromethyl)-1H-pyrimidin-2-one | 100.0 | 0.57 | 0.98 | 0.86 | 0.97 | 0.97 |
| GC | A1a | 1-(2-bromanyl-1-ethoxy-ethoxy)prop-2-enylbenzene | 100.0 | 0.55 | 0.82 | 0.10 | 0.77 | 0.67 |
| GC | A1a | 1,1,3-Trimethyl-3-(3-methyl-4-oxo-3,4-diphenylbutyl)urea | 100.0 | 0.47 | 0.87 | 0.74 | 0.96 |  |
| GC | A1a | Vanillin | 100.0 | 0.55 | 1.17 | 0.65 | 0.93 | 0.95 |
| GC | A1a | 3-O-ethyl 4-O-methyl 1-(carbamoylamino)-2-(2-ethoxy-2-oxoethyl)-5-methylpyrrole-3,4-dicarboxylate | 100.0 | 0.89 | 1.35 | 0.29 | 0.82 | 0.82 |
| GC | A1a | 2,4,5-triphenyl-1H-imidazole | 100.0 | 0.11 | 1.09 | 0.38 | 0.86 |  |
| GC | A1a | 1-(2-Hydroxyphenyl)-2-methyl-1-propanone | 100.0 | 0.82 | 1.02 | 0.89 | 0.97 | 0.97 |
| GC | A1a | 8-Methyl-8H-azepino[1,2-a]indole | 100.0 | 0.07 | 1.80 | 0.17 | 0.81 |  |
| GC | A1a | 7-p-anisylfuro[3,2-b]pyridine | 100.0 | 0.20 | 0.62 | 0.44 | 0.87 |  |
| GC | A1a | Dimethyl 5,6-dimethylene-7-oxabicyclo[2.2.1]hept-2-ene-2,3-dicarboxylate | 100.0 | 0.71 | 1.25 | 0.55 | 0.91 | 0.93 |
| GC | A1a | Ethyl 2-Hydroxy-(4-hydroxymethylphenyl)acetate | 100.0 | 0.28 | 0.96 | 0.91 | 0.97 |  |
| GC | A1a | Bis(3-tetrahydrofuryl) sulfide | 100.0 | 0.02 | 1.20 | 0.44 | 0.87 |  |
| GC | A1a | 1-(3-Phenylpropyl)piperidine | 100.0 | 0.12 | 0.89 | 0.49 | 0.89 |  |
| GC | A1a | 1(3H)-Isobenzofuranone, hexahydro-3-(1-methylethyl)-, (3.alpha.,3a.alpha.,7a.beta.)- | 100.0 | 0.44 | 1.13 | 0.19 | 0.81 |  |
| GC | A1a | [(E)-2-methylsulfanylethenyl]benzene | 100.0 | 0.23 | 1.13 | 0.62 | 0.93 |  |
| GC | A1a | 2-Methyl-6-nitrobenzaldehyde | 100.0 | 0.56 | 1.62 | 0.07 | 0.75 | 0.62 |
| GC | A1a | 1,9-Dimethylcarbazole | 100.0 | 0.51 | 1.46 | 0.55 | 0.91 | 0.93 |
| GC | A1a | 1-(1,3-dioxolan-2-yl)-1-cyclohexanecarboxaldehyde | 100.0 | 0.61 | 0.87 | 0.61 | 0.93 | 0.95 |
| GC | A1a | Dimethyl 4-[4'-(diethylamino)phenylethynyl]pyridine-2,6-dicarboxylate | 100.0 | 0.84 | 0.83 | 0.52 | 0.90 | 0.93 |
| GC | A1a | (2S,3S,4S,5R)-3,4,5-trimethoxy-6-oxidanyl-oxane-2-carboxylic acid | 100.0 | 0.50 | 1.50 | < 0.05* | 0.54 |  |
| GC | A1a | 4-Isopropyl-2,5-dimethoxybenzyl acetate | 100.0 | 0.40 | 0.74 | 0.56 | 0.91 |  |
| GC | A1a | 1-(4-Methylphenyl)-1-phenylethanol | 100.0 | 0.45 | 1.31 | 0.09 | 0.77 |  |
| GC | A1a | 2-Methyl-4'-(methylthio)-2-morpholinopropiophenone | 100.0 | 0.02 | 0.90 | 0.37 | 0.86 |  |
| GC | A1a | Isopropyl indolizine-2-carboxylate | 100.0 | 0.31 | 1.63 | < 0.05* | 0.72 |  |
| GC | A1a | 9H-Fluorene | 100.0 | 0.60 | 1.13 | 0.23 | 0.81 | 0.82 |
| GC | A1a | 1-(4-Methoxyphenyl)-N-methyl-ethanamine | 100.0 | 0.43 | 1.14 | 0.33 | 0.83 |  |
| GC | A1a | N-[6'-(Ethynylpyridin-2'-yl)]-acetamide | 100.0 | 0.58 | 1.45 | 0.13 | 0.77 | 0.69 |
| GC | A1a | Methyl p-hydroxydithiobenzoate | 100.0 | 0.74 | 0.92 | 0.51 | 0.89 | 0.93 |
| GC | A1a | 7-Methyl-3,4-dihydrobenzo[c]pyran-1H-ol | 100.0 | 0.31 | 1.12 | 0.64 | 0.93 |  |
| GC | A1a | 1-(Methylthio)naphthalene | 100.0 | 0.41 | 0.89 | 0.10 | 0.77 |  |
| GC | A1a | Methyl 1,2,3,4-tetrahydro-2-(3',4'-dimethoxybenzyl)-3-oxonaphthalene-2-carboxylate | 100.0 | 0.55 | 0.93 | 0.46 | 0.89 | 0.92 |
| GC | A1a | Dipentyl phthalate | 100.0 | 0.72 | 1.00 | 0.97 | 0.99 | 0.98 |
| GC | A1a | Resorcinol | 100.0 | 0.84 | 1.01 | 0.97 | 0.99 | 0.98 |
| GC | A1a | 2-(2-Hydroxy-2-methylpropyl)pyrrolidine | 100.0 | 0.48 | 1.30 | 0.43 | 0.87 |  |
| GC | A1a | 4,5-dihydro-2-benzofuran-5-ol | 100.0 | 0.68 | 1.51 | 0.05 | 0.74 | 0.55 |
| GC | A1a | 4-(Methylamino)benzonitrile | 100.0 | 0.12 | 1.27 | 0.30 | 0.82 |  |
| GC | A1a | 4-(Hexyloxy)phenol | 100.0 | 0.52 | 1.16 | 0.48 | 0.89 | 0.92 |
| GC | A1a | Neopentyl exo-1-methyl-2-oxabicyclo[3.1.0]hex-3-ene-6-sulfonate | 100.0 | 0.53 | 0.98 | 0.97 | 0.99 | 0.98 |
| GC | A1a | N'-formylbenzohydrazide | 100.0 | 0.22 | 0.77 | 0.19 | 0.81 |  |
| GC | A1a | diheptyl benzene-1,2-dicarboxylate | 100.0 | 0.05 | 1.14 | 0.27 | 0.82 |  |
| GC | A1a | (1S,8R)-9-(hydroxymethyl)-9-azabicyclo[6.2.0]decan-10-one | 100.0 | 0.24 | 1.33 | 0.60 | 0.92 |  |
| GC | A1a | Methyl salicylate | 100.0 | 0.92 | 1.56 | 0.33 | 0.83 | 0.84 |
| GC | A1a | Dicinnamyl ether | 100.0 | 0.41 | 1.64 | 0.26 | 0.82 |  |
| GC | A1a | [(3aS,4S,7aS)-2,2-dimethyl-3a,4,7,7a-tetrahydro-1,3-benzodioxol-4-yl]-tert-butyl-dimethyl-silane | 100.0 | 0.29 | 0.69 | 0.12 | 0.77 |  |
| GC | A1a | [(1S,3aR,4S,5R,7aS)-1-ethyl-5-ethynyl-2,3,3a,4,5,7a-hexahydro-1H-inden-4-yl]-(1H-pyrrol-2-yl)methanone | 100.0 | 0.55 | 0.95 | 0.67 | 0.94 | 0.95 |
| GC | A1a | 2,4-Diaza-3-methylbicyclo[3.2.0]heptane-1,5-dicarbonitrile | 100.0 | 0.20 | 0.95 | 0.77 | 0.96 |  |
| GC | A1a | 2-phenylpyridine | 100.0 | 0.73 | 1.22 | 0.20 | 0.81 | 0.80 |
| GC | A1a | Homosalate | 100.0 | 0.38 | 0.50 | 0.21 | 0.81 |  |
| GC | A1a | (S)-2-((2-Methoxyethoxy)methyl)pyrrolidine | 100.0 | 0.10 | 1.17 | 0.32 | 0.83 |  |
| GC | A1a | methyl 2-(3,5-dihydroxy-4-methylphenyl)acetate | 100.0 | 0.45 | 0.63 | 0.22 | 0.81 |  |
| GC | A1a | 2-[(1'-hydroxyethyl)carbonyl]thiophene | 100.0 | 0.57 | 1.11 | 0.71 | 0.95 | 0.97 |
| GC | A1a | Bis(2-ethylhexyl) terephthalate | 100.0 | 0.44 | 1.09 | 0.70 | 0.95 |  |
| GC | A1a | 3-[(t-Butyldimethylsilyl)oxy]-2-(4'-methoxybenzyloxy)-5-hexene | 100.0 | 0.44 | 1.01 | 0.92 | 0.97 |  |
| GC | A1a | (2-iminocyclohept[d]imidazol-1-yl)amine | 100.0 | 0.10 | 0.76 | 0.08 | 0.76 |  |
| GC | A1a | 3,4-Homotropilidene-3-methyl-2,6-dicarbonitrile | 100.0 | 0.46 | 1.33 | 0.56 | 0.91 |  |
| GC | A1a | 2,3,4,5,6-pentachlorophenol | 100.0 | 0.35 | 1.12 | 0.30 | 0.82 |  |
| GC | A1a | 3-Isopropyl-4-methoxy-1-(4-methoxyphenyl)-4-methylpent-1-yn-3-ol | 100.0 | 0.86 | 0.57 | 0.12 | 0.77 | 0.69 |
| GC | A1a | (3RS,5SR)-5-Dimethyl(phenyl)silyl-3-methyhexanonitrile | 100.0 | 0.30 | 0.87 | 0.63 | 0.93 |  |
| GC | A1a | methyl (9E)-9-methoxyiminononanoate | 100.0 | 0.71 | 1.17 | 0.65 | 0.93 | 0.95 |
| GC | A1a | 1-Naphthyl(veratryl)amine | 100.0 | 0.63 | 0.74 | 0.33 | 0.83 | 0.84 |
| GC | A1a | 1,4-Dimethyl-2-oxa-5-azabicyclo[2.2.2]octan-3,6-dione | 100.0 | 0.70 | 1.46 | 0.05 | 0.74 | 0.55 |
| GC | A1a | 3-Aminoacetophenone | 100.0 | 0.54 | 0.80 | 0.19 | 0.81 | 0.79 |
| GC | A1a | Ethyl (3-Ethylfuran-2-yl)phenylacetate | 100.0 | 0.41 | 1.35 | 0.25 | 0.82 |  |
| GC | A1a | 1,4-bis(hept-1-ynyl)benzene | 100.0 | 0.16 | 0.88 | 0.42 | 0.87 |  |
| GC | A1a | (E)-5-(1,3-benzodioxol-5-yl)-2-pentenal | 100.0 | 0.44 | 0.89 | 0.50 | 0.89 |  |
| GC | A1a | 1-allyl-2-nitro-cyclohexane | 100.0 | 0.07 | 0.80 | 0.35 | 0.84 |  |
| GC | A1a | 1-Ethyl-2,3,4,5-tetrahydro-1-benzazepine-2-carbonitrile | 100.0 | 0.04 | 0.95 | 0.59 | 0.92 |  |
| GC | A1a | 2-(Methylmercapto)benzothiazole | 100.0 | 0.04 | 1.00 | 1.00 | 1.00 |  |
| GC | A1a | {3,3-bis[4'-(Dimethylamino)phenyl]methyl}-tetrahydrofuran-2-one | 100.0 | 0.30 | 1.18 | 0.27 | 0.82 |  |
| GC | A1a | 2,2,4,4,6,6-hexamethyl-5-methylidenecyclohexane-1,3-dione | 100.0 | 0.18 | 0.91 | 0.74 | 0.96 |  |
| GC | A1a | 1-[(R)-phenyl(2-quinolylsulfanyl)methyl]cyclohexanol | 100.0 | 0.39 | 1.46 | 0.27 | 0.82 |  |
| GC | A1a | 2-methyl-5-propan-2-yl-1H-pyrrole-3-carbaldehyde | 100.0 | 0.44 | 1.21 | 0.19 | 0.81 |  |
| GC | A1a | 3,3-(Ethylenedioxy)-12-methoxyeudesm-7(11)-eno-13,6.alpha.-lactone | 100.0 | 0.05 | 0.81 | 0.14 | 0.77 |  |
| GC | A1a | (+-)-10,11-trans-11,12-trans-10,11-Dihydro-10-ethyl-12-hydroxy-4-propyl-6,6,11-trimethyl-2H,6H,12H-benzo[1,2-b:3,4:b':5,6-b\]tripyran-2-one"" | 100.0 | 0.25 | 0.95 | 0.73 | 0.96 |  |
| GC | A1a | methyl 3-hydroxy-2,2-dimethyl-3,3-diphenylpropanoate | 100.0 | 0.40 | 0.77 | 0.64 | 0.93 |  |
| GC | A1a | Phenyl 4-[bis(ethoxycarbonyl)but-3-ynyl]-2,3,4-trideoxy-.alpha.,L-glcero-pent-2-enopyranoside | 100.0 | 0.24 | 1.06 | 0.82 | 0.97 |  |
| GC | A1a | 6-methoxy-2,2,4-trimethyl-1H-quinoline | 100.0 | 0.86 | 1.24 | 0.24 | 0.82 | 0.82 |
| GC | A1a | 2-(3,4-Dinitrophenyl)-4-nitro-2H-1,2,3-triazole | 100.0 | 0.17 | 1.33 | 0.14 | 0.77 |  |
| GC | A1a | 7'-(iodomethyl)-1',7'-dimethylspiro[1,3-dioxolane-2,2'-bicyclo[2.2.1]heptane] | 100.0 | 0.19 | 1.24 | 0.40 | 0.86 |  |
| GC | A1a | Coumarin | 100.0 | 0.44 | 0.97 | 0.87 | 0.97 |  |
| GC | A1a | 3-(2-methyl-5-nitroimidazol-1-yl)propanenitrile | 100.0 | 0.46 | 1.05 | 0.59 | 0.92 |  |
| GC | A1a | Furmecyclox | 100.0 | 0.23 | 1.02 | 0.90 | 0.97 |  |
| GC | A1a | 3-(2,2-dimethylpropylidene)bicyclo[3.3.1]nonane-2,4-dione | 100.0 | 0.49 | 0.92 | 0.28 | 0.82 |  |
| GC | A1a | 4-(dimethylaminoazo)benzonitrile | 100.0 | 0.34 | 0.90 | 0.53 | 0.90 |  |
| GC | A1a | syn-2,5,5-trimethyl-2-[3-(phenylthio)propyl]tetrahydrofuran-3-ol | 100.0 | 0.18 | 0.88 | 0.15 | 0.78 |  |
| GC | A1a | 4-[(1S,6S)-Bicyclo[4.1.0]hept-1-yl]butyronitrile | 100.0 | 0.30 | 0.69 | 0.09 | 0.77 |  |
| GC | A1a | Acenaphthylene | 100.0 | 0.45 | 0.85 | 0.56 | 0.91 |  |
| GC | A1a | Amantadine | 100.0 | 0.49 | 0.94 | 0.89 | 0.97 |  |
| GC | A1a | [2-(2-bromopropanoyloxy)phenyl] 4-ethylbenzoate | 100.0 | 0.62 | 0.68 | 0.19 | 0.81 | 0.80 |
| GC | A1a | 5-[(4-methylphenoxy)methyl]-1,3,4-thiadiazol-2-amine | 100.0 | 0.33 | 1.10 | 0.79 | 0.97 |  |
| GC | A1a | (3aS,5S,11bS)-6,10,11-Trimethoxy-5,9-dimethyl-3,3a,5,11b-tetrahydro-1,4-dioxa-cyclopenta[a]anthracen-2-one | 100.0 | 0.37 | 0.95 | 0.16 | 0.79 |  |
| GC | A1a | (R,R)5-[3-(Trimethylsilyl)prop-2-yn-1-yl]-methyl-6-phenylcyclohexa-1,3-dien-1-carbaldehyde | 100.0 | 0.40 | 0.88 | 0.43 | 0.87 |  |
| GC | A1a | Hexyl salicylate | 100.0 | 0.63 | 1.04 | 0.94 | 0.98 | 0.98 |
| GC | A1a | (R)-2-{2-[6-Methoxy-3,4-dihydro-2H-naphthalen-(1E)-ylidene]-ethyl}-2-methyl-3-vinyl-cyclohexanone | 100.0 | 0.16 | 0.82 | 0.30 | 0.82 |  |
| GC | A1a | 5-Deoxy-5,5-difluoro-2-(formyloxy)-5-iodo-1,3,4-trimethoxy-D-Arabinitol | 100.0 | 0.67 | 1.64 | 0.29 | 0.82 | 0.82 |
| GC | A1a | 1-[4'-(Hydroxybenzyl]-6,7-dimethoxy-5-hydroxy-1,2,3,4-tetrahydro-isoquinoline | 100.0 | 0.62 | 2.18 | 0.15 | 0.79 | 0.72 |
| GC | A1a | 7-t-butyl-1-tetralol | 100.0 | 0.55 | 1.14 | 0.36 | 0.85 | 0.84 |
| GC | A1a | Acetanilide | 100.0 | 0.30 | 2.85 | < 0.05* | 0.62 |  |
| GC | A1a | (2'S,R,R)-(-)-3-[2'-(1-Ethyl-1-methoxypropy)-2-oxobipyrrolidinyl-3-yl]-3-hexanoic acid methyl ester | 100.0 | 0.17 | 1.45 | 0.25 | 0.82 |  |
| GC | A1a | 1-Methylphenanthrene | 100.0 | 0.38 | 1.59 | 0.28 | 0.82 |  |
| GC | A1a | 2,3,9,10,11-Pentamethoxy-5,6,7,8-tetrahydro-dibenzo[a,c]cyclooctene | 100.0 | 0.55 | 0.95 | 0.27 | 0.82 | 0.82 |
| GC | A1a | 1-(4-methoxyphenyl)-2,2-dimethylpropan-1-one | 100.0 | 0.48 | 0.95 | 0.78 | 0.96 |  |
| GC | A1a | 2,4-Dimethyl-3-(N-methylanilinomethyl)-3-pentanol | 100.0 | 0.31 | 1.03 | 0.87 | 0.97 |  |
| GC | A1a | Butyl Benzoate | 100.0 | 0.43 | 1.47 | 0.14 | 0.77 |  |
| GC | A1a | Diallyl maleate | 100.0 | 0.84 | 0.86 | 0.14 | 0.77 | 0.71 |
| GC | A1a | 2-(hydroxymethyl)-1-(3-methoxyphenyl)cyclopropanecarbonitrile | 100.0 | 0.09 | 1.21 | 0.36 | 0.85 |  |
| GC | A1a | 4-hydroxy-3,5-dimethoxybenzaldehyde | 100.0 | 0.43 | 1.34 | 0.41 | 0.87 |  |
| GC | A1a | 5-(4-Methoxyphenyl)-7-benzyl-1,11,11-trimethyl-5-aza-2-oxatetracyclo[6.5.0.0(3,6).010,12)]tridecan-4-one | 100.0 | 0.47 | 1.01 | 0.96 | 0.98 |  |
| GC | A1a | Binapacryl | 100.0 | 0.42 | 0.79 | 0.32 | 0.82 |  |
| GC | A1a | Caffeine | 100.0 | 0.51 | 0.97 | 0.48 | 0.89 | 0.92 |
| GC | A1a | delta-Tocopherol | 100.0 | 0.39 | 1.15 | 0.21 | 0.81 |  |
| GC | A1a | Phenylparaben | 100.0 | 0.50 | 1.07 | 0.67 | 0.94 |  |
| GC | A1a | 4-Methyl-1-(2,4,5-trimethylphenyl)-1-pentanone | 100.0 | 0.55 | 1.26 | 0.61 | 0.93 | 0.95 |
| GC | A1a | 1-(1H-indazol-3-yl)propanol | 100.0 | 0.62 | 0.72 | 0.28 | 0.82 | 0.82 |
| GC | A1a | Benzenecarbothioic acid, S-2-cyclopenten-1-yl ester | 100.0 | 0.77 | 1.01 | 0.96 | 0.98 | 0.98 |
| GC | A1a | 1-Naphthalenethiol | 100.0 | 0.75 | 1.31 | 0.35 | 0.84 | 0.84 |
| GC | A1a | 1-[6-(2-hydroxy-2-methyl-propyl)-2-pyridyl]-2-methyl-propan-2-ol | 100.0 | 0.76 | 1.09 | 0.76 | 0.96 | 0.97 |
| GC | A1a | 3-Methyl-1-phenyl-1-pentyn-3-ol | 100.0 | 0.40 | 1.23 | 0.49 | 0.89 |  |
| GC | A1a | Bicyclo[3.3.1]nonan-2-one, 1,4,5-trimethyl-9-methylene-, exo-(.+-.)- | 100.0 | 0.19 | 1.17 | 0.47 | 0.89 |  |
| GC | A1a | methyl (2S)-5-ethoxy-3,4-dihydro-2H-pyrrole-2-carboxylate | 100.0 | 0.29 | 1.49 | 0.10 | 0.77 |  |
| GC | A1a | dimethyl benzene-1,2-dicarboxylate | 100.0 | 0.16 | 1.42 | 0.10 | 0.77 |  |
| GC | A1a | Oxybenzone | 100.0 | 0.49 | 0.91 | 0.23 | 0.81 |  |
| GC | A1a | 3,3'-Diacetyl-1,1'-dimethyl-1H,1'H-2,2'-bisindolyl | 100.0 | 0.44 | 1.09 | 0.58 | 0.92 |  |
| GC | A1a | PCB 114 | 100.0 | 0.64 | 1.02 | 0.89 | 0.97 | 0.97 |
| GC | A1a | 2-(1,3-benzoxazol-2-yl)ethanol | 100.0 | 0.51 | 0.93 | 0.89 | 0.97 | 0.97 |
| GC | A1a | 6-tert-Butyl-2-phenyl-4-propan-2-ylidene-cyclopenta[c]pyrazole | 100.0 | 0.17 | 0.87 | 0.37 | 0.86 |  |
| GC | A1a | 2-(2-aminophenyl)-2-adamantanecarbonitrile | 100.0 | 0.32 | 1.36 | 0.07 | 0.75 |  |
| GC | A1a | 2,2-bis(prop-2-enyl)pyrrolidine | 100.0 | 0.04 | 0.90 | 0.58 | 0.91 |  |
| GC | A1a | 1-[4-(Dimethoxymethyl)phenyl]ethanone | 100.0 | 0.22 | 1.48 | 0.19 | 0.81 |  |
| GC | A1a | trans-11-Hydroxy-6-(hydroxymethyl)-2,3,10-trimethoxydibenzo[1a,4a:8a,12a]cyclooctadiene-7-carboxylic acid lactone | 100.0 | 0.26 | 1.11 | 0.16 | 0.80 |  |
| GC | A1a | gamma-Nonanolactone | 100.0 | 0.34 | 0.72 | 0.07 | 0.75 |  |
| GC | A1a | 1,2,2-Butanetricarboxylic acid, 4-(tetrahydro-2-methyl-2-furanyl)-, 2-ethyl ester | 100.0 | 0.42 | 0.82 | 0.34 | 0.83 |  |
| GC | A1a | 1-Furfurylpyrrole | 100.0 | 0.20 | 1.77 | 0.22 | 0.81 |  |
| GC | A1a | 3-phenylprop-2-yn-1-ol | 100.0 | 0.30 | 1.46 | 0.46 | 0.89 |  |
| GC | A1a | Mequinol | 100.0 | 0.41 | 1.25 | 0.60 | 0.92 |  |
| GC | A1a | (+-)-(1aR,2R)-1a-Methyl-2-phenylperhydrooxireno[2,3-c]furan | 100.0 | 0.15 | 0.94 | 0.86 | 0.97 |  |
| GC | A1a | Diphenyl sulfone | 100.0 | 0.12 | 0.93 | 0.77 | 0.96 |  |
| GC | A1a | methyl 6-(oxolan-2-yloxy)hexanoate | 100.0 | 0.22 | 1.02 | 0.88 | 0.97 |  |
| GC | A1a | Methyl paraben | 100.0 | 0.40 | 1.04 | 0.61 | 0.92 |  |
| GC | A1a | (1R(*),3S(*).5S(*).7S(*))-1,3-Dimethyl-7-hydroxy-6-oxabicyclo[3.2.1]octan-2-one | 100.0 | 0.04 | 1.06 | 0.87 | 0.97 |  |
| GC | A1a | 2,2,4,7-tetramethyl-3H-1-benzofuran | 100.0 | 0.53 | 2.94 | < 0.001* | 0.18 | 0.05 |
| GC | A1a | Propylidene-4,4-ethylenedioxycyclohexane | 100.0 | 0.14 | 0.92 | 0.81 | 0.97 |  |
| GC | A1a | 4-[(7-Fluoro-2,3-dihydro-1,4-benzodioxin-2-yl)methyl]morpholine | 100.0 | 0.45 | 1.33 | 0.26 | 0.82 |  |
| GC | A1a | (2,4,4-Trimethyl-6-oxo-cyclohex-1-enyl)-acetic acid ethyl ester | 100.0 | 0.24 | 0.84 | 0.23 | 0.81 |  |
| GC | A1a | 2-but-2-ynoxyoxane | 100.0 | 0.43 | 1.12 | 0.47 | 0.89 |  |
| GC | A1a | 2-(4-Chlorophenyl)-2-oxoethyl oct-2-ynoate | 100.0 | 0.38 | 1.37 | < 0.05* | 0.72 |  |
| GC | A1a | (1S)-3-(3'a-Hydroxy-7'a-methyl-5'-oxo-perhydroinden-1'-yl] acrylate | 100.0 | 0.48 | 1.43 | < 0.05* | 0.72 |  |
| GC | A1a | .alpha.-Fluoro-(p-methyl)chalcone | 100.0 | 0.71 | 0.96 | 0.76 | 0.96 | 0.97 |
| GC | A1a | Phenyl salicylate | 100.0 | 0.59 | 1.04 | 0.88 | 0.97 | 0.97 |
| GC | A1a | 2,2'-Methylenebis(4-methyl-6-tert-butylphenol) | 100.0 | 0.15 | 1.17 | 0.06 | 0.74 |  |
| GC | A1a | (4-nitrophenyl) 2-methylpropanoate | 100.0 | 0.42 | 0.92 | 0.77 | 0.96 |  |
| GC | A1a | 1,3-diethyl-1,3,3a,9a-tetrahydrobenzo[f][2]benzothiole-4,9-dione | 100.0 | 0.44 | 0.80 | 0.09 | 0.77 |  |
| GC | A1a | 7-anti-pentyl-3,3-dimethylnorbornano-1,2-endo-diol | 100.0 | 0.65 | 1.10 | 0.42 | 0.87 | 0.89 |
| GC | A1a | phenyl N-methylcarbamate | 100.0 | 0.51 | 2.10 | < 0.05* | 0.23 | 0.08 |
| GC | A1a | 15-(methoxymethoxy)-3-methylene-1-pentadecanol | 100.0 | 0.29 | 0.87 | 0.06 | 0.74 |  |
| GC | A1a | 2,7-Dimethoxy-1,3,4,5,6,8-hexamethyl-9-isopropyl-9H-xanthene | 100.0 | 0.28 | 1.08 | 0.54 | 0.91 |  |
| GC | A1a | 5-Methyl-2-(1H-pyrrol-2-yl)benzo[d]oxazole | 100.0 | 0.55 | 1.31 | 0.16 | 0.79 | 0.72 |
| GC | A1a | Pyracarbolid | 100.0 | 0.41 | 0.96 | 0.73 | 0.96 |  |
| GC | A1a | 2,2,4-Trimethylpentane-1,3-diyl dibenzoate | 100.0 | 0.24 | 0.90 | 0.29 | 0.82 |  |
| GC | A1a | 2-Phenyl[4,7]phenanthrolino[5,6-d]oxazole | 100.0 | 0.47 | 1.22 | 0.70 | 0.95 |  |
| GC | A1a | 4-(1,1,4,4,7-pentamethyltetralin-6-yl)sulfonylbenzoic acid | 100.0 | 0.71 | 0.79 | 0.43 | 0.87 | 0.91 |
| GC | A1a | (1-Methoxypentyl)benzene | 100.0 | 0.38 | 0.85 | 0.75 | 0.96 |  |
| GC | A1a | Methyl 3-Methyl-5-pentanoyl-2-furoate | 100.0 | 0.22 | 1.08 | 0.76 | 0.96 |  |
| GC | A1a | 2-(12'-methoxy-19'-norpodocarpa-8',11',13-trien-18'-yl)ethyl methanesulfonate | 100.0 | 0.42 | 1.20 | 0.34 | 0.83 |  |
| GC | A1a | 2,9-dihydro-\$b-carbolin-1-one | 100.0 | 0.10 | 1.13 | 0.55 | 0.91 |  |
| GC | A1a | 4-(1H-pyrazolyl-1-yl)indol-2-one | 100.0 | 0.17 | 0.97 | 0.80 | 0.97 |  |
| GC | A1a | (8R,9R)-Isocaryolane-8,9-diol | 100.0 | 0.43 | 0.80 | 0.09 | 0.77 |  |
| GC | A1a | methyl (E)-4-(1-hydroxy-3,4-dihydro-2H-triphenylen-1-yl)but-2-enoate | 100.0 | 0.66 | 1.14 | 0.66 | 0.93 | 0.95 |
| GC | A1a | Bis(2-ethylhexyl) phthalate | 100.0 | 0.19 | 1.10 | 0.49 | 0.89 |  |
| GC | A1a | 1,3-dimethyl-5-(2-methylpropylidene)-1,3-diazinane-2,4,6-trione | 100.0 | 0.25 | 1.29 | 0.32 | 0.83 |  |
| GC | A1a | alpha-Tocopherol | 100.0 | 0.34 | 0.95 | 0.70 | 0.95 |  |
| GC | A1a | (2-Hydroxyphenyl)acetonitrile | 100.0 | 0.18 | 1.46 | 0.12 | 0.77 |  |
| GC | A1a | 5-phenyl-3,4-dihydro-2H-pyrrole | 100.0 | 0.56 | 2.29 | < 0.05* | 0.72 | 0.53 |
| GC | A1a | methyl 3-[(1R,12bR)-3-acetyl-1,2,6,7,12,12b-hexahydroindolo[2,3-a]quinolizin-1-yl]-2-chloropropanoate | 100.0 | 0.81 | 1.14 | 0.63 | 0.93 | 0.95 |
| GC | A1a | (5S,6R)-5,6-Dihydroxy-cyclohex-3-enecarboxylic acid prop-2-ynyl ester | 100.0 | 0.22 | 1.24 | 0.30 | 0.82 |  |
| GC | A1a | Octocrylene | 100.0 | 0.06 | 0.94 | 0.44 | 0.87 |  |
| GC | A1a | 4-Methyl-4-(2-oxanyloxy)-2-pentanone | 100.0 | 0.20 | 0.89 | 0.54 | 0.91 |  |
| GC | A1a | 1-(cyclopentylmethyl)-4-methoxy-benzene | 100.0 | 0.68 | 0.55 | 0.13 | 0.77 | 0.69 |
| GC | A1a | Triethylene glycol diacetate | 100.0 | 0.20 | 0.94 | 0.44 | 0.87 |  |
| GC | A1a | (2,6-dimethyl-4-propyl-phenyl)amine | 100.0 | 0.26 | 1.02 | 0.94 | 0.97 |  |
| GC | A1a | 2-O-cyclohexyl 1-O-pentyl benzene-1,2-dicarboxylate | 100.0 | 0.75 | 0.99 | 0.91 | 0.97 | 0.97 |
| GC | A1a | 3-(2-(Piperidin-1-yl)acetamido)benzoic acid | 100.0 | 0.24 | 0.90 | 0.59 | 0.92 |  |
| GC | A1a | Morpholyl-2-oxocyclohexanecarboxamide | 100.0 | 0.55 | 0.91 | 0.47 | 0.89 | 0.92 |
| GC | A1a | 1-Methoxy-1-(4-methoxyphenyl)-4-phenylbutane | 100.0 | 0.41 | 0.66 | 0.30 | 0.82 |  |
| GC | A1a | 3,5,5-trimethyl-4-[(E)-3-oxidanylbut-1-enyl]cyclohex-3-en-1-ol | 100.0 | 0.42 | 0.76 | 0.19 | 0.81 |  |
| GC | A1a | Kikkanol F monoacetate | 100.0 | 0.37 | 0.67 | 0.08 | 0.76 |  |
| GC | A1a | 5,12-Dihydroxysterpyrene | 100.0 | 0.09 | 0.85 | 0.45 | 0.88 |  |
| GC | A1a | 4-Methylene-2-phenyl-tetrahydrofuran | 100.0 | 0.42 | 1.19 | 0.48 | 0.89 |  |
| GC | A1a | Phenyl benzoate | 100.0 | 0.34 | 1.49 | 0.46 | 0.89 |  |
| GC | A1a | 2-Isobutyryl-benzo[1,3]dioxol-5-carbaldehyde | 100.0 | 0.61 | 1.31 | 0.40 | 0.86 | 0.88 |
| GC | A1a | DL-trans-2-acetoxy-1-benzoyloxymethyl-3-[(E)-cinnamoyloxy]-4,6-cyclohexadiene | 100.0 | 0.18 | 0.82 | 0.21 | 0.81 |  |
| GC | A1a | methyl 2-hydroxy-2-(3-methoxyphenyl)acetate | 100.0 | 0.28 | 0.93 | 0.51 | 0.89 |  |
| GC | A1a | Methyl 2-(3',3'-dimethyl-1'-butyn-1'-yl)-1-cyclohexenecarboxylate | 100.0 | 0.91 | 1.37 | 0.35 | 0.84 | 0.84 |
| GC | A1a | 3,4-dihydro-2H-naphthalen-1-one | 100.0 | 0.44 | 1.24 | 0.47 | 0.89 |  |
| GC | A1a | 4-(1-Pyrrolidinyl)pyridazine | 100.0 | 0.13 | 1.03 | 0.88 | 0.97 |  |
| GC | A1a | Phenyl N-benzoyl-N-methyl-thiooxocarbamate | 100.0 | 0.34 | 1.11 | 0.54 | 0.91 |  |
| GC | A1a | Furaltadone | 100.0 | 0.07 | 0.80 | 0.51 | 0.89 |  |
| GC | A1a | Benzyl butyl phthalate | 100.0 | 0.41 | 1.20 | 0.26 | 0.82 |  |
| GC | A1a | 3-benzyl-4,7,7-trimethyl-3-azabicyclo[2.2.1]heptan-2-one | 100.0 | 0.41 | 0.97 | 0.94 | 0.97 |  |
| GC | A1a | Dicrotophos | 100.0 | 0.31 | 0.90 | 0.62 | 0.93 |  |
| GC | A1a | 1-ethyl-3-[(E)-hydroxyiminomethyl]pyridine-2-thione | 100.0 | 0.24 | 0.74 | 0.48 | 0.89 |  |
| GC | A1a | (R)-6-Phenyltetrahydro-2H-pyran-2-one | 100.0 | 0.09 | 1.25 | 0.38 | 0.86 |  |
| GC | A1a | Beta-BHC | 100.0 | 0.40 | 0.88 | 0.58 | 0.92 |  |
| GC | A1a | o-Phenoxybenzyl Alcohol | 100.0 | 0.54 | 1.97 | 0.10 | 0.77 | 0.67 |
| GC | A1a | 2,6-Diisopropylnaphthalene | 100.0 | 0.87 | 1.82 | 0.22 | 0.81 | 0.82 |
| GC | A1a | 4-[[[2-(Dimethylamino)ethyl]methylamino]methyl]phenol | 100.0 | 0.63 | 1.06 | 0.72 | 0.96 | 0.97 |
| GC | A1a | Benzothiazolone | 100.0 | 0.14 | 1.10 | 0.48 | 0.89 |  |
| GC | A1a | 2-[(4S)-2,2,4-trimethyl-1,3-dioxolan-4-yl]ethyl methanesulfonate | 100.0 | 0.24 | 0.97 | 0.81 | 0.97 |  |
| GC | A1a | Fluorene | 100.0 | 0.05 | 1.01 | 0.99 | 1.00 |  |
| GC | A1a | 8,9,11,12-Tetraimethoxy-1-methyl-1,2,3,4-dihydronaphtho[2,1-f]isoquinoline | 100.0 | 0.36 | 1.02 | 0.91 | 0.97 |  |
| GC | A1a | 1-(4-Chlorophenyl)-2-methylbut-3-en-1-ol | 100.0 | 0.13 | 1.39 | 0.12 | 0.77 |  |
| GC | A1a | n-Undecyl cyanide | 100.0 | 0.43 | 1.49 | 0.41 | 0.87 |  |
| GC | A1a | 4-[(2-methylacridin-9-yl)amino]-N-(1,3-thiazol-2-yl)benzenesulfonamide | 100.0 | 0.35 | 1.07 | 0.69 | 0.95 |  |
| GC | A1a | Octisalate | 100.0 | 0.52 | 1.34 | 0.63 | 0.93 | 0.95 |
| GC | A1a | 2-(4-Fluorophenyl)isoindoline | 100.0 | 0.65 | 1.37 | 0.08 | 0.76 | 0.64 |
| GC | A1a | Ethylparaben | 100.0 | 0.45 | 1.36 | 0.53 | 0.90 |  |
| GC | A1a | (2S,6R)-4-[3-(4-tert-butylphenyl)-2-methylpropyl]-2,6-dimethylmorpholine | 100.0 | 0.23 | 1.49 | 0.35 | 0.84 |  |
| GC | A1a | Octa-1,7-diyne-3,6-diol | 100.0 | 0.63 | 1.28 | 0.20 | 0.81 | 0.80 |
| GC | A1a | (E)-stilbene | 100.0 | 0.53 | 1.09 | 0.47 | 0.89 | 0.92 |
| GC | A1a | 2-butyl-6-methoxy-2,5-dihydropyridine | 100.0 | 0.42 | 1.10 | 0.55 | 0.91 |  |
| GC | A1a | 2-Methoxy-N,N-dimethyl-2-phenylacetamide | 100.0 | 0.41 | 1.35 | 0.12 | 0.77 |  |
| GC | A1a | .beta.-1,5-anhydro-2,3,4,6-tetra-O-methyl-1-C-phenyl-D-glucitol | 100.0 | 0.23 | 0.75 | < 0.05* | 0.72 |  |
| GC | A1a | 1-phenyl-2-sulfanylidene-3-pyridinecarboxylic acid | 100.0 | 0.08 | 1.22 | 0.55 | 0.91 |  |
| GC | A1a | Allyltri(4-N,N-dimethylaminophenyl)silane | 100.0 | 0.29 | 0.71 | 0.08 | 0.77 |  |
| GC | A1a | 7,10-Dioxo-8,9-dihydro[4]-paracyclophane | 100.0 | 0.63 | 0.86 | 0.60 | 0.92 | 0.95 |
| GC | A1a | Pyrolo[3,2-d]pyrimidin-2,4(1H,3H)-dione | 100.0 | 0.26 | 1.13 | 0.53 | 0.90 |  |
| GC | A1a | Mepivacaine | 100.0 | 0.02 | 1.27 | < 0.05* | 0.72 |  |
| GC | A1a | 4-Chlorophenol | 100.0 | 0.37 | 1.03 | 0.91 | 0.97 |  |
| GC | A1a | N-[(E)-3-cyano-1,1-dimethyl-allyl]acetamide | 100.0 | 0.60 | 0.84 | 0.49 | 0.89 | 0.92 |
| GC | A1a | Acetic Acid (+-)-(1.alpha.,3.beta.,4.beta.,6.alpha.,7.alpha.)-7-(3-Methyl-3-butenyl)-4-((E)-6-methyl-2,6-heptadienyl)-1-methylbicyclo[4.1.0]hept-3-yl Ester | 100.0 | 0.84 | 0.94 | 0.55 | 0.91 | 0.93 |
| GC | A1a | 2-Amino-2'-tert-butylthio-6-ethoxy[3,4']bipyridine-3,5-dicarbonitrile | 100.0 | 0.67 | 1.56 | < 0.05* | 0.51 | 0.18 |
| GC | A1a | Pyrene | 100.0 | 0.10 | 0.73 | 0.31 | 0.82 |  |
| GC | A1a | trans-2,3-di(methoxycarbonyl)-bicyclo[2.2.2]oct-5-ene | 100.0 | 0.41 | 0.73 | 0.37 | 0.86 |  |
| GC | A1a | 3-(4-Vinylbenzyloxy)benzyl alcohol | 100.0 | 0.55 | 0.86 | 0.85 | 0.97 | 0.97 |
| GC | A1a | methyl 2-pyrimidin-4-ylpropanoate | 100.0 | 0.63 | 1.40 | 0.11 | 0.77 | 0.67 |
| GC | A1a | 2-methyl-1H-benzimidazole | 100.0 | 0.45 | 1.57 | 0.13 | 0.77 |  |
| GC | A1a | (R)-2-N-Heptyl (S)-O-1-phenylethyl carbamate | 100.0 | 0.10 | 0.86 | 0.66 | 0.93 |  |
| GC | A1a | ethyl (2E,4E)-6-methoxy-6-sulfanylidenehexa-2,4-dienoate | 100.0 | 0.43 | 1.18 | 0.56 | 0.91 |  |
| GC | A1a | 1-(Biphenyl-4-yl)ethanone | 100.0 | 0.61 | 0.99 | 0.91 | 0.97 | 0.97 |
| GC | A1a | ethyl 2-((2r,4R,6S)-2,4,6-trimethyl-1,3-dioxan-2-yl)acetate | 100.0 | 0.31 | 1.04 | 0.76 | 0.96 |  |
| GC | A1a | 2-methyl-5-phenyl-1H-pyrrole | 100.0 | 0.49 | 1.84 | < 0.05* | 0.21 |  |
| GC | A1a | (6R,7R)-7-isopropyl-1,3-dihydro-1,5,5,6-tetramethyl-indan[5,6-c]furan | 100.0 | 0.46 | 1.13 | 0.29 | 0.82 |  |
| GC | A1a | 3,3,3-trifluoro-2-hydroxy-1-(4-methylphenyl)-1-propanone | 100.0 | 0.57 | 1.26 | 0.29 | 0.82 | 0.82 |
| GC | A1a | (4-methoxy-3-propan-2-yloxy-phenyl) ethanoate | 100.0 | 0.20 | 1.36 | 0.27 | 0.82 |  |
| GC | A1a | benzhydryl 2-(1-methyl-3-oxo-2-prop-2-enylcyclohexyl)acetate | 100.0 | 0.54 | 1.05 | 0.79 | 0.97 | 0.97 |
| GC | A1a | 9-Methyl-9,10-dihydroanthracene | 100.0 | 0.14 | 1.67 | 0.12 | 0.77 |  |
| GC | A1a | (3aR,5R,7aR)-1,1-Ethylidenedioxy-7a-methyl-3a,4,5,6,7,7a-hexahydroindan-5-ol | 100.0 | 0.53 | 0.89 | 0.57 | 0.91 | 0.94 |
| GC | A1a | N-(cyclohexylmethyl)morpholine | 100.0 | 0.18 | 1.06 | 0.76 | 0.96 |  |
| GC | A1a | (E)-.beta.-(tert-Butyldimethylsiloxy)-.beta.-(tert-butyldimethylsilyl)styrene | 100.0 | 0.29 | 1.08 | 0.80 | 0.97 |  |
| GC | A1a | 6-Heptadecylthymoquinone | 100.0 | 0.38 | 1.09 | 0.37 | 0.86 |  |
| GC | A1a | 2,5-Pyrrolidinedione, 1-[11-(4-morpholinyl)bicyclo[8.1.0]undec-11-yl]-, stereoisomer | 100.0 | 0.70 | 1.93 | 0.17 | 0.81 | 0.76 |
| GC | A1a | (Z)-3-(4-n-Butyl-3-thienyl)propenal | 100.0 | 0.59 | 1.03 | 0.89 | 0.97 | 0.97 |
| GC | A1a | (S)-(4-butoxyphenyl)(1-phthalimdoethyl)-ketone | 100.0 | 0.37 | 1.06 | 0.63 | 0.93 |  |
| GC | A1a | Acetyl tributyl citrate | 100.0 | 0.28 | 0.93 | 0.50 | 0.89 |  |
| GC | A1a | (4S,5R)-3-(3-methyl-1-oxobut-2-enyl)-4,5-diphenyl-2-oxazolidinone | 100.0 | 0.49 | 0.89 | 0.83 | 0.97 |  |
| GC | A1a | 3-Isopropoxy-5-methyl-1H-pyrazole | 100.0 | 0.52 | 0.93 | 0.85 | 0.97 | 0.97 |
| GC | A1a | N'-(2-methylpropyl)benzohydrazide | 100.0 | 0.46 | 1.22 | 0.43 | 0.87 |  |
| GC | A1a | 3-Butanoyloxy-2-(2-propenyl)tetrahydropyran | 100.0 | 0.17 | 1.00 | 0.98 | 1.00 |  |
| GC | A1a | 2-Phenoxyethyl propanoate | 100.0 | 0.34 | 0.96 | 0.90 | 0.97 |  |
| GC | A1a | Molinate | 100.0 | 0.15 | 1.12 | 0.43 | 0.87 |  |
| GC | A1a | 1-Decyclopentane-1,3-dione | 100.0 | 0.01 | 1.29 | 0.21 | 0.81 |  |
| GC | A1a | 1-(4-phenyl-1H-pyrrol-3-yl)propan-1-one | 100.0 | 0.48 | 0.83 | 0.40 | 0.86 |  |
| GC | A1a | Beta-Tocopherol | 100.0 | 0.55 | 1.02 | 0.74 | 0.96 | 0.97 |
| GC | A1a | N-pyridin-2-ylnitramide | 100.0 | 0.65 | 1.00 | 0.99 | 1.00 | 0.99 |
| GC | A1a | 2,2',3,4,4',5',6-Heptachlorobiphenyl | 100.0 | 0.25 | 1.01 | 0.95 | 0.98 |  |
| GC | A1a | Triethyl citrate | 100.0 | 0.39 | 0.91 | 0.58 | 0.92 |  |
| GC | A1a | (3R,8S,9S,10R,13S,14S,17S)-17-[(1S)-1-(dimethylamino)ethyl]-N-ethyl-N,10,13-trimethyl-2,3,4,7,8,9,11,12,14,15,16,17-dodecahydro-1H-cyclopenta[a]phenanthren-3-amine | 100.0 | 0.25 | 0.70 | 0.18 | 0.81 |  |
| GC | A1a | (2E,7E)-5-hydroxy-6,6,9-trimethyl-4-deca-2,7,9-trienone | 100.0 | 0.55 | 1.31 | 0.24 | 0.82 | 0.82 |
| GC | A1a | 2-Methyl-3-(trifluoromethyl)-1,4-naphthoquinone | 100.0 | 0.61 | 0.82 | 0.66 | 0.93 | 0.95 |
| GC | A1a | 1,3,5-Trimethyladamantane | 100.0 | 0.56 | 0.86 | 0.51 | 0.89 | 0.93 |
| GC | A1a | (1S)-1-[(3S,5S,8R,9S,10S,13S,14S,17S)-10,13-dimethyl-3-(propan-2-ylideneamino)-2,3,4,5,6,7,8,9,11,12,14,15,16,17-tetradecahydro-1H-cyclopenta[a]phenanthren-17-yl]-N,N-dimethylethanamine | 100.0 | 0.03 | 0.86 | 0.25 | 0.82 |  |
| GC | A1a | 2-Phenylethyl propionate | 100.0 | 0.39 | 0.91 | 0.23 | 0.81 |  |
| GC | A1a | (trans)-2-(Hydroxymethyl)-3-phenyltetrahydrofuran | 100.0 | 0.77 | 0.79 | 0.42 | 0.87 | 0.90 |
| GC | A1a | PCB 130 | 100.0 | 0.59 | 1.12 | 0.22 | 0.81 | 0.82 |
| GC | A1a | N,N-dimethyl-4-phenylbutanamide | 100.0 | 0.24 | 1.11 | 0.46 | 0.89 |  |
| GC | A1a | 2',6'-Dichloro-2,4-dimethylbenzanilide | 100.0 | 0.06 | 1.21 | 0.21 | 0.81 |  |
| GC | A1a | 4-(2-thenoyl)tetrahydrofuran-2-one | 100.0 | 0.43 | 1.34 | 0.62 | 0.93 |  |
| GC | A1a | 4a-Methyl-4,4a,5,6-tetrahydro-2(3H),7-indendione | 100.0 | 0.28 | 1.06 | 0.75 | 0.96 |  |
| GC | A1a | 1-methylquinolin-2(1H)-one | 100.0 | 0.56 | 1.43 | 0.51 | 0.89 | 0.93 |
| GC | A1a | (+)-Pinidine | 100.0 | 0.57 | 1.25 | 0.27 | 0.82 | 0.82 |
| GC | A1a | (3R*,4S*)-3-Ethyl-4-(trimethylsilyethynyl)azitidin-2-one | 100.0 | 0.03 | 1.27 | 0.18 | 0.81 |  |
| GC | A1a | 3-Methoxyphthalide | 100.0 | 0.11 | 1.38 | 0.14 | 0.77 |  |
| GC | A1a | methyloctyldiethoxysilane | 100.0 | 0.35 | 0.81 | 0.30 | 0.82 |  |
| GC | A1a | 1-Amino-3,4-dihydro-3,4-dimethyl-2-naphthalenecarbonitrile | 100.0 | 0.80 | 1.19 | 0.40 | 0.86 | 0.88 |
| GC | A1a | 2-Naphthalenamine | 100.0 | 0.59 | 0.95 | 0.85 | 0.97 | 0.97 |
| GC | A1a | 1,3-trans-4-Hydroxymethyl-1-methylcyclopentane-1,3-diol | 100.0 | 0.55 | 1.49 | < 0.05* | 0.51 | 0.18 |
| GC | A1a | 1-methoxy-4-[(E)-2-phenylethenyl]benzene | 100.0 | 0.69 | 1.00 | 0.95 | 0.98 | 0.98 |
| GC | A1a | 2-methyl-1,3-benzoxazole | 100.0 | 0.15 | 0.93 | 0.51 | 0.89 |  |
| GC | A1a | Diisobutyl phthalate | 100.0 | 0.47 | 0.85 | 0.37 | 0.86 |  |
| GC | A1a | 8-methylpyrrolo[1,2-a]quinoxaline | 100.0 | 0.07 | 1.04 | 0.87 | 0.97 |  |
| GC | A1a | 1-(2,4-Dimethylphenyl)but-3-en-1-ol | 100.0 | 0.38 | 1.21 | 0.62 | 0.93 |  |
| GC | A1a | 1,4,5,8-Tetramethylnaphthalene | 100.0 | 0.43 | 1.29 | 0.30 | 0.82 |  |
| GC | A1a | N-tert-Butylmaleimide | 100.0 | 0.23 | 0.98 | 0.85 | 0.97 |  |
| GC | A1a | 1-Piperidino-5-(2-hydroxy-4-nitrophenoxy)pentane | 100.0 | 0.24 | 0.53 | < 0.05* | 0.51 |  |
| GC | A1a | Diisopropyl phthalate | 100.0 | 0.75 | 1.13 | 0.48 | 0.89 | 0.92 |
| GC | A1a | Kanzonol P | 100.0 | 0.88 | 0.96 | 0.92 | 0.97 | 0.97 |
| GC | A1a | Hexachlorobenzene | 100.0 | 0.60 | 0.99 | 0.90 | 0.97 | 0.97 |
| GC | A1a | 5-Vinyloxyhept-6-enyl 2,2-dimethylpropanoate | 100.0 | 0.28 | 0.83 | 0.25 | 0.82 |  |
| GC | A1a | 1-Piperidino-5-(4-amino-2-methoxyphenoxy)ethane | 100.0 | 0.07 | 0.83 | 0.09 | 0.77 |  |
| GC | A1a | 1-chloro-4-[2,2,2-trichloro-1-(4-chlorophenyl)ethyl]benzene | 100.0 | 0.08 | 1.09 | 0.62 | 0.93 |  |
| GC | A1a | 3-(4-ketohexyl)cyclohex-2-en-1-one | 100.0 | 0.20 | 0.87 | 0.86 | 0.97 |  |
| GC | A1a | Tris(2-butoxyethyl) phosphate | 100.0 | 0.16 | 1.20 | 0.13 | 0.77 |  |
| GC | A1a | 2-(4-Methoxyphenoxy)benzothiophene | 100.0 | 0.47 | 0.87 | 0.67 | 0.94 |  |
| GC | A1a | Phenanthrene | 100.0 | 0.46 | 1.08 | 0.91 | 0.97 |  |
| GC | A1a | methyl 2-(5-methoxy-1,2-dimethylindol-3-yl)acetate | 100.0 | 0.50 | 0.86 | 0.34 | 0.83 | 0.84 |
| GC | A1a | 2'-Hydroxy-4',6'-dimethoxyacetophenone | 100.0 | 0.29 | 0.68 | 0.12 | 0.77 |  |
| GC | A1a | 2,2',3,4,4',5,5'-Heptachlorobiphenyl | 100.0 | 0.57 | 1.09 | 0.16 | 0.80 | 0.72 |
| GC | A1a | 3-Hexene-2,5-dione, 3-acetyl-, 5-(dimethylhydrazone) | 100.0 | 0.13 | 0.80 | 0.22 | 0.81 |  |
| GC | A1a | Captafol | 100.0 | 0.01 | 0.84 | 0.14 | 0.77 |  |
| GC | A1a | N-(2-Fluorobenzylidene)-2-fluorophenylmethylamine | 100.0 | 0.30 | 0.86 | 0.25 | 0.82 |  |
| GC | A1a | (2-ethyl-4,6-dimethylphenyl)methanol | 100.0 | 0.24 | 1.61 | 0.21 | 0.81 |  |
| GC | A1a | 2,2-Dimethyl-2H-chromene-6-carbaldehyde | 100.0 | 0.38 | 0.87 | 0.30 | 0.82 |  |
| GC | A1a | 3,3-Dimethyl-1,3-dihydro-1-isobenzofuranone | 100.0 | 0.24 | 2.59 | < 0.05* | 0.72 |  |
| GC | A1a | 3-Butenyl-5-propylindolizidine | 100.0 | 0.53 | 1.05 | 0.81 | 0.97 | 0.97 |
| GC | A1a | Di-n-propylphthalate | 100.0 | 0.31 | 0.81 | 0.16 | 0.80 |  |
| GC | A1a | 3,4-Diphenyl-6-methyl-7,8-(1,4-dimethoxybenzo)-9-oxatricyclo[4.2.1.0(2,5)]non-3-ene | 100.0 | 0.50 | 1.19 | < 0.05* | 0.74 |  |
| GC | A1a | 5-bicyclo[2.2.2]oct-2-enecarbonitrile | 100.0 | 0.56 | 1.90 | < 0.05* | 0.72 | 0.55 |
| GC | A1a | 1,6-dimethyl-4-propan-2-ylnaphthalene | 100.0 | 0.18 | 0.71 | 0.07 | 0.75 |  |
| GC | A1a | 1,2-Dicyano-4,5-di[(N,N-dimethylaminocarbonyl)methoxy]benzene | 100.0 | 0.17 | 0.78 | 0.11 | 0.77 |  |
| GC | A1a | 4-Hydroxyacetophenone | 100.0 | 0.55 | 0.99 | 0.90 | 0.97 | 0.97 |
| GC | A1a | 2-(2,3-dihydro-1H-indol-4-yl)ethanol | 100.0 | 0.03 | 1.04 | 0.67 | 0.94 |  |
| GC | A1a | (Z)-2-((2-Methylthio)styrylthio)aniline | 100.0 | 0.03 | 0.95 | 0.59 | 0.92 |  |
| GC | A1a | Carbaryl | 100.0 | 0.54 | 1.03 | 0.92 | 0.97 | 0.97 |
| GC | A1a | 4-Butylaniline | 100.0 | 0.52 | 0.74 | 0.08 | 0.77 | 0.64 |
| GC | A1a | Dibenzofuran | 100.0 | 0.11 | 0.96 | 0.87 | 0.97 |  |
| GC | A1a | 3-(2-methylbutyl)-1H-indole | 100.0 | 0.77 | 0.67 | 0.29 | 0.82 | 0.82 |
| GC | A1a | (2s)-n-tert-butoxycarbonyl-2-(4-(1-ethoxyethoxy)-1-hydroxybutyl)pyrrolidine | 100.0 | 0.02 | 0.80 | 0.06 | 0.74 |  |
| GC | A1a | (2RS,4SR)-4-(dimethylamino)-2,4-diphenyl-2-butanol | 100.0 | 0.20 | 0.99 | 0.92 | 0.97 |  |
| GC | A1a | 2-ethynylbenzaldehyde | 100.0 | 0.28 | 1.03 | 0.89 | 0.97 |  |
| GC | A1a | 1-Ethyl-4,5,8-trimethylnaphthalene | 100.0 | 0.79 | 1.36 | 0.06 | 0.74 | 0.55 |
| GC | A1a | Bisphenol A | 100.0 | 0.80 | 1.12 | 0.48 | 0.89 | 0.92 |
| GC | A1a | N-(1,2-diphenyl-2-phenylsulfanylethyl)acetamide | 100.0 | 0.37 | 0.77 | 0.36 | 0.85 |  |
| GC | A1a | Dehydro-botrydienol | 100.0 | 0.21 | 1.00 | 0.99 | 1.00 |  |
| GC | A1a | 2-methyl-2-[(E)-2-methyl-3-phenylprop-2-enyl]-1,3-dioxolane | 100.0 | 0.57 | 0.87 | 0.33 | 0.83 | 0.84 |
| GC | A1a | 5,5-Dicyano-2-methyl-3,4-dihydro-pyrrole | 100.0 | 0.43 | 1.23 | 0.40 | 0.86 |  |
| GC | A1a | Tetracyclo[6.2.1.1(3,6)]dodecane-2,7-dione | 100.0 | 0.67 | 0.95 | 0.81 | 0.97 | 0.97 |
| GC | A1a | 1-(2-piperidin-1-ylethyl)cyclohexan-1-ol | 100.0 | 0.55 | 0.93 | 0.87 | 0.97 | 0.97 |
| GC | A1a | 5-Phenylporphyrin | 100.0 | 0.81 | 0.61 | 0.09 | 0.77 | 0.64 |
| GC | A1a | 2-(2-Butoxyethyl)pyridine | 100.0 | 0.03 | 0.80 | 0.22 | 0.81 |  |
| GC | A1a | 1,2,3,3,4,4,5,5-Octamethylcyclopentene | 100.0 | 0.55 | 0.33 | < 0.05* | 0.51 | 0.18 |
| GC | A1a | 4-(2'-Hydroxyphenyl)non-1-ene | 100.0 | 0.47 | 0.81 | 0.21 | 0.81 |  |
| GC | A1a | N,N-Dimethyl-2,2-bis( 9'-anthracenyl)acetamide | 100.0 | 0.37 | 0.85 | 0.40 | 0.86 |  |
| GC | A1a | methyl (2S,3S)-4-(4-methoxyphenyl)-3-methyl-2-[(2-methylpropan-2-yl)oxycarbonylamino]-4-oxobutanoate | 100.0 | 0.12 | 1.06 | 0.69 | 0.95 |  |
| GC | A1a | ethyl 2-methylidene-4-(2,4,6-trimethylphenyl)butanoate | 100.0 | 0.35 | 1.26 | 0.07 | 0.75 |  |
| GC | A1a | (2S,6R)-4-[3-(4-tert-butylphenyl)-2-methylpropyl]-2,6-dimethylmorpholine | 100.0 | 0.03 | 1.04 | 0.70 | 0.95 |  |
| GC | A1a | 2,2',4,4',5,5'-Hexachlorobiphenyl | 100.0 | 0.47 | 1.07 | 0.42 | 0.87 |  |
| GC | A1a | (2E)-1-O-Acetyl-3,6-dimethyl-6-formyl-2-hepten-1-ol | 100.0 | 0.03 | 0.95 | 0.81 | 0.97 |  |
| GC | A1a | 2-(1-bromoethenyl)-5-methoxy-1,3-dimethylbenzene | 100.0 | 0.71 | 1.20 | 0.51 | 0.89 | 0.93 |
| GC | A1a | (1S,3R,4R)-2-[(S)-1-Phenylethylamino]-2-azabicyclo[2.2.1]heptane-3(S)-phenylmethanol | 100.0 | 0.21 | 0.89 | 0.21 | 0.81 |  |
| GC | A1a | N-(n-Butyl)-3-cyclohexenylpropiolamide | 100.0 | 0.44 | 0.82 | 0.56 | 0.91 |  |
| GC | A1a | 4-tert-Octylphenol | 100.0 | 0.33 | 0.90 | 0.27 | 0.82 |  |
| GC | A1a | 2-phenylpropanenitrile | 100.0 | 0.85 | 2.30 | < 0.05* | 0.72 | 0.48 |
| GC | A1a | (-)-(4a'R,8'S,8a'S,10a'S)-1',1',4a',8a'-Tetramethyl-3',4',4a',6',7',8',8a',9',10',10a'-decahydro-1'H-spiro[[1,3]dioxolane-2,2'-phenanthren]-8'-ol | 100.0 | 0.39 | 0.96 | 0.76 | 0.96 |  |
| GC | A1a | Benzoic acid, 2,4-dihydroxy-3-[[4-methoxy-2-(phenylmethoxy)-6-propylbenzoyl]oxy]-6-propyl-, methyl ester | 100.0 | 0.28 | 0.99 | 0.96 | 0.98 |  |
| GC | A1a | dimethyl 5-(1H-indol-3-yl)-2-methyl-3-methylsulfanyl-1,2,4-triazine-5,6-dicarboxylate | 100.0 | 0.27 | 0.89 | 0.43 | 0.87 |  |
| GC | A1a | 1,1-Dimethyl-1,3-dihydro-isobenzofuran-5-ol | 100.0 | 0.70 | 1.04 | 0.88 | 0.97 | 0.97 |
| GC | A1a | Methyl 1,3-dihydro-2H-isobenzofuran-4-carboxylate | 100.0 | 0.17 | 0.74 | 0.34 | 0.83 |  |
| GC | A1a | Diethyl Phthalate | 100.0 | 0.04 | 1.10 | 0.50 | 0.89 |  |
| GC | A1a | ethyl 2-hydroxy-2-(4-methylphenyl)-2-(5-oxocyclopenten-1-yl)acetate | 100.0 | 0.18 | 0.82 | 0.06 | 0.74 |  |
| GC | A1a | Methoxyethyl-3-(4-oxo-4H-1-benzopyran-3-yl)-acrylate | 100.0 | 0.42 | 0.96 | 0.93 | 0.97 |  |
| GC | A1a | Octahydro-5-pentylindolizine-8-methanol | 100.0 | 0.30 | 1.27 | 0.05 | 0.74 |  |
| GC | A1a | 1-prop-1-ynyl-1-cyclohex-2-enol | 100.0 | 0.79 | 1.28 | 0.50 | 0.89 | 0.93 |
| GC | A1a | .alpha.-Chloroformyl-4-ethoxyphenylhydrazine hydrochloride | 100.0 | 0.80 | 1.17 | 0.54 | 0.91 | 0.93 |
| GC | A1a | 2-Ethyl-N-methyl-1-phenyl-3-butenylamine | 100.0 | 0.43 | 1.76 | 0.26 | 0.82 |  |
| GC | A1a | 3-Methoxy-3-oxopropane-1,2-diyl dibutyrate | 100.0 | 0.05 | 0.66 | < 0.05* | 0.72 |  |
| GC | A1a | DEET | 100.0 | 0.15 | 0.55 | < 0.05* | 0.68 |  |
| GC | A1a | 3-(p-Methoxyphenyl)-5,6-dimethylindenone | 100.0 | 0.09 | 1.06 | 0.92 | 0.97 |  |
| GC | A1a | [2-(4-ethylbenzoyl)oxyphenyl] 2,6-difluorobenzoate | 100.0 | 0.63 | 0.72 | 0.34 | 0.83 | 0.84 |
| GC | A1a | 4-Ethylbenzoic acid, 3-chlorophenyl ester | 100.0 | 0.25 | 1.00 | 0.99 | 1.00 |  |
| GC | A1a | 5,6,8-Trimethoxy-7-methyl-2,3-diphenylquinoxaline | 100.0 | 0.74 | 0.89 | 0.68 | 0.94 | 0.96 |
| GC | A1a | Methfuroxam | 100.0 | 0.26 | 0.79 | 0.38 | 0.86 |  |
| GC | A1a | 2-(3,5-dichloro-2-methoxy-phenyl)-1H-pyrrole | 100.0 | 0.63 | 0.90 | 0.91 | 0.97 | 0.97 |
| GC | A1a | p-Tolyl Furan-2-Carboxylate | 100.0 | 0.34 | 1.16 | 0.69 | 0.95 |  |
| GC | A1a | 2-Methyl-2-(2'-propenyl)-cyclohexane-1,3-dione | 100.0 | 0.35 | 0.63 | < 0.05* | 0.51 |  |
| GC | A1a | 4,7-Dihydroxy-5-methylcoumarin | 100.0 | 0.78 | 0.82 | 0.39 | 0.86 | 0.88 |
| GC | A1a | (2S)-2-amino-2-(1,3-benzodioxol-5-yl)acetic acid methyl ester | 100.0 | 0.49 | 0.96 | 0.41 | 0.87 |  |
| GC | A1a | [(1R,2S,4aR,6R,8aR)-2,6-dimethyl-1,2,4a,5,6,7,8,8a-octahydronaphthalen-1-yl]methanol | 100.0 | 0.69 | 1.07 | 0.76 | 0.96 | 0.97 |
| GC | A1a | (4-methylphenyl) 2-hydroxybenzoate | 100.0 | 0.14 | 1.14 | 0.71 | 0.95 |  |
| GC | A1a | (1R,2S,6R)-2-Acetoxy-9-(hex-2-en-2-yl)bicyclo[4.3.0]nona-3,7-diene | 100.0 | 0.29 | 1.51 | 0.09 | 0.77 |  |
| GC | A1a | Methyl phenidate | 100.0 | 0.10 | 0.70 | < 0.05* | 0.68 |  |
| GC | A1a | 2-[(Phenylsulfanyl)methyl]oxirane | 100.0 | 0.26 | 1.07 | 0.85 | 0.97 |  |
| GC | A1a | endo-5,6-Di(acetyl)-7-acetoxymethylenebicyclo[2.2.1]hept-2-ene | 100.0 | 0.38 | 1.51 | 0.14 | 0.77 |  |
| GC | A1a | (E)-[(4S)-4-allyl-2,2-dimethyl-1,3-dioxan-5-ylidene]-[(2S)-2-(methoxymethyl)pyrrolidino]amine | 100.0 | 0.01 | 0.96 | 0.76 | 0.96 |  |
| GC | A1a | 3-(piperidin-1-ylmethyl)-2,3-dihydro-1,4-benzodioxin-6-yl Methanesulfonate | 100.0 | 0.25 | 1.02 | 0.93 | 0.97 |  |
| GC | A1a | 2,4-di-tert-butylphenol | 100.0 | 0.63 | 1.13 | 0.75 | 0.96 | 0.97 |
| GC | A1a | Cyclopropyl 2-aminobenzoate | 100.0 | 0.01 | 1.36 | 0.30 | 0.82 |  |
| GC | A1a | Methoxy-colorenone | 100.0 | 0.37 | 0.78 | 0.30 | 0.82 |  |
| GC | A1a | Methyl 1-(phenylsulfonyl)-3-(methoxycarbonyl)indole-2-acetate | 100.0 | 0.34 | 0.60 | < 0.05* | 0.51 |  |
| GC | A1a | (E)-1-cyclohexyl-2-hepten-1-one | 100.0 | 0.40 | 0.95 | 0.70 | 0.95 |  |
| GC | A1a | 3-(ethylamino)phenol | 100.0 | 0.52 | 1.04 | 0.82 | 0.97 | 0.97 |
| GC | A1a | N-ethyl-N-[(7-fluoro-2,3-dihydro-1,4-benzodioxin-2-yl)methyl]propan-1-amine | 100.0 | 0.12 | 0.97 | 0.90 | 0.97 |  |
| GC | A1a | 3-methylcyclohepta[b]furan-2-one | 100.0 | 0.17 | 1.39 | 0.25 | 0.82 |  |
| GC | A1a | methyl (2R,3R,3aR)-2-(2-methylpropanoyl)-2,3,3a,4,5,6-hexahydropyrrolo[1,2-b][1,2]oxazole-3-carboxylate | 100.0 | 0.08 | 0.92 | 0.67 | 0.94 |  |
| GC | A1a | N-[(7-fluoro-2,3-dihydro-1,4-benzodioxin-2-yl)methyl]-N-methylethanamine | 100.0 | 0.01 | 0.98 | 0.93 | 0.97 |  |
| GC | A1a | 6,9-Dimethoxy-5,5-dimethyl-3,3a,5,9b-tetrahydro-furo[3,2-c]isochromen-2-one | 100.0 | 0.15 | 0.96 | 0.71 | 0.95 |  |
| GC | A1a | 3-hydroxypropyl benzoate | 100.0 | 0.37 | 1.06 | 0.86 | 0.97 |  |
| GC | A1a | N-(2-Chloroethyl)-3-methoxybenzamide | 100.0 | 0.00 | 1.25 | 0.37 | 0.86 |  |
| GC | A1a | Octyl decyl phthalate | 100.0 | 0.38 | 0.93 | 0.52 | 0.90 |  |
| GC | A1a | [(2S)-2-[(1R,3aR,4S,7aR)-4-hydroxy-7a-methyl-1,2,3,3a,4,5,6,7-octahydroinden-1-yl]propyl] 4-methylbenzenesulfonate | 100.0 | 0.55 | 1.15 | 0.79 | 0.97 | 0.97 |
| GC | A1a | 4-methoxy-1-[methoxy(phenyl)methyl]-2-nitro-benzene | 100.0 | 0.09 | 0.85 | 0.14 | 0.77 |  |
| GC | A1a | Ibuprofen | 100.0 | 0.55 | 0.98 | 0.87 | 0.97 | 0.97 |
| GC | A1a | (+-)-5-[2-(7-Methoxyindol-3-yl)ethyl]-2,3,3a.beta.,6,7,7a.beta.-hexahydrofuro[3,2-c]pyridin-4(5H)-one | 100.0 | 0.41 | 1.13 | 0.54 | 0.91 |  |
| GC | A1a | 2,4,4'-Trichlorobiphenyl | 100.0 | 0.47 | 1.15 | 0.53 | 0.90 |  |
| GC | A1a | (E)-1-phenyl-4-hexen-1-one | 100.0 | 0.03 | 1.21 | 0.63 | 0.93 |  |
| GC | A1a | 2,2-Dimethyl-5-(prop-2'-ynyl)-1,3-dioxane-4,6-dione | 100.0 | 0.55 | 1.05 | 0.84 | 0.97 | 0.97 |
| GC | A1a | 2,5-dihydroxy-4-prop-2-enylbenzaldehyde | 100.0 | 0.27 | 0.95 | 0.76 | 0.96 |  |
| GC | A1a | 2(S)-Amino-3-methyl-N,N-dioctylbutyramide | 100.0 | 0.02 | 0.82 | 0.22 | 0.81 |  |
| GC | A1a | Ethyl (3-Methyl-6-oxo-1-phenyl-4,5,6,7-tetrahydro-1H-pyrazolo[3,4-b]pyridine-4-carboxamido)benzoate | 100.0 | 0.47 | 1.00 | 1.00 | 1.00 |  |
| GC | A1a | 3-(Chloromethyl)-4-hydroxy-4-phenyl-2-butanone | 100.0 | 0.58 | 0.99 | 0.96 | 0.98 | 0.98 |
| GC | A1a | 3-(Dimethylphenylsilyl)-2-methylpropionamide | 100.0 | 0.01 | 0.77 | 0.10 | 0.77 |  |
| GC | A1a | 4-tert-butyl-4-(1-methoxy-2-methyl-propan-2-yl)-1-methyl-azetidine-2,3-dione | 100.0 | 0.24 | 0.83 | < 0.05* | 0.64 |  |
| GC | A1a | (5S,8S)-8-Isopropyl-3-methoxy-5-methyl-1,2,3,4-tetrahydronaphthalen-2-carbaldehyde | 100.0 | 0.31 | 1.89 | < 0.05* | 0.72 |  |
| GC | A1a | 5-p-anisyloxypentan-1-ol | 100.0 | 0.23 | 1.80 | < 0.05* | 0.72 |  |
| GC | A1a | (2E,4E)-3-Phenylthiopenta-2,4-dienal | 100.0 | 0.47 | 0.75 | 0.14 | 0.77 |  |
| GC | A1a | (1R,2R)-1,2-diphenylethane-1,2-diol | 100.0 | 0.14 | 0.83 | 0.46 | 0.89 |  |
| GC | A1a | 4-Methylbenzyl vinyl ether | 100.0 | 0.73 | 1.03 | 0.82 | 0.97 | 0.97 |
| GC | A1a | 1-O-hexyl 2-O-propan-2-yl benzene-1,2-dicarboxylate | 100.0 | 0.73 | 1.22 | 0.19 | 0.81 | 0.79 |
| GC | A1a | methyl 2-methylbenzimidazole-1-carboxylate | 100.0 | 0.18 | 0.82 | 0.14 | 0.77 |  |
| GC | A1a | 3-Piperidin-1-yl-butan-2-ol | 100.0 | 0.43 | 0.97 | 0.63 | 0.93 |  |
| GC | A1a | (-)-(R,E)-5-(tert-Butylsulfonyl)hept-3-ene | 100.0 | 0.11 | 0.94 | 0.82 | 0.97 |  |
| GC | A1a | (Z)-4-Methylphenylazo tert-butyl sulfide | 100.0 | 0.52 | 3.19 | < 0.05* | 0.49 | 0.17 |
| GC | A1a | 10-Formyl-1,2,3,4,5,6-hexahydro-1,5-imino-9-methoxy-3,8,11-trimethyl-4-oxo-3-benzazocine | 100.0 | 0.66 | 0.92 | 0.60 | 0.92 | 0.95 |
| GC | A1a | 1-hydroxy-3-methyl-2-pyrrolecarbonitrile | 100.0 | 0.42 | 2.12 | < 0.05* | 0.51 |  |
| GC | A1a | (E)-2,2,5-triphenyl-4-pentenenitrile | 100.0 | 0.34 | 0.82 | 0.55 | 0.91 |  |
| GC | A1a | 4-Pentylphenol | 100.0 | 0.68 | 1.74 | 0.25 | 0.82 | 0.82 |
| GC | A1a | 2-[(E)-hydroxyiminomethyl]phenol | 100.0 | 0.58 | 1.28 | 0.67 | 0.94 | 0.95 |
| GC | A1a | Bis(2-ethylhexyl) adipate | 100.0 | 0.68 | 0.80 | 0.38 | 0.86 | 0.87 |
| GC | A1a | 4'-[(Methanesulfonyloxy)methyl]-[2,2' ; 6',6\]-terpyridyl"" | 100.0 | 0.66 | 1.17 | 0.53 | 0.90 | 0.93 |
| GC | A1a | (2S,3R,4R)-4-tert-butyl-2-phenyl-3-oxolanol | 100.0 | 0.52 | 0.88 | 0.47 | 0.89 | 0.92 |
| GC | A1a | 2-(2-nitrophenyl)-1H-benzimidazole | 100.0 | 0.77 | 1.13 | 0.84 | 0.97 | 0.97 |
| GC | A1a | 2-N-[2'-(Methoxymethyl)pyrrolidin-1'-yl]amino-3-methylheptane | 46.7 | 0.14 | 1.07 | 0.22 | 0.81 |  |
| GC | A1a | 1H-3,1-benzoxazine-2,4-dione | 91.7 | 0.55 | 0.91 | 0.16 | 0.79 | 0.72 |
| GC | A1a | 8-Methyl-8-(hydroxymethyl)-4,5,7,8-tetrahydro-6H-cyclohepta[b]furan | 67.8 | 0.29 | 0.98 | 0.81 | 0.97 |  |
| GC | A1a | 2-methyl-1H-pyrrole-3-carbonitrile | 96.0 | 0.42 | 1.03 | 0.68 | 0.94 |  |
| GC | A1a | 5,16-androstadiene-3.beta.-ol-acetate | 78.5 | 0.08 | 1.02 | 0.60 | 0.92 |  |
| GC | A1a | [1,2,4]Triazolo[1,5-a][1,3,5]triazin-7-amine, 1,2-dihydro-2-methyl-2-phenyl- | 98.7 | 0.60 | 1.09 | 0.11 | 0.77 | 0.67 |
| GC | A1a | (2R,3S)-{3-[4-[(Tetrahydropyran-2-yl)oxy]but-2(E)-enyl]oxiranyl}methanol | 84.1 | 0.06 | 1.01 | 0.88 | 0.97 |  |
| GC | A1a | 3-cyclohexyl-5-phenyl-1,3-oxazolidine-2,4-dione | 44.2 | 0.64 | 1.01 | 0.87 | 0.97 | 0.97 |
| GC | A1a | 2-Hydroxy-3-methylbicyclo[2.2.1]heptane-2-methanol | 99.7 | 0.16 | 1.07 | 0.71 | 0.95 |  |
| GC | A1a | 16,17-Epoxy-6,7-isopropylidenedioxy-1,15-dihydroxy-8,15-secomaoerystals A | 58.1 | 0.05 | 0.99 | 0.81 | 0.97 |  |
| GC | A1a | 2,5-Dichlorophenol | 84.9 | 0.44 | 0.85 | 0.07 | 0.75 |  |
| GC | A1a | 2-methyl-1-acenaphthylenecarbonitrile | 83.6 | 0.02 | 1.06 | 0.18 | 0.81 |  |
| GC | A1a | 10,11-Dimethoxy-12b-trimethylsilyl-5,6,7,8,12b,13a-hexahydrodibenzo[c,g]oxireno[2,3-e]azecin-5-one | 60.1 | 0.07 | 1.08 | 0.30 | 0.82 |  |
| GC | A1a | 1-(4-Chlorophenyl)-2-octanone | 59.5 | 0.24 | 0.96 | 0.07 | 0.75 |  |
| GC | A1a | (-)-2.beta.-Methoxy-2a.beta.,3,4,6,7,8,8a.beta.,8b.beta.-Octahydro-6,6,8b.beta.-trimethyl-2H-naphtho[1,8-bc]furan | 69.0 | 0.19 | 1.02 | 0.86 | 0.97 |  |
| GC | A1a | Ethyl 2,4-dimethylpyrido[2,3-a]benzofuran-3-carboxylate | 99.6 | 0.09 | 1.15 | < 0.05* | 0.72 |  |
| GC | A1a | [3'-Phenyl-2'-propenyl ] Allyl Ether | 99.9 | 0.53 | 0.95 | 0.66 | 0.93 | 0.95 |
| GC | A1a | l-Norleucine, N-methoxycarbonyl-, isohexyl ester | 99.4 | 0.56 | 0.95 | 0.66 | 0.93 | 0.95 |
| GC | A1a | 2,7-Dimethyl-10-n-propyl-4,5-dioxo-3,6,9-trioxa-3,4,5,6,9,10-hexahydroanthracene | 99.6 | 0.22 | 1.25 | 0.05 | 0.74 |  |
| GC | A1a | Benzo[b]furo[3,4-f][1,4]benzodioxin-1,3-dione, 3a,4,11a,11b-tetrahydro-, (3a.alpha.,11a.alpha.,11b.alpha.)- | 54.2 | 0.55 | 1.04 | 0.26 | 0.82 | 0.82 |
| GC | A1a | (1R*,2R*,5R*)-sec-butyl 5-hydroxy-2-methylcyclohexanecarboxylate | 78.6 | 0.29 | 0.94 | 0.14 | 0.77 |  |
| GC | A1a | 2,4,6-Tribromophenol | 89.3 | 0.37 | 0.93 | 0.19 | 0.81 |  |
| GC | A1a | 2,3-Dihydro-10-methyl-1H,7H,8H-benzo[ij]pyrano[3,2-c]]quinolizine-7,8-dione | 40.8 | 0.11 | 0.99 | 0.85 | 0.97 |  |
| GC | A1a | 1-(4'-Fluoro-2'-hydroxyphenyl)-2,2-dimethylpropan-1-one | 99.7 | 0.69 | 1.04 | 0.60 | 0.92 | 0.95 |
| GC | A1a | 2-Butyl-3-phenyl-5-isopropyl-4-pentylfuran | 77.7 | 0.29 | 1.01 | 0.90 | 0.97 |  |
| GC | A1a | 2,4,6,8-Tetramethylazulene | 92.1 | 0.53 | 1.09 | < 0.05* | 0.72 | 0.48 |
| GC | A1a | 2,2'-Diihydroxy-4'-methyl-5-methoxybiphenyl | 99.9 | 0.60 | 1.00 | 0.92 | 0.97 | 0.97 |
| GC | A1a | 9,10-seco-8b,10-dihydroxy-12-oxocalanolide A | 98.5 | 0.36 | 0.95 | 0.57 | 0.91 |  |
| GC | A1a | 1,2-Dihydrocyclopentafluorene | 69.8 | 0.40 | 1.04 | 0.34 | 0.83 |  |
| GC | A1a | 7-Acetoxy-8-ethenylquinoline | 97.0 | 0.63 | 1.02 | 0.86 | 0.97 | 0.97 |
| GC | A1a | 3-Amino-4,5-bis(p-biphenyl)-1H-pyrazolo[3,4-c]pyridazine | 98.3 | 0.02 | 0.98 | 0.32 | 0.83 |  |
| GC | A1a | 6-(1',1\-Biphenyl-4'-yl)-1,4,8-trimethylazulene"" | 99.0 | 0.46 | 0.97 | 0.70 | 0.95 |  |
| GC | A1a | 5-(Isobutyl)-1,4-pyrazine-2-carbonitrile | 90.6 | 0.36 | 1.07 | 0.62 | 0.93 |  |
| GC | A1a | N-Benzylbenzenamine | 96.9 | 0.04 | 1.00 | 0.94 | 0.97 |  |
| GC | A1a | (4'R,5'R)-Spiro[1,5-benzodithiepane-3,2'-[4.5]diethoxycarbonyl[1,3]dioxolane] | 48.1 | 0.29 | 1.05 | 0.48 | 0.89 |  |
| GC | A1a | Benz[c]acridine-7(2H)-one, 1,3,4,12-tetrahydro- | 99.2 | 0.20 | 1.00 | 0.98 | 1.00 |  |
| GC | A1a | 6-Ethoxycarbonyl-1-phenyl-1H-pyrazolo[4,3-c]pyridine-4-thione | 97.4 | 0.22 | 0.91 | 0.31 | 0.82 |  |
| GC | A1a | 1-(tert-Butyldimethylsilyloxy)-3-vinylpent-4-en-2-ol | 99.7 | 0.47 | 0.88 | 0.49 | 0.89 |  |
| GC | A1a | N-{4-[N'-(4-hydroxybenzylidene)hydrazino]benzyl}-2,2,2-trifluoroacetamide | 81.8 | 0.68 | 1.03 | 0.63 | 0.93 | 0.95 |
| GC | A1a | Benzoic acid, 3-hydroxy-2-(tetrahydro-2H-pyran-2-yl)-, methyl ester | 99.4 | 0.42 | 0.91 | 0.29 | 0.82 |  |
| GC | A1a | 6-Isobutanoyl-7-methoxycoumarin | 94.9 | 0.26 | 0.94 | 0.12 | 0.77 |  |
| GC | A1a | 1-methyl-4-phenyl-2-[(E)-2-phenylethenyl]pyrrole | 99.9 | 0.38 | 1.17 | 0.36 | 0.85 |  |
| GC | A1a | 1-hydroxy-3-methoxy-2-(1-methoxyethyl)-9,10-anthraquinone | 92.7 | 0.44 | 0.97 | 0.67 | 0.94 |  |
| GC | A1a | 2-Methyl-4-phenylpyrido[2,3-d]pyrimidin-5(8H)-one | 99.9 | 0.37 | 0.94 | 0.56 | 0.91 |  |
| GC | A1a | 2-(Pyrrolidin-1-yl)-1-(5,6,7,8-tetrahydronaphthalen-2- yl)pentan-1-one | 97.1 | 0.40 | 0.95 | 0.39 | 0.86 |  |
| GC | A1a | 2,3-Dihydro-1-(methylene)pyrrolo[1,2-a]indole-9-carbaldehyde N,N-dimethylhydrazone | 96.9 | 0.03 | 1.03 | 0.74 | 0.96 |  |
| GC | A1a | 1-methyl-2-oxidanylidene-pyridine-3-carbaldehyde | 81.3 | 0.38 | 1.04 | 0.80 | 0.97 |  |
| GC | A1a | 8-methoxy-2H-1-benzopyran-3-carboxylic acid methyl ester | 45.5 | 0.60 | 1.04 | 0.36 | 0.85 | 0.84 |
| GC | A1a | 1,3-Benzenedicarbonitrile | 52.9 | 0.67 | 1.11 | < 0.001* | 0.18 | 0.05 |
| GC | A1a | 7-methylpyrrolo[1,2-a]pyrazin-1-amine | 99.9 | 0.15 | 1.07 | 0.52 | 0.90 |  |
| GC | A1a | 2-[(5-bromo-2-methoxybenzoyl)amino]acetic acid | 64.9 | 0.42 | 0.95 | 0.31 | 0.82 |  |
| GC | A1a | Isopropyl 3-methylquinoxaline-2-carboxylate | 94.8 | 0.33 | 0.98 | 0.71 | 0.95 |  |
| GC | A1a | p,p'-DDE | 99.9 | 0.74 | 1.08 | 0.14 | 0.77 | 0.70 |
| GC | A1a | 4-hydroxy-3,4-dimethylnaphthalen-1-one | 99.9 | 0.57 | 1.40 | 0.08 | 0.77 | 0.64 |
| GC | A1a | Butanedioic acid, [1-(acetyloxy)-1,2,2-trimethylpropyl]-, diethyl ester | 85.6 | 0.31 | 1.00 | 0.93 | 0.97 |  |
| GC | A1a | Diethyl L-tartrate | 97.3 | 0.11 | 1.00 | 1.00 | 1.00 |  |
| GC | A1a | 5-(t-Butyl)-3,3-dimethyl-4-(benzyl)thiophen-2(3H)-one | 99.9 | 0.21 | 1.17 | 0.17 | 0.81 |  |
| GC | A1a | 2-Mercaptobenzothiazole | 99.9 | 0.13 | 1.04 | 0.43 | 0.87 |  |
| GC | A1a | 3-(5,7-Dioxo-5,7-dihydro-pyrrolo[3,4-b]pyrazin-6-yl)-propionic acid ethyl ester | 87.9 | 0.01 | 1.01 | 0.87 | 0.97 |  |
| GC | A1a | 2-Oxahexacyclo[7,5,1.0.0(3,13).0(5,12).0(10,14)]pentadecan-3-ol | 56.3 | 0.28 | 1.10 | 0.12 | 0.77 |  |
| GC | A1a | 10H-Phenoxazine | 57.8 | 0.03 | 0.98 | 0.74 | 0.96 |  |
| GC | A1a | 2-(furan-3-yl)-2-(2-iodoethyl)-1,3-dioxolane | 99.7 | 0.27 | 0.83 | < 0.05* | 0.72 |  |
| GC | A1a | 7-isopropyl-3,4-dihydro-2H-cyclohepta[b]pyrazine | 98.9 | 0.60 | 1.12 | 0.11 | 0.77 | 0.67 |
| GC | A1a | Acetic acid, [[1-(cyanohydroxymethyl)-2-naphthalenyl]oxy]-, ethyl ester | 92.8 | 0.23 | 1.06 | 0.15 | 0.78 |  |
| GC | A1a | Tris(2-ethylhexyl) trimellitate | 99.0 | 0.18 | 0.96 | 0.72 | 0.96 |  |
| GC | A1a | 1-Piperidineacetaldehyde, .alpha.-methyl-.alpha.-[[(tetrahydro-2H-pyran-2-yl)oxy]methyl]- | 96.3 | 0.09 | 1.12 | 0.07 | 0.75 |  |
| GC | A1a | 3,4-Diethoxy-2,5-dimethoxytetrahydrofuran | 94.5 | 0.23 | 0.95 | 0.38 | 0.86 |  |
| GC | A1a | 1-[2-(E)-5-(3-(1-Hydroxycyclohex-1-yl)-2-propynylidene)-1-cyclopenten-1-yl]ethynyl]cyclohexan-1-ol | 99.9 | 0.04 | 0.90 | 0.40 | 0.86 |  |
| GC | A1a | (2S,3S,4S)-2-[2'-((S)-4''-Benzyl-2''-oxo-oxazolidin-3''-yl)-2-oxo-ethyl]-N-cyclohexyl-5-cyano-3,4-dimethyl-piperidine | 87.0 | 0.04 | 0.98 | 0.48 | 0.89 |  |
| GC | A1a | Triphenyl phosphate | 99.7 | 0.04 | 0.96 | 0.61 | 0.93 |  |
| GC | A1a | (4R,5R)-2,2-dimethyl-4-(2-phenylethynyl)-5-vinyl-1,3-dioxolane | 89.8 | 0.36 | 1.01 | 0.91 | 0.97 |  |
| GC | A1a | N-{2-[(4-Acetylphenyl)ethynyl]phenyl}-2,2,2-trifluoroacetamide | 99.9 | 0.07 | 1.03 | 0.74 | 0.96 |  |
| GC | A1a | (3R,3aR,6S,6aR)-3-(3,4-dimethoxyphenyl)-6-(3,4,5-trimethoxyphenyl)-1,3,3a,4,6,6a-hexahydrofuro[3,4-c]furan | 99.8 | 0.44 | 1.03 | 0.58 | 0.91 |  |
| GC | A1a | Methyl endo-9-acetyl-7-methyltricyclo[5.2.2.0(1,5)]undec-5-ene-6-carboxylate | 58.8 | 0.19 | 1.00 | 1.00 | 1.00 |  |
| GC | A1a | 2-ethoxy-3,1-benzoxazin-4-one | 99.8 | 0.12 | 1.04 | 0.79 | 0.97 |  |
| GC | A1a | 9-methyl-9-phenyl-9,10-dihydrophenanthrene | 74.8 | 0.83 | 1.00 | 0.95 | 0.98 | 0.98 |
| GC | A1a | (Z)-Phenylazo tert-butyl sulfide | 99.8 | 0.86 | 1.19 | 0.27 | 0.82 | 0.82 |
| GC | A1a | 4-Chloro-2-(6-chlorohex-1-yn-1-yl)pyridine | 88.4 | 0.67 | 1.02 | 0.63 | 0.93 | 0.95 |
| GC | A1a | Thiazolo[5,4-f]quinoline-2-carbonitrile | 98.7 | 0.47 | 1.09 | 0.17 | 0.81 |  |
| GC | A1a | 1,4,5,6-Tetrahydro-1-(4',6'-heptadienoyl)-pyridine | 97.4 | 0.09 | 0.94 | 0.59 | 0.92 |  |
| GC | A1a | (3S,5R,8aS)-3-butyl-5-propyl-1,2,3,5,6,7,8,8a-octahydroindolizine | 96.9 | 0.04 | 0.91 | 0.33 | 0.83 |  |
| GC | A1a | 4,6,7-trimethylinden-1-one | 97.1 | 0.44 | 1.22 | 0.07 | 0.75 |  |
| GC | A1a | methyl 1-butyl-2-oxocycloheptane-1-carboxylate | 99.7 | 0.37 | 1.06 | 0.72 | 0.96 |  |
| GC | A1a | 4-Acetoxy-10,12-dimethyl-14-oxatricyclo[7.4.0.0(6,7)]tetradecane | 99.4 | 0.39 | 0.95 | 0.34 | 0.83 |  |
| GC | A1a | 2,3,4,4a,5,6,6a,11b-Octahydro-6a-hydroperoxy-8-methoxy-7,9,10-trimethyl-1H-benzo[a]carbazole | 97.2 | 0.47 | 0.89 | 0.13 | 0.77 |  |
| GC | A1a | 7-Cyano-2-oxatricyclo[5.3.1.0(1,11)]undec-1(10)-en-9-one | 99.9 | 0.27 | 0.72 | 0.23 | 0.81 |  |
| GC | A1a | 1,2,4-Trichlorobenzene | 99.5 | 0.66 | 1.05 | 0.81 | 0.97 | 0.97 |
| GC | A1a | Apocynin | 99.3 | 0.04 | 0.85 | 0.22 | 0.81 |  |
| GC | A1a | 2-Acetoxy-3-chlorophenol | 72.0 | 0.81 | 0.96 | 0.35 | 0.84 | 0.84 |
| GC | A1a | 4,5-Dihydrofuro[1,2-a]naphthalene | 99.9 | 0.19 | 1.06 | 0.68 | 0.94 |  |
| GC | A1a | 2-tert-Butylfuro[4,5-d][1,3]dioxan-6-one | 97.7 | 0.77 | 1.24 | 0.11 | 0.77 | 0.67 |
| GC | A1a | 10-(p-Tolyl)benzo[h]quinoline | 78.3 | 0.30 | 1.00 | 0.88 | 0.97 |  |
| GC | A1a | Isoxaben | 99.2 | 0.53 | 0.95 | 0.52 | 0.90 | 0.93 |
| GC | A1a | 1-(5-Methyl-2-oxo-2,5-dihydrofuran-3-yl)-9-(tert-butyldimethylsiloxy)nonan-8-ol | 94.4 | 0.25 | 0.95 | 0.40 | 0.87 |  |
| GC | A1a | 3-butylcyclohexan-1-one | 94.4 | 0.02 | 0.95 | 0.57 | 0.91 |  |
| GC | A1a | 6,7-Dimethoxy-1-[(p-tolyl)sulfinyl]methyl-1-(trifluoromethyl)-1,2,3,4-tetrahydroisoquinoline | 99.9 | 0.44 | 1.03 | 0.85 | 0.97 |  |
| GC | A1a | 2-[Chloro(diphenyl)methyl]pyridine | 84.3 | 0.48 | 1.03 | 0.70 | 0.95 |  |
| GC | A1a | 1-[(1S,2R)-2-propoxycyclopentyl]hexane-1,2-dione | 74.7 | 0.12 | 1.08 | < 0.05* | 0.51 |  |
| GC | A1a | Dodecyltriethoxysilane | 99.8 | 0.32 | 0.99 | 0.93 | 0.97 |  |
| GC | A1a | N-(1-methyl-2,5-dioxo-3-[1]benzopyrano[4,3-b]pyridinyl)acetamide | 98.2 | 0.03 | 0.89 | 0.25 | 0.82 |  |
| GC | A1a | 6-Phenyl-2-pyridinamine | 76.5 | 0.54 | 1.10 | 0.05 | 0.74 | 0.55 |
| GC | A1a | 4-(3-ethoxyphenyl)-N,N-diethyl-aniline | 67.5 | 0.50 | 1.05 | 0.29 | 0.82 | 0.82 |
| GC | A1a | (4R,4aR,8aR)-4,8a-dimethyl-1,2,3,4,7,8-hexahydronaphthalen-4a-ol | 99.7 | 0.28 | 0.84 | 0.33 | 0.83 |  |
| GC | A1a | Padimate O | 71.2 | 0.20 | 1.02 | 0.65 | 0.93 |  |
| GC | A1a | (2E)-2-(phenylmethylene)-3-benzofuranone | 69.1 | 0.01 | 0.93 | 0.28 | 0.82 |  |
| GC | A1a | methyl 2-(4-methoxyphenyl)propanoate | 91.2 | 0.64 | 0.98 | 0.69 | 0.95 | 0.96 |
| GC | A1a | (8-acetoxy-1-cyano-7-ethoxy-6-methyl-5-isoquinolyl) acetate | 66.8 | 0.27 | 0.90 | < 0.05* | 0.72 |  |
| GC | A1a | 15-Nonacosanone | 85.9 | 0.20 | 0.95 | 0.34 | 0.83 |  |
| GC | A1a | 1,4:5,8-Dimethanonaphthalene, 6-butyl-1,2,3,4,4a,5,8,8a-octahydro-, (1.alpha.,4.alpha.,4a.beta.,5.beta.,8.beta.,8a.beta.)- | 97.7 | 0.17 | 1.12 | 0.26 | 0.82 |  |
| GC | A1a | (Z)-2-Ethyl-5-bromo-2-dicyanomethyladamantane | 59.3 | 0.08 | 1.04 | 0.26 | 0.82 |  |
| GC | A1a | 3-(5,5,6-Trimethylbicyclo[2.2.1]hept-2-ylidene)-2-propanone oxime | 99.9 | 0.02 | 1.02 | 0.89 | 0.97 |  |
| GC | A1a | (3,6-dimethyl-4-oxidanylidene-2H-chromen-3-yl) nitrate | 91.5 | 0.01 | 1.44 | < 0.05* | 0.58 |  |
| GC | A1a | 6,7-dimethoxy-1-(methoxymethyl)-3,4-dihydroisoquinoline | 99.9 | 0.07 | 1.04 | 0.53 | 0.90 |  |
| GC | A1a | Methyl (E)-3-methoxy-4-oxo-4-phenylbut-2-enoate | 87.7 | 0.26 | 0.94 | 0.39 | 0.86 |  |
| GC | A1a | 3,3,7-trimethyl-5-nitro-1H-indol-2-one | 83.1 | 0.68 | 1.00 | 0.98 | 0.99 | 0.98 |
| GC | A1a | 1-hydroxy-1-methyl-tetralin-6-carbaldehyde | 99.9 | 0.53 | 1.68 | < 0.05* | 0.72 | 0.55 |
| GC | A1a | Acetosyringon | 51.6 | 0.16 | 0.97 | 0.44 | 0.87 |  |
| GC | A1a | Methyl (S)-1-(4,4-diphenylbut-3-en-1-yl)pyrrolidine-2-carboxylate | 99.9 | 0.26 | 0.96 | 0.77 | 0.96 |  |
| GC | A1a | Octabenzone | 97.5 | 0.36 | 0.99 | 0.92 | 0.97 |  |
| GC | A1a | Diphenyl(1,3,5-trimethyl-1H-pyrazol-4-yl)amine | 98.7 | 0.36 | 1.08 | 0.22 | 0.81 |  |
| GC | A1a | Octicizer | 91.4 | 0.02 | 1.01 | 0.91 | 0.97 |  |
| GC | A1a | 6-Methoxy-2-phenyl-2-(phenylthio)-3-benzofuranone | 92.8 | 0.02 | 0.97 | 0.75 | 0.96 |  |
| GC | A1a | 2-Acetonyl-3-acetyl-5-methoxy-1,4-naphthoquinone | 82.0 | 0.65 | 1.03 | 0.48 | 0.89 | 0.92 |
| GC | A1a | Bumetrizole | 95.8 | 0.17 | 1.24 | 0.07 | 0.75 |  |
| GC | A1a | (2SR,3aRS,6aRS,10aRS)-1,7,7,10a-Tetramethyl-2-phenyl-2,3,3a,6,6a,7,8,9,10,10a-decahydro-4-oxabenz[e]azulen-5-one | 75.8 | 0.25 | 1.02 | 0.84 | 0.97 |  |
| GC | A1a | Tri(2-ethylhexyl) phosphate | 98.1 | 0.23 | 1.06 | 0.48 | 0.89 |  |
| GC | A1a | 2-[(E)-pentadec-1-enyl]benzene-1,3-dicarbaldehyde | 53.3 | 0.17 | 1.04 | 0.42 | 0.87 |  |
| GC | A1a | 1-(6-Methoxypyridin-2-yl)propan-2-one | 99.5 | 0.39 | 1.16 | 0.21 | 0.81 |  |
| GC | A1a | (R)-4-Isobutyloxazolidin-2-one | 99.8 | 0.37 | 1.14 | 0.28 | 0.82 |  |
| GC | A1a | methyl 2-(2,4,6-trimethylphenyl)acetate | 98.1 | 0.01 | 0.95 | 0.65 | 0.93 |  |
| GC | A1a | 2-[(4-Chlorobenzoyl)(2-ethoxycarbonylbenzyl)methylene]-1H-imidazole | 65.3 | 0.26 | 1.05 | 0.27 | 0.82 |  |
| GC | A1a | 3-(2-Chloromethylphenyl)-2,2-dimethylpropionaldehyde | 99.7 | 0.22 | 0.93 | 0.66 | 0.93 |  |
| GC | A1a | (+)-(2S,3R)-4-Benzyloxy-3-(tert-butyldimethylsiloxy)-2-methylbutanol | 84.1 | 0.54 | 1.11 | 0.13 | 0.77 | 0.69 |
| GC | A1a | cis-3,5-[di(t-Butyl)silanedioxy]cyclopent-1-ene | 76.9 | 0.87 | 1.04 | 0.41 | 0.87 | 0.88 |
| GC | A1a | 4-hydroperoxy-2,3,4-trimethylnaphthalen-1-one | 69.4 | 0.19 | 0.94 | 0.21 | 0.81 |  |
| GC | A1a | Ethanethioic acid, S-bicyclo[2.2.1]hept-2-en-7-yl ester, anti- | 79.1 | 0.37 | 0.98 | 0.75 | 0.96 |  |
| GC | A1a | 1-(benzo[d][1,3]dioxol-5-yl)-2-(4-methylpiperidin-1-yl)pentan-1-one | 99.7 | 0.36 | 1.04 | 0.65 | 0.93 |  |
| GC | A1a | Propylparaben | 99.9 | 0.44 | 1.04 | 0.56 | 0.91 |  |
| GC | A1a | 2,4-Dihydroxy-5-methoxy-6-methyl-tetrahydropyran | 93.3 | 0.24 | 1.15 | 0.11 | 0.77 |  |
| GC | A1a | Octinoxate | 99.5 | 0.21 | 1.01 | 0.91 | 0.97 |  |
| GC | A1a | (Z)-13-Ethylene-9,10-dimethoxy-2,3-(methylenedioxy)-8,14-cycloberbine | 85.8 | 0.41 | 0.99 | 0.87 | 0.97 |  |
| GC | A1a | 1,7-Dimethylxanthine | 60.7 | 0.22 | 0.99 | 0.73 | 0.96 |  |
| GC | A1a | 5-carboxylated pyrrole derivative of 4,4-dimethyl-2-(N-methylpyrrol-2-yl)oxazoline | 99.9 | 0.92 | 1.04 | 0.73 | 0.96 | 0.97 |
| GC | A1a | Pestalolide | 99.9 | 0.53 | 0.71 | 0.27 | 0.82 | 0.82 |
| GC | A1a | Methyl (1R*,5S*)-1-Hydroxy-1-methylspiro[4.5]dec-7-ene-8-carboxylate | 95.8 | 0.08 | 1.04 | 0.43 | 0.87 |  |
| GC | A1a | (3-methylphenyl) 3-nitrobenzoate | 93.5 | 0.40 | 1.05 | 0.31 | 0.82 |  |
| GC | A1a | 2-(3-furanyl)-1H-benzimidazole | 53.1 | 0.56 | 1.09 | 0.33 | 0.83 | 0.84 |
| GC | A1a | (1R,3S)-3-phenylcyclohexanol | 99.9 | 0.44 | 0.93 | 0.49 | 0.89 |  |
| GC | A1a | 9-[1-(3-methylphenyl)ethylidene]fluorene | 63.8 | 0.48 | 1.02 | 0.31 | 0.82 |  |
| GC | A1a | 2,3,3',4,4',5'-Hexachlorobiphenyl | 96.1 | 0.58 | 1.06 | 0.22 | 0.81 | 0.82 |
| GC | A1a | (2-iodanyl-2-triethylsilyl-ethyl) 2,2,2-tris(fluoranyl)ethanoate | 95.9 | 0.03 | 0.95 | 0.63 | 0.93 |  |
| GC | A1a | 4-ethyl-4-methyl-5-methylene-1,3-dioxolan-2-one | 99.4 | 0.34 | 0.92 | 0.56 | 0.91 |  |
| GC | A1a | (4aS,8aS)-5,5,8a-trimethyl-1-methylene-3,4,4a,6,7,8-hexahydronaphthalen-2-one | 99.9 | 0.17 | 0.85 | < 0.05* | 0.62 |  |
| GC | A1a | Dibutyl phthalate | 97.2 | 0.64 | 1.20 | < 0.05* | 0.58 | 0.30 |
| GC | A1a | 7-Methoxy-2,5-dimethyl-chromone | 96.2 | 0.61 | 0.98 | 0.59 | 0.92 | 0.95 |
| GC | A1a | (R)-(S)-.alpha.-Methoxyphenylacetic acid (NPA) | 99.4 | 0.41 | 0.88 | 0.08 | 0.76 |  |
| GC | A1a | 4-Ethyl-3-methyl-1,2,4-thiadiazol-5-one | 95.3 | 0.04 | 1.04 | 0.74 | 0.96 |  |
| GC | A1a | 3-Aminochrysene | 99.9 | 0.31 | 1.28 | 0.11 | 0.77 |  |
| GC | A1a | 2-(tert-Butyl-dimethyl-silanyloxy)-3-methyl-cyclopent-2-enone | 61.7 | 0.15 | 1.08 | 0.15 | 0.79 |  |
| GC | A1a | 10-(3-Acetoxyprop-2-yl)-3,7-dimethylbicyclo[4.4.0]dec-2-ene | 99.7 | 0.62 | 0.93 | 0.27 | 0.82 | 0.82 |
| GC | A1a | (Z)-2-Phenyl-1-(triethylsilyl)-1,4-pentadiene | 99.7 | 0.03 | 1.08 | 0.65 | 0.93 |  |
| GC | A1a | (2,4-dimethoxy-3-methyl-phenyl) acetate | 68.2 | 0.30 | 0.99 | 0.30 | 0.82 |  |
| GC | A1a | Niloticin | 52.2 | 0.39 | 1.07 | 0.20 | 0.81 |  |
| GC | A1a | Methyl 4-[2-(2-Chloro-3-cyanopyrid-4-yl)ethyl]benzoate | 79.4 | 0.28 | 1.00 | 0.98 | 0.99 |  |
| GC | A1a | 1,4-Dideoxy-1,4-imino-D-arabinitol | 99.6 | 0.01 | 0.98 | 0.77 | 0.96 |  |
| GC | A1a | Triisobutyl phosphate | 99.7 | 0.64 | 0.90 | 0.64 | 0.93 | 0.95 |
| GC | A1a | (2'R*,3'R*,.alpha.S)-2,3-Epoxyoctyl .alpha.-methoxyphenylacetate | 99.8 | 0.06 | 0.82 | < 0.05* | 0.72 |  |
| GC | A1a | 5-Prop-2-ynyl-cyclohex-1-enylamine | 98.5 | 0.15 | 1.02 | 0.88 | 0.97 |  |
| GC | A1a | 2-{[Dimethyl(3-methylbut-2-en-1-yl)silyl]methyl}allyl Acetate | 48.4 | 0.16 | 0.91 | 0.22 | 0.81 |  |
| GC | A1a | 7a-Carboxymethyl-1-methylene-1,4,5,6,7,7a-hexahydro-2H-indene | 96.7 | 0.34 | 0.89 | 0.28 | 0.82 |  |
| GC | A1a | (5S,6R)-5,6-dimethyl-3,4,5,6-tetrahydro-2H-cyclopenta[b]pyran-7-one | 99.4 | 0.68 | 1.06 | 0.32 | 0.83 | 0.84 |
| GC | A1a | 9-phenylcarbazole | 88.4 | 0.17 | 1.06 | 0.21 | 0.81 |  |
| GC | A1a | (S)-2-(4-Methoxyphenyl)-3-methylbutan-1-ol | 89.4 | 0.11 | 1.09 | 0.42 | 0.87 |  |
| GC | A1a | 1,4-Naphthalenedione, 5-[(acetyloxy)methyl]-4a,5-dihydro-2-methoxy-4a-methyl-, trans- | 97.0 | 0.08 | 1.11 | 0.26 | 0.82 |  |
| GC | A1a | 7-Phenylethyl-4,6-dioxa-5-carbonyl-spiro[2,5]-7-octene | 93.2 | 0.51 | 0.94 | 0.23 | 0.81 | 0.82 |
| GC | A1a | 1-Cyano-1-(t-butyl)-2-aminoethene | 99.3 | 0.43 | 1.23 | 0.14 | 0.77 |  |
| GC | A1a | 2-Carboxy-4-methylbicyclo[2.2.2]oct-2-en-1-ol | 99.7 | 0.64 | 0.68 | < 0.05* | 0.54 | 0.26 |
| GC | A1a | 3-isopropyl-2-(9H-xanthen-9-yl)-1H-indole | 52.9 | 0.05 | 1.03 | 0.48 | 0.89 |  |
| GC | A1a | 5,5-diethyl-1,3-diazinane-2,4,6-trione | 93.3 | 0.05 | 0.92 | 0.42 | 0.87 |  |
| GC | A1a | 6-Deoxy-1,2-O-isopropylidene-5-O-thiocarbonyl-.alpha.,D-glucofuranose | 95.5 | 0.42 | 1.02 | 0.45 | 0.88 |  |
| GC | A1a | 4,5-dimethyl-3-(2-methylpropyl)-2,3-dihydroisoindol-1-one | 97.4 | 0.10 | 1.18 | 0.11 | 0.77 |  |
| GC | A1a | (E)-Methyl undec-2-en-4-ynoate | 59.0 | 0.01 | 1.00 | 0.89 | 0.97 |  |
| GC | A1a | Tributyl citrate | 89.0 | 0.31 | 1.02 | 0.64 | 0.93 |  |
| GC | A1a | Ethyl 3-phenyl-3-(2\,4\""-dinitrophenylhydrazono)-2-(1'-methylpiperidin-2'-ylidene)propionate"" | 78.7 | 0.08 | 0.97 | 0.21 | 0.81 |  |
| GC | A1a | 3-[tert-butyl(diphenyl)silyl]prop-2-yn-1-ol | 82.5 | 0.37 | 0.94 | < 0.05* | 0.72 |  |
| GC | A1a | (3Z,9Z)-3,9-Dodecadiene-2,5,8,11-tetraone | 96.7 | 0.35 | 0.96 | 0.39 | 0.86 |  |
| GC | A1a | 5-Acetoxy-2,7-dimethylchromone | 99.9 | 0.36 | 0.94 | 0.77 | 0.96 |  |
| GC | A1a | 4-octoxybenzaldehyde | 57.4 | 0.08 | 0.94 | 0.68 | 0.94 |  |
| GC | A1a | 3-(cyclopenten-1-yl)prop-2-ynyl methanesulfonate | 94.6 | 0.03 | 1.01 | 0.93 | 0.97 |  |
| GC | A1a | 3-hydroxy-3-(2-methoxycyclohexyl)-2-methylisoindol-1-one | 56.2 | 0.03 | 1.06 | 0.13 | 0.77 |  |
| GC | A1a | Ethyl 3-nitro-4,7-dihydro-4,7-ethano-2H-isoindole-1-carboxylate | 82.3 | 0.82 | 1.09 | < 0.05* | 0.72 | 0.55 |
| GC | A1a | 2,4-Dimethoxy-6-methylphenol | 99.8 | 0.34 | 0.99 | 0.86 | 0.97 |  |
| GC | A1a | (1,1,1-trichloro-2-methoxypropan-2-yl)oxycyclohexane | 56.6 | 0.14 | 0.97 | 0.76 | 0.96 |  |
| GC | A1a | Piperine | 99.8 | 0.40 | 0.96 | 0.17 | 0.81 |  |
| GC | A1a | 2-Oxo-2H-chromen-7-yl Acetate | 99.8 | 0.53 | 1.29 | 0.07 | 0.75 | 0.62 |
| GC | A1a | 2-[(3R)-3,7-dimethyloct-6-enylidene]malononitrile | 77.2 | 0.68 | 1.02 | 0.56 | 0.91 | 0.93 |
| GC | A1a | 5-tert-butyl-2-(4-tert-butylphenyl)-1,3-dinitro-benzene | 95.2 | 0.41 | 1.13 | 0.21 | 0.81 |  |
| GC | A1a | Furo[3',4':6,7]naphtho[2,3-d]-1,3-dioxole, 5,5a,6,8,8a,9-hexahydro-, (5.alpha.,5a.beta.,8a.beta.)- | 94.4 | 0.40 | 0.98 | 0.80 | 0.97 |  |
| GC | A1a | Thiophen-3-yl 2-aminobenzoate | 74.6 | 0.13 | 1.02 | 0.78 | 0.96 |  |
| GC | A1a | 3,4-Dihydro-5,8-dimethoxy-1,3-dimethyl-1H-2-benzopyran | 99.8 | 0.31 | 1.05 | 0.68 | 0.94 |  |
| GC | A1a | 2-Amino-3-cyano-6-methyl-4H-chromene | 42.1 | 0.60 | 1.01 | 0.86 | 0.97 | 0.97 |
| GC | A1a | (1S,3S)-2-iodanyl-2,3-dihydro-1H-indene-1,3-diol | 79.2 | 0.14 | 0.98 | 0.78 | 0.96 |  |
| GC | A1a | 1-(Phenylethynyl)-1-benzylcyclopropane | 92.1 | 0.07 | 1.06 | 0.50 | 0.89 |  |
| GC | A1a | Bromoxynil | 46.4 | 0.41 | 0.97 | 0.43 | 0.87 |  |
| GC | A1a | (6S)-6-undecyl-2-oxanone | 96.0 | 0.12 | 0.90 | 0.21 | 0.81 |  |
| GC | A1a | 1,1-Diformylcyclohex-3-ene | 99.8 | 0.64 | 1.07 | 0.73 | 0.96 | 0.97 |
| LC | A1b | Potentially chlorinated | 26.3 | NA | 0.90 | 0.82 | 0.98 |  |
| GC | A1b | 2H-Furo[2,3-h]-1-benzopyran-2-one | 27.4 | NA | 0.92 | 0.62 | 0.84 |  |
| GC | A1b | Aceclofenac | 26.5 | NA | 1.29 | 0.14 | 0.73 |  |
| GC | A1b | 6-phenyl-2,3,4,5-tetrahydropyridine | 39.6 | NA | 1.08 | 0.55 | 0.84 |  |
| GC | A1b | 7-Methyl-8-hydroxy-9-oxo-1-azabicyclo[4.3.0]non-7-ene | 25.7 | NA | 0.85 | 0.34 | 0.75 |  |
| GC | A1b | 2-(6-bromo-1,3-benzodioxol-5-yl)-1-(2-bromophenyl)ethanone | 31.6 | NA | 1.35 | < 0.05* | 0.62 |  |
| GC | A1b | 3-(Methylthio)propyl isothiocyanate | 27.7 | NA | 1.10 | 0.44 | 0.78 |  |
| GC | A1b | 4,5-Dichloro-10-(4-nitrobenzyl)-10H-anthracen-9-one | 15.7 | NA | 0.84 | 0.40 | 0.75 |  |
| GC | A1b | 2,3,6-trimethoxydibenz[b,f]oxepine | 38.4 | 0.62 | 1.14 | 0.40 | 0.75 | 0.40 |
| GC | A1b | Hydrastine | 12.2 | NA | 0.70 | 0.18 | 0.73 |  |
| GC | A1b | 1-methyl-12-phenyl-12-azatricyclo[6.3.1.02,7]dodeca-2,4,6,10-tetraen-9-one | 38.9 | NA | 0.93 | 0.59 | 0.84 |  |
| GC | A1b | ethyl 2-[6-(dimethoxymethyl)-1,3-benzodioxol-5-yl]-2-oxoacetate | 18.3 | NA | 1.18 | 0.26 | 0.73 |  |
| GC | A1b | 6-(2-pyridinyl)-2-pyridinecarboxaldehyde | 39.1 | 0.03 | 1.05 | 0.70 | 0.92 |  |
| GC | A1b | (1E,2E)-1-{2-[3-(Trifluoromethyl)phenyl]hydrazono}non-2-en-4-ol | 40.0 | 0.27 | 0.84 | 0.22 | 0.73 |  |
| GC | A1b | Methyl 4'-(1,1-dimethylprop-2-ynyloxy)-2'-methoxycinnamate | 29.3 | NA | 0.88 | 0.40 | 0.75 |  |
| GC | A1b | 6-Bromo-3-(3,4-dimethoxyphenethyl)quinazoline-2,4(1H,3H)-dione | 15.9 | NA | 1.02 | 0.92 | 1.00 |  |
| GC | A1b | 2-(2-methoxyethyl)-3,4-dihydronaphthalen-1(2H)-one | 17.8 | NA | 1.21 | 0.27 | 0.73 |  |
| GC | A1b | 2-(3-Nitrophenyl)-2,3-dihydro-1H-quinazolin-4-one | 27.5 | NA | 1.17 | 0.58 | 0.84 |  |
| GC | A1b | 5-(But-3'-en-1'-yl)-6-methylizidine | 20.2 | NA | 1.00 | 1.00 | 1.00 |  |
| GC | A1b | 2,6-Bis(4-t-butylphenyl)phenol | 13.0 | NA | 1.53 | 0.17 | 0.73 |  |
| GC | A1b | 4-[2-(1,3-Dithianyl)]-4-(2-ethoxycarbonylpyrrol-4-yl)butyric acid | 16.0 | NA | 0.82 | 0.30 | 0.75 |  |
| GC | A1b | anti-14-bromo-5,8-dimethoxy-16-methyl-2,11-dithia[3]paracyclo[3](2,4)thiophenophane | 20.8 | NA | 1.00 | 1.00 | 1.00 |  |
| GC | A1b | spiro[2,2,Dicyano-4-[(3-vinyl)phenyl]cyclobutane-1,1'-bis[1,2,5]thiadiazolodicyanocyclohexanemethane | 32.5 | NA | 0.80 | 0.12 | 0.73 |  |
| GC | A1b | 3-Iodophenol | 29.2 | NA | 1.65 | < 0.05* | 0.64 |  |
| GC | A1b | 2-(1-adamantyl)-3,3-bis(trifluoromethyl)spiro[1,4,2-dithiazolidine-5,2'-adamantane] | 34.6 | NA | 1.04 | 0.76 | 0.94 |  |
| GC | A1b | Diflubenzuron | 21.5 | NA | 0.99 | 0.94 | 1.00 |  |
| GC | A1b | (S,R)-N-(1-Methyl-3-butynyl)-N-(1-phenylethyl)amine | 24.9 | NA | 0.76 | 0.20 | 0.73 |  |
| GC | A1b | 3-Adamantyl-5-(t-butyl)-3-hydroxybenzaldehyde | 28.8 | NA | 0.86 | 0.24 | 0.73 |  |
| GC | A1b | N-(1,3-Diphenyl-2-propynyl)-phenyl carbamate | 18.5 | NA | 1.02 | 0.92 | 1.00 |  |
| GC | A1b | spiro[2,2,Dicyano-4-[(4-vinyl)phenyl]cyclobutane-1,1'-bis[1,2,5]thiadiazolodicyanocyclohexanemethane | 23.0 | NA | 0.92 | 0.60 | 0.84 |  |
| LC | A2a | Pipecolate | 98.7 | 0.33 | 0.91 | < 0.05* | 0.19 |  |
| LC | A2a | Glycochenodeoxycholic acid | 99.9 | 0.35 | 1.09 | 0.20 | 0.58 |  |
| LC | A2a | Pipecolate | 95.5 | 0.38 | 1.09 | 0.29 | 0.58 |  |
| LC | A2a | 5-oxo-D-Proline | 98.9 | 0.24 | 1.02 | 0.89 | 0.94 |  |
| LC | A2a | Cortisone | 99.9 | 0.20 | 0.86 | 0.42 | 0.67 |  |
| LC | A2a | Pipecolate | 97.2 | 0.38 | 1.15 | 0.22 | 0.58 |  |
| LC | A2a | Cortisol | 99.9 | 0.28 | 0.99 | 0.94 | 0.94 |  |
| LC | A2a | Cortisol | 99.9 | 0.20 | 0.98 | 0.87 | 0.94 |  |
| GC | A2b | 4-Thiazolidinecarboxylic acid | 38.1 | NA | 1.06 | 0.62 | 0.62 |  |
| LC | A3a | Tyrosine | 100.0 | 0.20 | 1.06 | 0.65 | 0.93 |  |
| LC | A3a | LysoPE (22:6) | 100.0 | 0.49 | 0.92 | 0.58 | 0.93 |  |
| LC | A3a | L-Tryptophan | 100.0 | 0.13 | 0.99 | 0.95 | 0.99 |  |
| LC | A3a | L-Proline | 100.0 | 0.56 | 1.05 | 0.75 | 0.96 | 0.86 |
| LC | A3a | Alanine/Sarcosine | 100.0 | 0.27 | 1.26 | 0.29 | 0.82 |  |
| LC | A3a | Creatine | 100.0 | 0.76 | 1.16 | 0.10 | 0.70 | 0.36 |
| LC | A3a | L-Carnitine | 100.0 | 0.40 | 0.89 | 0.63 | 0.93 |  |
| LC | A3a | ACar 5:0 | 100.0 | 0.52 | 0.97 | 0.81 | 0.97 | 0.87 |
| LC | A3a | L-Histidine | 100.0 | 0.35 | 0.83 | 0.30 | 0.82 |  |
| LC | A3a | 4-Aminobutanoic acid/2-Amino-2-methylpropanoic acid/3-Aminobutanoic acid | 100.0 | 0.62 | 0.90 | 0.57 | 0.93 | 0.71 |
| LC | A3a | Betaine | 100.0 | 0.52 | 0.77 | 0.21 | 0.78 | 0.56 |
| LC | A3a | LysoPC(22:5) | 100.0 | 0.42 | 1.01 | 0.96 | 0.99 |  |
| LC | A3a | LPC 0:0/22:6 | 100.0 | 0.51 | 0.91 | 0.49 | 0.93 | 0.70 |
| LC | A3a | Trimethylamine N-oxide | 100.0 | 0.18 | 1.04 | 0.56 | 0.93 |  |
| LC | A3a | L-Valine | 100.0 | 0.26 | 0.73 | 0.18 | 0.75 |  |
| LC | A3a | Hypoxanthine | 100.0 | 0.17 | 1.04 | 0.58 | 0.93 |  |
| LC | A3a | L-Glutamine | 100.0 | 0.14 | 1.41 | 0.13 | 0.70 |  |
| LC | A3a | LysoPE (18:2) | 100.0 | 0.31 | 1.35 | < 0.05* | 0.43 |  |
| LC | A3a | LYSOPC (18:2) | 100.0 | 0.33 | 1.01 | 0.96 | 0.99 |  |
| LC | A3a | LYSOPC (16:3) | 100.0 | 0.42 | 1.06 | 0.64 | 0.93 |  |
| LC | A3a | LYSOPE (18:1) | 100.0 | 0.26 | 1.37 | < 0.05* | 0.36 |  |
| LC | A3a | LysoPE (20:4) | 100.0 | 0.43 | 1.11 | 0.56 | 0.93 |  |
| LC | A3a | PC(16:0_20:4) | 99.0 | 0.48 | 0.88 | 0.53 | 0.93 |  |
| LC | A3a | 7a-hydroxy-3-oxo-4-cholestenoic acid (7-Hoca) | 99.9 | 0.47 | 1.08 | 0.66 | 0.94 |  |
| LC | A3a | SM d34:2 | 94.7 | 0.09 | 0.80 | 0.21 | 0.78 |  |
| LC | A3a | PC (32:2) | 96.9 | 0.37 | 1.03 | 0.78 | 0.96 |  |
| LC | A3a | ACar 10:1 | 99.9 | 0.50 | 0.82 | < 0.05* | 0.43 | 0.19 |
| LC | A3a | LYSOPC (20:4) | 97.4 | 0.45 | 1.00 | 0.98 | 1.00 |  |
| LC | A3a | Deoxycarnitine-4-butyric acid betaine | 99.9 | 0.63 | 0.85 | 0.41 | 0.93 | 0.69 |
| LC | A3a | LYSOPC (16:3) | 91.9 | 0.34 | 1.06 | 0.60 | 0.93 |  |
| LC | A3a | PC (36:4) | 47.8 | 0.06 | 0.98 | 0.55 | 0.93 |  |
| LC | A3a | LysoPC(20:2) | 99.3 | 0.28 | 1.10 | 0.53 | 0.93 |  |
| LC | A3a | PC (36:2) | 94.2 | 0.23 | 1.06 | 0.64 | 0.93 |  |
| LC | A3a | N-alpha-acetyl-L-lysine | 98.7 | 0.24 | 1.25 | 0.25 | 0.81 |  |
| LC | A3a | LYSOPC (16:1) | 99.9 | 0.54 | 1.31 | 0.07 | 0.68 | 0.31 |
| LC | A3a | ACar 11:1 | 99.7 | 0.40 | 0.85 | 0.17 | 0.74 |  |
| LC | A3a | Serotonin | 98.2 | 0.26 | 1.00 | 0.99 | 1.00 |  |
| LC | A3a | Cholesterol | 97.0 | 0.21 | 0.66 | 0.09 | 0.70 |  |
| LC | A3a | LysoPC(P-16:0) | 99.8 | 0.41 | 0.90 | 0.52 | 0.93 |  |
| LC | A3a | Docosahexaenoic acid | 99.9 | 0.47 | 0.79 | < 0.05* | 0.36 |  |
| LC | A3a | Glycolate | 96.6 | 0.03 | 1.09 | 0.49 | 0.93 |  |
| LC | A3a | Lauroylcarnitine | 99.9 | 0.42 | 0.90 | 0.30 | 0.82 |  |
| LC | A3a | LYSOPC (18:2) | 99.7 | 0.13 | 1.00 | 0.96 | 0.99 |  |
| LC | A3a | Trigonelline | 99.3 | 0.60 | 1.04 | 0.53 | 0.93 | 0.70 |
| LC | A3a | L-gamma-Glutamyl-L-leucine | 95.9 | 0.16 | 1.03 | 0.82 | 0.97 |  |
| LC | A3a | PC (36:3) | 87.0 | 0.13 | 0.89 | 0.29 | 0.82 |  |
| LC | A3a | LysoPC (18:3) | 98.2 | 0.39 | 1.16 | 0.18 | 0.74 |  |
| LC | A3a | ACar 4:0 | 81.3 | 0.69 | 0.95 | 0.45 | 0.93 | 0.70 |
| LC | A3a | PC (36:4) | 93.7 | 0.03 | 1.02 | 0.70 | 0.95 |  |
| LC | A3a | Plasmalogen PC(38:4) or PC(O-38:5) | 93.4 | 0.18 | 0.93 | 0.58 | 0.93 |  |
| LC | A3a | Taurine | 95.5 | 0.11 | 0.98 | 0.85 | 0.99 |  |
| LC | A3a | PC (38:5) | 99.2 | 0.36 | 1.09 | 0.63 | 0.93 |  |
| LC | A3a | LysoPE (18:2) | 72.0 | 0.27 | 1.05 | 0.52 | 0.93 |  |
| LC | A3a | 4-Guanidinobutanoic acid | 98.1 | 0.52 | 1.26 | < 0.05* | 0.36 | 0.12 |
| LC | A3a | ACar 10:1 | 99.1 | 0.50 | 0.83 | 0.06 | 0.62 |  |
| LC | A3a | LYSOPC (16:1) | 52.9 | 0.37 | 1.22 | < 0.05* | 0.43 |  |
| LC | A3a | ACar 9:0 | 99.9 | 0.59 | 1.03 | 0.78 | 0.96 | 0.86 |
| LC | A3a | Plasmalogen PC(36:4) or PC(O-36:5) | 97.2 | 0.38 | 1.01 | 0.94 | 0.99 |  |
| LC | A3a | Uric acid | 98.9 | 0.42 | 0.95 | 0.74 | 0.96 |  |
| LC | A3a | LYSOPC (18:0) | 95.3 | 0.14 | 1.15 | 0.38 | 0.93 |  |
| LC | A3a | LYSOPC (16:1) | 99.9 | 0.19 | 0.92 | 0.14 | 0.70 |  |
| LC | A3a | Kynurenine | 94.2 | 0.38 | 0.97 | 0.69 | 0.95 |  |
| LC | A3a | 5-Valerolactone | 69.0 | 0.24 | 1.07 | 0.46 | 0.93 |  |
| LC | A3a | ACar 4:0 | 94.1 | 0.46 | 1.04 | 0.59 | 0.93 |  |
| LC | A3a | Acetylcarnitine (C2) | 99.1 | 0.43 | 0.99 | 0.97 | 1.00 |  |
| LC | A3a | LysoPC (16:0) | 99.6 | 0.26 | 0.52 | 0.17 | 0.74 |  |
| LC | A3a | ACar 8:0 | 99.9 | 0.51 | 0.91 | 0.32 | 0.82 | 0.56 |
| LC | A3a | Linoleic acid | 94.2 | 0.02 | 0.89 | 0.10 | 0.70 |  |
| LC | A3a | LYSOPC (16:1) | 61.1 | 0.52 | 1.09 | < 0.05* | 0.43 | 0.19 |
| LC | A3a | 5,6-Dihydrouracil | 99.2 | 0.34 | 2.06 | < 0.05* | 0.36 |  |
| LC | A3a | Decanoyl-L-Carnitine (C10:0) | 99.9 | 0.45 | 0.91 | 0.26 | 0.81 |  |
| LC | A3a | L-Serine | 99.5 | 0.27 | 0.95 | 0.75 | 0.96 |  |
| LC | A3a | 1-methylnicotinamide | 99.9 | 0.41 | 1.02 | 0.80 | 0.96 |  |
| LC | A3a | LYSOPC (14:0) | 99.8 | 0.43 | 1.12 | 0.43 | 0.93 |  |
| LC | A3a | ACar 8:1 | 99.6 | 0.45 | 0.83 | < 0.05* | 0.43 |  |
| LC | A3a | LYSOPC (20:4) | 99.9 | 0.42 | 1.14 | 0.59 | 0.93 |  |
| LC | A3a | LYSOPC (18:0) | 99.5 | 0.27 | 0.93 | 0.67 | 0.94 |  |
| LC | A3a | L-Methionine | 98.8 | 0.18 | 0.91 | 0.51 | 0.93 |  |
| LC | A3a | Propionyl-L-carnitine (C3:0) | 99.9 | 0.43 | 1.12 | 0.41 | 0.93 |  |
| LC | A3a | PC (34:3) | 79.2 | 0.07 | 0.97 | 0.64 | 0.93 |  |
| LC | A3a | L-Tyrosine | 98.7 | 0.24 | 0.96 | 0.79 | 0.96 |  |
| LC | A3a | PC 38:6 | 97.4 | 0.55 | 0.90 | 0.47 | 0.93 | 0.70 |
| LC | A3a | ACar 18:1 | 99.3 | 0.38 | 1.00 | 1.00 | 1.00 |  |
| LC | A3a | LYSOPC (17:0) | 99.9 | 0.42 | 0.99 | 0.94 | 0.99 |  |
| LC | A3a | Theophylline | 98.8 | 0.53 | 0.88 | < 0.05* | 0.43 | 0.19 |
| LC | A3a | Glycocholic acid | 93.6 | 0.40 | 1.03 | 0.71 | 0.96 |  |
| LC | A3a | PC (34:1) | 96.3 | 0.19 | 1.15 | 0.16 | 0.74 |  |
| LC | A3a | PC (30:0) | 97.4 | 0.33 | 1.08 | 0.46 | 0.93 |  |
| LC | A3a | PC (38:3) | 86.1 | 0.07 | 0.99 | 0.91 | 0.99 |  |
| LC | A3a | 3,5-Tetradecadiencarnitine (C14:2) | 99.9 | 0.45 | 0.86 | 0.10 | 0.70 |  |
| LC | A3a | Indolelactic acid | 99.9 | 0.41 | 1.30 | 0.11 | 0.70 |  |
| LC | A3a | Dehydroepiandrosterone | 54.8 | 0.98 | 1.01 | 0.93 | 0.99 | 0.93 |
| LC | A3a | Paraxanthine | 99.6 | 0.50 | 0.94 | 0.22 | 0.79 |  |
| LC | A3a | LYSOPC (20:3) | 97.7 | 0.21 | 0.90 | 0.47 | 0.93 |  |
| LC | A3a | Uracil | 98.8 | 0.30 | 1.00 | 0.99 | 1.00 |  |
| LC | A3a | Threonic acid | 98.9 | 0.17 | 1.07 | 0.32 | 0.82 |  |
| LC | A3a | LYSOPC (20:3) | 99.9 | 0.42 | 1.19 | 0.30 | 0.82 |  |
| LC | A3a | Retinol | 99.0 | 0.45 | 1.29 | 0.13 | 0.70 |  |
| LC | A3a | LYSOPC (20:3) | 47.1 | 0.08 | 0.97 | 0.68 | 0.94 |  |
| LC | A3a | PC (34:2) | 94.9 | 0.15 | 1.11 | 0.64 | 0.93 |  |
| LC | A3a | LYSOPC (18:4) | 99.9 | 0.51 | 1.34 | < 0.05* | 0.43 | 0.19 |
| LC | A3a | PC (36:4) | 97.1 | 0.29 | 1.07 | 0.56 | 0.93 |  |
| LC | A3a | Nepsilon, nepsilon, nepsilon-trimethyllysine | 99.9 | 0.40 | 1.16 | 0.26 | 0.81 |  |
| LC | A3a | PC(32:1) | 96.7 | 0.49 | 1.26 | < 0.05* | 0.36 |  |
| LC | A3a | L-Threonine/L-Homoserine | 98.4 | 0.32 | 0.78 | 0.11 | 0.70 |  |
| LC | A3a | PC (40:5) | 90.6 | 0.14 | 1.02 | 0.79 | 0.96 |  |
| LC | A3a | Theobromine | 99.9 | 0.39 | 0.92 | 0.15 | 0.74 |  |
| LC | A3a | Phe-Phe | 99.9 | 0.28 | 0.95 | 0.26 | 0.81 |  |
| LC | A3a | Creatinine | 99.9 | 0.60 | 0.79 | 0.28 | 0.82 | 0.56 |
| LC | A3a | Tyrosine | 99.9 | 0.22 | 1.10 | 0.55 | 0.93 |  |
| LC | A3a | Undecanoylcarnitine | 99.8 | 0.34 | 0.96 | 0.76 | 0.96 |  |
| LC | A3a | Plasmalogen PC(36:3) or PC(O-36:4) | 93.6 | 0.16 | 0.99 | 0.96 | 0.99 |  |
| LC | A3a | Diacylglycerol (C34:2) | 79.8 | 0.21 | 1.10 | 0.12 | 0.70 |  |
| LC | A3a | Dimethylsulfone | 99.9 | 0.44 | 0.99 | 0.94 | 0.99 |  |
| LC | A3a | Indole-3-propionic acid (IPA) | 99.5 | 0.45 | 0.96 | 0.57 | 0.93 |  |
| LC | A3a | PC (36:3) | 87.3 | 0.01 | 0.98 | 0.88 | 0.99 |  |
| LC | A3a | ACar 5:0 | 93.8 | 0.29 | 1.17 | 0.21 | 0.78 |  |
| LC | A3a | Glycodeoxycholic acid | 89.5 | 0.52 | 0.93 | 0.13 | 0.70 | 0.42 |
| LC | A3a | PC (40:6) | 95.1 | 0.28 | 0.94 | 0.51 | 0.93 |  |
| LC | A3a | Tetradecanoylcarnitine (C14:0) | 99.4 | 0.30 | 1.05 | 0.76 | 0.96 |  |
| GC | A3a | 3-Hydroxy-dithiobutanoic acid | 100.0 | 0.55 | 1.25 | 0.31 | 0.82 | 0.56 |
| GC | A3a | Tetradecanoic acid | 100.0 | 0.22 | 0.85 | < 0.05* | 0.47 |  |
| GC | A3a | (9E,12E)-octadeca-9,12-dienoic acid | 100.0 | 0.70 | 0.77 | 0.14 | 0.70 | 0.42 |
| GC | A3a | Indole | 100.0 | 0.61 | 1.75 | < 0.05* | 0.36 | 0.12 |
| GC | A3a | Skatole | 100.0 | 0.57 | 0.88 | 0.32 | 0.82 | 0.56 |
| GC | A3a | (5S)-5-ethyl-5-[(1R)-1-hydroxyethyl]-2-oxolanone | 100.0 | 0.26 | 0.86 | 0.46 | 0.93 |  |
| GC | A3a | 8-Hydroxy-4Z,9-decadienoic acid | 100.0 | 0.30 | 1.09 | 0.50 | 0.93 |  |
| GC | A3a | 2-Hydroxy-4-phenyl-butyric acid ethyl ester | 100.0 | 0.51 | 0.88 | 0.53 | 0.93 | 0.70 |
| GC | A3a | 4-oxo-6,7,8,9-tetrahydro-pyrido[1,2-a]pyrimidine | 100.0 | 0.18 | 1.13 | 0.35 | 0.87 |  |
| GC | A3a | 3-[(2S)-2-pyrrolidin-1-iumyl]propanoate | 100.0 | 0.46 | 1.34 | 0.19 | 0.75 |  |
| GC | A3a | .beta.-apo-8'-carotenal | 100.0 | 0.53 | 1.01 | 0.89 | 0.99 | 0.92 |
| GC | A3a | 3-methyl-2-pentyl-1H-pyrrole | 100.0 | 0.51 | 0.87 | 0.25 | 0.81 | 0.56 |
| GC | A3a | methyl 2-(1,3-dimethyl-2-oxoimidazolidin-4-yl)acetate | 100.0 | 0.02 | 0.99 | 0.94 | 0.99 |  |
| GC | A3a | 2-phenylacetic acid | 100.0 | 0.51 | 0.81 | 0.26 | 0.81 | 0.56 |
| GC | A3a | 6-[6'-Hydroxy-1'-hexyn-1'-yl]-1H-purine | 100.0 | 0.06 | 0.96 | 0.96 | 0.99 |  |
| GC | A3a | 5,6-Dioxoheptanoic acid | 100.0 | 0.46 | 1.32 | 0.13 | 0.70 |  |
| GC | A3a | 2-amino-7-methyl-3H-purin-6-one | 100.0 | 0.74 | 0.82 | 0.67 | 0.94 | 0.81 |
| GC | A3a | Decanoic acid | 100.0 | 0.36 | 0.99 | 0.89 | 0.99 |  |
| GC | A3a | (S)-2-Ethyl-5-oxotetrahydrofuran-2-carboxylic acid | 100.0 | 0.48 | 0.98 | 0.91 | 0.99 |  |
| GC | A3a | 2-Ethylhexanoic acid | 100.0 | 0.33 | 1.06 | 0.77 | 0.96 |  |
| GC | A3a | (2S)-2-Amino-3-(1H-2-indolyl)propionic acid (L-isotrytophan) | 99.9 | 0.40 | 0.92 | 0.48 | 0.93 |  |
| GC | A3a | 12-Oxooctadec-9-enoic acid | 89.1 | 0.25 | 1.10 | 0.08 | 0.70 |  |
| GC | A3a | (E)-2-phenyl-3-(2-pyridinyl)-2-propenoic acid | 59.5 | 0.30 | 1.09 | 0.17 | 0.74 |  |
| GC | A3a | 1,5-Anhydro-D-fructose | 55.2 | 0.27 | 1.02 | 0.75 | 0.96 |  |
| GC | A3a | (E)-6-hydroxyhept-4-enoic acid | 92.3 | 0.16 | 1.03 | 0.60 | 0.93 |  |
| GC | A3a | 1,5-Anhydro-2-deoxy-2-(methoximino)-D-arabino-hexitol | 42.1 | 0.20 | 0.99 | 0.62 | 0.93 |  |
| GC | A3a | 2-(Aminomethyl)-5,5-dimethyl-1,3-thiazolidine-4-carboxylic acid | 55.5 | 0.20 | 1.01 | 0.79 | 0.96 |  |
| GC | A3a | 11,15'-Dihydrooxepin-.beta.,.beta.-carotene | 99.9 | 0.46 | 1.01 | 0.73 | 0.96 |  |
| GC | A3a | (S)-5-Methyl-2-(dimethylamino)-1H-imidazol-4(5H)-one | 51.1 | NA | 1.00 | 0.96 | 0.99 |  |
| GC | A3a | 4,5-Secocholest-8-en-3-one, 5-ethylidene-14-methyl- | 99.7 | 0.36 | 1.10 | 0.25 | 0.81 |  |
| LC | A3b | PC (36:2) | 36.6 | 0.02 | 1.01 | 0.94 | 0.95 |  |
| LC | A3b | Cotinine | 27.7 | NA | 1.69 | < 0.001** | < 0.001 |  |
| LC | A3b | LYSOPC (20:3) | 10.0 | NA | 0.79 | 0.20 | 0.28 |  |
| LC | A3b | Cotinine secondary peak | 36.3 | 0.76 | 1.34 | < 0.05** | 0.04 | 0.01 |
| LC | A3b | LYSOPC (20:4) | 36.6 | NA | 1.19 | 0.15 | 0.28 |  |
| LC | A3b | 2-Hydroxyphenylacetic acid | 24.1 | NA | 1.32 | 0.20 | 0.28 |  |
| GC | A3b | Guadiscoline | 27.6 | NA | 1.01 | 0.95 | 0.95 |  |

**Table S5:**
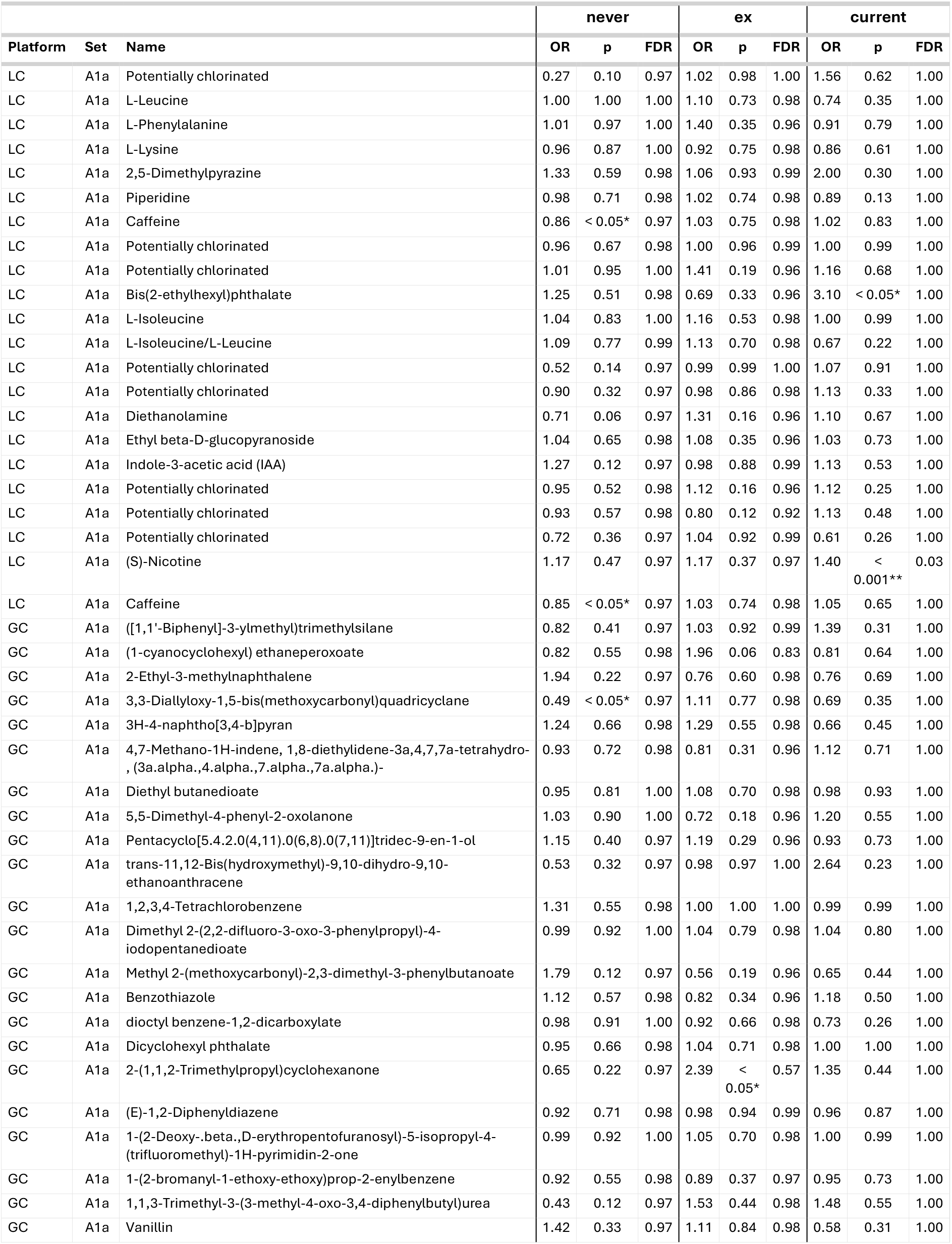
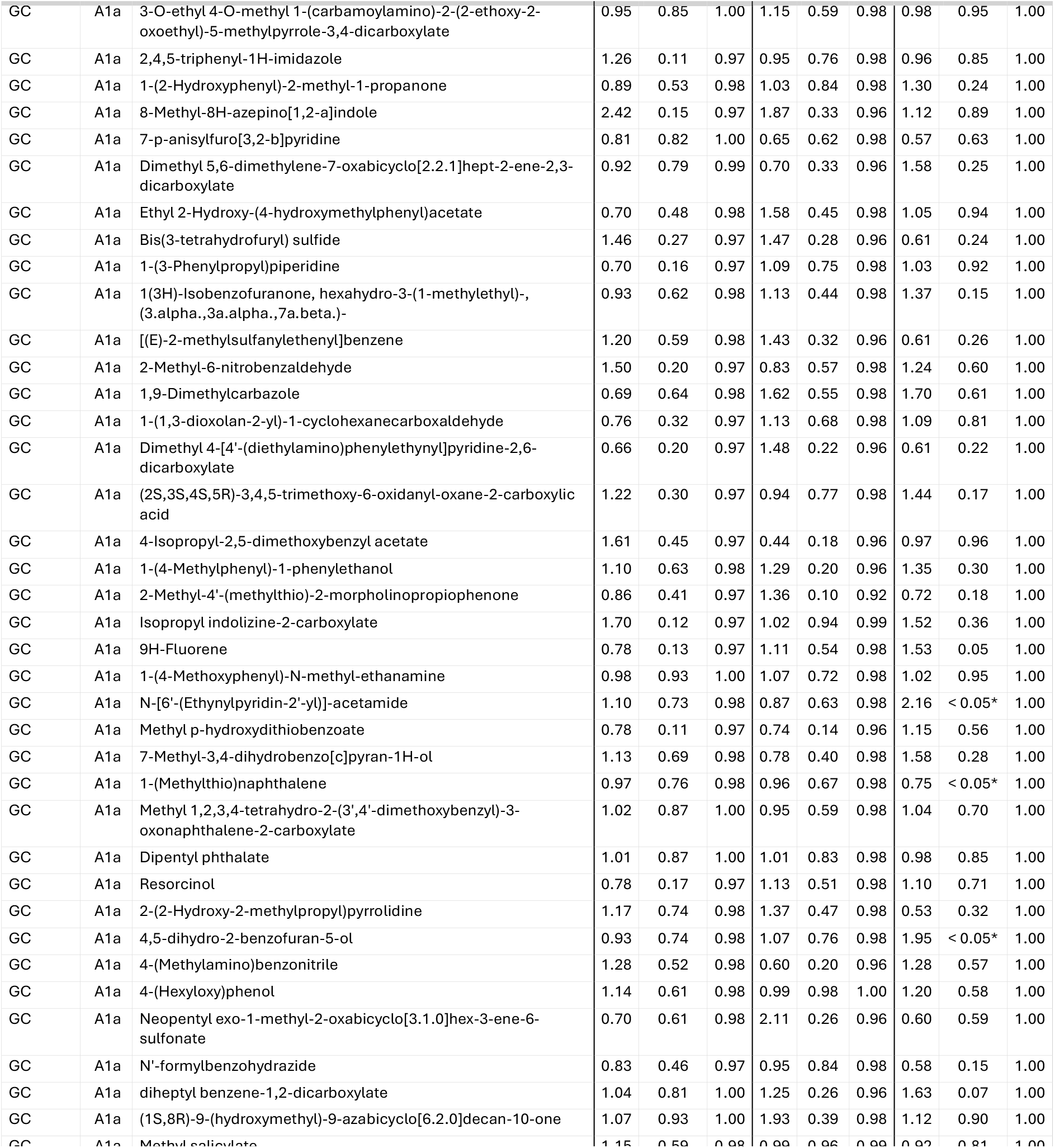
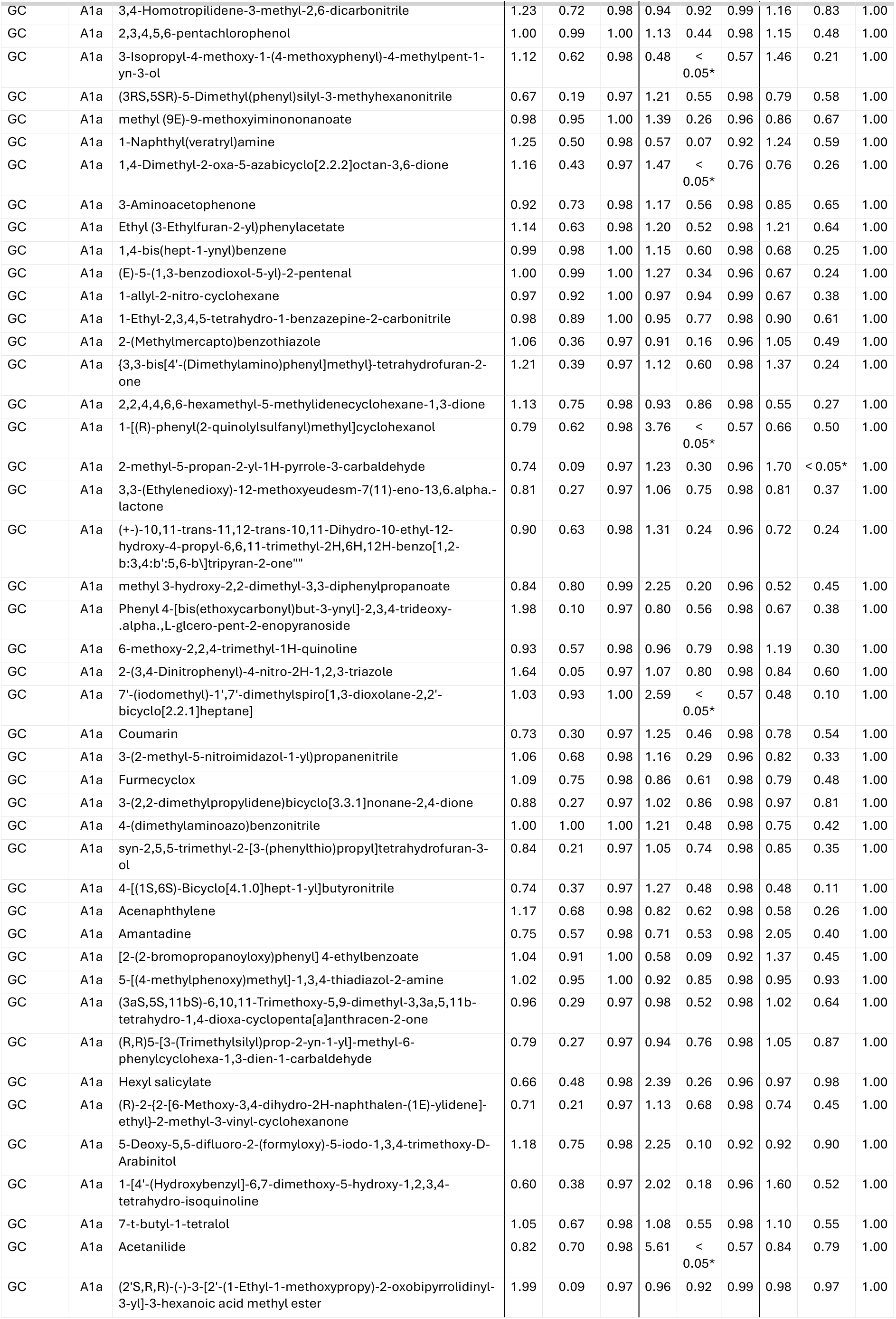
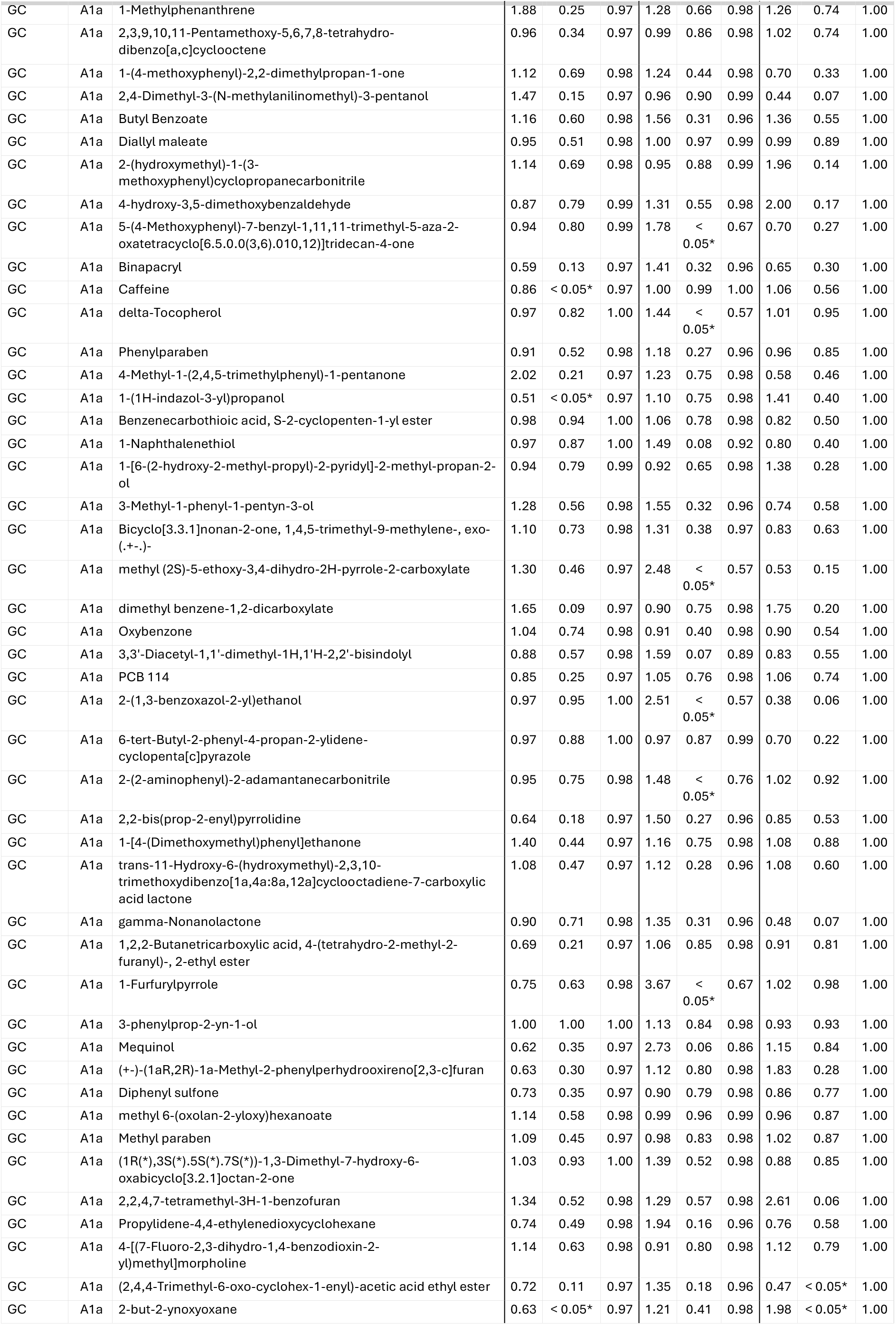
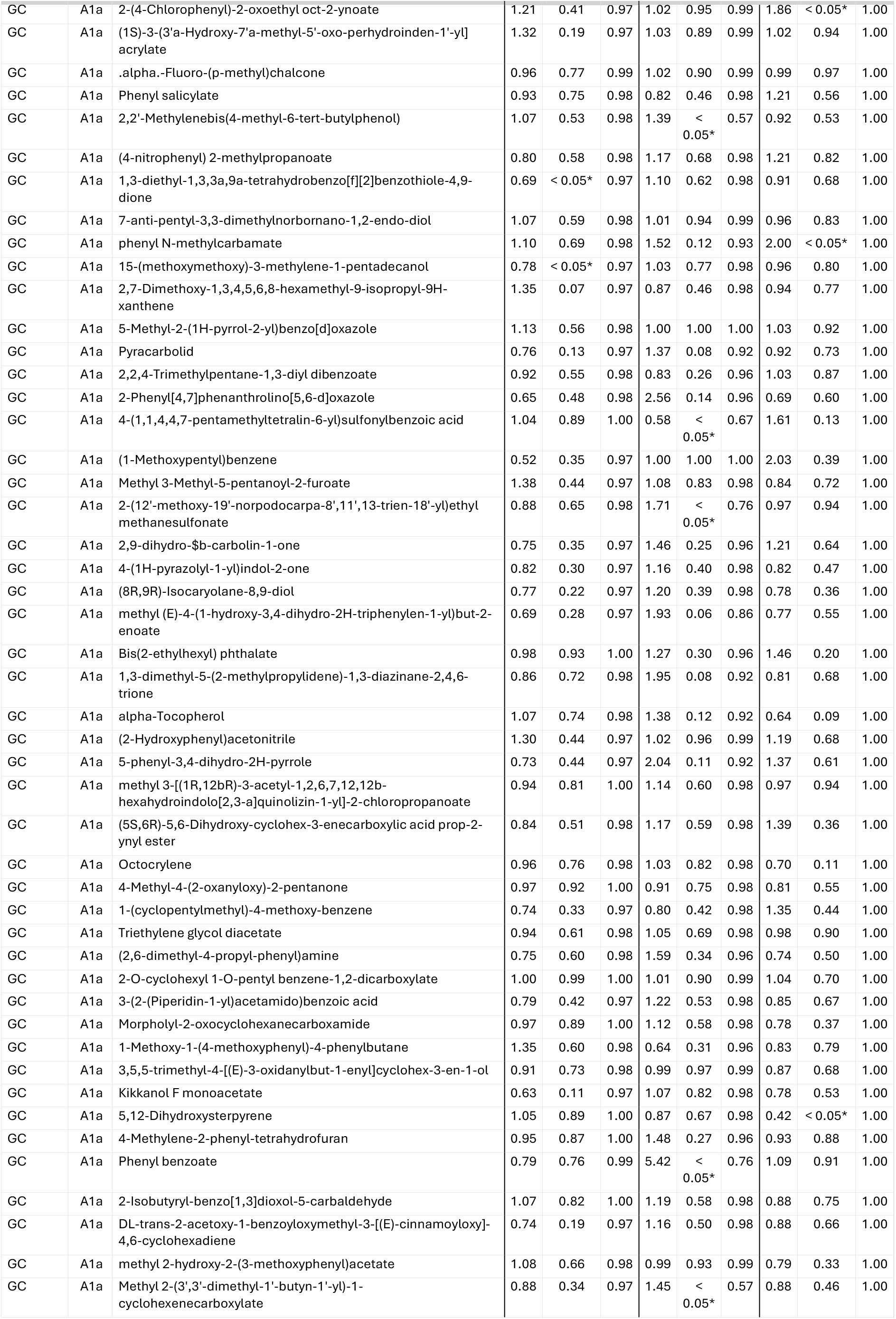
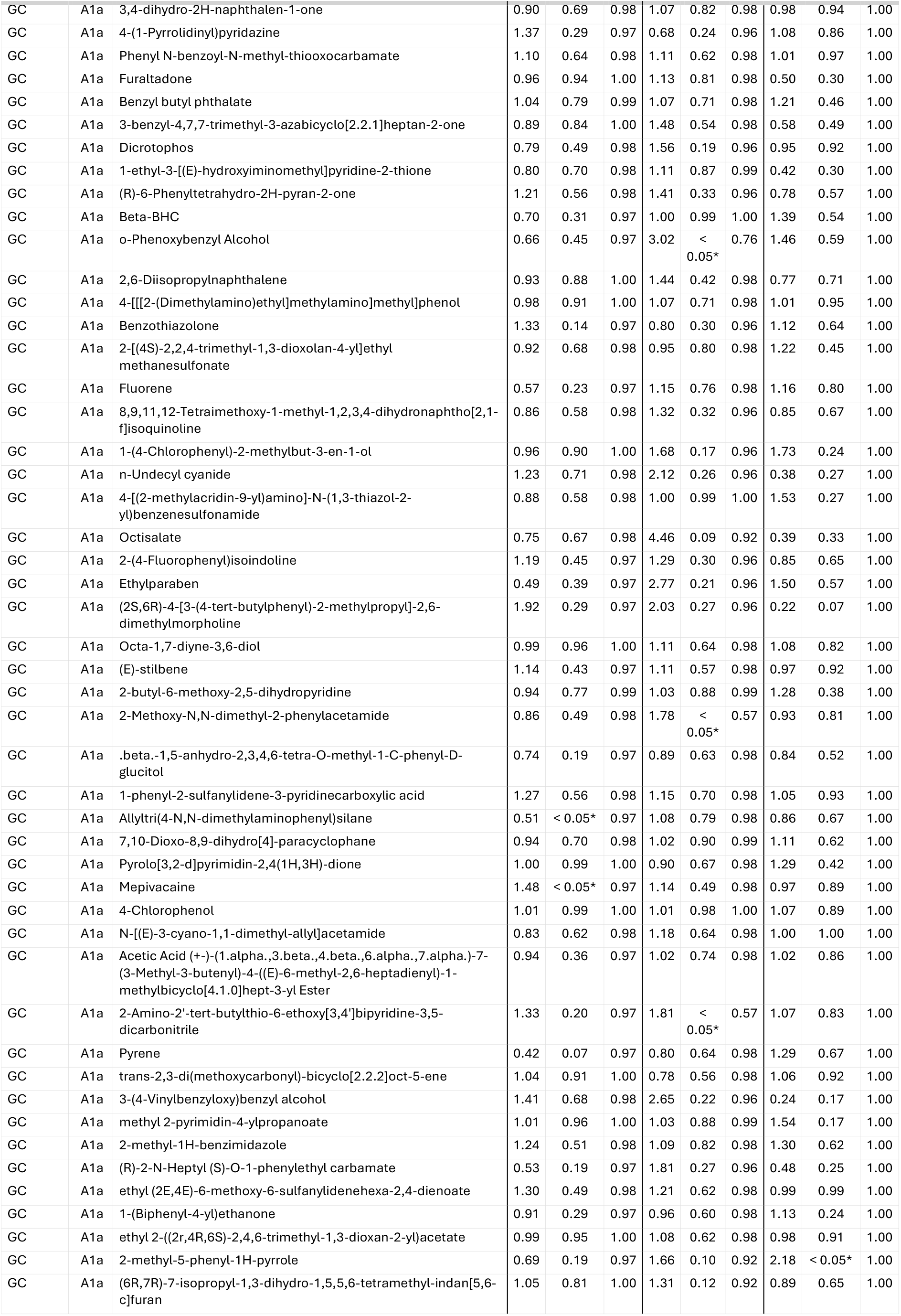
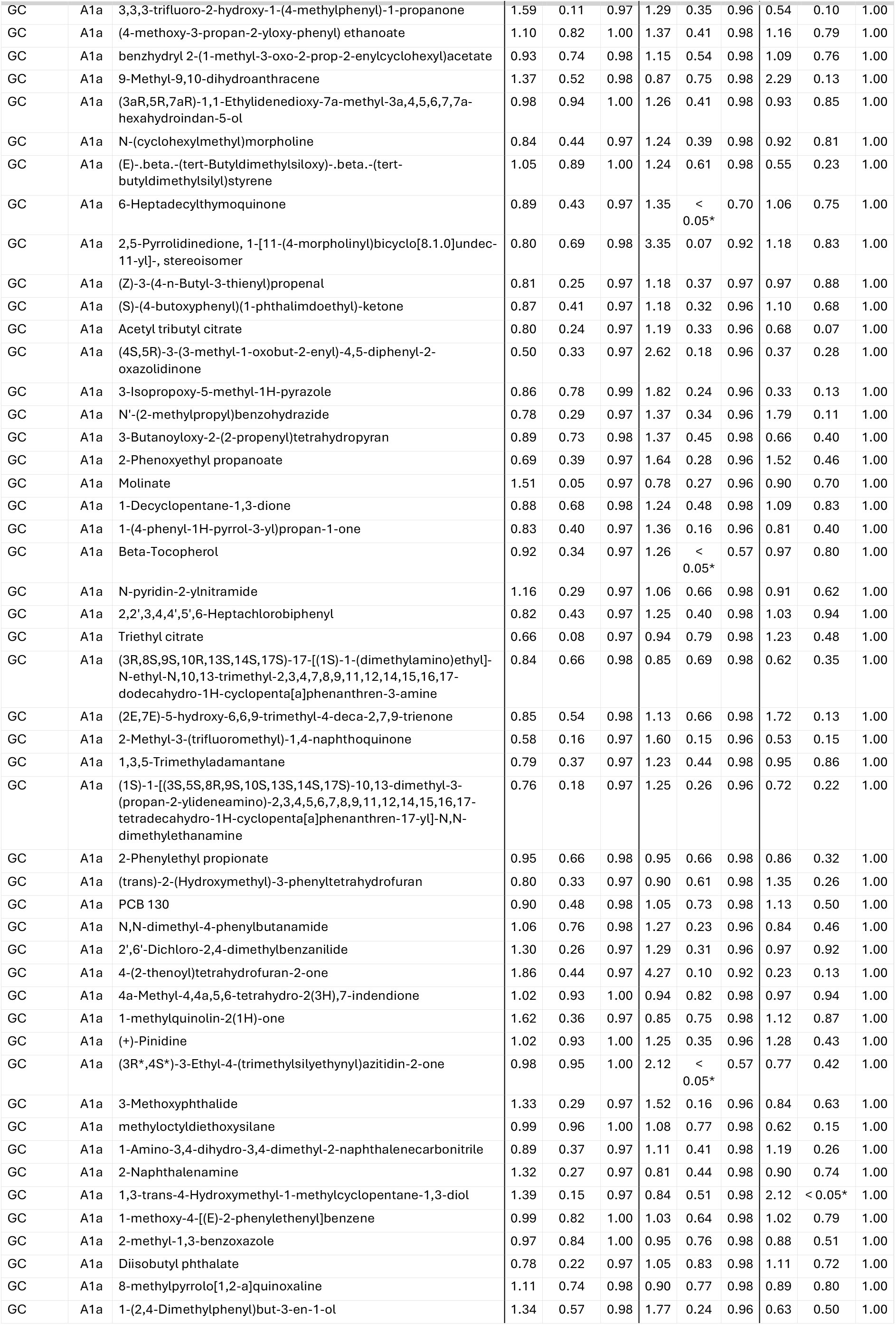
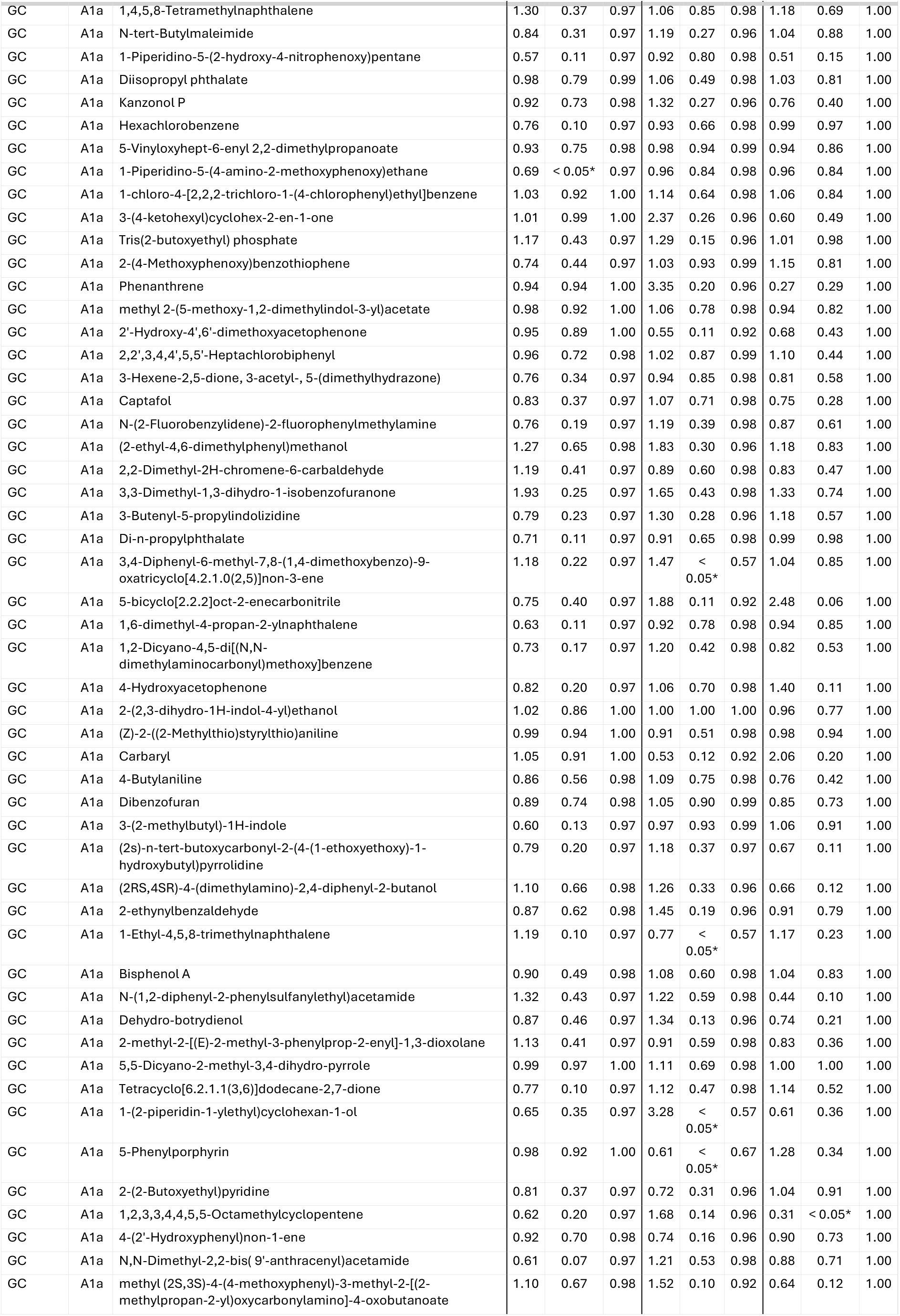
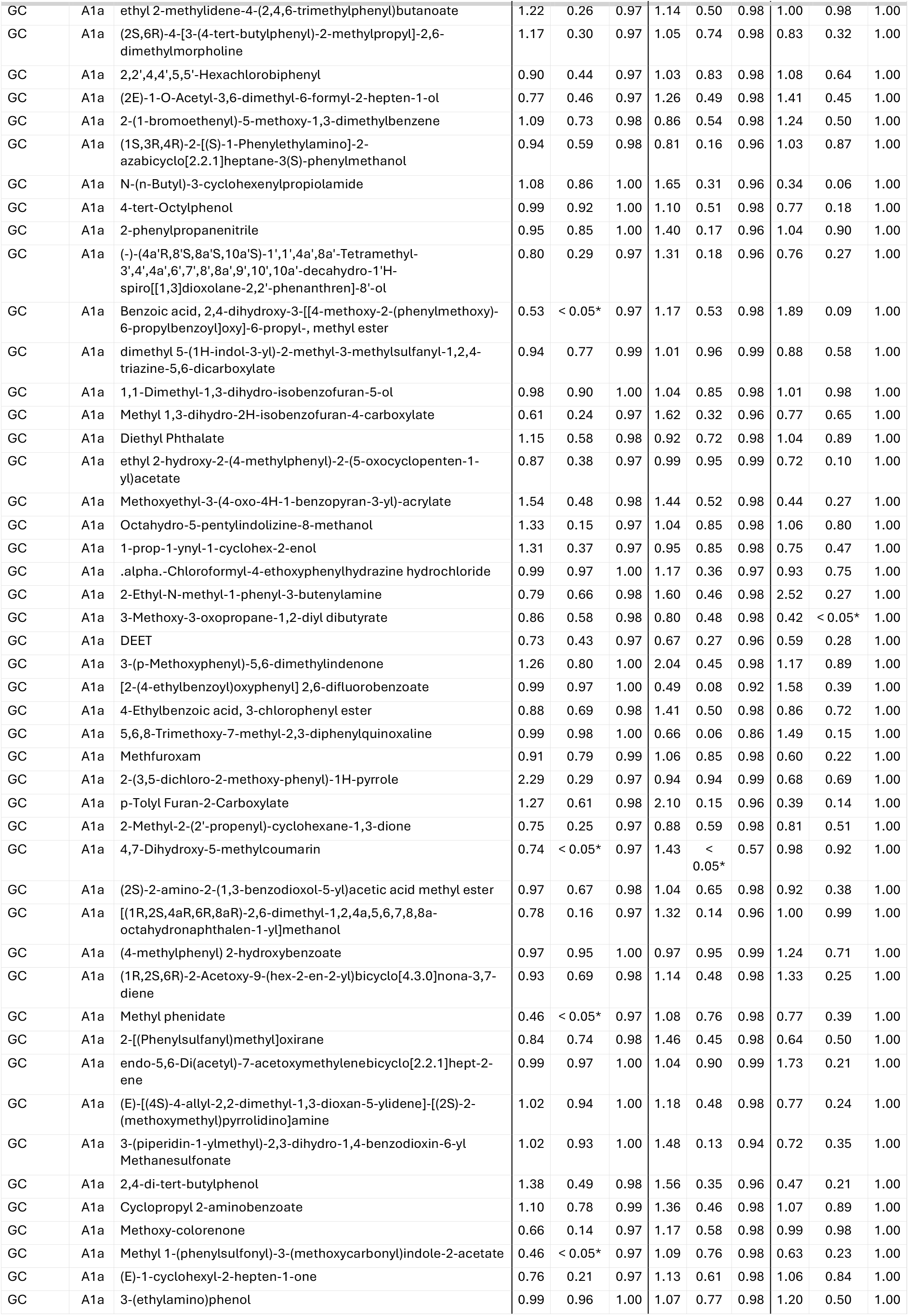
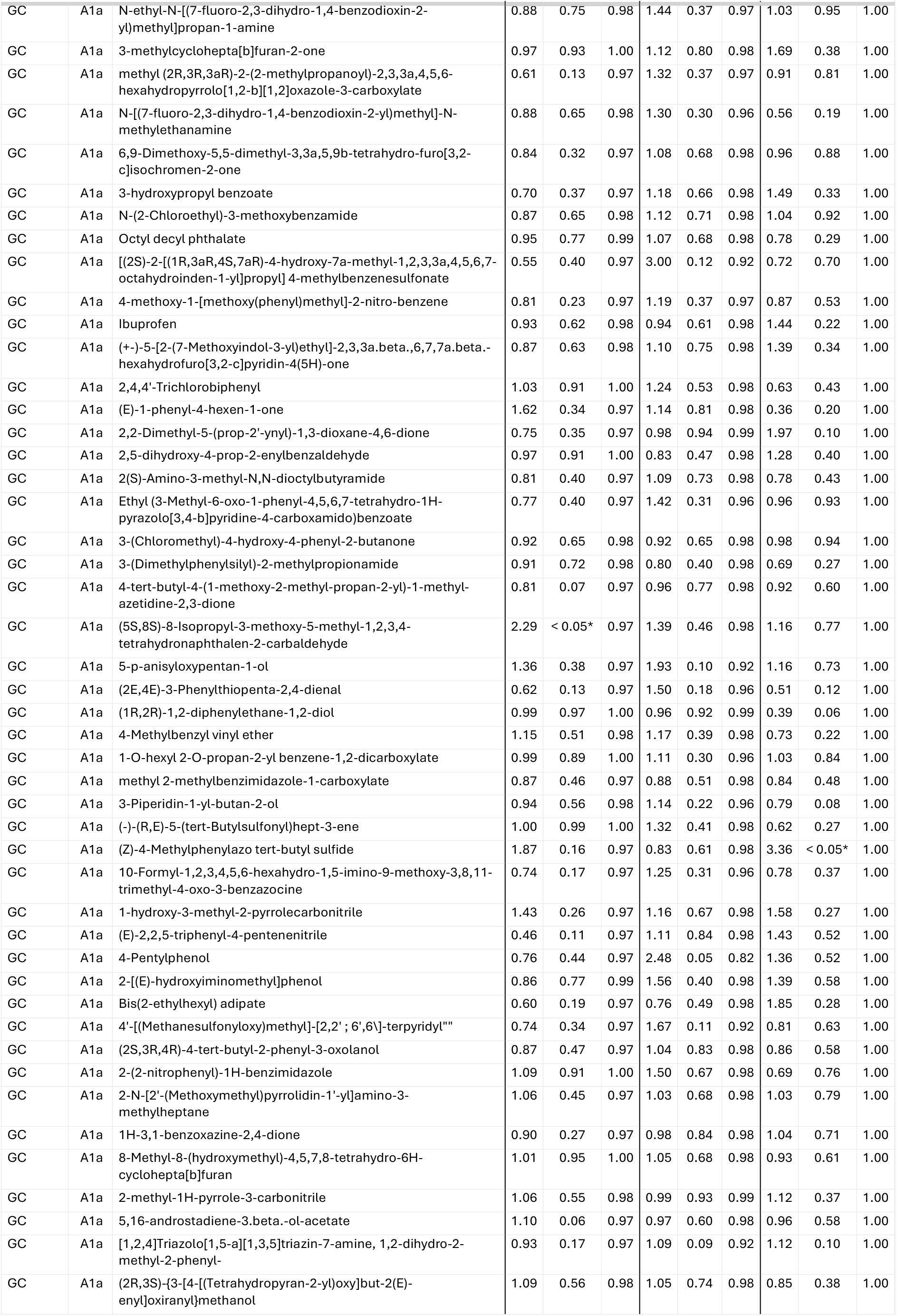
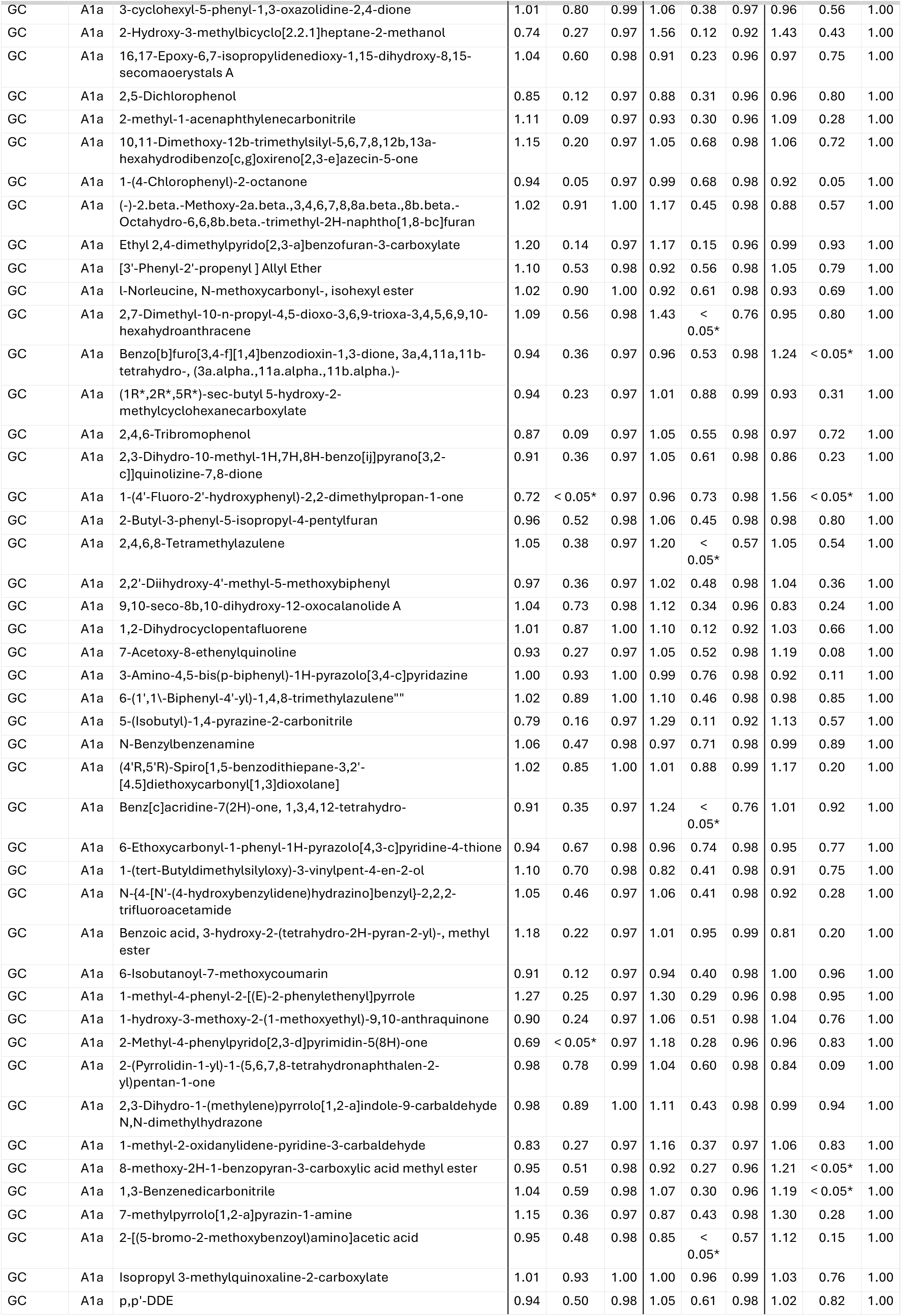
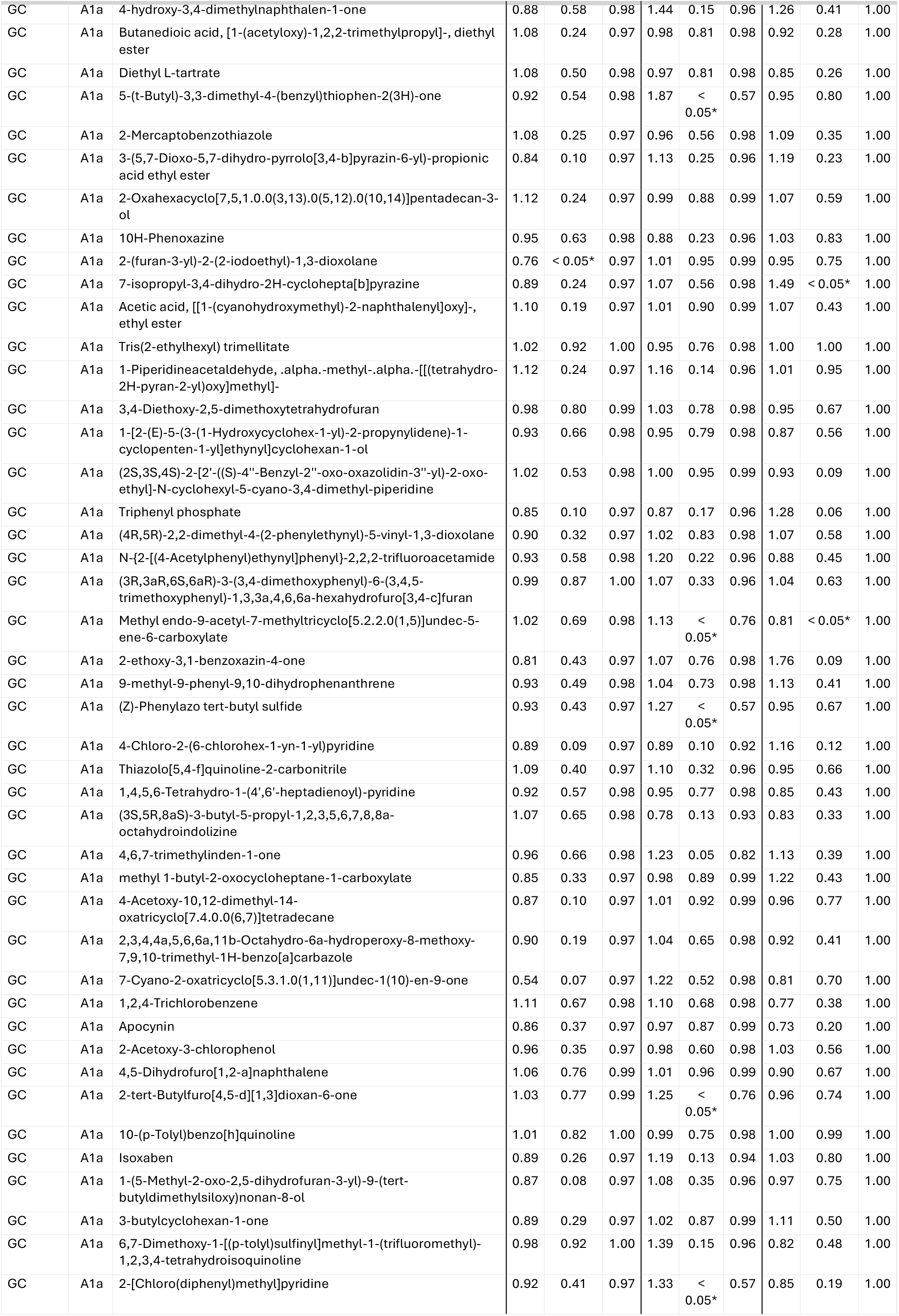
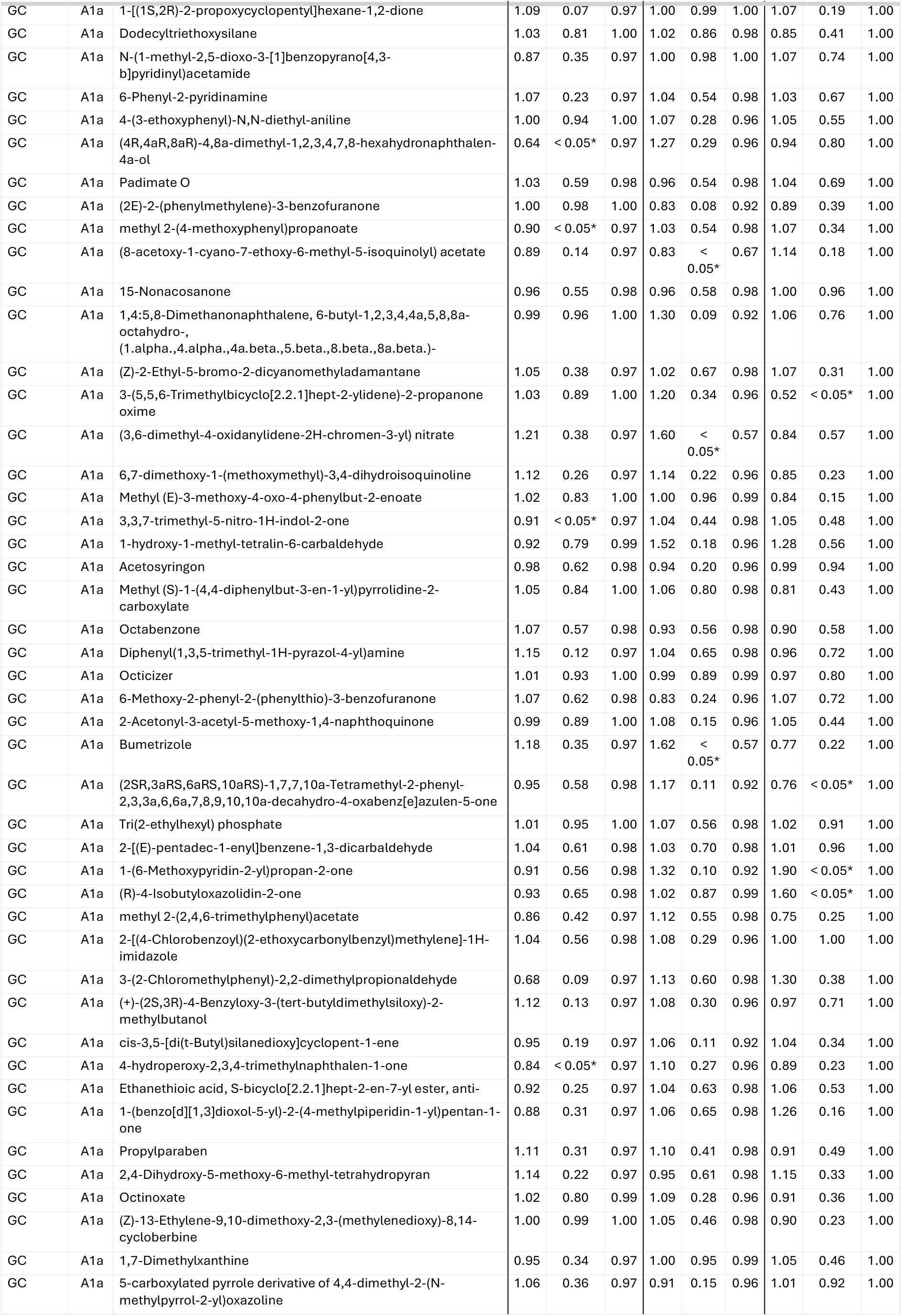
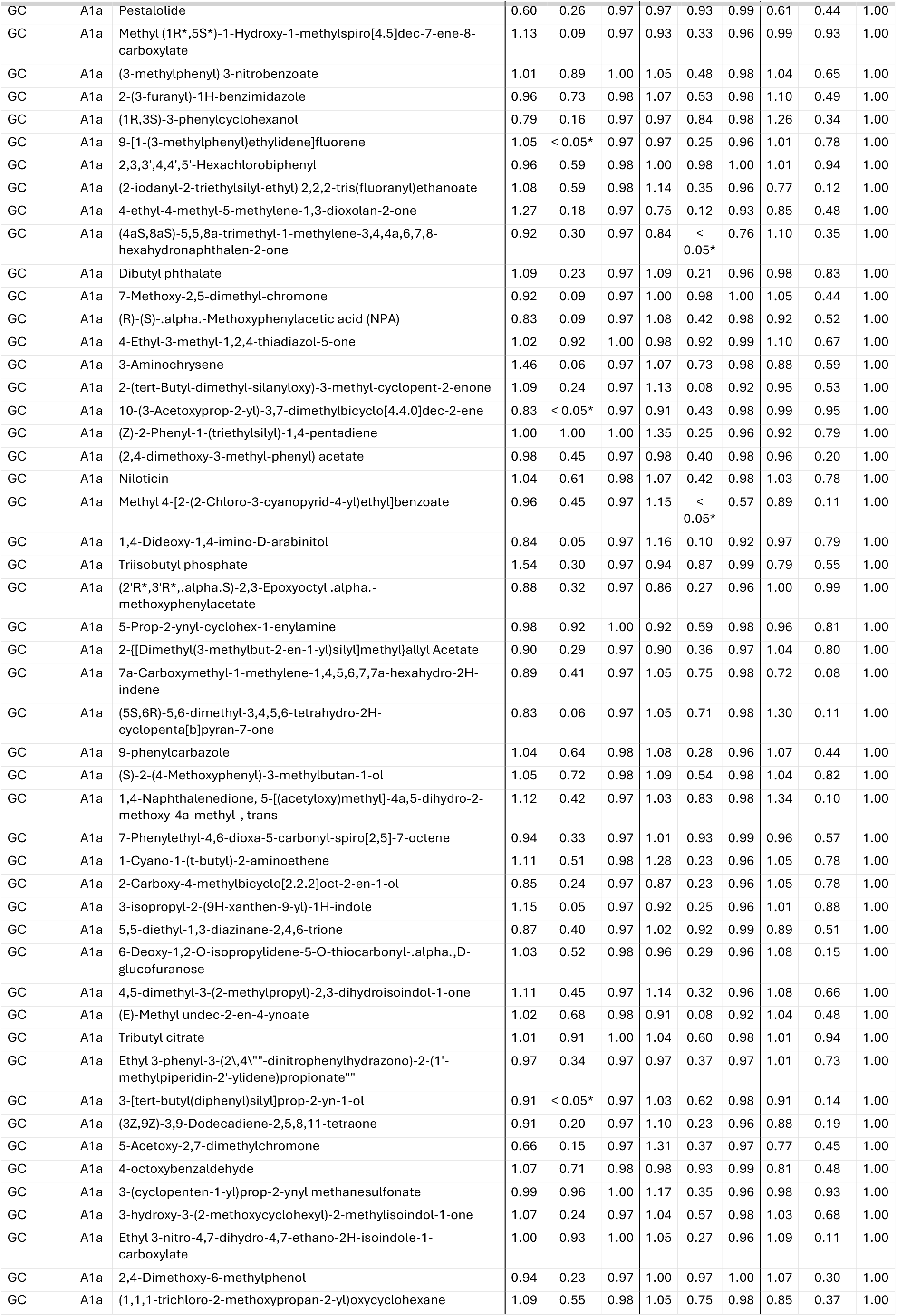
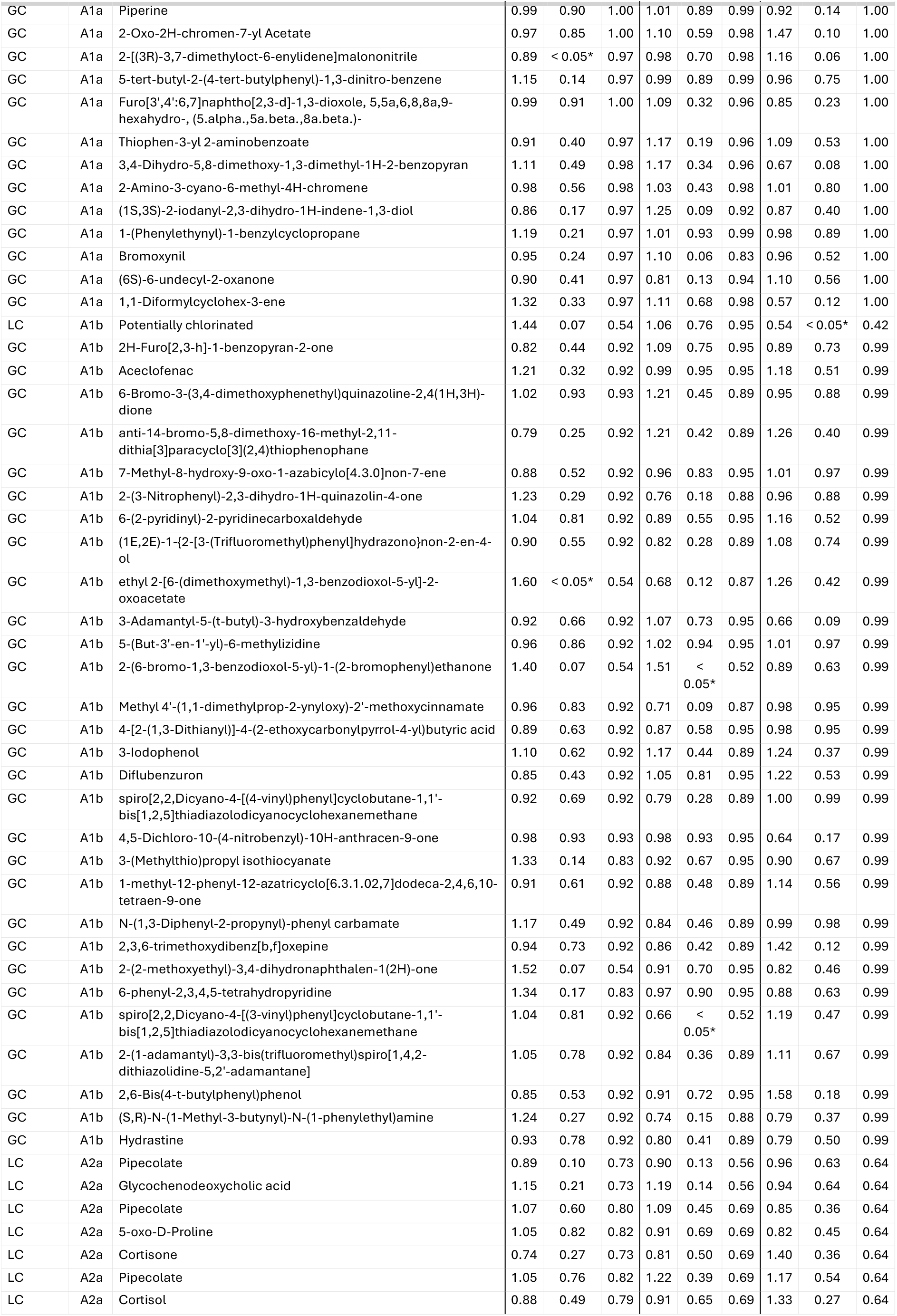
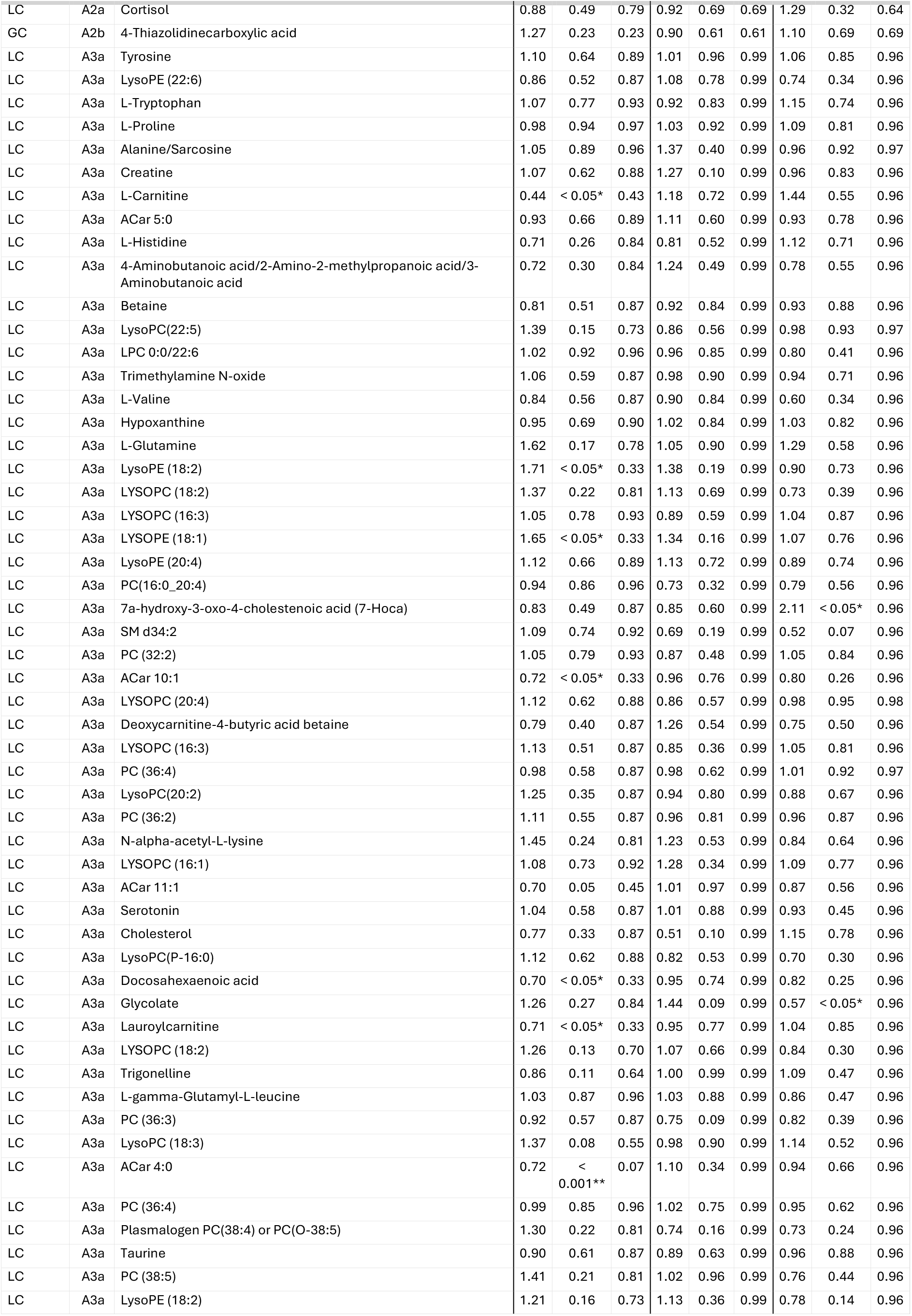
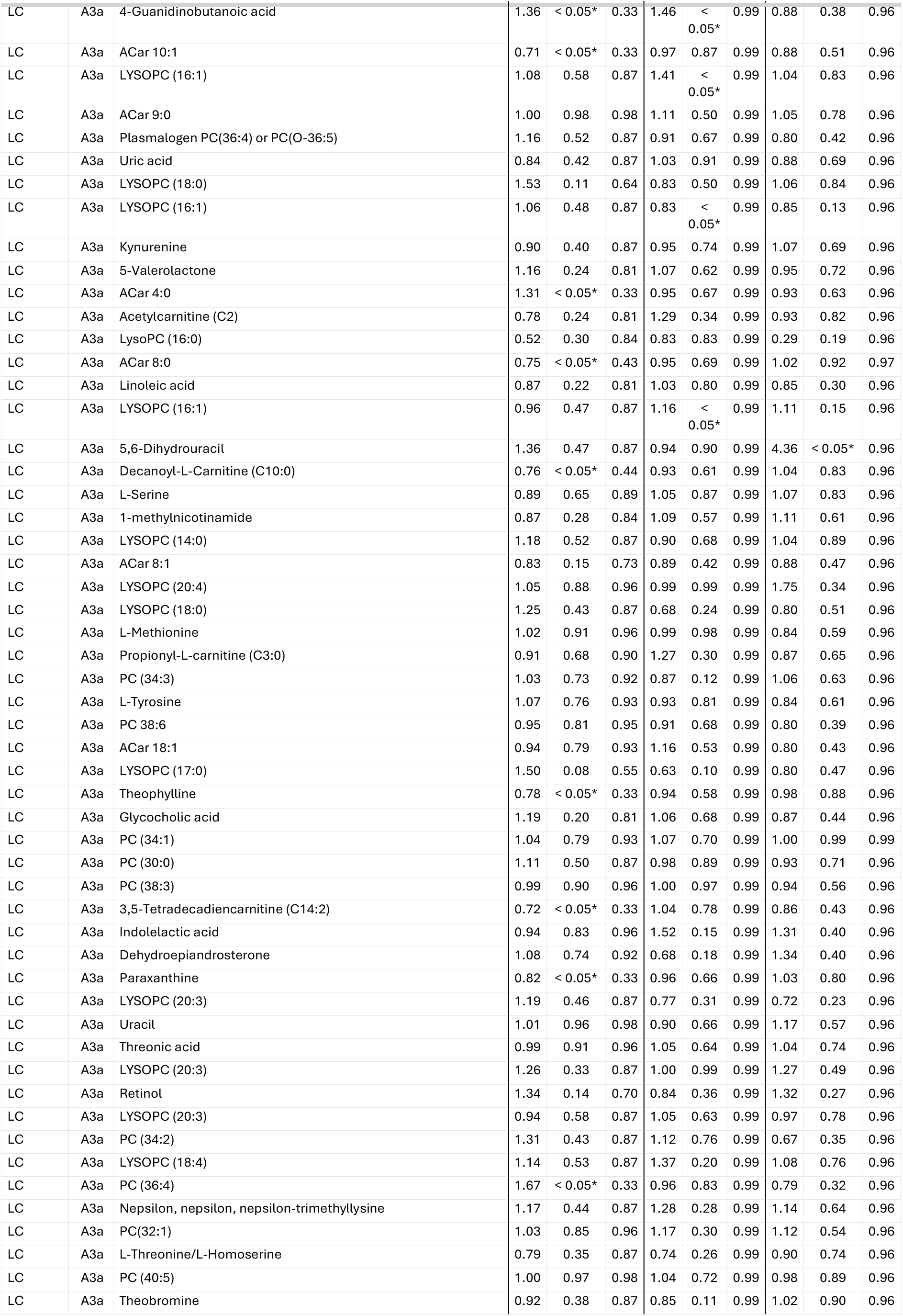
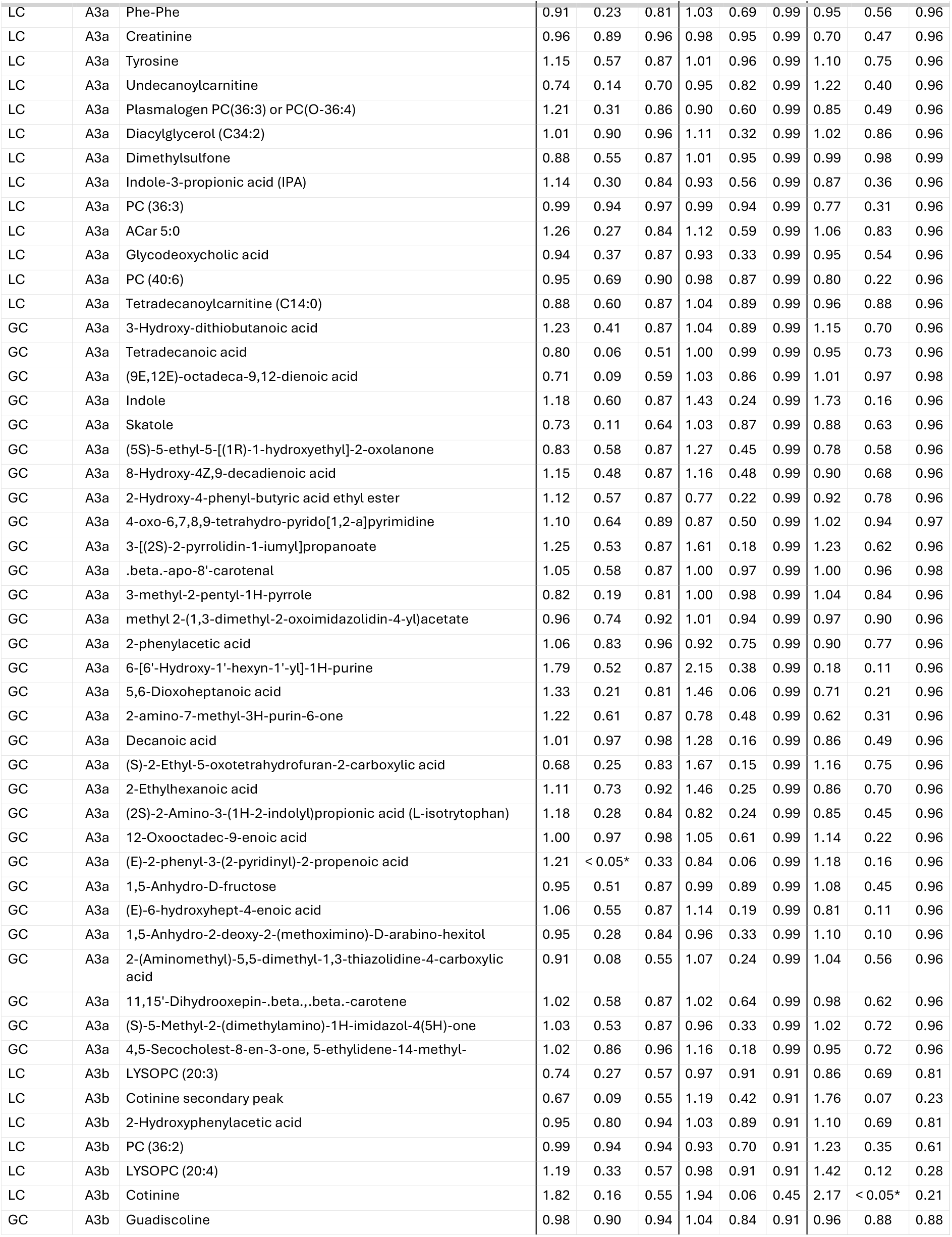
results for the initially annotated peaks in the smoker stratified analysis. Set A describes the annotated metabolites, Set B indicates the initially unidentified features/clusters. 1, 2, 3 describes the exogenous, endogenous, both endogenous/exogenously derived metabolites respectively. The last letter indicates if the feature was analyzed as continuous variable (lower case ‘a’) or as binary variable of detected, not detected (lower case ‘b’). % = proportion of samples case-control study annotation was detected; OR = odds ratio; p = p-value; FDR = false discovery rate.

|  |  |  | never |  |  | ex |  |  | current |  |  |
|---|---|---|---|---|---|---|---|---|---|---|---|
| Platform | Set | Name | OR | p | FDR | OR | p | FDR | OR | p | FDR |
| LC | A1a | Potentially chlorinated | 0.27 | 0.10 | 0.97 | 1.02 | 0.98 | 1.00 | 1.56 | 0.62 | 1.00 |
| LC | A1a | L-Leucine | 1.00 | 1.00 | 1.00 | 1.10 | 0.73 | 0.98 | 0.74 | 0.35 | 1.00 |
| LC | A1a | L-Phenylalanine | 1.01 | 0.97 | 1.00 | 1.40 | 0.35 | 0.96 | 0.91 | 0.79 | 1.00 |
| LC | A1a | L-Lysine | 0.96 | 0.87 | 1.00 | 0.92 | 0.75 | 0.98 | 0.86 | 0.61 | 1.00 |
| LC | A1a | 2,5-Dimethylpyrazine | 1.33 | 0.59 | 0.98 | 1.06 | 0.93 | 0.99 | 2.00 | 0.30 | 1.00 |
| LC | A1a | Piperidine | 0.98 | 0.71 | 0.98 | 1.02 | 0.74 | 0.98 | 0.89 | 0.13 | 1.00 |
| LC | A1a | Caffeine | 0.86 | < 0.05* | 0.97 | 1.03 | 0.75 | 0.98 | 1.02 | 0.83 | 1.00 |
| LC | A1a | Potentially chlorinated | 0.96 | 0.67 | 0.98 | 1.00 | 0.96 | 0.99 | 1.00 | 0.99 | 1.00 |
| LC | A1a | Potentially chlorinated | 1.01 | 0.95 | 1.00 | 1.41 | 0.19 | 0.96 | 1.16 | 0.68 | 1.00 |
| LC | A1a | Bis(2-ethylhexyl)phthalate | 1.25 | 0.51 | 0.98 | 0.69 | 0.33 | 0.96 | 3.10 | < 0.05* | 1.00 |
| LC | A1a | L-Isoleucine | 1.04 | 0.83 | 1.00 | 1.16 | 0.53 | 0.98 | 1.00 | 0.99 | 1.00 |
| LC | A1a | L-Isoleucine/L-Leucine | 1.09 | 0.77 | 0.99 | 1.13 | 0.70 | 0.98 | 0.67 | 0.22 | 1.00 |
| LC | A1a | Potentially chlorinated | 0.52 | 0.14 | 0.97 | 0.99 | 0.99 | 1.00 | 1.07 | 0.91 | 1.00 |
| LC | A1a | Potentially chlorinated | 0.90 | 0.32 | 0.97 | 0.98 | 0.86 | 0.98 | 1.13 | 0.33 | 1.00 |
| LC | A1a | Diethanolamine | 0.71 | 0.06 | 0.97 | 1.31 | 0.16 | 0.96 | 1.10 | 0.67 | 1.00 |
| LC | A1a | Ethyl beta-D-glucopyranoside | 1.04 | 0.65 | 0.98 | 1.08 | 0.35 | 0.96 | 1.03 | 0.73 | 1.00 |
| LC | A1a | Indole-3-acetic acid (IAA) | 1.27 | 0.12 | 0.97 | 0.98 | 0.88 | 0.99 | 1.13 | 0.53 | 1.00 |
| LC | A1a | Potentially chlorinated | 0.95 | 0.52 | 0.98 | 1.12 | 0.16 | 0.96 | 1.12 | 0.25 | 1.00 |
| LC | A1a | Potentially chlorinated | 0.93 | 0.57 | 0.98 | 0.80 | 0.12 | 0.92 | 1.13 | 0.48 | 1.00 |
| LC | A1a | Potentially chlorinated | 0.72 | 0.36 | 0.97 | 1.04 | 0.92 | 0.99 | 0.61 | 0.26 | 1.00 |
| LC | A1a | (S)-Nicotine | 1.17 | 0.47 | 0.97 | 1.17 | 0.37 | 0.97 | 1.40 | < 0.001** | 0.03 |
| LC | A1a | Caffeine | 0.85 | < 0.05* | 0.97 | 1.03 | 0.74 | 0.98 | 1.05 | 0.65 | 1.00 |
| GC | A1a | ([1,1'-Biphenyl]-3-ylmethyl)trimethylsilane | 0.82 | 0.41 | 0.97 | 1.03 | 0.92 | 0.99 | 1.39 | 0.31 | 1.00 |
| GC | A1a | (1-cyanocyclohexyl) ethaneperoxoate | 0.82 | 0.55 | 0.98 | 1.96 | 0.06 | 0.83 | 0.81 | 0.64 | 1.00 |
| GC | A1a | 2-Ethyl-3-methylnaphthalene | 1.94 | 0.22 | 0.97 | 0.76 | 0.60 | 0.98 | 0.76 | 0.69 | 1.00 |
| GC | A1a | 3,3-Diallyloxy-1,5-bis(methoxycarbonyl)quadricyclane | 0.49 | < 0.05* | 0.97 | 1.11 | 0.77 | 0.98 | 0.69 | 0.35 | 1.00 |
| GC | A1a | 3H-4-naphtho[3,4-b]pyran | 1.24 | 0.66 | 0.98 | 1.29 | 0.55 | 0.98 | 0.66 | 0.45 | 1.00 |
| GC | A1a | 4,7-Methano-1H-indene, 1,8-diethylidene-3a,4,7,7a-tetrahydro-, (3a.alpha.,4.alpha.,7.alpha.,7a.alpha.)- | 0.93 | 0.72 | 0.98 | 0.81 | 0.31 | 0.96 | 1.12 | 0.71 | 1.00 |
| GC | A1a | Diethyl butanedioate | 0.95 | 0.81 | 1.00 | 1.08 | 0.70 | 0.98 | 0.98 | 0.93 | 1.00 |
| GC | A1a | 5,5-Dimethyl-4-phenyl-2-oxolanone | 1.03 | 0.90 | 1.00 | 0.72 | 0.18 | 0.96 | 1.20 | 0.55 | 1.00 |
| GC | A1a | Pentacyclo[5.4.2.0(4,11).0(6,8).0(7,11)]tridec-9-en-1-ol | 1.15 | 0.40 | 0.97 | 1.19 | 0.29 | 0.96 | 0.93 | 0.73 | 1.00 |
| GC | A1a | trans-11,12-Bis(hydroxymethyl)-9,10-dihydro-9,10-ethanoanthracene | 0.53 | 0.32 | 0.97 | 0.98 | 0.97 | 1.00 | 2.64 | 0.23 | 1.00 |
| GC | A1a | 1,2,3,4-Tetrachlorobenzene | 1.31 | 0.55 | 0.98 | 1.00 | 1.00 | 1.00 | 0.99 | 0.99 | 1.00 |
| GC | A1a | Dimethyl 2-(2,2-difluoro-3-oxo-3-phenylpropyl)-4-iodopentanedioate | 0.99 | 0.92 | 1.00 | 1.04 | 0.79 | 0.98 | 1.04 | 0.80 | 1.00 |
| GC | A1a | Methyl 2-(methoxycarbonyl)-2,3-dimethyl-3-phenylbutanoate | 1.79 | 0.12 | 0.97 | 0.56 | 0.19 | 0.96 | 0.65 | 0.44 | 1.00 |
| GC | A1a | Benzothiazole | 1.12 | 0.57 | 0.98 | 0.82 | 0.34 | 0.96 | 1.18 | 0.50 | 1.00 |
| GC | A1a | dioctyl benzene-1,2-dicarboxylate | 0.98 | 0.91 | 1.00 | 0.92 | 0.66 | 0.98 | 0.73 | 0.26 | 1.00 |
| GC | A1a | Dicyclohexyl phthalate | 0.95 | 0.66 | 0.98 | 1.04 | 0.71 | 0.98 | 1.00 | 1.00 | 1.00 |
| GC | A1a | 2-(1,1,2-Trimethylpropyl)cyclohexanone | 0.65 | 0.22 | 0.97 | 2.39 | < 0.05* | 0.57 | 1.35 | 0.44 | 1.00 |
| GC | A1a | (E)-1,2-Diphenyldiazene | 0.92 | 0.71 | 0.98 | 0.98 | 0.94 | 0.99 | 0.96 | 0.87 | 1.00 |
| GC | A1a | 1-(2-Deoxy-.beta.,D-erythropentofuranosyl)-5-isopropyl-4-(trifluoromethyl)-1H-pyrimidin-2-one | 0.99 | 0.92 | 1.00 | 1.05 | 0.70 | 0.98 | 1.00 | 0.99 | 1.00 |
| GC | A1a | 1-(2-bromanyl-1-ethoxy-ethoxy)prop-2-enylbenzene | 0.92 | 0.55 | 0.98 | 0.89 | 0.37 | 0.97 | 0.95 | 0.73 | 1.00 |
| GC | A1a | 1,1,3-Trimethyl-3-(3-methyl-4-oxo-3,4-diphenylbutyl)urea | 0.43 | 0.12 | 0.97 | 1.53 | 0.44 | 0.98 | 1.48 | 0.55 | 1.00 |
| GC | A1a | Vanillin | 1.42 | 0.33 | 0.97 | 1.11 | 0.84 | 0.98 | 0.58 | 0.31 | 1.00 |
| GC | A1a | 3-O-ethyl 4-O-methyl 1-(carbamoylamino)-2-(2-ethoxy-2-oxoethyl)-5-methylpyrrole-3,4-dicarboxylate | 0.95 | 0.85 | 1.00 | 1.15 | 0.59 | 0.98 | 0.98 | 0.95 | 1.00 |
| GC | A1a | 2,4,5-triphenyl-1H-imidazole | 1.26 | 0.11 | 0.97 | 0.95 | 0.76 | 0.98 | 0.96 | 0.85 | 1.00 |
| GC | A1a | 1-(2-Hydroxyphenyl)-2-methyl-1-propanone | 0.89 | 0.53 | 0.98 | 1.03 | 0.84 | 0.98 | 1.30 | 0.24 | 1.00 |
| GC | A1a | 8-Methyl-8H-azepino[1,2-a]indole | 2.42 | 0.15 | 0.97 | 1.87 | 0.33 | 0.96 | 1.12 | 0.89 | 1.00 |
| GC | A1a | 7-p-anisylfuro[3,2-b]pyridine | 0.81 | 0.82 | 1.00 | 0.65 | 0.62 | 0.98 | 0.57 | 0.63 | 1.00 |
| GC | A1a | Dimethyl 5,6-dimethylene-7-oxabicyclo[2.2.1]hept-2-ene-2,3-dicarboxylate | 0.92 | 0.79 | 0.99 | 0.70 | 0.33 | 0.96 | 1.58 | 0.25 | 1.00 |
| GC | A1a | Ethyl 2-Hydroxy-(4-hydroxymethylphenyl)acetate | 0.70 | 0.48 | 0.98 | 1.58 | 0.45 | 0.98 | 1.05 | 0.94 | 1.00 |
| GC | A1a | Bis(3-tetrahydrofuryl) sulfide | 1.46 | 0.27 | 0.97 | 1.47 | 0.28 | 0.96 | 0.61 | 0.24 | 1.00 |
| GC | A1a | 1-(3-Phenylpropyl)piperidine | 0.70 | 0.16 | 0.97 | 1.09 | 0.75 | 0.98 | 1.03 | 0.92 | 1.00 |
| GC | A1a | 1(3H)-Isobenzofuranone, hexahydro-3-(1-methylethyl)-, (3.alpha.,3a.alpha.,7a.beta.)- | 0.93 | 0.62 | 0.98 | 1.13 | 0.44 | 0.98 | 1.37 | 0.15 | 1.00 |
| GC | A1a | [(E)-2-methylsulfanylethenyl]benzene | 1.20 | 0.59 | 0.98 | 1.43 | 0.32 | 0.96 | 0.61 | 0.26 | 1.00 |
| GC | A1a | 2-Methyl-6-nitrobenzaldehyde | 1.50 | 0.20 | 0.97 | 0.83 | 0.57 | 0.98 | 1.24 | 0.60 | 1.00 |
| GC | A1a | 1,9-Dimethylcarbazole | 0.69 | 0.64 | 0.98 | 1.62 | 0.55 | 0.98 | 1.70 | 0.61 | 1.00 |
| GC | A1a | 1-(1,3-dioxolan-2-yl)-1-cyclohexanecarboxaldehyde | 0.76 | 0.32 | 0.97 | 1.13 | 0.68 | 0.98 | 1.09 | 0.81 | 1.00 |
| GC | A1a | Dimethyl 4-[4'-(diethylamino)phenylethynyl]pyridine-2,6-dicarboxylate | 0.66 | 0.20 | 0.97 | 1.48 | 0.22 | 0.96 | 0.61 | 0.22 | 1.00 |
| GC | A1a | (2S,3S,4S,5R)-3,4,5-trimethoxy-6-oxidanyl-oxane-2-carboxylic acid | 1.22 | 0.30 | 0.97 | 0.94 | 0.77 | 0.98 | 1.44 | 0.17 | 1.00 |
| GC | A1a | 4-Isopropyl-2,5-dimethoxybenzyl acetate | 1.61 | 0.45 | 0.97 | 0.44 | 0.18 | 0.96 | 0.97 | 0.96 | 1.00 |
| GC | A1a | 1-(4-Methylphenyl)-1-phenylethanol | 1.10 | 0.63 | 0.98 | 1.29 | 0.20 | 0.96 | 1.35 | 0.30 | 1.00 |
| GC | A1a | 2-Methyl-4'-(methylthio)-2-morpholinopropiophenone | 0.86 | 0.41 | 0.97 | 1.36 | 0.10 | 0.92 | 0.72 | 0.18 | 1.00 |
| GC | A1a | Isopropyl indolizine-2-carboxylate | 1.70 | 0.12 | 0.97 | 1.02 | 0.94 | 0.99 | 1.52 | 0.36 | 1.00 |
| GC | A1a | 9H-Fluorene | 0.78 | 0.13 | 0.97 | 1.11 | 0.54 | 0.98 | 1.53 | 0.05 | 1.00 |
| GC | A1a | 1-(4-Methoxyphenyl)-N-methyl-ethanamine | 0.98 | 0.93 | 1.00 | 1.07 | 0.72 | 0.98 | 1.02 | 0.95 | 1.00 |
| GC | A1a | N-[6'-(Ethynylpyridin-2'-yl)]-acetamide | 1.10 | 0.73 | 0.98 | 0.87 | 0.63 | 0.98 | 2.16 | < 0.05* | 1.00 |
| GC | A1a | Methyl p-hydroxydithiobenzoate | 0.78 | 0.11 | 0.97 | 0.74 | 0.14 | 0.96 | 1.15 | 0.56 | 1.00 |
| GC | A1a | 7-Methyl-3,4-dihydrobenzo[c]pyran-1H-ol | 1.13 | 0.69 | 0.98 | 0.78 | 0.40 | 0.98 | 1.58 | 0.28 | 1.00 |
| GC | A1a | 1-(Methylthio)naphthalene | 0.97 | 0.76 | 0.98 | 0.96 | 0.67 | 0.98 | 0.75 | < 0.05* | 1.00 |
| GC | A1a | Methyl 1,2,3,4-tetrahydro-2-(3',4'-dimethoxybenzyl)-3-oxonaphthalene-2-carboxylate | 1.02 | 0.87 | 1.00 | 0.95 | 0.59 | 0.98 | 1.04 | 0.70 | 1.00 |
| GC | A1a | Dipentyl phthalate | 1.01 | 0.87 | 1.00 | 1.01 | 0.83 | 0.98 | 0.98 | 0.85 | 1.00 |
| GC | A1a | Resorcinol | 0.78 | 0.17 | 0.97 | 1.13 | 0.51 | 0.98 | 1.10 | 0.71 | 1.00 |
| GC | A1a | 2-(2-Hydroxy-2-methylpropyl)pyrrolidine | 1.17 | 0.74 | 0.98 | 1.37 | 0.47 | 0.98 | 0.53 | 0.32 | 1.00 |
| GC | A1a | 4,5-dihydro-2-benzofuran-5-ol | 0.93 | 0.74 | 0.98 | 1.07 | 0.76 | 0.98 | 1.95 | < 0.05* | 1.00 |
| GC | A1a | 4-(Methylamino)benzonitrile | 1.28 | 0.52 | 0.98 | 0.60 | 0.20 | 0.96 | 1.28 | 0.57 | 1.00 |
| GC | A1a | 4-(Hexyloxy)phenol | 1.14 | 0.61 | 0.98 | 0.99 | 0.98 | 1.00 | 1.20 | 0.58 | 1.00 |
| GC | A1a | Neopentyl exo-1-methyl-2-oxabicyclo[3.1.0]hex-3-ene-6-sulfonate | 0.70 | 0.61 | 0.98 | 2.11 | 0.26 | 0.96 | 0.60 | 0.59 | 1.00 |
| GC | A1a | N'-formylbenzohydrazide | 0.83 | 0.46 | 0.97 | 0.95 | 0.84 | 0.98 | 0.58 | 0.15 | 1.00 |
| GC | A1a | diheptyl benzene-1,2-dicarboxylate | 1.04 | 0.81 | 1.00 | 1.25 | 0.26 | 0.96 | 1.63 | 0.07 | 1.00 |
| GC | A1a | (1S,8R)-9-(hydroxymethyl)-9-azabicyclo[6.2.0]decan-10-one | 1.07 | 0.93 | 1.00 | 1.93 | 0.39 | 0.98 | 1.12 | 0.90 | 1.00 |
| GC | A1a | Methyl salicylate | 1.15 | 0.59 | 0.98 | 0.99 | 0.96 | 0.99 | 0.92 | 0.81 | 1.00 |
| GC | A1a | 3,4-Homotropilidene-3-methyl-2,6-dicarbonitrile | 1.23 | 0.72 | 0.98 | 0.94 | 0.92 | 0.99 | 1.16 | 0.83 | 1.00 |
| GC | A1a | 2,3,4,5,6-pentachlorophenol | 1.00 | 0.99 | 1.00 | 1.13 | 0.44 | 0.98 | 1.15 | 0.48 | 1.00 |
| GC | A1a | 3-Isopropyl-4-methoxy-1-(4-methoxyphenyl)-4-methylpent-1-yn-3-ol | 1.12 | 0.62 | 0.98 | 0.48 | < 0.05* | 0.57 | 1.46 | 0.21 | 1.00 |
| GC | A1a | (3RS,5SR)-5-Dimethyl(phenyl)silyl-3-methyhexanonitrile | 0.67 | 0.19 | 0.97 | 1.21 | 0.55 | 0.98 | 0.79 | 0.58 | 1.00 |
| GC | A1a | methyl (9E)-9-methoxyiminononanoate | 0.98 | 0.95 | 1.00 | 1.39 | 0.26 | 0.96 | 0.86 | 0.67 | 1.00 |
| GC | A1a | 1-Naphthyl(veratryl)amine | 1.25 | 0.50 | 0.98 | 0.57 | 0.07 | 0.92 | 1.24 | 0.59 | 1.00 |
| GC | A1a | 1,4-Dimethyl-2-oxa-5-azabicyclo[2.2.2]octan-3,6-dione | 1.16 | 0.43 | 0.97 | 1.47 | < 0.05* | 0.76 | 0.76 | 0.26 | 1.00 |
| GC | A1a | 3-Aminoacetophenone | 0.92 | 0.73 | 0.98 | 1.17 | 0.56 | 0.98 | 0.85 | 0.65 | 1.00 |
| GC | A1a | Ethyl (3-Ethylfuran-2-yl)phenylacetate | 1.14 | 0.63 | 0.98 | 1.20 | 0.52 | 0.98 | 1.21 | 0.64 | 1.00 |
| GC | A1a | 1,4-bis(hept-1-ynyl)benzene | 0.99 | 0.98 | 1.00 | 1.15 | 0.60 | 0.98 | 0.68 | 0.25 | 1.00 |
| GC | A1a | (E)-5-(1,3-benzodioxol-5-yl)-2-pentenal | 1.00 | 0.99 | 1.00 | 1.27 | 0.34 | 0.96 | 0.67 | 0.24 | 1.00 |
| GC | A1a | 1-allyl-2-nitro-cyclohexane | 0.97 | 0.92 | 1.00 | 0.97 | 0.94 | 0.99 | 0.67 | 0.38 | 1.00 |
| GC | A1a | 1-Ethyl-2,3,4,5-tetrahydro-1-benzazepine-2-carbonitrile | 0.98 | 0.89 | 1.00 | 0.95 | 0.77 | 0.98 | 0.90 | 0.61 | 1.00 |
| GC | A1a | 2-(Methylmercapto)benzothiazole | 1.06 | 0.36 | 0.97 | 0.91 | 0.16 | 0.96 | 1.05 | 0.49 | 1.00 |
| GC | A1a | {3,3-bis[4'-(Dimethylamino)phenyl]methyl}-tetrahydrofuran-2-one | 1.21 | 0.39 | 0.97 | 1.12 | 0.60 | 0.98 | 1.37 | 0.24 | 1.00 |
| GC | A1a | 2,2,4,4,6,6-hexamethyl-5-methylidenecyclohexane-1,3-dione | 1.13 | 0.75 | 0.98 | 0.93 | 0.86 | 0.98 | 0.55 | 0.27 | 1.00 |
| GC | A1a | 1-[(R)-phenyl(2-quinolylsulfanyl)methyl]cyclohexanol | 0.79 | 0.62 | 0.98 | 3.76 | < 0.05* | 0.57 | 0.66 | 0.50 | 1.00 |
| GC | A1a | 2-methyl-5-propan-2-yl-1H-pyrrole-3-carbaldehyde | 0.74 | 0.09 | 0.97 | 1.23 | 0.30 | 0.96 | 1.70 | < 0.05* | 1.00 |
| GC | A1a | 3,3-(Ethylenedioxy)-12-methoxyeudesm-7(11)-eno-13,6.alpha.-lactone | 0.81 | 0.27 | 0.97 | 1.06 | 0.75 | 0.98 | 0.81 | 0.37 | 1.00 |
| GC | A1a | (+-)-10,11-trans-11,12-trans-10,11-Dihydro-10-ethyl-12-hydroxy-4-propyl-6,6,11-trimethyl-2H,6H,12H-benzo[1,2-b:3,4:b':5,6-b\]tripyran-2-one"" | 0.90 | 0.63 | 0.98 | 1.31 | 0.24 | 0.96 | 0.72 | 0.24 | 1.00 |
| GC | A1a | methyl 3-hydroxy-2,2-dimethyl-3,3-diphenylpropanoate | 0.84 | 0.80 | 0.99 | 2.25 | 0.20 | 0.96 | 0.52 | 0.45 | 1.00 |
| GC | A1a | Phenyl 4-[bis(ethoxycarbonyl)but-3-ynyl]-2,3,4-trideoxy-.alpha.,L-glcero-pent-2-enopyranoside | 1.98 | 0.10 | 0.97 | 0.80 | 0.56 | 0.98 | 0.67 | 0.38 | 1.00 |
| GC | A1a | 6-methoxy-2,2,4-trimethyl-1H-quinoline | 0.93 | 0.57 | 0.98 | 0.96 | 0.79 | 0.98 | 1.19 | 0.30 | 1.00 |
| GC | A1a | 2-(3,4-Dinitrophenyl)-4-nitro-2H-1,2,3-triazole | 1.64 | 0.05 | 0.97 | 1.07 | 0.80 | 0.98 | 0.84 | 0.60 | 1.00 |
| GC | A1a | 7'-(iodomethyl)-1',7'-dimethylspiro[1,3-dioxolane-2,2'-bicyclo[2.2.1]heptane] | 1.03 | 0.93 | 1.00 | 2.59 | < 0.05* | 0.57 | 0.48 | 0.10 | 1.00 |
| GC | A1a | Coumarin | 0.73 | 0.30 | 0.97 | 1.25 | 0.46 | 0.98 | 0.78 | 0.54 | 1.00 |
| GC | A1a | 3-(2-methyl-5-nitroimidazol-1-yl)propanenitrile | 1.06 | 0.68 | 0.98 | 1.16 | 0.29 | 0.96 | 0.82 | 0.33 | 1.00 |
| GC | A1a | Furmecyclox | 1.09 | 0.75 | 0.98 | 0.86 | 0.61 | 0.98 | 0.79 | 0.48 | 1.00 |
| GC | A1a | 3-(2,2-dimethylpropylidene)bicyclo[3.3.1]nonane-2,4-dione | 0.88 | 0.27 | 0.97 | 1.02 | 0.86 | 0.98 | 0.97 | 0.81 | 1.00 |
| GC | A1a | 4-(dimethylaminoazo)benzonitrile | 1.00 | 1.00 | 1.00 | 1.21 | 0.48 | 0.98 | 0.75 | 0.42 | 1.00 |
| GC | A1a | syn-2,5,5-trimethyl-2-[3-(phenylthio)propyl]tetrahydrofuran-3-ol | 0.84 | 0.21 | 0.97 | 1.05 | 0.74 | 0.98 | 0.85 | 0.35 | 1.00 |
| GC | A1a | 4-[(1S,6S)-Bicyclo[4.1.0]hept-1-yl]butyronitrile | 0.74 | 0.37 | 0.97 | 1.27 | 0.48 | 0.98 | 0.48 | 0.11 | 1.00 |
| GC | A1a | Acenaphthylene | 1.17 | 0.68 | 0.98 | 0.82 | 0.62 | 0.98 | 0.58 | 0.26 | 1.00 |
| GC | A1a | Amantadine | 0.75 | 0.57 | 0.98 | 0.71 | 0.53 | 0.98 | 2.05 | 0.40 | 1.00 |
| GC | A1a | [2-(2-bromopropanoyloxy)phenyl] 4-ethylbenzoate | 1.04 | 0.91 | 1.00 | 0.58 | 0.09 | 0.92 | 1.37 | 0.45 | 1.00 |
| GC | A1a | 5-[(4-methylphenoxy)methyl]-1,3,4-thiadiazol-2-amine | 1.02 | 0.95 | 1.00 | 0.92 | 0.85 | 0.98 | 0.95 | 0.93 | 1.00 |
| GC | A1a | (3aS,5S,11bS)-6,10,11-Trimethoxy-5,9-dimethyl-3,3a,5,11b-tetrahydro-1,4-dioxa-cyclopenta[a]anthracen-2-one | 0.96 | 0.29 | 0.97 | 0.98 | 0.52 | 0.98 | 1.02 | 0.64 | 1.00 |
| GC | A1a | (R,R)5-[3-(Trimethylsilyl)prop-2-yn-1-yl]-methyl-6-phenylcyclohexa-1,3-dien-1-carbaldehyde | 0.79 | 0.27 | 0.97 | 0.94 | 0.76 | 0.98 | 1.05 | 0.87 | 1.00 |
| GC | A1a | Hexyl salicylate | 0.66 | 0.48 | 0.98 | 2.39 | 0.26 | 0.96 | 0.97 | 0.98 | 1.00 |
| GC | A1a | (R)-2-{2-[6-Methoxy-3,4-dihydro-2H-naphthalen-(1E)-ylidene]-ethyl}-2-methyl-3-vinyl-cyclohexanone | 0.71 | 0.21 | 0.97 | 1.13 | 0.68 | 0.98 | 0.74 | 0.45 | 1.00 |
| GC | A1a | 5-Deoxy-5,5-difluoro-2-(formyloxy)-5-iodo-1,3,4-trimethoxy-D-Arabinitol | 1.18 | 0.75 | 0.98 | 2.25 | 0.10 | 0.92 | 0.92 | 0.90 | 1.00 |
| GC | A1a | 1-[4'-(Hydroxybenzyl]-6,7-dimethoxy-5-hydroxy-1,2,3,4-tetrahydro-isoquinoline | 0.60 | 0.38 | 0.97 | 2.02 | 0.18 | 0.96 | 1.60 | 0.52 | 1.00 |
| GC | A1a | 7-t-butyl-1-tetralol | 1.05 | 0.67 | 0.98 | 1.08 | 0.55 | 0.98 | 1.10 | 0.55 | 1.00 |
| GC | A1a | Acetanilide | 0.82 | 0.70 | 0.98 | 5.61 | < 0.05* | 0.57 | 0.84 | 0.79 | 1.00 |
| GC | A1a | (2'S,R,R)-(-)-3-[2'-(1-Ethyl-1-methoxypropy)-2-oxobipyrrolidinyl-3-yl]-3-hexanoic acid methyl ester | 1.99 | 0.09 | 0.97 | 0.96 | 0.92 | 0.99 | 0.98 | 0.97 | 1.00 |
| GC | A1a | 1-Methylphenanthrene | 1.88 | 0.25 | 0.97 | 1.28 | 0.66 | 0.98 | 1.26 | 0.74 | 1.00 |
| GC | A1a | 2,3,9,10,11-Pentamethoxy-5,6,7,8-tetrahydro-dibenzo[a,c]cyclooctene | 0.96 | 0.34 | 0.97 | 0.99 | 0.86 | 0.98 | 1.02 | 0.74 | 1.00 |
| GC | A1a | 1-(4-methoxyphenyl)-2,2-dimethylpropan-1-one | 1.12 | 0.69 | 0.98 | 1.24 | 0.44 | 0.98 | 0.70 | 0.33 | 1.00 |
| GC | A1a | 2,4-Dimethyl-3-(N-methylanilinomethyl)-3-pentanol | 1.47 | 0.15 | 0.97 | 0.96 | 0.90 | 0.99 | 0.44 | 0.07 | 1.00 |
| GC | A1a | Butyl Benzoate | 1.16 | 0.60 | 0.98 | 1.56 | 0.31 | 0.96 | 1.36 | 0.55 | 1.00 |
| GC | A1a | Diallyl maleate | 0.95 | 0.51 | 0.98 | 1.00 | 0.97 | 0.99 | 0.99 | 0.89 | 1.00 |
| GC | A1a | 2-(hydroxymethyl)-1-(3-methoxyphenyl)cyclopropanecarbonitrile | 1.14 | 0.69 | 0.98 | 0.95 | 0.88 | 0.99 | 1.96 | 0.14 | 1.00 |
| GC | A1a | 4-hydroxy-3,5-dimethoxybenzaldehyde | 0.87 | 0.79 | 0.99 | 1.31 | 0.55 | 0.98 | 2.00 | 0.17 | 1.00 |
| GC | A1a | 5-(4-Methoxyphenyl)-7-benzyl-1,11,11-trimethyl-5-aza-2-oxatetracyclo[6.5.0.0(3,6).010,12)]tridecan-4-one | 0.94 | 0.80 | 0.99 | 1.78 | < 0.05* | 0.67 | 0.70 | 0.27 | 1.00 |
| GC | A1a | Binapacryl | 0.59 | 0.13 | 0.97 | 1.41 | 0.32 | 0.96 | 0.65 | 0.30 | 1.00 |
| GC | A1a | Caffeine | 0.86 | < 0.05* | 0.97 | 1.00 | 0.99 | 1.00 | 1.06 | 0.56 | 1.00 |
| GC | A1a | delta-Tocopherol | 0.97 | 0.82 | 1.00 | 1.44 | < 0.05* | 0.57 | 1.01 | 0.95 | 1.00 |
| GC | A1a | Phenylparaben | 0.91 | 0.52 | 0.98 | 1.18 | 0.27 | 0.96 | 0.96 | 0.85 | 1.00 |
| GC | A1a | 4-Methyl-1-(2,4,5-trimethylphenyl)-1-pentanone | 2.02 | 0.21 | 0.97 | 1.23 | 0.75 | 0.98 | 0.58 | 0.46 | 1.00 |
| GC | A1a | 1-(1H-indazol-3-yl)propanol | 0.51 | < 0.05* | 0.97 | 1.10 | 0.75 | 0.98 | 1.41 | 0.40 | 1.00 |
| GC | A1a | Benzenecarbothioic acid, S-2-cyclopenten-1-yl ester | 0.98 | 0.94 | 1.00 | 1.06 | 0.78 | 0.98 | 0.82 | 0.50 | 1.00 |
| GC | A1a | 1-Naphthalenethiol | 0.97 | 0.87 | 1.00 | 1.49 | 0.08 | 0.92 | 0.80 | 0.40 | 1.00 |
| GC | A1a | 1-[6-(2-hydroxy-2-methyl-propyl)-2-pyridyl]-2-methyl-propan-2-ol | 0.94 | 0.79 | 0.99 | 0.92 | 0.65 | 0.98 | 1.38 | 0.28 | 1.00 |
| GC | A1a | 3-Methyl-1-phenyl-1-pentyn-3-ol | 1.28 | 0.56 | 0.98 | 1.55 | 0.32 | 0.96 | 0.74 | 0.58 | 1.00 |
| GC | A1a | Bicyclo[3.3.1]nonan-2-one, 1,4,5-trimethyl-9-methylene-, exo-(.+-.)- | 1.10 | 0.73 | 0.98 | 1.31 | 0.38 | 0.97 | 0.83 | 0.63 | 1.00 |
| GC | A1a | methyl (2S)-5-ethoxy-3,4-dihydro-2H-pyrrole-2-carboxylate | 1.30 | 0.46 | 0.97 | 2.48 | < 0.05* | 0.57 | 0.53 | 0.15 | 1.00 |
| GC | A1a | dimethyl benzene-1,2-dicarboxylate | 1.65 | 0.09 | 0.97 | 0.90 | 0.75 | 0.98 | 1.75 | 0.20 | 1.00 |
| GC | A1a | Oxybenzone | 1.04 | 0.74 | 0.98 | 0.91 | 0.40 | 0.98 | 0.90 | 0.54 | 1.00 |
| GC | A1a | 3,3'-Diacetyl-1,1'-dimethyl-1H,1'H-2,2'-bisindolyl | 0.88 | 0.57 | 0.98 | 1.59 | 0.07 | 0.89 | 0.83 | 0.55 | 1.00 |
| GC | A1a | PCB 114 | 0.85 | 0.25 | 0.97 | 1.05 | 0.76 | 0.98 | 1.06 | 0.74 | 1.00 |
| GC | A1a | 2-(1,3-benzoxazol-2-yl)ethanol | 0.97 | 0.95 | 1.00 | 2.51 | < 0.05* | 0.57 | 0.38 | 0.06 | 1.00 |
| GC | A1a | 6-tert-Butyl-2-phenyl-4-propan-2-ylidene-cyclopenta[c]pyrazole | 0.97 | 0.88 | 1.00 | 0.97 | 0.87 | 0.99 | 0.70 | 0.22 | 1.00 |
| GC | A1a | 2-(2-aminophenyl)-2-adamantanecarbonitrile | 0.95 | 0.75 | 0.98 | 1.48 | < 0.05* | 0.76 | 1.02 | 0.92 | 1.00 |
| GC | A1a | 2,2-bis(prop-2-enyl)pyrrolidine | 0.64 | 0.18 | 0.97 | 1.50 | 0.27 | 0.96 | 0.85 | 0.53 | 1.00 |
| GC | A1a | 1-[4-(Dimethoxymethyl)phenyl]ethanone | 1.40 | 0.44 | 0.97 | 1.16 | 0.75 | 0.98 | 1.08 | 0.88 | 1.00 |
| GC | A1a | trans-11-Hydroxy-6-(hydroxymethyl)-2,3,10-trimethoxydibenzo[1a,4a:8a,12a]cyclooctadiene-7-carboxylic acid lactone | 1.08 | 0.47 | 0.97 | 1.12 | 0.28 | 0.96 | 1.08 | 0.60 | 1.00 |
| GC | A1a | gamma-Nonanolactone | 0.90 | 0.71 | 0.98 | 1.35 | 0.31 | 0.96 | 0.48 | 0.07 | 1.00 |
| GC | A1a | 1,2,2-Butanetricarboxylic acid, 4-(tetrahydro-2-methyl-2-furanyl)-, 2-ethyl ester | 0.69 | 0.21 | 0.97 | 1.06 | 0.85 | 0.98 | 0.91 | 0.81 | 1.00 |
| GC | A1a | 1-Furfurylpyrrole | 0.75 | 0.63 | 0.98 | 3.67 | < 0.05* | 0.67 | 1.02 | 0.98 | 1.00 |
| GC | A1a | 3-phenylprop-2-yn-1-ol | 1.00 | 1.00 | 1.00 | 1.13 | 0.84 | 0.98 | 0.93 | 0.93 | 1.00 |
| GC | A1a | Mequinol | 0.62 | 0.35 | 0.97 | 2.73 | 0.06 | 0.86 | 1.15 | 0.84 | 1.00 |
| GC | A1a | (+-)-(1aR,2R)-1a-Methyl-2-phenylperhydrooxireno[2,3-c]furan | 0.63 | 0.30 | 0.97 | 1.12 | 0.80 | 0.98 | 1.83 | 0.28 | 1.00 |
| GC | A1a | Diphenyl sulfone | 0.73 | 0.35 | 0.97 | 0.90 | 0.79 | 0.98 | 0.86 | 0.77 | 1.00 |
| GC | A1a | methyl 6-(oxolan-2-yloxy)hexanoate | 1.14 | 0.58 | 0.98 | 0.99 | 0.96 | 0.99 | 0.96 | 0.87 | 1.00 |
| GC | A1a | Methyl paraben | 1.09 | 0.45 | 0.97 | 0.98 | 0.83 | 0.98 | 1.02 | 0.87 | 1.00 |
| GC | A1a | (1R(*),3S(*).5S(*).7S(*))-1,3-Dimethyl-7-hydroxy-6-oxabicyclo[3.2.1]octan-2-one | 1.03 | 0.93 | 1.00 | 1.39 | 0.52 | 0.98 | 0.88 | 0.85 | 1.00 |
| GC | A1a | 2,2,4,7-tetramethyl-3H-1-benzofuran | 1.34 | 0.52 | 0.98 | 1.29 | 0.57 | 0.98 | 2.61 | 0.06 | 1.00 |
| GC | A1a | Propylidene-4,4-ethylenedioxycyclohexane | 0.74 | 0.49 | 0.98 | 1.94 | 0.16 | 0.96 | 0.76 | 0.58 | 1.00 |
| GC | A1a | 4-[(7-Fluoro-2,3-dihydro-1,4-benzodioxin-2-yl)methyl]morpholine | 1.14 | 0.63 | 0.98 | 0.91 | 0.80 | 0.98 | 1.12 | 0.79 | 1.00 |
| GC | A1a | (2,4,4-Trimethyl-6-oxo-cyclohex-1-enyl)-acetic acid ethyl ester | 0.72 | 0.11 | 0.97 | 1.35 | 0.18 | 0.96 | 0.47 | < 0.05* | 1.00 |
| GC | A1a | 2-but-2-ynoxyoxane | 0.63 | < 0.05* | 0.97 | 1.21 | 0.41 | 0.98 | 1.98 | < 0.05* | 1.00 |
| GC | A1a | 2-(4-Chlorophenyl)-2-oxoethyl oct-2-ynoate | 1.21 | 0.41 | 0.97 | 1.02 | 0.95 | 0.99 | 1.86 | < 0.05* | 1.00 |
| GC | A1a | (1S)-3-(3'a-Hydroxy-7'a-methyl-5'-oxo-perhydroinden-1'-yl] acrylate | 1.32 | 0.19 | 0.97 | 1.03 | 0.89 | 0.99 | 1.02 | 0.94 | 1.00 |
| GC | A1a | .alpha.-Fluoro-(p-methyl)chalcone | 0.96 | 0.77 | 0.99 | 1.02 | 0.90 | 0.99 | 0.99 | 0.97 | 1.00 |
| GC | A1a | Phenyl salicylate | 0.93 | 0.75 | 0.98 | 0.82 | 0.46 | 0.98 | 1.21 | 0.56 | 1.00 |
| GC | A1a | 2,2'-Methylenebis(4-methyl-6-tert-butylphenol) | 1.07 | 0.53 | 0.98 | 1.39 | < 0.05* | 0.57 | 0.92 | 0.53 | 1.00 |
| GC | A1a | (4-nitrophenyl) 2-methylpropanoate | 0.80 | 0.58 | 0.98 | 1.17 | 0.68 | 0.98 | 1.21 | 0.82 | 1.00 |
| GC | A1a | 1,3-diethyl-1,3,3a,9a-tetrahydrobenzo[f][2]benzothiole-4,9-dione | 0.69 | < 0.05* | 0.97 | 1.10 | 0.62 | 0.98 | 0.91 | 0.68 | 1.00 |
| GC | A1a | 7-anti-pentyl-3,3-dimethylnorbornano-1,2-endo-diol | 1.07 | 0.59 | 0.98 | 1.01 | 0.94 | 0.99 | 0.96 | 0.83 | 1.00 |
| GC | A1a | phenyl N-methylcarbamate | 1.10 | 0.69 | 0.98 | 1.52 | 0.12 | 0.93 | 2.00 | < 0.05* | 1.00 |
| GC | A1a | 15-(methoxymethoxy)-3-methylene-1-pentadecanol | 0.78 | < 0.05* | 0.97 | 1.03 | 0.77 | 0.98 | 0.96 | 0.80 | 1.00 |
| GC | A1a | 2,7-Dimethoxy-1,3,4,5,6,8-hexamethyl-9-isopropyl-9H-xanthene | 1.35 | 0.07 | 0.97 | 0.87 | 0.46 | 0.98 | 0.94 | 0.77 | 1.00 |
| GC | A1a | 5-Methyl-2-(1H-pyrrol-2-yl)benzo[d]oxazole | 1.13 | 0.56 | 0.98 | 1.00 | 1.00 | 1.00 | 1.03 | 0.92 | 1.00 |
| GC | A1a | Pyracarbolid | 0.76 | 0.13 | 0.97 | 1.37 | 0.08 | 0.92 | 0.92 | 0.73 | 1.00 |
| GC | A1a | 2,2,4-Trimethylpentane-1,3-diyl dibenzoate | 0.92 | 0.55 | 0.98 | 0.83 | 0.26 | 0.96 | 1.03 | 0.87 | 1.00 |
| GC | A1a | 2-Phenyl[4,7]phenanthrolino[5,6-d]oxazole | 0.65 | 0.48 | 0.98 | 2.56 | 0.14 | 0.96 | 0.69 | 0.60 | 1.00 |
| GC | A1a | 4-(1,1,4,4,7-pentamethyltetralin-6-yl)sulfonylbenzoic acid | 1.04 | 0.89 | 1.00 | 0.58 | < 0.05* | 0.67 | 1.61 | 0.13 | 1.00 |
| GC | A1a | (1-Methoxypentyl)benzene | 0.52 | 0.35 | 0.97 | 1.00 | 1.00 | 1.00 | 2.03 | 0.39 | 1.00 |
| GC | A1a | Methyl 3-Methyl-5-pentanoyl-2-furoate | 1.38 | 0.44 | 0.97 | 1.08 | 0.83 | 0.98 | 0.84 | 0.72 | 1.00 |
| GC | A1a | 2-(12'-methoxy-19'-norpodocarpa-8',11',13-trien-18'-yl)ethyl methanesulfonate | 0.88 | 0.65 | 0.98 | 1.71 | < 0.05* | 0.76 | 0.97 | 0.94 | 1.00 |
| GC | A1a | 2,9-dihydro-\$b-carbolin-1-one | 0.75 | 0.35 | 0.97 | 1.46 | 0.25 | 0.96 | 1.21 | 0.64 | 1.00 |
| GC | A1a | 4-(1H-pyrazolyl-1-yl)indol-2-one | 0.82 | 0.30 | 0.97 | 1.16 | 0.40 | 0.98 | 0.82 | 0.47 | 1.00 |
| GC | A1a | (8R,9R)-Isocaryolane-8,9-diol | 0.77 | 0.22 | 0.97 | 1.20 | 0.39 | 0.98 | 0.78 | 0.36 | 1.00 |
| GC | A1a | methyl (E)-4-(1-hydroxy-3,4-dihydro-2H-triphenylen-1-yl)but-2-enoate | 0.69 | 0.28 | 0.97 | 1.93 | 0.06 | 0.86 | 0.77 | 0.55 | 1.00 |
| GC | A1a | Bis(2-ethylhexyl) phthalate | 0.98 | 0.93 | 1.00 | 1.27 | 0.30 | 0.96 | 1.46 | 0.20 | 1.00 |
| GC | A1a | 1,3-dimethyl-5-(2-methylpropylidene)-1,3-diazinane-2,4,6-trione | 0.86 | 0.72 | 0.98 | 1.95 | 0.08 | 0.92 | 0.81 | 0.68 | 1.00 |
| GC | A1a | alpha-Tocopherol | 1.07 | 0.74 | 0.98 | 1.38 | 0.12 | 0.92 | 0.64 | 0.09 | 1.00 |
| GC | A1a | (2-Hydroxyphenyl)acetonitrile | 1.30 | 0.44 | 0.97 | 1.02 | 0.96 | 0.99 | 1.19 | 0.68 | 1.00 |
| GC | A1a | 5-phenyl-3,4-dihydro-2H-pyrrole | 0.73 | 0.44 | 0.97 | 2.04 | 0.11 | 0.92 | 1.37 | 0.61 | 1.00 |
| GC | A1a | methyl 3-[(1R,12bR)-3-acetyl-1,2,6,7,12,12b-hexahydroindolo[2,3-a]quinolizin-1-yl]-2-chloropropanoate | 0.94 | 0.81 | 1.00 | 1.14 | 0.60 | 0.98 | 0.97 | 0.94 | 1.00 |
| GC | A1a | (5S,6R)-5,6-Dihydroxy-cyclohex-3-enecarboxylic acid prop-2-ynyl ester | 0.84 | 0.51 | 0.98 | 1.17 | 0.59 | 0.98 | 1.39 | 0.36 | 1.00 |
| GC | A1a | Octocrylene | 0.96 | 0.76 | 0.98 | 1.03 | 0.82 | 0.98 | 0.70 | 0.11 | 1.00 |
| GC | A1a | 4-Methyl-4-(2-oxanyloxy)-2-pentanone | 0.97 | 0.92 | 1.00 | 0.91 | 0.75 | 0.98 | 0.81 | 0.55 | 1.00 |
| GC | A1a | 1-(cyclopentylmethyl)-4-methoxy-benzene | 0.74 | 0.33 | 0.97 | 0.80 | 0.42 | 0.98 | 1.35 | 0.44 | 1.00 |
| GC | A1a | Triethylene glycol diacetate | 0.94 | 0.61 | 0.98 | 1.05 | 0.69 | 0.98 | 0.98 | 0.90 | 1.00 |
| GC | A1a | (2,6-dimethyl-4-propyl-phenyl)amine | 0.75 | 0.60 | 0.98 | 1.59 | 0.34 | 0.96 | 0.74 | 0.50 | 1.00 |
| GC | A1a | 2-O-cyclohexyl 1-O-pentyl benzene-1,2-dicarboxylate | 1.00 | 0.99 | 1.00 | 1.01 | 0.90 | 0.99 | 1.04 | 0.70 | 1.00 |
| GC | A1a | 3-(2-(Piperidin-1-yl)acetamido)benzoic acid | 0.79 | 0.42 | 0.97 | 1.22 | 0.53 | 0.98 | 0.85 | 0.67 | 1.00 |
| GC | A1a | Morpholyl-2-oxocyclohexanecarboxamide | 0.97 | 0.89 | 1.00 | 1.12 | 0.58 | 0.98 | 0.78 | 0.37 | 1.00 |
| GC | A1a | 1-Methoxy-1-(4-methoxyphenyl)-4-phenylbutane | 1.35 | 0.60 | 0.98 | 0.64 | 0.31 | 0.96 | 0.83 | 0.79 | 1.00 |
| GC | A1a | 3,5,5-trimethyl-4-[(E)-3-oxidanylbut-1-enyl]cyclohex-3-en-1-ol | 0.91 | 0.73 | 0.98 | 0.99 | 0.97 | 0.99 | 0.87 | 0.68 | 1.00 |
| GC | A1a | Kikkanol F monoacetate | 0.63 | 0.11 | 0.97 | 1.07 | 0.82 | 0.98 | 0.78 | 0.53 | 1.00 |
| GC | A1a | 5,12-Dihydroxysterpyrene | 1.05 | 0.89 | 1.00 | 0.87 | 0.67 | 0.98 | 0.42 | < 0.05* | 1.00 |
| GC | A1a | 4-Methylene-2-phenyl-tetrahydrofuran | 0.95 | 0.87 | 1.00 | 1.48 | 0.27 | 0.96 | 0.93 | 0.88 | 1.00 |
| GC | A1a | Phenyl benzoate | 0.79 | 0.76 | 0.99 | 5.42 | < 0.05* | 0.76 | 1.09 | 0.91 | 1.00 |
| GC | A1a | 2-Isobutyryl-benzo[1,3]dioxol-5-carbaldehyde | 1.07 | 0.82 | 1.00 | 1.19 | 0.58 | 0.98 | 0.88 | 0.75 | 1.00 |
| GC | A1a | DL-trans-2-acetoxy-1-benzoyloxymethyl-3-[(E)-cinnamoyloxy]-4,6-cyclohexadiene | 0.74 | 0.19 | 0.97 | 1.16 | 0.50 | 0.98 | 0.88 | 0.66 | 1.00 |
| GC | A1a | methyl 2-hydroxy-2-(3-methoxyphenyl)acetate | 1.08 | 0.66 | 0.98 | 0.99 | 0.93 | 0.99 | 0.79 | 0.33 | 1.00 |
| GC | A1a | Methyl 2-(3',3'-dimethyl-1'-butyn-1'-yl)-1-cyclohexenecarboxylate | 0.88 | 0.34 | 0.97 | 1.45 | < 0.05* | 0.57 | 0.88 | 0.46 | 1.00 |
| GC | A1a | 3,4-dihydro-2H-naphthalen-1-one | 0.90 | 0.69 | 0.98 | 1.07 | 0.82 | 0.98 | 0.98 | 0.94 | 1.00 |
| GC | A1a | 4-(1-Pyrrolidinyl)pyridazine | 1.37 | 0.29 | 0.97 | 0.68 | 0.24 | 0.96 | 1.08 | 0.86 | 1.00 |
| GC | A1a | Phenyl N-benzoyl-N-methyl-thiooxocarbamate | 1.10 | 0.64 | 0.98 | 1.11 | 0.62 | 0.98 | 1.01 | 0.97 | 1.00 |
| GC | A1a | Furaltadone | 0.96 | 0.94 | 1.00 | 1.13 | 0.81 | 0.98 | 0.50 | 0.30 | 1.00 |
| GC | A1a | Benzyl butyl phthalate | 1.04 | 0.79 | 0.99 | 1.07 | 0.71 | 0.98 | 1.21 | 0.46 | 1.00 |
| GC | A1a | 3-benzyl-4,7,7-trimethyl-3-azabicyclo[2.2.1]heptan-2-one | 0.89 | 0.84 | 1.00 | 1.48 | 0.54 | 0.98 | 0.58 | 0.49 | 1.00 |
| GC | A1a | Dicrotophos | 0.79 | 0.49 | 0.98 | 1.56 | 0.19 | 0.96 | 0.95 | 0.92 | 1.00 |
| GC | A1a | 1-ethyl-3-[(E)-hydroxyiminomethyl]pyridine-2-thione | 0.80 | 0.70 | 0.98 | 1.11 | 0.87 | 0.99 | 0.42 | 0.30 | 1.00 |
| GC | A1a | (R)-6-Phenyltetrahydro-2H-pyran-2-one | 1.21 | 0.56 | 0.98 | 1.41 | 0.33 | 0.96 | 0.78 | 0.57 | 1.00 |
| GC | A1a | Beta-BHC | 0.70 | 0.31 | 0.97 | 1.00 | 0.99 | 1.00 | 1.39 | 0.54 | 1.00 |
| GC | A1a | o-Phenoxybenzyl Alcohol | 0.66 | 0.45 | 0.97 | 3.02 | < 0.05* | 0.76 | 1.46 | 0.59 | 1.00 |
| GC | A1a | 2,6-Diisopropylnaphthalene | 0.93 | 0.88 | 1.00 | 1.44 | 0.42 | 0.98 | 0.77 | 0.71 | 1.00 |
| GC | A1a | 4-[[[2-(Dimethylamino)ethyl]methylamino]methyl]phenol | 0.98 | 0.91 | 1.00 | 1.07 | 0.71 | 0.98 | 1.01 | 0.95 | 1.00 |
| GC | A1a | Benzothiazolone | 1.33 | 0.14 | 0.97 | 0.80 | 0.30 | 0.96 | 1.12 | 0.64 | 1.00 |
| GC | A1a | 2-[(4S)-2,2,4-trimethyl-1,3-dioxolan-4-yl]ethyl methanesulfonate | 0.92 | 0.68 | 0.98 | 0.95 | 0.80 | 0.98 | 1.22 | 0.45 | 1.00 |
| GC | A1a | Fluorene | 0.57 | 0.23 | 0.97 | 1.15 | 0.76 | 0.98 | 1.16 | 0.80 | 1.00 |
| GC | A1a | 8,9,11,12-Tetraimethoxy-1-methyl-1,2,3,4-dihydronaphtho[2,1-f]isoquinoline | 0.86 | 0.58 | 0.98 | 1.32 | 0.32 | 0.96 | 0.85 | 0.67 | 1.00 |
| GC | A1a | 1-(4-Chlorophenyl)-2-methylbut-3-en-1-ol | 0.96 | 0.90 | 1.00 | 1.68 | 0.17 | 0.96 | 1.73 | 0.24 | 1.00 |
| GC | A1a | n-Undecyl cyanide | 1.23 | 0.71 | 0.98 | 2.12 | 0.26 | 0.96 | 0.38 | 0.27 | 1.00 |
| GC | A1a | 4-[(2-methylacridin-9-yl)amino]-N-(1,3-thiazol-2-yl)benzenesulfonamide | 0.88 | 0.58 | 0.98 | 1.00 | 0.99 | 1.00 | 1.53 | 0.27 | 1.00 |
| GC | A1a | Octisalate | 0.75 | 0.67 | 0.98 | 4.46 | 0.09 | 0.92 | 0.39 | 0.33 | 1.00 |
| GC | A1a | 2-(4-Fluorophenyl)isoindoline | 1.19 | 0.45 | 0.97 | 1.29 | 0.30 | 0.96 | 0.85 | 0.65 | 1.00 |
| GC | A1a | Ethylparaben | 0.49 | 0.39 | 0.97 | 2.77 | 0.21 | 0.96 | 1.50 | 0.57 | 1.00 |
| GC | A1a | (2S,6R)-4-[3-(4-tert-butylphenyl)-2-methylpropyl]-2,6-dimethylmorpholine | 1.92 | 0.29 | 0.97 | 2.03 | 0.27 | 0.96 | 0.22 | 0.07 | 1.00 |
| GC | A1a | Octa-1,7-diyne-3,6-diol | 0.99 | 0.96 | 1.00 | 1.11 | 0.64 | 0.98 | 1.08 | 0.82 | 1.00 |
| GC | A1a | (E)-stilbene | 1.14 | 0.43 | 0.97 | 1.11 | 0.57 | 0.98 | 0.97 | 0.92 | 1.00 |
| GC | A1a | 2-butyl-6-methoxy-2,5-dihydropyridine | 0.94 | 0.77 | 0.99 | 1.03 | 0.88 | 0.99 | 1.28 | 0.38 | 1.00 |
| GC | A1a | 2-Methoxy-N,N-dimethyl-2-phenylacetamide | 0.86 | 0.49 | 0.98 | 1.78 | < 0.05* | 0.57 | 0.93 | 0.81 | 1.00 |
| GC | A1a | .beta.-1,5-anhydro-2,3,4,6-tetra-O-methyl-1-C-phenyl-D-glucitol | 0.74 | 0.19 | 0.97 | 0.89 | 0.63 | 0.98 | 0.84 | 0.52 | 1.00 |
| GC | A1a | 1-phenyl-2-sulfanylidene-3-pyridinecarboxylic acid | 1.27 | 0.56 | 0.98 | 1.15 | 0.70 | 0.98 | 1.05 | 0.93 | 1.00 |
| GC | A1a | Allyltri(4-N,N-dimethylaminophenyl)silane | 0.51 | < 0.05* | 0.97 | 1.08 | 0.79 | 0.98 | 0.86 | 0.67 | 1.00 |
| GC | A1a | 7,10-Dioxo-8,9-dihydro[4]-paracyclophane | 0.94 | 0.70 | 0.98 | 1.02 | 0.90 | 0.99 | 1.11 | 0.62 | 1.00 |
| GC | A1a | Pyrolo[3,2-d]pyrimidin-2,4(1H,3H)-dione | 1.00 | 0.99 | 1.00 | 0.90 | 0.67 | 0.98 | 1.29 | 0.42 | 1.00 |
| GC | A1a | Mepivacaine | 1.48 | < 0.05* | 0.97 | 1.14 | 0.49 | 0.98 | 0.97 | 0.89 | 1.00 |
| GC | A1a | 4-Chlorophenol | 1.01 | 0.99 | 1.00 | 1.01 | 0.98 | 1.00 | 1.07 | 0.89 | 1.00 |
| GC | A1a | N-[(E)-3-cyano-1,1-dimethyl-allyl]acetamide | 0.83 | 0.62 | 0.98 | 1.18 | 0.64 | 0.98 | 1.00 | 1.00 | 1.00 |
| GC | A1a | Acetic Acid (+-)-(1.alpha.,3.beta.,4.beta.,6.alpha.,7.alpha.)-7-(3-Methyl-3-butenyl)-4-((E)-6-methyl-2,6-heptadienyl)-1-methylbicyclo[4.1.0]hept-3-yl Ester | 0.94 | 0.36 | 0.97 | 1.02 | 0.74 | 0.98 | 1.02 | 0.86 | 1.00 |
| GC | A1a | 2-Amino-2'-tert-butylthio-6-ethoxy[3,4']bipyridine-3,5-dicarbonitrile | 1.33 | 0.20 | 0.97 | 1.81 | < 0.05* | 0.57 | 1.07 | 0.83 | 1.00 |
| GC | A1a | Pyrene | 0.42 | 0.07 | 0.97 | 0.80 | 0.64 | 0.98 | 1.29 | 0.67 | 1.00 |
| GC | A1a | trans-2,3-di(methoxycarbonyl)-bicyclo[2.2.2]oct-5-ene | 1.04 | 0.91 | 1.00 | 0.78 | 0.56 | 0.98 | 1.06 | 0.92 | 1.00 |
| GC | A1a | 3-(4-Vinylbenzyloxy)benzyl alcohol | 1.41 | 0.68 | 0.98 | 2.65 | 0.22 | 0.96 | 0.24 | 0.17 | 1.00 |
| GC | A1a | methyl 2-pyrimidin-4-ylpropanoate | 1.01 | 0.96 | 1.00 | 1.03 | 0.88 | 0.99 | 1.54 | 0.17 | 1.00 |
| GC | A1a | 2-methyl-1H-benzimidazole | 1.24 | 0.51 | 0.98 | 1.09 | 0.82 | 0.98 | 1.30 | 0.62 | 1.00 |
| GC | A1a | (R)-2-N-Heptyl (S)-O-1-phenylethyl carbamate | 0.53 | 0.19 | 0.97 | 1.81 | 0.27 | 0.96 | 0.48 | 0.25 | 1.00 |
| GC | A1a | ethyl (2E,4E)-6-methoxy-6-sulfanylidenehexa-2,4-dienoate | 1.30 | 0.49 | 0.98 | 1.21 | 0.62 | 0.98 | 0.99 | 0.99 | 1.00 |
| GC | A1a | 1-(Biphenyl-4-yl)ethanone | 0.91 | 0.29 | 0.97 | 0.96 | 0.60 | 0.98 | 1.13 | 0.24 | 1.00 |
| GC | A1a | ethyl 2-((2r,4R,6S)-2,4,6-trimethyl-1,3-dioxan-2-yl)acetate | 0.99 | 0.95 | 1.00 | 1.08 | 0.62 | 0.98 | 0.98 | 0.91 | 1.00 |
| GC | A1a | 2-methyl-5-phenyl-1H-pyrrole | 0.69 | 0.19 | 0.97 | 1.66 | 0.10 | 0.92 | 2.18 | < 0.05* | 1.00 |
| GC | A1a | (6R,7R)-7-isopropyl-1,3-dihydro-1,5,5,6-tetramethyl-indan[5,6-c]furan | 1.05 | 0.81 | 1.00 | 1.31 | 0.12 | 0.92 | 0.89 | 0.65 | 1.00 |
| GC | A1a | 3,3,3-trifluoro-2-hydroxy-1-(4-methylphenyl)-1-propanone | 1.59 | 0.11 | 0.97 | 1.29 | 0.35 | 0.96 | 0.54 | 0.10 | 1.00 |
| GC | A1a | (4-methoxy-3-propan-2-yloxy-phenyl) ethanoate | 1.10 | 0.82 | 1.00 | 1.37 | 0.41 | 0.98 | 1.16 | 0.79 | 1.00 |
| GC | A1a | benzhydryl 2-(1-methyl-3-oxo-2-prop-2-enylcyclohexyl)acetate | 0.93 | 0.74 | 0.98 | 1.15 | 0.54 | 0.98 | 1.09 | 0.76 | 1.00 |
| GC | A1a | 9-Methyl-9,10-dihydroanthracene | 1.37 | 0.52 | 0.98 | 0.87 | 0.75 | 0.98 | 2.29 | 0.13 | 1.00 |
| GC | A1a | (3aR,5R,7aR)-1,1-Ethylidenedioxy-7a-methyl-3a,4,5,6,7,7a-hexahydroindan-5-ol | 0.98 | 0.94 | 1.00 | 1.26 | 0.41 | 0.98 | 0.93 | 0.85 | 1.00 |
| GC | A1a | N-(cyclohexylmethyl)morpholine | 0.84 | 0.44 | 0.97 | 1.24 | 0.39 | 0.98 | 0.92 | 0.81 | 1.00 |
| GC | A1a | (E)-.beta.-(tert-Butyldimethylsiloxy)-.beta.-(tert-butyldimethylsilyl)styrene | 1.05 | 0.89 | 1.00 | 1.24 | 0.61 | 0.98 | 0.55 | 0.23 | 1.00 |
| GC | A1a | 6-Heptadecylthymoquinone | 0.89 | 0.43 | 0.97 | 1.35 | < 0.05* | 0.70 | 1.06 | 0.75 | 1.00 |
| GC | A1a | 2,5-Pyrrolidinedione, 1-[11-(4-morpholinyl)bicyclo[8.1.0]undec-11-yl]-, stereoisomer | 0.80 | 0.69 | 0.98 | 3.35 | 0.07 | 0.92 | 1.18 | 0.83 | 1.00 |
| GC | A1a | (Z)-3-(4-n-Butyl-3-thienyl)propenal | 0.81 | 0.25 | 0.97 | 1.18 | 0.37 | 0.97 | 0.97 | 0.88 | 1.00 |
| GC | A1a | (S)-(4-butoxyphenyl)(1-phthalimdoethyl)-ketone | 0.87 | 0.41 | 0.97 | 1.18 | 0.32 | 0.96 | 1.10 | 0.68 | 1.00 |
| GC | A1a | Acetyl tributyl citrate | 0.80 | 0.24 | 0.97 | 1.19 | 0.33 | 0.96 | 0.68 | 0.07 | 1.00 |
| GC | A1a | (4S,5R)-3-(3-methyl-1-oxobut-2-enyl)-4,5-diphenyl-2-oxazolidinone | 0.50 | 0.33 | 0.97 | 2.62 | 0.18 | 0.96 | 0.37 | 0.28 | 1.00 |
| GC | A1a | 3-Isopropoxy-5-methyl-1H-pyrazole | 0.86 | 0.78 | 0.99 | 1.82 | 0.24 | 0.96 | 0.33 | 0.13 | 1.00 |
| GC | A1a | N'-(2-methylpropyl)benzohydrazide | 0.78 | 0.29 | 0.97 | 1.37 | 0.34 | 0.96 | 1.79 | 0.11 | 1.00 |
| GC | A1a | 3-Butanoyloxy-2-(2-propenyl)tetrahydropyran | 0.89 | 0.73 | 0.98 | 1.37 | 0.45 | 0.98 | 0.66 | 0.40 | 1.00 |
| GC | A1a | 2-Phenoxyethyl propanoate | 0.69 | 0.39 | 0.97 | 1.64 | 0.28 | 0.96 | 1.52 | 0.46 | 1.00 |
| GC | A1a | Molinate | 1.51 | 0.05 | 0.97 | 0.78 | 0.27 | 0.96 | 0.90 | 0.70 | 1.00 |
| GC | A1a | 1-Decyclopentane-1,3-dione | 0.88 | 0.68 | 0.98 | 1.24 | 0.48 | 0.98 | 1.09 | 0.83 | 1.00 |
| GC | A1a | 1-(4-phenyl-1H-pyrrol-3-yl)propan-1-one | 0.83 | 0.40 | 0.97 | 1.36 | 0.16 | 0.96 | 0.81 | 0.40 | 1.00 |
| GC | A1a | Beta-Tocopherol | 0.92 | 0.34 | 0.97 | 1.26 | < 0.05* | 0.57 | 0.97 | 0.80 | 1.00 |
| GC | A1a | N-pyridin-2-ylnitramide | 1.16 | 0.29 | 0.97 | 1.06 | 0.66 | 0.98 | 0.91 | 0.62 | 1.00 |
| GC | A1a | 2,2',3,4,4',5',6-Heptachlorobiphenyl | 0.82 | 0.43 | 0.97 | 1.25 | 0.40 | 0.98 | 1.03 | 0.94 | 1.00 |
| GC | A1a | Triethyl citrate | 0.66 | 0.08 | 0.97 | 0.94 | 0.79 | 0.98 | 1.23 | 0.48 | 1.00 |
| GC | A1a | (3R,8S,9S,10R,13S,14S,17S)-17-[(1S)-1-(dimethylamino)ethyl]-N-ethyl-N,10,13-trimethyl-2,3,4,7,8,9,11,12,14,15,16,17-dodecahydro-1H-cyclopenta[a]phenanthren-3-amine | 0.84 | 0.66 | 0.98 | 0.85 | 0.69 | 0.98 | 0.62 | 0.35 | 1.00 |
| GC | A1a | (2E,7E)-5-hydroxy-6,6,9-trimethyl-4-deca-2,7,9-trienone | 0.85 | 0.54 | 0.98 | 1.13 | 0.66 | 0.98 | 1.72 | 0.13 | 1.00 |
| GC | A1a | 2-Methyl-3-(trifluoromethyl)-1,4-naphthoquinone | 0.58 | 0.16 | 0.97 | 1.60 | 0.15 | 0.96 | 0.53 | 0.15 | 1.00 |
| GC | A1a | 1,3,5-Trimethyladamantane | 0.79 | 0.37 | 0.97 | 1.23 | 0.44 | 0.98 | 0.95 | 0.86 | 1.00 |
| GC | A1a | (1S)-1-[(3S,5S,8R,9S,10S,13S,14S,17S)-10,13-dimethyl-3-(propan-2-ylideneamino)-2,3,4,5,6,7,8,9,11,12,14,15,16,17-tetradecahydro-1H-cyclopenta[a]phenanthren-17-yl]-N,N-dimethylethanamine | 0.76 | 0.18 | 0.97 | 1.25 | 0.26 | 0.96 | 0.72 | 0.22 | 1.00 |
| GC | A1a | 2-Phenylethyl propionate | 0.95 | 0.66 | 0.98 | 0.95 | 0.66 | 0.98 | 0.86 | 0.32 | 1.00 |
| GC | A1a | (trans)-2-(Hydroxymethyl)-3-phenyltetrahydrofuran | 0.80 | 0.33 | 0.97 | 0.90 | 0.61 | 0.98 | 1.35 | 0.26 | 1.00 |
| GC | A1a | PCB 130 | 0.90 | 0.48 | 0.98 | 1.05 | 0.73 | 0.98 | 1.13 | 0.50 | 1.00 |
| GC | A1a | N,N-dimethyl-4-phenylbutanamide | 1.06 | 0.76 | 0.98 | 1.27 | 0.23 | 0.96 | 0.84 | 0.46 | 1.00 |
| GC | A1a | 2',6'-Dichloro-2,4-dimethylbenzanilide | 1.30 | 0.26 | 0.97 | 1.29 | 0.31 | 0.96 | 0.97 | 0.92 | 1.00 |
| GC | A1a | 4-(2-thenoyl)tetrahydrofuran-2-one | 1.86 | 0.44 | 0.97 | 4.27 | 0.10 | 0.92 | 0.23 | 0.13 | 1.00 |
| GC | A1a | 4a-Methyl-4,4a,5,6-tetrahydro-2(3H),7-indendione | 1.02 | 0.93 | 1.00 | 0.94 | 0.82 | 0.98 | 0.97 | 0.94 | 1.00 |
| GC | A1a | 1-methylquinolin-2(1H)-one | 1.62 | 0.36 | 0.97 | 0.85 | 0.75 | 0.98 | 1.12 | 0.87 | 1.00 |
| GC | A1a | (+)-Pinidine | 1.02 | 0.93 | 1.00 | 1.25 | 0.35 | 0.96 | 1.28 | 0.43 | 1.00 |
| GC | A1a | (3R*,4S*)-3-Ethyl-4-(trimethylsilyethynyl)azitidin-2-one | 0.98 | 0.95 | 1.00 | 2.12 | < 0.05* | 0.57 | 0.77 | 0.42 | 1.00 |
| GC | A1a | 3-Methoxyphthalide | 1.33 | 0.29 | 0.97 | 1.52 | 0.16 | 0.96 | 0.84 | 0.63 | 1.00 |
| GC | A1a | methyloctyldiethoxysilane | 0.99 | 0.96 | 1.00 | 1.08 | 0.77 | 0.98 | 0.62 | 0.15 | 1.00 |
| GC | A1a | 1-Amino-3,4-dihydro-3,4-dimethyl-2-naphthalenecarbonitrile | 0.89 | 0.37 | 0.97 | 1.11 | 0.41 | 0.98 | 1.19 | 0.26 | 1.00 |
| GC | A1a | 2-Naphthalenamine | 1.32 | 0.27 | 0.97 | 0.81 | 0.44 | 0.98 | 0.90 | 0.74 | 1.00 |
| GC | A1a | 1,3-trans-4-Hydroxymethyl-1-methylcyclopentane-1,3-diol | 1.39 | 0.15 | 0.97 | 0.84 | 0.51 | 0.98 | 2.12 | < 0.05* | 1.00 |
| GC | A1a | 1-methoxy-4-[(E)-2-phenylethenyl]benzene | 0.99 | 0.82 | 1.00 | 1.03 | 0.64 | 0.98 | 1.02 | 0.79 | 1.00 |
| GC | A1a | 2-methyl-1,3-benzoxazole | 0.97 | 0.84 | 1.00 | 0.95 | 0.76 | 0.98 | 0.88 | 0.51 | 1.00 |
| GC | A1a | Diisobutyl phthalate | 0.78 | 0.22 | 0.97 | 1.05 | 0.83 | 0.98 | 1.11 | 0.72 | 1.00 |
| GC | A1a | 8-methylpyrrolo[1,2-a]quinoxaline | 1.11 | 0.74 | 0.98 | 0.90 | 0.77 | 0.98 | 0.89 | 0.80 | 1.00 |
| GC | A1a | 1-(2,4-Dimethylphenyl)but-3-en-1-ol | 1.34 | 0.57 | 0.98 | 1.77 | 0.24 | 0.96 | 0.63 | 0.50 | 1.00 |
| GC | A1a | 1,4,5,8-Tetramethylnaphthalene | 1.30 | 0.37 | 0.97 | 1.06 | 0.85 | 0.98 | 1.18 | 0.69 | 1.00 |
| GC | A1a | N-tert-Butylmaleimide | 0.84 | 0.31 | 0.97 | 1.19 | 0.27 | 0.96 | 1.04 | 0.88 | 1.00 |
| GC | A1a | 1-Piperidino-5-(2-hydroxy-4-nitrophenoxy)pentane | 0.57 | 0.11 | 0.97 | 0.92 | 0.80 | 0.98 | 0.51 | 0.15 | 1.00 |
| GC | A1a | Diisopropyl phthalate | 0.98 | 0.79 | 0.99 | 1.06 | 0.49 | 0.98 | 1.03 | 0.81 | 1.00 |
| GC | A1a | Kanzonol P | 0.92 | 0.73 | 0.98 | 1.32 | 0.27 | 0.96 | 0.76 | 0.40 | 1.00 |
| GC | A1a | Hexachlorobenzene | 0.76 | 0.10 | 0.97 | 0.93 | 0.66 | 0.98 | 0.99 | 0.97 | 1.00 |
| GC | A1a | 5-Vinyloxyhept-6-enyl 2,2-dimethylpropanoate | 0.93 | 0.75 | 0.98 | 0.98 | 0.94 | 0.99 | 0.94 | 0.86 | 1.00 |
| GC | A1a | 1-Piperidino-5-(4-amino-2-methoxyphenoxy)ethane | 0.69 | < 0.05* | 0.97 | 0.96 | 0.84 | 0.98 | 0.96 | 0.84 | 1.00 |
| GC | A1a | 1-chloro-4-[2,2,2-trichloro-1-(4-chlorophenyl)ethyl]benzene | 1.03 | 0.92 | 1.00 | 1.14 | 0.64 | 0.98 | 1.06 | 0.84 | 1.00 |
| GC | A1a | 3-(4-ketohexyl)cyclohex-2-en-1-one | 1.01 | 0.99 | 1.00 | 2.37 | 0.26 | 0.96 | 0.60 | 0.49 | 1.00 |
| GC | A1a | Tris(2-butoxyethyl) phosphate | 1.17 | 0.43 | 0.97 | 1.29 | 0.15 | 0.96 | 1.01 | 0.98 | 1.00 |
| GC | A1a | 2-(4-Methoxyphenoxy)benzothiophene | 0.74 | 0.44 | 0.97 | 1.03 | 0.93 | 0.99 | 1.15 | 0.81 | 1.00 |
| GC | A1a | Phenanthrene | 0.94 | 0.94 | 1.00 | 3.35 | 0.20 | 0.96 | 0.27 | 0.29 | 1.00 |
| GC | A1a | methyl 2-(5-methoxy-1,2-dimethylindol-3-yl)acetate | 0.98 | 0.92 | 1.00 | 1.06 | 0.78 | 0.98 | 0.94 | 0.82 | 1.00 |
| GC | A1a | 2'-Hydroxy-4',6'-dimethoxyacetophenone | 0.95 | 0.89 | 1.00 | 0.55 | 0.11 | 0.92 | 0.68 | 0.43 | 1.00 |
| GC | A1a | 2,2',3,4,4',5,5'-Heptachlorobiphenyl | 0.96 | 0.72 | 0.98 | 1.02 | 0.87 | 0.99 | 1.10 | 0.44 | 1.00 |
| GC | A1a | 3-Hexene-2,5-dione, 3-acetyl-, 5-(dimethylhydrazone) | 0.76 | 0.34 | 0.97 | 0.94 | 0.85 | 0.98 | 0.81 | 0.58 | 1.00 |
| GC | A1a | Captafol | 0.83 | 0.37 | 0.97 | 1.07 | 0.71 | 0.98 | 0.75 | 0.28 | 1.00 |
| GC | A1a | N-(2-Fluorobenzylidene)-2-fluorophenylmethylamine | 0.76 | 0.19 | 0.97 | 1.19 | 0.39 | 0.98 | 0.87 | 0.61 | 1.00 |
| GC | A1a | (2-ethyl-4,6-dimethylphenyl)methanol | 1.27 | 0.65 | 0.98 | 1.83 | 0.30 | 0.96 | 1.18 | 0.83 | 1.00 |
| GC | A1a | 2,2-Dimethyl-2H-chromene-6-carbaldehyde | 1.19 | 0.41 | 0.97 | 0.89 | 0.60 | 0.98 | 0.83 | 0.47 | 1.00 |
| GC | A1a | 3,3-Dimethyl-1,3-dihydro-1-isobenzofuranone | 1.93 | 0.25 | 0.97 | 1.65 | 0.43 | 0.98 | 1.33 | 0.74 | 1.00 |
| GC | A1a | 3-Butenyl-5-propylindolizidine | 0.79 | 0.23 | 0.97 | 1.30 | 0.28 | 0.96 | 1.18 | 0.57 | 1.00 |
| GC | A1a | Di-n-propylphthalate | 0.71 | 0.11 | 0.97 | 0.91 | 0.65 | 0.98 | 0.99 | 0.98 | 1.00 |
| GC | A1a | 3,4-Diphenyl-6-methyl-7,8-(1,4-dimethoxybenzo)-9-oxatricyclo[4.2.1.0(2,5)]non-3-ene | 1.18 | 0.22 | 0.97 | 1.47 | < 0.05* | 0.57 | 1.04 | 0.85 | 1.00 |
| GC | A1a | 5-bicyclo[2.2.2]oct-2-enecarbonitrile | 0.75 | 0.40 | 0.97 | 1.88 | 0.11 | 0.92 | 2.48 | 0.06 | 1.00 |
| GC | A1a | 1,6-dimethyl-4-propan-2-ylnaphthalene | 0.63 | 0.11 | 0.97 | 0.92 | 0.78 | 0.98 | 0.94 | 0.85 | 1.00 |
| GC | A1a | 1,2-Dicyano-4,5-di[(N,N-dimethylaminocarbonyl)methoxy]benzene | 0.73 | 0.17 | 0.97 | 1.20 | 0.42 | 0.98 | 0.82 | 0.53 | 1.00 |
| GC | A1a | 4-Hydroxyacetophenone | 0.82 | 0.20 | 0.97 | 1.06 | 0.70 | 0.98 | 1.40 | 0.11 | 1.00 |
| GC | A1a | 2-(2,3-dihydro-1H-indol-4-yl)ethanol | 1.02 | 0.86 | 1.00 | 1.00 | 1.00 | 1.00 | 0.96 | 0.77 | 1.00 |
| GC | A1a | (Z)-2-((2-Methylthio)styrylthio)aniline | 0.99 | 0.94 | 1.00 | 0.91 | 0.51 | 0.98 | 0.98 | 0.94 | 1.00 |
| GC | A1a | Carbaryl | 1.05 | 0.91 | 1.00 | 0.53 | 0.12 | 0.92 | 2.06 | 0.20 | 1.00 |
| GC | A1a | 4-Butylaniline | 0.86 | 0.56 | 0.98 | 1.09 | 0.75 | 0.98 | 0.76 | 0.42 | 1.00 |
| GC | A1a | Dibenzofuran | 0.89 | 0.74 | 0.98 | 1.05 | 0.90 | 0.99 | 0.85 | 0.73 | 1.00 |
| GC | A1a | 3-(2-methylbutyl)-1H-indole | 0.60 | 0.13 | 0.97 | 0.97 | 0.93 | 0.99 | 1.06 | 0.91 | 1.00 |
| GC | A1a | (2s)-n-tert-butoxycarbonyl-2-(4-(1-ethoxyethoxy)-1-hydroxybutyl)pyrrolidine | 0.79 | 0.20 | 0.97 | 1.18 | 0.37 | 0.97 | 0.67 | 0.11 | 1.00 |
| GC | A1a | (2RS,4SR)-4-(dimethylamino)-2,4-diphenyl-2-butanol | 1.10 | 0.66 | 0.98 | 1.26 | 0.33 | 0.96 | 0.66 | 0.12 | 1.00 |
| GC | A1a | 2-ethynylbenzaldehyde | 0.87 | 0.62 | 0.98 | 1.45 | 0.19 | 0.96 | 0.91 | 0.79 | 1.00 |
| GC | A1a | 1-Ethyl-4,5,8-trimethylnaphthalene | 1.19 | 0.10 | 0.97 | 0.77 | < 0.05* | 0.57 | 1.17 | 0.23 | 1.00 |
| GC | A1a | Bisphenol A | 0.90 | 0.49 | 0.98 | 1.08 | 0.60 | 0.98 | 1.04 | 0.83 | 1.00 |
| GC | A1a | N-(1,2-diphenyl-2-phenylsulfanylethyl)acetamide | 1.32 | 0.43 | 0.97 | 1.22 | 0.59 | 0.98 | 0.44 | 0.10 | 1.00 |
| GC | A1a | Dehydro-botrydienol | 0.87 | 0.46 | 0.97 | 1.34 | 0.13 | 0.96 | 0.74 | 0.21 | 1.00 |
| GC | A1a | 2-methyl-2-[(E)-2-methyl-3-phenylprop-2-enyl]-1,3-dioxolane | 1.13 | 0.41 | 0.97 | 0.91 | 0.59 | 0.98 | 0.83 | 0.36 | 1.00 |
| GC | A1a | 5,5-Dicyano-2-methyl-3,4-dihydro-pyrrole | 0.99 | 0.97 | 1.00 | 1.11 | 0.69 | 0.98 | 1.00 | 1.00 | 1.00 |
| GC | A1a | Tetracyclo[6.2.1.1(3,6)]dodecane-2,7-dione | 0.77 | 0.10 | 0.97 | 1.12 | 0.47 | 0.98 | 1.14 | 0.52 | 1.00 |
| GC | A1a | 1-(2-piperidin-1-ylethyl)cyclohexan-1-ol | 0.65 | 0.35 | 0.97 | 3.28 | < 0.05* | 0.57 | 0.61 | 0.36 | 1.00 |
| GC | A1a | 5-Phenylporphyrin | 0.98 | 0.92 | 1.00 | 0.61 | < 0.05* | 0.67 | 1.28 | 0.34 | 1.00 |
| GC | A1a | 2-(2-Butoxyethyl)pyridine | 0.81 | 0.37 | 0.97 | 0.72 | 0.31 | 0.96 | 1.04 | 0.91 | 1.00 |
| GC | A1a | 1,2,3,3,4,4,5,5-Octamethylcyclopentene | 0.62 | 0.20 | 0.97 | 1.68 | 0.14 | 0.96 | 0.31 | < 0.05* | 1.00 |
| GC | A1a | 4-(2'-Hydroxyphenyl)non-1-ene | 0.92 | 0.70 | 0.98 | 0.74 | 0.16 | 0.96 | 0.90 | 0.73 | 1.00 |
| GC | A1a | N,N-Dimethyl-2,2-bis( 9'-anthracenyl)acetamide | 0.61 | 0.07 | 0.97 | 1.21 | 0.53 | 0.98 | 0.88 | 0.71 | 1.00 |
| GC | A1a | methyl (2S,3S)-4-(4-methoxyphenyl)-3-methyl-2-[(2-methylpropan-2-yl)oxycarbonylamino]-4-oxobutanoate | 1.10 | 0.67 | 0.98 | 1.52 | 0.10 | 0.92 | 0.64 | 0.12 | 1.00 |
| GC | A1a | ethyl 2-methylidene-4-(2,4,6-trimethylphenyl)butanoate | 1.22 | 0.26 | 0.97 | 1.14 | 0.50 | 0.98 | 1.00 | 0.98 | 1.00 |
| GC | A1a | (2S,6R)-4-[3-(4-tert-butylphenyl)-2-methylpropyl]-2,6-dimethylmorpholine | 1.17 | 0.30 | 0.97 | 1.05 | 0.74 | 0.98 | 0.83 | 0.32 | 1.00 |
| GC | A1a | 2,2',4,4',5,5'-Hexachlorobiphenyl | 0.90 | 0.44 | 0.97 | 1.03 | 0.83 | 0.98 | 1.08 | 0.64 | 1.00 |
| GC | A1a | (2E)-1-O-Acetyl-3,6-dimethyl-6-formyl-2-hepten-1-ol | 0.77 | 0.46 | 0.97 | 1.26 | 0.49 | 0.98 | 1.41 | 0.45 | 1.00 |
| GC | A1a | 2-(1-bromoethenyl)-5-methoxy-1,3-dimethylbenzene | 1.09 | 0.73 | 0.98 | 0.86 | 0.54 | 0.98 | 1.24 | 0.50 | 1.00 |
| GC | A1a | (1S,3R,4R)-2-[(S)-1-Phenylethylamino]-2-azabicyclo[2.2.1]heptane-3(S)-phenylmethanol | 0.94 | 0.59 | 0.98 | 0.81 | 0.16 | 0.96 | 1.03 | 0.87 | 1.00 |
| GC | A1a | N-(n-Butyl)-3-cyclohexenylpropiolamide | 1.08 | 0.86 | 1.00 | 1.65 | 0.31 | 0.96 | 0.34 | 0.06 | 1.00 |
| GC | A1a | 4-tert-Octylphenol | 0.99 | 0.92 | 1.00 | 1.10 | 0.51 | 0.98 | 0.77 | 0.18 | 1.00 |
| GC | A1a | 2-phenylpropanenitrile | 0.95 | 0.85 | 1.00 | 1.40 | 0.17 | 0.96 | 1.04 | 0.90 | 1.00 |
| GC | A1a | (-)-(4a'R,8'S,8a'S,10a'S)-1',1',4a',8a'-Tetramethyl-3',4',4a',6',7',8',8a',9',10',10a'-decahydro-1'H-spiro[[1,3]dioxolane-2,2'-phenanthren]-8'-ol | 0.80 | 0.29 | 0.97 | 1.31 | 0.18 | 0.96 | 0.76 | 0.27 | 1.00 |
| GC | A1a | Benzoic acid, 2,4-dihydroxy-3-[[4-methoxy-2-(phenylmethoxy)-6-propylbenzoyl]oxy]-6-propyl-, methyl ester | 0.53 | < 0.05* | 0.97 | 1.17 | 0.53 | 0.98 | 1.89 | 0.09 | 1.00 |
| GC | A1a | dimethyl 5-(1H-indol-3-yl)-2-methyl-3-methylsulfanyl-1,2,4-triazine-5,6-dicarboxylate | 0.94 | 0.77 | 0.99 | 1.01 | 0.96 | 0.99 | 0.88 | 0.58 | 1.00 |
| GC | A1a | 1,1-Dimethyl-1,3-dihydro-isobenzofuran-5-ol | 0.98 | 0.90 | 1.00 | 1.04 | 0.85 | 0.98 | 1.01 | 0.98 | 1.00 |
| GC | A1a | Methyl 1,3-dihydro-2H-isobenzofuran-4-carboxylate | 0.61 | 0.24 | 0.97 | 1.62 | 0.32 | 0.96 | 0.77 | 0.65 | 1.00 |
| GC | A1a | Diethyl Phthalate | 1.15 | 0.58 | 0.98 | 0.92 | 0.72 | 0.98 | 1.04 | 0.89 | 1.00 |
| GC | A1a | ethyl 2-hydroxy-2-(4-methylphenyl)-2-(5-oxocyclopenten-1-yl)acetate | 0.87 | 0.38 | 0.97 | 0.99 | 0.95 | 0.99 | 0.72 | 0.10 | 1.00 |
| GC | A1a | Methoxyethyl-3-(4-oxo-4H-1-benzopyran-3-yl)-acrylate | 1.54 | 0.48 | 0.98 | 1.44 | 0.52 | 0.98 | 0.44 | 0.27 | 1.00 |
| GC | A1a | Octahydro-5-pentylindolizine-8-methanol | 1.33 | 0.15 | 0.97 | 1.04 | 0.85 | 0.98 | 1.06 | 0.80 | 1.00 |
| GC | A1a | 1-prop-1-ynyl-1-cyclohex-2-enol | 1.31 | 0.37 | 0.97 | 0.95 | 0.85 | 0.98 | 0.75 | 0.47 | 1.00 |
| GC | A1a | .alpha.-Chloroformyl-4-ethoxyphenylhydrazine hydrochloride | 0.99 | 0.97 | 1.00 | 1.17 | 0.36 | 0.97 | 0.93 | 0.75 | 1.00 |
| GC | A1a | 2-Ethyl-N-methyl-1-phenyl-3-butenylamine | 0.79 | 0.66 | 0.98 | 1.60 | 0.46 | 0.98 | 2.52 | 0.27 | 1.00 |
| GC | A1a | 3-Methoxy-3-oxopropane-1,2-diyl dibutyrate | 0.86 | 0.58 | 0.98 | 0.80 | 0.48 | 0.98 | 0.42 | < 0.05* | 1.00 |
| GC | A1a | DEET | 0.73 | 0.43 | 0.97 | 0.67 | 0.27 | 0.96 | 0.59 | 0.28 | 1.00 |
| GC | A1a | 3-(p-Methoxyphenyl)-5,6-dimethylindenone | 1.26 | 0.80 | 1.00 | 2.04 | 0.45 | 0.98 | 1.17 | 0.89 | 1.00 |
| GC | A1a | [2-(4-ethylbenzoyl)oxyphenyl] 2,6-difluorobenzoate | 0.99 | 0.97 | 1.00 | 0.49 | 0.08 | 0.92 | 1.58 | 0.39 | 1.00 |
| GC | A1a | 4-Ethylbenzoic acid, 3-chlorophenyl ester | 0.88 | 0.69 | 0.98 | 1.41 | 0.50 | 0.98 | 0.86 | 0.72 | 1.00 |
| GC | A1a | 5,6,8-Trimethoxy-7-methyl-2,3-diphenylquinoxaline | 0.99 | 0.98 | 1.00 | 0.66 | 0.06 | 0.86 | 1.49 | 0.15 | 1.00 |
| GC | A1a | Methfuroxam | 0.91 | 0.79 | 0.99 | 1.06 | 0.85 | 0.98 | 0.60 | 0.22 | 1.00 |
| GC | A1a | 2-(3,5-dichloro-2-methoxy-phenyl)-1H-pyrrole | 2.29 | 0.29 | 0.97 | 0.94 | 0.94 | 0.99 | 0.68 | 0.69 | 1.00 |
| GC | A1a | p-Tolyl Furan-2-Carboxylate | 1.27 | 0.61 | 0.98 | 2.10 | 0.15 | 0.96 | 0.39 | 0.14 | 1.00 |
| GC | A1a | 2-Methyl-2-(2'-propenyl)-cyclohexane-1,3-dione | 0.75 | 0.25 | 0.97 | 0.88 | 0.59 | 0.98 | 0.81 | 0.51 | 1.00 |
| GC | A1a | 4,7-Dihydroxy-5-methylcoumarin | 0.74 | < 0.05* | 0.97 | 1.43 | < 0.05* | 0.57 | 0.98 | 0.92 | 1.00 |
| GC | A1a | (2S)-2-amino-2-(1,3-benzodioxol-5-yl)acetic acid methyl ester | 0.97 | 0.67 | 0.98 | 1.04 | 0.65 | 0.98 | 0.92 | 0.38 | 1.00 |
| GC | A1a | [(1R,2S,4aR,6R,8aR)-2,6-dimethyl-1,2,4a,5,6,7,8,8a-octahydronaphthalen-1-yl]methanol | 0.78 | 0.16 | 0.97 | 1.32 | 0.14 | 0.96 | 1.00 | 0.99 | 1.00 |
| GC | A1a | (4-methylphenyl) 2-hydroxybenzoate | 0.97 | 0.95 | 1.00 | 0.97 | 0.95 | 0.99 | 1.24 | 0.71 | 1.00 |
| GC | A1a | (1R,2S,6R)-2-Acetoxy-9-(hex-2-en-2-yl)bicyclo[4.3.0]nona-3,7-diene | 0.93 | 0.69 | 0.98 | 1.14 | 0.48 | 0.98 | 1.33 | 0.25 | 1.00 |
| GC | A1a | Methyl phenidate | 0.46 | < 0.05* | 0.97 | 1.08 | 0.76 | 0.98 | 0.77 | 0.39 | 1.00 |
| GC | A1a | 2-[(Phenylsulfanyl)methyl]oxirane | 0.84 | 0.74 | 0.98 | 1.46 | 0.45 | 0.98 | 0.64 | 0.50 | 1.00 |
| GC | A1a | endo-5,6-Di(acetyl)-7-acetoxymethylenebicyclo[2.2.1]hept-2-ene | 0.99 | 0.97 | 1.00 | 1.04 | 0.90 | 0.99 | 1.73 | 0.21 | 1.00 |
| GC | A1a | (E)-[(4S)-4-allyl-2,2-dimethyl-1,3-dioxan-5-ylidene]-[(2S)-2-(methoxymethyl)pyrrolidino]amine | 1.02 | 0.94 | 1.00 | 1.18 | 0.48 | 0.98 | 0.77 | 0.24 | 1.00 |
| GC | A1a | 3-(piperidin-1-ylmethyl)-2,3-dihydro-1,4-benzodioxin-6-yl Methanesulfonate | 1.02 | 0.93 | 1.00 | 1.48 | 0.13 | 0.94 | 0.72 | 0.35 | 1.00 |
| GC | A1a | 2,4-di-tert-butylphenol | 1.38 | 0.49 | 0.98 | 1.56 | 0.35 | 0.96 | 0.47 | 0.21 | 1.00 |
| GC | A1a | Cyclopropyl 2-aminobenzoate | 1.10 | 0.78 | 0.99 | 1.36 | 0.46 | 0.98 | 1.07 | 0.89 | 1.00 |
| GC | A1a | Methoxy-colorenone | 0.66 | 0.14 | 0.97 | 1.17 | 0.58 | 0.98 | 0.99 | 0.98 | 1.00 |
| GC | A1a | Methyl 1-(phenylsulfonyl)-3-(methoxycarbonyl)indole-2-acetate | 0.46 | < 0.05* | 0.97 | 1.09 | 0.76 | 0.98 | 0.63 | 0.23 | 1.00 |
| GC | A1a | (E)-1-cyclohexyl-2-hepten-1-one | 0.76 | 0.21 | 0.97 | 1.13 | 0.61 | 0.98 | 1.06 | 0.84 | 1.00 |
| GC | A1a | 3-(ethylamino)phenol | 0.99 | 0.96 | 1.00 | 1.07 | 0.77 | 0.98 | 1.20 | 0.50 | 1.00 |
| GC | A1a | N-ethyl-N-[(7-fluoro-2,3-dihydro-1,4-benzodioxin-2-yl)methyl]propan-1-amine | 0.88 | 0.75 | 0.98 | 1.44 | 0.37 | 0.97 | 1.03 | 0.95 | 1.00 |
| GC | A1a | 3-methylcyclohepta[b]furan-2-one | 0.97 | 0.93 | 1.00 | 1.12 | 0.80 | 0.98 | 1.69 | 0.38 | 1.00 |
| GC | A1a | methyl (2R,3R,3aR)-2-(2-methylpropanoyl)-2,3,3a,4,5,6-hexahydropyrrolo[1,2-b][1,2]oxazole-3-carboxylate | 0.61 | 0.13 | 0.97 | 1.32 | 0.37 | 0.97 | 0.91 | 0.81 | 1.00 |
| GC | A1a | N-[(7-fluoro-2,3-dihydro-1,4-benzodioxin-2-yl)methyl]-N-methylethanamine | 0.88 | 0.65 | 0.98 | 1.30 | 0.30 | 0.96 | 0.56 | 0.19 | 1.00 |
| GC | A1a | 6,9-Dimethoxy-5,5-dimethyl-3,3a,5,9b-tetrahydro-furo[3,2-c]isochromen-2-one | 0.84 | 0.32 | 0.97 | 1.08 | 0.68 | 0.98 | 0.96 | 0.88 | 1.00 |
| GC | A1a | 3-hydroxypropyl benzoate | 0.70 | 0.37 | 0.97 | 1.18 | 0.66 | 0.98 | 1.49 | 0.33 | 1.00 |
| GC | A1a | N-(2-Chloroethyl)-3-methoxybenzamide | 0.87 | 0.65 | 0.98 | 1.12 | 0.71 | 0.98 | 1.04 | 0.92 | 1.00 |
| GC | A1a | Octyl decyl phthalate | 0.95 | 0.77 | 0.99 | 1.07 | 0.68 | 0.98 | 0.78 | 0.29 | 1.00 |
| GC | A1a | [(2S)-2-[(1R,3aR,4S,7aR)-4-hydroxy-7a-methyl-1,2,3,3a,4,5,6,7-octahydroinden-1-yl]propyl] 4-methylbenzenesulfonate | 0.55 | 0.40 | 0.97 | 3.00 | 0.12 | 0.92 | 0.72 | 0.70 | 1.00 |
| GC | A1a | 4-methoxy-1-[methoxy(phenyl)methyl]-2-nitro-benzene | 0.81 | 0.23 | 0.97 | 1.19 | 0.37 | 0.97 | 0.87 | 0.53 | 1.00 |
| GC | A1a | Ibuprofen | 0.93 | 0.62 | 0.98 | 0.94 | 0.61 | 0.98 | 1.44 | 0.22 | 1.00 |
| GC | A1a | (+-)-5-[2-(7-Methoxyindol-3-yl)ethyl]-2,3,3a.beta.,6,7,7a.beta.-hexahydrofuro[3,2-c]pyridin-4(5H)-one | 0.87 | 0.63 | 0.98 | 1.10 | 0.75 | 0.98 | 1.39 | 0.34 | 1.00 |
| GC | A1a | 2,4,4'-Trichlorobiphenyl | 1.03 | 0.91 | 1.00 | 1.24 | 0.53 | 0.98 | 0.63 | 0.43 | 1.00 |
| GC | A1a | (E)-1-phenyl-4-hexen-1-one | 1.62 | 0.34 | 0.97 | 1.14 | 0.81 | 0.98 | 0.36 | 0.20 | 1.00 |
| GC | A1a | 2,2-Dimethyl-5-(prop-2'-ynyl)-1,3-dioxane-4,6-dione | 0.75 | 0.35 | 0.97 | 0.98 | 0.94 | 0.99 | 1.97 | 0.10 | 1.00 |
| GC | A1a | 2,5-dihydroxy-4-prop-2-enylbenzaldehyde | 0.97 | 0.91 | 1.00 | 0.83 | 0.47 | 0.98 | 1.28 | 0.40 | 1.00 |
| GC | A1a | 2(S)-Amino-3-methyl-N,N-dioctylbutyramide | 0.81 | 0.40 | 0.97 | 1.09 | 0.73 | 0.98 | 0.78 | 0.43 | 1.00 |
| GC | A1a | Ethyl (3-Methyl-6-oxo-1-phenyl-4,5,6,7-tetrahydro-1H-pyrazolo[3,4-b]pyridine-4-carboxamido)benzoate | 0.77 | 0.40 | 0.97 | 1.42 | 0.31 | 0.96 | 0.96 | 0.93 | 1.00 |
| GC | A1a | 3-(Chloromethyl)-4-hydroxy-4-phenyl-2-butanone | 0.92 | 0.65 | 0.98 | 0.92 | 0.65 | 0.98 | 0.98 | 0.94 | 1.00 |
| GC | A1a | 3-(Dimethylphenylsilyl)-2-methylpropionamide | 0.91 | 0.72 | 0.98 | 0.80 | 0.40 | 0.98 | 0.69 | 0.27 | 1.00 |
| GC | A1a | 4-tert-butyl-4-(1-methoxy-2-methyl-propan-2-yl)-1-methyl-azetidine-2,3-dione | 0.81 | 0.07 | 0.97 | 0.96 | 0.77 | 0.98 | 0.92 | 0.60 | 1.00 |
| GC | A1a | (5S,8S)-8-Isopropyl-3-methoxy-5-methyl-1,2,3,4-tetrahydronaphthalen-2-carbaldehyde | 2.29 | < 0.05* | 0.97 | 1.39 | 0.46 | 0.98 | 1.16 | 0.77 | 1.00 |
| GC | A1a | 5-p-anisyloxypentan-1-ol | 1.36 | 0.38 | 0.97 | 1.93 | 0.10 | 0.92 | 1.16 | 0.73 | 1.00 |
| GC | A1a | (2E,4E)-3-Phenylthiopenta-2,4-dienal | 0.62 | 0.13 | 0.97 | 1.50 | 0.18 | 0.96 | 0.51 | 0.12 | 1.00 |
| GC | A1a | (1R,2R)-1,2-diphenylethane-1,2-diol | 0.99 | 0.97 | 1.00 | 0.96 | 0.92 | 0.99 | 0.39 | 0.06 | 1.00 |
| GC | A1a | 4-Methylbenzyl vinyl ether | 1.15 | 0.51 | 0.98 | 1.17 | 0.39 | 0.98 | 0.73 | 0.22 | 1.00 |
| GC | A1a | 1-O-hexyl 2-O-propan-2-yl benzene-1,2-dicarboxylate | 0.99 | 0.89 | 1.00 | 1.11 | 0.30 | 0.96 | 1.03 | 0.84 | 1.00 |
| GC | A1a | methyl 2-methylbenzimidazole-1-carboxylate | 0.87 | 0.46 | 0.97 | 0.88 | 0.51 | 0.98 | 0.84 | 0.48 | 1.00 |
| GC | A1a | 3-Piperidin-1-yl-butan-2-ol | 0.94 | 0.56 | 0.98 | 1.14 | 0.22 | 0.96 | 0.79 | 0.08 | 1.00 |
| GC | A1a | (-)-(R,E)-5-(tert-Butylsulfonyl)hept-3-ene | 1.00 | 0.99 | 1.00 | 1.32 | 0.41 | 0.98 | 0.62 | 0.27 | 1.00 |
| GC | A1a | (Z)-4-Methylphenylazo tert-butyl sulfide | 1.87 | 0.16 | 0.97 | 0.83 | 0.61 | 0.98 | 3.36 | < 0.05* | 1.00 |
| GC | A1a | 10-Formyl-1,2,3,4,5,6-hexahydro-1,5-imino-9-methoxy-3,8,11-trimethyl-4-oxo-3-benzazocine | 0.74 | 0.17 | 0.97 | 1.25 | 0.31 | 0.96 | 0.78 | 0.37 | 1.00 |
| GC | A1a | 1-hydroxy-3-methyl-2-pyrrolecarbonitrile | 1.43 | 0.26 | 0.97 | 1.16 | 0.67 | 0.98 | 1.58 | 0.27 | 1.00 |
| GC | A1a | (E)-2,2,5-triphenyl-4-pentenenitrile | 0.46 | 0.11 | 0.97 | 1.11 | 0.84 | 0.98 | 1.43 | 0.52 | 1.00 |
| GC | A1a | 4-Pentylphenol | 0.76 | 0.44 | 0.97 | 2.48 | 0.05 | 0.82 | 1.36 | 0.52 | 1.00 |
| GC | A1a | 2-[(E)-hydroxyiminomethyl]phenol | 0.86 | 0.77 | 0.99 | 1.56 | 0.40 | 0.98 | 1.39 | 0.58 | 1.00 |
| GC | A1a | Bis(2-ethylhexyl) adipate | 0.60 | 0.19 | 0.97 | 0.76 | 0.49 | 0.98 | 1.85 | 0.28 | 1.00 |
| GC | A1a | 4'-[(Methanesulfonyloxy)methyl]-[2,2' ; 6',6\]-terpyridyl"" | 0.74 | 0.34 | 0.97 | 1.67 | 0.11 | 0.92 | 0.81 | 0.63 | 1.00 |
| GC | A1a | (2S,3R,4R)-4-tert-butyl-2-phenyl-3-oxolanol | 0.87 | 0.47 | 0.97 | 1.04 | 0.83 | 0.98 | 0.86 | 0.58 | 1.00 |
| GC | A1a | 2-(2-nitrophenyl)-1H-benzimidazole | 1.09 | 0.91 | 1.00 | 1.50 | 0.67 | 0.98 | 0.69 | 0.76 | 1.00 |
| GC | A1a | 2-N-[2'-(Methoxymethyl)pyrrolidin-1'-yl]amino-3-methylheptane | 1.06 | 0.45 | 0.97 | 1.03 | 0.68 | 0.98 | 1.03 | 0.79 | 1.00 |
| GC | A1a | 1H-3,1-benzoxazine-2,4-dione | 0.90 | 0.27 | 0.97 | 0.98 | 0.84 | 0.98 | 1.04 | 0.71 | 1.00 |
| GC | A1a | 8-Methyl-8-(hydroxymethyl)-4,5,7,8-tetrahydro-6H-cyclohepta[b]furan | 1.01 | 0.95 | 1.00 | 1.05 | 0.68 | 0.98 | 0.93 | 0.61 | 1.00 |
| GC | A1a | 2-methyl-1H-pyrrole-3-carbonitrile | 1.06 | 0.55 | 0.98 | 0.99 | 0.93 | 0.99 | 1.12 | 0.37 | 1.00 |
| GC | A1a | 5,16-androstadiene-3.beta.-ol-acetate | 1.10 | 0.06 | 0.97 | 0.97 | 0.60 | 0.98 | 0.96 | 0.58 | 1.00 |
| GC | A1a | [1,2,4]Triazolo[1,5-a][1,3,5]triazin-7-amine, 1,2-dihydro-2-methyl-2-phenyl- | 0.93 | 0.17 | 0.97 | 1.09 | 0.09 | 0.92 | 1.12 | 0.10 | 1.00 |
| GC | A1a | (2R,3S)-{3-[4-[(Tetrahydropyran-2-yl)oxy]but-2(E)-enyl]oxiranyl}methanol | 1.09 | 0.56 | 0.98 | 1.05 | 0.74 | 0.98 | 0.85 | 0.38 | 1.00 |
| GC | A1a | 3-cyclohexyl-5-phenyl-1,3-oxazolidine-2,4-dione | 1.01 | 0.80 | 0.99 | 1.06 | 0.38 | 0.97 | 0.96 | 0.56 | 1.00 |
| GC | A1a | 2-Hydroxy-3-methylbicyclo[2.2.1]heptane-2-methanol | 0.74 | 0.27 | 0.97 | 1.56 | 0.12 | 0.92 | 1.43 | 0.43 | 1.00 |
| GC | A1a | 16,17-Epoxy-6,7-isopropylidenedioxy-1,15-dihydroxy-8,15-secomaoerystals A | 1.04 | 0.60 | 0.98 | 0.91 | 0.23 | 0.96 | 0.97 | 0.75 | 1.00 |
| GC | A1a | 2,5-Dichlorophenol | 0.85 | 0.12 | 0.97 | 0.88 | 0.31 | 0.96 | 0.96 | 0.80 | 1.00 |
| GC | A1a | 2-methyl-1-acenaphthylenecarbonitrile | 1.11 | 0.09 | 0.97 | 0.93 | 0.30 | 0.96 | 1.09 | 0.28 | 1.00 |
| GC | A1a | 10,11-Dimethoxy-12b-trimethylsilyl-5,6,7,8,12b,13a-hexahydrodibenzo[c,g]oxireno[2,3-e]azecin-5-one | 1.15 | 0.20 | 0.97 | 1.05 | 0.68 | 0.98 | 1.06 | 0.72 | 1.00 |
| GC | A1a | 1-(4-Chlorophenyl)-2-octanone | 0.94 | 0.05 | 0.97 | 0.99 | 0.68 | 0.98 | 0.92 | 0.05 | 1.00 |
| GC | A1a | (-)-2.beta.-Methoxy-2a.beta.,3,4,6,7,8,8a.beta.,8b.beta.-Octahydro-6,6,8b.beta.-trimethyl-2H-naphtho[1,8-bc]furan | 1.02 | 0.91 | 1.00 | 1.17 | 0.45 | 0.98 | 0.88 | 0.57 | 1.00 |
| GC | A1a | Ethyl 2,4-dimethylpyrido[2,3-a]benzofuran-3-carboxylate | 1.20 | 0.14 | 0.97 | 1.17 | 0.15 | 0.96 | 0.99 | 0.93 | 1.00 |
| GC | A1a | [3'-Phenyl-2'-propenyl ] Allyl Ether | 1.10 | 0.53 | 0.98 | 0.92 | 0.56 | 0.98 | 1.05 | 0.79 | 1.00 |
| GC | A1a | l-Norleucine, N-methoxycarbonyl-, isohexyl ester | 1.02 | 0.90 | 1.00 | 0.92 | 0.61 | 0.98 | 0.93 | 0.69 | 1.00 |
| GC | A1a | 2,7-Dimethyl-10-n-propyl-4,5-dioxo-3,6,9-trioxa-3,4,5,6,9,10-hexahydroanthracene | 1.09 | 0.56 | 0.98 | 1.43 | < 0.05* | 0.76 | 0.95 | 0.80 | 1.00 |
| GC | A1a | Benzo[b]furo[3,4-f][1,4]benzodioxin-1,3-dione, 3a,4,11a,11b-tetrahydro-, (3a.alpha.,11a.alpha.,11b.alpha.)- | 0.94 | 0.36 | 0.97 | 0.96 | 0.53 | 0.98 | 1.24 | < 0.05* | 1.00 |
| GC | A1a | (1R*,2R*,5R*)-sec-butyl 5-hydroxy-2-methylcyclohexanecarboxylate | 0.94 | 0.23 | 0.97 | 1.01 | 0.88 | 0.99 | 0.93 | 0.31 | 1.00 |
| GC | A1a | 2,4,6-Tribromophenol | 0.87 | 0.09 | 0.97 | 1.05 | 0.55 | 0.98 | 0.97 | 0.72 | 1.00 |
| GC | A1a | 2,3-Dihydro-10-methyl-1H,7H,8H-benzo[ij]pyrano[3,2-c]]quinolizine-7,8-dione | 0.91 | 0.36 | 0.97 | 1.05 | 0.61 | 0.98 | 0.86 | 0.23 | 1.00 |
| GC | A1a | 1-(4'-Fluoro-2'-hydroxyphenyl)-2,2-dimethylpropan-1-one | 0.72 | < 0.05* | 0.97 | 0.96 | 0.73 | 0.98 | 1.56 | < 0.05* | 1.00 |
| GC | A1a | 2-Butyl-3-phenyl-5-isopropyl-4-pentylfuran | 0.96 | 0.52 | 0.98 | 1.06 | 0.45 | 0.98 | 0.98 | 0.80 | 1.00 |
| GC | A1a | 2,4,6,8-Tetramethylazulene | 1.05 | 0.38 | 0.97 | 1.20 | < 0.05* | 0.57 | 1.05 | 0.54 | 1.00 |
| GC | A1a | 2,2'-Diihydroxy-4'-methyl-5-methoxybiphenyl | 0.97 | 0.36 | 0.97 | 1.02 | 0.48 | 0.98 | 1.04 | 0.36 | 1.00 |
| GC | A1a | 9,10-seco-8b,10-dihydroxy-12-oxocalanolide A | 1.04 | 0.73 | 0.98 | 1.12 | 0.34 | 0.96 | 0.83 | 0.24 | 1.00 |
| GC | A1a | 1,2-Dihydrocyclopentafluorene | 1.01 | 0.87 | 1.00 | 1.10 | 0.12 | 0.92 | 1.03 | 0.66 | 1.00 |
| GC | A1a | 7-Acetoxy-8-ethenylquinoline | 0.93 | 0.27 | 0.97 | 1.05 | 0.52 | 0.98 | 1.19 | 0.08 | 1.00 |
| GC | A1a | 3-Amino-4,5-bis(p-biphenyl)-1H-pyrazolo[3,4-c]pyridazine | 1.00 | 0.93 | 1.00 | 0.99 | 0.76 | 0.98 | 0.92 | 0.11 | 1.00 |
| GC | A1a | 6-(1',1\-Biphenyl-4'-yl)-1,4,8-trimethylazulene"" | 1.02 | 0.89 | 1.00 | 1.10 | 0.46 | 0.98 | 0.98 | 0.85 | 1.00 |
| GC | A1a | 5-(Isobutyl)-1,4-pyrazine-2-carbonitrile | 0.79 | 0.16 | 0.97 | 1.29 | 0.11 | 0.92 | 1.13 | 0.57 | 1.00 |
| GC | A1a | N-Benzylbenzenamine | 1.06 | 0.47 | 0.98 | 0.97 | 0.71 | 0.98 | 0.99 | 0.89 | 1.00 |
| GC | A1a | (4'R,5'R)-Spiro[1,5-benzodithiepane-3,2'-[4.5]diethoxycarbonyl[1,3]dioxolane] | 1.02 | 0.85 | 1.00 | 1.01 | 0.88 | 0.99 | 1.17 | 0.20 | 1.00 |
| GC | A1a | Benz[c]acridine-7(2H)-one, 1,3,4,12-tetrahydro- | 0.91 | 0.35 | 0.97 | 1.24 | < 0.05* | 0.76 | 1.01 | 0.92 | 1.00 |
| GC | A1a | 6-Ethoxycarbonyl-1-phenyl-1H-pyrazolo[4,3-c]pyridine-4-thione | 0.94 | 0.67 | 0.98 | 0.96 | 0.74 | 0.98 | 0.95 | 0.77 | 1.00 |
| GC | A1a | 1-(tert-Butyldimethylsilyloxy)-3-vinylpent-4-en-2-ol | 1.10 | 0.70 | 0.98 | 0.82 | 0.41 | 0.98 | 0.91 | 0.75 | 1.00 |
| GC | A1a | N-{4-[N'-(4-hydroxybenzylidene)hydrazino]benzyl}-2,2,2-trifluoroacetamide | 1.05 | 0.46 | 0.97 | 1.06 | 0.41 | 0.98 | 0.92 | 0.28 | 1.00 |
| GC | A1a | Benzoic acid, 3-hydroxy-2-(tetrahydro-2H-pyran-2-yl)-, methyl ester | 1.18 | 0.22 | 0.97 | 1.01 | 0.95 | 0.99 | 0.81 | 0.20 | 1.00 |
| GC | A1a | 6-Isobutanoyl-7-methoxycoumarin | 0.91 | 0.12 | 0.97 | 0.94 | 0.40 | 0.98 | 1.00 | 0.96 | 1.00 |
| GC | A1a | 1-methyl-4-phenyl-2-[(E)-2-phenylethenyl]pyrrole | 1.27 | 0.25 | 0.97 | 1.30 | 0.29 | 0.96 | 0.98 | 0.95 | 1.00 |
| GC | A1a | 1-hydroxy-3-methoxy-2-(1-methoxyethyl)-9,10-anthraquinone | 0.90 | 0.24 | 0.97 | 1.06 | 0.51 | 0.98 | 1.04 | 0.76 | 1.00 |
| GC | A1a | 2-Methyl-4-phenylpyrido[2,3-d]pyrimidin-5(8H)-one | 0.69 | < 0.05* | 0.97 | 1.18 | 0.28 | 0.96 | 0.96 | 0.83 | 1.00 |
| GC | A1a | 2-(Pyrrolidin-1-yl)-1-(5,6,7,8-tetrahydronaphthalen-2-yl)pentan-1-one | 0.98 | 0.78 | 0.99 | 1.04 | 0.60 | 0.98 | 0.84 | 0.09 | 1.00 |
| GC | A1a | 2,3-Dihydro-1-(methylene)pyrrolo[1,2-a]indole-9-carbaldehyde N,N-dimethylhydrazone | 0.98 | 0.89 | 1.00 | 1.11 | 0.43 | 0.98 | 0.99 | 0.94 | 1.00 |
| GC | A1a | 1-methyl-2-oxidanylidene-pyridine-3-carbaldehyde | 0.83 | 0.27 | 0.97 | 1.16 | 0.37 | 0.97 | 1.06 | 0.83 | 1.00 |
| GC | A1a | 8-methoxy-2H-1-benzopyran-3-carboxylic acid methyl ester | 0.95 | 0.51 | 0.98 | 0.92 | 0.27 | 0.96 | 1.21 | < 0.05* | 1.00 |
| GC | A1a | 1,3-Benzenedicarbonitrile | 1.04 | 0.59 | 0.98 | 1.07 | 0.30 | 0.96 | 1.19 | < 0.05* | 1.00 |
| GC | A1a | 7-methylpyrrolo[1,2-a]pyrazin-1-amine | 1.15 | 0.36 | 0.97 | 0.87 | 0.43 | 0.98 | 1.30 | 0.28 | 1.00 |
| GC | A1a | 2-[(5-bromo-2-methoxybenzoyl)amino]acetic acid | 0.95 | 0.48 | 0.98 | 0.85 | < 0.05* | 0.57 | 1.12 | 0.15 | 1.00 |
| GC | A1a | Isopropyl 3-methylquinoxaline-2-carboxylate | 1.01 | 0.93 | 1.00 | 1.00 | 0.96 | 0.99 | 1.03 | 0.76 | 1.00 |
| GC | A1a | p,p'-DDE | 0.94 | 0.50 | 0.98 | 1.05 | 0.61 | 0.98 | 1.02 | 0.82 | 1.00 |
| GC | A1a | 4-hydroxy-3,4-dimethylnaphthalen-1-one | 0.88 | 0.58 | 0.98 | 1.44 | 0.15 | 0.96 | 1.26 | 0.41 | 1.00 |
| GC | A1a | Butanedioic acid, [1-(acetyloxy)-1,2,2-trimethylpropyl]-, diethyl ester | 1.08 | 0.24 | 0.97 | 0.98 | 0.81 | 0.98 | 0.92 | 0.28 | 1.00 |
| GC | A1a | Diethyl L-tartrate | 1.08 | 0.50 | 0.98 | 0.97 | 0.81 | 0.98 | 0.85 | 0.26 | 1.00 |
| GC | A1a | 5-(t-Butyl)-3,3-dimethyl-4-(benzyl)thiophen-2(3H)-one | 0.92 | 0.54 | 0.98 | 1.87 | < 0.05* | 0.57 | 0.95 | 0.80 | 1.00 |
| GC | A1a | 2-Mercaptobenzothiazole | 1.08 | 0.25 | 0.97 | 0.96 | 0.56 | 0.98 | 1.09 | 0.35 | 1.00 |
| GC | A1a | 3-(5,7-Dioxo-5,7-dihydro-pyrrolo[3,4-b]pyrazin-6-yl)-propionic acid ethyl ester | 0.84 | 0.10 | 0.97 | 1.13 | 0.25 | 0.96 | 1.19 | 0.23 | 1.00 |
| GC | A1a | 2-Oxahexacyclo[7,5,1.0.0(3,13).0(5,12).0(10,14)]pentadecan-3-ol | 1.12 | 0.24 | 0.97 | 0.99 | 0.88 | 0.99 | 1.07 | 0.59 | 1.00 |
| GC | A1a | 10H-Phenoxazine | 0.95 | 0.63 | 0.98 | 0.88 | 0.23 | 0.96 | 1.03 | 0.83 | 1.00 |
| GC | A1a | 2-(furan-3-yl)-2-(2-iodoethyl)-1,3-dioxolane | 0.76 | < 0.05* | 0.97 | 1.01 | 0.95 | 0.99 | 0.95 | 0.75 | 1.00 |
| GC | A1a | 7-isopropyl-3,4-dihydro-2H-cyclohepta[b]pyrazine | 0.89 | 0.24 | 0.97 | 1.07 | 0.56 | 0.98 | 1.49 | < 0.05* | 1.00 |
| GC | A1a | Acetic acid, [[1-(cyanohydroxymethyl)-2-naphthalenyl]oxy]-, ethyl ester | 1.10 | 0.19 | 0.97 | 1.01 | 0.90 | 0.99 | 1.07 | 0.43 | 1.00 |
| GC | A1a | Tris(2-ethylhexyl) trimellitate | 1.02 | 0.92 | 1.00 | 0.95 | 0.76 | 0.98 | 1.00 | 1.00 | 1.00 |
| GC | A1a | 1-Piperidineacetaldehyde, .alpha.-methyl-.alpha.-[[(tetrahydro-2H-pyran-2-yl)oxy]methyl]- | 1.12 | 0.24 | 0.97 | 1.16 | 0.14 | 0.96 | 1.01 | 0.95 | 1.00 |
| GC | A1a | 3,4-Diethoxy-2,5-dimethoxytetrahydrofuran | 0.98 | 0.80 | 0.99 | 1.03 | 0.78 | 0.98 | 0.95 | 0.67 | 1.00 |
| GC | A1a | 1-[2-(E)-5-(3-(1-Hydroxycyclohex-1-yl)-2-propynylidene)-1-cyclopenten-1-yl]ethynyl]cyclohexan-1-ol | 0.93 | 0.66 | 0.98 | 0.95 | 0.79 | 0.98 | 0.87 | 0.56 | 1.00 |
| GC | A1a | (2S,3S,4S)-2-[2'-((S)-4''-Benzyl-2''-oxo-oxazolidin-3''-yl)-2-oxo-ethyl]-N-cyclohexyl-5-cyano-3,4-dimethyl-piperidine | 1.02 | 0.53 | 0.98 | 1.00 | 0.95 | 0.99 | 0.93 | 0.09 | 1.00 |
| GC | A1a | Triphenyl phosphate | 0.85 | 0.10 | 0.97 | 0.87 | 0.17 | 0.96 | 1.28 | 0.06 | 1.00 |
| GC | A1a | (4R,5R)-2,2-dimethyl-4-(2-phenylethynyl)-5-vinyl-1,3-dioxolane | 0.90 | 0.32 | 0.97 | 1.02 | 0.83 | 0.98 | 1.07 | 0.58 | 1.00 |
| GC | A1a | N-{2-[(4-Acetylphenyl)ethynyl]phenyl}-2,2,2-trifluoroacetamide | 0.93 | 0.58 | 0.98 | 1.20 | 0.22 | 0.96 | 0.88 | 0.45 | 1.00 |
| GC | A1a | (3R,3aR,6S,6aR)-3-(3,4-dimethoxyphenyl)-6-(3,4,5-trimethoxyphenyl)-1,3,3a,4,6,6a-hexahydrofuro[3,4-c]furan | 0.99 | 0.87 | 1.00 | 1.07 | 0.33 | 0.96 | 1.04 | 0.63 | 1.00 |
| GC | A1a | Methyl endo-9-acetyl-7-methyltricyclo[5.2.2.0(1,5)]undec-5-ene-6-carboxylate | 1.02 | 0.69 | 0.98 | 1.13 | < 0.05* | 0.76 | 0.81 | < 0.05* | 1.00 |
| GC | A1a | 2-ethoxy-3,1-benzoxazin-4-one | 0.81 | 0.43 | 0.97 | 1.07 | 0.76 | 0.98 | 1.76 | 0.09 | 1.00 |
| GC | A1a | 9-methyl-9-phenyl-9,10-dihydrophenanthrene | 0.93 | 0.49 | 0.98 | 1.04 | 0.73 | 0.98 | 1.13 | 0.41 | 1.00 |
| GC | A1a | (Z)-Phenylazo tert-butyl sulfide | 0.93 | 0.43 | 0.97 | 1.27 | < 0.05* | 0.57 | 0.95 | 0.67 | 1.00 |
| GC | A1a | 4-Chloro-2-(6-chlorohex-1-yn-1-yl)pyridine | 0.89 | 0.09 | 0.97 | 0.89 | 0.10 | 0.92 | 1.16 | 0.12 | 1.00 |
| GC | A1a | Thiazolo[5,4-f]quinoline-2-carbonitrile | 1.09 | 0.40 | 0.97 | 1.10 | 0.32 | 0.96 | 0.95 | 0.66 | 1.00 |
| GC | A1a | 1,4,5,6-Tetrahydro-1-(4',6'-heptadienoyl)-pyridine | 0.92 | 0.57 | 0.98 | 0.95 | 0.77 | 0.98 | 0.85 | 0.43 | 1.00 |
| GC | A1a | (3S,5R,8aS)-3-butyl-5-propyl-1,2,3,5,6,7,8,8a-octahydroindolizine | 1.07 | 0.65 | 0.98 | 0.78 | 0.13 | 0.93 | 0.83 | 0.33 | 1.00 |
| GC | A1a | 4,6,7-trimethylinden-1-one | 0.96 | 0.66 | 0.98 | 1.23 | 0.05 | 0.82 | 1.13 | 0.39 | 1.00 |
| GC | A1a | methyl 1-butyl-2-oxocycloheptane-1-carboxylate | 0.85 | 0.33 | 0.97 | 0.98 | 0.89 | 0.99 | 1.22 | 0.43 | 1.00 |
| GC | A1a | 4-Acetoxy-10,12-dimethyl-14-oxatricyclo[7.4.0.0(6,7)]tetradecane | 0.87 | 0.10 | 0.97 | 1.01 | 0.92 | 0.99 | 0.96 | 0.77 | 1.00 |
| GC | A1a | 2,3,4,4a,5,6,6a,11b-Octahydro-6a-hydroperoxy-8-methoxy-7,9,10-trimethyl-1H-benzo[a]carbazole | 0.90 | 0.19 | 0.97 | 1.04 | 0.65 | 0.98 | 0.92 | 0.41 | 1.00 |
| GC | A1a | 7-Cyano-2-oxatricyclo[5.3.1.0(1,11)]undec-1(10)-en-9-one | 0.54 | 0.07 | 0.97 | 1.22 | 0.52 | 0.98 | 0.81 | 0.70 | 1.00 |
| GC | A1a | 1,2,4-Trichlorobenzene | 1.11 | 0.67 | 0.98 | 1.10 | 0.68 | 0.98 | 0.77 | 0.38 | 1.00 |
| GC | A1a | Apocynin | 0.86 | 0.37 | 0.97 | 0.97 | 0.87 | 0.99 | 0.73 | 0.20 | 1.00 |
| GC | A1a | 2-Acetoxy-3-chlorophenol | 0.96 | 0.35 | 0.97 | 0.98 | 0.60 | 0.98 | 1.03 | 0.56 | 1.00 |
| GC | A1a | 4,5-Dihydrofuro[1,2-a]naphthalene | 1.06 | 0.76 | 0.99 | 1.01 | 0.96 | 0.99 | 0.90 | 0.67 | 1.00 |
| GC | A1a | 2-tert-Butylfuro[4,5-d][1,3]dioxan-6-one | 1.03 | 0.77 | 0.99 | 1.25 | < 0.05* | 0.76 | 0.96 | 0.74 | 1.00 |
| GC | A1a | 10-(p-Tolyl)benzo[h]quinoline | 1.01 | 0.82 | 1.00 | 0.99 | 0.75 | 0.98 | 1.00 | 0.99 | 1.00 |
| GC | A1a | Isoxaben | 0.89 | 0.26 | 0.97 | 1.19 | 0.13 | 0.94 | 1.03 | 0.80 | 1.00 |
| GC | A1a | 1-(5-Methyl-2-oxo-2,5-dihydrofuran-3-yl)-9-(tert-butyldimethylsiloxy)nonan-8-ol | 0.87 | 0.08 | 0.97 | 1.08 | 0.35 | 0.96 | 0.97 | 0.75 | 1.00 |
| GC | A1a | 3-butylcyclohexan-1-one | 0.89 | 0.29 | 0.97 | 1.02 | 0.87 | 0.99 | 1.11 | 0.50 | 1.00 |
| GC | A1a | 6,7-Dimethoxy-1-[(p-tolyl)sulfinyl]methyl-1-(trifluoromethyl)-1,2,3,4-tetrahydroisoquinoline | 0.98 | 0.92 | 1.00 | 1.39 | 0.15 | 0.96 | 0.82 | 0.48 | 1.00 |
| GC | A1a | 2-[Chloro(diphenyl)methyl]pyridine | 0.92 | 0.41 | 0.97 | 1.33 | < 0.05* | 0.57 | 0.85 | 0.19 | 1.00 |
| GC | A1a | 1-[(1S,2R)-2-propoxycyclopentyl]hexane-1,2-dione | 1.09 | 0.07 | 0.97 | 1.00 | 0.99 | 1.00 | 1.07 | 0.19 | 1.00 |
| GC | A1a | Dodecyltriethoxysilane | 1.03 | 0.81 | 1.00 | 1.02 | 0.86 | 0.98 | 0.85 | 0.41 | 1.00 |
| GC | A1a | N-(1-methyl-2,5-dioxo-3-[1]benzopyrano[4,3-b]pyridinyl)acetamide | 0.87 | 0.35 | 0.97 | 1.00 | 0.98 | 1.00 | 1.07 | 0.74 | 1.00 |
| GC | A1a | 6-Phenyl-2-pyridinamine | 1.07 | 0.23 | 0.97 | 1.04 | 0.54 | 0.98 | 1.03 | 0.67 | 1.00 |
| GC | A1a | 4-(3-ethoxyphenyl)-N,N-diethyl-aniline | 1.00 | 0.94 | 1.00 | 1.07 | 0.28 | 0.96 | 1.05 | 0.55 | 1.00 |
| GC | A1a | (4R,4aR,8aR)-4,8a-dimethyl-1,2,3,4,7,8-hexahydronaphthalen-4a-ol | 0.64 | < 0.05* | 0.97 | 1.27 | 0.29 | 0.96 | 0.94 | 0.80 | 1.00 |
| GC | A1a | Padimate O | 1.03 | 0.59 | 0.98 | 0.96 | 0.54 | 0.98 | 1.04 | 0.69 | 1.00 |
| GC | A1a | (2E)-2-(phenylmethylene)-3-benzofuranone | 1.00 | 0.98 | 1.00 | 0.83 | 0.08 | 0.92 | 0.89 | 0.39 | 1.00 |
| GC | A1a | methyl 2-(4-methoxyphenyl)propanoate | 0.90 | < 0.05* | 0.97 | 1.03 | 0.54 | 0.98 | 1.07 | 0.34 | 1.00 |
| GC | A1a | (8-acetoxy-1-cyano-7-ethoxy-6-methyl-5-isoquinolyl) acetate | 0.89 | 0.14 | 0.97 | 0.83 | < 0.05* | 0.67 | 1.14 | 0.18 | 1.00 |
| GC | A1a | 15-Nonacosanone | 0.96 | 0.55 | 0.98 | 0.96 | 0.58 | 0.98 | 1.00 | 0.96 | 1.00 |
| GC | A1a | 1,4:5,8-Dimethanonaphthalene, 6-butyl-1,2,3,4,4a,5,8,8a-octahydro-, (1.alpha.,4.alpha.,4a.beta.,5.beta.,8.beta.,8a.beta.)- | 0.99 | 0.96 | 1.00 | 1.30 | 0.09 | 0.92 | 1.06 | 0.76 | 1.00 |
| GC | A1a | (Z)-2-Ethyl-5-bromo-2-dicyanomethyladamantane | 1.05 | 0.38 | 0.97 | 1.02 | 0.67 | 0.98 | 1.07 | 0.31 | 1.00 |
| GC | A1a | 3-(5,5,6-Trimethylbicyclo[2.2.1]hept-2-ylidene)-2-propanone oxime | 1.03 | 0.89 | 1.00 | 1.20 | 0.34 | 0.96 | 0.52 | < 0.05* | 1.00 |
| GC | A1a | (3,6-dimethyl-4-oxidanylidene-2H-chromen-3-yl) nitrate | 1.21 | 0.38 | 0.97 | 1.60 | < 0.05* | 0.57 | 0.84 | 0.57 | 1.00 |
| GC | A1a | 6,7-dimethoxy-1-(methoxymethyl)-3,4-dihydroisoquinoline | 1.12 | 0.26 | 0.97 | 1.14 | 0.22 | 0.96 | 0.85 | 0.23 | 1.00 |
| GC | A1a | Methyl (E)-3-methoxy-4-oxo-4-phenylbut-2-enoate | 1.02 | 0.83 | 1.00 | 1.00 | 0.96 | 0.99 | 0.84 | 0.15 | 1.00 |
| GC | A1a | 3,3,7-trimethyl-5-nitro-1H-indol-2-one | 0.91 | < 0.05* | 0.97 | 1.04 | 0.44 | 0.98 | 1.05 | 0.48 | 1.00 |
| GC | A1a | 1-hydroxy-1-methyl-tetralin-6-carbaldehyde | 0.92 | 0.79 | 0.99 | 1.52 | 0.18 | 0.96 | 1.28 | 0.56 | 1.00 |
| GC | A1a | Acetosyringon | 0.98 | 0.62 | 0.98 | 0.94 | 0.20 | 0.96 | 0.99 | 0.94 | 1.00 |
| GC | A1a | Methyl (S)-1-(4,4-diphenylbut-3-en-1-yl)pyrrolidine-2-carboxylate | 1.05 | 0.84 | 1.00 | 1.06 | 0.80 | 0.98 | 0.81 | 0.43 | 1.00 |
| GC | A1a | Octabenzone | 1.07 | 0.57 | 0.98 | 0.93 | 0.56 | 0.98 | 0.90 | 0.58 | 1.00 |
| GC | A1a | Diphenyl(1,3,5-trimethyl-1H-pyrazol-4-yl)amine | 1.15 | 0.12 | 0.97 | 1.04 | 0.65 | 0.98 | 0.96 | 0.72 | 1.00 |
| GC | A1a | Octicizer | 1.01 | 0.93 | 1.00 | 0.99 | 0.89 | 0.99 | 0.97 | 0.80 | 1.00 |
| GC | A1a | 6-Methoxy-2-phenyl-2-(phenylthio)-3-benzofuranone | 1.07 | 0.62 | 0.98 | 0.83 | 0.24 | 0.96 | 1.07 | 0.72 | 1.00 |
| GC | A1a | 2-Acetonyl-3-acetyl-5-methoxy-1,4-naphthoquinone | 0.99 | 0.89 | 1.00 | 1.08 | 0.15 | 0.96 | 1.05 | 0.44 | 1.00 |
| GC | A1a | Bumetrizole | 1.18 | 0.35 | 0.97 | 1.62 | < 0.05* | 0.57 | 0.77 | 0.22 | 1.00 |
| GC | A1a | (2SR,3aRS,6aRS,10aRS)-1,7,7,10a-Tetramethyl-2-phenyl-2,3,3a,6,6a,7,8,9,10,10a-decahydro-4-oxabenz[e]azulen-5-one | 0.95 | 0.58 | 0.98 | 1.17 | 0.11 | 0.92 | 0.76 | < 0.05* | 1.00 |
| GC | A1a | Tri(2-ethylhexyl) phosphate | 1.01 | 0.95 | 1.00 | 1.07 | 0.56 | 0.98 | 1.02 | 0.91 | 1.00 |
| GC | A1a | 2-[(E)-pentadec-1-enyl]benzene-1,3-dicarbaldehyde | 1.04 | 0.61 | 0.98 | 1.03 | 0.70 | 0.98 | 1.01 | 0.96 | 1.00 |
| GC | A1a | 1-(6-Methoxypyridin-2-yl)propan-2-one | 0.91 | 0.56 | 0.98 | 1.32 | 0.10 | 0.92 | 1.90 | < 0.05* | 1.00 |
| GC | A1a | (R)-4-Isobutyloxazolidin-2-one | 0.93 | 0.65 | 0.98 | 1.02 | 0.87 | 0.99 | 1.60 | < 0.05* | 1.00 |
| GC | A1a | methyl 2-(2,4,6-trimethylphenyl)acetate | 0.86 | 0.42 | 0.97 | 1.12 | 0.55 | 0.98 | 0.75 | 0.25 | 1.00 |
| GC | A1a | 2-[(4-Chlorobenzoyl)(2-ethoxycarbonylbenzyl)methylene]-1H-imidazole | 1.04 | 0.56 | 0.98 | 1.08 | 0.29 | 0.96 | 1.00 | 1.00 | 1.00 |
| GC | A1a | 3-(2-Chloromethylphenyl)-2,2-dimethylpropionaldehyde | 0.68 | 0.09 | 0.97 | 1.13 | 0.60 | 0.98 | 1.30 | 0.38 | 1.00 |
| GC | A1a | (+)-(2S,3R)-4-Benzyloxy-3-(tert-butyldimethylsiloxy)-2-methylbutanol | 1.12 | 0.13 | 0.97 | 1.08 | 0.30 | 0.96 | 0.97 | 0.71 | 1.00 |
| GC | A1a | cis-3,5-[di(t-Butyl)silanedioxy]cyclopent-1-ene | 0.95 | 0.19 | 0.97 | 1.06 | 0.11 | 0.92 | 1.04 | 0.34 | 1.00 |
| GC | A1a | 4-hydroperoxy-2,3,4-trimethylnaphthalen-1-one | 0.84 | < 0.05* | 0.97 | 1.10 | 0.27 | 0.96 | 0.89 | 0.23 | 1.00 |
| GC | A1a | Ethanethioic acid, S-bicyclo[2.2.1]hept-2-en-7-yl ester, anti- | 0.92 | 0.25 | 0.97 | 1.04 | 0.63 | 0.98 | 1.06 | 0.53 | 1.00 |
| GC | A1a | 1-(benzo[d][1,3]dioxol-5-yl)-2-(4-methylpiperidin-1-yl)pentan-1-one | 0.88 | 0.31 | 0.97 | 1.06 | 0.65 | 0.98 | 1.26 | 0.16 | 1.00 |
| GC | A1a | Propylparaben | 1.11 | 0.31 | 0.97 | 1.10 | 0.41 | 0.98 | 0.91 | 0.49 | 1.00 |
| GC | A1a | 2,4-Dihydroxy-5-methoxy-6-methyl-tetrahydropyran | 1.14 | 0.22 | 0.97 | 0.95 | 0.61 | 0.98 | 1.15 | 0.33 | 1.00 |
| GC | A1a | Octinoxate | 1.02 | 0.80 | 0.99 | 1.09 | 0.28 | 0.96 | 0.91 | 0.36 | 1.00 |
| GC | A1a | (Z)-13-Ethylene-9,10-dimethoxy-2,3-(methylenedioxy)-8,14-cycloberbine | 1.00 | 0.99 | 1.00 | 1.05 | 0.46 | 0.98 | 0.90 | 0.23 | 1.00 |
| GC | A1a | 1,7-Dimethylxanthine | 0.95 | 0.34 | 0.97 | 1.00 | 0.95 | 0.99 | 1.05 | 0.46 | 1.00 |
| GC | A1a | 5-carboxylated pyrrole derivative of 4,4-dimethyl-2-(N-methylpyrrol-2-yl)oxazoline | 1.06 | 0.36 | 0.97 | 0.91 | 0.15 | 0.96 | 1.01 | 0.92 | 1.00 |
| GC | A1a | Pestalolide | 0.60 | 0.26 | 0.97 | 0.97 | 0.93 | 0.99 | 0.61 | 0.44 | 1.00 |
| GC | A1a | Methyl (1R*,5S*)-1-Hydroxy-1-methylspiro[4.5]dec-7-ene-8-carboxylate | 1.13 | 0.09 | 0.97 | 0.93 | 0.33 | 0.96 | 0.99 | 0.93 | 1.00 |
| GC | A1a | (3-methylphenyl) 3-nitrobenzoate | 1.01 | 0.89 | 1.00 | 1.05 | 0.48 | 0.98 | 1.04 | 0.65 | 1.00 |
| GC | A1a | 2-(3-furanyl)-1H-benzimidazole | 0.96 | 0.73 | 0.98 | 1.07 | 0.53 | 0.98 | 1.10 | 0.49 | 1.00 |
| GC | A1a | (1R,3S)-3-phenylcyclohexanol | 0.79 | 0.16 | 0.97 | 0.97 | 0.84 | 0.98 | 1.26 | 0.34 | 1.00 |
| GC | A1a | 9-[1-(3-methylphenyl)ethylidene]fluorene | 1.05 | < 0.05* | 0.97 | 0.97 | 0.25 | 0.96 | 1.01 | 0.78 | 1.00 |
| GC | A1a | 2,3,3',4,4',5'-Hexachlorobiphenyl | 0.96 | 0.59 | 0.98 | 1.00 | 0.98 | 1.00 | 1.01 | 0.94 | 1.00 |
| GC | A1a | (2-iodanyl-2-triethylsilyl-ethyl) 2,2,2-tris(fluoranyl)ethanoate | 1.08 | 0.59 | 0.98 | 1.14 | 0.35 | 0.96 | 0.77 | 0.12 | 1.00 |
| GC | A1a | 4-ethyl-4-methyl-5-methylene-1,3-dioxolan-2-one | 1.27 | 0.18 | 0.97 | 0.75 | 0.12 | 0.93 | 0.85 | 0.48 | 1.00 |
| GC | A1a | (4aS,8aS)-5,5,8a-trimethyl-1-methylene-3,4,4a,6,7,8-hexahydronaphthalen-2-one | 0.92 | 0.30 | 0.97 | 0.84 | < 0.05* | 0.76 | 1.10 | 0.35 | 1.00 |
| GC | A1a | Dibutyl phthalate | 1.09 | 0.23 | 0.97 | 1.09 | 0.21 | 0.96 | 0.98 | 0.83 | 1.00 |
| GC | A1a | 7-Methoxy-2,5-dimethyl-chromone | 0.92 | 0.09 | 0.97 | 1.00 | 0.98 | 1.00 | 1.05 | 0.44 | 1.00 |
| GC | A1a | (R)-(S)-.alpha.-Methoxyphenylacetic acid (NPA) | 0.83 | 0.09 | 0.97 | 1.08 | 0.42 | 0.98 | 0.92 | 0.52 | 1.00 |
| GC | A1a | 4-Ethyl-3-methyl-1,2,4-thiadiazol-5-one | 1.02 | 0.92 | 1.00 | 0.98 | 0.92 | 0.99 | 1.10 | 0.67 | 1.00 |
| GC | A1a | 3-Aminochrysene | 1.46 | 0.06 | 0.97 | 1.07 | 0.73 | 0.98 | 0.88 | 0.59 | 1.00 |
| GC | A1a | 2-(tert-Butyl-dimethyl-silanyloxy)-3-methyl-cyclopent-2-enone | 1.09 | 0.24 | 0.97 | 1.13 | 0.08 | 0.92 | 0.95 | 0.53 | 1.00 |
| GC | A1a | 10-(3-Acetoxyprop-2-yl)-3,7-dimethylbicyclo[4.4.0]dec-2-ene | 0.83 | < 0.05* | 0.97 | 0.91 | 0.43 | 0.98 | 0.99 | 0.95 | 1.00 |
| GC | A1a | (Z)-2-Phenyl-1-(triethylsilyl)-1,4-pentadiene | 1.00 | 1.00 | 1.00 | 1.35 | 0.25 | 0.96 | 0.92 | 0.79 | 1.00 |
| GC | A1a | (2,4-dimethoxy-3-methyl-phenyl) acetate | 0.98 | 0.45 | 0.97 | 0.98 | 0.40 | 0.98 | 0.96 | 0.20 | 1.00 |
| GC | A1a | Niloticin | 1.04 | 0.61 | 0.98 | 1.07 | 0.42 | 0.98 | 1.03 | 0.78 | 1.00 |
| GC | A1a | Methyl 4-[2-(2-Chloro-3-cyanopyrid-4-yl)ethyl]benzoate | 0.96 | 0.45 | 0.97 | 1.15 | < 0.05* | 0.57 | 0.89 | 0.11 | 1.00 |
| GC | A1a | 1,4-Dideoxy-1,4-imino-D-arabinitol | 0.84 | 0.05 | 0.97 | 1.16 | 0.10 | 0.92 | 0.97 | 0.79 | 1.00 |
| GC | A1a | Triisobutyl phosphate | 1.54 | 0.30 | 0.97 | 0.94 | 0.87 | 0.99 | 0.79 | 0.55 | 1.00 |
| GC | A1a | (2'R*,3'R*,.alpha.S)-2,3-Epoxyoctyl .alpha.-methoxyphenylacetate | 0.88 | 0.32 | 0.97 | 0.86 | 0.27 | 0.96 | 1.00 | 0.99 | 1.00 |
| GC | A1a | 5-Prop-2-ynyl-cyclohex-1-enylamine | 0.98 | 0.92 | 1.00 | 0.92 | 0.59 | 0.98 | 0.96 | 0.81 | 1.00 |
| GC | A1a | 2-{[Dimethyl(3-methylbut-2-en-1-yl)silyl]methyl}allyl Acetate | 0.90 | 0.29 | 0.97 | 0.90 | 0.36 | 0.97 | 1.04 | 0.80 | 1.00 |
| GC | A1a | 7a-Carboxymethyl-1-methylene-1,4,5,6,7,7a-hexahydro-2H-indene | 0.89 | 0.41 | 0.97 | 1.05 | 0.75 | 0.98 | 0.72 | 0.08 | 1.00 |
| GC | A1a | (5S,6R)-5,6-dimethyl-3,4,5,6-tetrahydro-2H-cyclopenta[b]pyran-7-one | 0.83 | 0.06 | 0.97 | 1.05 | 0.71 | 0.98 | 1.30 | 0.11 | 1.00 |
| GC | A1a | 9-phenylcarbazole | 1.04 | 0.64 | 0.98 | 1.08 | 0.28 | 0.96 | 1.07 | 0.44 | 1.00 |
| GC | A1a | (S)-2-(4-Methoxyphenyl)-3-methylbutan-1-ol | 1.05 | 0.72 | 0.98 | 1.09 | 0.54 | 0.98 | 1.04 | 0.82 | 1.00 |
| GC | A1a | 1,4-Naphthalenedione, 5-[(acetyloxy)methyl]-4a,5-dihydro-2-methoxy-4a-methyl-, trans- | 1.12 | 0.42 | 0.97 | 1.03 | 0.83 | 0.98 | 1.34 | 0.10 | 1.00 |
| GC | A1a | 7-Phenylethyl-4,6-dioxa-5-carbonyl-spiro[2,5]-7-octene | 0.94 | 0.33 | 0.97 | 1.01 | 0.93 | 0.99 | 0.96 | 0.57 | 1.00 |
| GC | A1a | 1-Cyano-1-(t-butyl)-2-aminoethene | 1.11 | 0.51 | 0.98 | 1.28 | 0.23 | 0.96 | 1.05 | 0.78 | 1.00 |
| GC | A1a | 2-Carboxy-4-methylbicyclo[2.2.2]oct-2-en-1-ol | 0.85 | 0.24 | 0.97 | 0.87 | 0.23 | 0.96 | 1.05 | 0.78 | 1.00 |
| GC | A1a | 3-isopropyl-2-(9H-xanthen-9-yl)-1H-indole | 1.15 | 0.05 | 0.97 | 0.92 | 0.25 | 0.96 | 1.01 | 0.88 | 1.00 |
| GC | A1a | 5,5-diethyl-1,3-diazinane-2,4,6-trione | 0.87 | 0.40 | 0.97 | 1.02 | 0.92 | 0.99 | 0.89 | 0.51 | 1.00 |
| GC | A1a | 6-Deoxy-1,2-O-isopropylidene-5-O-thiocarbonyl-.alpha.,D-glucofuranose | 1.03 | 0.52 | 0.98 | 0.96 | 0.29 | 0.96 | 1.08 | 0.15 | 1.00 |
| GC | A1a | 4,5-dimethyl-3-(2-methylpropyl)-2,3-dihydroisoindol-1-one | 1.11 | 0.45 | 0.97 | 1.14 | 0.32 | 0.96 | 1.08 | 0.66 | 1.00 |
| GC | A1a | (E)-Methyl undec-2-en-4-ynoate | 1.02 | 0.68 | 0.98 | 0.91 | 0.08 | 0.92 | 1.04 | 0.48 | 1.00 |
| GC | A1a | Tributyl citrate | 1.01 | 0.91 | 1.00 | 1.04 | 0.60 | 0.98 | 1.01 | 0.94 | 1.00 |
| GC | A1a | Ethyl 3-phenyl-3-(2\,4\""-dinitrophenylhydrazono)-2-(1'-methylpiperidin-2'-ylidene)propionate"" | 0.97 | 0.34 | 0.97 | 0.97 | 0.37 | 0.97 | 1.01 | 0.73 | 1.00 |
| GC | A1a | 3-[tert-butyl(diphenyl)silyl]prop-2-yn-1-ol | 0.91 | < 0.05* | 0.97 | 1.03 | 0.62 | 0.98 | 0.91 | 0.14 | 1.00 |
| GC | A1a | (3Z,9Z)-3,9-Dodecadiene-2,5,8,11-tetraone | 0.91 | 0.20 | 0.97 | 1.10 | 0.23 | 0.96 | 0.88 | 0.19 | 1.00 |
| GC | A1a | 5-Acetoxy-2,7-dimethylchromone | 0.66 | 0.15 | 0.97 | 1.31 | 0.37 | 0.97 | 0.77 | 0.45 | 1.00 |
| GC | A1a | 4-octoxybenzaldehyde | 1.07 | 0.71 | 0.98 | 0.98 | 0.93 | 0.99 | 0.81 | 0.48 | 1.00 |
| GC | A1a | 3-(cyclopenten-1-yl)prop-2-ynyl methanesulfonate | 0.99 | 0.96 | 1.00 | 1.17 | 0.35 | 0.96 | 0.98 | 0.93 | 1.00 |
| GC | A1a | 3-hydroxy-3-(2-methoxycyclohexyl)-2-methylisoindol-1-one | 1.07 | 0.24 | 0.97 | 1.04 | 0.57 | 0.98 | 1.03 | 0.68 | 1.00 |
| GC | A1a | Ethyl 3-nitro-4,7-dihydro-4,7-ethano-2H-isoindole-1-carboxylate | 1.00 | 0.93 | 1.00 | 1.05 | 0.27 | 0.96 | 1.09 | 0.11 | 1.00 |
| GC | A1a | 2,4-Dimethoxy-6-methylphenol | 0.94 | 0.23 | 0.97 | 1.00 | 0.97 | 1.00 | 1.07 | 0.30 | 1.00 |
| GC | A1a | (1,1,1-trichloro-2-methoxypropan-2-yl)oxycyclohexane | 1.09 | 0.55 | 0.98 | 1.05 | 0.75 | 0.98 | 0.85 | 0.37 | 1.00 |
| GC | A1a | Piperine | 0.99 | 0.90 | 1.00 | 1.01 | 0.89 | 0.99 | 0.92 | 0.14 | 1.00 |
| GC | A1a | 2-Oxo-2H-chromen-7-yl Acetate | 0.97 | 0.85 | 1.00 | 1.10 | 0.59 | 0.98 | 1.47 | 0.10 | 1.00 |
| GC | A1a | 2-[(3R)-3,7-dimethyloct-6-enylidene]malononitrile | 0.89 | < 0.05* | 0.97 | 0.98 | 0.70 | 0.98 | 1.16 | 0.06 | 1.00 |
| GC | A1a | 5-tert-butyl-2-(4-tert-butylphenyl)-1,3-dinitro-benzene | 1.15 | 0.14 | 0.97 | 0.99 | 0.89 | 0.99 | 0.96 | 0.75 | 1.00 |
| GC | A1a | Furo[3',4':6,7]naphtho[2,3-d]-1,3-dioxole, 5,5a,6,8,8a,9-hexahydro-, (5.alpha.,5a.beta.,8a.beta.)- | 0.99 | 0.91 | 1.00 | 1.09 | 0.32 | 0.96 | 0.85 | 0.23 | 1.00 |
| GC | A1a | Thiophen-3-yl 2-aminobenzoate | 0.91 | 0.40 | 0.97 | 1.17 | 0.19 | 0.96 | 1.09 | 0.53 | 1.00 |
| GC | A1a | 3,4-Dihydro-5,8-dimethoxy-1,3-dimethyl-1H-2-benzopyran | 1.11 | 0.49 | 0.98 | 1.17 | 0.34 | 0.96 | 0.67 | 0.08 | 1.00 |
| GC | A1a | 2-Amino-3-cyano-6-methyl-4H-chromene | 0.98 | 0.56 | 0.98 | 1.03 | 0.43 | 0.98 | 1.01 | 0.80 | 1.00 |
| GC | A1a | (1S,3S)-2-iodanyl-2,3-dihydro-1H-indene-1,3-diol | 0.86 | 0.17 | 0.97 | 1.25 | 0.09 | 0.92 | 0.87 | 0.40 | 1.00 |
| GC | A1a | 1-(Phenylethynyl)-1-benzylcyclopropane | 1.19 | 0.21 | 0.97 | 1.01 | 0.93 | 0.99 | 0.98 | 0.89 | 1.00 |
| GC | A1a | Bromoxynil | 0.95 | 0.24 | 0.97 | 1.10 | 0.06 | 0.83 | 0.96 | 0.52 | 1.00 |
| GC | A1a | (6S)-6-undecyl-2-oxanone | 0.90 | 0.41 | 0.97 | 0.81 | 0.13 | 0.94 | 1.10 | 0.56 | 1.00 |
| GC | A1a | 1,1-Diformylcyclohex-3-ene | 1.32 | 0.33 | 0.97 | 1.11 | 0.68 | 0.98 | 0.57 | 0.12 | 1.00 |
| LC | A1b | Potentially chlorinated | 1.44 | 0.07 | 0.54 | 1.06 | 0.76 | 0.95 | 0.54 | < 0.05* | 0.42 |
| GC | A1b | 2H-Furo[2,3-h]-1-benzopyran-2-one | 0.82 | 0.44 | 0.92 | 1.09 | 0.75 | 0.95 | 0.89 | 0.73 | 0.99 |
| GC | A1b | Aceclofenac | 1.21 | 0.32 | 0.92 | 0.99 | 0.95 | 0.95 | 1.18 | 0.51 | 0.99 |
| GC | A1b | 6-Bromo-3-(3,4-dimethoxyphenethyl)quinazoline-2,4(1H,3H)-dione | 1.02 | 0.93 | 0.93 | 1.21 | 0.45 | 0.89 | 0.95 | 0.88 | 0.99 |
| GC | A1b | anti-14-bromo-5,8-dimethoxy-16-methyl-2,11-dithia[3]paracyclo[3](2,4)thiophenophane | 0.79 | 0.25 | 0.92 | 1.21 | 0.42 | 0.89 | 1.26 | 0.40 | 0.99 |
| GC | A1b | 7-Methyl-8-hydroxy-9-oxo-1-azabicyclo[4.3.0]non-7-ene | 0.88 | 0.52 | 0.92 | 0.96 | 0.83 | 0.95 | 1.01 | 0.97 | 0.99 |
| GC | A1b | 2-(3-Nitrophenyl)-2,3-dihydro-1H-quinazolin-4-one | 1.23 | 0.29 | 0.92 | 0.76 | 0.18 | 0.88 | 0.96 | 0.88 | 0.99 |
| GC | A1b | 6-(2-pyridinyl)-2-pyridinecarboxaldehyde | 1.04 | 0.81 | 0.92 | 0.89 | 0.55 | 0.95 | 1.16 | 0.52 | 0.99 |
| GC | A1b | (1E,2E)-1-{2-[3-(Trifluoromethyl)phenyl]hydrazono}non-2-en-4-ol | 0.90 | 0.55 | 0.92 | 0.82 | 0.28 | 0.89 | 1.08 | 0.74 | 0.99 |
| GC | A1b | ethyl 2-[6-(dimethoxymethyl)-1,3-benzodioxol-5-yl]-2-oxoacetate | 1.60 | < 0.05* | 0.54 | 0.68 | 0.12 | 0.87 | 1.26 | 0.42 | 0.99 |
| GC | A1b | 3-Adamantyl-5-(t-butyl)-3-hydroxybenzaldehyde | 0.92 | 0.66 | 0.92 | 1.07 | 0.73 | 0.95 | 0.66 | 0.09 | 0.99 |
| GC | A1b | 5-(But-3'-en-1'-yl)-6-methylizidine | 0.96 | 0.86 | 0.92 | 1.02 | 0.94 | 0.95 | 1.01 | 0.97 | 0.99 |
| GC | A1b | 2-(6-bromo-1,3-benzodioxol-5-yl)-1-(2-bromophenyl)ethanone | 1.40 | 0.07 | 0.54 | 1.51 | < 0.05* | 0.52 | 0.89 | 0.63 | 0.99 |
| GC | A1b | Methyl 4'-(1,1-dimethylprop-2-ynyloxy)-2'-methoxycinnamate | 0.96 | 0.83 | 0.92 | 0.71 | 0.09 | 0.87 | 0.98 | 0.95 | 0.99 |
| GC | A1b | 4-[2-(1,3-Dithianyl)]-4-(2-ethoxycarbonylpyrrol-4-yl)butyric acid | 0.89 | 0.63 | 0.92 | 0.87 | 0.58 | 0.95 | 0.98 | 0.95 | 0.99 |
| GC | A1b | 3-Iodophenol | 1.10 | 0.62 | 0.92 | 1.17 | 0.44 | 0.89 | 1.24 | 0.37 | 0.99 |
| GC | A1b | Diflubenzuron | 0.85 | 0.43 | 0.92 | 1.05 | 0.81 | 0.95 | 1.22 | 0.53 | 0.99 |
| GC | A1b | spiro[2,2,Dicyano-4-[(4-vinyl)phenyl]cyclobutane-1,1'-bis[1,2,5]thiadiazolodicyanocyclohexanemethane | 0.92 | 0.69 | 0.92 | 0.79 | 0.28 | 0.89 | 1.00 | 0.99 | 0.99 |
| GC | A1b | 4,5-Dichloro-10-(4-nitrobenzyl)-10H-anthracen-9-one | 0.98 | 0.93 | 0.93 | 0.98 | 0.93 | 0.95 | 0.64 | 0.17 | 0.99 |
| GC | A1b | 3-(Methylthio)propyl isothiocyanate | 1.33 | 0.14 | 0.83 | 0.92 | 0.67 | 0.95 | 0.90 | 0.67 | 0.99 |
| GC | A1b | 1-methyl-12-phenyl-12-azatricyclo[6.3.1.02,7]dodeca-2,4,6,10-tetraen-9-one | 0.91 | 0.61 | 0.92 | 0.88 | 0.48 | 0.89 | 1.14 | 0.56 | 0.99 |
| GC | A1b | N-(1,3-Diphenyl-2-propynyl)-phenyl carbamate | 1.17 | 0.49 | 0.92 | 0.84 | 0.46 | 0.89 | 0.99 | 0.98 | 0.99 |
| GC | A1b | 2,3,6-trimethoxydibenz[b,f]oxepine | 0.94 | 0.73 | 0.92 | 0.86 | 0.42 | 0.89 | 1.42 | 0.12 | 0.99 |
| GC | A1b | 2-(2-methoxyethyl)-3,4-dihydronaphthalen-1(2H)-one | 1.52 | 0.07 | 0.54 | 0.91 | 0.70 | 0.95 | 0.82 | 0.46 | 0.99 |
| GC | A1b | 6-phenyl-2,3,4,5-tetrahydropyridine | 1.34 | 0.17 | 0.83 | 0.97 | 0.90 | 0.95 | 0.88 | 0.63 | 0.99 |
| GC | A1b | spiro[2,2,Dicyano-4-[(3-vinyl)phenyl]cyclobutane-1,1'-bis[1,2,5]thiadiazolodicyanocyclohexanemethane | 1.04 | 0.81 | 0.92 | 0.66 | < 0.05* | 0.52 | 1.19 | 0.47 | 0.99 |
| GC | A1b | 2-(1-adamantyl)-3,3-bis(trifluoromethyl)spiro[1,4,2-dithiazolidine-5,2'-adamantane] | 1.05 | 0.78 | 0.92 | 0.84 | 0.36 | 0.89 | 1.11 | 0.67 | 0.99 |
| GC | A1b | 2,6-Bis(4-t-butylphenyl)phenol | 0.85 | 0.53 | 0.92 | 0.91 | 0.72 | 0.95 | 1.58 | 0.18 | 0.99 |
| GC | A1b | (S,R)-N-(1-Methyl-3-butynyl)-N-(1-phenylethyl)amine | 1.24 | 0.27 | 0.92 | 0.74 | 0.15 | 0.88 | 0.79 | 0.37 | 0.99 |
| GC | A1b | Hydrastine | 0.93 | 0.78 | 0.92 | 0.80 | 0.41 | 0.89 | 0.79 | 0.50 | 0.99 |
| LC | A2a | Pipecolate | 0.89 | 0.10 | 0.73 | 0.90 | 0.13 | 0.56 | 0.96 | 0.63 | 0.64 |
| LC | A2a | Glycochenodeoxycholic acid | 1.15 | 0.21 | 0.73 | 1.19 | 0.14 | 0.56 | 0.94 | 0.64 | 0.64 |
| LC | A2a | Pipecolate | 1.07 | 0.60 | 0.80 | 1.09 | 0.45 | 0.69 | 0.85 | 0.36 | 0.64 |
| LC | A2a | 5-oxo-D-Proline | 1.05 | 0.82 | 0.82 | 0.91 | 0.69 | 0.69 | 0.82 | 0.45 | 0.64 |
| LC | A2a | Cortisone | 0.74 | 0.27 | 0.73 | 0.81 | 0.50 | 0.69 | 1.40 | 0.36 | 0.64 |
| LC | A2a | Pipecolate | 1.05 | 0.76 | 0.82 | 1.22 | 0.39 | 0.69 | 1.17 | 0.54 | 0.64 |
| LC | A2a | Cortisol | 0.88 | 0.49 | 0.79 | 0.91 | 0.65 | 0.69 | 1.33 | 0.27 | 0.64 |
| LC | A2a | Cortisol | 0.88 | 0.49 | 0.79 | 0.92 | 0.69 | 0.69 | 1.29 | 0.32 | 0.64 |
| GC | A2b | 4-Thiazolidinecarboxylic acid | 1.27 | 0.23 | 0.23 | 0.90 | 0.61 | 0.61 | 1.10 | 0.69 | 0.69 |
| LC | A3a | Tyrosine | 1.10 | 0.64 | 0.89 | 1.01 | 0.96 | 0.99 | 1.06 | 0.85 | 0.96 |
| LC | A3a | LysoPE (22:6) | 0.86 | 0.52 | 0.87 | 1.08 | 0.78 | 0.99 | 0.74 | 0.34 | 0.96 |
| LC | A3a | L-Tryptophan | 1.07 | 0.77 | 0.93 | 0.92 | 0.83 | 0.99 | 1.15 | 0.74 | 0.96 |
| LC | A3a | L-Proline | 0.98 | 0.94 | 0.97 | 1.03 | 0.92 | 0.99 | 1.09 | 0.81 | 0.96 |
| LC | A3a | Alanine/Sarcosine | 1.05 | 0.89 | 0.96 | 1.37 | 0.40 | 0.99 | 0.96 | 0.92 | 0.97 |
| LC | A3a | Creatine | 1.07 | 0.62 | 0.88 | 1.27 | 0.10 | 0.99 | 0.96 | 0.83 | 0.96 |
| LC | A3a | L-Carnitine | 0.44 | < 0.05* | 0.43 | 1.18 | 0.72 | 0.99 | 1.44 | 0.55 | 0.96 |
| LC | A3a | ACar 5:0 | 0.93 | 0.66 | 0.89 | 1.11 | 0.60 | 0.99 | 0.93 | 0.78 | 0.96 |
| LC | A3a | L-Histidine | 0.71 | 0.26 | 0.84 | 0.81 | 0.52 | 0.99 | 1.12 | 0.71 | 0.96 |
| LC | A3a | 4-Aminobutanoic acid/2-Amino-2-methylpropanoic acid/3-Aminobutanoic acid | 0.72 | 0.30 | 0.84 | 1.24 | 0.49 | 0.99 | 0.78 | 0.55 | 0.96 |
| LC | A3a | Betaine | 0.81 | 0.51 | 0.87 | 0.92 | 0.84 | 0.99 | 0.93 | 0.88 | 0.96 |
| LC | A3a | LysoPC(22:5) | 1.39 | 0.15 | 0.73 | 0.86 | 0.56 | 0.99 | 0.98 | 0.93 | 0.97 |
| LC | A3a | LPC 0:0/22:6 | 1.02 | 0.92 | 0.96 | 0.96 | 0.85 | 0.99 | 0.80 | 0.41 | 0.96 |
| LC | A3a | Trimethylamine N-oxide | 1.06 | 0.59 | 0.87 | 0.98 | 0.90 | 0.99 | 0.94 | 0.71 | 0.96 |
| LC | A3a | L-Valine | 0.84 | 0.56 | 0.87 | 0.90 | 0.84 | 0.99 | 0.60 | 0.34 | 0.96 |
| LC | A3a | Hypoxanthine | 0.95 | 0.69 | 0.90 | 1.02 | 0.84 | 0.99 | 1.03 | 0.82 | 0.96 |
| LC | A3a | L-Glutamine | 1.62 | 0.17 | 0.78 | 1.05 | 0.90 | 0.99 | 1.29 | 0.58 | 0.96 |
| LC | A3a | LysoPE (18:2) | 1.71 | < 0.05* | 0.33 | 1.38 | 0.19 | 0.99 | 0.90 | 0.73 | 0.96 |
| LC | A3a | LYSOPC (18:2) | 1.37 | 0.22 | 0.81 | 1.13 | 0.69 | 0.99 | 0.73 | 0.39 | 0.96 |
| LC | A3a | LYSOPC (16:3) | 1.05 | 0.78 | 0.93 | 0.89 | 0.59 | 0.99 | 1.04 | 0.87 | 0.96 |
| LC | A3a | LYSOPE (18:1) | 1.65 | < 0.05* | 0.33 | 1.34 | 0.16 | 0.99 | 1.07 | 0.76 | 0.96 |
| LC | A3a | LysoPE (20:4) | 1.12 | 0.66 | 0.89 | 1.13 | 0.72 | 0.99 | 0.89 | 0.74 | 0.96 |
| LC | A3a | PC(16:0_20:4) | 0.94 | 0.86 | 0.96 | 0.73 | 0.32 | 0.99 | 0.79 | 0.56 | 0.96 |
| LC | A3a | 7a-hydroxy-3-oxo-4-cholestenoic acid (7-Hoca) | 0.83 | 0.49 | 0.87 | 0.85 | 0.60 | 0.99 | 2.11 | < 0.05* | 0.96 |
| LC | A3a | SM d34:2 | 1.09 | 0.74 | 0.92 | 0.69 | 0.19 | 0.99 | 0.52 | 0.07 | 0.96 |
| LC | A3a | PC (32:2) | 1.05 | 0.79 | 0.93 | 0.87 | 0.48 | 0.99 | 1.05 | 0.84 | 0.96 |
| LC | A3a | ACar 10:1 | 0.72 | < 0.05* | 0.33 | 0.96 | 0.76 | 0.99 | 0.80 | 0.26 | 0.96 |
| LC | A3a | LYSOPC (20:4) | 1.12 | 0.62 | 0.88 | 0.86 | 0.57 | 0.99 | 0.98 | 0.95 | 0.98 |
| LC | A3a | Deoxycarnitine-4-butyric acid betaine | 0.79 | 0.40 | 0.87 | 1.26 | 0.54 | 0.99 | 0.75 | 0.50 | 0.96 |
| LC | A3a | LYSOPC (16:3) | 1.13 | 0.51 | 0.87 | 0.85 | 0.36 | 0.99 | 1.05 | 0.81 | 0.96 |
| LC | A3a | PC (36:4) | 0.98 | 0.58 | 0.87 | 0.98 | 0.62 | 0.99 | 1.01 | 0.92 | 0.97 |
| LC | A3a | LysoPC(20:2) | 1.25 | 0.35 | 0.87 | 0.94 | 0.80 | 0.99 | 0.88 | 0.67 | 0.96 |
| LC | A3a | PC (36:2) | 1.11 | 0.55 | 0.87 | 0.96 | 0.81 | 0.99 | 0.96 | 0.87 | 0.96 |
| LC | A3a | N-alpha-acetyl-L-lysine | 1.45 | 0.24 | 0.81 | 1.23 | 0.53 | 0.99 | 0.84 | 0.64 | 0.96 |
| LC | A3a | LYSOPC (16:1) | 1.08 | 0.73 | 0.92 | 1.28 | 0.34 | 0.99 | 1.09 | 0.77 | 0.96 |
| LC | A3a | ACar 11:1 | 0.70 | 0.05 | 0.45 | 1.01 | 0.97 | 0.99 | 0.87 | 0.56 | 0.96 |
| LC | A3a | Serotonin | 1.04 | 0.58 | 0.87 | 1.01 | 0.88 | 0.99 | 0.93 | 0.45 | 0.96 |
| LC | A3a | Cholesterol | 0.77 | 0.33 | 0.87 | 0.51 | 0.10 | 0.99 | 1.15 | 0.78 | 0.96 |
| LC | A3a | LysoPC(P-16:0) | 1.12 | 0.62 | 0.88 | 0.82 | 0.53 | 0.99 | 0.70 | 0.30 | 0.96 |
| LC | A3a | Docosahexaenoic acid | 0.70 | < 0.05* | 0.33 | 0.95 | 0.74 | 0.99 | 0.82 | 0.25 | 0.96 |
| LC | A3a | Glycolate | 1.26 | 0.27 | 0.84 | 1.44 | 0.09 | 0.99 | 0.57 | < 0.05* | 0.96 |
| LC | A3a | Lauroylcarnitine | 0.71 | < 0.05* | 0.33 | 0.95 | 0.77 | 0.99 | 1.04 | 0.85 | 0.96 |
| LC | A3a | LYSOPC (18:2) | 1.26 | 0.13 | 0.70 | 1.07 | 0.66 | 0.99 | 0.84 | 0.30 | 0.96 |
| LC | A3a | Trigonelline | 0.86 | 0.11 | 0.64 | 1.00 | 0.99 | 0.99 | 1.09 | 0.47 | 0.96 |
| LC | A3a | L-gamma-Glutamyl-L-leucine | 1.03 | 0.87 | 0.96 | 1.03 | 0.88 | 0.99 | 0.86 | 0.47 | 0.96 |
| LC | A3a | PC (36:3) | 0.92 | 0.57 | 0.87 | 0.75 | 0.09 | 0.99 | 0.82 | 0.39 | 0.96 |
| LC | A3a | LysoPC (18:3) | 1.37 | 0.08 | 0.55 | 0.98 | 0.90 | 0.99 | 1.14 | 0.52 | 0.96 |
| LC | A3a | ACar 4:0 | 0.72 | < 0.001** | 0.07 | 1.10 | 0.34 | 0.99 | 0.94 | 0.66 | 0.96 |
| LC | A3a | PC (36:4) | 0.99 | 0.85 | 0.96 | 1.02 | 0.75 | 0.99 | 0.95 | 0.62 | 0.96 |
| LC | A3a | Plasmalogen PC(38:4) or PC(O-38:5) | 1.30 | 0.22 | 0.81 | 0.74 | 0.16 | 0.99 | 0.73 | 0.24 | 0.96 |
| LC | A3a | Taurine | 0.90 | 0.61 | 0.87 | 0.89 | 0.63 | 0.99 | 0.96 | 0.88 | 0.96 |
| LC | A3a | PC (38:5) | 1.41 | 0.21 | 0.81 | 1.02 | 0.96 | 0.99 | 0.76 | 0.44 | 0.96 |
| LC | A3a | LysoPE (18:2) | 1.21 | 0.16 | 0.73 | 1.13 | 0.36 | 0.99 | 0.78 | 0.14 | 0.96 |
| LC | A3a | 4-Guanidinobutanoic acid | 1.36 | < 0.05* | 0.33 | 1.46 | < 0.05* | 0.99 | 0.88 | 0.38 | 0.96 |
| LC | A3a | ACar 10:1 | 0.71 | < 0.05* | 0.33 | 0.97 | 0.87 | 0.99 | 0.88 | 0.51 | 0.96 |
| LC | A3a | LYSOPC (16:1) | 1.08 | 0.58 | 0.87 | 1.41 | < 0.05* | 0.99 | 1.04 | 0.83 | 0.96 |
| LC | A3a | ACar 9:0 | 1.00 | 0.98 | 0.98 | 1.11 | 0.50 | 0.99 | 1.05 | 0.78 | 0.96 |
| LC | A3a | Plasmalogen PC(36:4) or PC(O-36:5) | 1.16 | 0.52 | 0.87 | 0.91 | 0.67 | 0.99 | 0.80 | 0.42 | 0.96 |
| LC | A3a | Uric acid | 0.84 | 0.42 | 0.87 | 1.03 | 0.91 | 0.99 | 0.88 | 0.69 | 0.96 |
| LC | A3a | LYSOPC (18:0) | 1.53 | 0.11 | 0.64 | 0.83 | 0.50 | 0.99 | 1.06 | 0.84 | 0.96 |
| LC | A3a | LYSOPC (16:1) | 1.06 | 0.48 | 0.87 | 0.83 | < 0.05* | 0.99 | 0.85 | 0.13 | 0.96 |
| LC | A3a | Kynurenine | 0.90 | 0.40 | 0.87 | 0.95 | 0.74 | 0.99 | 1.07 | 0.69 | 0.96 |
| LC | A3a | 5-Valerolactone | 1.16 | 0.24 | 0.81 | 1.07 | 0.62 | 0.99 | 0.95 | 0.72 | 0.96 |
| LC | A3a | ACar 4:0 | 1.31 | < 0.05* | 0.33 | 0.95 | 0.67 | 0.99 | 0.93 | 0.63 | 0.96 |
| LC | A3a | Acetylcarnitine (C2) | 0.78 | 0.24 | 0.81 | 1.29 | 0.34 | 0.99 | 0.93 | 0.82 | 0.96 |
| LC | A3a | LysoPC (16:0) | 0.52 | 0.30 | 0.84 | 0.83 | 0.83 | 0.99 | 0.29 | 0.19 | 0.96 |
| LC | A3a | ACar 8:0 | 0.75 | < 0.05* | 0.43 | 0.95 | 0.69 | 0.99 | 1.02 | 0.92 | 0.97 |
| LC | A3a | Linoleic acid | 0.87 | 0.22 | 0.81 | 1.03 | 0.80 | 0.99 | 0.85 | 0.30 | 0.96 |
| LC | A3a | LYSOPC (16:1) | 0.96 | 0.47 | 0.87 | 1.16 | < 0.05* | 0.99 | 1.11 | 0.15 | 0.96 |
| LC | A3a | 5,6-Dihydrouracil | 1.36 | 0.47 | 0.87 | 0.94 | 0.90 | 0.99 | 4.36 | < 0.05* | 0.96 |
| LC | A3a | Decanoyl-L-Carnitine (C10:0) | 0.76 | < 0.05* | 0.44 | 0.93 | 0.61 | 0.99 | 1.04 | 0.83 | 0.96 |
| LC | A3a | L-Serine | 0.89 | 0.65 | 0.89 | 1.05 | 0.87 | 0.99 | 1.07 | 0.83 | 0.96 |
| LC | A3a | 1-methylnicotinamide | 0.87 | 0.28 | 0.84 | 1.09 | 0.57 | 0.99 | 1.11 | 0.61 | 0.96 |
| LC | A3a | LYSOPC (14:0) | 1.18 | 0.52 | 0.87 | 0.90 | 0.68 | 0.99 | 1.04 | 0.89 | 0.96 |
| LC | A3a | ACar 8:1 | 0.83 | 0.15 | 0.73 | 0.89 | 0.42 | 0.99 | 0.88 | 0.47 | 0.96 |
| LC | A3a | LYSOPC (20:4) | 1.05 | 0.88 | 0.96 | 0.99 | 0.99 | 0.99 | 1.75 | 0.34 | 0.96 |
| LC | A3a | LYSOPC (18:0) | 1.25 | 0.43 | 0.87 | 0.68 | 0.24 | 0.99 | 0.80 | 0.51 | 0.96 |
| LC | A3a | L-Methionine | 1.02 | 0.91 | 0.96 | 0.99 | 0.98 | 0.99 | 0.84 | 0.59 | 0.96 |
| LC | A3a | Propionyl-L-carnitine (C3:0) | 0.91 | 0.68 | 0.90 | 1.27 | 0.30 | 0.99 | 0.87 | 0.65 | 0.96 |
| LC | A3a | PC (34:3) | 1.03 | 0.73 | 0.92 | 0.87 | 0.12 | 0.99 | 1.06 | 0.63 | 0.96 |
| LC | A3a | L-Tyrosine | 1.07 | 0.76 | 0.93 | 0.93 | 0.81 | 0.99 | 0.84 | 0.61 | 0.96 |
| LC | A3a | PC 38:6 | 0.95 | 0.81 | 0.95 | 0.91 | 0.68 | 0.99 | 0.80 | 0.39 | 0.96 |
| LC | A3a | ACar 18:1 | 0.94 | 0.79 | 0.93 | 1.16 | 0.53 | 0.99 | 0.80 | 0.43 | 0.96 |
| LC | A3a | LYSOPC (17:0) | 1.50 | 0.08 | 0.55 | 0.63 | 0.10 | 0.99 | 0.80 | 0.47 | 0.96 |
| LC | A3a | Theophylline | 0.78 | < 0.05* | 0.33 | 0.94 | 0.58 | 0.99 | 0.98 | 0.88 | 0.96 |
| LC | A3a | Glycocholic acid | 1.19 | 0.20 | 0.81 | 1.06 | 0.68 | 0.99 | 0.87 | 0.44 | 0.96 |
| LC | A3a | PC (34:1) | 1.04 | 0.79 | 0.93 | 1.07 | 0.70 | 0.99 | 1.00 | 0.99 | 0.99 |
| LC | A3a | PC (30:0) | 1.11 | 0.50 | 0.87 | 0.98 | 0.89 | 0.99 | 0.93 | 0.71 | 0.96 |
| LC | A3a | PC (38:3) | 0.99 | 0.90 | 0.96 | 1.00 | 0.97 | 0.99 | 0.94 | 0.56 | 0.96 |
| LC | A3a | 3,5-Tetradecadiencarnitine (C14:2) | 0.72 | < 0.05* | 0.33 | 1.04 | 0.78 | 0.99 | 0.86 | 0.43 | 0.96 |
| LC | A3a | Indolelactic acid | 0.94 | 0.83 | 0.96 | 1.52 | 0.15 | 0.99 | 1.31 | 0.40 | 0.96 |
| LC | A3a | Dehydroepiandrosterone | 1.08 | 0.74 | 0.92 | 0.68 | 0.18 | 0.99 | 1.34 | 0.40 | 0.96 |
| LC | A3a | Paraxanthine | 0.82 | < 0.05* | 0.33 | 0.96 | 0.66 | 0.99 | 1.03 | 0.80 | 0.96 |
| LC | A3a | LYSOPC (20:3) | 1.19 | 0.46 | 0.87 | 0.77 | 0.31 | 0.99 | 0.72 | 0.23 | 0.96 |
| LC | A3a | Uracil | 1.01 | 0.96 | 0.98 | 0.90 | 0.66 | 0.99 | 1.17 | 0.57 | 0.96 |
| LC | A3a | Threonic acid | 0.99 | 0.91 | 0.96 | 1.05 | 0.64 | 0.99 | 1.04 | 0.74 | 0.96 |
| LC | A3a | LYSOPC (20:3) | 1.26 | 0.33 | 0.87 | 1.00 | 0.99 | 0.99 | 1.27 | 0.49 | 0.96 |
| LC | A3a | Retinol | 1.34 | 0.14 | 0.70 | 0.84 | 0.36 | 0.99 | 1.32 | 0.27 | 0.96 |
| LC | A3a | LYSOPC (20:3) | 0.94 | 0.58 | 0.87 | 1.05 | 0.63 | 0.99 | 0.97 | 0.78 | 0.96 |
| LC | A3a | PC (34:2) | 1.31 | 0.43 | 0.87 | 1.12 | 0.76 | 0.99 | 0.67 | 0.35 | 0.96 |
| LC | A3a | LYSOPC (18:4) | 1.14 | 0.53 | 0.87 | 1.37 | 0.20 | 0.99 | 1.08 | 0.76 | 0.96 |
| LC | A3a | PC (36:4) | 1.67 | < 0.05* | 0.33 | 0.96 | 0.83 | 0.99 | 0.79 | 0.32 | 0.96 |
| LC | A3a | Nepsilon, nepsilon, nepsilon-trimethyllysine | 1.17 | 0.44 | 0.87 | 1.28 | 0.28 | 0.99 | 1.14 | 0.64 | 0.96 |
| LC | A3a | PC(32:1) | 1.03 | 0.85 | 0.96 | 1.17 | 0.30 | 0.99 | 1.12 | 0.54 | 0.96 |
| LC | A3a | L-Threonine/L-Homoserine | 0.79 | 0.35 | 0.87 | 0.74 | 0.26 | 0.99 | 0.90 | 0.74 | 0.96 |
| LC | A3a | PC (40:5) | 1.00 | 0.97 | 0.98 | 1.04 | 0.72 | 0.99 | 0.98 | 0.89 | 0.96 |
| LC | A3a | Theobromine | 0.92 | 0.38 | 0.87 | 0.85 | 0.11 | 0.99 | 1.02 | 0.90 | 0.96 |
| LC | A3a | Phe-Phe | 0.91 | 0.23 | 0.81 | 1.03 | 0.69 | 0.99 | 0.95 | 0.56 | 0.96 |
| LC | A3a | Creatinine | 0.96 | 0.89 | 0.96 | 0.98 | 0.95 | 0.99 | 0.70 | 0.47 | 0.96 |
| LC | A3a | Tyrosine | 1.15 | 0.57 | 0.87 | 1.01 | 0.96 | 0.99 | 1.10 | 0.75 | 0.96 |
| LC | A3a | Undecanoylcarnitine | 0.74 | 0.14 | 0.70 | 0.95 | 0.82 | 0.99 | 1.22 | 0.40 | 0.96 |
| LC | A3a | Plasmalogen PC(36:3) or PC(O-36:4) | 1.21 | 0.31 | 0.86 | 0.90 | 0.60 | 0.99 | 0.85 | 0.49 | 0.96 |
| LC | A3a | Diacylglycerol (C34:2) | 1.01 | 0.90 | 0.96 | 1.11 | 0.32 | 0.99 | 1.02 | 0.86 | 0.96 |
| LC | A3a | Dimethylsulfone | 0.88 | 0.55 | 0.87 | 1.01 | 0.95 | 0.99 | 0.99 | 0.98 | 0.99 |
| LC | A3a | Indole-3-propionic acid (IPA) | 1.14 | 0.30 | 0.84 | 0.93 | 0.56 | 0.99 | 0.87 | 0.36 | 0.96 |
| LC | A3a | PC (36:3) | 0.99 | 0.94 | 0.97 | 0.99 | 0.94 | 0.99 | 0.77 | 0.31 | 0.96 |
| LC | A3a | ACar 5:0 | 1.26 | 0.27 | 0.84 | 1.12 | 0.59 | 0.99 | 1.06 | 0.83 | 0.96 |
| LC | A3a | Glycodeoxycholic acid | 0.94 | 0.37 | 0.87 | 0.93 | 0.33 | 0.99 | 0.95 | 0.54 | 0.96 |
| LC | A3a | PC (40:6) | 0.95 | 0.69 | 0.90 | 0.98 | 0.87 | 0.99 | 0.80 | 0.22 | 0.96 |
| LC | A3a | Tetradecanoylcarnitine (C14:0) | 0.88 | 0.60 | 0.87 | 1.04 | 0.89 | 0.99 | 0.96 | 0.88 | 0.96 |
| GC | A3a | 3-Hydroxy-dithiobutanoic acid | 1.23 | 0.41 | 0.87 | 1.04 | 0.89 | 0.99 | 1.15 | 0.70 | 0.96 |
| GC | A3a | Tetradecanoic acid | 0.80 | 0.06 | 0.51 | 1.00 | 0.99 | 0.99 | 0.95 | 0.73 | 0.96 |
| GC | A3a | (9E,12E)-octadeca-9,12-dienoic acid | 0.71 | 0.09 | 0.59 | 1.03 | 0.86 | 0.99 | 1.01 | 0.97 | 0.98 |
| GC | A3a | Indole | 1.18 | 0.60 | 0.87 | 1.43 | 0.24 | 0.99 | 1.73 | 0.16 | 0.96 |
| GC | A3a | Skatole | 0.73 | 0.11 | 0.64 | 1.03 | 0.87 | 0.99 | 0.88 | 0.63 | 0.96 |
| GC | A3a | (5S)-5-ethyl-5-[(1R)-1-hydroxyethyl]-2-oxolanone | 0.83 | 0.58 | 0.87 | 1.27 | 0.45 | 0.99 | 0.78 | 0.58 | 0.96 |
| GC | A3a | 8-Hydroxy-4Z,9-decadienoic acid | 1.15 | 0.48 | 0.87 | 1.16 | 0.48 | 0.99 | 0.90 | 0.68 | 0.96 |
| GC | A3a | 2-Hydroxy-4-phenyl-butyric acid ethyl ester | 1.12 | 0.57 | 0.87 | 0.77 | 0.22 | 0.99 | 0.92 | 0.78 | 0.96 |
| GC | A3a | 4-oxo-6,7,8,9-tetrahydro-pyrido[1,2-a]pyrimidine | 1.10 | 0.64 | 0.89 | 0.87 | 0.50 | 0.99 | 1.02 | 0.94 | 0.97 |
| GC | A3a | 3-[(2S)-2-pyrrolidin-1-iumyl]propanoate | 1.25 | 0.53 | 0.87 | 1.61 | 0.18 | 0.99 | 1.23 | 0.62 | 0.96 |
| GC | A3a | .beta.-apo-8'-carotenal | 1.05 | 0.58 | 0.87 | 1.00 | 0.97 | 0.99 | 1.00 | 0.96 | 0.98 |
| GC | A3a | 3-methyl-2-pentyl-1H-pyrrole | 0.82 | 0.19 | 0.81 | 1.00 | 0.98 | 0.99 | 1.04 | 0.84 | 0.96 |
| GC | A3a | methyl 2-(1,3-dimethyl-2-oxoimidazolidin-4-yl)acetate | 0.96 | 0.74 | 0.92 | 1.01 | 0.94 | 0.99 | 0.97 | 0.90 | 0.96 |
| GC | A3a | 2-phenylacetic acid | 1.06 | 0.83 | 0.96 | 0.92 | 0.75 | 0.99 | 0.90 | 0.77 | 0.96 |
| GC | A3a | 6-[6'-Hydroxy-1'-hexyn-1'-yl]-1H-purine | 1.79 | 0.52 | 0.87 | 2.15 | 0.38 | 0.99 | 0.18 | 0.11 | 0.96 |
| GC | A3a | 5,6-Dioxoheptanoic acid | 1.33 | 0.21 | 0.81 | 1.46 | 0.06 | 0.99 | 0.71 | 0.21 | 0.96 |
| GC | A3a | 2-amino-7-methyl-3H-purin-6-one | 1.22 | 0.61 | 0.87 | 0.78 | 0.48 | 0.99 | 0.62 | 0.31 | 0.96 |
| GC | A3a | Decanoic acid | 1.01 | 0.97 | 0.98 | 1.28 | 0.16 | 0.99 | 0.86 | 0.49 | 0.96 |
| GC | A3a | (S)-2-Ethyl-5-oxotetrahydrofuran-2-carboxylic acid | 0.68 | 0.25 | 0.83 | 1.67 | 0.15 | 0.99 | 1.16 | 0.75 | 0.96 |
| GC | A3a | 2-Ethylhexanoic acid | 1.11 | 0.73 | 0.92 | 1.46 | 0.25 | 0.99 | 0.86 | 0.70 | 0.96 |
| GC | A3a | (2S)-2-Amino-3-(1H-2-indolyl)propionic acid (L-isotrytophan) | 1.18 | 0.28 | 0.84 | 0.82 | 0.24 | 0.99 | 0.85 | 0.45 | 0.96 |
| GC | A3a | 12-Oxooctadec-9-enoic acid | 1.00 | 0.97 | 0.98 | 1.05 | 0.61 | 0.99 | 1.14 | 0.22 | 0.96 |
| GC | A3a | (E)-2-phenyl-3-(2-pyridinyl)-2-propenoic acid | 1.21 | < 0.05* | 0.33 | 0.84 | 0.06 | 0.99 | 1.18 | 0.16 | 0.96 |
| GC | A3a | 1,5-Anhydro-D-fructose | 0.95 | 0.51 | 0.87 | 0.99 | 0.89 | 0.99 | 1.08 | 0.45 | 0.96 |
| GC | A3a | (E)-6-hydroxyhept-4-enoic acid | 1.06 | 0.55 | 0.87 | 1.14 | 0.19 | 0.99 | 0.81 | 0.11 | 0.96 |
| GC | A3a | 1,5-Anhydro-2-deoxy-2-(methoximino)-D-arabino-hexitol | 0.95 | 0.28 | 0.84 | 0.96 | 0.33 | 0.99 | 1.10 | 0.10 | 0.96 |
| GC | A3a | 2-(Aminomethyl)-5,5-dimethyl-1,3-thiazolidine-4-carboxylic acid | 0.91 | 0.08 | 0.55 | 1.07 | 0.24 | 0.99 | 1.04 | 0.56 | 0.96 |
| GC | A3a | 11,15'-Dihydrooxepin-.beta.,.beta.-carotene | 1.02 | 0.58 | 0.87 | 1.02 | 0.64 | 0.99 | 0.98 | 0.62 | 0.96 |
| GC | A3a | (S)-5-Methyl-2-(dimethylamino)-1H-imidazol-4(5H)-one | 1.03 | 0.53 | 0.87 | 0.96 | 0.33 | 0.99 | 1.02 | 0.72 | 0.96 |
| GC | A3a | 4,5-Secocholest-8-en-3-one, 5-ethylidene-14-methyl- | 1.02 | 0.86 | 0.96 | 1.16 | 0.18 | 0.99 | 0.95 | 0.72 | 0.96 |
| LC | A3b | LYSOPC (20:3) | 0.74 | 0.27 | 0.57 | 0.97 | 0.91 | 0.91 | 0.86 | 0.69 | 0.81 |
| LC | A3b | Cotinine secondary peak | 0.67 | 0.09 | 0.55 | 1.19 | 0.42 | 0.91 | 1.76 | 0.07 | 0.23 |
| LC | A3b | 2-Hydroxyphenylacetic acid | 0.95 | 0.80 | 0.94 | 1.03 | 0.89 | 0.91 | 1.10 | 0.69 | 0.81 |
| LC | A3b | PC (36:2) | 0.99 | 0.94 | 0.94 | 0.93 | 0.70 | 0.91 | 1.23 | 0.35 | 0.61 |
| LC | A3b | LYSOPC (20:4) | 1.19 | 0.33 | 0.57 | 0.98 | 0.91 | 0.91 | 1.42 | 0.12 | 0.28 |
| LC | A3b | Cotinine | 1.82 | 0.16 | 0.55 | 1.94 | 0.06 | 0.45 | 2.17 | < 0.05* | 0.21 |
| GC | A3b | Guadiscoline | 0.98 | 0.90 | 0.94 | 1.04 | 0.84 | 0.91 | 0.96 | 0.88 | 0.88 |

**Table S6:**
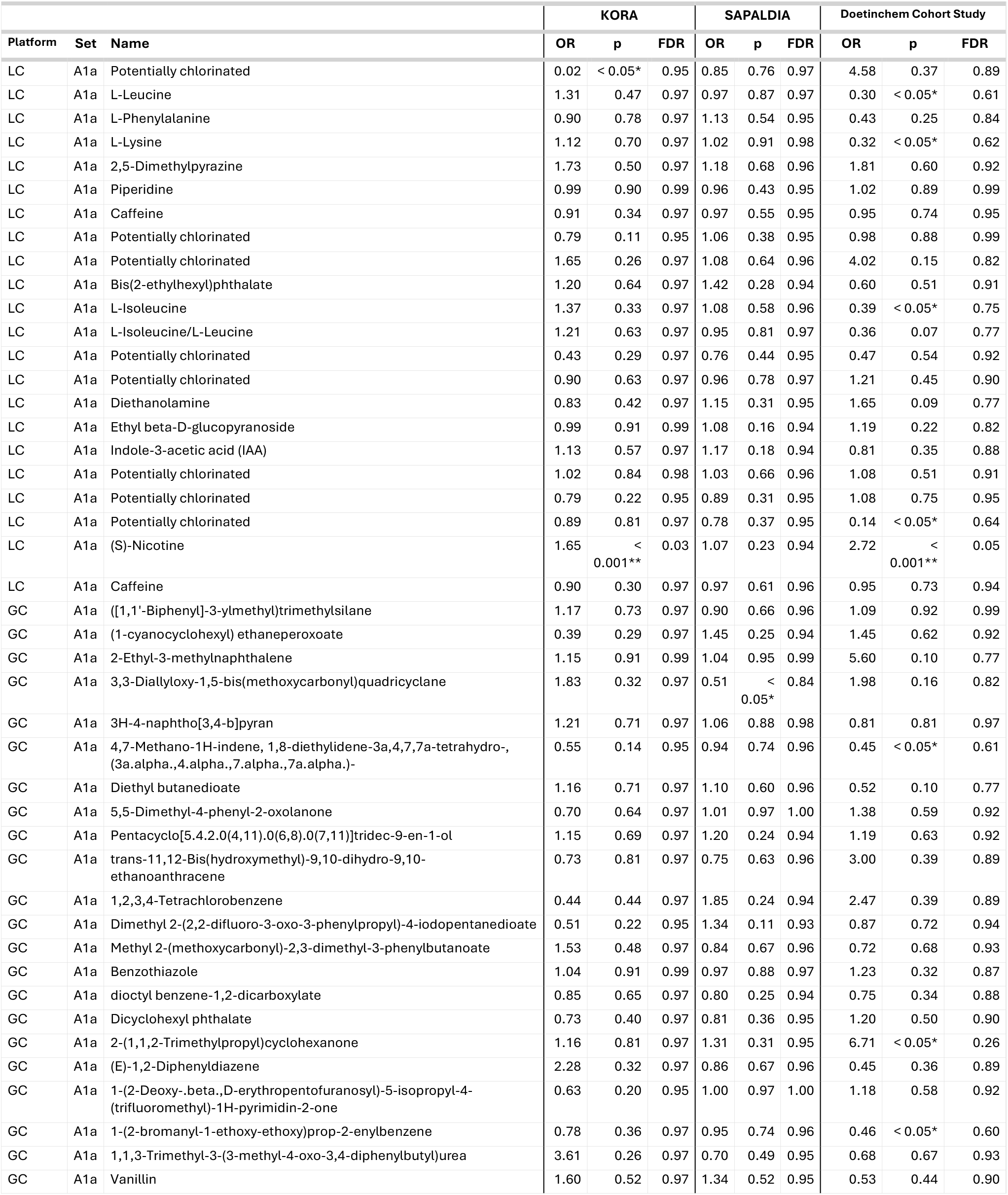
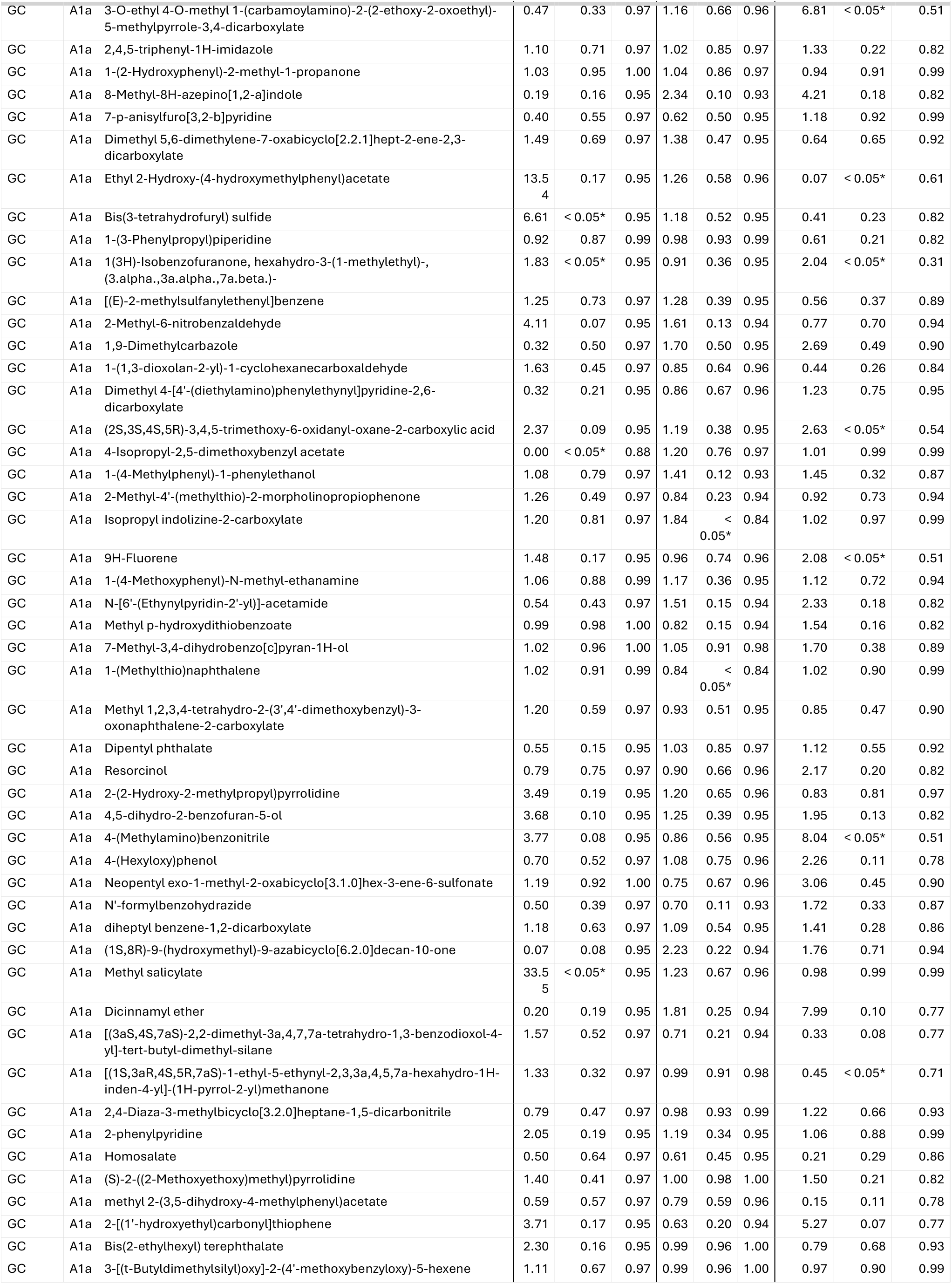
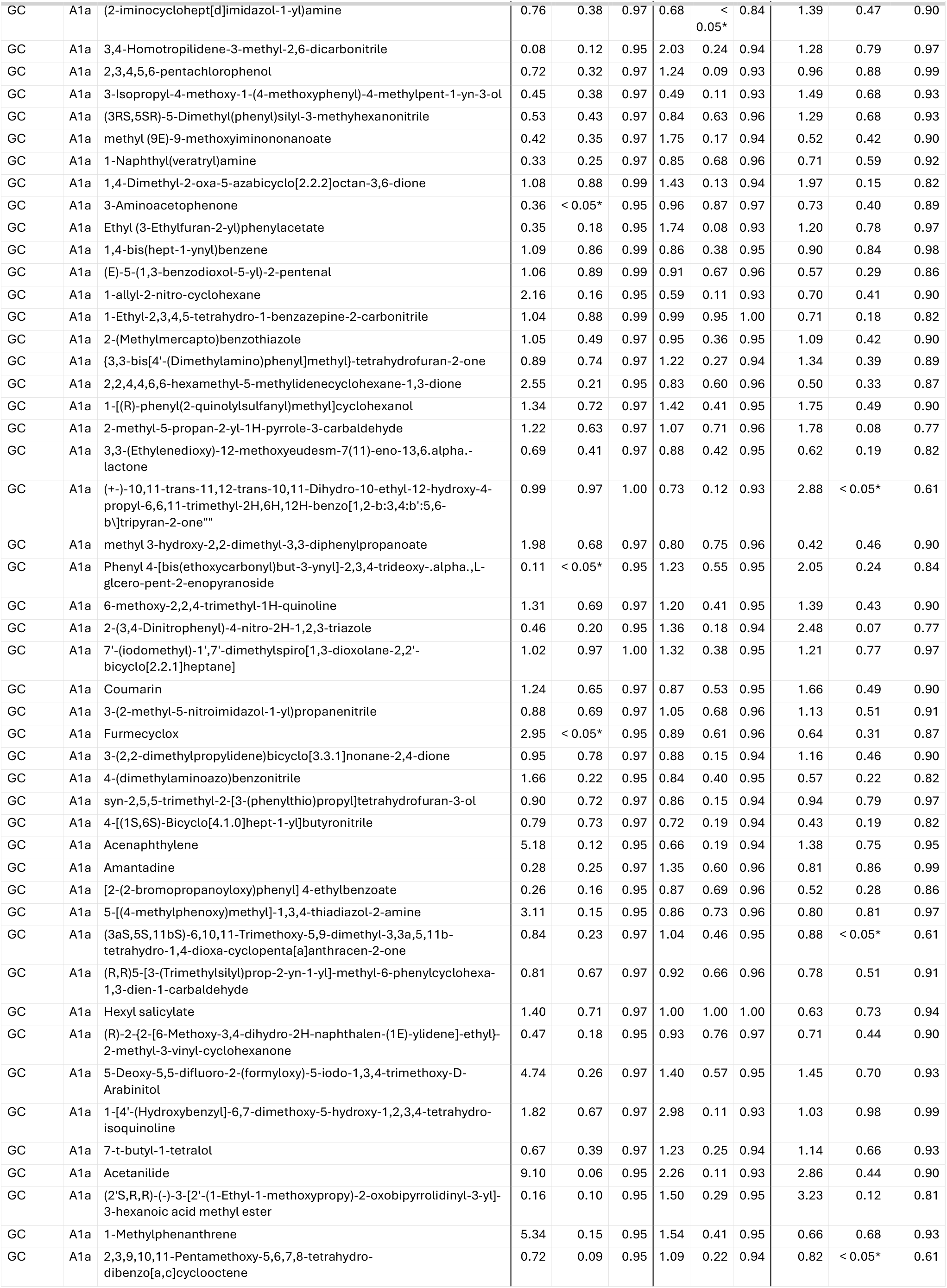
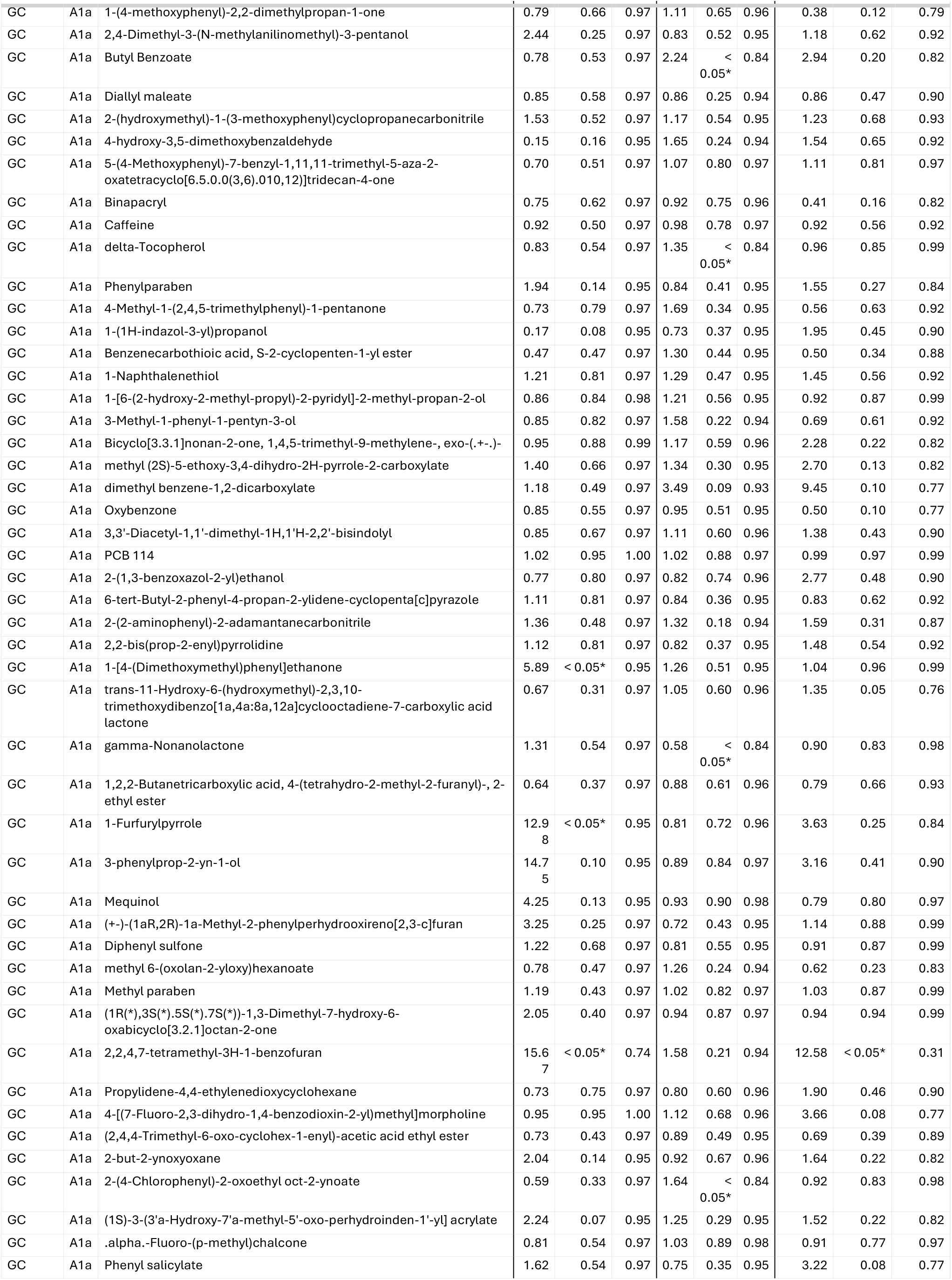
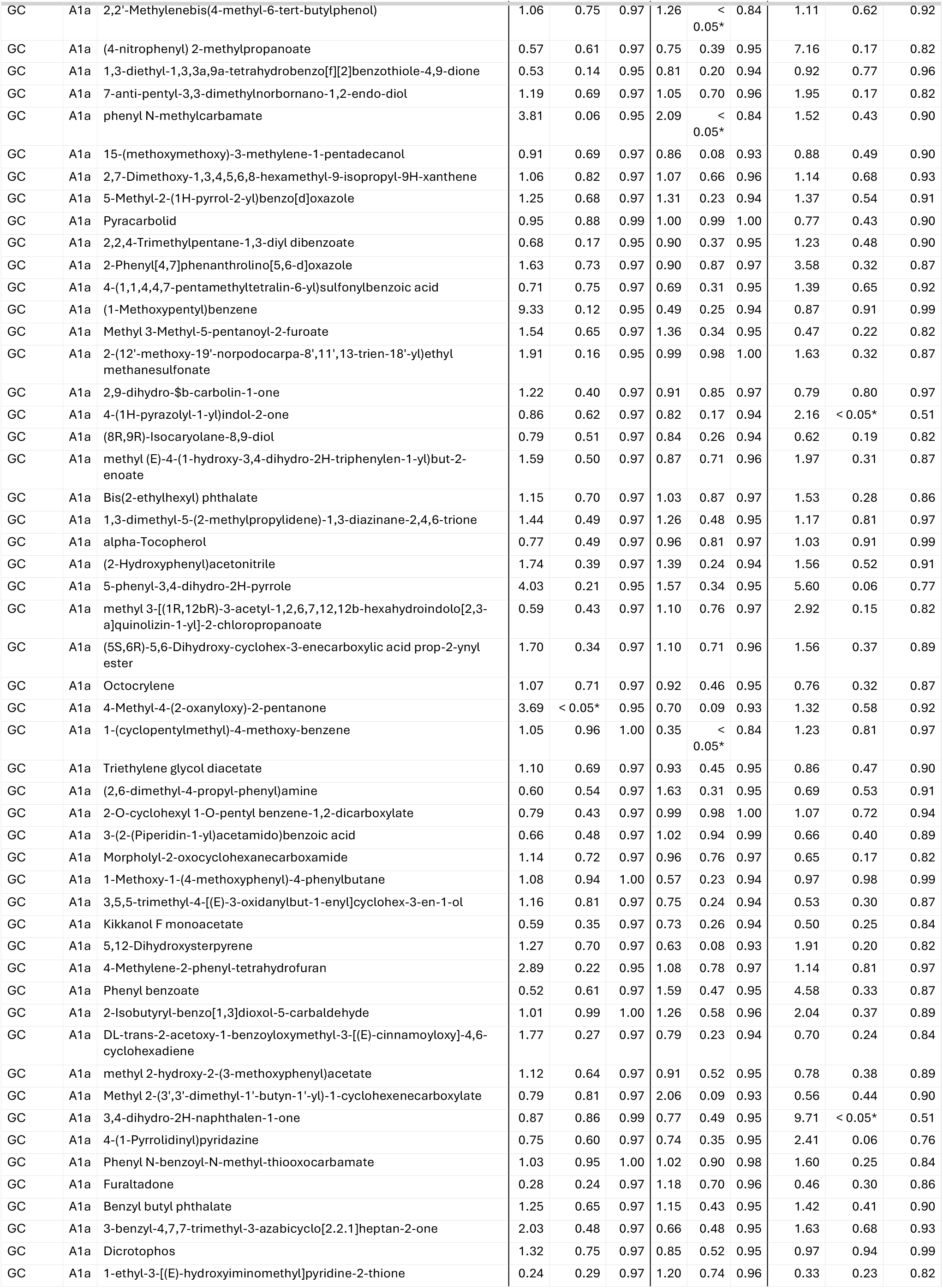
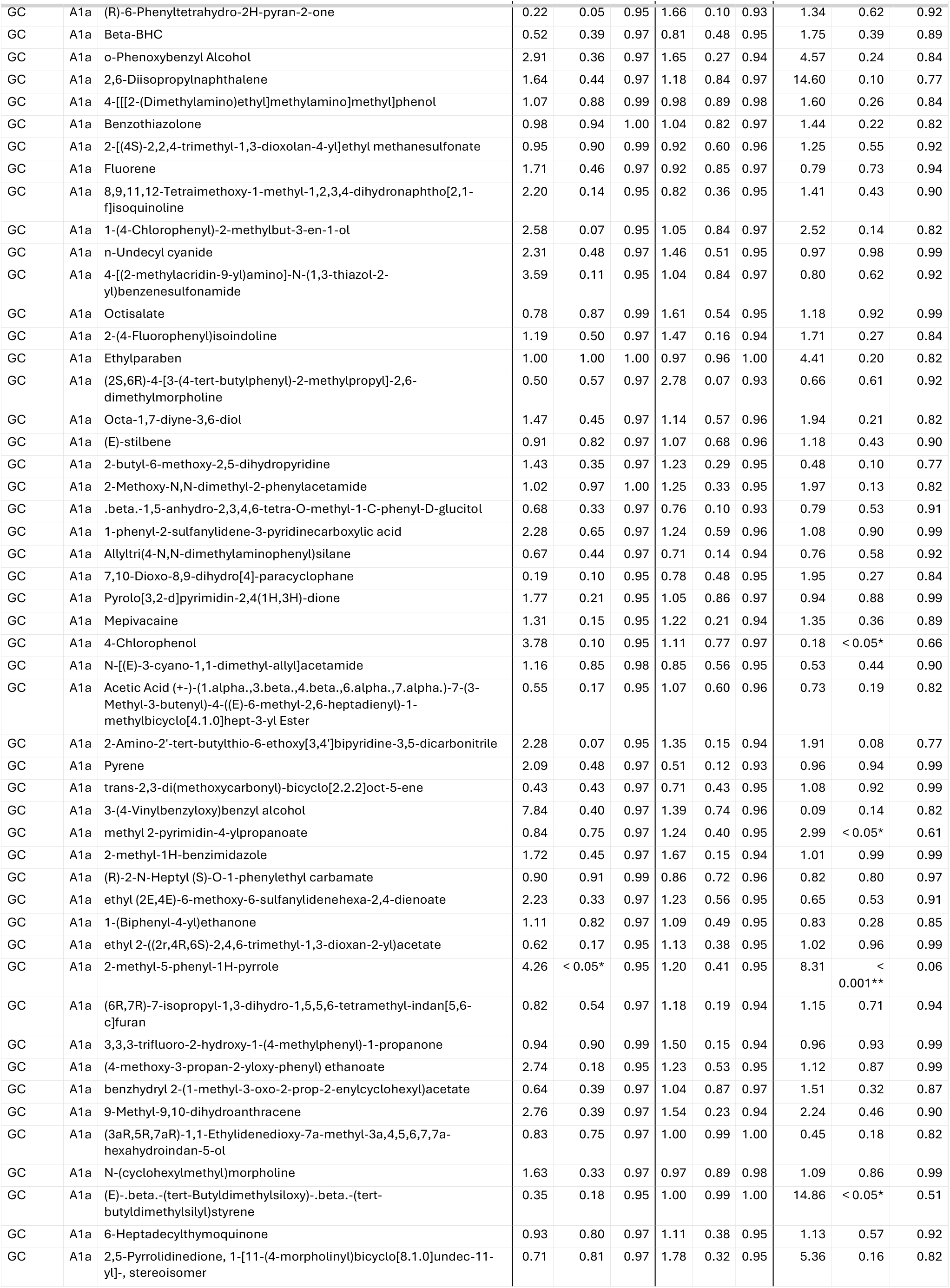
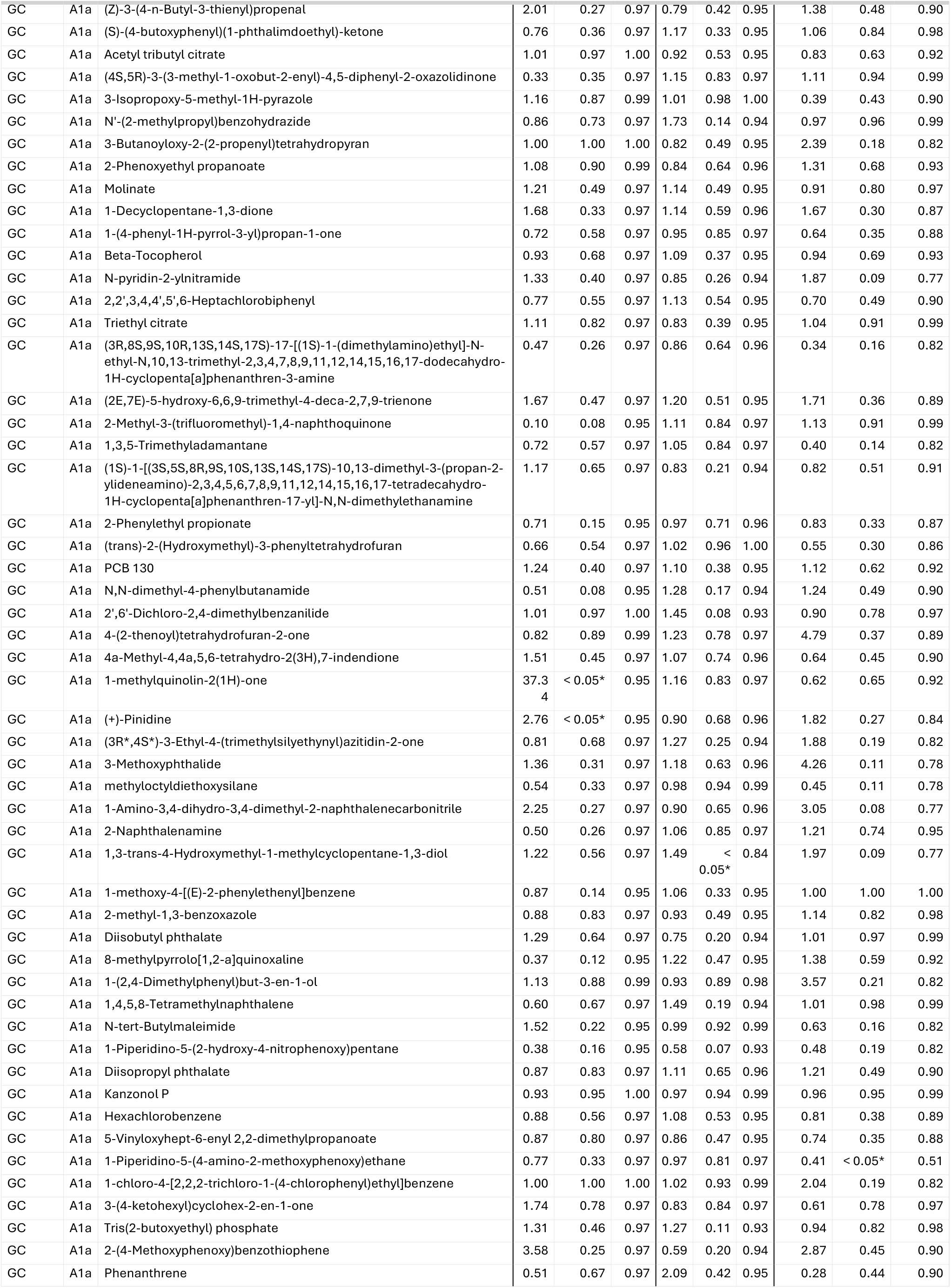
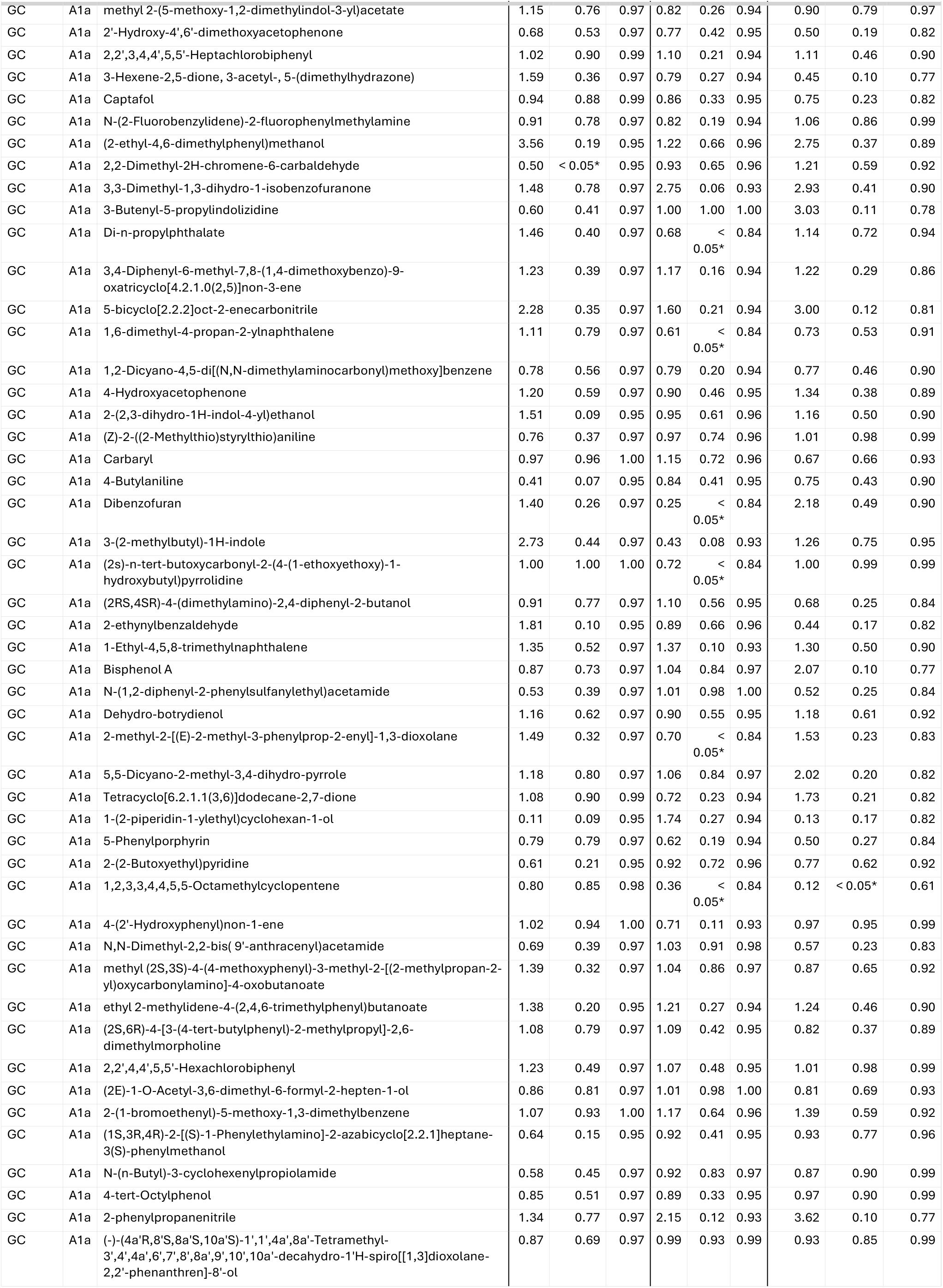
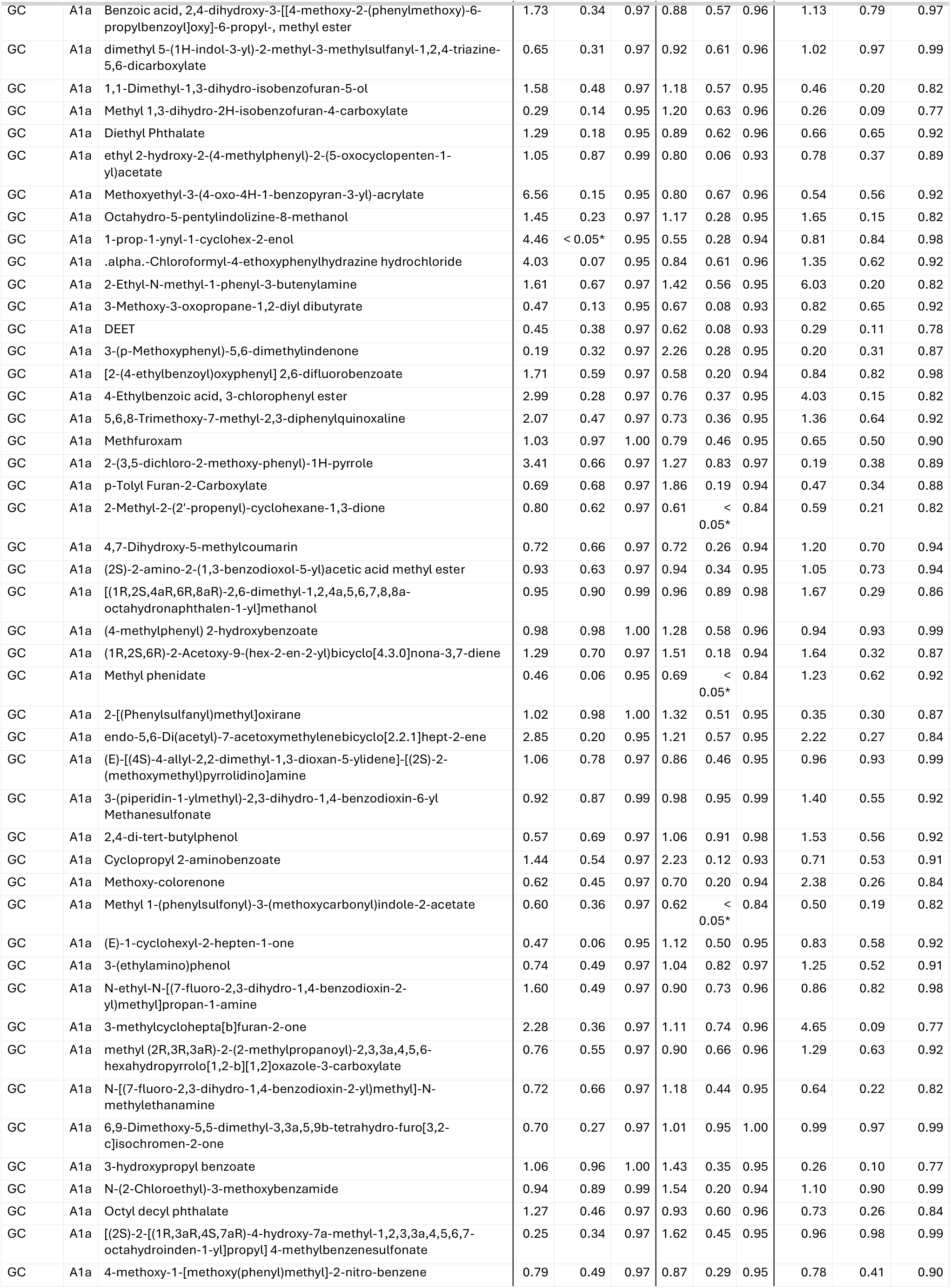
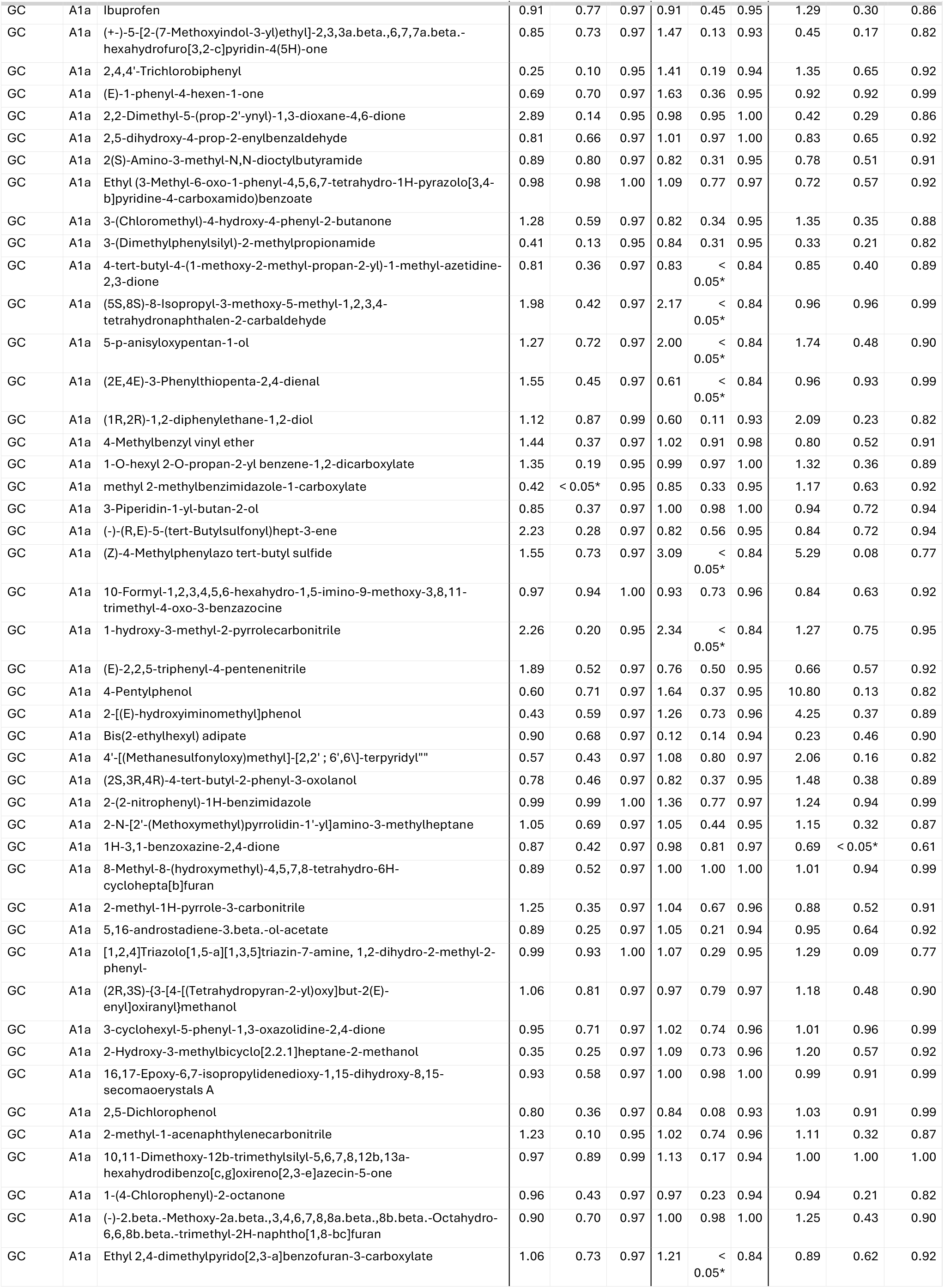
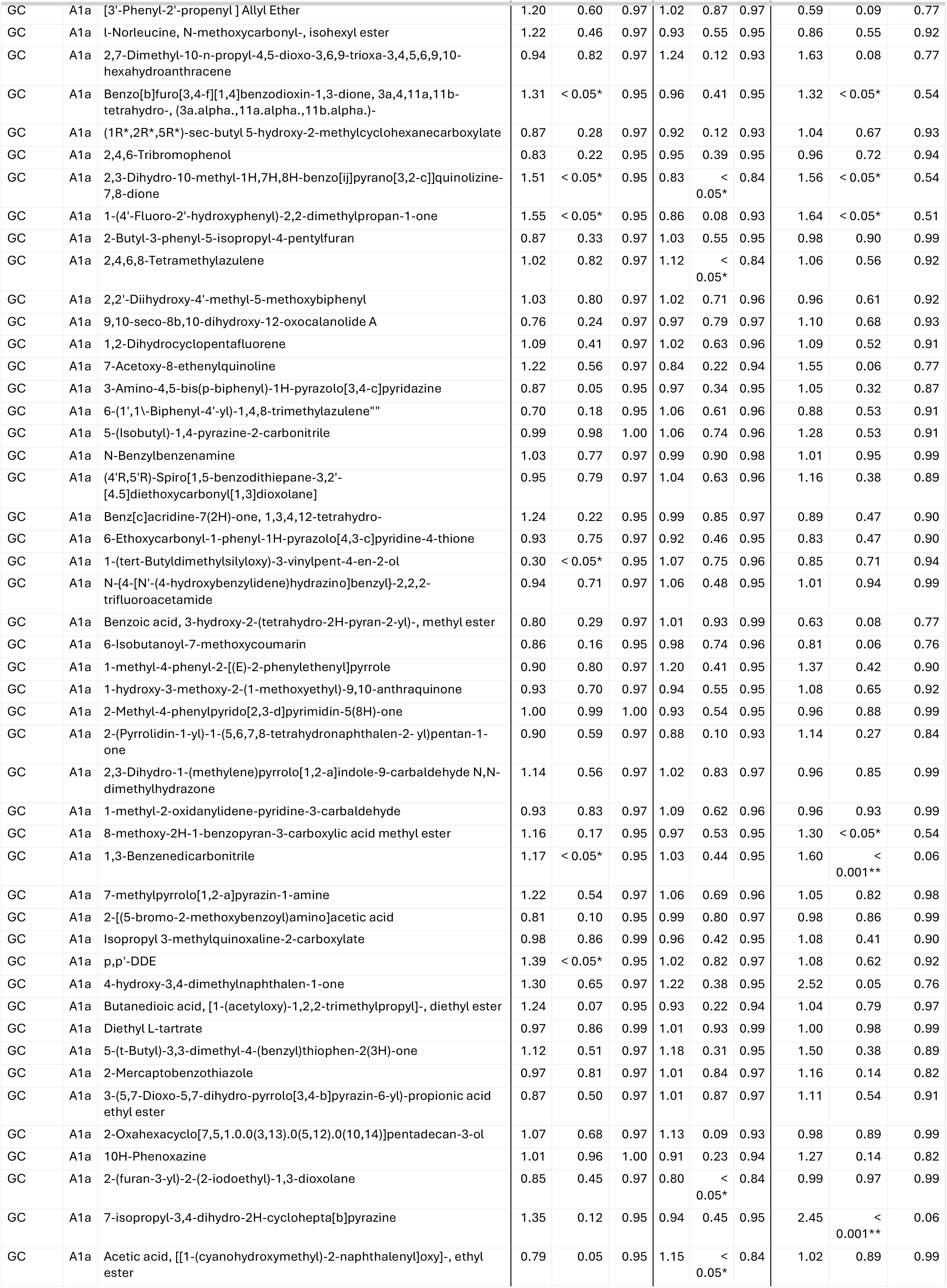
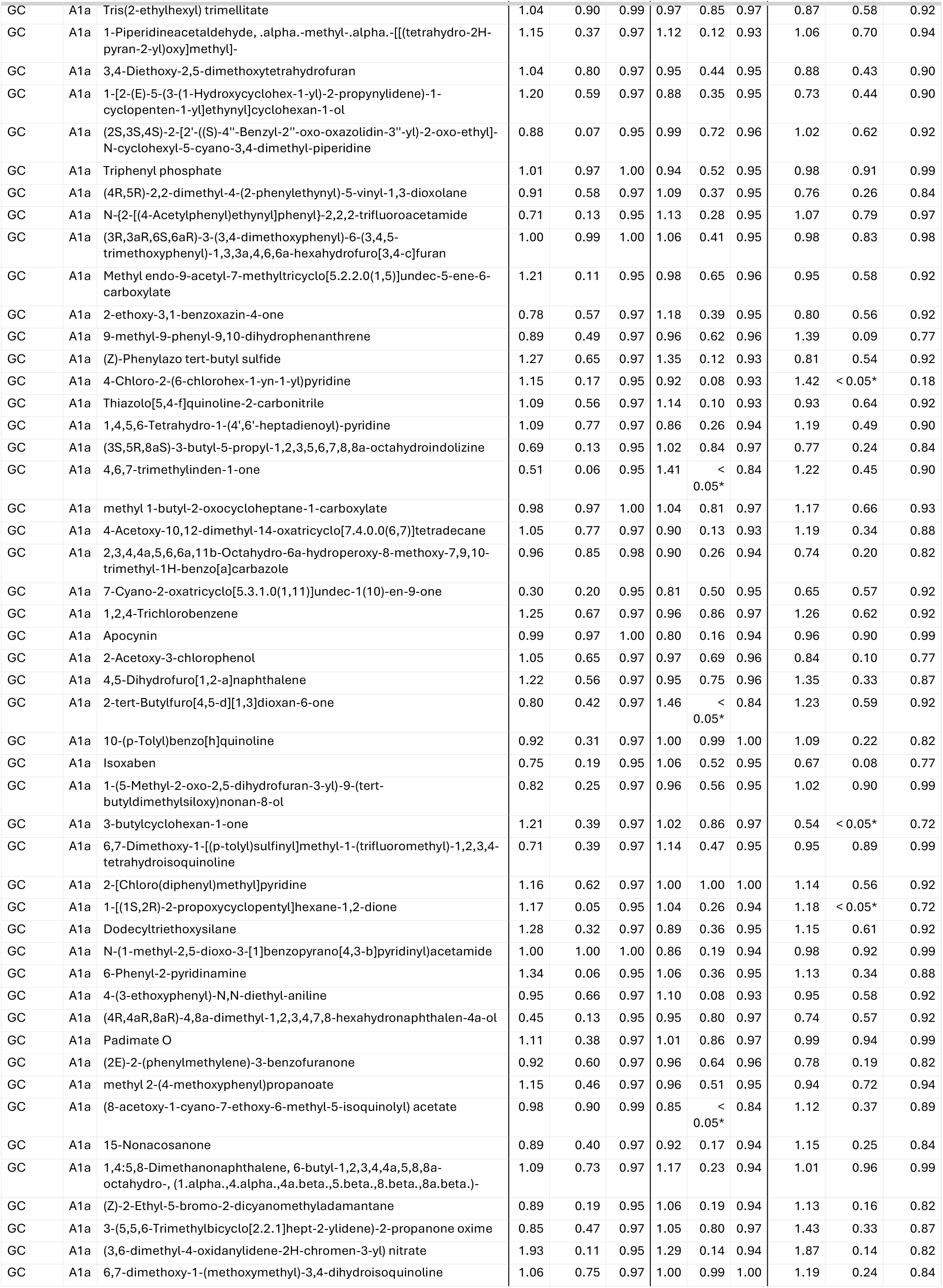
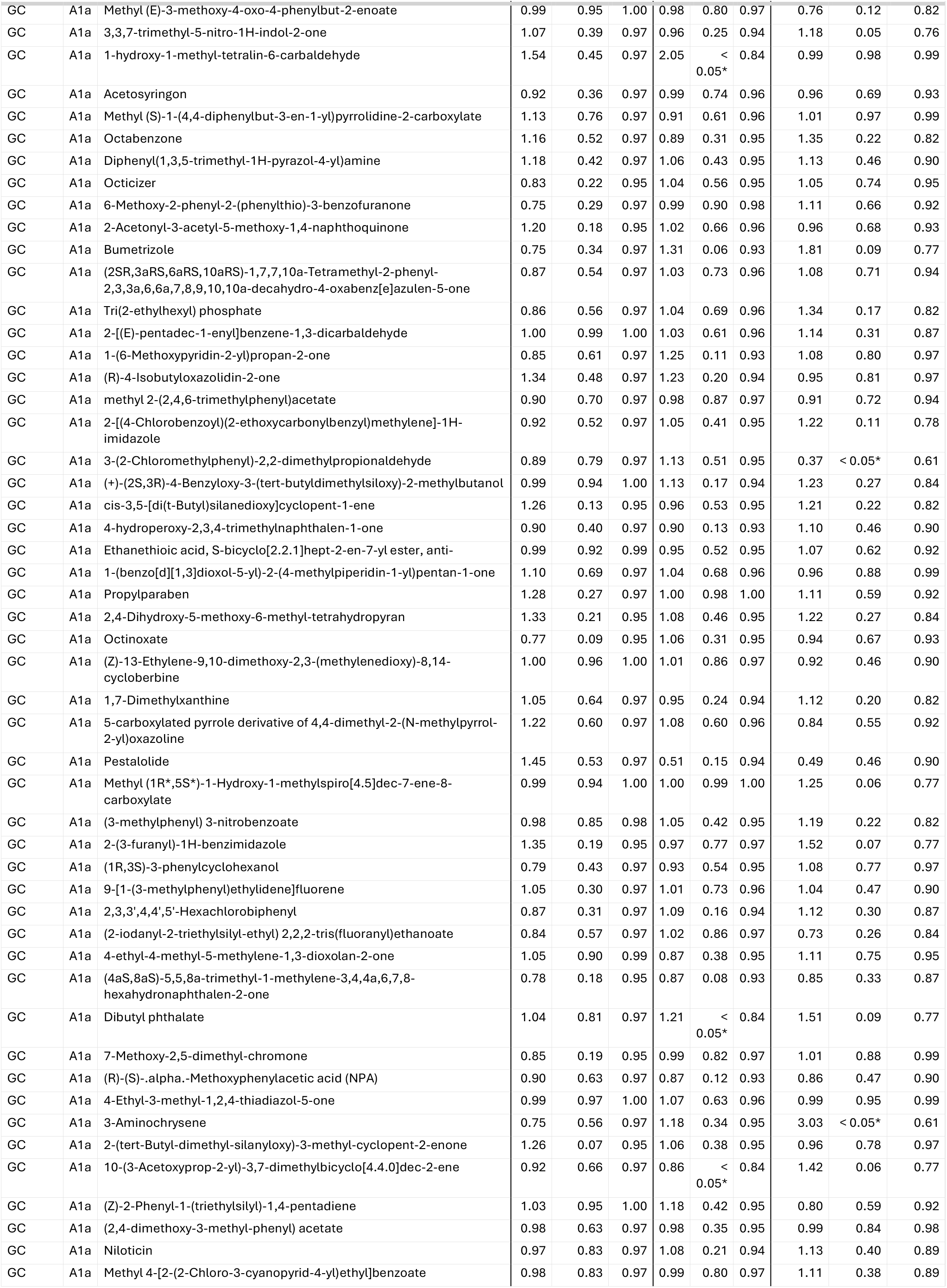
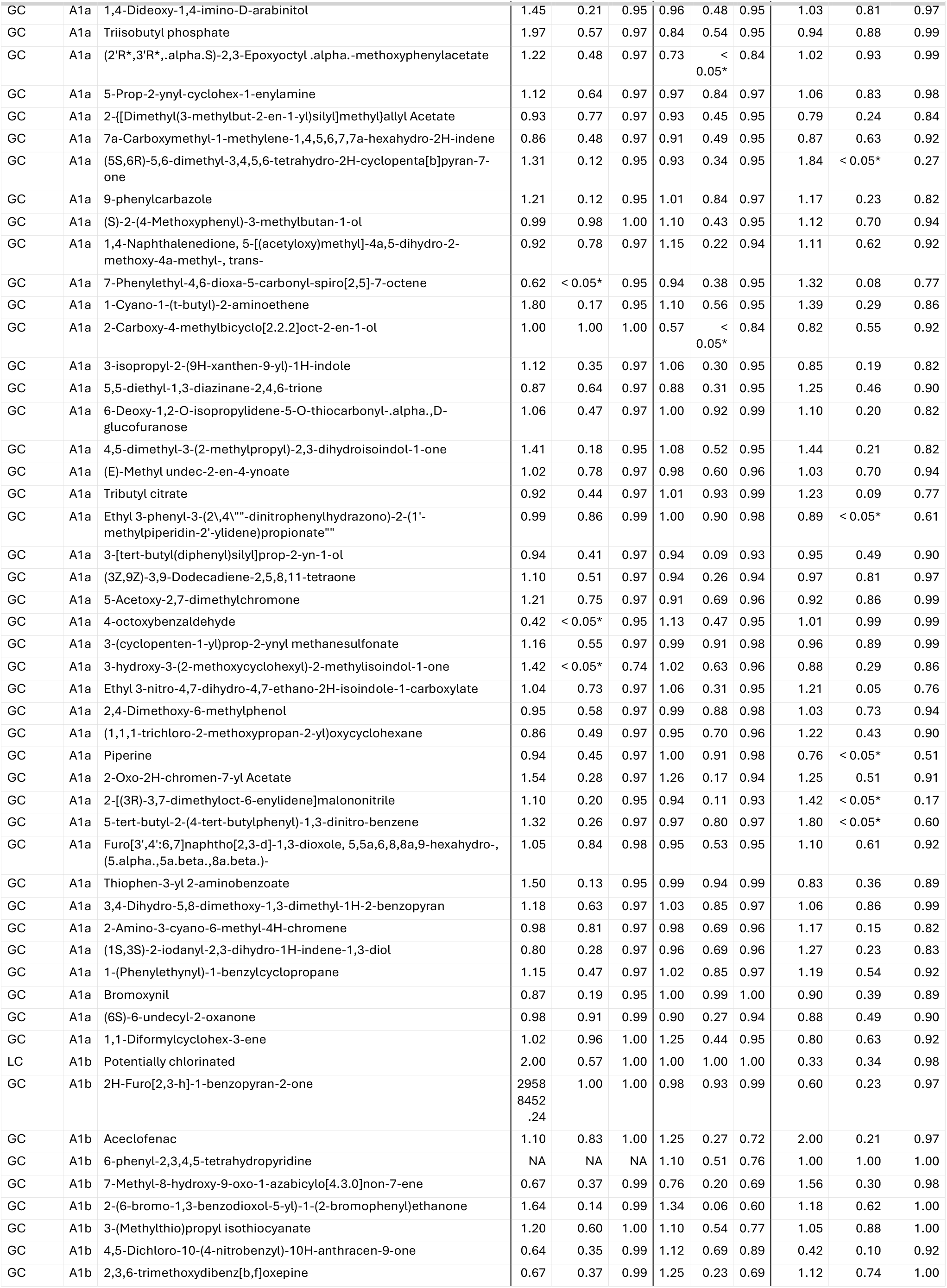
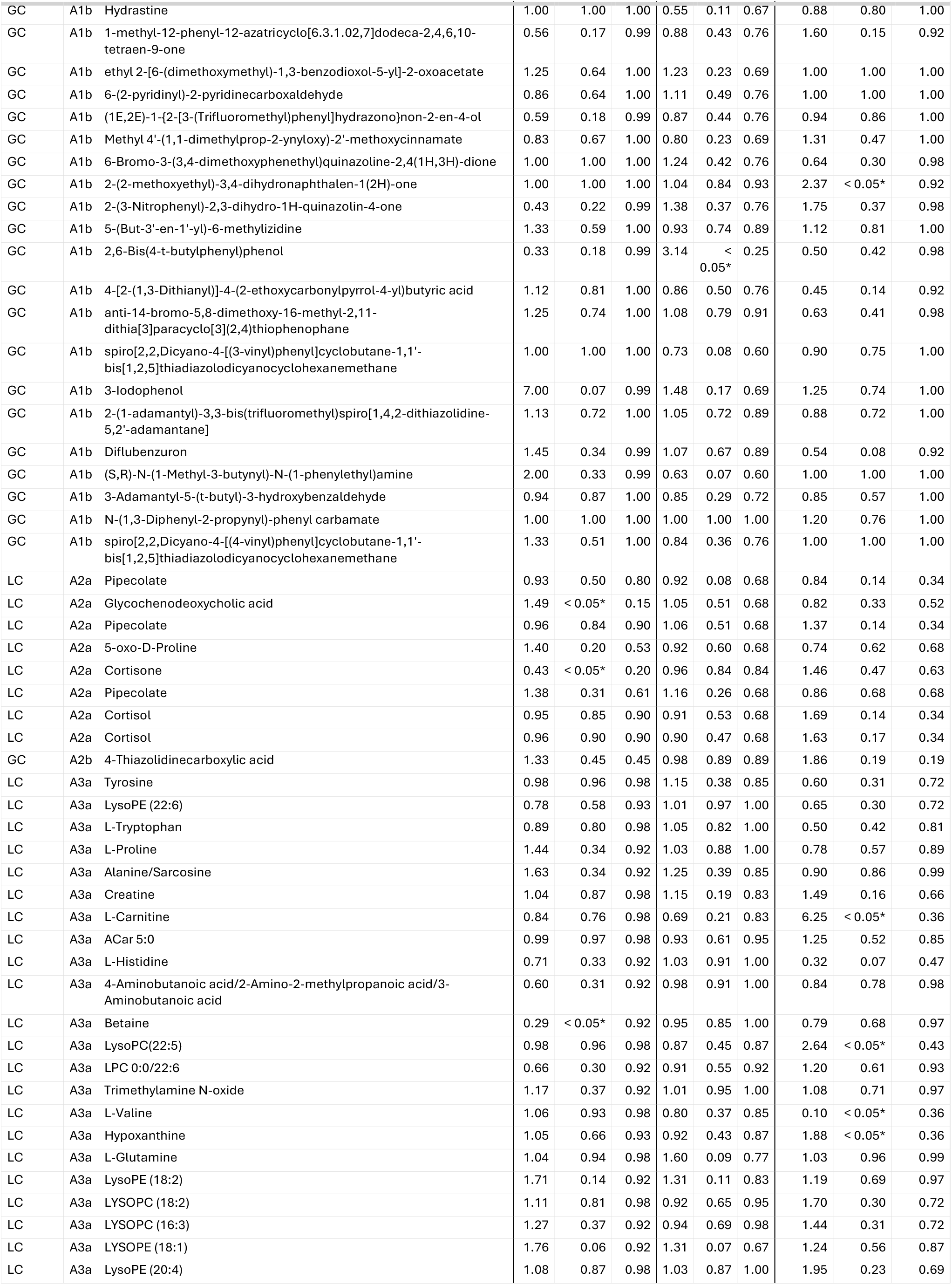
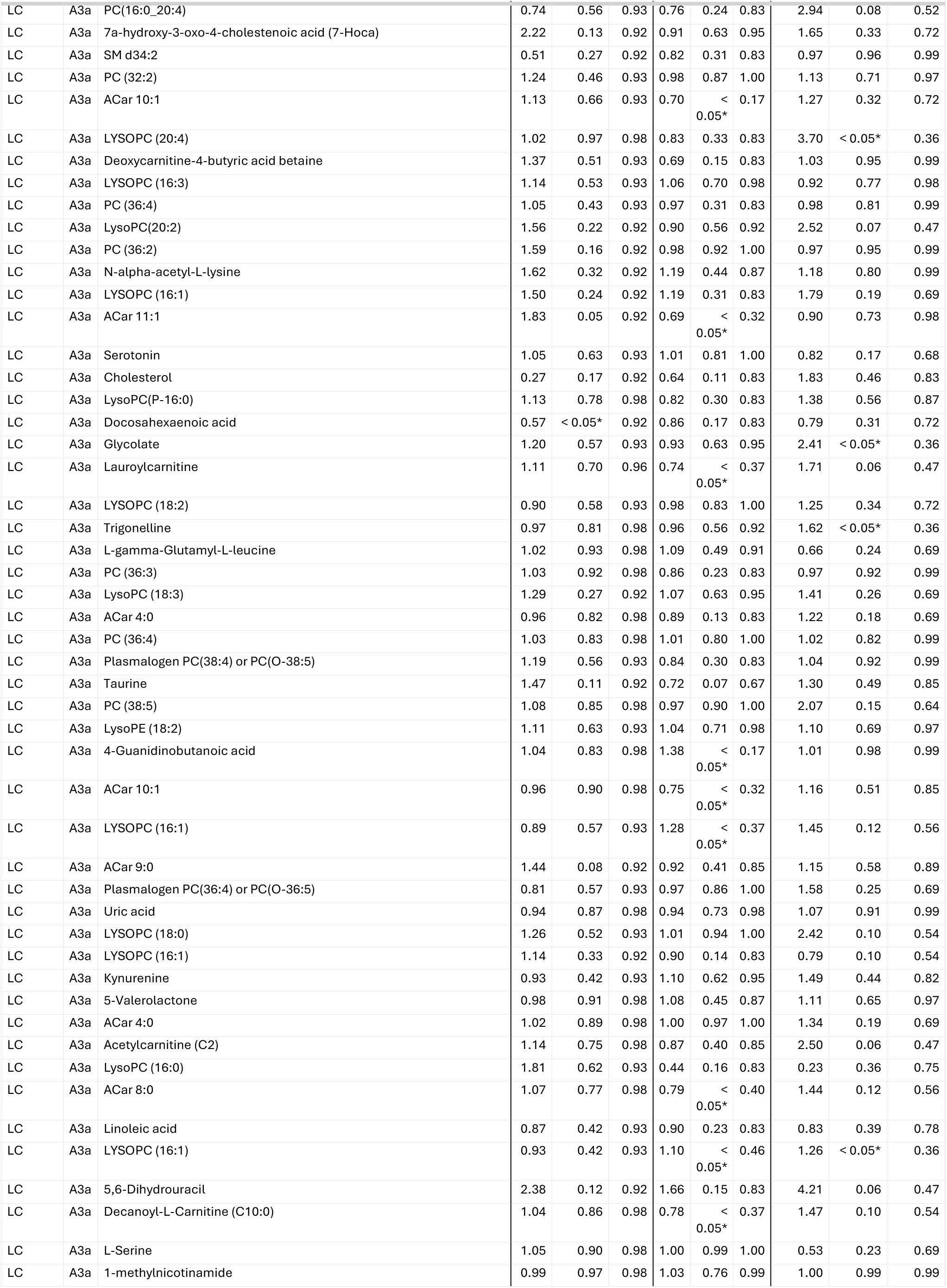
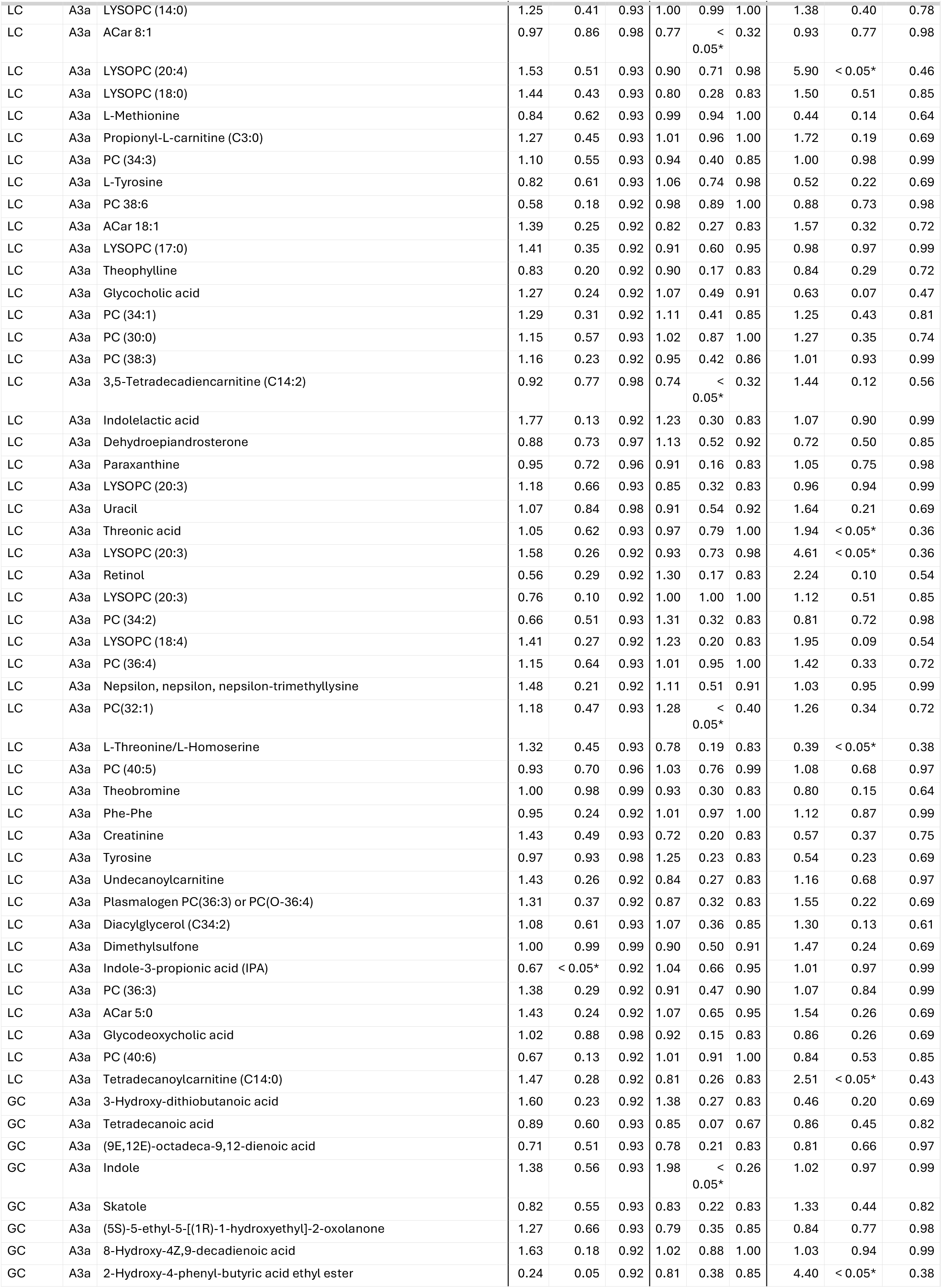
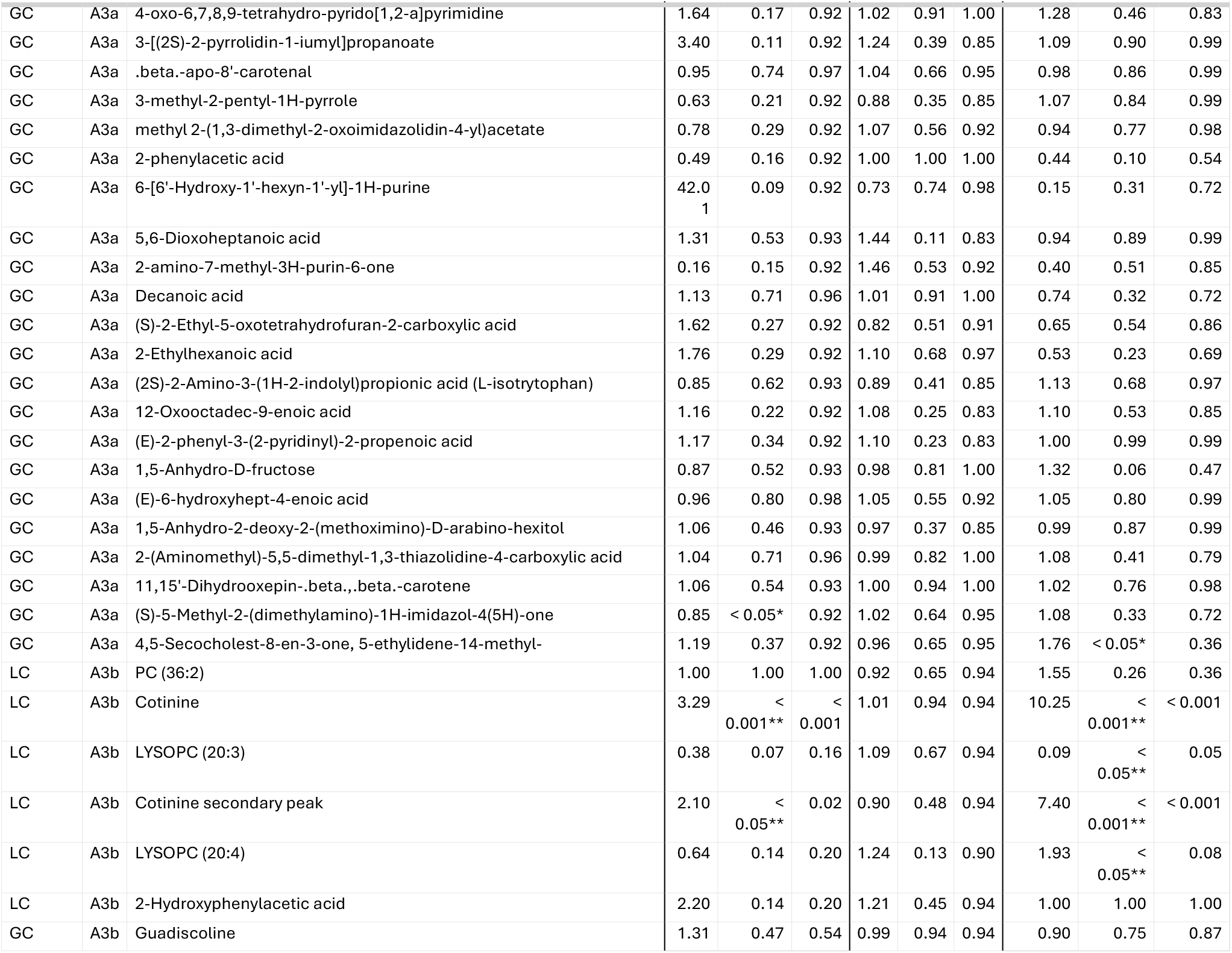
results for the initially annotated peaks in the cohort stratified analysis. Set A describes the annotated metabolites, Set B indicates the initially unidentified features/clusters. 1, 2, 3 describes the exogenous, endogenous, both endogenous/exogenously derived metabolites respectively. The last letter indicates if the feature was analyzed as continuous variable (lower case ‘a’) or as binary variable of detected, not detected (lower case ‘b’). % = proportion of samples case-control study annotation was detected; OR = odds ratio; p = p-value; FDR = false discovery rate.

|  |  |  | KORA |  |  | SAPALDIA |  |  | Doetinchem Cohort Study |  |  |
|---|---|---|---|---|---|---|---|---|---|---|---|
| Platform | Set | Name | OR | p | FDR | OR | p | FDR | OR | p | FDR |
| LC | A1a | Potentially chlorinated | 0.02 | < 0.05* | 0.95 | 0.85 | 0.76 | 0.97 | 4.58 | 0.37 | 0.89 |
| LC | A1a | L-Leucine | 1.31 | 0.47 | 0.97 | 0.97 | 0.87 | 0.97 | 0.30 | < 0.05* | 0.61 |
| LC | A1a | L-Phenylalanine | 0.90 | 0.78 | 0.97 | 1.13 | 0.54 | 0.95 | 0.43 | 0.25 | 0.84 |
| LC | A1a | L-Lysine | 1.12 | 0.70 | 0.97 | 1.02 | 0.91 | 0.98 | 0.32 | < 0.05* | 0.62 |
| LC | A1a | 2,5-Dimethylpyrazine | 1.73 | 0.50 | 0.97 | 1.18 | 0.68 | 0.96 | 1.81 | 0.60 | 0.92 |
| LC | A1a | Piperidine | 0.99 | 0.90 | 0.99 | 0.96 | 0.43 | 0.95 | 1.02 | 0.89 | 0.99 |
| LC | A1a | Caffeine | 0.91 | 0.34 | 0.97 | 0.97 | 0.55 | 0.95 | 0.95 | 0.74 | 0.95 |
| LC | A1a | Potentially chlorinated | 0.79 | 0.11 | 0.95 | 1.06 | 0.38 | 0.95 | 0.98 | 0.88 | 0.99 |
| LC | A1a | Potentially chlorinated | 1.65 | 0.26 | 0.97 | 1.08 | 0.64 | 0.96 | 4.02 | 0.15 | 0.82 |
| LC | A1a | Bis(2-ethylhexyl)phthalate | 1.20 | 0.64 | 0.97 | 1.42 | 0.28 | 0.94 | 0.60 | 0.51 | 0.91 |
| LC | A1a | L-Isoleucine | 1.37 | 0.33 | 0.97 | 1.08 | 0.58 | 0.96 | 0.39 | < 0.05* | 0.75 |
| LC | A1a | L-Isoleucine/L-Leucine | 1.21 | 0.63 | 0.97 | 0.95 | 0.81 | 0.97 | 0.36 | 0.07 | 0.77 |
| LC | A1a | Potentially chlorinated | 0.43 | 0.29 | 0.97 | 0.76 | 0.44 | 0.95 | 0.47 | 0.54 | 0.92 |
| LC | A1a | Potentially chlorinated | 0.90 | 0.63 | 0.97 | 0.96 | 0.78 | 0.97 | 1.21 | 0.45 | 0.90 |
| LC | A1a | Diethanolamine | 0.83 | 0.42 | 0.97 | 1.15 | 0.31 | 0.95 | 1.65 | 0.09 | 0.77 |
| LC | A1a | Ethyl beta-D-glucopyranoside | 0.99 | 0.91 | 0.99 | 1.08 | 0.16 | 0.94 | 1.19 | 0.22 | 0.82 |
| LC | A1a | Indole-3-acetic acid (IAA) | 1.13 | 0.57 | 0.97 | 1.17 | 0.18 | 0.94 | 0.81 | 0.35 | 0.88 |
| LC | A1a | Potentially chlorinated | 1.02 | 0.84 | 0.98 | 1.03 | 0.66 | 0.96 | 1.08 | 0.51 | 0.91 |
| LC | A1a | Potentially chlorinated | 0.79 | 0.22 | 0.95 | 0.89 | 0.31 | 0.95 | 1.08 | 0.75 | 0.95 |
| LC | A1a | Potentially chlorinated | 0.89 | 0.81 | 0.97 | 0.78 | 0.37 | 0.95 | 0.14 | < 0.05* | 0.64 |
| LC | A1a | (S)-Nicotine | 1.65 | < 0.001** | 0.03 | 1.07 | 0.23 | 0.94 | 2.72 | < 0.001** | 0.05 |
| LC | A1a | Caffeine | 0.90 | 0.30 | 0.97 | 0.97 | 0.61 | 0.96 | 0.95 | 0.73 | 0.94 |
| GC | A1a | ([1,1'-Biphenyl]-3-ylmethyl)trimethylsilane | 1.17 | 0.73 | 0.97 | 0.90 | 0.66 | 0.96 | 1.09 | 0.92 | 0.99 |
| GC | A1a | (1-cyanocyclohexyl) ethaneperoxoate | 0.39 | 0.29 | 0.97 | 1.45 | 0.25 | 0.94 | 1.45 | 0.62 | 0.92 |
| GC | A1a | 2-Ethyl-3-methylnaphthalene | 1.15 | 0.91 | 0.99 | 1.04 | 0.95 | 0.99 | 5.60 | 0.10 | 0.77 |
| GC | A1a | 3,3-Diallyloxy-1,5-bis(methoxycarbonyl)quadricyclane | 1.83 | 0.32 | 0.97 | 0.51 | < 0.05* | 0.84 | 1.98 | 0.16 | 0.82 |
| GC | A1a | 3H-4-naphtho[3,4-b]pyran | 1.21 | 0.71 | 0.97 | 1.06 | 0.88 | 0.98 | 0.81 | 0.81 | 0.97 |
| GC | A1a | 4,7-Methano-1H-indene, 1,8-diethylidene-3a,4,7,7a-tetrahydro-, (3a.alpha.,4.alpha.,7.alpha.,7a.alpha.)- | 0.55 | 0.14 | 0.95 | 0.94 | 0.74 | 0.96 | 0.45 | < 0.05* | 0.61 |
| GC | A1a | Diethyl butanedioate | 1.16 | 0.71 | 0.97 | 1.10 | 0.60 | 0.96 | 0.52 | 0.10 | 0.77 |
| GC | A1a | 5,5-Dimethyl-4-phenyl-2-oxolanone | 0.70 | 0.64 | 0.97 | 1.01 | 0.97 | 1.00 | 1.38 | 0.59 | 0.92 |
| GC | A1a | Pentacyclo[5.4.2.0(4,11).0(6,8).0(7,11)]tridec-9-en-1-ol | 1.15 | 0.69 | 0.97 | 1.20 | 0.24 | 0.94 | 1.19 | 0.63 | 0.92 |
| GC | A1a | trans-11,12-Bis(hydroxymethyl)-9,10-dihydro-9,10-ethanoanthracene | 0.73 | 0.81 | 0.97 | 0.75 | 0.63 | 0.96 | 3.00 | 0.39 | 0.89 |
| GC | A1a | 1,2,3,4-Tetrachlorobenzene | 0.44 | 0.44 | 0.97 | 1.85 | 0.24 | 0.94 | 2.47 | 0.39 | 0.89 |
| GC | A1a | Dimethyl 2-(2,2-difluoro-3-oxo-3-phenylpropyl)-4-iodopentanedioate | 0.51 | 0.22 | 0.95 | 1.34 | 0.11 | 0.93 | 0.87 | 0.72 | 0.94 |
| GC | A1a | Methyl 2-(methoxycarbonyl)-2,3-dimethyl-3-phenylbutanoate | 1.53 | 0.48 | 0.97 | 0.84 | 0.67 | 0.96 | 0.72 | 0.68 | 0.93 |
| GC | A1a | Benzothiazole | 1.04 | 0.91 | 0.99 | 0.97 | 0.88 | 0.97 | 1.23 | 0.32 | 0.87 |
| GC | A1a | dioctyl benzene-1,2-dicarboxylate | 0.85 | 0.65 | 0.97 | 0.80 | 0.25 | 0.94 | 0.75 | 0.34 | 0.88 |
| GC | A1a | Dicyclohexyl phthalate | 0.73 | 0.40 | 0.97 | 0.81 | 0.36 | 0.95 | 1.20 | 0.50 | 0.90 |
| GC | A1a | 2-(1,1,2-Trimethylpropyl)cyclohexanone | 1.16 | 0.81 | 0.97 | 1.31 | 0.31 | 0.95 | 6.71 | < 0.05* | 0.26 |
| GC | A1a | (E)-1,2-Diphenyldiazene | 2.28 | 0.32 | 0.97 | 0.86 | 0.67 | 0.96 | 0.45 | 0.36 | 0.89 |
| GC | A1a | 1-(2-Deoxy-.beta.,D-erythropentofuranosyl)-5-isopropyl-4-(trifluoromethyl)-1H-pyrimidin-2-one | 0.63 | 0.20 | 0.95 | 1.00 | 0.97 | 1.00 | 1.18 | 0.58 | 0.92 |
| GC | A1a | 1-(2-bromanyl-1-ethoxy-ethoxy)prop-2-enylbenzene | 0.78 | 0.36 | 0.97 | 0.95 | 0.74 | 0.96 | 0.46 | < 0.05* | 0.60 |
| GC | A1a | 1,1,3-Trimethyl-3-(3-methyl-4-oxo-3,4-diphenylbutyl)urea | 3.61 | 0.26 | 0.97 | 0.70 | 0.49 | 0.95 | 0.68 | 0.67 | 0.93 |
| GC | A1a | Vanillin | 1.60 | 0.52 | 0.97 | 1.34 | 0.52 | 0.95 | 0.53 | 0.44 | 0.90 |
| GC | A1a | 3-O-ethyl 4-O-methyl 1-(carbamoylamino)-2-(2-ethoxy-2-oxoethyl)-5-methylpyrrole-3,4-dicarboxylate | 0.47 | 0.33 | 0.97 | 1.16 | 0.66 | 0.96 | 6.81 | < 0.05* | 0.51 |
| GC | A1a | 2,4,5-triphenyl-1H-imidazole | 1.10 | 0.71 | 0.97 | 1.02 | 0.85 | 0.97 | 1.33 | 0.22 | 0.82 |
| GC | A1a | 1-(2-Hydroxyphenyl)-2-methyl-1-propanone | 1.03 | 0.95 | 1.00 | 1.04 | 0.86 | 0.97 | 0.94 | 0.91 | 0.99 |
| GC | A1a | 8-Methyl-8H-azepino[1,2-a]indole | 0.19 | 0.16 | 0.95 | 2.34 | 0.10 | 0.93 | 4.21 | 0.18 | 0.82 |
| GC | A1a | 7-p-anisylfuro[3,2-b]pyridine | 0.40 | 0.55 | 0.97 | 0.62 | 0.50 | 0.95 | 1.18 | 0.92 | 0.99 |
| GC | A1a | Dimethyl 5,6-dimethylene-7-oxabicyclo[2.2.1]hept-2-ene-2,3-dicarboxylate | 1.49 | 0.69 | 0.97 | 1.38 | 0.47 | 0.95 | 0.64 | 0.65 | 0.92 |
| GC | A1a | Ethyl 2-Hydroxy-(4-hydroxymethylphenyl)acetate | 13.54 | 0.17 | 0.95 | 1.26 | 0.58 | 0.96 | 0.07 | < 0.05* | 0.61 |
| GC | A1a | Bis(3-tetrahydrofuryl) sulfide | 6.61 | < 0.05* | 0.95 | 1.18 | 0.52 | 0.95 | 0.41 | 0.23 | 0.82 |
| GC | A1a | 1-(3-Phenylpropyl)piperidine | 0.92 | 0.87 | 0.99 | 0.98 | 0.93 | 0.99 | 0.61 | 0.21 | 0.82 |
| GC | A1a | 1(3H)-Isobenzofuranone, hexahydro-3-(1-methylethyl)-, (3.alpha.,3a.alpha.,7a.beta.)- | 1.83 | < 0.05* | 0.95 | 0.91 | 0.36 | 0.95 | 2.04 | < 0.05* | 0.31 |
| GC | A1a | [(E)-2-methylsulfanylethenyl]benzene | 1.25 | 0.73 | 0.97 | 1.28 | 0.39 | 0.95 | 0.56 | 0.37 | 0.89 |
| GC | A1a | 2-Methyl-6-nitrobenzaldehyde | 4.11 | 0.07 | 0.95 | 1.61 | 0.13 | 0.94 | 0.77 | 0.70 | 0.94 |
| GC | A1a | 1,9-Dimethylcarbazole | 0.32 | 0.50 | 0.97 | 1.70 | 0.50 | 0.95 | 2.69 | 0.49 | 0.90 |
| GC | A1a | 1-(1,3-dioxolan-2-yl)-1-cyclohexanecarboxaldehyde | 1.63 | 0.45 | 0.97 | 0.85 | 0.64 | 0.96 | 0.44 | 0.26 | 0.84 |
| GC | A1a | Dimethyl 4-[4'-(diethylamino)phenylethynyl]pyridine-2,6-dicarboxylate | 0.32 | 0.21 | 0.95 | 0.86 | 0.67 | 0.96 | 1.23 | 0.75 | 0.95 |
| GC | A1a | (2S,3S,4S,5R)-3,4,5-trimethoxy-6-oxidanyl-oxane-2-carboxylic acid | 2.37 | 0.09 | 0.95 | 1.19 | 0.38 | 0.95 | 2.63 | < 0.05* | 0.54 |
| GC | A1a | 4-Isopropyl-2,5-dimethoxybenzyl acetate | 0.00 | < 0.05* | 0.88 | 1.20 | 0.76 | 0.97 | 1.01 | 0.99 | 0.99 |
| GC | A1a | 1-(4-Methylphenyl)-1-phenylethanol | 1.08 | 0.79 | 0.97 | 1.41 | 0.12 | 0.93 | 1.45 | 0.32 | 0.87 |
| GC | A1a | 2-Methyl-4'-(methylthio)-2-morpholinopropiophenone | 1.26 | 0.49 | 0.97 | 0.84 | 0.23 | 0.94 | 0.92 | 0.73 | 0.94 |
| GC | A1a | Isopropyl indolizine-2-carboxylate | 1.20 | 0.81 | 0.97 | 1.84 | < 0.05* | 0.84 | 1.02 | 0.97 | 0.99 |
| GC | A1a | 9H-Fluorene | 1.48 | 0.17 | 0.95 | 0.96 | 0.74 | 0.96 | 2.08 | < 0.05* | 0.51 |
| GC | A1a | 1-(4-Methoxyphenyl)-N-methyl-ethanamine | 1.06 | 0.88 | 0.99 | 1.17 | 0.36 | 0.95 | 1.12 | 0.72 | 0.94 |
| GC | A1a | N-[6'-(Ethynylpyridin-2'-yl)]-acetamide | 0.54 | 0.43 | 0.97 | 1.51 | 0.15 | 0.94 | 2.33 | 0.18 | 0.82 |
| GC | A1a | Methyl p-hydroxydithiobenzoate | 0.99 | 0.98 | 1.00 | 0.82 | 0.15 | 0.94 | 1.54 | 0.16 | 0.82 |
| GC | A1a | 7-Methyl-3,4-dihydrobenzo[c]pyran-1H-ol | 1.02 | 0.96 | 1.00 | 1.05 | 0.91 | 0.98 | 1.70 | 0.38 | 0.89 |
| GC | A1a | 1-(Methylthio)naphthalene | 1.02 | 0.91 | 0.99 | 0.84 | < 0.05* | 0.84 | 1.02 | 0.90 | 0.99 |
| GC | A1a | Methyl 1,2,3,4-tetrahydro-2-(3',4'-dimethoxybenzyl)-3-oxonaphthalene-2-carboxylate | 1.20 | 0.59 | 0.97 | 0.93 | 0.51 | 0.95 | 0.85 | 0.47 | 0.90 |
| GC | A1a | Dipentyl phthalate | 0.55 | 0.15 | 0.95 | 1.03 | 0.85 | 0.97 | 1.12 | 0.55 | 0.92 |
| GC | A1a | Resorcinol | 0.79 | 0.75 | 0.97 | 0.90 | 0.66 | 0.96 | 2.17 | 0.20 | 0.82 |
| GC | A1a | 2-(2-Hydroxy-2-methylpropyl)pyrrolidine | 3.49 | 0.19 | 0.95 | 1.20 | 0.65 | 0.96 | 0.83 | 0.81 | 0.97 |
| GC | A1a | 4,5-dihydro-2-benzofuran-5-ol | 3.68 | 0.10 | 0.95 | 1.25 | 0.39 | 0.95 | 1.95 | 0.13 | 0.82 |
| GC | A1a | 4-(Methylamino)benzonitrile | 3.77 | 0.08 | 0.95 | 0.86 | 0.56 | 0.95 | 8.04 | < 0.05* | 0.51 |
| GC | A1a | 4-(Hexyloxy)phenol | 0.70 | 0.52 | 0.97 | 1.08 | 0.75 | 0.96 | 2.26 | 0.11 | 0.78 |
| GC | A1a | Neopentyl exo-1-methyl-2-oxabicyclo[3.1.0]hex-3-ene-6-sulfonate | 1.19 | 0.92 | 1.00 | 0.75 | 0.67 | 0.96 | 3.06 | 0.45 | 0.90 |
| GC | A1a | N'-formylbenzohydrazide | 0.50 | 0.39 | 0.97 | 0.70 | 0.11 | 0.93 | 1.72 | 0.33 | 0.87 |
| GC | A1a | diheptyl benzene-1,2-dicarboxylate | 1.18 | 0.63 | 0.97 | 1.09 | 0.54 | 0.95 | 1.41 | 0.28 | 0.86 |
| GC | A1a | (1S,8R)-9-(hydroxymethyl)-9-azabicyclo[6.2.0]decan-10-one | 0.07 | 0.08 | 0.95 | 2.23 | 0.22 | 0.94 | 1.76 | 0.71 | 0.94 |
| GC | A1a | Methyl salicylate | 33.55 | < 0.05* | 0.95 | 1.23 | 0.67 | 0.96 | 0.98 | 0.99 | 0.99 |
| GC | A1a | Dicinnamyl ether | 0.20 | 0.19 | 0.95 | 1.81 | 0.25 | 0.94 | 7.99 | 0.10 | 0.77 |
| GC | A1a | [(3aS,4S,7aS)-2,2-dimethyl-3a,4,7,7a-tetrahydro-1,3-benzodioxol-4-yl]-tert-butyl-dimethyl-silane | 1.57 | 0.52 | 0.97 | 0.71 | 0.21 | 0.94 | 0.33 | 0.08 | 0.77 |
| GC | A1a | [(1S,3aR,4S,5R,7aS)-1-ethyl-5-ethynyl-2,3,3a,4,5,7a-hexahydro-1H-inden-4-yl]-(1H-pyrrol-2-yl)methanone | 1.33 | 0.32 | 0.97 | 0.99 | 0.91 | 0.98 | 0.45 | < 0.05* | 0.71 |
| GC | A1a | 2,4-Diaza-3-methylbicyclo[3.2.0]heptane-1,5-dicarbonitrile | 0.79 | 0.47 | 0.97 | 0.98 | 0.93 | 0.99 | 1.22 | 0.66 | 0.93 |
| GC | A1a | 2-phenylpyridine | 2.05 | 0.19 | 0.95 | 1.19 | 0.34 | 0.95 | 1.06 | 0.88 | 0.99 |
| GC | A1a | Homosalate | 0.50 | 0.64 | 0.97 | 0.61 | 0.45 | 0.95 | 0.21 | 0.29 | 0.86 |
| GC | A1a | (S)-2-((2-Methoxyethoxy)methyl)pyrrolidine | 1.40 | 0.41 | 0.97 | 1.00 | 0.98 | 1.00 | 1.50 | 0.21 | 0.82 |
| GC | A1a | methyl 2-(3,5-dihydroxy-4-methylphenyl)acetate | 0.59 | 0.57 | 0.97 | 0.79 | 0.59 | 0.96 | 0.15 | 0.11 | 0.78 |
| GC | A1a | 2-[(1'-hydroxyethyl)carbonyl]thiophene | 3.71 | 0.17 | 0.95 | 0.63 | 0.20 | 0.94 | 5.27 | 0.07 | 0.77 |
| GC | A1a | Bis(2-ethylhexyl) terephthalate | 2.30 | 0.16 | 0.95 | 0.99 | 0.96 | 1.00 | 0.79 | 0.68 | 0.93 |
| GC | A1a | 3-[(t-Butyldimethylsilyl)oxy]-2-(4'-methoxybenzyloxy)-5-hexene | 1.11 | 0.67 | 0.97 | 0.99 | 0.96 | 1.00 | 0.97 | 0.90 | 0.99 |
| GC | A1a | (2-iminocyclohept[d]imidazol-1-yl)amine | 0.76 | 0.38 | 0.97 | 0.68 | < 0.05* | 0.84 | 1.39 | 0.47 | 0.90 |
| GC | A1a | 3,4-Homotropilidene-3-methyl-2,6-dicarbonitrile | 0.08 | 0.12 | 0.95 | 2.03 | 0.24 | 0.94 | 1.28 | 0.79 | 0.97 |
| GC | A1a | 2,3,4,5,6-pentachlorophenol | 0.72 | 0.32 | 0.97 | 1.24 | 0.09 | 0.93 | 0.96 | 0.88 | 0.99 |
| GC | A1a | 3-Isopropyl-4-methoxy-1-(4-methoxyphenyl)-4-methylpent-1-yn-3-ol | 0.45 | 0.38 | 0.97 | 0.49 | 0.11 | 0.93 | 1.49 | 0.68 | 0.93 |
| GC | A1a | (3RS,5SR)-5-Dimethyl(phenyl)silyl-3-methyhexanonitrile | 0.53 | 0.43 | 0.97 | 0.84 | 0.63 | 0.96 | 1.29 | 0.68 | 0.93 |
| GC | A1a | methyl (9E)-9-methoxyiminononanoate | 0.42 | 0.35 | 0.97 | 1.75 | 0.17 | 0.94 | 0.52 | 0.42 | 0.90 |
| GC | A1a | 1-Naphthyl(veratryl)amine | 0.33 | 0.25 | 0.97 | 0.85 | 0.68 | 0.96 | 0.71 | 0.59 | 0.92 |
| GC | A1a | 1,4-Dimethyl-2-oxa-5-azabicyclo[2.2.2]octan-3,6-dione | 1.08 | 0.88 | 0.99 | 1.43 | 0.13 | 0.94 | 1.97 | 0.15 | 0.82 |
| GC | A1a | 3-Aminoacetophenone | 0.36 | < 0.05* | 0.95 | 0.96 | 0.87 | 0.97 | 0.73 | 0.40 | 0.89 |
| GC | A1a | Ethyl (3-Ethylfuran-2-yl)phenylacetate | 0.35 | 0.18 | 0.95 | 1.74 | 0.08 | 0.93 | 1.20 | 0.78 | 0.97 |
| GC | A1a | 1,4-bis(hept-1-ynyl)benzene | 1.09 | 0.86 | 0.99 | 0.86 | 0.38 | 0.95 | 0.90 | 0.84 | 0.98 |
| GC | A1a | (E)-5-(1,3-benzodioxol-5-yl)-2-pentenal | 1.06 | 0.89 | 0.99 | 0.91 | 0.67 | 0.96 | 0.57 | 0.29 | 0.86 |
| GC | A1a | 1-allyl-2-nitro-cyclohexane | 2.16 | 0.16 | 0.95 | 0.59 | 0.11 | 0.93 | 0.70 | 0.41 | 0.90 |
| GC | A1a | 1-Ethyl-2,3,4,5-tetrahydro-1-benzazepine-2-carbonitrile | 1.04 | 0.88 | 0.99 | 0.99 | 0.95 | 1.00 | 0.71 | 0.18 | 0.82 |
| GC | A1a | 2-(Methylmercapto)benzothiazole | 1.05 | 0.49 | 0.97 | 0.95 | 0.36 | 0.95 | 1.09 | 0.42 | 0.90 |
| GC | A1a | {3,3-bis[4'-(Dimethylamino)phenyl]methyl}-tetrahydrofuran-2-one | 0.89 | 0.74 | 0.97 | 1.22 | 0.27 | 0.94 | 1.34 | 0.39 | 0.89 |
| GC | A1a | 2,2,4,4,6,6-hexamethyl-5-methylidenecyclohexane-1,3-dione | 2.55 | 0.21 | 0.95 | 0.83 | 0.60 | 0.96 | 0.50 | 0.33 | 0.87 |
| GC | A1a | 1-[(R)-phenyl(2-quinolylsulfanyl)methyl]cyclohexanol | 1.34 | 0.72 | 0.97 | 1.42 | 0.41 | 0.95 | 1.75 | 0.49 | 0.90 |
| GC | A1a | 2-methyl-5-propan-2-yl-1H-pyrrole-3-carbaldehyde | 1.22 | 0.63 | 0.97 | 1.07 | 0.71 | 0.96 | 1.78 | 0.08 | 0.77 |
| GC | A1a | 3,3-(Ethylenedioxy)-12-methoxyeudesm-7(11)-eno-13,6.alpha.-lactone | 0.69 | 0.41 | 0.97 | 0.88 | 0.42 | 0.95 | 0.62 | 0.19 | 0.82 |
| GC | A1a | (+-)-10,11-trans-11,12-trans-10,11-Dihydro-10-ethyl-12-hydroxy-4-propyl-6,6,11-trimethyl-2H,6H,12H-benzo[1,2-b:3,4:b':5,6-b\]tripyran-2-one"" | 0.99 | 0.97 | 1.00 | 0.73 | 0.12 | 0.93 | 2.88 | < 0.05* | 0.61 |
| GC | A1a | methyl 3-hydroxy-2,2-dimethyl-3,3-diphenylpropanoate | 1.98 | 0.68 | 0.97 | 0.80 | 0.75 | 0.96 | 0.42 | 0.46 | 0.90 |
| GC | A1a | Phenyl 4-[bis(ethoxycarbonyl)but-3-ynyl]-2,3,4-trideoxy-.alpha.,L-glcero-pent-2-enopyranoside | 0.11 | < 0.05* | 0.95 | 1.23 | 0.55 | 0.95 | 2.05 | 0.24 | 0.84 |
| GC | A1a | 6-methoxy-2,2,4-trimethyl-1H-quinoline | 1.31 | 0.69 | 0.97 | 1.20 | 0.41 | 0.95 | 1.39 | 0.43 | 0.90 |
| GC | A1a | 2-(3,4-Dinitrophenyl)-4-nitro-2H-1,2,3-triazole | 0.46 | 0.20 | 0.95 | 1.36 | 0.18 | 0.94 | 2.48 | 0.07 | 0.77 |
| GC | A1a | 7'-(iodomethyl)-1',7'-dimethylspiro[1,3-dioxolane-2,2'-bicyclo[2.2.1]heptane] | 1.02 | 0.97 | 1.00 | 1.32 | 0.38 | 0.95 | 1.21 | 0.77 | 0.97 |
| GC | A1a | Coumarin | 1.24 | 0.65 | 0.97 | 0.87 | 0.53 | 0.95 | 1.66 | 0.49 | 0.90 |
| GC | A1a | 3-(2-methyl-5-nitroimidazol-1-yl)propanenitrile | 0.88 | 0.69 | 0.97 | 1.05 | 0.68 | 0.96 | 1.13 | 0.51 | 0.91 |
| GC | A1a | Furmecyclox | 2.95 | < 0.05* | 0.95 | 0.89 | 0.61 | 0.96 | 0.64 | 0.31 | 0.87 |
| GC | A1a | 3-(2,2-dimethylpropylidene)bicyclo[3.3.1]nonane-2,4-dione | 0.95 | 0.78 | 0.97 | 0.88 | 0.15 | 0.94 | 1.16 | 0.46 | 0.90 |
| GC | A1a | 4-(dimethylaminoazo)benzonitrile | 1.66 | 0.22 | 0.95 | 0.84 | 0.40 | 0.95 | 0.57 | 0.22 | 0.82 |
| GC | A1a | syn-2,5,5-trimethyl-2-[3-(phenylthio)propyl]tetrahydrofuran-3-ol | 0.90 | 0.72 | 0.97 | 0.86 | 0.15 | 0.94 | 0.94 | 0.79 | 0.97 |
| GC | A1a | 4-[(1S,6S)-Bicyclo[4.1.0]hept-1-yl]butyronitrile | 0.79 | 0.73 | 0.97 | 0.72 | 0.19 | 0.94 | 0.43 | 0.19 | 0.82 |
| GC | A1a | Acenaphthylene | 5.18 | 0.12 | 0.95 | 0.66 | 0.19 | 0.94 | 1.38 | 0.75 | 0.95 |
| GC | A1a | Amantadine | 0.28 | 0.25 | 0.97 | 1.35 | 0.60 | 0.96 | 0.81 | 0.86 | 0.99 |
| GC | A1a | [2-(2-bromopropanoyloxy)phenyl] 4-ethylbenzoate | 0.26 | 0.16 | 0.95 | 0.87 | 0.69 | 0.96 | 0.52 | 0.28 | 0.86 |
| GC | A1a | 5-[(4-methylphenoxy)methyl]-1,3,4-thiadiazol-2-amine | 3.11 | 0.15 | 0.95 | 0.86 | 0.73 | 0.96 | 0.80 | 0.81 | 0.97 |
| GC | A1a | (3aS,5S,11bS)-6,10,11-Trimethoxy-5,9-dimethyl-3,3a,5,11b-tetrahydro-1,4-dioxa-cyclopenta[a]anthracen-2-one | 0.84 | 0.23 | 0.97 | 1.04 | 0.46 | 0.95 | 0.88 | < 0.05* | 0.61 |
| GC | A1a | (R,R)5-[3-(Trimethylsilyl)prop-2-yn-1-yl]-methyl-6-phenylcyclohexa-1,3-dien-1-carbaldehyde | 0.81 | 0.67 | 0.97 | 0.92 | 0.66 | 0.96 | 0.78 | 0.51 | 0.91 |
| GC | A1a | Hexyl salicylate | 1.40 | 0.71 | 0.97 | 1.00 | 1.00 | 1.00 | 0.63 | 0.73 | 0.94 |
| GC | A1a | (R)-2-{2-[6-Methoxy-3,4-dihydro-2H-naphthalen-(1E)-ylidene]-ethyl}-2-methyl-3-vinyl-cyclohexanone | 0.47 | 0.18 | 0.95 | 0.93 | 0.76 | 0.97 | 0.71 | 0.44 | 0.90 |
| GC | A1a | 5-Deoxy-5,5-difluoro-2-(formyloxy)-5-iodo-1,3,4-trimethoxy-D-Arabinitol | 4.74 | 0.26 | 0.97 | 1.40 | 0.57 | 0.95 | 1.45 | 0.70 | 0.93 |
| GC | A1a | 1-[4'-(Hydroxybenzyl]-6,7-dimethoxy-5-hydroxy-1,2,3,4-tetrahydro-isoquinoline | 1.82 | 0.67 | 0.97 | 2.98 | 0.11 | 0.93 | 1.03 | 0.98 | 0.99 |
| GC | A1a | 7-t-butyl-1-tetralol | 0.67 | 0.39 | 0.97 | 1.23 | 0.25 | 0.94 | 1.14 | 0.66 | 0.93 |
| GC | A1a | Acetanilide | 9.10 | 0.06 | 0.95 | 2.26 | 0.11 | 0.93 | 2.86 | 0.44 | 0.90 |
| GC | A1a | (2'S,R,R)-(-)-3-[2'-(1-Ethyl-1-methoxypropy)-2-oxobipyrrolidinyl-3-yl]-3-hexanoic acid methyl ester | 0.16 | 0.10 | 0.95 | 1.50 | 0.29 | 0.95 | 3.23 | 0.12 | 0.81 |
| GC | A1a | 1-Methylphenanthrene | 5.34 | 0.15 | 0.95 | 1.54 | 0.41 | 0.95 | 0.66 | 0.68 | 0.93 |
| GC | A1a | 2,3,9,10,11-Pentamethoxy-5,6,7,8-tetrahydro-dibenzo[a,c]cyclooctene | 0.72 | 0.09 | 0.95 | 1.09 | 0.22 | 0.94 | 0.82 | < 0.05* | 0.61 |
| GC | A1a | 1-(4-methoxyphenyl)-2,2-dimethylpropan-1-one | 0.79 | 0.66 | 0.97 | 1.11 | 0.65 | 0.96 | 0.38 | 0.12 | 0.79 |
| GC | A1a | 2,4-Dimethyl-3-(N-methylanilinomethyl)-3-pentanol | 2.44 | 0.25 | 0.97 | 0.83 | 0.52 | 0.95 | 1.18 | 0.62 | 0.92 |
| GC | A1a | Butyl Benzoate | 0.78 | 0.53 | 0.97 | 2.24 | < 0.05* | 0.84 | 2.94 | 0.20 | 0.82 |
| GC | A1a | Diallyl maleate | 0.85 | 0.58 | 0.97 | 0.86 | 0.25 | 0.94 | 0.86 | 0.47 | 0.90 |
| GC | A1a | 2-(hydroxymethyl)-1-(3-methoxyphenyl)cyclopropanecarbonitrile | 1.53 | 0.52 | 0.97 | 1.17 | 0.54 | 0.95 | 1.23 | 0.68 | 0.93 |
| GC | A1a | 4-hydroxy-3,5-dimethoxybenzaldehyde | 0.15 | 0.16 | 0.95 | 1.65 | 0.24 | 0.94 | 1.54 | 0.65 | 0.92 |
| GC | A1a | 5-(4-Methoxyphenyl)-7-benzyl-1,11,11-trimethyl-5-aza-2-oxatetracyclo[6.5.0.0(3,6).010,12)]tridecan-4-one | 0.70 | 0.51 | 0.97 | 1.07 | 0.80 | 0.97 | 1.11 | 0.81 | 0.97 |
| GC | A1a | Binapacryl | 0.75 | 0.62 | 0.97 | 0.92 | 0.75 | 0.96 | 0.41 | 0.16 | 0.82 |
| GC | A1a | Caffeine | 0.92 | 0.50 | 0.97 | 0.98 | 0.78 | 0.97 | 0.92 | 0.56 | 0.92 |
| GC | A1a | delta-Tocopherol | 0.83 | 0.54 | 0.97 | 1.35 | < 0.05* | 0.84 | 0.96 | 0.85 | 0.99 |
| GC | A1a | Phenylparaben | 1.94 | 0.14 | 0.95 | 0.84 | 0.41 | 0.95 | 1.55 | 0.27 | 0.84 |
| GC | A1a | 4-Methyl-1-(2,4,5-trimethylphenyl)-1-pentanone | 0.73 | 0.79 | 0.97 | 1.69 | 0.34 | 0.95 | 0.56 | 0.63 | 0.92 |
| GC | A1a | 1-(1H-indazol-3-yl)propanol | 0.17 | 0.08 | 0.95 | 0.73 | 0.37 | 0.95 | 1.95 | 0.45 | 0.90 |
| GC | A1a | Benzenecarbothioic acid, S-2-cyclopenten-1-yl ester | 0.47 | 0.47 | 0.97 | 1.30 | 0.44 | 0.95 | 0.50 | 0.34 | 0.88 |
| GC | A1a | 1-Naphthalenethiol | 1.21 | 0.81 | 0.97 | 1.29 | 0.47 | 0.95 | 1.45 | 0.56 | 0.92 |
| GC | A1a | 1-[6-(2-hydroxy-2-methyl-propyl)-2-pyridyl]-2-methyl-propan-2-ol | 0.86 | 0.84 | 0.98 | 1.21 | 0.56 | 0.95 | 0.92 | 0.87 | 0.99 |
| GC | A1a | 3-Methyl-1-phenyl-1-pentyn-3-ol | 0.85 | 0.82 | 0.97 | 1.58 | 0.22 | 0.94 | 0.69 | 0.61 | 0.92 |
| GC | A1a | Bicyclo[3.3.1]nonan-2-one, 1,4,5-trimethyl-9-methylene-, exo-(.+-.)- | 0.95 | 0.88 | 0.99 | 1.17 | 0.59 | 0.96 | 2.28 | 0.22 | 0.82 |
| GC | A1a | methyl (2S)-5-ethoxy-3,4-dihydro-2H-pyrrole-2-carboxylate | 1.40 | 0.66 | 0.97 | 1.34 | 0.30 | 0.95 | 2.70 | 0.13 | 0.82 |
| GC | A1a | dimethyl benzene-1,2-dicarboxylate | 1.18 | 0.49 | 0.97 | 3.49 | 0.09 | 0.93 | 9.45 | 0.10 | 0.77 |
| GC | A1a | Oxybenzone | 0.85 | 0.55 | 0.97 | 0.95 | 0.51 | 0.95 | 0.50 | 0.10 | 0.77 |
| GC | A1a | 3,3'-Diacetyl-1,1'-dimethyl-1H,1'H-2,2'-bisindolyl | 0.85 | 0.67 | 0.97 | 1.11 | 0.60 | 0.96 | 1.38 | 0.43 | 0.90 |
| GC | A1a | PCB 114 | 1.02 | 0.95 | 1.00 | 1.02 | 0.88 | 0.97 | 0.99 | 0.97 | 0.99 |
| GC | A1a | 2-(1,3-benzoxazol-2-yl)ethanol | 0.77 | 0.80 | 0.97 | 0.82 | 0.74 | 0.96 | 2.77 | 0.48 | 0.90 |
| GC | A1a | 6-tert-Butyl-2-phenyl-4-propan-2-ylidene-cyclopenta[c]pyrazole | 1.11 | 0.81 | 0.97 | 0.84 | 0.36 | 0.95 | 0.83 | 0.62 | 0.92 |
| GC | A1a | 2-(2-aminophenyl)-2-adamantanecarbonitrile | 1.36 | 0.48 | 0.97 | 1.32 | 0.18 | 0.94 | 1.59 | 0.31 | 0.87 |
| GC | A1a | 2,2-bis(prop-2-enyl)pyrrolidine | 1.12 | 0.81 | 0.97 | 0.82 | 0.37 | 0.95 | 1.48 | 0.54 | 0.92 |
| GC | A1a | 1-[4-(Dimethoxymethyl)phenyl]ethanone | 5.89 | < 0.05* | 0.95 | 1.26 | 0.51 | 0.95 | 1.04 | 0.96 | 0.99 |
| GC | A1a | trans-11-Hydroxy-6-(hydroxymethyl)-2,3,10-trimethoxydibenzo[1a,4a:8a,12a]cyclooctadiene-7-carboxylic acid lactone | 0.67 | 0.31 | 0.97 | 1.05 | 0.60 | 0.96 | 1.35 | 0.05 | 0.76 |
| GC | A1a | gamma-Nonanolactone | 1.31 | 0.54 | 0.97 | 0.58 | < 0.05* | 0.84 | 0.90 | 0.83 | 0.98 |
| GC | A1a | 1,2,2-Butanetricarboxylic acid, 4-(tetrahydro-2-methyl-2-furanyl)-, 2-ethyl ester | 0.64 | 0.37 | 0.97 | 0.88 | 0.61 | 0.96 | 0.79 | 0.66 | 0.93 |
| GC | A1a | 1-Furfurylpyrrole | 12.98 | < 0.05* | 0.95 | 0.81 | 0.72 | 0.96 | 3.63 | 0.25 | 0.84 |
| GC | A1a | 3-phenylprop-2-yn-1-ol | 14.75 | 0.10 | 0.95 | 0.89 | 0.84 | 0.97 | 3.16 | 0.41 | 0.90 |
| GC | A1a | Mequinol | 4.25 | 0.13 | 0.95 | 0.93 | 0.90 | 0.98 | 0.79 | 0.80 | 0.97 |
| GC | A1a | (+-)-(1aR,2R)-1a-Methyl-2-phenylperhydrooxireno[2,3-c]furan | 3.25 | 0.25 | 0.97 | 0.72 | 0.43 | 0.95 | 1.14 | 0.88 | 0.99 |
| GC | A1a | Diphenyl sulfone | 1.22 | 0.68 | 0.97 | 0.81 | 0.55 | 0.95 | 0.91 | 0.87 | 0.99 |
| GC | A1a | methyl 6-(oxolan-2-yloxy)hexanoate | 0.78 | 0.47 | 0.97 | 1.26 | 0.24 | 0.94 | 0.62 | 0.23 | 0.83 |
| GC | A1a | Methyl paraben | 1.19 | 0.43 | 0.97 | 1.02 | 0.82 | 0.97 | 1.03 | 0.87 | 0.99 |
| GC | A1a | (1R(*),3S(*).5S(*).7S(*))-1,3-Dimethyl-7-hydroxy-6-oxabicyclo[3.2.1]octan-2-one | 2.05 | 0.40 | 0.97 | 0.94 | 0.87 | 0.97 | 0.94 | 0.94 | 0.99 |
| GC | A1a | 2,2,4,7-tetramethyl-3H-1-benzofuran | 15.67 | < 0.05* | 0.74 | 1.58 | 0.21 | 0.94 | 12.58 | < 0.05* | 0.31 |
| GC | A1a | Propylidene-4,4-ethylenedioxycyclohexane | 0.73 | 0.75 | 0.97 | 0.80 | 0.60 | 0.96 | 1.90 | 0.46 | 0.90 |
| GC | A1a | 4-[(7-Fluoro-2,3-dihydro-1,4-benzodioxin-2-yl)methyl]morpholine | 0.95 | 0.95 | 1.00 | 1.12 | 0.68 | 0.96 | 3.66 | 0.08 | 0.77 |
| GC | A1a | (2,4,4-Trimethyl-6-oxo-cyclohex-1-enyl)-acetic acid ethyl ester | 0.73 | 0.43 | 0.97 | 0.89 | 0.49 | 0.95 | 0.69 | 0.39 | 0.89 |
| GC | A1a | 2-but-2-ynoxyoxane | 2.04 | 0.14 | 0.95 | 0.92 | 0.67 | 0.96 | 1.64 | 0.22 | 0.82 |
| GC | A1a | 2-(4-Chlorophenyl)-2-oxoethyl oct-2-ynoate | 0.59 | 0.33 | 0.97 | 1.64 | < 0.05* | 0.84 | 0.92 | 0.83 | 0.98 |
| GC | A1a | (1S)-3-(3'a-Hydroxy-7'a-methyl-5'-oxo-perhydroinden-1'-yl] acrylate | 2.24 | 0.07 | 0.95 | 1.25 | 0.29 | 0.95 | 1.52 | 0.22 | 0.82 |
| GC | A1a | .alpha.-Fluoro-(p-methyl)chalcone | 0.81 | 0.54 | 0.97 | 1.03 | 0.89 | 0.98 | 0.91 | 0.77 | 0.97 |
| GC | A1a | Phenyl salicylate | 1.62 | 0.54 | 0.97 | 0.75 | 0.35 | 0.95 | 3.22 | 0.08 | 0.77 |
| GC | A1a | 2,2'-Methylenebis(4-methyl-6-tert-butylphenol) | 1.06 | 0.75 | 0.97 | 1.26 | < 0.05* | 0.84 | 1.11 | 0.62 | 0.92 |
| GC | A1a | (4-nitrophenyl) 2-methylpropanoate | 0.57 | 0.61 | 0.97 | 0.75 | 0.39 | 0.95 | 7.16 | 0.17 | 0.82 |
| GC | A1a | 1,3-diethyl-1,3,3a,9a-tetrahydrobenzo[f][2]benzothiole-4,9-dione | 0.53 | 0.14 | 0.95 | 0.81 | 0.20 | 0.94 | 0.92 | 0.77 | 0.96 |
| GC | A1a | 7-anti-pentyl-3,3-dimethylnorbornano-1,2-endo-diol | 1.19 | 0.69 | 0.97 | 1.05 | 0.70 | 0.96 | 1.95 | 0.17 | 0.82 |
| GC | A1a | phenyl N-methylcarbamate | 3.81 | 0.06 | 0.95 | 2.09 | < 0.05* | 0.84 | 1.52 | 0.43 | 0.90 |
| GC | A1a | 15-(methoxymethoxy)-3-methylene-1-pentadecanol | 0.91 | 0.69 | 0.97 | 0.86 | 0.08 | 0.93 | 0.88 | 0.49 | 0.90 |
| GC | A1a | 2,7-Dimethoxy-1,3,4,5,6,8-hexamethyl-9-isopropyl-9H-xanthene | 1.06 | 0.82 | 0.97 | 1.07 | 0.66 | 0.96 | 1.14 | 0.68 | 0.93 |
| GC | A1a | 5-Methyl-2-(1H-pyrrol-2-yl)benzo[d]oxazole | 1.25 | 0.68 | 0.97 | 1.31 | 0.23 | 0.94 | 1.37 | 0.54 | 0.91 |
| GC | A1a | Pyracarbolid | 0.95 | 0.88 | 0.99 | 1.00 | 0.99 | 1.00 | 0.77 | 0.43 | 0.90 |
| GC | A1a | 2,2,4-Trimethylpentane-1,3-diyl dibenzoate | 0.68 | 0.17 | 0.95 | 0.90 | 0.37 | 0.95 | 1.23 | 0.48 | 0.90 |
| GC | A1a | 2-Phenyl[4,7]phenanthrolino[5,6-d]oxazole | 1.63 | 0.73 | 0.97 | 0.90 | 0.87 | 0.97 | 3.58 | 0.32 | 0.87 |
| GC | A1a | 4-(1,1,4,4,7-pentamethyltetralin-6-yl)sulfonylbenzoic acid | 0.71 | 0.75 | 0.97 | 0.69 | 0.31 | 0.95 | 1.39 | 0.65 | 0.92 |
| GC | A1a | (1-Methoxypentyl)benzene | 9.33 | 0.12 | 0.95 | 0.49 | 0.25 | 0.94 | 0.87 | 0.91 | 0.99 |
| GC | A1a | Methyl 3-Methyl-5-pentanoyl-2-furoate | 1.54 | 0.65 | 0.97 | 1.36 | 0.34 | 0.95 | 0.47 | 0.22 | 0.82 |
| GC | A1a | 2-(12'-methoxy-19'-norpodocarpa-8',11',13-trien-18'-yl)ethyl methanesulfonate | 1.91 | 0.16 | 0.95 | 0.99 | 0.98 | 1.00 | 1.63 | 0.32 | 0.87 |
| GC | A1a | 2,9-dihydro-\$b-carbolin-1-one | 1.22 | 0.40 | 0.97 | 0.91 | 0.85 | 0.97 | 0.79 | 0.80 | 0.97 |
| GC | A1a | 4-(1H-pyrazolyl-1-yl)indol-2-one | 0.86 | 0.62 | 0.97 | 0.82 | 0.17 | 0.94 | 2.16 | < 0.05* | 0.51 |
| GC | A1a | (8R,9R)-Isocaryolane-8,9-diol | 0.79 | 0.51 | 0.97 | 0.84 | 0.26 | 0.94 | 0.62 | 0.19 | 0.82 |
| GC | A1a | methyl (E)-4-(1-hydroxy-3,4-dihydro-2H-triphenylen-1-yl)but-2-enoate | 1.59 | 0.50 | 0.97 | 0.87 | 0.71 | 0.96 | 1.97 | 0.31 | 0.87 |
| GC | A1a | Bis(2-ethylhexyl) phthalate | 1.15 | 0.70 | 0.97 | 1.03 | 0.87 | 0.97 | 1.53 | 0.28 | 0.86 |
| GC | A1a | 1,3-dimethyl-5-(2-methylpropylidene)-1,3-diazinane-2,4,6-trione | 1.44 | 0.49 | 0.97 | 1.26 | 0.48 | 0.95 | 1.17 | 0.81 | 0.97 |
| GC | A1a | alpha-Tocopherol | 0.77 | 0.49 | 0.97 | 0.96 | 0.81 | 0.97 | 1.03 | 0.91 | 0.99 |
| GC | A1a | (2-Hydroxyphenyl)acetonitrile | 1.74 | 0.39 | 0.97 | 1.39 | 0.24 | 0.94 | 1.56 | 0.52 | 0.91 |
| GC | A1a | 5-phenyl-3,4-dihydro-2H-pyrrole | 4.03 | 0.21 | 0.95 | 1.57 | 0.34 | 0.95 | 5.60 | 0.06 | 0.77 |
| GC | A1a | methyl 3-[(1R,12bR)-3-acetyl-1,2,6,7,12,12b-hexahydroindolo[2,3-a]quinolizin-1-yl]-2-chloropropanoate | 0.59 | 0.43 | 0.97 | 1.10 | 0.76 | 0.97 | 2.92 | 0.15 | 0.82 |
| GC | A1a | (5S,6R)-5,6-Dihydroxy-cyclohex-3-enecarboxylic acid prop-2-ynyl ester | 1.70 | 0.34 | 0.97 | 1.10 | 0.71 | 0.96 | 1.56 | 0.37 | 0.89 |
| GC | A1a | Octocrylene | 1.07 | 0.71 | 0.97 | 0.92 | 0.46 | 0.95 | 0.76 | 0.32 | 0.87 |
| GC | A1a | 4-Methyl-4-(2-oxanyloxy)-2-pentanone | 3.69 | < 0.05* | 0.95 | 0.70 | 0.09 | 0.93 | 1.32 | 0.58 | 0.92 |
| GC | A1a | 1-(cyclopentylmethyl)-4-methoxy-benzene | 1.05 | 0.96 | 1.00 | 0.35 | < 0.05* | 0.84 | 1.23 | 0.81 | 0.97 |
| GC | A1a | Triethylene glycol diacetate | 1.10 | 0.69 | 0.97 | 0.93 | 0.45 | 0.95 | 0.86 | 0.47 | 0.90 |
| GC | A1a | (2,6-dimethyl-4-propyl-phenyl)amine | 0.60 | 0.54 | 0.97 | 1.63 | 0.31 | 0.95 | 0.69 | 0.53 | 0.91 |
| GC | A1a | 2-O-cyclohexyl 1-O-pentyl benzene-1,2-dicarboxylate | 0.79 | 0.43 | 0.97 | 0.99 | 0.98 | 1.00 | 1.07 | 0.72 | 0.94 |
| GC | A1a | 3-(2-(Piperidin-1-yl)acetamido)benzoic acid | 0.66 | 0.48 | 0.97 | 1.02 | 0.94 | 0.99 | 0.66 | 0.40 | 0.89 |
| GC | A1a | Morpholyl-2-oxocyclohexanecarboxamide | 1.14 | 0.72 | 0.97 | 0.96 | 0.76 | 0.97 | 0.65 | 0.17 | 0.82 |
| GC | A1a | 1-Methoxy-1-(4-methoxyphenyl)-4-phenylbutane | 1.08 | 0.94 | 1.00 | 0.57 | 0.23 | 0.94 | 0.97 | 0.98 | 0.99 |
| GC | A1a | 3,5,5-trimethyl-4-[(E)-3-oxidanylbut-1-enyl]cyclohex-3-en-1-ol | 1.16 | 0.81 | 0.97 | 0.75 | 0.24 | 0.94 | 0.53 | 0.30 | 0.87 |
| GC | A1a | Kikkanol F monoacetate | 0.59 | 0.35 | 0.97 | 0.73 | 0.26 | 0.94 | 0.50 | 0.25 | 0.84 |
| GC | A1a | 5,12-Dihydroxysterpyrene | 1.27 | 0.70 | 0.97 | 0.63 | 0.08 | 0.93 | 1.91 | 0.20 | 0.82 |
| GC | A1a | 4-Methylene-2-phenyl-tetrahydrofuran | 2.89 | 0.22 | 0.95 | 1.08 | 0.78 | 0.97 | 1.14 | 0.81 | 0.97 |
| GC | A1a | Phenyl benzoate | 0.52 | 0.61 | 0.97 | 1.59 | 0.47 | 0.95 | 4.58 | 0.33 | 0.87 |
| GC | A1a | 2-Isobutyryl-benzo[1,3]dioxol-5-carbaldehyde | 1.01 | 0.99 | 1.00 | 1.26 | 0.58 | 0.96 | 2.04 | 0.37 | 0.89 |
| GC | A1a | DL-trans-2-acetoxy-1-benzoyloxymethyl-3-[(E)-cinnamoyloxy]-4,6-cyclohexadiene | 1.77 | 0.27 | 0.97 | 0.79 | 0.23 | 0.94 | 0.70 | 0.24 | 0.84 |
| GC | A1a | methyl 2-hydroxy-2-(3-methoxyphenyl)acetate | 1.12 | 0.64 | 0.97 | 0.91 | 0.52 | 0.95 | 0.78 | 0.38 | 0.89 |
| GC | A1a | Methyl 2-(3',3'-dimethyl-1'-butyn-1'-yl)-1-cyclohexenecarboxylate | 0.79 | 0.81 | 0.97 | 2.06 | 0.09 | 0.93 | 0.56 | 0.44 | 0.90 |
| GC | A1a | 3,4-dihydro-2H-naphthalen-1-one | 0.87 | 0.86 | 0.99 | 0.77 | 0.49 | 0.95 | 9.71 | < 0.05* | 0.51 |
| GC | A1a | 4-(1-Pyrrolidinyl)pyridazine | 0.75 | 0.60 | 0.97 | 0.74 | 0.35 | 0.95 | 2.41 | 0.06 | 0.76 |
| GC | A1a | Phenyl N-benzoyl-N-methyl-thiooxocarbamate | 1.03 | 0.95 | 1.00 | 1.02 | 0.90 | 0.98 | 1.60 | 0.25 | 0.84 |
| GC | A1a | Furaltadone | 0.28 | 0.24 | 0.97 | 1.18 | 0.70 | 0.96 | 0.46 | 0.30 | 0.86 |
| GC | A1a | Benzyl butyl phthalate | 1.25 | 0.65 | 0.97 | 1.15 | 0.43 | 0.95 | 1.42 | 0.41 | 0.90 |
| GC | A1a | 3-benzyl-4,7,7-trimethyl-3-azabicyclo[2.2.1]heptan-2-one | 2.03 | 0.48 | 0.97 | 0.66 | 0.48 | 0.95 | 1.63 | 0.68 | 0.93 |
| GC | A1a | Dicrotophos | 1.32 | 0.75 | 0.97 | 0.85 | 0.52 | 0.95 | 0.97 | 0.94 | 0.99 |
| GC | A1a | 1-ethyl-3-[(E)-hydroxyiminomethyl]pyridine-2-thione | 0.24 | 0.29 | 0.97 | 1.20 | 0.74 | 0.96 | 0.33 | 0.23 | 0.82 |
| GC | A1a | (R)-6-Phenyltetrahydro-2H-pyran-2-one | 0.22 | 0.05 | 0.95 | 1.66 | 0.10 | 0.93 | 1.34 | 0.62 | 0.92 |
| GC | A1a | Beta-BHC | 0.52 | 0.39 | 0.97 | 0.81 | 0.48 | 0.95 | 1.75 | 0.39 | 0.89 |
| GC | A1a | o-Phenoxybenzyl Alcohol | 2.91 | 0.36 | 0.97 | 1.65 | 0.27 | 0.94 | 4.57 | 0.24 | 0.84 |
| GC | A1a | 2,6-Diisopropylnaphthalene | 1.64 | 0.44 | 0.97 | 1.18 | 0.84 | 0.97 | 14.60 | 0.10 | 0.77 |
| GC | A1a | 4-[[[2-(Dimethylamino)ethyl]methylamino]methyl]phenol | 1.07 | 0.88 | 0.99 | 0.98 | 0.89 | 0.98 | 1.60 | 0.26 | 0.84 |
| GC | A1a | Benzothiazolone | 0.98 | 0.94 | 1.00 | 1.04 | 0.82 | 0.97 | 1.44 | 0.22 | 0.82 |
| GC | A1a | 2-[(4S)-2,2,4-trimethyl-1,3-dioxolan-4-yl]ethyl methanesulfonate | 0.95 | 0.90 | 0.99 | 0.92 | 0.60 | 0.96 | 1.25 | 0.55 | 0.92 |
| GC | A1a | Fluorene | 1.71 | 0.46 | 0.97 | 0.92 | 0.85 | 0.97 | 0.79 | 0.73 | 0.94 |
| GC | A1a | 8,9,11,12-Tetraimethoxy-1-methyl-1,2,3,4-dihydronaphtho[2,1-f]isoquinoline | 2.20 | 0.14 | 0.95 | 0.82 | 0.36 | 0.95 | 1.41 | 0.43 | 0.90 |
| GC | A1a | 1-(4-Chlorophenyl)-2-methylbut-3-en-1-ol | 2.58 | 0.07 | 0.95 | 1.05 | 0.84 | 0.97 | 2.52 | 0.14 | 0.82 |
| GC | A1a | n-Undecyl cyanide | 2.31 | 0.48 | 0.97 | 1.46 | 0.51 | 0.95 | 0.97 | 0.98 | 0.99 |
| GC | A1a | 4-[(2-methylacridin-9-yl)amino]-N-(1,3-thiazol-2-yl)benzenesulfonamide | 3.59 | 0.11 | 0.95 | 1.04 | 0.84 | 0.97 | 0.80 | 0.62 | 0.92 |
| GC | A1a | Octisalate | 0.78 | 0.87 | 0.99 | 1.61 | 0.54 | 0.95 | 1.18 | 0.92 | 0.99 |
| GC | A1a | 2-(4-Fluorophenyl)isoindoline | 1.19 | 0.50 | 0.97 | 1.47 | 0.16 | 0.94 | 1.71 | 0.27 | 0.84 |
| GC | A1a | Ethylparaben | 1.00 | 1.00 | 1.00 | 0.97 | 0.96 | 1.00 | 4.41 | 0.20 | 0.82 |
| GC | A1a | (2S,6R)-4-[3-(4-tert-butylphenyl)-2-methylpropyl]-2,6-dimethylmorpholine | 0.50 | 0.57 | 0.97 | 2.78 | 0.07 | 0.93 | 0.66 | 0.61 | 0.92 |
| GC | A1a | Octa-1,7-diyne-3,6-diol | 1.47 | 0.45 | 0.97 | 1.14 | 0.57 | 0.96 | 1.94 | 0.21 | 0.82 |
| GC | A1a | (E)-stilbene | 0.91 | 0.82 | 0.97 | 1.07 | 0.68 | 0.96 | 1.18 | 0.43 | 0.90 |
| GC | A1a | 2-butyl-6-methoxy-2,5-dihydropyridine | 1.43 | 0.35 | 0.97 | 1.23 | 0.29 | 0.95 | 0.48 | 0.10 | 0.77 |
| GC | A1a | 2-Methoxy-N,N-dimethyl-2-phenylacetamide | 1.02 | 0.97 | 1.00 | 1.25 | 0.33 | 0.95 | 1.97 | 0.13 | 0.82 |
| GC | A1a | .beta.-1,5-anhydro-2,3,4,6-tetra-O-methyl-1-C-phenyl-D-glucitol | 0.68 | 0.33 | 0.97 | 0.76 | 0.10 | 0.93 | 0.79 | 0.53 | 0.91 |
| GC | A1a | 1-phenyl-2-sulfanylidene-3-pyridinecarboxylic acid | 2.28 | 0.65 | 0.97 | 1.24 | 0.59 | 0.96 | 1.08 | 0.90 | 0.99 |
| GC | A1a | Allyltri(4-N,N-dimethylaminophenyl)silane | 0.67 | 0.44 | 0.97 | 0.71 | 0.14 | 0.94 | 0.76 | 0.58 | 0.92 |
| GC | A1a | 7,10-Dioxo-8,9-dihydro[4]-paracyclophane | 0.19 | 0.10 | 0.95 | 0.78 | 0.48 | 0.95 | 1.95 | 0.27 | 0.84 |
| GC | A1a | Pyrolo[3,2-d]pyrimidin-2,4(1H,3H)-dione | 1.77 | 0.21 | 0.95 | 1.05 | 0.86 | 0.97 | 0.94 | 0.88 | 0.99 |
| GC | A1a | Mepivacaine | 1.31 | 0.15 | 0.95 | 1.22 | 0.21 | 0.94 | 1.35 | 0.36 | 0.89 |
| GC | A1a | 4-Chlorophenol | 3.78 | 0.10 | 0.95 | 1.11 | 0.77 | 0.97 | 0.18 | < 0.05* | 0.66 |
| GC | A1a | N-[(E)-3-cyano-1,1-dimethyl-allyl]acetamide | 1.16 | 0.85 | 0.98 | 0.85 | 0.56 | 0.95 | 0.53 | 0.44 | 0.90 |
| GC | A1a | Acetic Acid (+-)-(1.alpha.,3.beta.,4.beta.,6.alpha.,7.alpha.)-7-(3-Methyl-3-butenyl)-4-((E)-6-methyl-2,6-heptadienyl)-1-methylbicyclo[4.1.0]hept-3-yl Ester | 0.55 | 0.17 | 0.95 | 1.07 | 0.60 | 0.96 | 0.73 | 0.19 | 0.82 |
| GC | A1a | 2-Amino-2'-tert-butylthio-6-ethoxy[3,4']bipyridine-3,5-dicarbonitrile | 2.28 | 0.07 | 0.95 | 1.35 | 0.15 | 0.94 | 1.91 | 0.08 | 0.77 |
| GC | A1a | Pyrene | 2.09 | 0.48 | 0.97 | 0.51 | 0.12 | 0.93 | 0.96 | 0.94 | 0.99 |
| GC | A1a | trans-2,3-di(methoxycarbonyl)-bicyclo[2.2.2]oct-5-ene | 0.43 | 0.43 | 0.97 | 0.71 | 0.43 | 0.95 | 1.08 | 0.92 | 0.99 |
| GC | A1a | 3-(4-Vinylbenzyloxy)benzyl alcohol | 7.84 | 0.40 | 0.97 | 1.39 | 0.74 | 0.96 | 0.09 | 0.14 | 0.82 |
| GC | A1a | methyl 2-pyrimidin-4-ylpropanoate | 0.84 | 0.75 | 0.97 | 1.24 | 0.40 | 0.95 | 2.99 | < 0.05* | 0.61 |
| GC | A1a | 2-methyl-1H-benzimidazole | 1.72 | 0.45 | 0.97 | 1.67 | 0.15 | 0.94 | 1.01 | 0.99 | 0.99 |
| GC | A1a | (R)-2-N-Heptyl (S)-O-1-phenylethyl carbamate | 0.90 | 0.91 | 0.99 | 0.86 | 0.72 | 0.96 | 0.82 | 0.80 | 0.97 |
| GC | A1a | ethyl (2E,4E)-6-methoxy-6-sulfanylidenehexa-2,4-dienoate | 2.23 | 0.33 | 0.97 | 1.23 | 0.56 | 0.95 | 0.65 | 0.53 | 0.91 |
| GC | A1a | 1-(Biphenyl-4-yl)ethanone | 1.11 | 0.82 | 0.97 | 1.09 | 0.49 | 0.95 | 0.83 | 0.28 | 0.85 |
| GC | A1a | ethyl 2-((2r,4R,6S)-2,4,6-trimethyl-1,3-dioxan-2-yl)acetate | 0.62 | 0.17 | 0.95 | 1.13 | 0.38 | 0.95 | 1.02 | 0.96 | 0.99 |
| GC | A1a | 2-methyl-5-phenyl-1H-pyrrole | 4.26 | < 0.05* | 0.95 | 1.20 | 0.41 | 0.95 | 8.31 | < 0.001** | 0.06 |
| GC | A1a | (6R,7R)-7-isopropyl-1,3-dihydro-1,5,5,6-tetramethyl-indan[5,6-c]furan | 0.82 | 0.54 | 0.97 | 1.18 | 0.19 | 0.94 | 1.15 | 0.71 | 0.94 |
| GC | A1a | 3,3,3-trifluoro-2-hydroxy-1-(4-methylphenyl)-1-propanone | 0.94 | 0.90 | 0.99 | 1.50 | 0.15 | 0.94 | 0.96 | 0.93 | 0.99 |
| GC | A1a | (4-methoxy-3-propan-2-yloxy-phenyl) ethanoate | 2.74 | 0.18 | 0.95 | 1.23 | 0.53 | 0.95 | 1.12 | 0.87 | 0.99 |
| GC | A1a | benzhydryl 2-(1-methyl-3-oxo-2-prop-2-enylcyclohexyl)acetate | 0.64 | 0.39 | 0.97 | 1.04 | 0.87 | 0.97 | 1.51 | 0.32 | 0.87 |
| GC | A1a | 9-Methyl-9,10-dihydroanthracene | 2.76 | 0.39 | 0.97 | 1.54 | 0.23 | 0.94 | 2.24 | 0.46 | 0.90 |
| GC | A1a | (3aR,5R,7aR)-1,1-Ethylidenedioxy-7a-methyl-3a,4,5,6,7,7a-hexahydroindan-5-ol | 0.83 | 0.75 | 0.97 | 1.00 | 0.99 | 1.00 | 0.45 | 0.18 | 0.82 |
| GC | A1a | N-(cyclohexylmethyl)morpholine | 1.63 | 0.33 | 0.97 | 0.97 | 0.89 | 0.98 | 1.09 | 0.86 | 0.99 |
| GC | A1a | (E)-.beta.-(tert-Butyldimethylsiloxy)-.beta.-(tert-butyldimethylsilyl)styrene | 0.35 | 0.18 | 0.95 | 1.00 | 0.99 | 1.00 | 14.86 | < 0.05* | 0.51 |
| GC | A1a | 6-Heptadecylthymoquinone | 0.93 | 0.80 | 0.97 | 1.11 | 0.38 | 0.95 | 1.13 | 0.57 | 0.92 |
| GC | A1a | 2,5-Pyrrolidinedione, 1-[11-(4-morpholinyl)bicyclo[8.1.0]undec-11-yl]-, stereoisomer | 0.71 | 0.81 | 0.97 | 1.78 | 0.32 | 0.95 | 5.36 | 0.16 | 0.82 |
| GC | A1a | (Z)-3-(4-n-Butyl-3-thienyl)propenal | 2.01 | 0.27 | 0.97 | 0.79 | 0.42 | 0.95 | 1.38 | 0.48 | 0.90 |
| GC | A1a | (S)-(4-butoxyphenyl)(1-phthalimdoethyl)-ketone | 0.76 | 0.36 | 0.97 | 1.17 | 0.33 | 0.95 | 1.06 | 0.84 | 0.98 |
| GC | A1a | Acetyl tributyl citrate | 1.01 | 0.97 | 1.00 | 0.92 | 0.53 | 0.95 | 0.83 | 0.63 | 0.92 |
| GC | A1a | (4S,5R)-3-(3-methyl-1-oxobut-2-enyl)-4,5-diphenyl-2-oxazolidinone | 0.33 | 0.35 | 0.97 | 1.15 | 0.83 | 0.97 | 1.11 | 0.94 | 0.99 |
| GC | A1a | 3-Isopropoxy-5-methyl-1H-pyrazole | 1.16 | 0.87 | 0.99 | 1.01 | 0.98 | 1.00 | 0.39 | 0.43 | 0.90 |
| GC | A1a | N'-(2-methylpropyl)benzohydrazide | 0.86 | 0.73 | 0.97 | 1.73 | 0.14 | 0.94 | 0.97 | 0.96 | 0.99 |
| GC | A1a | 3-Butanoyloxy-2-(2-propenyl)tetrahydropyran | 1.00 | 1.00 | 1.00 | 0.82 | 0.49 | 0.95 | 2.39 | 0.18 | 0.82 |
| GC | A1a | 2-Phenoxyethyl propanoate | 1.08 | 0.90 | 0.99 | 0.84 | 0.64 | 0.96 | 1.31 | 0.68 | 0.93 |
| GC | A1a | Molinate | 1.21 | 0.49 | 0.97 | 1.14 | 0.49 | 0.95 | 0.91 | 0.80 | 0.97 |
| GC | A1a | 1-Decyclopentane-1,3-dione | 1.68 | 0.33 | 0.97 | 1.14 | 0.59 | 0.96 | 1.67 | 0.30 | 0.87 |
| GC | A1a | 1-(4-phenyl-1H-pyrrol-3-yl)propan-1-one | 0.72 | 0.58 | 0.97 | 0.95 | 0.85 | 0.97 | 0.64 | 0.35 | 0.88 |
| GC | A1a | Beta-Tocopherol | 0.93 | 0.68 | 0.97 | 1.09 | 0.37 | 0.95 | 0.94 | 0.69 | 0.93 |
| GC | A1a | N-pyridin-2-ylnitramide | 1.33 | 0.40 | 0.97 | 0.85 | 0.26 | 0.94 | 1.87 | 0.09 | 0.77 |
| GC | A1a | 2,2',3,4,4',5',6-Heptachlorobiphenyl | 0.77 | 0.55 | 0.97 | 1.13 | 0.54 | 0.95 | 0.70 | 0.49 | 0.90 |
| GC | A1a | Triethyl citrate | 1.11 | 0.82 | 0.97 | 0.83 | 0.39 | 0.95 | 1.04 | 0.91 | 0.99 |
| GC | A1a | (3R,8S,9S,10R,13S,14S,17S)-17-[(1S)-1-(dimethylamino)ethyl]-N-ethyl-N,10,13-trimethyl-2,3,4,7,8,9,11,12,14,15,16,17-dodecahydro-1H-cyclopenta[a]phenanthren-3-amine | 0.47 | 0.26 | 0.97 | 0.86 | 0.64 | 0.96 | 0.34 | 0.16 | 0.82 |
| GC | A1a | (2E,7E)-5-hydroxy-6,6,9-trimethyl-4-deca-2,7,9-trienone | 1.67 | 0.47 | 0.97 | 1.20 | 0.51 | 0.95 | 1.71 | 0.36 | 0.89 |
| GC | A1a | 2-Methyl-3-(trifluoromethyl)-1,4-naphthoquinone | 0.10 | 0.08 | 0.95 | 1.11 | 0.84 | 0.97 | 1.13 | 0.91 | 0.99 |
| GC | A1a | 1,3,5-Trimethyladamantane | 0.72 | 0.57 | 0.97 | 1.05 | 0.84 | 0.97 | 0.40 | 0.14 | 0.82 |
| GC | A1a | (1S)-1-[(3S,5S,8R,9S,10S,13S,14S,17S)-10,13-dimethyl-3-(propan-2-ylideneamino)-2,3,4,5,6,7,8,9,11,12,14,15,16,17-tetradecahydro-1H-cyclopenta[a]phenanthren-17-yl]-N,N-dimethylethanamine | 1.17 | 0.65 | 0.97 | 0.83 | 0.21 | 0.94 | 0.82 | 0.51 | 0.91 |
| GC | A1a | 2-Phenylethyl propionate | 0.71 | 0.15 | 0.95 | 0.97 | 0.71 | 0.96 | 0.83 | 0.33 | 0.87 |
| GC | A1a | (trans)-2-(Hydroxymethyl)-3-phenyltetrahydrofuran | 0.66 | 0.54 | 0.97 | 1.02 | 0.96 | 1.00 | 0.55 | 0.30 | 0.86 |
| GC | A1a | PCB 130 | 1.24 | 0.40 | 0.97 | 1.10 | 0.38 | 0.95 | 1.12 | 0.62 | 0.92 |
| GC | A1a | N,N-dimethyl-4-phenylbutanamide | 0.51 | 0.08 | 0.95 | 1.28 | 0.17 | 0.94 | 1.24 | 0.49 | 0.90 |
| GC | A1a | 2',6'-Dichloro-2,4-dimethylbenzanilide | 1.01 | 0.97 | 1.00 | 1.45 | 0.08 | 0.93 | 0.90 | 0.78 | 0.97 |
| GC | A1a | 4-(2-thenoyl)tetrahydrofuran-2-one | 0.82 | 0.89 | 0.99 | 1.23 | 0.78 | 0.97 | 4.79 | 0.37 | 0.89 |
| GC | A1a | 4a-Methyl-4,4a,5,6-tetrahydro-2(3H),7-indendione | 1.51 | 0.45 | 0.97 | 1.07 | 0.74 | 0.96 | 0.64 | 0.45 | 0.90 |
| GC | A1a | 1-methylquinolin-2(1H)-one | 37.34 | < 0.05* | 0.95 | 1.16 | 0.83 | 0.97 | 0.62 | 0.65 | 0.92 |
| GC | A1a | (+)-Pinidine | 2.76 | < 0.05* | 0.95 | 0.90 | 0.68 | 0.96 | 1.82 | 0.27 | 0.84 |
| GC | A1a | (3R*,4S*)-3-Ethyl-4-(trimethylsilyethynyl)azitidin-2-one | 0.81 | 0.68 | 0.97 | 1.27 | 0.25 | 0.94 | 1.88 | 0.19 | 0.82 |
| GC | A1a | 3-Methoxyphthalide | 1.36 | 0.31 | 0.97 | 1.18 | 0.63 | 0.96 | 4.26 | 0.11 | 0.78 |
| GC | A1a | methyloctyldiethoxysilane | 0.54 | 0.33 | 0.97 | 0.98 | 0.94 | 0.99 | 0.45 | 0.11 | 0.78 |
| GC | A1a | 1-Amino-3,4-dihydro-3,4-dimethyl-2-naphthalenecarbonitrile | 2.25 | 0.27 | 0.97 | 0.90 | 0.65 | 0.96 | 3.05 | 0.08 | 0.77 |
| GC | A1a | 2-Naphthalenamine | 0.50 | 0.26 | 0.97 | 1.06 | 0.85 | 0.97 | 1.21 | 0.74 | 0.95 |
| GC | A1a | 1,3-trans-4-Hydroxymethyl-1-methylcyclopentane-1,3-diol | 1.22 | 0.56 | 0.97 | 1.49 | < 0.05* | 0.84 | 1.97 | 0.09 | 0.77 |
| GC | A1a | 1-methoxy-4-[(E)-2-phenylethenyl]benzene | 0.87 | 0.14 | 0.95 | 1.06 | 0.33 | 0.95 | 1.00 | 1.00 | 1.00 |
| GC | A1a | 2-methyl-1,3-benzoxazole | 0.88 | 0.83 | 0.97 | 0.93 | 0.49 | 0.95 | 1.14 | 0.82 | 0.98 |
| GC | A1a | Diisobutyl phthalate | 1.29 | 0.64 | 0.97 | 0.75 | 0.20 | 0.94 | 1.01 | 0.97 | 0.99 |
| GC | A1a | 8-methylpyrrolo[1,2-a]quinoxaline | 0.37 | 0.12 | 0.95 | 1.22 | 0.47 | 0.95 | 1.38 | 0.59 | 0.92 |
| GC | A1a | 1-(2,4-Dimethylphenyl)but-3-en-1-ol | 1.13 | 0.88 | 0.99 | 0.93 | 0.89 | 0.98 | 3.57 | 0.21 | 0.82 |
| GC | A1a | 1,4,5,8-Tetramethylnaphthalene | 0.60 | 0.67 | 0.97 | 1.49 | 0.19 | 0.94 | 1.01 | 0.98 | 0.99 |
| GC | A1a | N-tert-Butylmaleimide | 1.52 | 0.22 | 0.95 | 0.99 | 0.92 | 0.99 | 0.63 | 0.16 | 0.82 |
| GC | A1a | 1-Piperidino-5-(2-hydroxy-4-nitrophenoxy)pentane | 0.38 | 0.16 | 0.95 | 0.58 | 0.07 | 0.93 | 0.48 | 0.19 | 0.82 |
| GC | A1a | Diisopropyl phthalate | 0.87 | 0.83 | 0.97 | 1.11 | 0.65 | 0.96 | 1.21 | 0.49 | 0.90 |
| GC | A1a | Kanzonol P | 0.93 | 0.95 | 1.00 | 0.97 | 0.94 | 0.99 | 0.96 | 0.95 | 0.99 |
| GC | A1a | Hexachlorobenzene | 0.88 | 0.56 | 0.97 | 1.08 | 0.53 | 0.95 | 0.81 | 0.38 | 0.89 |
| GC | A1a | 5-Vinyloxyhept-6-enyl 2,2-dimethylpropanoate | 0.87 | 0.80 | 0.97 | 0.86 | 0.47 | 0.95 | 0.74 | 0.35 | 0.88 |
| GC | A1a | 1-Piperidino-5-(4-amino-2-methoxyphenoxy)ethane | 0.77 | 0.33 | 0.97 | 0.97 | 0.81 | 0.97 | 0.41 | < 0.05* | 0.51 |
| GC | A1a | 1-chloro-4-[2,2,2-trichloro-1-(4-chlorophenyl)ethyl]benzene | 1.00 | 1.00 | 1.00 | 1.02 | 0.93 | 0.99 | 2.04 | 0.19 | 0.82 |
| GC | A1a | 3-(4-ketohexyl)cyclohex-2-en-1-one | 1.74 | 0.78 | 0.97 | 0.83 | 0.84 | 0.97 | 0.61 | 0.78 | 0.97 |
| GC | A1a | Tris(2-butoxyethyl) phosphate | 1.31 | 0.46 | 0.97 | 1.27 | 0.11 | 0.93 | 0.94 | 0.82 | 0.98 |
| GC | A1a | 2-(4-Methoxyphenoxy)benzothiophene | 3.58 | 0.25 | 0.97 | 0.59 | 0.20 | 0.94 | 2.87 | 0.45 | 0.90 |
| GC | A1a | Phenanthrene | 0.51 | 0.67 | 0.97 | 2.09 | 0.42 | 0.95 | 0.28 | 0.44 | 0.90 |
| GC | A1a | methyl 2-(5-methoxy-1,2-dimethylindol-3-yl)acetate | 1.15 | 0.76 | 0.97 | 0.82 | 0.26 | 0.94 | 0.90 | 0.79 | 0.97 |
| GC | A1a | 2'-Hydroxy-4',6'-dimethoxyacetophenone | 0.68 | 0.53 | 0.97 | 0.77 | 0.42 | 0.95 | 0.50 | 0.19 | 0.82 |
| GC | A1a | 2,2',3,4,4',5,5'-Heptachlorobiphenyl | 1.02 | 0.90 | 0.99 | 1.10 | 0.21 | 0.94 | 1.11 | 0.46 | 0.90 |
| GC | A1a | 3-Hexene-2,5-dione, 3-acetyl-, 5-(dimethylhydrazone) | 1.59 | 0.36 | 0.97 | 0.79 | 0.27 | 0.94 | 0.45 | 0.10 | 0.77 |
| GC | A1a | Captafol | 0.94 | 0.88 | 0.99 | 0.86 | 0.33 | 0.95 | 0.75 | 0.23 | 0.82 |
| GC | A1a | N-(2-Fluorobenzylidene)-2-fluorophenylmethylamine | 0.91 | 0.78 | 0.97 | 0.82 | 0.19 | 0.94 | 1.06 | 0.86 | 0.99 |
| GC | A1a | (2-ethyl-4,6-dimethylphenyl)methanol | 3.56 | 0.19 | 0.95 | 1.22 | 0.66 | 0.96 | 2.75 | 0.37 | 0.89 |
| GC | A1a | 2,2-Dimethyl-2H-chromene-6-carbaldehyde | 0.50 | < 0.05* | 0.95 | 0.93 | 0.65 | 0.96 | 1.21 | 0.59 | 0.92 |
| GC | A1a | 3,3-Dimethyl-1,3-dihydro-1-isobenzofuranone | 1.48 | 0.78 | 0.97 | 2.75 | 0.06 | 0.93 | 2.93 | 0.41 | 0.90 |
| GC | A1a | 3-Butenyl-5-propylindolizidine | 0.60 | 0.41 | 0.97 | 1.00 | 1.00 | 1.00 | 3.03 | 0.11 | 0.78 |
| GC | A1a | Di-n-propylphthalate | 1.46 | 0.40 | 0.97 | 0.68 | < 0.05* | 0.84 | 1.14 | 0.72 | 0.94 |
| GC | A1a | 3,4-Diphenyl-6-methyl-7,8-(1,4-dimethoxybenzo)-9-oxatricyclo[4.2.1.0(2,5)]non-3-ene | 1.23 | 0.39 | 0.97 | 1.17 | 0.16 | 0.94 | 1.22 | 0.29 | 0.86 |
| GC | A1a | 5-bicyclo[2.2.2]oct-2-enecarbonitrile | 2.28 | 0.35 | 0.97 | 1.60 | 0.21 | 0.94 | 3.00 | 0.12 | 0.81 |
| GC | A1a | 1,6-dimethyl-4-propan-2-ylnaphthalene | 1.11 | 0.79 | 0.97 | 0.61 | < 0.05* | 0.84 | 0.73 | 0.53 | 0.91 |
| GC | A1a | 1,2-Dicyano-4,5-di[(N,N-dimethylaminocarbonyl)methoxy]benzene | 0.78 | 0.56 | 0.97 | 0.79 | 0.20 | 0.94 | 0.77 | 0.46 | 0.90 |
| GC | A1a | 4-Hydroxyacetophenone | 1.20 | 0.59 | 0.97 | 0.90 | 0.46 | 0.95 | 1.34 | 0.38 | 0.89 |
| GC | A1a | 2-(2,3-dihydro-1H-indol-4-yl)ethanol | 1.51 | 0.09 | 0.95 | 0.95 | 0.61 | 0.96 | 1.16 | 0.50 | 0.90 |
| GC | A1a | (Z)-2-((2-Methylthio)styrylthio)aniline | 0.76 | 0.37 | 0.97 | 0.97 | 0.74 | 0.96 | 1.01 | 0.98 | 0.99 |
| GC | A1a | Carbaryl | 0.97 | 0.96 | 1.00 | 1.15 | 0.72 | 0.96 | 0.67 | 0.66 | 0.93 |
| GC | A1a | 4-Butylaniline | 0.41 | 0.07 | 0.95 | 0.84 | 0.41 | 0.95 | 0.75 | 0.43 | 0.90 |
| GC | A1a | Dibenzofuran | 1.40 | 0.26 | 0.97 | 0.25 | < 0.05* | 0.84 | 2.18 | 0.49 | 0.90 |
| GC | A1a | 3-(2-methylbutyl)-1H-indole | 2.73 | 0.44 | 0.97 | 0.43 | 0.08 | 0.93 | 1.26 | 0.75 | 0.95 |
| GC | A1a | (2s)-n-tert-butoxycarbonyl-2-(4-(1-ethoxyethoxy)-1-hydroxybutyl)pyrrolidine | 1.00 | 1.00 | 1.00 | 0.72 | < 0.05* | 0.84 | 1.00 | 0.99 | 0.99 |
| GC | A1a | (2RS,4SR)-4-(dimethylamino)-2,4-diphenyl-2-butanol | 0.91 | 0.77 | 0.97 | 1.10 | 0.56 | 0.95 | 0.68 | 0.25 | 0.84 |
| GC | A1a | 2-ethynylbenzaldehyde | 1.81 | 0.10 | 0.95 | 0.89 | 0.66 | 0.96 | 0.44 | 0.17 | 0.82 |
| GC | A1a | 1-Ethyl-4,5,8-trimethylnaphthalene | 1.35 | 0.52 | 0.97 | 1.37 | 0.10 | 0.93 | 1.30 | 0.50 | 0.90 |
| GC | A1a | Bisphenol A | 0.87 | 0.73 | 0.97 | 1.04 | 0.84 | 0.97 | 2.07 | 0.10 | 0.77 |
| GC | A1a | N-(1,2-diphenyl-2-phenylsulfanylethyl)acetamide | 0.53 | 0.39 | 0.97 | 1.01 | 0.98 | 1.00 | 0.52 | 0.25 | 0.84 |
| GC | A1a | Dehydro-botrydienol | 1.16 | 0.62 | 0.97 | 0.90 | 0.55 | 0.95 | 1.18 | 0.61 | 0.92 |
| GC | A1a | 2-methyl-2-[(E)-2-methyl-3-phenylprop-2-enyl]-1,3-dioxolane | 1.49 | 0.32 | 0.97 | 0.70 | < 0.05* | 0.84 | 1.53 | 0.23 | 0.83 |
| GC | A1a | 5,5-Dicyano-2-methyl-3,4-dihydro-pyrrole | 1.18 | 0.80 | 0.97 | 1.06 | 0.84 | 0.97 | 2.02 | 0.20 | 0.82 |
| GC | A1a | Tetracyclo[6.2.1.1(3,6)]dodecane-2,7-dione | 1.08 | 0.90 | 0.99 | 0.72 | 0.23 | 0.94 | 1.73 | 0.21 | 0.82 |
| GC | A1a | 1-(2-piperidin-1-ylethyl)cyclohexan-1-ol | 0.11 | 0.09 | 0.95 | 1.74 | 0.27 | 0.94 | 0.13 | 0.17 | 0.82 |
| GC | A1a | 5-Phenylporphyrin | 0.79 | 0.79 | 0.97 | 0.62 | 0.19 | 0.94 | 0.50 | 0.27 | 0.84 |
| GC | A1a | 2-(2-Butoxyethyl)pyridine | 0.61 | 0.21 | 0.95 | 0.92 | 0.72 | 0.96 | 0.77 | 0.62 | 0.92 |
| GC | A1a | 1,2,3,3,4,4,5,5-Octamethylcyclopentene | 0.80 | 0.85 | 0.98 | 0.36 | < 0.05* | 0.84 | 0.12 | < 0.05* | 0.61 |
| GC | A1a | 4-(2'-Hydroxyphenyl)non-1-ene | 1.02 | 0.94 | 1.00 | 0.71 | 0.11 | 0.93 | 0.97 | 0.95 | 0.99 |
| GC | A1a | N,N-Dimethyl-2,2-bis( 9'-anthracenyl)acetamide | 0.69 | 0.39 | 0.97 | 1.03 | 0.91 | 0.98 | 0.57 | 0.23 | 0.83 |
| GC | A1a | methyl (2S,3S)-4-(4-methoxyphenyl)-3-methyl-2-[(2-methylpropan-2-yl)oxycarbonylamino]-4-oxobutanoate | 1.39 | 0.32 | 0.97 | 1.04 | 0.86 | 0.97 | 0.87 | 0.65 | 0.92 |
| GC | A1a | ethyl 2-methylidene-4-(2,4,6-trimethylphenyl)butanoate | 1.38 | 0.20 | 0.95 | 1.21 | 0.27 | 0.94 | 1.24 | 0.46 | 0.90 |
| GC | A1a | (2S,6R)-4-[3-(4-tert-butylphenyl)-2-methylpropyl]-2,6-dimethylmorpholine | 1.08 | 0.79 | 0.97 | 1.09 | 0.42 | 0.95 | 0.82 | 0.37 | 0.89 |
| GC | A1a | 2,2',4,4',5,5'-Hexachlorobiphenyl | 1.23 | 0.49 | 0.97 | 1.07 | 0.48 | 0.95 | 1.01 | 0.98 | 0.99 |
| GC | A1a | (2E)-1-O-Acetyl-3,6-dimethyl-6-formyl-2-hepten-1-ol | 0.86 | 0.81 | 0.97 | 1.01 | 0.98 | 1.00 | 0.81 | 0.69 | 0.93 |
| GC | A1a | 2-(1-bromoethenyl)-5-methoxy-1,3-dimethylbenzene | 1.07 | 0.93 | 1.00 | 1.17 | 0.64 | 0.96 | 1.39 | 0.59 | 0.92 |
| GC | A1a | (1S,3R,4R)-2-[(S)-1-Phenylethylamino]-2-azabicyclo[2.2.1]heptane-3(S)-phenylmethanol | 0.64 | 0.15 | 0.95 | 0.92 | 0.41 | 0.95 | 0.93 | 0.77 | 0.96 |
| GC | A1a | N-(n-Butyl)-3-cyclohexenylpropiolamide | 0.58 | 0.45 | 0.97 | 0.92 | 0.83 | 0.97 | 0.87 | 0.90 | 0.99 |
| GC | A1a | 4-tert-Octylphenol | 0.85 | 0.51 | 0.97 | 0.89 | 0.33 | 0.95 | 0.97 | 0.90 | 0.99 |
| GC | A1a | 2-phenylpropanenitrile | 1.34 | 0.77 | 0.97 | 2.15 | 0.12 | 0.93 | 3.62 | 0.10 | 0.77 |
| GC | A1a | (-)-(4a'R,8'S,8a'S,10a'S)-1',1',4a',8a'-Tetramethyl-3',4',4a',6',7',8',8a',9',10',10a'-decahydro-1'H-spiro[[1,3]dioxolane-2,2'-phenanthren]-8'-ol | 0.87 | 0.69 | 0.97 | 0.99 | 0.93 | 0.99 | 0.93 | 0.85 | 0.99 |
| GC | A1a | Benzoic acid, 2,4-dihydroxy-3-[[4-methoxy-2-(phenylmethoxy)-6-propylbenzoyl]oxy]-6-propyl-, methyl ester | 1.73 | 0.34 | 0.97 | 0.88 | 0.57 | 0.96 | 1.13 | 0.79 | 0.97 |
| GC | A1a | dimethyl 5-(1H-indol-3-yl)-2-methyl-3-methylsulfanyl-1,2,4-triazine-5,6-dicarboxylate | 0.65 | 0.31 | 0.97 | 0.92 | 0.61 | 0.96 | 1.02 | 0.97 | 0.99 |
| GC | A1a | 1,1-Dimethyl-1,3-dihydro-isobenzofuran-5-ol | 1.58 | 0.48 | 0.97 | 1.18 | 0.57 | 0.95 | 0.46 | 0.20 | 0.82 |
| GC | A1a | Methyl 1,3-dihydro-2H-isobenzofuran-4-carboxylate | 0.29 | 0.14 | 0.95 | 1.20 | 0.63 | 0.96 | 0.26 | 0.09 | 0.77 |
| GC | A1a | Diethyl Phthalate | 1.29 | 0.18 | 0.95 | 0.89 | 0.62 | 0.96 | 0.66 | 0.65 | 0.92 |
| GC | A1a | ethyl 2-hydroxy-2-(4-methylphenyl)-2-(5-oxocyclopenten-1-yl)acetate | 1.05 | 0.87 | 0.99 | 0.80 | 0.06 | 0.93 | 0.78 | 0.37 | 0.89 |
| GC | A1a | Methoxyethyl-3-(4-oxo-4H-1-benzopyran-3-yl)-acrylate | 6.56 | 0.15 | 0.95 | 0.80 | 0.67 | 0.96 | 0.54 | 0.56 | 0.92 |
| GC | A1a | Octahydro-5-pentylindolizine-8-methanol | 1.45 | 0.23 | 0.97 | 1.17 | 0.28 | 0.95 | 1.65 | 0.15 | 0.82 |
| GC | A1a | 1-prop-1-ynyl-1-cyclohex-2-enol | 4.46 | < 0.05* | 0.95 | 0.55 | 0.28 | 0.94 | 0.81 | 0.84 | 0.98 |
| GC | A1a | .alpha.-Chloroformyl-4-ethoxyphenylhydrazine hydrochloride | 4.03 | 0.07 | 0.95 | 0.84 | 0.61 | 0.96 | 1.35 | 0.62 | 0.92 |
| GC | A1a | 2-Ethyl-N-methyl-1-phenyl-3-butenylamine | 1.61 | 0.67 | 0.97 | 1.42 | 0.56 | 0.95 | 6.03 | 0.20 | 0.82 |
| GC | A1a | 3-Methoxy-3-oxopropane-1,2-diyl dibutyrate | 0.47 | 0.13 | 0.95 | 0.67 | 0.08 | 0.93 | 0.82 | 0.65 | 0.92 |
| GC | A1a | DEET | 0.45 | 0.38 | 0.97 | 0.62 | 0.08 | 0.93 | 0.29 | 0.11 | 0.78 |
| GC | A1a | 3-(p-Methoxyphenyl)-5,6-dimethylindenone | 0.19 | 0.32 | 0.97 | 2.26 | 0.28 | 0.95 | 0.20 | 0.31 | 0.87 |
| GC | A1a | [2-(4-ethylbenzoyl)oxyphenyl] 2,6-difluorobenzoate | 1.71 | 0.59 | 0.97 | 0.58 | 0.20 | 0.94 | 0.84 | 0.82 | 0.98 |
| GC | A1a | 4-Ethylbenzoic acid, 3-chlorophenyl ester | 2.99 | 0.28 | 0.97 | 0.76 | 0.37 | 0.95 | 4.03 | 0.15 | 0.82 |
| GC | A1a | 5,6,8-Trimethoxy-7-methyl-2,3-diphenylquinoxaline | 2.07 | 0.47 | 0.97 | 0.73 | 0.36 | 0.95 | 1.36 | 0.64 | 0.92 |
| GC | A1a | Methfuroxam | 1.03 | 0.97 | 1.00 | 0.79 | 0.46 | 0.95 | 0.65 | 0.50 | 0.90 |
| GC | A1a | 2-(3,5-dichloro-2-methoxy-phenyl)-1H-pyrrole | 3.41 | 0.66 | 0.97 | 1.27 | 0.83 | 0.97 | 0.19 | 0.38 | 0.89 |
| GC | A1a | p-Tolyl Furan-2-Carboxylate | 0.69 | 0.68 | 0.97 | 1.86 | 0.19 | 0.94 | 0.47 | 0.34 | 0.88 |
| GC | A1a | 2-Methyl-2-(2'-propenyl)-cyclohexane-1,3-dione | 0.80 | 0.62 | 0.97 | 0.61 | < 0.05* | 0.84 | 0.59 | 0.21 | 0.82 |
| GC | A1a | 4,7-Dihydroxy-5-methylcoumarin | 0.72 | 0.66 | 0.97 | 0.72 | 0.26 | 0.94 | 1.20 | 0.70 | 0.94 |
| GC | A1a | (2S)-2-amino-2-(1,3-benzodioxol-5-yl)acetic acid methyl ester | 0.93 | 0.63 | 0.97 | 0.94 | 0.34 | 0.95 | 1.05 | 0.73 | 0.94 |
| GC | A1a | [(1R,2S,4aR,6R,8aR)-2,6-dimethyl-1,2,4a,5,6,7,8,8a-octahydronaphthalen-1-yl]methanol | 0.95 | 0.90 | 0.99 | 0.96 | 0.89 | 0.98 | 1.67 | 0.29 | 0.86 |
| GC | A1a | (4-methylphenyl) 2-hydroxybenzoate | 0.98 | 0.98 | 1.00 | 1.28 | 0.58 | 0.96 | 0.94 | 0.93 | 0.99 |
| GC | A1a | (1R,2S,6R)-2-Acetoxy-9-(hex-2-en-2-yl)bicyclo[4.3.0]nona-3,7-diene | 1.29 | 0.70 | 0.97 | 1.51 | 0.18 | 0.94 | 1.64 | 0.32 | 0.87 |
| GC | A1a | Methyl phenidate | 0.46 | 0.06 | 0.95 | 0.69 | < 0.05* | 0.84 | 1.23 | 0.62 | 0.92 |
| GC | A1a | 2-[(Phenylsulfanyl)methyl]oxirane | 1.02 | 0.98 | 1.00 | 1.32 | 0.51 | 0.95 | 0.35 | 0.30 | 0.87 |
| GC | A1a | endo-5,6-Di(acetyl)-7-acetoxymethylenebicyclo[2.2.1]hept-2-ene | 2.85 | 0.20 | 0.95 | 1.21 | 0.57 | 0.95 | 2.22 | 0.27 | 0.84 |
| GC | A1a | (E)-[(4S)-4-allyl-2,2-dimethyl-1,3-dioxan-5-ylidene]-[(2S)-2-(methoxymethyl)pyrrolidino]amine | 1.06 | 0.78 | 0.97 | 0.86 | 0.46 | 0.95 | 0.96 | 0.93 | 0.99 |
| GC | A1a | 3-(piperidin-1-ylmethyl)-2,3-dihydro-1,4-benzodioxin-6-yl Methanesulfonate | 0.92 | 0.87 | 0.99 | 0.98 | 0.95 | 0.99 | 1.40 | 0.55 | 0.92 |
| GC | A1a | 2,4-di-tert-butylphenol | 0.57 | 0.69 | 0.97 | 1.06 | 0.91 | 0.98 | 1.53 | 0.56 | 0.92 |
| GC | A1a | Cyclopropyl 2-aminobenzoate | 1.44 | 0.54 | 0.97 | 2.23 | 0.12 | 0.93 | 0.71 | 0.53 | 0.91 |
| GC | A1a | Methoxy-colorenone | 0.62 | 0.45 | 0.97 | 0.70 | 0.20 | 0.94 | 2.38 | 0.26 | 0.84 |
| GC | A1a | Methyl 1-(phenylsulfonyl)-3-(methoxycarbonyl)indole-2-acetate | 0.60 | 0.36 | 0.97 | 0.62 | < 0.05* | 0.84 | 0.50 | 0.19 | 0.82 |
| GC | A1a | (E)-1-cyclohexyl-2-hepten-1-one | 0.47 | 0.06 | 0.95 | 1.12 | 0.50 | 0.95 | 0.83 | 0.58 | 0.92 |
| GC | A1a | 3-(ethylamino)phenol | 0.74 | 0.49 | 0.97 | 1.04 | 0.82 | 0.97 | 1.25 | 0.52 | 0.91 |
| GC | A1a | N-ethyl-N-[(7-fluoro-2,3-dihydro-1,4-benzodioxin-2-yl)methyl]propan-1-amine | 1.60 | 0.49 | 0.97 | 0.90 | 0.73 | 0.96 | 0.86 | 0.82 | 0.98 |
| GC | A1a | 3-methylcyclohepta[b]furan-2-one | 2.28 | 0.36 | 0.97 | 1.11 | 0.74 | 0.96 | 4.65 | 0.09 | 0.77 |
| GC | A1a | methyl (2R,3R,3aR)-2-(2-methylpropanoyl)-2,3,3a,4,5,6-hexahydropyrrolo[1,2-b][1,2]oxazole-3-carboxylate | 0.76 | 0.55 | 0.97 | 0.90 | 0.66 | 0.96 | 1.29 | 0.63 | 0.92 |
| GC | A1a | N-[(7-fluoro-2,3-dihydro-1,4-benzodioxin-2-yl)methyl]-N-methylethanamine | 0.72 | 0.66 | 0.97 | 1.18 | 0.44 | 0.95 | 0.64 | 0.22 | 0.82 |
| GC | A1a | 6,9-Dimethoxy-5,5-dimethyl-3,3a,5,9b-tetrahydro-furo[3,2-c]isochromen-2-one | 0.70 | 0.27 | 0.97 | 1.01 | 0.95 | 1.00 | 0.99 | 0.97 | 0.99 |
| GC | A1a | 3-hydroxypropyl benzoate | 1.06 | 0.96 | 1.00 | 1.43 | 0.35 | 0.95 | 0.26 | 0.10 | 0.77 |
| GC | A1a | N-(2-Chloroethyl)-3-methoxybenzamide | 0.94 | 0.89 | 0.99 | 1.54 | 0.20 | 0.94 | 1.10 | 0.90 | 0.99 |
| GC | A1a | Octyl decyl phthalate | 1.27 | 0.46 | 0.97 | 0.93 | 0.60 | 0.96 | 0.73 | 0.26 | 0.84 |
| GC | A1a | [(2S)-2-[(1R,3aR,4S,7aR)-4-hydroxy-7a-methyl-1,2,3,3a,4,5,6,7-octahydroinden-1-yl]propyl] 4-methylbenzenesulfonate | 0.25 | 0.34 | 0.97 | 1.62 | 0.45 | 0.95 | 0.96 | 0.98 | 0.99 |
| GC | A1a | 4-methoxy-1-[methoxy(phenyl)methyl]-2-nitro-benzene | 0.79 | 0.49 | 0.97 | 0.87 | 0.29 | 0.95 | 0.78 | 0.41 | 0.90 |
| GC | A1a | Ibuprofen | 0.91 | 0.77 | 0.97 | 0.91 | 0.45 | 0.95 | 1.29 | 0.30 | 0.86 |
| GC | A1a | (+-)-5-[2-(7-Methoxyindol-3-yl)ethyl]-2,3,3a.beta.,6,7,7a.beta.-hexahydrofuro[3,2-c]pyridin-4(5H)-one | 0.85 | 0.73 | 0.97 | 1.47 | 0.13 | 0.93 | 0.45 | 0.17 | 0.82 |
| GC | A1a | 2,4,4'-Trichlorobiphenyl | 0.25 | 0.10 | 0.95 | 1.41 | 0.19 | 0.94 | 1.35 | 0.65 | 0.92 |
| GC | A1a | (E)-1-phenyl-4-hexen-1-one | 0.69 | 0.70 | 0.97 | 1.63 | 0.36 | 0.95 | 0.92 | 0.92 | 0.99 |
| GC | A1a | 2,2-Dimethyl-5-(prop-2'-ynyl)-1,3-dioxane-4,6-dione | 2.89 | 0.14 | 0.95 | 0.98 | 0.95 | 1.00 | 0.42 | 0.29 | 0.86 |
| GC | A1a | 2,5-dihydroxy-4-prop-2-enylbenzaldehyde | 0.81 | 0.66 | 0.97 | 1.01 | 0.97 | 1.00 | 0.83 | 0.65 | 0.92 |
| GC | A1a | 2(S)-Amino-3-methyl-N,N-dioctylbutyramide | 0.89 | 0.80 | 0.97 | 0.82 | 0.31 | 0.95 | 0.78 | 0.51 | 0.91 |
| GC | A1a | Ethyl (3-Methyl-6-oxo-1-phenyl-4,5,6,7-tetrahydro-1H-pyrazolo[3,4-b]pyridine-4-carboxamido)benzoate | 0.98 | 0.98 | 1.00 | 1.09 | 0.77 | 0.97 | 0.72 | 0.57 | 0.92 |
| GC | A1a | 3-(Chloromethyl)-4-hydroxy-4-phenyl-2-butanone | 1.28 | 0.59 | 0.97 | 0.82 | 0.34 | 0.95 | 1.35 | 0.35 | 0.88 |
| GC | A1a | 3-(Dimethylphenylsilyl)-2-methylpropionamide | 0.41 | 0.13 | 0.95 | 0.84 | 0.31 | 0.95 | 0.33 | 0.21 | 0.82 |
| GC | A1a | 4-tert-butyl-4-(1-methoxy-2-methyl-propan-2-yl)-1-methyl-azetidine-2,3-dione | 0.81 | 0.36 | 0.97 | 0.83 | < 0.05* | 0.84 | 0.85 | 0.40 | 0.89 |
| GC | A1a | (5S,8S)-8-Isopropyl-3-methoxy-5-methyl-1,2,3,4-tetrahydronaphthalen-2-carbaldehyde | 1.98 | 0.42 | 0.97 | 2.17 | < 0.05* | 0.84 | 0.96 | 0.96 | 0.99 |
| GC | A1a | 5-p-anisyloxypentan-1-ol | 1.27 | 0.72 | 0.97 | 2.00 | < 0.05* | 0.84 | 1.74 | 0.48 | 0.90 |
| GC | A1a | (2E,4E)-3-Phenylthiopenta-2,4-dienal | 1.55 | 0.45 | 0.97 | 0.61 | < 0.05* | 0.84 | 0.96 | 0.93 | 0.99 |
| GC | A1a | (1R,2R)-1,2-diphenylethane-1,2-diol | 1.12 | 0.87 | 0.99 | 0.60 | 0.11 | 0.93 | 2.09 | 0.23 | 0.82 |
| GC | A1a | 4-Methylbenzyl vinyl ether | 1.44 | 0.37 | 0.97 | 1.02 | 0.91 | 0.98 | 0.80 | 0.52 | 0.91 |
| GC | A1a | 1-O-hexyl 2-O-propan-2-yl benzene-1,2-dicarboxylate | 1.35 | 0.19 | 0.95 | 0.99 | 0.97 | 1.00 | 1.32 | 0.36 | 0.89 |
| GC | A1a | methyl 2-methylbenzimidazole-1-carboxylate | 0.42 | < 0.05* | 0.95 | 0.85 | 0.33 | 0.95 | 1.17 | 0.63 | 0.92 |
| GC | A1a | 3-Piperidin-1-yl-butan-2-ol | 0.85 | 0.37 | 0.97 | 1.00 | 0.98 | 1.00 | 0.94 | 0.72 | 0.94 |
| GC | A1a | (-)-(R,E)-5-(tert-Butylsulfonyl)hept-3-ene | 2.23 | 0.28 | 0.97 | 0.82 | 0.56 | 0.95 | 0.84 | 0.72 | 0.94 |
| GC | A1a | (Z)-4-Methylphenylazo tert-butyl sulfide | 1.55 | 0.73 | 0.97 | 3.09 | < 0.05* | 0.84 | 5.29 | 0.08 | 0.77 |
| GC | A1a | 10-Formyl-1,2,3,4,5,6-hexahydro-1,5-imino-9-methoxy-3,8,11-trimethyl-4-oxo-3-benzazocine | 0.97 | 0.94 | 1.00 | 0.93 | 0.73 | 0.96 | 0.84 | 0.63 | 0.92 |
| GC | A1a | 1-hydroxy-3-methyl-2-pyrrolecarbonitrile | 2.26 | 0.20 | 0.95 | 2.34 | < 0.05* | 0.84 | 1.27 | 0.75 | 0.95 |
| GC | A1a | (E)-2,2,5-triphenyl-4-pentenenitrile | 1.89 | 0.52 | 0.97 | 0.76 | 0.50 | 0.95 | 0.66 | 0.57 | 0.92 |
| GC | A1a | 4-Pentylphenol | 0.60 | 0.71 | 0.97 | 1.64 | 0.37 | 0.95 | 10.80 | 0.13 | 0.82 |
| GC | A1a | 2-[(E)-hydroxyiminomethyl]phenol | 0.43 | 0.59 | 0.97 | 1.26 | 0.73 | 0.96 | 4.25 | 0.37 | 0.89 |
| GC | A1a | Bis(2-ethylhexyl) adipate | 0.90 | 0.68 | 0.97 | 0.12 | 0.14 | 0.94 | 0.23 | 0.46 | 0.90 |
| GC | A1a | 4'-[(Methanesulfonyloxy)methyl]-[2,2' ; 6',6\]-terpyridyl"" | 0.57 | 0.43 | 0.97 | 1.08 | 0.80 | 0.97 | 2.06 | 0.16 | 0.82 |
| GC | A1a | (2S,3R,4R)-4-tert-butyl-2-phenyl-3-oxolanol | 0.78 | 0.46 | 0.97 | 0.82 | 0.37 | 0.95 | 1.48 | 0.38 | 0.89 |
| GC | A1a | 2-(2-nitrophenyl)-1H-benzimidazole | 0.99 | 0.99 | 1.00 | 1.36 | 0.77 | 0.97 | 1.24 | 0.94 | 0.99 |
| GC | A1a | 2-N-[2'-(Methoxymethyl)pyrrolidin-1'-yl]amino-3-methylheptane | 1.05 | 0.69 | 0.97 | 1.05 | 0.44 | 0.95 | 1.15 | 0.32 | 0.87 |
| GC | A1a | 1H-3,1-benzoxazine-2,4-dione | 0.87 | 0.42 | 0.97 | 0.98 | 0.81 | 0.97 | 0.69 | < 0.05* | 0.61 |
| GC | A1a | 8-Methyl-8-(hydroxymethyl)-4,5,7,8-tetrahydro-6H-cyclohepta[b]furan | 0.89 | 0.52 | 0.97 | 1.00 | 1.00 | 1.00 | 1.01 | 0.94 | 0.99 |
| GC | A1a | 2-methyl-1H-pyrrole-3-carbonitrile | 1.25 | 0.35 | 0.97 | 1.04 | 0.67 | 0.96 | 0.88 | 0.52 | 0.91 |
| GC | A1a | 5,16-androstadiene-3.beta.-ol-acetate | 0.89 | 0.25 | 0.97 | 1.05 | 0.21 | 0.94 | 0.95 | 0.64 | 0.92 |
| GC | A1a | [1,2,4]Triazolo[1,5-a][1,3,5]triazin-7-amine, 1,2-dihydro-2-methyl-2-phenyl- | 0.99 | 0.93 | 1.00 | 1.07 | 0.29 | 0.95 | 1.29 | 0.09 | 0.77 |
| GC | A1a | (2R,3S)-{3-[4-[(Tetrahydropyran-2-yl)oxy]but-2(E)-enyl]oxiranyl}methanol | 1.06 | 0.81 | 0.97 | 0.97 | 0.79 | 0.97 | 1.18 | 0.48 | 0.90 |
| GC | A1a | 3-cyclohexyl-5-phenyl-1,3-oxazolidine-2,4-dione | 0.95 | 0.71 | 0.97 | 1.02 | 0.74 | 0.96 | 1.01 | 0.96 | 0.99 |
| GC | A1a | 2-Hydroxy-3-methylbicyclo[2.2.1]heptane-2-methanol | 0.35 | 0.25 | 0.97 | 1.09 | 0.73 | 0.96 | 1.20 | 0.57 | 0.92 |
| GC | A1a | 16,17-Epoxy-6,7-isopropylidenedioxy-1,15-dihydroxy-8,15-secomaoerystals A | 0.93 | 0.58 | 0.97 | 1.00 | 0.98 | 1.00 | 0.99 | 0.91 | 0.99 |
| GC | A1a | 2,5-Dichlorophenol | 0.80 | 0.36 | 0.97 | 0.84 | 0.08 | 0.93 | 1.03 | 0.91 | 0.99 |
| GC | A1a | 2-methyl-1-acenaphthylenecarbonitrile | 1.23 | 0.10 | 0.95 | 1.02 | 0.74 | 0.96 | 1.11 | 0.32 | 0.87 |
| GC | A1a | 10,11-Dimethoxy-12b-trimethylsilyl-5,6,7,8,12b,13a-hexahydrodibenzo[c,g]oxireno[2,3-e]azecin-5-one | 0.97 | 0.89 | 0.99 | 1.13 | 0.17 | 0.94 | 1.00 | 1.00 | 1.00 |
| GC | A1a | 1-(4-Chlorophenyl)-2-octanone | 0.96 | 0.43 | 0.97 | 0.97 | 0.23 | 0.94 | 0.94 | 0.21 | 0.82 |
| GC | A1a | (-)-2.beta.-Methoxy-2a.beta.,3,4,6,7,8,8a.beta.,8b.beta.-Octahydro-6,6,8b.beta.-trimethyl-2H-naphtho[1,8-bc]furan | 0.90 | 0.70 | 0.97 | 1.00 | 0.98 | 1.00 | 1.25 | 0.43 | 0.90 |
| GC | A1a | Ethyl 2,4-dimethylpyrido[2,3-a]benzofuran-3-carboxylate | 1.06 | 0.73 | 0.97 | 1.21 | < 0.05* | 0.84 | 0.89 | 0.62 | 0.92 |
| GC | A1a | [3'-Phenyl-2'-propenyl ] Allyl Ether | 1.20 | 0.60 | 0.97 | 1.02 | 0.87 | 0.97 | 0.59 | 0.09 | 0.77 |
| GC | A1a | l-Norleucine, N-methoxycarbonyl-, isohexyl ester | 1.22 | 0.46 | 0.97 | 0.93 | 0.55 | 0.95 | 0.86 | 0.55 | 0.92 |
| GC | A1a | 2,7-Dimethyl-10-n-propyl-4,5-dioxo-3,6,9-trioxa-3,4,5,6,9,10-hexahydroanthracene | 0.94 | 0.82 | 0.97 | 1.24 | 0.12 | 0.93 | 1.63 | 0.08 | 0.77 |
| GC | A1a | Benzo[b]furo[3,4-f][1,4]benzodioxin-1,3-dione, 3a,4,11a,11b-tetrahydro-, (3a.alpha.,11a.alpha.,11b.alpha.)- | 1.31 | < 0.05* | 0.95 | 0.96 | 0.41 | 0.95 | 1.32 | < 0.05* | 0.54 |
| GC | A1a | (1R*,2R*,5R*)-sec-butyl 5-hydroxy-2-methylcyclohexanecarboxylate | 0.87 | 0.28 | 0.97 | 0.92 | 0.12 | 0.93 | 1.04 | 0.67 | 0.93 |
| GC | A1a | 2,4,6-Tribromophenol | 0.83 | 0.22 | 0.95 | 0.95 | 0.39 | 0.95 | 0.96 | 0.72 | 0.94 |
| GC | A1a | 2,3-Dihydro-10-methyl-1H,7H,8H-benzo[ij]pyrano[3,2-c]]quinolizine-7,8-dione | 1.51 | < 0.05* | 0.95 | 0.83 | < 0.05* | 0.84 | 1.56 | < 0.05* | 0.54 |
| GC | A1a | 1-(4'-Fluoro-2'-hydroxyphenyl)-2,2-dimethylpropan-1-one | 1.55 | < 0.05* | 0.95 | 0.86 | 0.08 | 0.93 | 1.64 | < 0.05* | 0.51 |
| GC | A1a | 2-Butyl-3-phenyl-5-isopropyl-4-pentylfuran | 0.87 | 0.33 | 0.97 | 1.03 | 0.55 | 0.95 | 0.98 | 0.90 | 0.99 |
| GC | A1a | 2,4,6,8-Tetramethylazulene | 1.02 | 0.82 | 0.97 | 1.12 | < 0.05* | 0.84 | 1.06 | 0.56 | 0.92 |
| GC | A1a | 2,2'-Diihydroxy-4'-methyl-5-methoxybiphenyl | 1.03 | 0.80 | 0.97 | 1.02 | 0.71 | 0.96 | 0.96 | 0.61 | 0.92 |
| GC | A1a | 9,10-seco-8b,10-dihydroxy-12-oxocalanolide A | 0.76 | 0.24 | 0.97 | 0.97 | 0.79 | 0.97 | 1.10 | 0.68 | 0.93 |
| GC | A1a | 1,2-Dihydrocyclopentafluorene | 1.09 | 0.41 | 0.97 | 1.02 | 0.63 | 0.96 | 1.09 | 0.52 | 0.91 |
| GC | A1a | 7-Acetoxy-8-ethenylquinoline | 1.22 | 0.56 | 0.97 | 0.84 | 0.22 | 0.94 | 1.55 | 0.06 | 0.77 |
| GC | A1a | 3-Amino-4,5-bis(p-biphenyl)-1H-pyrazolo[3,4-c]pyridazine | 0.87 | 0.05 | 0.95 | 0.97 | 0.34 | 0.95 | 1.05 | 0.32 | 0.87 |
| GC | A1a | 6-(1',1\-Biphenyl-4'-yl)-1,4,8-trimethylazulene"" | 0.70 | 0.18 | 0.95 | 1.06 | 0.61 | 0.96 | 0.88 | 0.53 | 0.91 |
| GC | A1a | 5-(Isobutyl)-1,4-pyrazine-2-carbonitrile | 0.99 | 0.98 | 1.00 | 1.06 | 0.74 | 0.96 | 1.28 | 0.53 | 0.91 |
| GC | A1a | N-Benzylbenzenamine | 1.03 | 0.77 | 0.97 | 0.99 | 0.90 | 0.98 | 1.01 | 0.95 | 0.99 |
| GC | A1a | (4'R,5'R)-Spiro[1,5-benzodithiepane-3,2'-[4.5]diethoxycarbonyl[1,3]dioxolane] | 0.95 | 0.79 | 0.97 | 1.04 | 0.63 | 0.96 | 1.16 | 0.38 | 0.89 |
| GC | A1a | Benz[c]acridine-7(2H)-one, 1,3,4,12-tetrahydro- | 1.24 | 0.22 | 0.95 | 0.99 | 0.85 | 0.97 | 0.89 | 0.47 | 0.90 |
| GC | A1a | 6-Ethoxycarbonyl-1-phenyl-1H-pyrazolo[4,3-c]pyridine-4-thione | 0.93 | 0.75 | 0.97 | 0.92 | 0.46 | 0.95 | 0.83 | 0.47 | 0.90 |
| GC | A1a | 1-(tert-Butyldimethylsilyloxy)-3-vinylpent-4-en-2-ol | 0.30 | < 0.05* | 0.95 | 1.07 | 0.75 | 0.96 | 0.85 | 0.71 | 0.94 |
| GC | A1a | N-{4-[N'-(4-hydroxybenzylidene)hydrazino]benzyl}-2,2,2-trifluoroacetamide | 0.94 | 0.71 | 0.97 | 1.06 | 0.48 | 0.95 | 1.01 | 0.94 | 0.99 |
| GC | A1a | Benzoic acid, 3-hydroxy-2-(tetrahydro-2H-pyran-2-yl)-, methyl ester | 0.80 | 0.29 | 0.97 | 1.01 | 0.93 | 0.99 | 0.63 | 0.08 | 0.77 |
| GC | A1a | 6-Isobutanoyl-7-methoxycoumarin | 0.86 | 0.16 | 0.95 | 0.98 | 0.74 | 0.96 | 0.81 | 0.06 | 0.76 |
| GC | A1a | 1-methyl-4-phenyl-2-[(E)-2-phenylethenyl]pyrrole | 0.90 | 0.80 | 0.97 | 1.20 | 0.41 | 0.95 | 1.37 | 0.42 | 0.90 |
| GC | A1a | 1-hydroxy-3-methoxy-2-(1-methoxyethyl)-9,10-anthraquinone | 0.93 | 0.70 | 0.97 | 0.94 | 0.55 | 0.95 | 1.08 | 0.65 | 0.92 |
| GC | A1a | 2-Methyl-4-phenylpyrido[2,3-d]pyrimidin-5(8H)-one | 1.00 | 0.99 | 1.00 | 0.93 | 0.54 | 0.95 | 0.96 | 0.88 | 0.99 |
| GC | A1a | 2-(Pyrrolidin-1-yl)-1-(5,6,7,8-tetrahydronaphthalen-2- yl)pentan-1-one | 0.90 | 0.59 | 0.97 | 0.88 | 0.10 | 0.93 | 1.14 | 0.27 | 0.84 |
| GC | A1a | 2,3-Dihydro-1-(methylene)pyrrolo[1,2-a]indole-9-carbaldehyde N,N-dimethylhydrazone | 1.14 | 0.56 | 0.97 | 1.02 | 0.83 | 0.97 | 0.96 | 0.85 | 0.99 |
| GC | A1a | 1-methyl-2-oxidanylidene-pyridine-3-carbaldehyde | 0.93 | 0.83 | 0.97 | 1.09 | 0.62 | 0.96 | 0.96 | 0.93 | 0.99 |
| GC | A1a | 8-methoxy-2H-1-benzopyran-3-carboxylic acid methyl ester | 1.16 | 0.17 | 0.95 | 0.97 | 0.53 | 0.95 | 1.30 | < 0.05* | 0.54 |
| GC | A1a | 1,3-Benzenedicarbonitrile | 1.17 | < 0.05* | 0.95 | 1.03 | 0.44 | 0.95 | 1.60 | < 0.001** | 0.06 |
| GC | A1a | 7-methylpyrrolo[1,2-a]pyrazin-1-amine | 1.22 | 0.54 | 0.97 | 1.06 | 0.69 | 0.96 | 1.05 | 0.82 | 0.98 |
| GC | A1a | 2-[(5-bromo-2-methoxybenzoyl)amino]acetic acid | 0.81 | 0.10 | 0.95 | 0.99 | 0.80 | 0.97 | 0.98 | 0.86 | 0.99 |
| GC | A1a | Isopropyl 3-methylquinoxaline-2-carboxylate | 0.98 | 0.86 | 0.99 | 0.96 | 0.42 | 0.95 | 1.08 | 0.41 | 0.90 |
| GC | A1a | p,p'-DDE | 1.39 | < 0.05* | 0.95 | 1.02 | 0.82 | 0.97 | 1.08 | 0.62 | 0.92 |
| GC | A1a | 4-hydroxy-3,4-dimethylnaphthalen-1-one | 1.30 | 0.65 | 0.97 | 1.22 | 0.38 | 0.95 | 2.52 | 0.05 | 0.76 |
| GC | A1a | Butanedioic acid, [1-(acetyloxy)-1,2,2-trimethylpropyl]-, diethyl ester | 1.24 | 0.07 | 0.95 | 0.93 | 0.22 | 0.94 | 1.04 | 0.79 | 0.97 |
| GC | A1a | Diethyl L-tartrate | 0.97 | 0.86 | 0.99 | 1.01 | 0.93 | 0.99 | 1.00 | 0.98 | 0.99 |
| GC | A1a | 5-(t-Butyl)-3,3-dimethyl-4-(benzyl)thiophen-2(3H)-one | 1.12 | 0.51 | 0.97 | 1.18 | 0.31 | 0.95 | 1.50 | 0.38 | 0.89 |
| GC | A1a | 2-Mercaptobenzothiazole | 0.97 | 0.81 | 0.97 | 1.01 | 0.84 | 0.97 | 1.16 | 0.14 | 0.82 |
| GC | A1a | 3-(5,7-Dioxo-5,7-dihydro-pyrrolo[3,4-b]pyrazin-6-yl)-propionic acid ethyl ester | 0.87 | 0.50 | 0.97 | 1.01 | 0.87 | 0.97 | 1.11 | 0.54 | 0.91 |
| GC | A1a | 2-Oxahexacyclo[7,5,1.0.0(3,13).0(5,12).0(10,14)]pentadecan-3-ol | 1.07 | 0.68 | 0.97 | 1.13 | 0.09 | 0.93 | 0.98 | 0.89 | 0.99 |
| GC | A1a | 10H-Phenoxazine | 1.01 | 0.96 | 1.00 | 0.91 | 0.23 | 0.94 | 1.27 | 0.14 | 0.82 |
| GC | A1a | 2-(furan-3-yl)-2-(2-iodoethyl)-1,3-dioxolane | 0.85 | 0.45 | 0.97 | 0.80 | < 0.05* | 0.84 | 0.99 | 0.97 | 0.99 |
| GC | A1a | 7-isopropyl-3,4-dihydro-2H-cyclohepta[b]pyrazine | 1.35 | 0.12 | 0.95 | 0.94 | 0.45 | 0.95 | 2.45 | < 0.001** | 0.06 |
| GC | A1a | Acetic acid, [[1-(cyanohydroxymethyl)-2-naphthalenyl]oxy]-, ethyl ester | 0.79 | 0.05 | 0.95 | 1.15 | < 0.05* | 0.84 | 1.02 | 0.89 | 0.99 |
| GC | A1a | Tris(2-ethylhexyl) trimellitate | 1.04 | 0.90 | 0.99 | 0.97 | 0.85 | 0.97 | 0.87 | 0.58 | 0.92 |
| GC | A1a | 1-Piperidineacetaldehyde, .alpha.-methyl-.alpha.-[[(tetrahydro-2H-pyran-2-yl)oxy]methyl]- | 1.15 | 0.37 | 0.97 | 1.12 | 0.12 | 0.93 | 1.06 | 0.70 | 0.94 |
| GC | A1a | 3,4-Diethoxy-2,5-dimethoxytetrahydrofuran | 1.04 | 0.80 | 0.97 | 0.95 | 0.44 | 0.95 | 0.88 | 0.43 | 0.90 |
| GC | A1a | 1-[2-(E)-5-(3-(1-Hydroxycyclohex-1-yl)-2-propynylidene)-1-cyclopenten-1-yl]ethynyl]cyclohexan-1-ol | 1.20 | 0.59 | 0.97 | 0.88 | 0.35 | 0.95 | 0.73 | 0.44 | 0.90 |
| GC | A1a | (2S,3S,4S)-2-[2'-((S)-4''-Benzyl-2''-oxo-oxazolidin-3''-yl)-2-oxo-ethyl]-N-cyclohexyl-5-cyano-3,4-dimethyl-piperidine | 0.88 | 0.07 | 0.95 | 0.99 | 0.72 | 0.96 | 1.02 | 0.62 | 0.92 |
| GC | A1a | Triphenyl phosphate | 1.01 | 0.97 | 1.00 | 0.94 | 0.52 | 0.95 | 0.98 | 0.91 | 0.99 |
| GC | A1a | (4R,5R)-2,2-dimethyl-4-(2-phenylethynyl)-5-vinyl-1,3-dioxolane | 0.91 | 0.58 | 0.97 | 1.09 | 0.37 | 0.95 | 0.76 | 0.26 | 0.84 |
| GC | A1a | N-{2-[(4-Acetylphenyl)ethynyl]phenyl}-2,2,2-trifluoroacetamide | 0.71 | 0.13 | 0.95 | 1.13 | 0.28 | 0.95 | 1.07 | 0.79 | 0.97 |
| GC | A1a | (3R,3aR,6S,6aR)-3-(3,4-dimethoxyphenyl)-6-(3,4,5-trimethoxyphenyl)-1,3,3a,4,6,6a-hexahydrofuro[3,4-c]furan | 1.00 | 0.99 | 1.00 | 1.06 | 0.41 | 0.95 | 0.98 | 0.83 | 0.98 |
| GC | A1a | Methyl endo-9-acetyl-7-methyltricyclo[5.2.2.0(1,5)]undec-5-ene-6-carboxylate | 1.21 | 0.11 | 0.95 | 0.98 | 0.65 | 0.96 | 0.95 | 0.58 | 0.92 |
| GC | A1a | 2-ethoxy-3,1-benzoxazin-4-one | 0.78 | 0.57 | 0.97 | 1.18 | 0.39 | 0.95 | 0.80 | 0.56 | 0.92 |
| GC | A1a | 9-methyl-9-phenyl-9,10-dihydrophenanthrene | 0.89 | 0.49 | 0.97 | 0.96 | 0.62 | 0.96 | 1.39 | 0.09 | 0.77 |
| GC | A1a | (Z)-Phenylazo tert-butyl sulfide | 1.27 | 0.65 | 0.97 | 1.35 | 0.12 | 0.93 | 0.81 | 0.54 | 0.92 |
| GC | A1a | 4-Chloro-2-(6-chlorohex-1-yn-1-yl)pyridine | 1.15 | 0.17 | 0.95 | 0.92 | 0.08 | 0.93 | 1.42 | < 0.05* | 0.18 |
| GC | A1a | Thiazolo[5,4-f]quinoline-2-carbonitrile | 1.09 | 0.56 | 0.97 | 1.14 | 0.10 | 0.93 | 0.93 | 0.64 | 0.92 |
| GC | A1a | 1,4,5,6-Tetrahydro-1-(4',6'-heptadienoyl)-pyridine | 1.09 | 0.77 | 0.97 | 0.86 | 0.26 | 0.94 | 1.19 | 0.49 | 0.90 |
| GC | A1a | (3S,5R,8aS)-3-butyl-5-propyl-1,2,3,5,6,7,8,8a-octahydroindolizine | 0.69 | 0.13 | 0.95 | 1.02 | 0.84 | 0.97 | 0.77 | 0.24 | 0.84 |
| GC | A1a | 4,6,7-trimethylinden-1-one | 0.51 | 0.06 | 0.95 | 1.41 | < 0.05* | 0.84 | 1.22 | 0.45 | 0.90 |
| GC | A1a | methyl 1-butyl-2-oxocycloheptane-1-carboxylate | 0.98 | 0.97 | 1.00 | 1.04 | 0.81 | 0.97 | 1.17 | 0.66 | 0.93 |
| GC | A1a | 4-Acetoxy-10,12-dimethyl-14-oxatricyclo[7.4.0.0(6,7)]tetradecane | 1.05 | 0.77 | 0.97 | 0.90 | 0.13 | 0.93 | 1.19 | 0.34 | 0.88 |
| GC | A1a | 2,3,4,4a,5,6,6a,11b-Octahydro-6a-hydroperoxy-8-methoxy-7,9,10-trimethyl-1H-benzo[a]carbazole | 0.96 | 0.85 | 0.98 | 0.90 | 0.26 | 0.94 | 0.74 | 0.20 | 0.82 |
| GC | A1a | 7-Cyano-2-oxatricyclo[5.3.1.0(1,11)]undec-1(10)-en-9-one | 0.30 | 0.20 | 0.95 | 0.81 | 0.50 | 0.95 | 0.65 | 0.57 | 0.92 |
| GC | A1a | 1,2,4-Trichlorobenzene | 1.25 | 0.67 | 0.97 | 0.96 | 0.86 | 0.97 | 1.26 | 0.62 | 0.92 |
| GC | A1a | Apocynin | 0.99 | 0.97 | 1.00 | 0.80 | 0.16 | 0.94 | 0.96 | 0.90 | 0.99 |
| GC | A1a | 2-Acetoxy-3-chlorophenol | 1.05 | 0.65 | 0.97 | 0.97 | 0.69 | 0.96 | 0.84 | 0.10 | 0.77 |
| GC | A1a | 4,5-Dihydrofuro[1,2-a]naphthalene | 1.22 | 0.56 | 0.97 | 0.95 | 0.75 | 0.96 | 1.35 | 0.33 | 0.87 |
| GC | A1a | 2-tert-Butylfuro[4,5-d][1,3]dioxan-6-one | 0.80 | 0.42 | 0.97 | 1.46 | < 0.05* | 0.84 | 1.23 | 0.59 | 0.92 |
| GC | A1a | 10-(p-Tolyl)benzo[h]quinoline | 0.92 | 0.31 | 0.97 | 1.00 | 0.99 | 1.00 | 1.09 | 0.22 | 0.82 |
| GC | A1a | Isoxaben | 0.75 | 0.19 | 0.95 | 1.06 | 0.52 | 0.95 | 0.67 | 0.08 | 0.77 |
| GC | A1a | 1-(5-Methyl-2-oxo-2,5-dihydrofuran-3-yl)-9-(tert-butyldimethylsiloxy)nonan-8-ol | 0.82 | 0.25 | 0.97 | 0.96 | 0.56 | 0.95 | 1.02 | 0.90 | 0.99 |
| GC | A1a | 3-butylcyclohexan-1-one | 1.21 | 0.39 | 0.97 | 1.02 | 0.86 | 0.97 | 0.54 | < 0.05* | 0.72 |
| GC | A1a | 6,7-Dimethoxy-1-[(p-tolyl)sulfinyl]methyl-1-(trifluoromethyl)-1,2,3,4-tetrahydroisoquinoline | 0.71 | 0.39 | 0.97 | 1.14 | 0.47 | 0.95 | 0.95 | 0.89 | 0.99 |
| GC | A1a | 2-[Chloro(diphenyl)methyl]pyridine | 1.16 | 0.62 | 0.97 | 1.00 | 1.00 | 1.00 | 1.14 | 0.56 | 0.92 |
| GC | A1a | 1-[(1S,2R)-2-propoxycyclopentyl]hexane-1,2-dione | 1.17 | 0.05 | 0.95 | 1.04 | 0.26 | 0.94 | 1.18 | < 0.05* | 0.72 |
| GC | A1a | Dodecyltriethoxysilane | 1.28 | 0.32 | 0.97 | 0.89 | 0.36 | 0.95 | 1.15 | 0.61 | 0.92 |
| GC | A1a | N-(1-methyl-2,5-dioxo-3-[1]benzopyrano[4,3-b]pyridinyl)acetamide | 1.00 | 1.00 | 1.00 | 0.86 | 0.19 | 0.94 | 0.98 | 0.92 | 0.99 |
| GC | A1a | 6-Phenyl-2-pyridinamine | 1.34 | 0.06 | 0.95 | 1.06 | 0.36 | 0.95 | 1.13 | 0.34 | 0.88 |
| GC | A1a | 4-(3-ethoxyphenyl)-N,N-diethyl-aniline | 0.95 | 0.66 | 0.97 | 1.10 | 0.08 | 0.93 | 0.95 | 0.58 | 0.92 |
| GC | A1a | (4R,4aR,8aR)-4,8a-dimethyl-1,2,3,4,7,8-hexahydronaphthalen-4a-ol | 0.45 | 0.13 | 0.95 | 0.95 | 0.80 | 0.97 | 0.74 | 0.57 | 0.92 |
| GC | A1a | Padimate O | 1.11 | 0.38 | 0.97 | 1.01 | 0.86 | 0.97 | 0.99 | 0.94 | 0.99 |
| GC | A1a | (2E)-2-(phenylmethylene)-3-benzofuranone | 0.92 | 0.60 | 0.97 | 0.96 | 0.64 | 0.96 | 0.78 | 0.19 | 0.82 |
| GC | A1a | methyl 2-(4-methoxyphenyl)propanoate | 1.15 | 0.46 | 0.97 | 0.96 | 0.51 | 0.95 | 0.94 | 0.72 | 0.94 |
| GC | A1a | (8-acetoxy-1-cyano-7-ethoxy-6-methyl-5-isoquinolyl) acetate | 0.98 | 0.90 | 0.99 | 0.85 | < 0.05* | 0.84 | 1.12 | 0.37 | 0.89 |
| GC | A1a | 15-Nonacosanone | 0.89 | 0.40 | 0.97 | 0.92 | 0.17 | 0.94 | 1.15 | 0.25 | 0.84 |
| GC | A1a | 1,4:5,8-Dimethanonaphthalene, 6-butyl-1,2,3,4,4a,5,8,8a-octahydro-, (1.alpha.,4.alpha.,4a.beta.,5.beta.,8.beta.,8a.beta.)- | 1.09 | 0.73 | 0.97 | 1.17 | 0.23 | 0.94 | 1.01 | 0.96 | 0.99 |
| GC | A1a | (Z)-2-Ethyl-5-bromo-2-dicyanomethyladamantane | 0.89 | 0.19 | 0.95 | 1.06 | 0.19 | 0.94 | 1.13 | 0.16 | 0.82 |
| GC | A1a | 3-(5,5,6-Trimethylbicyclo[2.2.1]hept-2-ylidene)-2-propanone oxime | 0.85 | 0.47 | 0.97 | 1.05 | 0.80 | 0.97 | 1.43 | 0.33 | 0.87 |
| GC | A1a | (3,6-dimethyl-4-oxidanylidene-2H-chromen-3-yl) nitrate | 1.93 | 0.11 | 0.95 | 1.29 | 0.14 | 0.94 | 1.87 | 0.14 | 0.82 |
| GC | A1a | 6,7-dimethoxy-1-(methoxymethyl)-3,4-dihydroisoquinoline | 1.06 | 0.75 | 0.97 | 1.00 | 0.99 | 1.00 | 1.19 | 0.24 | 0.84 |
| GC | A1a | Methyl (E)-3-methoxy-4-oxo-4-phenylbut-2-enoate | 0.99 | 0.95 | 1.00 | 0.98 | 0.80 | 0.97 | 0.76 | 0.12 | 0.82 |
| GC | A1a | 3,3,7-trimethyl-5-nitro-1H-indol-2-one | 1.07 | 0.39 | 0.97 | 0.96 | 0.25 | 0.94 | 1.18 | 0.05 | 0.76 |
| GC | A1a | 1-hydroxy-1-methyl-tetralin-6-carbaldehyde | 1.54 | 0.45 | 0.97 | 2.05 | < 0.05* | 0.84 | 0.99 | 0.98 | 0.99 |
| GC | A1a | Acetosyringon | 0.92 | 0.36 | 0.97 | 0.99 | 0.74 | 0.96 | 0.96 | 0.69 | 0.93 |
| GC | A1a | Methyl (S)-1-(4,4-diphenylbut-3-en-1-yl)pyrrolidine-2-carboxylate | 1.13 | 0.76 | 0.97 | 0.91 | 0.61 | 0.96 | 1.01 | 0.97 | 0.99 |
| GC | A1a | Octabenzone | 1.16 | 0.52 | 0.97 | 0.89 | 0.31 | 0.95 | 1.35 | 0.22 | 0.82 |
| GC | A1a | Diphenyl(1,3,5-trimethyl-1H-pyrazol-4-yl)amine | 1.18 | 0.42 | 0.97 | 1.06 | 0.43 | 0.95 | 1.13 | 0.46 | 0.90 |
| GC | A1a | Octicizer | 0.83 | 0.22 | 0.95 | 1.04 | 0.56 | 0.95 | 1.05 | 0.74 | 0.95 |
| GC | A1a | 6-Methoxy-2-phenyl-2-(phenylthio)-3-benzofuranone | 0.75 | 0.29 | 0.97 | 0.99 | 0.90 | 0.98 | 1.11 | 0.66 | 0.92 |
| GC | A1a | 2-Acetonyl-3-acetyl-5-methoxy-1,4-naphthoquinone | 1.20 | 0.18 | 0.95 | 1.02 | 0.66 | 0.96 | 0.96 | 0.68 | 0.93 |
| GC | A1a | Bumetrizole | 0.75 | 0.34 | 0.97 | 1.31 | 0.06 | 0.93 | 1.81 | 0.09 | 0.77 |
| GC | A1a | (2SR,3aRS,6aRS,10aRS)-1,7,7,10a-Tetramethyl-2-phenyl-2,3,3a,6,6a,7,8,9,10,10a-decahydro-4-oxabenz[e]azulen-5-one | 0.87 | 0.54 | 0.97 | 1.03 | 0.73 | 0.96 | 1.08 | 0.71 | 0.94 |
| GC | A1a | Tri(2-ethylhexyl) phosphate | 0.86 | 0.56 | 0.97 | 1.04 | 0.69 | 0.96 | 1.34 | 0.17 | 0.82 |
| GC | A1a | 2-[(E)-pentadec-1-enyl]benzene-1,3-dicarbaldehyde | 1.00 | 0.99 | 1.00 | 1.03 | 0.61 | 0.96 | 1.14 | 0.31 | 0.87 |
| GC | A1a | 1-(6-Methoxypyridin-2-yl)propan-2-one | 0.85 | 0.61 | 0.97 | 1.25 | 0.11 | 0.93 | 1.08 | 0.80 | 0.97 |
| GC | A1a | (R)-4-Isobutyloxazolidin-2-one | 1.34 | 0.48 | 0.97 | 1.23 | 0.20 | 0.94 | 0.95 | 0.81 | 0.97 |
| GC | A1a | methyl 2-(2,4,6-trimethylphenyl)acetate | 0.90 | 0.70 | 0.97 | 0.98 | 0.87 | 0.97 | 0.91 | 0.72 | 0.94 |
| GC | A1a | 2-[(4-Chlorobenzoyl)(2-ethoxycarbonylbenzyl)methylene]-1H-imidazole | 0.92 | 0.52 | 0.97 | 1.05 | 0.41 | 0.95 | 1.22 | 0.11 | 0.78 |
| GC | A1a | 3-(2-Chloromethylphenyl)-2,2-dimethylpropionaldehyde | 0.89 | 0.79 | 0.97 | 1.13 | 0.51 | 0.95 | 0.37 | < 0.05* | 0.61 |
| GC | A1a | (+)-(2S,3R)-4-Benzyloxy-3-(tert-butyldimethylsiloxy)-2-methylbutanol | 0.99 | 0.94 | 1.00 | 1.13 | 0.17 | 0.94 | 1.23 | 0.27 | 0.84 |
| GC | A1a | cis-3,5-[di(t-Butyl)silanedioxy]cyclopent-1-ene | 1.26 | 0.13 | 0.95 | 0.96 | 0.53 | 0.95 | 1.21 | 0.22 | 0.82 |
| GC | A1a | 4-hydroperoxy-2,3,4-trimethylnaphthalen-1-one | 0.90 | 0.40 | 0.97 | 0.90 | 0.13 | 0.93 | 1.10 | 0.46 | 0.90 |
| GC | A1a | Ethanethioic acid, S-bicyclo[2.2.1]hept-2-en-7-yl ester, anti- | 0.99 | 0.92 | 0.99 | 0.95 | 0.52 | 0.95 | 1.07 | 0.62 | 0.92 |
| GC | A1a | 1-(benzo[d][1,3]dioxol-5-yl)-2-(4-methylpiperidin-1-yl)pentan-1-one | 1.10 | 0.69 | 0.97 | 1.04 | 0.68 | 0.96 | 0.96 | 0.88 | 0.99 |
| GC | A1a | Propylparaben | 1.28 | 0.27 | 0.97 | 1.00 | 0.98 | 1.00 | 1.11 | 0.59 | 0.92 |
| GC | A1a | 2,4-Dihydroxy-5-methoxy-6-methyl-tetrahydropyran | 1.33 | 0.21 | 0.95 | 1.08 | 0.46 | 0.95 | 1.22 | 0.27 | 0.84 |
| GC | A1a | Octinoxate | 0.77 | 0.09 | 0.95 | 1.06 | 0.31 | 0.95 | 0.94 | 0.67 | 0.93 |
| GC | A1a | (Z)-13-Ethylene-9,10-dimethoxy-2,3-(methylenedioxy)-8,14-cycloberbine | 1.00 | 0.96 | 1.00 | 1.01 | 0.86 | 0.97 | 0.92 | 0.46 | 0.90 |
| GC | A1a | 1,7-Dimethylxanthine | 1.05 | 0.64 | 0.97 | 0.95 | 0.24 | 0.94 | 1.12 | 0.20 | 0.82 |
| GC | A1a | 5-carboxylated pyrrole derivative of 4,4-dimethyl-2-(N-methylpyrrol-2-yl)oxazoline | 1.22 | 0.60 | 0.97 | 1.08 | 0.60 | 0.96 | 0.84 | 0.55 | 0.92 |
| GC | A1a | Pestalolide | 1.45 | 0.53 | 0.97 | 0.51 | 0.15 | 0.94 | 0.49 | 0.46 | 0.90 |
| GC | A1a | Methyl (1R*,5S*)-1-Hydroxy-1-methylspiro[4.5]dec-7-ene-8-carboxylate | 0.99 | 0.94 | 1.00 | 1.00 | 0.99 | 1.00 | 1.25 | 0.06 | 0.77 |
| GC | A1a | (3-methylphenyl) 3-nitrobenzoate | 0.98 | 0.85 | 0.98 | 1.05 | 0.42 | 0.95 | 1.19 | 0.22 | 0.82 |
| GC | A1a | 2-(3-furanyl)-1H-benzimidazole | 1.35 | 0.19 | 0.95 | 0.97 | 0.77 | 0.97 | 1.52 | 0.07 | 0.77 |
| GC | A1a | (1R,3S)-3-phenylcyclohexanol | 0.79 | 0.43 | 0.97 | 0.93 | 0.54 | 0.95 | 1.08 | 0.77 | 0.97 |
| GC | A1a | 9-[1-(3-methylphenyl)ethylidene]fluorene | 1.05 | 0.30 | 0.97 | 1.01 | 0.73 | 0.96 | 1.04 | 0.47 | 0.90 |
| GC | A1a | 2,3,3',4,4',5'-Hexachlorobiphenyl | 0.87 | 0.31 | 0.97 | 1.09 | 0.16 | 0.94 | 1.12 | 0.30 | 0.87 |
| GC | A1a | (2-iodanyl-2-triethylsilyl-ethyl) 2,2,2-tris(fluoranyl)ethanoate | 0.84 | 0.57 | 0.97 | 1.02 | 0.86 | 0.97 | 0.73 | 0.26 | 0.84 |
| GC | A1a | 4-ethyl-4-methyl-5-methylene-1,3-dioxolan-2-one | 1.05 | 0.90 | 0.99 | 0.87 | 0.38 | 0.95 | 1.11 | 0.75 | 0.95 |
| GC | A1a | (4aS,8aS)-5,5,8a-trimethyl-1-methylene-3,4,4a,6,7,8-hexahydronaphthalen-2-one | 0.78 | 0.18 | 0.95 | 0.87 | 0.08 | 0.93 | 0.85 | 0.33 | 0.87 |
| GC | A1a | Dibutyl phthalate | 1.04 | 0.81 | 0.97 | 1.21 | < 0.05* | 0.84 | 1.51 | 0.09 | 0.77 |
| GC | A1a | 7-Methoxy-2,5-dimethyl-chromone | 0.85 | 0.19 | 0.95 | 0.99 | 0.82 | 0.97 | 1.01 | 0.88 | 0.99 |
| GC | A1a | (R)-(S)-.alpha.-Methoxyphenylacetic acid (NPA) | 0.90 | 0.63 | 0.97 | 0.87 | 0.12 | 0.93 | 0.86 | 0.47 | 0.90 |
| GC | A1a | 4-Ethyl-3-methyl-1,2,4-thiadiazol-5-one | 0.99 | 0.97 | 1.00 | 1.07 | 0.63 | 0.96 | 0.99 | 0.95 | 0.99 |
| GC | A1a | 3-Aminochrysene | 0.75 | 0.56 | 0.97 | 1.18 | 0.34 | 0.95 | 3.03 | < 0.05* | 0.61 |
| GC | A1a | 2-(tert-Butyl-dimethyl-silanyloxy)-3-methyl-cyclopent-2-enone | 1.26 | 0.07 | 0.95 | 1.06 | 0.38 | 0.95 | 0.96 | 0.78 | 0.97 |
| GC | A1a | 10-(3-Acetoxyprop-2-yl)-3,7-dimethylbicyclo[4.4.0]dec-2-ene | 0.92 | 0.66 | 0.97 | 0.86 | < 0.05* | 0.84 | 1.42 | 0.06 | 0.77 |
| GC | A1a | (Z)-2-Phenyl-1-(triethylsilyl)-1,4-pentadiene | 1.03 | 0.95 | 1.00 | 1.18 | 0.42 | 0.95 | 0.80 | 0.59 | 0.92 |
| GC | A1a | (2,4-dimethoxy-3-methyl-phenyl) acetate | 0.98 | 0.63 | 0.97 | 0.98 | 0.35 | 0.95 | 0.99 | 0.84 | 0.98 |
| GC | A1a | Niloticin | 0.97 | 0.83 | 0.97 | 1.08 | 0.21 | 0.94 | 1.13 | 0.40 | 0.89 |
| GC | A1a | Methyl 4-[2-(2-Chloro-3-cyanopyrid-4-yl)ethyl]benzoate | 0.98 | 0.83 | 0.97 | 0.99 | 0.80 | 0.97 | 1.11 | 0.38 | 0.89 |
| GC | A1a | 1,4-Dideoxy-1,4-imino-D-arabinitol | 1.45 | 0.21 | 0.95 | 0.96 | 0.48 | 0.95 | 1.03 | 0.81 | 0.97 |
| GC | A1a | Triisobutyl phosphate | 1.97 | 0.57 | 0.97 | 0.84 | 0.54 | 0.95 | 0.94 | 0.88 | 0.99 |
| GC | A1a | (2'R*,3'R*,.alpha.S)-2,3-Epoxyoctyl .alpha.-methoxyphenylacetate | 1.22 | 0.48 | 0.97 | 0.73 | < 0.05* | 0.84 | 1.02 | 0.93 | 0.99 |
| GC | A1a | 5-Prop-2-ynyl-cyclohex-1-enylamine | 1.12 | 0.64 | 0.97 | 0.97 | 0.84 | 0.97 | 1.06 | 0.83 | 0.98 |
| GC | A1a | 2-{[Dimethyl(3-methylbut-2-en-1-yl)silyl]methyl}allyl Acetate | 0.93 | 0.77 | 0.97 | 0.93 | 0.45 | 0.95 | 0.79 | 0.24 | 0.84 |
| GC | A1a | 7a-Carboxymethyl-1-methylene-1,4,5,6,7,7a-hexahydro-2H-indene | 0.86 | 0.48 | 0.97 | 0.91 | 0.49 | 0.95 | 0.87 | 0.63 | 0.92 |
| GC | A1a | (5S,6R)-5,6-dimethyl-3,4,5,6-tetrahydro-2H-cyclopenta[b]pyran-7-one | 1.31 | 0.12 | 0.95 | 0.93 | 0.34 | 0.95 | 1.84 | < 0.05* | 0.27 |
| GC | A1a | 9-phenylcarbazole | 1.21 | 0.12 | 0.95 | 1.01 | 0.84 | 0.97 | 1.17 | 0.23 | 0.82 |
| GC | A1a | (S)-2-(4-Methoxyphenyl)-3-methylbutan-1-ol | 0.99 | 0.98 | 1.00 | 1.10 | 0.43 | 0.95 | 1.12 | 0.70 | 0.94 |
| GC | A1a | 1,4-Naphthalenedione, 5-[(acetyloxy)methyl]-4a,5-dihydro-2-methoxy-4a-methyl-, trans- | 0.92 | 0.78 | 0.97 | 1.15 | 0.22 | 0.94 | 1.11 | 0.62 | 0.92 |
| GC | A1a | 7-Phenylethyl-4,6-dioxa-5-carbonyl-spiro[2,5]-7-octene | 0.62 | < 0.05* | 0.95 | 0.94 | 0.38 | 0.95 | 1.32 | 0.08 | 0.77 |
| GC | A1a | 1-Cyano-1-(t-butyl)-2-aminoethene | 1.80 | 0.17 | 0.95 | 1.10 | 0.56 | 0.95 | 1.39 | 0.29 | 0.86 |
| GC | A1a | 2-Carboxy-4-methylbicyclo[2.2.2]oct-2-en-1-ol | 1.00 | 1.00 | 1.00 | 0.57 | < 0.05* | 0.84 | 0.82 | 0.55 | 0.92 |
| GC | A1a | 3-isopropyl-2-(9H-xanthen-9-yl)-1H-indole | 1.12 | 0.35 | 0.97 | 1.06 | 0.30 | 0.95 | 0.85 | 0.19 | 0.82 |
| GC | A1a | 5,5-diethyl-1,3-diazinane-2,4,6-trione | 0.87 | 0.64 | 0.97 | 0.88 | 0.31 | 0.95 | 1.25 | 0.46 | 0.90 |
| GC | A1a | 6-Deoxy-1,2-O-isopropylidene-5-O-thiocarbonyl-.alpha.,D-glucofuranose | 1.06 | 0.47 | 0.97 | 1.00 | 0.92 | 0.99 | 1.10 | 0.20 | 0.82 |
| GC | A1a | 4,5-dimethyl-3-(2-methylpropyl)-2,3-dihydroisoindol-1-one | 1.41 | 0.18 | 0.95 | 1.08 | 0.52 | 0.95 | 1.44 | 0.21 | 0.82 |
| GC | A1a | (E)-Methyl undec-2-en-4-ynoate | 1.02 | 0.78 | 0.97 | 0.98 | 0.60 | 0.96 | 1.03 | 0.70 | 0.94 |
| GC | A1a | Tributyl citrate | 0.92 | 0.44 | 0.97 | 1.01 | 0.93 | 0.99 | 1.23 | 0.09 | 0.77 |
| GC | A1a | Ethyl 3-phenyl-3-(2\,4\""-dinitrophenylhydrazono)-2-(1'-methylpiperidin-2'-ylidene)propionate"" | 0.99 | 0.86 | 0.99 | 1.00 | 0.90 | 0.98 | 0.89 | < 0.05* | 0.61 |
| GC | A1a | 3-[tert-butyl(diphenyl)silyl]prop-2-yn-1-ol | 0.94 | 0.41 | 0.97 | 0.94 | 0.09 | 0.93 | 0.95 | 0.49 | 0.90 |
| GC | A1a | (3Z,9Z)-3,9-Dodecadiene-2,5,8,11-tetraone | 1.10 | 0.51 | 0.97 | 0.94 | 0.26 | 0.94 | 0.97 | 0.81 | 0.97 |
| GC | A1a | 5-Acetoxy-2,7-dimethylchromone | 1.21 | 0.75 | 0.97 | 0.91 | 0.69 | 0.96 | 0.92 | 0.86 | 0.99 |
| GC | A1a | 4-octoxybenzaldehyde | 0.42 | < 0.05* | 0.95 | 1.13 | 0.47 | 0.95 | 1.01 | 0.99 | 0.99 |
| GC | A1a | 3-(cyclopenten-1-yl)prop-2-ynyl methanesulfonate | 1.16 | 0.55 | 0.97 | 0.99 | 0.91 | 0.98 | 0.96 | 0.89 | 0.99 |
| GC | A1a | 3-hydroxy-3-(2-methoxycyclohexyl)-2-methylisoindol-1-one | 1.42 | < 0.05* | 0.74 | 1.02 | 0.63 | 0.96 | 0.88 | 0.29 | 0.86 |
| GC | A1a | Ethyl 3-nitro-4,7-dihydro-4,7-ethano-2H-isoindole-1-carboxylate | 1.04 | 0.73 | 0.97 | 1.06 | 0.31 | 0.95 | 1.21 | 0.05 | 0.76 |
| GC | A1a | 2,4-Dimethoxy-6-methylphenol | 0.95 | 0.58 | 0.97 | 0.99 | 0.88 | 0.98 | 1.03 | 0.73 | 0.94 |
| GC | A1a | (1,1,1-trichloro-2-methoxypropan-2-yl)oxycyclohexane | 0.86 | 0.49 | 0.97 | 0.95 | 0.70 | 0.96 | 1.22 | 0.43 | 0.90 |
| GC | A1a | Piperine | 0.94 | 0.45 | 0.97 | 1.00 | 0.91 | 0.98 | 0.76 | < 0.05* | 0.51 |
| GC | A1a | 2-Oxo-2H-chromen-7-yl Acetate | 1.54 | 0.28 | 0.97 | 1.26 | 0.17 | 0.94 | 1.25 | 0.51 | 0.91 |
| GC | A1a | 2-[(3R)-3,7-dimethyloct-6-enylidene]malononitrile | 1.10 | 0.20 | 0.95 | 0.94 | 0.11 | 0.93 | 1.42 | < 0.05* | 0.17 |
| GC | A1a | 5-tert-butyl-2-(4-tert-butylphenyl)-1,3-dinitro-benzene | 1.32 | 0.26 | 0.97 | 0.97 | 0.80 | 0.97 | 1.80 | < 0.05* | 0.60 |
| GC | A1a | Furo[3',4':6,7]naphtho[2,3-d]-1,3-dioxole, 5,5a,6,8,8a,9-hexahydro-, (5.alpha.,5a.beta.,8a.beta.)- | 1.05 | 0.84 | 0.98 | 0.95 | 0.53 | 0.95 | 1.10 | 0.61 | 0.92 |
| GC | A1a | Thiophen-3-yl 2-aminobenzoate | 1.50 | 0.13 | 0.95 | 0.99 | 0.94 | 0.99 | 0.83 | 0.36 | 0.89 |
| GC | A1a | 3,4-Dihydro-5,8-dimethoxy-1,3-dimethyl-1H-2-benzopyran | 1.18 | 0.63 | 0.97 | 1.03 | 0.85 | 0.97 | 1.06 | 0.86 | 0.99 |
| GC | A1a | 2-Amino-3-cyano-6-methyl-4H-chromene | 0.98 | 0.81 | 0.97 | 0.98 | 0.69 | 0.96 | 1.17 | 0.15 | 0.82 |
| GC | A1a | (1S,3S)-2-iodanyl-2,3-dihydro-1H-indene-1,3-diol | 0.80 | 0.28 | 0.97 | 0.96 | 0.69 | 0.96 | 1.27 | 0.23 | 0.83 |
| GC | A1a | 1-(Phenylethynyl)-1-benzylcyclopropane | 1.15 | 0.47 | 0.97 | 1.02 | 0.85 | 0.97 | 1.19 | 0.54 | 0.92 |
| GC | A1a | Bromoxynil | 0.87 | 0.19 | 0.95 | 1.00 | 0.99 | 1.00 | 0.90 | 0.39 | 0.89 |
| GC | A1a | (6S)-6-undecyl-2-oxanone | 0.98 | 0.91 | 0.99 | 0.90 | 0.27 | 0.94 | 0.88 | 0.49 | 0.90 |
| GC | A1a | 1,1-Diformylcyclohex-3-ene | 1.02 | 0.96 | 1.00 | 1.25 | 0.44 | 0.95 | 0.80 | 0.63 | 0.92 |
| LC | A1b | Potentially chlorinated | 2.00 | 0.57 | 1.00 | 1.00 | 1.00 | 1.00 | 0.33 | 0.34 | 0.98 |
| GC | A1b | 2H-Furo[2,3-h]-1-benzopyran-2-one | 29588452.24 | 1.00 | 1.00 | 0.98 | 0.93 | 0.99 | 0.60 | 0.23 | 0.97 |
| GC | A1b | Aceclofenac | 1.10 | 0.83 | 1.00 | 1.25 | 0.27 | 0.72 | 2.00 | 0.21 | 0.97 |
| GC | A1b | 6-phenyl-2,3,4,5-tetrahydropyridine | NA | NA | NA | 1.10 | 0.51 | 0.76 | 1.00 | 1.00 | 1.00 |
| GC | A1b | 7-Methyl-8-hydroxy-9-oxo-1-azabicyclo[4.3.0]non-7-ene | 0.67 | 0.37 | 0.99 | 0.76 | 0.20 | 0.69 | 1.56 | 0.30 | 0.98 |
| GC | A1b | 2-(6-bromo-1,3-benzodioxol-5-yl)-1-(2-bromophenyl)ethanone | 1.64 | 0.14 | 0.99 | 1.34 | 0.06 | 0.60 | 1.18 | 0.62 | 1.00 |
| GC | A1b | 3-(Methylthio)propyl isothiocyanate | 1.20 | 0.60 | 1.00 | 1.10 | 0.54 | 0.77 | 1.05 | 0.88 | 1.00 |
| GC | A1b | 4,5-Dichloro-10-(4-nitrobenzyl)-10H-anthracen-9-one | 0.64 | 0.35 | 0.99 | 1.12 | 0.69 | 0.89 | 0.42 | 0.10 | 0.92 |
| GC | A1b | 2,3,6-trimethoxydibenz[b,f]oxepine | 0.67 | 0.37 | 0.99 | 1.25 | 0.23 | 0.69 | 1.12 | 0.74 | 1.00 |
| GC | A1b | Hydrastine | 1.00 | 1.00 | 1.00 | 0.55 | 0.11 | 0.67 | 0.88 | 0.80 | 1.00 |
| GC | A1b | 1-methyl-12-phenyl-12-azatricyclo[6.3.1.02,7]dodeca-2,4,6,10-tetraen-9-one | 0.56 | 0.17 | 0.99 | 0.88 | 0.43 | 0.76 | 1.60 | 0.15 | 0.92 |
| GC | A1b | ethyl 2-[6-(dimethoxymethyl)-1,3-benzodioxol-5-yl]-2-oxoacetate | 1.25 | 0.64 | 1.00 | 1.23 | 0.23 | 0.69 | 1.00 | 1.00 | 1.00 |
| GC | A1b | 6-(2-pyridinyl)-2-pyridinecarboxaldehyde | 0.86 | 0.64 | 1.00 | 1.11 | 0.49 | 0.76 | 1.00 | 1.00 | 1.00 |
| GC | A1b | (1E,2E)-1-{2-[3-(Trifluoromethyl)phenyl]hydrazono}non-2-en-4-ol | 0.59 | 0.18 | 0.99 | 0.87 | 0.44 | 0.76 | 0.94 | 0.86 | 1.00 |
| GC | A1b | Methyl 4'-(1,1-dimethylprop-2-ynyloxy)-2'-methoxycinnamate | 0.83 | 0.67 | 1.00 | 0.80 | 0.23 | 0.69 | 1.31 | 0.47 | 1.00 |
| GC | A1b | 6-Bromo-3-(3,4-dimethoxyphenethyl)quinazoline-2,4(1H,3H)-dione | 1.00 | 1.00 | 1.00 | 1.24 | 0.42 | 0.76 | 0.64 | 0.30 | 0.98 |
| GC | A1b | 2-(2-methoxyethyl)-3,4-dihydronaphthalen-1(2H)-one | 1.00 | 1.00 | 1.00 | 1.04 | 0.84 | 0.93 | 2.37 | < 0.05* | 0.92 |
| GC | A1b | 2-(3-Nitrophenyl)-2,3-dihydro-1H-quinazolin-4-one | 0.43 | 0.22 | 0.99 | 1.38 | 0.37 | 0.76 | 1.75 | 0.37 | 0.98 |
| GC | A1b | 5-(But-3'-en-1'-yl)-6-methylizidine | 1.33 | 0.59 | 1.00 | 0.93 | 0.74 | 0.89 | 1.12 | 0.81 | 1.00 |
| GC | A1b | 2,6-Bis(4-t-butylphenyl)phenol | 0.33 | 0.18 | 0.99 | 3.14 | < 0.05* | 0.25 | 0.50 | 0.42 | 0.98 |
| GC | A1b | 4-[2-(1,3-Dithianyl)]-4-(2-ethoxycarbonylpyrrol-4-yl)butyric acid | 1.12 | 0.81 | 1.00 | 0.86 | 0.50 | 0.76 | 0.45 | 0.14 | 0.92 |
| GC | A1b | anti-14-bromo-5,8-dimethoxy-16-methyl-2,11-dithia[3]paracyclo[3](2,4)thiophenophane | 1.25 | 0.74 | 1.00 | 1.08 | 0.79 | 0.91 | 0.63 | 0.41 | 0.98 |
| GC | A1b | spiro[2,2,Dicyano-4-[(3-vinyl)phenyl]cyclobutane-1,1'-bis[1,2,5]thiadiazolodicyanocyclohexanemethane | 1.00 | 1.00 | 1.00 | 0.73 | 0.08 | 0.60 | 0.90 | 0.75 | 1.00 |
| GC | A1b | 3-Iodophenol | 7.00 | 0.07 | 0.99 | 1.48 | 0.17 | 0.69 | 1.25 | 0.74 | 1.00 |
| GC | A1b | 2-(1-adamantyl)-3,3-bis(trifluoromethyl)spiro[1,4,2-dithiazolidine-5,2'-adamantane] | 1.13 | 0.72 | 1.00 | 1.05 | 0.72 | 0.89 | 0.88 | 0.72 | 1.00 |
| GC | A1b | Diflubenzuron | 1.45 | 0.34 | 0.99 | 1.07 | 0.67 | 0.89 | 0.54 | 0.08 | 0.92 |
| GC | A1b | (S,R)-N-(1-Methyl-3-butynyl)-N-(1-phenylethyl)amine | 2.00 | 0.33 | 0.99 | 0.63 | 0.07 | 0.60 | 1.00 | 1.00 | 1.00 |
| GC | A1b | 3-Adamantyl-5-(t-butyl)-3-hydroxybenzaldehyde | 0.94 | 0.87 | 1.00 | 0.85 | 0.29 | 0.72 | 0.85 | 0.57 | 1.00 |
| GC | A1b | N-(1,3-Diphenyl-2-propynyl)-phenyl carbamate | 1.00 | 1.00 | 1.00 | 1.00 | 1.00 | 1.00 | 1.20 | 0.76 | 1.00 |
| GC | A1b | spiro[2,2,Dicyano-4-[(4-vinyl)phenyl]cyclobutane-1,1'-bis[1,2,5]thiadiazolodicyanocyclohexanemethane | 1.33 | 0.51 | 1.00 | 0.84 | 0.36 | 0.76 | 1.00 | 1.00 | 1.00 |
| LC | A2a | Pipecolate | 0.93 | 0.50 | 0.80 | 0.92 | 0.08 | 0.68 | 0.84 | 0.14 | 0.34 |
| LC | A2a | Glycochenodeoxycholic acid | 1.49 | < 0.05* | 0.15 | 1.05 | 0.51 | 0.68 | 0.82 | 0.33 | 0.52 |
| LC | A2a | Pipecolate | 0.96 | 0.84 | 0.90 | 1.06 | 0.51 | 0.68 | 1.37 | 0.14 | 0.34 |
| LC | A2a | 5-oxo-D-Proline | 1.40 | 0.20 | 0.53 | 0.92 | 0.60 | 0.68 | 0.74 | 0.62 | 0.68 |
| LC | A2a | Cortisone | 0.43 | < 0.05* | 0.20 | 0.96 | 0.84 | 0.84 | 1.46 | 0.47 | 0.63 |
| LC | A2a | Pipecolate | 1.38 | 0.31 | 0.61 | 1.16 | 0.26 | 0.68 | 0.86 | 0.68 | 0.68 |
| LC | A2a | Cortisol | 0.95 | 0.85 | 0.90 | 0.91 | 0.53 | 0.68 | 1.69 | 0.14 | 0.34 |
| LC | A2a | Cortisol | 0.96 | 0.90 | 0.90 | 0.90 | 0.47 | 0.68 | 1.63 | 0.17 | 0.34 |
| GC | A2b | 4-Thiazolidinecarboxylic acid | 1.33 | 0.45 | 0.45 | 0.98 | 0.89 | 0.89 | 1.86 | 0.19 | 0.19 |
| LC | A3a | Tyrosine | 0.98 | 0.96 | 0.98 | 1.15 | 0.38 | 0.85 | 0.60 | 0.31 | 0.72 |
| LC | A3a | LysoPE (22:6) | 0.78 | 0.58 | 0.93 | 1.01 | 0.97 | 1.00 | 0.65 | 0.30 | 0.72 |
| LC | A3a | L-Tryptophan | 0.89 | 0.80 | 0.98 | 1.05 | 0.82 | 1.00 | 0.50 | 0.42 | 0.81 |
| LC | A3a | L-Proline | 1.44 | 0.34 | 0.92 | 1.03 | 0.88 | 1.00 | 0.78 | 0.57 | 0.89 |
| LC | A3a | Alanine/Sarcosine | 1.63 | 0.34 | 0.92 | 1.25 | 0.39 | 0.85 | 0.90 | 0.86 | 0.99 |
| LC | A3a | Creatine | 1.04 | 0.87 | 0.98 | 1.15 | 0.19 | 0.83 | 1.49 | 0.16 | 0.66 |
| LC | A3a | L-Carnitine | 0.84 | 0.76 | 0.98 | 0.69 | 0.21 | 0.83 | 6.25 | < 0.05* | 0.36 |
| LC | A3a | ACar 5:0 | 0.99 | 0.97 | 0.98 | 0.93 | 0.61 | 0.95 | 1.25 | 0.52 | 0.85 |
| LC | A3a | L-Histidine | 0.71 | 0.33 | 0.92 | 1.03 | 0.91 | 1.00 | 0.32 | 0.07 | 0.47 |
| LC | A3a | 4-Aminobutanoic acid/2-Amino-2-methylpropanoic acid/3-Aminobutanoic acid | 0.60 | 0.31 | 0.92 | 0.98 | 0.91 | 1.00 | 0.84 | 0.78 | 0.98 |
| LC | A3a | Betaine | 0.29 | < 0.05* | 0.92 | 0.95 | 0.85 | 1.00 | 0.79 | 0.68 | 0.97 |
| LC | A3a | LysoPC(22:5) | 0.98 | 0.96 | 0.98 | 0.87 | 0.45 | 0.87 | 2.64 | < 0.05* | 0.43 |
| LC | A3a | LPC 0:0/22:6 | 0.66 | 0.30 | 0.92 | 0.91 | 0.55 | 0.92 | 1.20 | 0.61 | 0.93 |
| LC | A3a | Trimethylamine N-oxide | 1.17 | 0.37 | 0.92 | 1.01 | 0.95 | 1.00 | 1.08 | 0.71 | 0.97 |
| LC | A3a | L-Valine | 1.06 | 0.93 | 0.98 | 0.80 | 0.37 | 0.85 | 0.10 | < 0.05* | 0.36 |
| LC | A3a | Hypoxanthine | 1.05 | 0.66 | 0.93 | 0.92 | 0.43 | 0.87 | 1.88 | < 0.05* | 0.36 |
| LC | A3a | L-Glutamine | 1.04 | 0.94 | 0.98 | 1.60 | 0.09 | 0.77 | 1.03 | 0.96 | 0.99 |
| LC | A3a | LysoPE (18:2) | 1.71 | 0.14 | 0.92 | 1.31 | 0.11 | 0.83 | 1.19 | 0.69 | 0.97 |
| LC | A3a | LYSOPC (18:2) | 1.11 | 0.81 | 0.98 | 0.92 | 0.65 | 0.95 | 1.70 | 0.30 | 0.72 |
| LC | A3a | LYSOPC (16:3) | 1.27 | 0.37 | 0.92 | 0.94 | 0.69 | 0.98 | 1.44 | 0.31 | 0.72 |
| LC | A3a | LYSOPE (18:1) | 1.76 | 0.06 | 0.92 | 1.31 | 0.07 | 0.67 | 1.24 | 0.56 | 0.87 |
| LC | A3a | LysoPE (20:4) | 1.08 | 0.87 | 0.98 | 1.03 | 0.87 | 1.00 | 1.95 | 0.23 | 0.69 |
| LC | A3a | PC(16:0_20:4) | 0.74 | 0.56 | 0.93 | 0.76 | 0.24 | 0.83 | 2.94 | 0.08 | 0.52 |
| LC | A3a | 7a-hydroxy-3-oxo-4-cholestenoic acid (7-Hoca) | 2.22 | 0.13 | 0.92 | 0.91 | 0.63 | 0.95 | 1.65 | 0.33 | 0.72 |
| LC | A3a | SM d34:2 | 0.51 | 0.27 | 0.92 | 0.82 | 0.31 | 0.83 | 0.97 | 0.96 | 0.99 |
| LC | A3a | PC (32:2) | 1.24 | 0.46 | 0.93 | 0.98 | 0.87 | 1.00 | 1.13 | 0.71 | 0.97 |
| LC | A3a | ACar 10:1 | 1.13 | 0.66 | 0.93 | 0.70 | < 0.05* | 0.17 | 1.27 | 0.32 | 0.72 |
| LC | A3a | LYSOPC (20:4) | 1.02 | 0.97 | 0.98 | 0.83 | 0.33 | 0.83 | 3.70 | < 0.05* | 0.36 |
| LC | A3a | Deoxycarnitine-4-butyric acid betaine | 1.37 | 0.51 | 0.93 | 0.69 | 0.15 | 0.83 | 1.03 | 0.95 | 0.99 |
| LC | A3a | LYSOPC (16:3) | 1.14 | 0.53 | 0.93 | 1.06 | 0.70 | 0.98 | 0.92 | 0.77 | 0.98 |
| LC | A3a | PC (36:4) | 1.05 | 0.43 | 0.93 | 0.97 | 0.31 | 0.83 | 0.98 | 0.81 | 0.99 |
| LC | A3a | LysoPC(20:2) | 1.56 | 0.22 | 0.92 | 0.90 | 0.56 | 0.92 | 2.52 | 0.07 | 0.47 |
| LC | A3a | PC (36:2) | 1.59 | 0.16 | 0.92 | 0.98 | 0.92 | 1.00 | 0.97 | 0.95 | 0.99 |
| LC | A3a | N-alpha-acetyl-L-lysine | 1.62 | 0.32 | 0.92 | 1.19 | 0.44 | 0.87 | 1.18 | 0.80 | 0.99 |
| LC | A3a | LYSOPC (16:1) | 1.50 | 0.24 | 0.92 | 1.19 | 0.31 | 0.83 | 1.79 | 0.19 | 0.69 |
| LC | A3a | ACar 11:1 | 1.83 | 0.05 | 0.92 | 0.69 | < 0.05* | 0.32 | 0.90 | 0.73 | 0.98 |
| LC | A3a | Serotonin | 1.05 | 0.63 | 0.93 | 1.01 | 0.81 | 1.00 | 0.82 | 0.17 | 0.68 |
| LC | A3a | Cholesterol | 0.27 | 0.17 | 0.92 | 0.64 | 0.11 | 0.83 | 1.83 | 0.46 | 0.83 |
| LC | A3a | LysoPC(P-16:0) | 1.13 | 0.78 | 0.98 | 0.82 | 0.30 | 0.83 | 1.38 | 0.56 | 0.87 |
| LC | A3a | Docosahexaenoic acid | 0.57 | < 0.05* | 0.92 | 0.86 | 0.17 | 0.83 | 0.79 | 0.31 | 0.72 |
| LC | A3a | Glycolate | 1.20 | 0.57 | 0.93 | 0.93 | 0.63 | 0.95 | 2.41 | < 0.05* | 0.36 |
| LC | A3a | Lauroylcarnitine | 1.11 | 0.70 | 0.96 | 0.74 | < 0.05* | 0.37 | 1.71 | 0.06 | 0.47 |
| LC | A3a | LYSOPC (18:2) | 0.90 | 0.58 | 0.93 | 0.98 | 0.83 | 1.00 | 1.25 | 0.34 | 0.72 |
| LC | A3a | Trigonelline | 0.97 | 0.81 | 0.98 | 0.96 | 0.56 | 0.92 | 1.62 | < 0.05* | 0.36 |
| LC | A3a | L-gamma-Glutamyl-L-leucine | 1.02 | 0.93 | 0.98 | 1.09 | 0.49 | 0.91 | 0.66 | 0.24 | 0.69 |
| LC | A3a | PC (36:3) | 1.03 | 0.92 | 0.98 | 0.86 | 0.23 | 0.83 | 0.97 | 0.92 | 0.99 |
| LC | A3a | LysoPC (18:3) | 1.29 | 0.27 | 0.92 | 1.07 | 0.63 | 0.95 | 1.41 | 0.26 | 0.69 |
| LC | A3a | ACar 4:0 | 0.96 | 0.82 | 0.98 | 0.89 | 0.13 | 0.83 | 1.22 | 0.18 | 0.69 |
| LC | A3a | PC (36:4) | 1.03 | 0.83 | 0.98 | 1.01 | 0.80 | 1.00 | 1.02 | 0.82 | 0.99 |
| LC | A3a | Plasmalogen PC(38:4) or PC(O-38:5) | 1.19 | 0.56 | 0.93 | 0.84 | 0.30 | 0.83 | 1.04 | 0.92 | 0.99 |
| LC | A3a | Taurine | 1.47 | 0.11 | 0.92 | 0.72 | 0.07 | 0.67 | 1.30 | 0.49 | 0.85 |
| LC | A3a | PC (38:5) | 1.08 | 0.85 | 0.98 | 0.97 | 0.90 | 1.00 | 2.07 | 0.15 | 0.64 |
| LC | A3a | LysoPE (18:2) | 1.11 | 0.63 | 0.93 | 1.04 | 0.71 | 0.98 | 1.10 | 0.69 | 0.97 |
| LC | A3a | 4-Guanidinobutanoic acid | 1.04 | 0.83 | 0.98 | 1.38 | < 0.05* | 0.17 | 1.01 | 0.98 | 0.99 |
| LC | A3a | ACar 10:1 | 0.96 | 0.90 | 0.98 | 0.75 | < 0.05* | 0.32 | 1.16 | 0.51 | 0.85 |
| LC | A3a | LYSOPC (16:1) | 0.89 | 0.57 | 0.93 | 1.28 | < 0.05* | 0.37 | 1.45 | 0.12 | 0.56 |
| LC | A3a | ACar 9:0 | 1.44 | 0.08 | 0.92 | 0.92 | 0.41 | 0.85 | 1.15 | 0.58 | 0.89 |
| LC | A3a | Plasmalogen PC(36:4) or PC(O-36:5) | 0.81 | 0.57 | 0.93 | 0.97 | 0.86 | 1.00 | 1.58 | 0.25 | 0.69 |
| LC | A3a | Uric acid | 0.94 | 0.87 | 0.98 | 0.94 | 0.73 | 0.98 | 1.07 | 0.91 | 0.99 |
| LC | A3a | LYSOPC (18:0) | 1.26 | 0.52 | 0.93 | 1.01 | 0.94 | 1.00 | 2.42 | 0.10 | 0.54 |
| LC | A3a | LYSOPC (16:1) | 1.14 | 0.33 | 0.92 | 0.90 | 0.14 | 0.83 | 0.79 | 0.10 | 0.54 |
| LC | A3a | Kynurenine | 0.93 | 0.42 | 0.93 | 1.10 | 0.62 | 0.95 | 1.49 | 0.44 | 0.82 |
| LC | A3a | 5-Valerolactone | 0.98 | 0.91 | 0.98 | 1.08 | 0.45 | 0.87 | 1.11 | 0.65 | 0.97 |
| LC | A3a | ACar 4:0 | 1.02 | 0.89 | 0.98 | 1.00 | 0.97 | 1.00 | 1.34 | 0.19 | 0.69 |
| LC | A3a | Acetylcarnitine (C2) | 1.14 | 0.75 | 0.98 | 0.87 | 0.40 | 0.85 | 2.50 | 0.06 | 0.47 |
| LC | A3a | LysoPC (16:0) | 1.81 | 0.62 | 0.93 | 0.44 | 0.16 | 0.83 | 0.23 | 0.36 | 0.75 |
| LC | A3a | ACar 8:0 | 1.07 | 0.77 | 0.98 | 0.79 | < 0.05* | 0.40 | 1.44 | 0.12 | 0.56 |
| LC | A3a | Linoleic acid | 0.87 | 0.42 | 0.93 | 0.90 | 0.23 | 0.83 | 0.83 | 0.39 | 0.78 |
| LC | A3a | LYSOPC (16:1) | 0.93 | 0.42 | 0.93 | 1.10 | < 0.05* | 0.46 | 1.26 | < 0.05* | 0.36 |
| LC | A3a | 5,6-Dihydrouracil | 2.38 | 0.12 | 0.92 | 1.66 | 0.15 | 0.83 | 4.21 | 0.06 | 0.47 |
| LC | A3a | Decanoyl-L-Carnitine (C10:0) | 1.04 | 0.86 | 0.98 | 0.78 | < 0.05* | 0.37 | 1.47 | 0.10 | 0.54 |
| LC | A3a | L-Serine | 1.05 | 0.90 | 0.98 | 1.00 | 0.99 | 1.00 | 0.53 | 0.23 | 0.69 |
| LC | A3a | 1-methylnicotinamide | 0.99 | 0.97 | 0.98 | 1.03 | 0.76 | 0.99 | 1.00 | 0.99 | 0.99 |
| LC | A3a | LYSOPC (14:0) | 1.25 | 0.41 | 0.93 | 1.00 | 0.99 | 1.00 | 1.38 | 0.40 | 0.78 |
| LC | A3a | ACar 8:1 | 0.97 | 0.86 | 0.98 | 0.77 | < 0.05* | 0.32 | 0.93 | 0.77 | 0.98 |
| LC | A3a | LYSOPC (20:4) | 1.53 | 0.51 | 0.93 | 0.90 | 0.71 | 0.98 | 5.90 | < 0.05* | 0.46 |
| LC | A3a | LYSOPC (18:0) | 1.44 | 0.43 | 0.93 | 0.80 | 0.28 | 0.83 | 1.50 | 0.51 | 0.85 |
| LC | A3a | L-Methionine | 0.84 | 0.62 | 0.93 | 0.99 | 0.94 | 1.00 | 0.44 | 0.14 | 0.64 |
| LC | A3a | Propionyl-L-carnitine (C3:0) | 1.27 | 0.45 | 0.93 | 1.01 | 0.96 | 1.00 | 1.72 | 0.19 | 0.69 |
| LC | A3a | PC (34:3) | 1.10 | 0.55 | 0.93 | 0.94 | 0.40 | 0.85 | 1.00 | 0.98 | 0.99 |
| LC | A3a | L-Tyrosine | 0.82 | 0.61 | 0.93 | 1.06 | 0.74 | 0.98 | 0.52 | 0.22 | 0.69 |
| LC | A3a | PC 38:6 | 0.58 | 0.18 | 0.92 | 0.98 | 0.89 | 1.00 | 0.88 | 0.73 | 0.98 |
| LC | A3a | ACar 18:1 | 1.39 | 0.25 | 0.92 | 0.82 | 0.27 | 0.83 | 1.57 | 0.32 | 0.72 |
| LC | A3a | LYSOPC (17:0) | 1.41 | 0.35 | 0.92 | 0.91 | 0.60 | 0.95 | 0.98 | 0.97 | 0.99 |
| LC | A3a | Theophylline | 0.83 | 0.20 | 0.92 | 0.90 | 0.17 | 0.83 | 0.84 | 0.29 | 0.72 |
| LC | A3a | Glycocholic acid | 1.27 | 0.24 | 0.92 | 1.07 | 0.49 | 0.91 | 0.63 | 0.07 | 0.47 |
| LC | A3a | PC (34:1) | 1.29 | 0.31 | 0.92 | 1.11 | 0.41 | 0.85 | 1.25 | 0.43 | 0.81 |
| LC | A3a | PC (30:0) | 1.15 | 0.57 | 0.93 | 1.02 | 0.87 | 1.00 | 1.27 | 0.35 | 0.74 |
| LC | A3a | PC (38:3) | 1.16 | 0.23 | 0.92 | 0.95 | 0.42 | 0.86 | 1.01 | 0.93 | 0.99 |
| LC | A3a | 3,5-Tetradecadiencarnitine (C14:2) | 0.92 | 0.77 | 0.98 | 0.74 | < 0.05* | 0.32 | 1.44 | 0.12 | 0.56 |
| LC | A3a | Indolelactic acid | 1.77 | 0.13 | 0.92 | 1.23 | 0.30 | 0.83 | 1.07 | 0.90 | 0.99 |
| LC | A3a | Dehydroepiandrosterone | 0.88 | 0.73 | 0.97 | 1.13 | 0.52 | 0.92 | 0.72 | 0.50 | 0.85 |
| LC | A3a | Paraxanthine | 0.95 | 0.72 | 0.96 | 0.91 | 0.16 | 0.83 | 1.05 | 0.75 | 0.98 |
| LC | A3a | LYSOPC (20:3) | 1.18 | 0.66 | 0.93 | 0.85 | 0.32 | 0.83 | 0.96 | 0.94 | 0.99 |
| LC | A3a | Uracil | 1.07 | 0.84 | 0.98 | 0.91 | 0.54 | 0.92 | 1.64 | 0.21 | 0.69 |
| LC | A3a | Threonic acid | 1.05 | 0.62 | 0.93 | 0.97 | 0.79 | 1.00 | 1.94 | < 0.05* | 0.36 |
| LC | A3a | LYSOPC (20:3) | 1.58 | 0.26 | 0.92 | 0.93 | 0.73 | 0.98 | 4.61 | < 0.05* | 0.36 |
| LC | A3a | Retinol | 0.56 | 0.29 | 0.92 | 1.30 | 0.17 | 0.83 | 2.24 | 0.10 | 0.54 |
| LC | A3a | LYSOPC (20:3) | 0.76 | 0.10 | 0.92 | 1.00 | 1.00 | 1.00 | 1.12 | 0.51 | 0.85 |
| LC | A3a | PC (34:2) | 0.66 | 0.51 | 0.93 | 1.31 | 0.32 | 0.83 | 0.81 | 0.72 | 0.98 |
| LC | A3a | LYSOPC (18:4) | 1.41 | 0.27 | 0.92 | 1.23 | 0.20 | 0.83 | 1.95 | 0.09 | 0.54 |
| LC | A3a | PC (36:4) | 1.15 | 0.64 | 0.93 | 1.01 | 0.95 | 1.00 | 1.42 | 0.33 | 0.72 |
| LC | A3a | Nepsilon, nepsilon, nepsilon-trimethyllysine | 1.48 | 0.21 | 0.92 | 1.11 | 0.51 | 0.91 | 1.03 | 0.95 | 0.99 |
| LC | A3a | PC(32:1) | 1.18 | 0.47 | 0.93 | 1.28 | < 0.05* | 0.40 | 1.26 | 0.34 | 0.72 |
| LC | A3a | L-Threonine/L-Homoserine | 1.32 | 0.45 | 0.93 | 0.78 | 0.19 | 0.83 | 0.39 | < 0.05* | 0.38 |
| LC | A3a | PC (40:5) | 0.93 | 0.70 | 0.96 | 1.03 | 0.76 | 0.99 | 1.08 | 0.68 | 0.97 |
| LC | A3a | Theobromine | 1.00 | 0.98 | 0.99 | 0.93 | 0.30 | 0.83 | 0.80 | 0.15 | 0.64 |
| LC | A3a | Phe-Phe | 0.95 | 0.24 | 0.92 | 1.01 | 0.97 | 1.00 | 1.12 | 0.87 | 0.99 |
| LC | A3a | Creatinine | 1.43 | 0.49 | 0.93 | 0.72 | 0.20 | 0.83 | 0.57 | 0.37 | 0.75 |
| LC | A3a | Tyrosine | 0.97 | 0.93 | 0.98 | 1.25 | 0.23 | 0.83 | 0.54 | 0.23 | 0.69 |
| LC | A3a | Undecanoylcarnitine | 1.43 | 0.26 | 0.92 | 0.84 | 0.27 | 0.83 | 1.16 | 0.68 | 0.97 |
| LC | A3a | Plasmalogen PC(36:3) or PC(O-36:4) | 1.31 | 0.37 | 0.92 | 0.87 | 0.32 | 0.83 | 1.55 | 0.22 | 0.69 |
| LC | A3a | Diacylglycerol (C34:2) | 1.08 | 0.61 | 0.93 | 1.07 | 0.36 | 0.85 | 1.30 | 0.13 | 0.61 |
| LC | A3a | Dimethylsulfone | 1.00 | 0.99 | 0.99 | 0.90 | 0.50 | 0.91 | 1.47 | 0.24 | 0.69 |
| LC | A3a | Indole-3-propionic acid (IPA) | 0.67 | < 0.05* | 0.92 | 1.04 | 0.66 | 0.95 | 1.01 | 0.97 | 0.99 |
| LC | A3a | PC (36:3) | 1.38 | 0.29 | 0.92 | 0.91 | 0.47 | 0.90 | 1.07 | 0.84 | 0.99 |
| LC | A3a | ACar 5:0 | 1.43 | 0.24 | 0.92 | 1.07 | 0.65 | 0.95 | 1.54 | 0.26 | 0.69 |
| LC | A3a | Glycodeoxycholic acid | 1.02 | 0.88 | 0.98 | 0.92 | 0.15 | 0.83 | 0.86 | 0.26 | 0.69 |
| LC | A3a | PC (40:6) | 0.67 | 0.13 | 0.92 | 1.01 | 0.91 | 1.00 | 0.84 | 0.53 | 0.85 |
| LC | A3a | Tetradecanoylcarnitine (C14:0) | 1.47 | 0.28 | 0.92 | 0.81 | 0.26 | 0.83 | 2.51 | < 0.05* | 0.43 |
| GC | A3a | 3-Hydroxy-dithiobutanoic acid | 1.60 | 0.23 | 0.92 | 1.38 | 0.27 | 0.83 | 0.46 | 0.20 | 0.69 |
| GC | A3a | Tetradecanoic acid | 0.89 | 0.60 | 0.93 | 0.85 | 0.07 | 0.67 | 0.86 | 0.45 | 0.82 |
| GC | A3a | (9E,12E)-octadeca-9,12-dienoic acid | 0.71 | 0.51 | 0.93 | 0.78 | 0.21 | 0.83 | 0.81 | 0.66 | 0.97 |
| GC | A3a | Indole | 1.38 | 0.56 | 0.93 | 1.98 | < 0.05* | 0.26 | 1.02 | 0.97 | 0.99 |
| GC | A3a | Skatole | 0.82 | 0.55 | 0.93 | 0.83 | 0.22 | 0.83 | 1.33 | 0.44 | 0.82 |
| GC | A3a | (5S)-5-ethyl-5-[(1R)-1-hydroxyethyl]-2-oxolanone | 1.27 | 0.66 | 0.93 | 0.79 | 0.35 | 0.85 | 0.84 | 0.77 | 0.98 |
| GC | A3a | 8-Hydroxy-4Z,9-decadienoic acid | 1.63 | 0.18 | 0.92 | 1.02 | 0.88 | 1.00 | 1.03 | 0.94 | 0.99 |
| GC | A3a | 2-Hydroxy-4-phenyl-butyric acid ethyl ester | 0.24 | 0.05 | 0.92 | 0.81 | 0.38 | 0.85 | 4.40 | < 0.05* | 0.38 |
| GC | A3a | 4-oxo-6,7,8,9-tetrahydro-pyrido[1,2-a]pyrimidine | 1.64 | 0.17 | 0.92 | 1.02 | 0.91 | 1.00 | 1.28 | 0.46 | 0.83 |
| GC | A3a | 3-[(2S)-2-pyrrolidin-1-iumyl]propanoate | 3.40 | 0.11 | 0.92 | 1.24 | 0.39 | 0.85 | 1.09 | 0.90 | 0.99 |
| GC | A3a | .beta.-apo-8'-carotenal | 0.95 | 0.74 | 0.97 | 1.04 | 0.66 | 0.95 | 0.98 | 0.86 | 0.99 |
| GC | A3a | 3-methyl-2-pentyl-1H-pyrrole | 0.63 | 0.21 | 0.92 | 0.88 | 0.35 | 0.85 | 1.07 | 0.84 | 0.99 |
| GC | A3a | methyl 2-(1,3-dimethyl-2-oxoimidazolidin-4-yl)acetate | 0.78 | 0.29 | 0.92 | 1.07 | 0.56 | 0.92 | 0.94 | 0.77 | 0.98 |
| GC | A3a | 2-phenylacetic acid | 0.49 | 0.16 | 0.92 | 1.00 | 1.00 | 1.00 | 0.44 | 0.10 | 0.54 |
| GC | A3a | 6-[6'-Hydroxy-1'-hexyn-1'-yl]-1H-purine | 42.01 | 0.09 | 0.92 | 0.73 | 0.74 | 0.98 | 0.15 | 0.31 | 0.72 |
| GC | A3a | 5,6-Dioxoheptanoic acid | 1.31 | 0.53 | 0.93 | 1.44 | 0.11 | 0.83 | 0.94 | 0.89 | 0.99 |
| GC | A3a | 2-amino-7-methyl-3H-purin-6-one | 0.16 | 0.15 | 0.92 | 1.46 | 0.53 | 0.92 | 0.40 | 0.51 | 0.85 |
| GC | A3a | Decanoic acid | 1.13 | 0.71 | 0.96 | 1.01 | 0.91 | 1.00 | 0.74 | 0.32 | 0.72 |
| GC | A3a | (S)-2-Ethyl-5-oxotetrahydrofuran-2-carboxylic acid | 1.62 | 0.27 | 0.92 | 0.82 | 0.51 | 0.91 | 0.65 | 0.54 | 0.86 |
| GC | A3a | 2-Ethylhexanoic acid | 1.76 | 0.29 | 0.92 | 1.10 | 0.68 | 0.97 | 0.53 | 0.23 | 0.69 |
| GC | A3a | (2S)-2-Amino-3-(1H-2-indolyl)propionic acid (L-isotrytophan) | 0.85 | 0.62 | 0.93 | 0.89 | 0.41 | 0.85 | 1.13 | 0.68 | 0.97 |
| GC | A3a | 12-Oxooctadec-9-enoic acid | 1.16 | 0.22 | 0.92 | 1.08 | 0.25 | 0.83 | 1.10 | 0.53 | 0.85 |
| GC | A3a | (E)-2-phenyl-3-(2-pyridinyl)-2-propenoic acid | 1.17 | 0.34 | 0.92 | 1.10 | 0.23 | 0.83 | 1.00 | 0.99 | 0.99 |
| GC | A3a | 1,5-Anhydro-D-fructose | 0.87 | 0.52 | 0.93 | 0.98 | 0.81 | 1.00 | 1.32 | 0.06 | 0.47 |
| GC | A3a | (E)-6-hydroxyhept-4-enoic acid | 0.96 | 0.80 | 0.98 | 1.05 | 0.55 | 0.92 | 1.05 | 0.80 | 0.99 |
| GC | A3a | 1,5-Anhydro-2-deoxy-2-(methoximino)-D-arabino-hexitol | 1.06 | 0.46 | 0.93 | 0.97 | 0.37 | 0.85 | 0.99 | 0.87 | 0.99 |
| GC | A3a | 2-(Aminomethyl)-5,5-dimethyl-1,3-thiazolidine-4-carboxylic acid | 1.04 | 0.71 | 0.96 | 0.99 | 0.82 | 1.00 | 1.08 | 0.41 | 0.79 |
| GC | A3a | 11,15'-Dihydrooxepin-.beta.,.beta.-carotene | 1.06 | 0.54 | 0.93 | 1.00 | 0.94 | 1.00 | 1.02 | 0.76 | 0.98 |
| GC | A3a | (S)-5-Methyl-2-(dimethylamino)-1H-imidazol-4(5H)-one | 0.85 | < 0.05* | 0.92 | 1.02 | 0.64 | 0.95 | 1.08 | 0.33 | 0.72 |
| GC | A3a | 4,5-Secocholest-8-en-3-one, 5-ethylidene-14-methyl- | 1.19 | 0.37 | 0.92 | 0.96 | 0.65 | 0.95 | 1.76 | < 0.05* | 0.36 |
| LC | A3b | PC (36:2) | 1.00 | 1.00 | 1.00 | 0.92 | 0.65 | 0.94 | 1.55 | 0.26 | 0.36 |
| LC | A3b | Cotinine | 3.29 | < 0.001** | < 0.001 | 1.01 | 0.94 | 0.94 | 10.25 | < 0.001** | < 0.001 |
| LC | A3b | LYSOPC (20:3) | 0.38 | 0.07 | 0.16 | 1.09 | 0.67 | 0.94 | 0.09 | < 0.05** | 0.05 |
| LC | A3b | Cotinine secondary peak | 2.10 | < 0.05** | 0.02 | 0.90 | 0.48 | 0.94 | 7.40 | < 0.001** | < 0.001 |
| LC | A3b | LYSOPC (20:4) | 0.64 | 0.14 | 0.20 | 1.24 | 0.13 | 0.90 | 1.93 | < 0.05** | 0.08 |
| LC | A3b | 2-Hydroxyphenylacetic acid | 2.20 | 0.14 | 0.20 | 1.21 | 0.45 | 0.94 | 1.00 | 1.00 | 1.00 |
| GC | A3b | Guadiscoline | 1.31 | 0.47 | 0.54 | 0.99 | 0.94 | 0.94 | 0.90 | 0.75 | 0.87 |

**Table S7a:** results Mummichog pathway analysis in pooled analysis. FDR = false discovery rate.

| **Pathway** | **Nr. members pathway** | **Matches** | **Matches in enriched set** | **Expected** | **p-value** | **FDR** |
|---|---|---|---|---|---|---|
| Fatty acid activation | 74 | 12 | 7 | 1.610 | 7.61e-05 | 0.00106 |
| D4&E4-neuroprostanes formation | 37 | 10 | 9 | 0.403 | 3.76e-02 | 0.25800 |
| Fatty Acid Metabolism | 63 | 6 | 4 | 1.080 | 5.53e-02 | 0.25800 |
| Vitamin A (retinol) metabolism | 67 | 16 | 7 | 0.806 | 1.50e-01 | 0.52600 |
| Purine metabolism | 80 | 7 | 2 | 0.941 | 1.95e-01 | 0.54700 |
| Arachidonic acid metabolism | 95 | 32 | 11 | 1.340 | 3.36e-01 | 0.73400 |
| Methionine and cysteine metabolism | 94 | 5 | 1 | 0.672 | 4.71e-01 | 0.73400 |
| Lysine metabolism | 52 | 5 | 1 | 0.672 | 4.71e-01 | 0.73400 |
| Linoleate metabolism | 46 | 17 | 4 | 1.750 | 4.72e-01 | 0.73400 |
| Urea cycle/amino group metabolism | 85 | 5 | 1 | 0.806 | 5.35e-01 | 0.74900 |
| Putative anti-Inflammatory metabolites formation from EPA | 27 | 12 | 1 | 1.080 | 6.42e-01 | 0.78100 |
| Tyrosine metabolism | 160 | 20 | 2 | 2.550 | 6.90e-01 | 0.78100 |
| Prostaglandin formation from arachidonate | 78 | 15 | 1 | 1.340 | 7.25e-01 | 0.78100 |
| Vitamin E metabolism | 54 | 13 | 1 | 2.280 | 8.93e-01 | 0.89300 |

**Table S7b:**
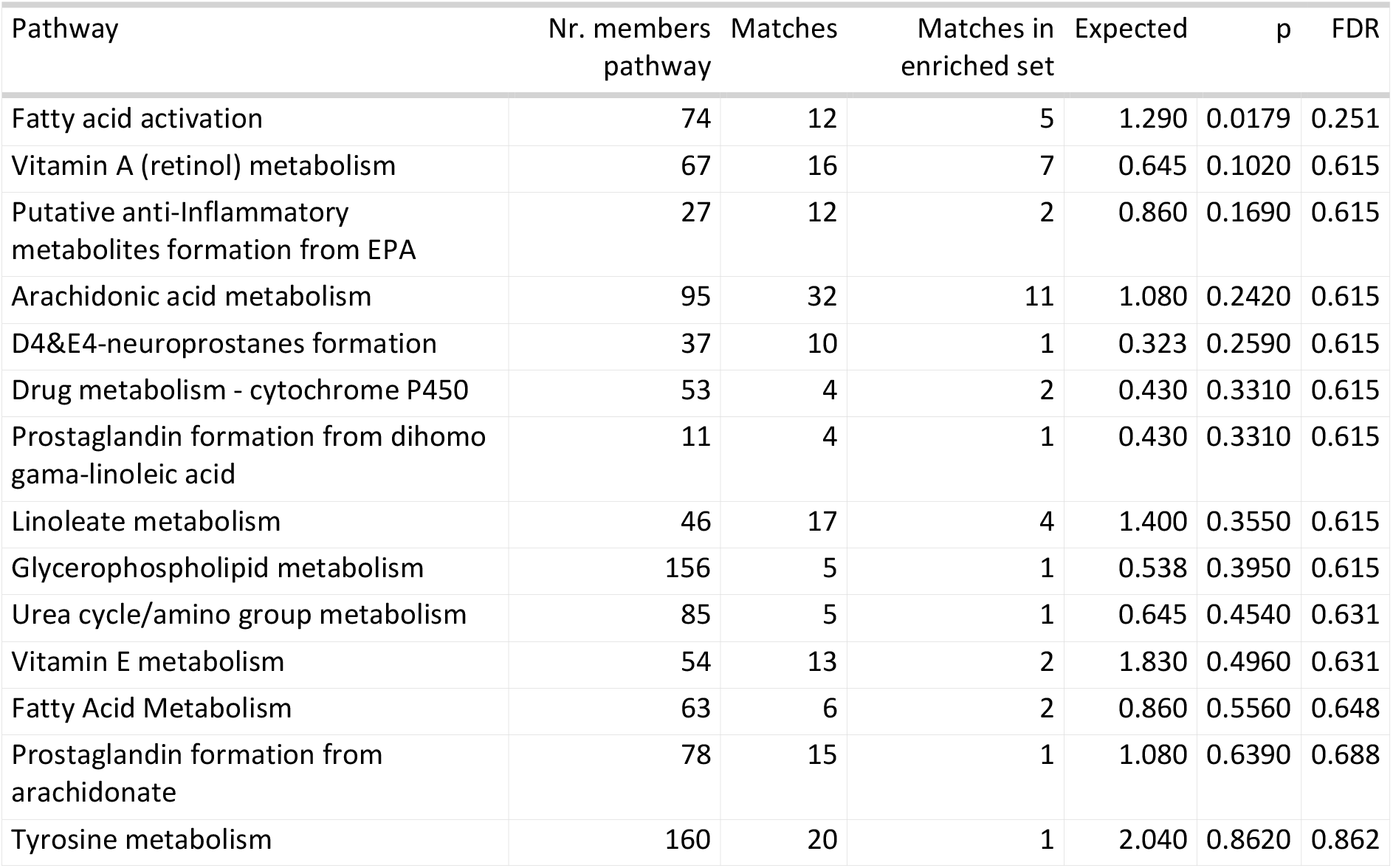
results Mummichog pathway analysis in never-smokers analysis. FDR = false discovery rate.

**Table S7c:** fatty acid activation putative annotation results in pooled analysis and result of follow-up. RT = retention time; mz = mass-to-charge ratio (m/z).

| mz | RT | Putative annotation Mummichog | Putative annotation Mummichog inferred name | Putative matched form | In enriched set | Empirical compound Mummichog | Notes from follow-up |
|---|---|---|---|---|---|---|---|
| 326.2196 | 7.4256682 | arachd | Arachidonic acid | M+Na [1+] | TRUE | EC0004 |  |
| 326.2222 | 7.236089 | arachd | Arachidonic acid | M+Na [1+] | TRUE | EC0004 | This feature was annotated as Arachidonic acid (level 1) in follow-up |
| 350.2203 | 7.3548465 | crvnc | Cervonic acid | M+Na [1+] | TRUE | EC000129 | This feature was annotated as Docosahexaenoic acid + Na (level 1) in follow-up (synonym of Cervonic acid) |
| 350.2219 | 7.2134323 | crvnc | Cervonic acid | M+Na [1+] | TRUE | EC000129 |  |
| 350.222 | 7.2105017 | crvnc | Cervonic acid | M+Na [1+] | TRUE | EC000129 |  |
| 326.2196 | 7.4256682 | eicostet | Eicosatetranoic acid | M+Na [1+] | TRUE | EC0004 | Eicosatetranoic acid synonym of Arachidonic acid |
| 326.2222 | 7.236089 | eicostet | Eicosatetranoic acid | M+Na [1+] | TRUE | EC0004 | Eicosatetranoic acid synonym of Arachidonic acid |
| 254.2241 | 6.6171193 | hdcea | Hexadecenoate | M+H [1+] | TRUE | EC000205 |  |
| 254.2242 | 6.7480006 | hdcea | Hexadecenoate | M+H [1+] | TRUE | EC000205 |  |
| 254.2244 | 6.803694 | hdcea | Hexadecenoate | M+H [1+] | TRUE | EC000205 |  |
| 254.2248 | 6.8706675 | hdcea | Hexadecenoate | M+H [1+] | TRUE | EC000205 |  |
| 254.2253 | 7.1457825 | hdcea | Hexadecenoate | M+H [1+] | TRUE | EC000206 |  |
| 276.2053 | 7.1141734 | hdcea | Hexadecenoate | M+Na [1+] | TRUE | EC000206 | The correlation with the M+H does not seem too high to justify M+Na |
| 280.2389 | 7.2579775 | lneldc | Linoelaidic acid (all trans C18:2) | M+H [1+] | TRUE | EC000207 | This feature was annotated as Linoleic acid (level 1) in follow-up |
| 280.2398 | 6.9345326 | lneldc | Linoelaidic acid (all trans C18:2) | M+H [1+] | TRUE | EC000208 |  |
| 280.2421 | 7.1483483 | lneldc | Linoelaidic acid (all trans C18:2) | M+H [1+] | TRUE | EC000207 |  |
| 302.2218 | 7.251964 | lneldc | Linoelaidic acid (all trans C18:2) | M+Na [1+] | TRUE | EC000207 | The correlation with the M+H of Linoelaidic acid does not justify M+H-M+Na relationship |
| 356.2685 | 6.4492984 | tethex3 | Tetracosahexaenoic acid, n-3 | M+H [1+] | TRUE | EC000157 |  |
| 356.2698 | 6.7619915 | tethex3 | Tetracosahexaenoic acid, n-4 | M+H [1+] | TRUE | EC000158 |  |
| 302.2218 | 7.251964 | tmndnc | Timnodonic acid C20:5, n-3 | M+H [1+] | TRUE | EC000150 | The correlation with the M+Na of Timnodonic acid does not justify M+H-M+Na relationship |
| 324.2039 | 7.2596874 | tmndnc | Timnodonic acid C20:5, n-4 | M+Na [1+] | TRUE | EC000150 | The 15 ppm between masses 324.2039 and 324.2088 is large to justify the two corresponding to the same formula |
| 324.2088 | 7.1051755 | tmndnc | Timnodonic acid C20:5, n-5 | M+Na [1+] | TRUE | EC000150 | The 15 ppm between masses 324.2039 and 324.2088 is large to justify the two corresponding to the same formula |
| 330.2582 | 7.2962317 | clpnd | Clupanodonic acid | M+H [1+] | FALSE | EC000203 |  |
| 330.2582 | 7.2962317 | dcsptn1 | Docosa-4,7,10,13,16-pentaenoic acid (n-6) | M+H [1+] | FALSE | EC000203 | Docosa-4,7,10,13,16-pentaenoic acid (n-6) synonym of Clupanodonic acid |
| 282.2561 | 7.062082 | ocdcea | Octadecenoate (n-C18:1) | M+H [1+] | FALSE | EC000209 | Octadecenoate (n-C18:1) synonym of Vaccenic acid |
| 282.2565 | 7.1216617 | ocdcea | Octadecenoate (n-C18:1) | M+H [1+] | FALSE | EC000209 | Octadecenoate (n-C18:1) synonym of Vaccenic acid |
| 282.2575 | 7.014573 | ocdcea | Octadecenoate (n-C18:1) | M+H [1+] | FALSE | EC000209 | Octadecenoate (n-C18:1) synonym of Vaccenic acid |
| 298.1882 | 7.1691628 | strdnc | Stearidonic acid C18:4, n-3 | M+Na [1+] | FALSE | EC000211 |  |
| 282.2561 | 7.062082 | vacc | Vaccenic acid | M+H [1+] | FALSE | EC000209 |  |
| 282.2565 | 7.1216617 | vacc | Vaccenic acid | M+H [1+] | FALSE | EC000209 |  |
| 282.2575 | 7.014573 | vacc | Vaccenic acid | M+H [1+] | FALSE | EC000209 |  |

**Table S8:** all results that were significant in an analysis, and their corresponding annotation level. Set A describes the annotated metabolites, Set B indicates the initially unidentified features/clusters. 1, 2, 3 describes the exogenous, endogenous, both endogenous/exogenously derived metabolites respectively. The last letter indicates if the feature was analyzed as continuous variable (lower case ‘a’) or as binary variable of detected, not detected (lower case ‘b’). GC = gas-chromatography; LC = liquid-chromatography; ICC = intra-class correlation coefficient; RT = retention time.

| Platform | Set | monoisotopic mass | RT | Identity | FDR significant in which analysis? | Annotation level |
|---|---|---|---|---|---|---|
| GC | A1a |  |  | phenyl N-methylcarbamate | After ICC filter at FDR < 0.1 | Level 2 |
| GC | A1a |  |  | 2,2,4,7-tetramethyl-3H-1-benzofuran | Pooled at FDR < 0.2; after ICC filter at FDR < 0.1 | Level 2 |
| GC | A1a |  |  | 7-isopropyl-3,4-dihydro-2H-cyclohepta[b]pyrazine | Doetinchem Cohort Study at FDR < 0.1 | Level 2 |
| GC | A1a |  |  | 4-Chloro-2-(6-chlorohex-1-yn-1-yl)pyridine | Doetinchem Cohort Study at FDR < 0.2 | Level 2 |
| GC | A1a |  |  | 2-[(3R)-3,7-dimethyloct-6-enylidene]malononitrile | Doetinchem Cohort Study at FDR < 0.2 | Level 2 |
| GC | A1a |  |  | 2-methyl-5-phenyl-1H-pyrrole | Doetinchem Cohort Study at FDR < 0.1 | Level 2 |
| GC | A1a |  |  | 1,3-Benzenedicarbonitrile | Pooled at FDR < 0.2; Doetinchem Cohort Study at FDR < 0.1; after ICC filter at FDR < 0.1 | Level 1 |
| GC | A2b |  |  | 4-Thiazolidinecarboxylic acid | Doetinchem Cohort Study at FDR < 0.2 | Level 2 |
| GC | Bb |  |  | C5155 | After ICC filter at FDR < 0.1 | - |
| GC | Bb |  |  | C5609 | Ex-smokers analysis at FDR < 0.2 | - |
| LC | Bb | 85.9463 | 0.90 | - | Ex-smokers analysis at FDR < 0.2 | - |
| LC | A2a | 129.0791 | 0.74 | - | Pooled analysis at FDR < 0.2 | - |
| LC |  | 145.0839 | 0.83 | - | SAPALDIA at FDR < 0.2 | - |
| LC | A1a | 162.1132 | 0.92 | Nicotine | Pooled at FDR < 0.1; KORA at FDR < 0.1; Doetinchem Cohort Study at FDR < 0.1; current-smokers at FDR < 0.1; after ICC filter at FDR < 0.1 | Level 1 |
| LC | A3b | 176.0942 | 0.98 | Cotinine secondary peak | Pooled at FDR < 0.1; KORA at FDR < 0.1; Doetinchem Cohort Study at FDR < 0.1; after ICC filter at FDR < 0.1 | - |
| LC | A3b | 176.0942 | 1.56 | Cotinine | Pooled at FDR < 0.1; KORA at FDR < 0.1; Doetinchem Cohort Study at FDR < 0.1; | Level 1 |
| LC | Bb | 192.0892 | 0.92 | - | Pooled at FDR < 0.1; KORA at FDR < 0.1; Doetinchem Cohort Study at FDR < 0.1; | - |
| LC | A3a | 231.1470 | 2.13 | ACar 4:0 | Never-smokers analysis at FDR < 0.1 | Level 1 |
| LC | A3a | 313.2245 | 5.08 | ACar 10:1 | SAPALDIA at FDR < 0.2 | Level 2 |
| LC | A2a | 360.1935 | 5.00 | Cortisone | KORA at FDR < 0.2 | Level 1 |
| LC | A2a | 471.2946 | 6.40 | Glycochenodeoxycholic acid | KORA at FDR < 0.2 | Level 1 |
| LC | A3b | 543.3313 | 6.83 | LysoPC (20:4) | Doetinchem Cohort Study at FDR < 0.1 | Level 2 |
| LC | A3b | 545.3458 | 6.94 | LysoPC (20:3) | KORA at FDR < 0.2; Doetinchem Cohort Study at FDR < 0.1 | Level 2 |

## Supporting Information I

Chromatograms and isotope patterns from the study’s quality-control sample, and pure chemical standards of cotinine and serotonin. *Bottom*: Chromatograms and isotope patterns from representative study samples containing or lacking the cotinine peak (bottom).

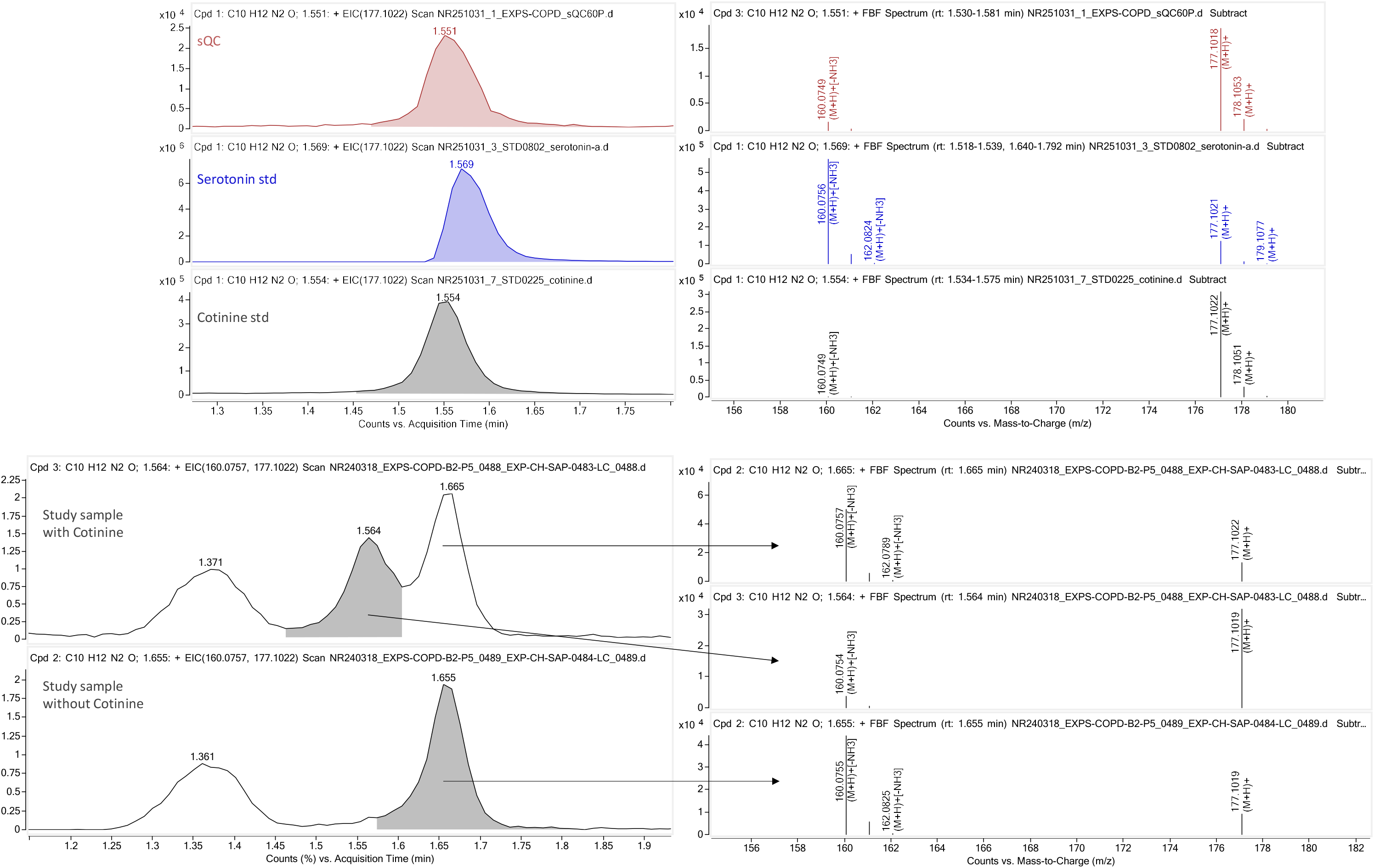

